# Predicting nutrition and environmental factors associated with female reproductive disorders using a knowledge graph and random forests

**DOI:** 10.1101/2023.07.14.23292679

**Authors:** Lauren E Chan, Elena Casiraghi, Tim Putman, Justin Reese, Quaker E. Harmon, Kevin Schaper, Harshad Hedge, Giorgio Valentini, Charles Schmitt, Alison Motsinger-Reif, Janet E Hall, Christopher J Mungall, Peter N Robinson, Melissa A Haendel

## Abstract

**Objective:** Female reproductive disorders (FRDs) are common health conditions that may present with significant symptoms. Diet and environment are potential areas for FRD interventions. We utilized a knowledge graph (KG) method to predict factors associated with common FRDs (e.g., endometriosis, ovarian cyst, and uterine fibroids).

**Materials and Methods:** We harmonized survey data from the Personalized Environment and Genes Study on internal and external environmental exposures and health conditions with biomedical ontology content. We merged the harmonized data and ontologies with supplemental nutrient and agricultural chemical data to create a KG. We analyzed the KG by embedding edges and applying a random forest for edge prediction to identify variables potentially associated with FRDs. We also conducted logistic regression analysis for comparison.

**Results:** Across 9765 PEGS respondents, the KG analysis resulted in 8535 significant predicted links between FRDs and chemicals, phenotypes, and diseases. Amongst these links, 32 were exact matches when compared with the logistic regression results, including comorbidities, medications, foods, and occupational exposures.

**Discussion:** Mechanistic underpinnings of predicted links documented in the literature may support some of our findings. Our KG methods are useful for predicting possible associations in large, survey-based datasets with added information on directionality and magnitude of effect from logistic regression. These results should not be construed as causal, but can support hypothesis generation.

**Conclusion:** This investigation enabled the generation of hypotheses on a variety of potential links between FRDs and exposures. Future investigations should prospectively evaluate the variables hypothesized to impact FRDs.

## INTRODUCTION

Female reproductive disorders (FRDs) such as endometriosis, uterine fibroids, and ovarian cysts significantly affect physical and emotional health, disability, and fertility for women and those assigned female at birth.[1] FRDs fall into a category of conditions that are often misdiagnosed and have prolonged diagnostic timeframes and limited therapeutic options.[2,3] Prevalence of common FRDs such as endometriosis is often underestimated given the clinical difficulty of identifying the condition without invasive laparoscopic surgery and the often years-long lag between symptom onset and diagnosis.[2,4] Due to their widespread prevalence and substantial impact on daily life, ways to more easily identify FRDs as well as viable therapeutic approaches for FRDs are highly sought after.[5–7] Diet and environment have been proposed as potential intervention opportunities for FRDs,[8,9] but standard clinical recommendations on diet and exposures are limited. Focusing on modifiable features such as diet, lifestyle factors, and environmental exposures may offer new options for individuals and care providers to manage these common conditions and improve outcomes. We present an innovative approach for assessing survey-based data to predict links between nutrition, environmental exposures, comorbidity, and medication and three common FRDs, namely endometriosis, uterine fibroids, and ovarian cysts.

### Common FRDs

Endometriosis is the extrauterine growth of endometrial tissue (also called lesions) with hallmark symptoms that include pelvic pain, dysuria, dysmenorrhea, and sub- or infertility.[10] This FRD is estimated to occur in 10% of women.[11] Delays in diagnosis are common with endometriosis, and many individuals wait years for a conclusive diagnosis.[2,4] Accordingly, estimates of prevalence vary widely and are likely inaccurate. An estimated 35-50% of individuals diagnosed with endometriosis experience pain and/or infertility,[5] but approximately 20-25% of individuals with endometriosis do not experience pelvic pain.[5,12,13] Because symptoms can be inconsistent, clinical diagnosis is difficult. Endometriosis is often diagnosed during treatment for fertility issues.[14,15] Endometriosis can present similarly to other gynecological disorders including primary dysmenorrhea, pelvic inflammatory disease, and pelvic adhesions presenting as chronic pelvic pain, painful menses, tubal pregnancies, and infertility.[2,3] Due to its inconsistent presentation, surgical visualization is needed to definitively diagnose endometriosis, which is a barrier to diagnosis and treatment.[2]

Uterine fibroids, also called leiomyomas, are common benign tumors estimated to be present in 70-80% of women by the age of menopause,[16] and approximately 20-25% of those individuals present with clinical symptoms.[17] The fibroids are composed of smooth muscle cells and fibrous extracellular matrix that is overproduced and creates tumors within the myometrium.[18] Many women with fibroids are not clinically diagnosed. Some have no symptoms, and some live with significantly burdensome symptoms without a clinical diagnosis. The high prevalence of undiagnosed fibroids means that prevalence may be underestimated when determined using clinical records. Common fibroid symptoms include heavy menses, pelvic pain, anemia, urinary incontinence, and infertility.[18–20] With symptomatic fibroids, pregnancy complications (placenta previa, intrauterine growth restriction, increased need for cesarean section) can be more common.[21] Diagnosis of fibroids is usually accomplished with a variety of imaging techniques, including transvaginal ultrasound, hysterosalpingography, saline infusion sonography, hysteroscopy, and MRI.[21–23]

Ovarian cysts affect approximately one in 25 women.[7] There are multiple types of ovarian cysts, but functional cysts are the most prevalent. Functional cysts occur when a follicle forms in the ovary but no ovulation ensues and the follicle does not rupture, creating a cyst.[24] The most frequently reported symptoms of ovarian cysts are pelvic pain, abdominal pressure, bloating, and infertility although asymptomatic ovarian cysts can occur.[25,26] Asymptomatic ovarian cysts can be left untreated and may not require intervention, with some cysts disappearing naturally. However, cysts affecting fertility, pelvic anatomy, or quality of life in a significant way can be surgically removed.[27]

### Ontologies

Ontologies are a methodology for standardizing terminology in a computable fashion to support the creation of logical axioms between related terms. Prominent ontologies in the biomedical sciences include the Gene Ontology[28] and the Human Phenotype Ontology,[29] with many others related to foods, chemicals, and diseases.[30–32] Knowledge graphs (KGs) are a method for representing knowledge such as ontology content and instance level data in a graph structure in which nodes and edges are explicitly connected via semantic relationships.[33] Because of their innate high dimensionality, data inquiries can be conducted using KGs. However, the dimensionality of KGs can be reduced through embedding so they can support other analytic methodologies.[34] In our investigation, we aligned heterogeneous data regarding health, environment, and internal exposures to ontology content for ingestion into a KG, which was subsequently embedded and analyzed using machine learning techniques.

## METHODS

### Data Sets

The primary data for this project came from the Personalized Environment and Genes Study (PEGS, formerly known as the Environmental Polymorphisms Registry) conducted by the National Institute of Environmental Health Sciences (NIEHS),[35,36] which includes data from three respondent surveys, the Health and Exposure (self-reported diseases and phenotypes), Internal Exposome (foods, medications, supplements, and ingested exposures), and External Exposome (environmental exposures) surveys. Survey respondents are adult (aged 18 years or more) residents of North Carolina recruited for voluntary participation through health providers or events such as health fairs. The data included in this investigation were collected between 2012 and 2020. This investigation was approved and deemed research with no human subjects (Category 4 exemption) by the Oregon State University (IRB-2021-1207).

Additional publicly available data were included in this investigation. Agricultural Chemical Usage Program (ACUP) data from the United States Department of Agriculture (USDA) on fungicides, pesticides, and other chemicals applied to agricultural crops during 2016-2020 was included for all relevant questions in the PEGS data sets (e.g., data on chemicals applied to carrots was included as PEGS inquires about consumption of carrots). ACUP data were not included if there was no related PEGS question, and not all PEGS questions about diet had related ACUP data (e.g., consumption of combination foods such as hamburgers or foods without crop components, such as meat). Nutrient data for Foundation Foods from the USDA Food Data Central (FDC) was included when available with references to the FoodOn ontology.[32] This allowed for direct mapping to the selected ontology alignment (e.g., a survey question on intake of cottage cheese mapped to FOODON:03303720; and ‘cottage cheese (lowfat)’ mapped to FDC ID: 328841 and FDC nutritional content for ‘Cheese, cottage, lowfat, 2% milkfat’).

### Knowledge Graph Data Preparation

Combined, the PEGS surveys comprise 1842 questions. We assessed the survey questions for ontology alignment based on existing ontology content and complexity of the survey question as well as the primary topic area. We focused on questions related to diseases, phenotypes, dietary exposures, and environmental exposures. We then aligned feasible survey questions of interest (n = 341, with 135 from the External Exposure Survey, 131 from the Internal Exposure Survey, and 75 from the Health and Exposure Survey) to ontology terminology. An ontology curator (author LC) manually reviewed the data to map the PEGS survey questions to the coordinating ontology content. Free-response components of the PEGS surveys and other data sets, including USDA ACUP data, were mapped to ontology terms using semi-automated curation with OntoRunNER,[37] followed by supplemental manual review by the curator. The ‘survey question label’ selected for free response questions was assigned the mapped ontology term value of the response due to the list aggregation used to process data via OntoRunNER. When necessary, we requested new ontology terms in efforts to support the mappings needed for this data alignment. Primary requests were made to the Food Ontology (FoodOn)[32] and the Environmental Conditions, Treatments, and Exposures Ontology (ECTO).[38]

### Creating a KG

We created the KG for this project with an extract, transform, load (ETL) pipeline constructed using the Knowledge Graph Hub project KG-template.[39] The KG-template offers a skeleton structure of data download, transformation, and merge scripts that we customized for this project. This pipeline was developed using Python (Version 3.9.10) and Koza,[40] a data transformation framework constructed by the Monarch Initiative. Transformations included the alignment of self-reported data for questions of interest with the ontology mappings generated manually or semi-automatically as described in Figure 1.

**Figure 1.**
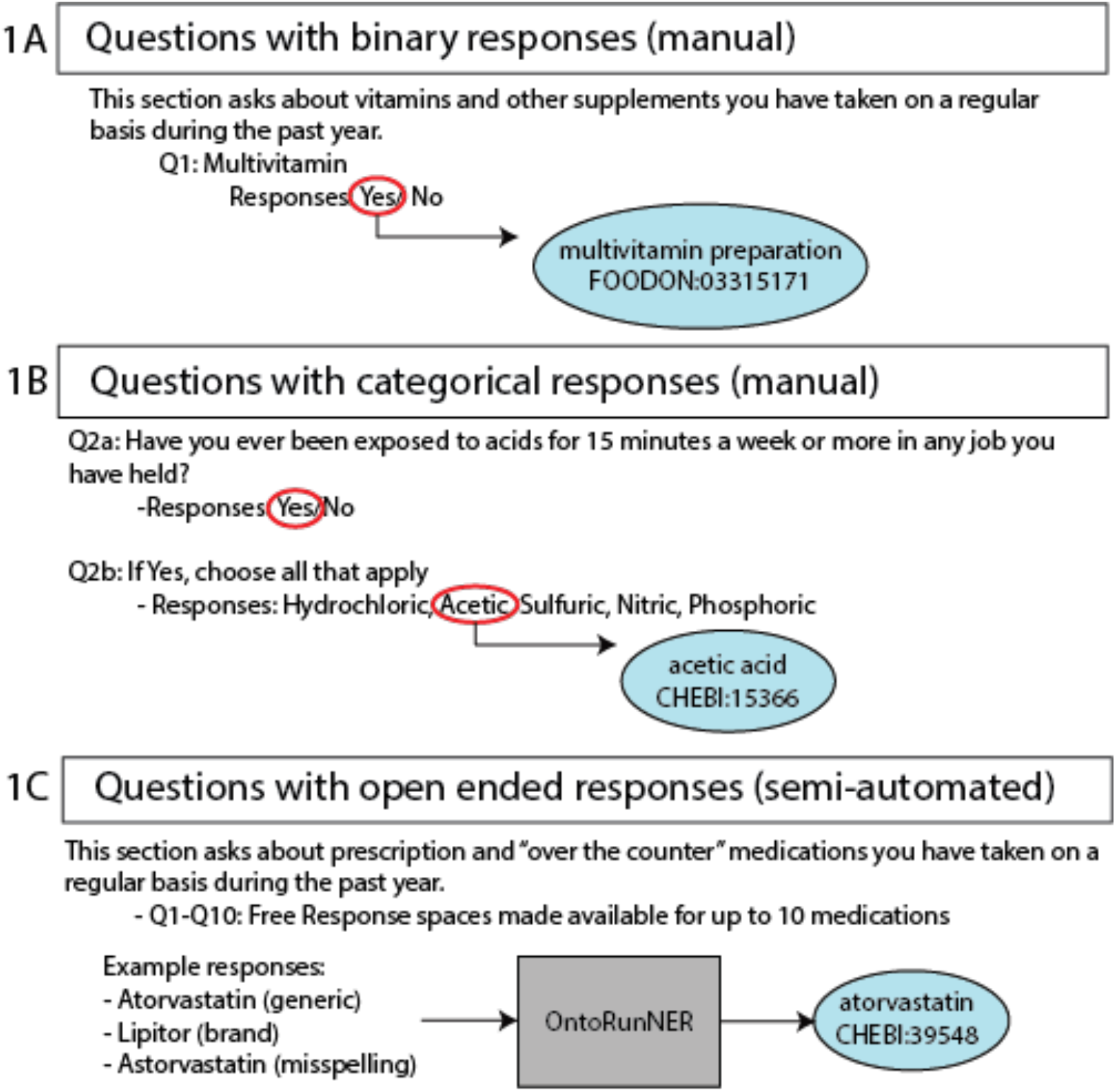
Translating survey questions to ontology content. In efforts to coordinate PEGS survey questions with ontology content, a combination of manual and semi-automated mappings were conducted. For questions with binary or categorical, finite responses, manual curation was used to align a single ontology term to the question (binary) or to each variable response option (categorical) (Fig 1A/B). For free response questions, the named entity recognition tool, OntoRuNER was used to create mappings to ontology terms for unique answer fields (Fig 1C). Ontology abbreviations: FOODON, Food Ontology. CHEBI, Chemical Entities of Biological Interest.

We conducted each data transformation (e.g., disease, phenotype, medication, food) with a unique script that asserted the correct “predicate” (e.g., the phenotype transform created assertions such as ‘Person:1234’ ‘has phenotype’ ‘uterine leiomyoma’). We followed this process for all PEGS data and all supplemental data on food, chemical usage, and nutrient content. Figure 2 provides an example of the full mapping and transformation process, in which reusable nodes were generated for a respondent’s unique ID and their survey responses. In turn, all questions answered by a respondent were mapped to the same respondent node using their ID. Similarly, all respondents who answered the same question were mapped to the same question response node. In addition to the transformed respondent data, the full content of relevant ontologies (Human Phenotype Ontology (HPO), Mondo Disease Ontology (Mondo), Medical Actions Ontology (MAxO), Gene Ontology (GO), Environment Ontology (ENVO), Chemical Entities of Biological Interest (ChEBI), ECTO, and FoodOn) was merged to create the KG. Within the KG structure, each ontology term or survey participant was considered a “node”, with all relationships between each node considered an “edge”.

**Figure 2.**
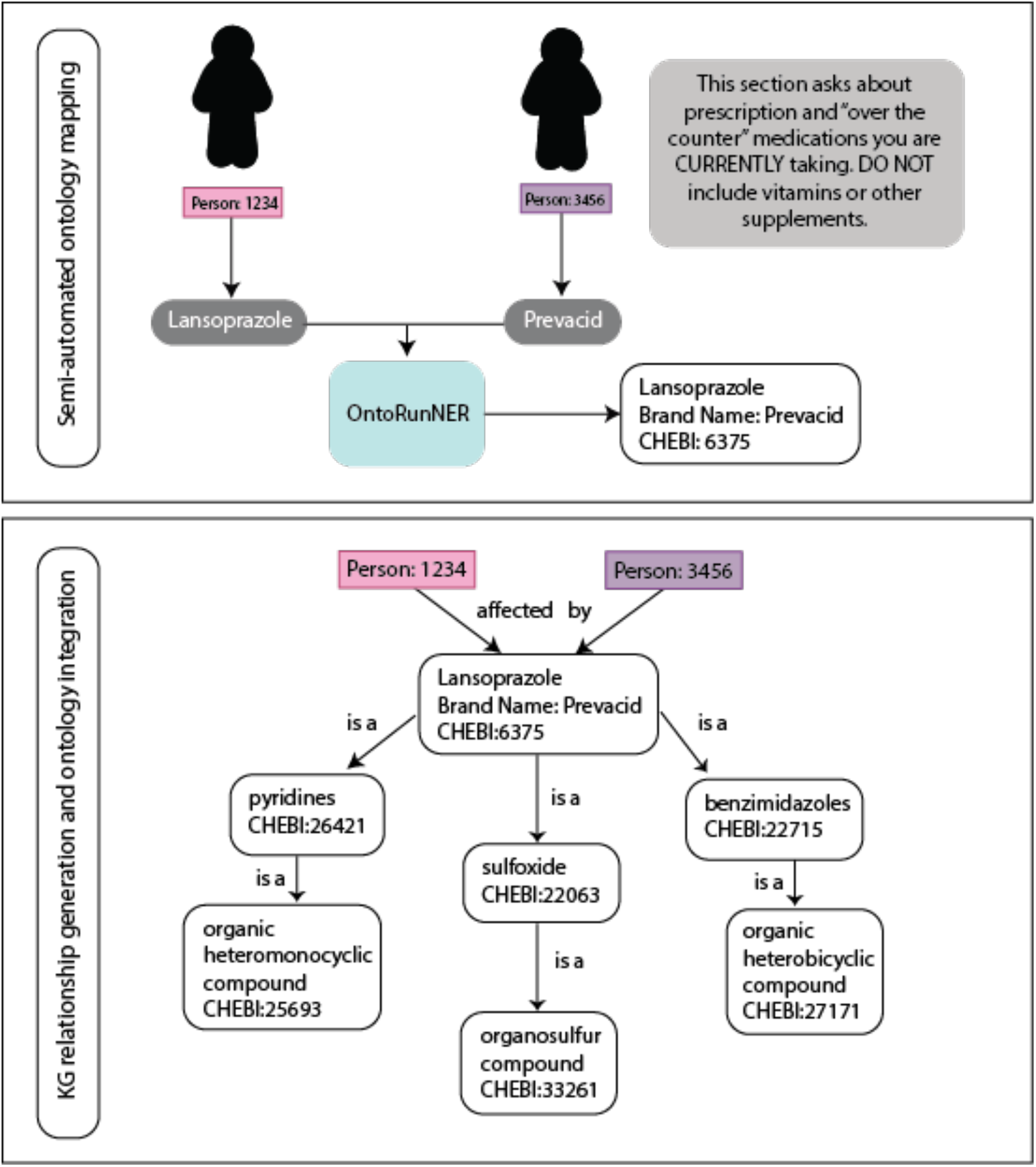
Coordinating respondent data to ontology content. Following completion of a survey question, the responses are used to generate an appropriate mapping of the response to an ontology term. During this process, nodes are established for each respondent as well as each positively answered survey question. Only unique nodes are generated, meaning only one node is created for each respondent and each survey question. Ontology terms have a corresponding hierarchy within the ontology that is also coordinated to the survey question and response. Unique “transformation” steps for each question type (e.g., medication, environmental exposure, disease) are used to then create a three part relationship including a subject, predicate, and object. As seen in this example question regarding medication usage, following a response of “lansoprazole”, Person:1234 had their response mapped using the semi-automated OntoRunNER tool to the appropriate ontology term and then the transformation step created a relationship result of “Person:1234 affected by Lansoprazole (CHEBI:6375)”. Given lansoprazole is contained within a hierarchy in the Chemical Entities of Biological Interest ontology, it is subsequently associated with a variety of terms in the taxonomy. As ontology content that is identified to have the same label, or shared synonyms will be mapped to the same node within a KG, a positive response of lansoprazole usage by the brand name Prevacid, similarly allows for the resulting relationship of “Person:3456 affected by lansoprazole”.

### Embedding the KG

As with many KGs, the KG for this project was a high-dimensional object with a large number of nodes and edges, making it less amenable to machine learning. Lower-dimension forms of a KG allow for improved generalization of knowledge, as the latent representation places dissimilar nodes farther away from one another and nodes with greater similarity closer to each other. To reduce the dimensionality of the KG in preparation for machine-learning techniques, we embedded the KG using Graph Representation leArning, Predictions and Evaluation (GRAPE)[41] and its embedding library. We used only the largest component of the KG, which eliminated data from 691 (7.1%) survey respondents due to insufficient data. The generated embedded representations included ontology terms, exposures, clinical variables, FRDs, and respondents. As such, the resulting representations embedded the topological relationships between the different types of entities populating the KG in a Euclidean space. Additional details can be found in the Supplemental Methods.

For the following machine learning methods, we generated two edge-embedding versions, a training embedding and a full data embedding. The training embedding included a ‘Training’ portion comprising 70% of the graph and a ‘Test’ portion comprising the remaining 30%. We created the test portion by selecting and holding out edges that, when removed from the full embedding, did not create a new component and thus kept the primary component of the graph intact. This avoided a biased estimation of the edge prediction results for the test set (see the GRAPE github repository for a full description of the method[42]). Edges in the training set were not specifically selected as “positive” responses (e.g., edges documenting an FRD-variable relationship), in efforts to train the model for edge prediction based on the entire topology of the graph. The full embedding included all available data. Figure 3 summarizes the analytical methods.

**Figure 3:**
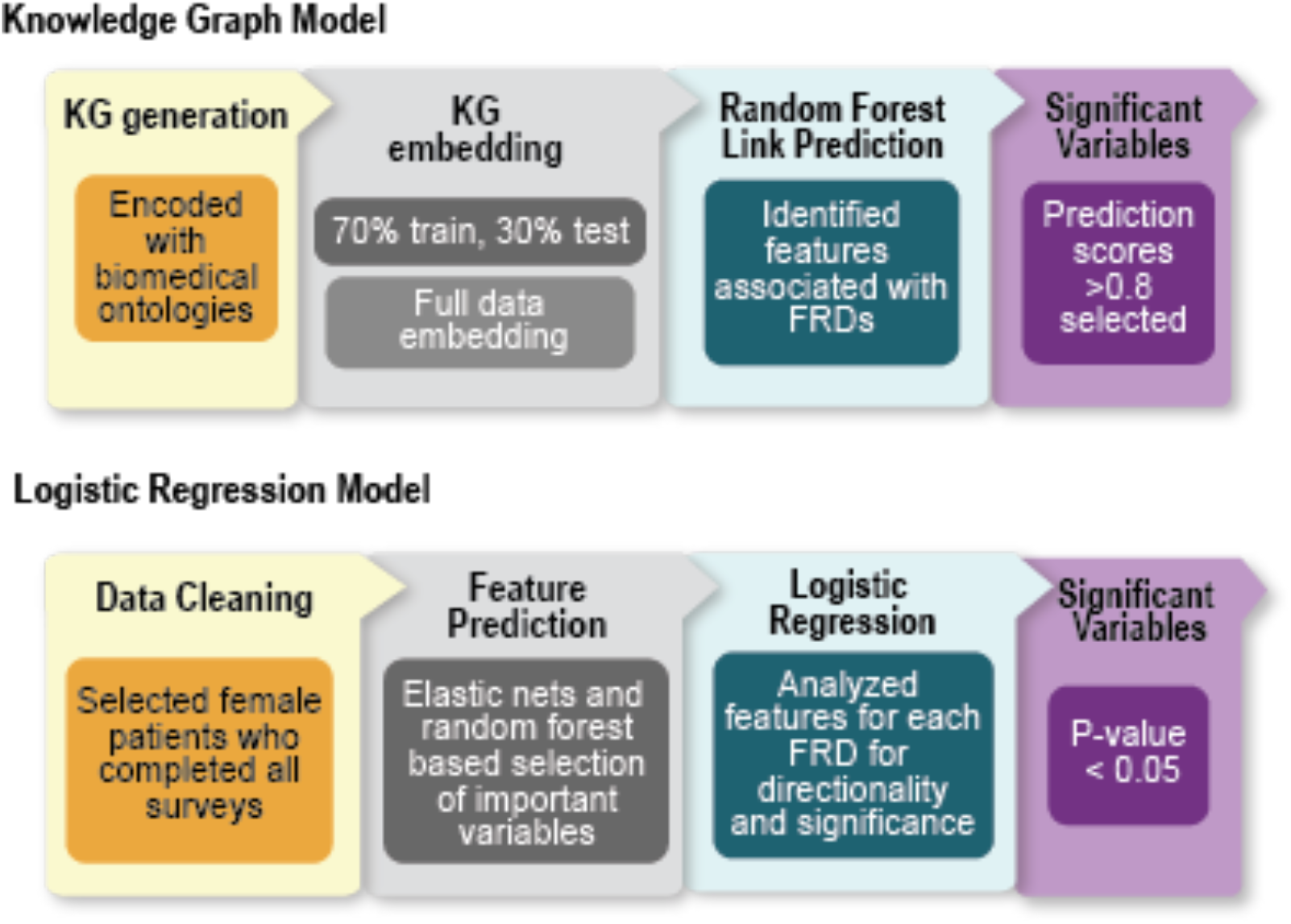
Computational methods overview. Two comparative analytical approaches were used to evaluate the Personal Environment and Genes Study survey data regarding internal and external exposures and personal health. The KG model included encoding all survey data with biomedical ontology content and creation of a KG structure, followed by embedding of the KG to create a low dimensional format for use in the random forest model to assess predicted links between FRDs of interest and exposures or health variables. The comparison logistic regression analysis system supported data interpretation by including data cleaning, application of elastic nets to improve regularization, a random forest based feature importance selection (explainability), and logistic regression to evaluate significance and directionality (interpretability) of the associations between exposures, health conditions, and FRDs. Ontology acronyms: Chemical Entities of Biological Interest (ChEBI), Environmental Conditions, Treatments, and Exposures Ontology (ECTO), Gene Ontology (GO), Human Phenotype Ontology (HP), Medical Actions Ontology (MAxO), Mondo Disease Ontology (Mondo).

### Machine Learning Analyses

Random forests (RF)[43] are machine-learning classifiers used for computing medical predictions due to their inherent explainability and interpretability and the availability of methods (although preliminary) to convert them into a checklist of rules.[44,45]

Our primary machine-learning task was generating link predictions between variables (e.g., food, nutrient, environmental exposure, disease, phenotype) and the FDRs of interest. We then trained an RF model (501 trees, 15 maximum depth) using the embeddings of the training data (with holdouts). The standard machine-learning performance metrics indicated the model was trained successfully and suitable for our analysis (area under the receiver operating characteristic (AUROC) = 0.915 for the ‘Test’ portion of the training data). To produce actionable results, we then retrained the model on the full dataset to obtain a set of predicted links between the FRDs and other variables. In the output, predicted links were represented by two node values—the “source” (independent variable) and “destination” (dependent variable) nodes of the link—and a “prediction” score indicating the strength of the predicted link between the two nodes. Utilizing the full graph embedding, we selected prediction outcomes from the model that included an FRD (e.g., endometriosis, ovarian cysts, uterine fibroids) as the “source” and the resulting “destination”. We retained pairs with a prediction score >0.8, resulting in a list of predicted variables for each FRD of interest.

### Logistic regression analysis

For additional comparison of our KG findings, we conducted a secondary analysis using elastic nets, RFs, and logistic regression models to provide feature explanations (in terms of feature importance in prediction) and interpretations (in terms of the directionality of risk scores associated with each feature). We conducted this analysis in R, version 4.2.2. We cleaned the primary PEGS data on health conditions and internal and external exposures to include female participants only. We then excluded participants who did not complete all three surveys to improve data quality, given the lower response rates to the Internal and External Exposure Surveys versus the Health and Exposure Survey. For the regression analysis, we utilized only survey questions that aligned with the KG analysis (see KG Data Preparation) to maintain consistency and enable comparison. We imputed missing data using the missForest algorithm, which has exhibited superior performance in previous work.[44,46]

To select the features with the strongest relationships with the FRDs of interest, we leveraged an explainable machine-learning technique,[47] to account for the class imbalance affecting the FRD datasets and to produce both importance scores and their directionality concerning the risk of disease. We developed a model that applied a first step of supervised feature selection on the training set and then selected features used to train an RF classifier. The model then computed permutation-based feature importance scores based on the RF classifier that were used to select the most important variables for FRD prediction. Features regarded as important by an RF are not characterized by directionality and magnitude, which is important for a medical context.[48] To assess these characteristics, we then trained logistic regression classifiers, whose learned odds ratios and *P* values indicate the significance and directionality of risk scores. We ran the model three times, each time utilizing a different FRD as the primary outcome. We adjusted the *P* values obtained in the logistic regression analyses for endometriosis, ovarian cysts, and uterine fibroids using Bonferroni correction to account for the family-wise false discovery rate (FDR).

Based on the KG and logistic regression model results, we identified the most influential features for each FRD. We compared both the KG and logistic regression outputs for exact matches for each FRD. Details of additional methods can be found in the Supplemental Methods.

## RESULTS

A total of 16039 surveys were completed (External Exposome = 3579, Internal Exposome = 3034, Health and Exposure = 9426) by 9765 unique individuals, including 2773 individuals who completed all three surveys. In the study population, there was reported prevalence of 7% for endometriosis, 15% for uterine fibroids, and 13% for ovarian cysts. Translation keys for all survey questions of interest and their coordinating ontology content, including OntoRunNER generated mappings, can be found in Supplementary Tables 1A-D (Supp Table 1D is also available on github[49]). The majority of survey respondents were female, with an average age between 49.9 and 54 years depending on the survey (Table 1). Further information such as race/ethnicity, pregnancy history, age at menarche, and health care access level were not available in this dataset.

**Table 1.** Demographics for PEGS survey data.

|  | Health and Exposure<br>(n = 9426) | External Exposures<br>(n = 3579) | Internal Exposures (n<br>= 3034) |
| --- | --- | --- | --- |
| BMI (mean,<br>% missingness) | 28.1 (0.02) | NA | NA |
| Gender (% female,<br>% missingness) | 67.1% (0) | NA | NA |
| Age (mean,<br>% missingness) | 49.9 years (0.01) | 54 years (0) | 53.9 years (0) |

The KG created for this project has 308.60K heterogeneous nodes and 696.68K edges in total. The graph contains 28.44K connected components (of which 28.41K are disconnected nodes), with the largest one containing 280.03K nodes and the smallest one containing a single node. Figure 4 shows the resulting full graph embedding after selecting for the largest component and excluding single note components.

**Figure 4.**
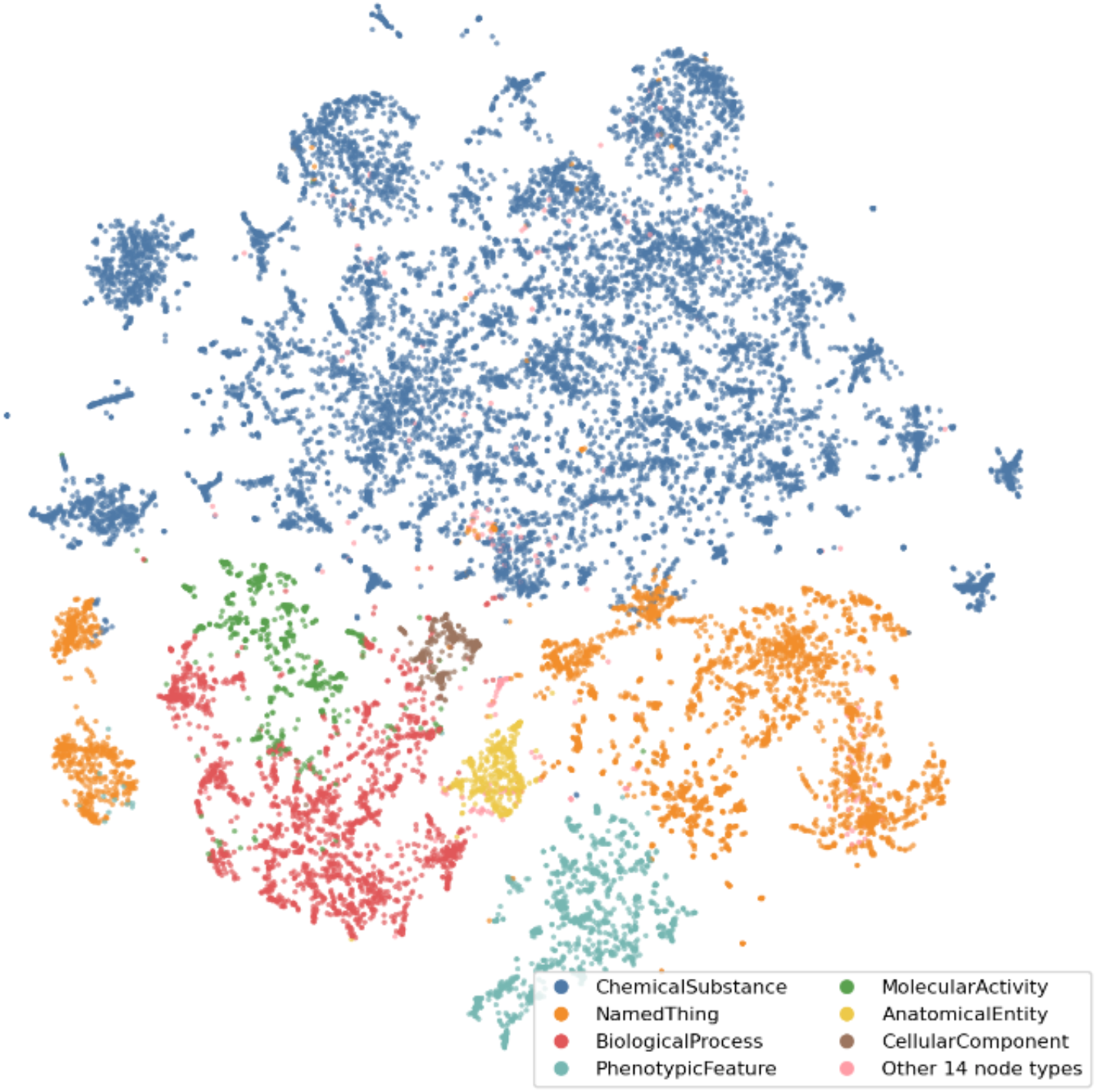
KG visualization. The KG embedding produced using the GRAPE DeepWalkSkipGram method, displays a variety of node types represented in the graph. Distinct separation between the colors within the embedding displays the clustering of similar node types.

We identified a list of significant variable features from both the KG and logistic regression analyses. All survey labels were coded for a “Yes” response to the question, indicating the presence of an exposure or condition. Table 2A-C shows the significant features (P < 0.05) identified from logistic regression. Supplemental Tables 2A-C provide a full list of variables identified from logistic regression. A full list of significant variables discovered as part of the KG link prediction methodology can be found in Supplemental Table 3 (Supp Table 3 is also available on Github[50]). Significant features from both analyses are indicated in bold in Tables 2A-C below.

**Table 2A-C.**
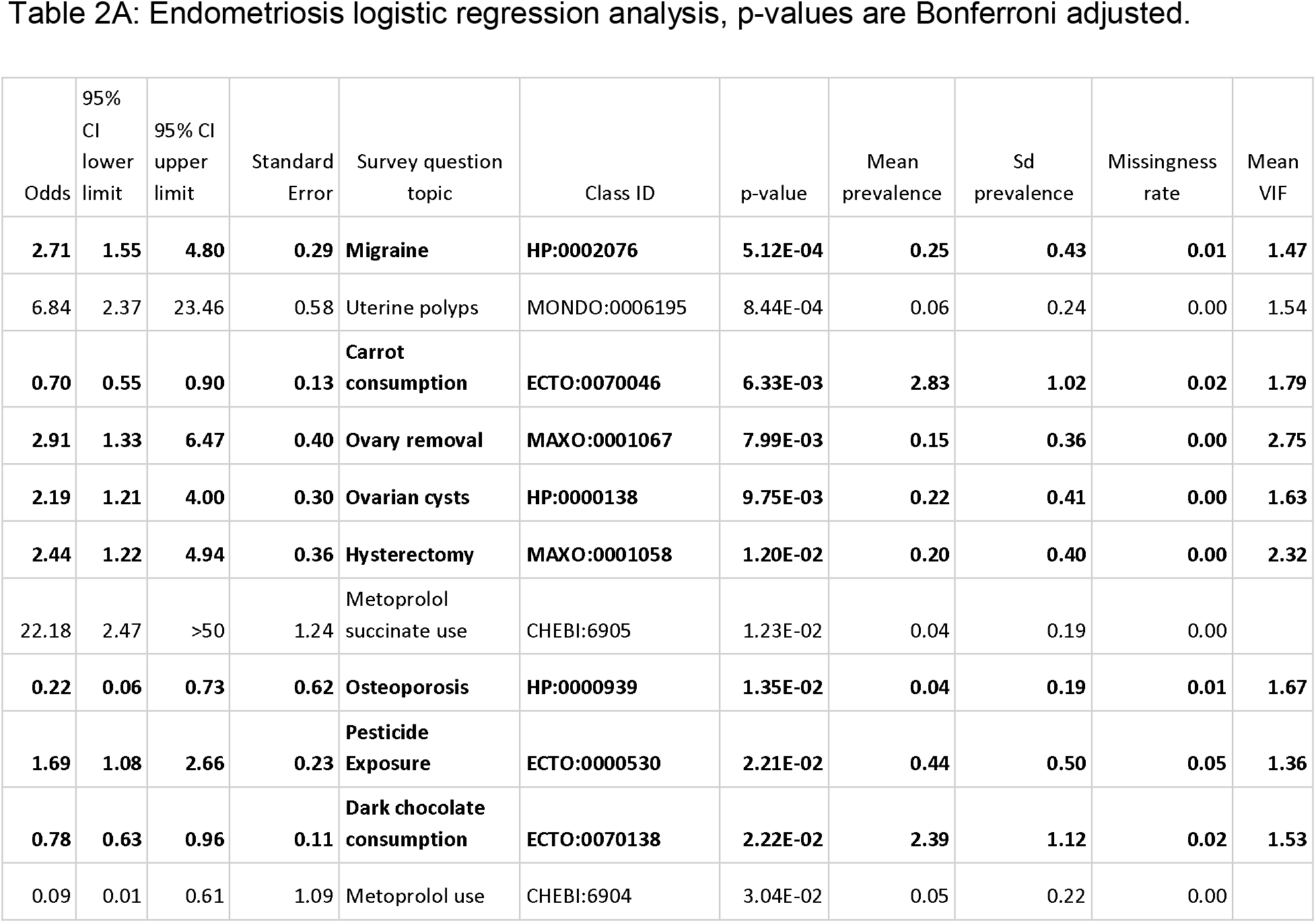
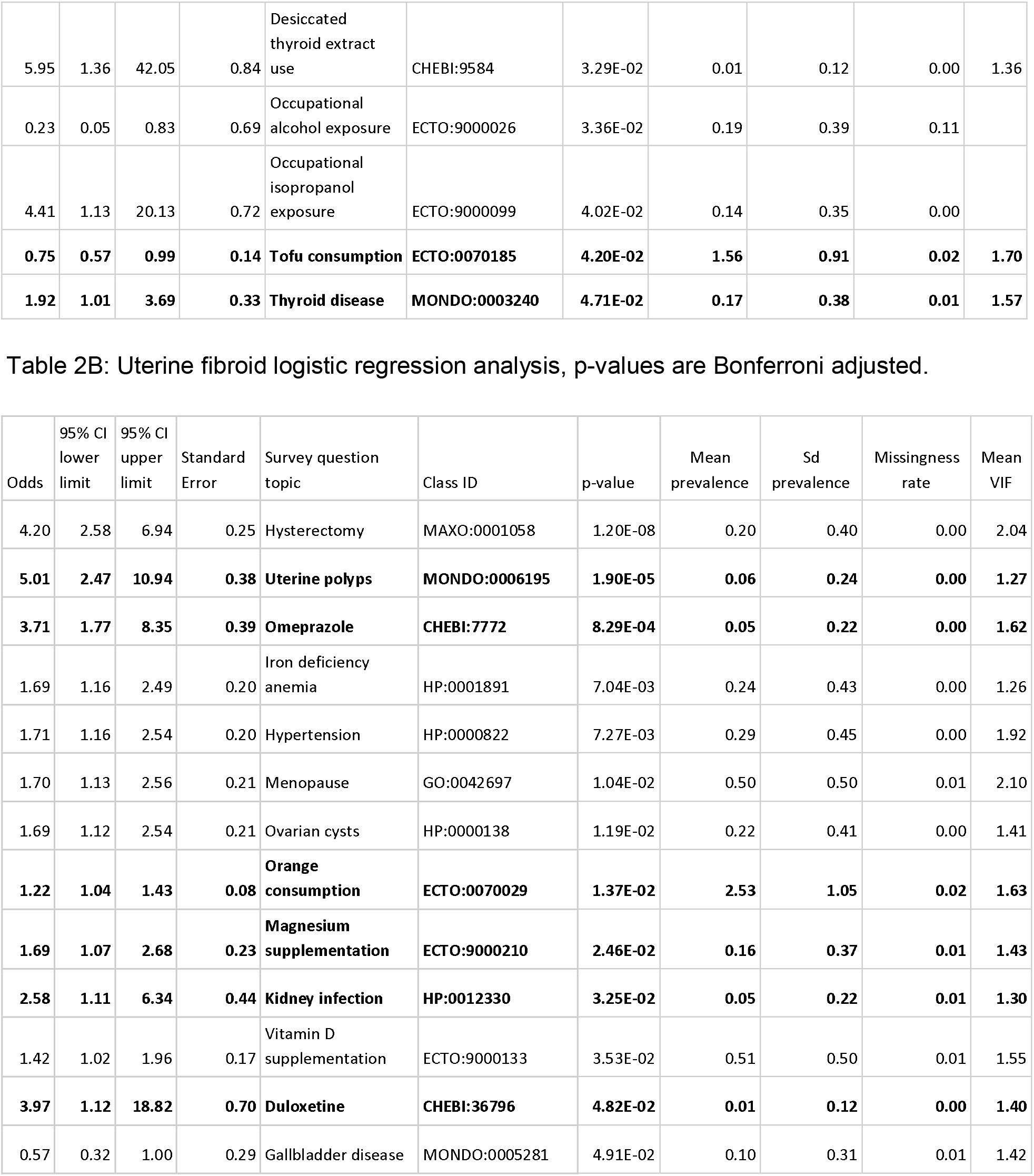
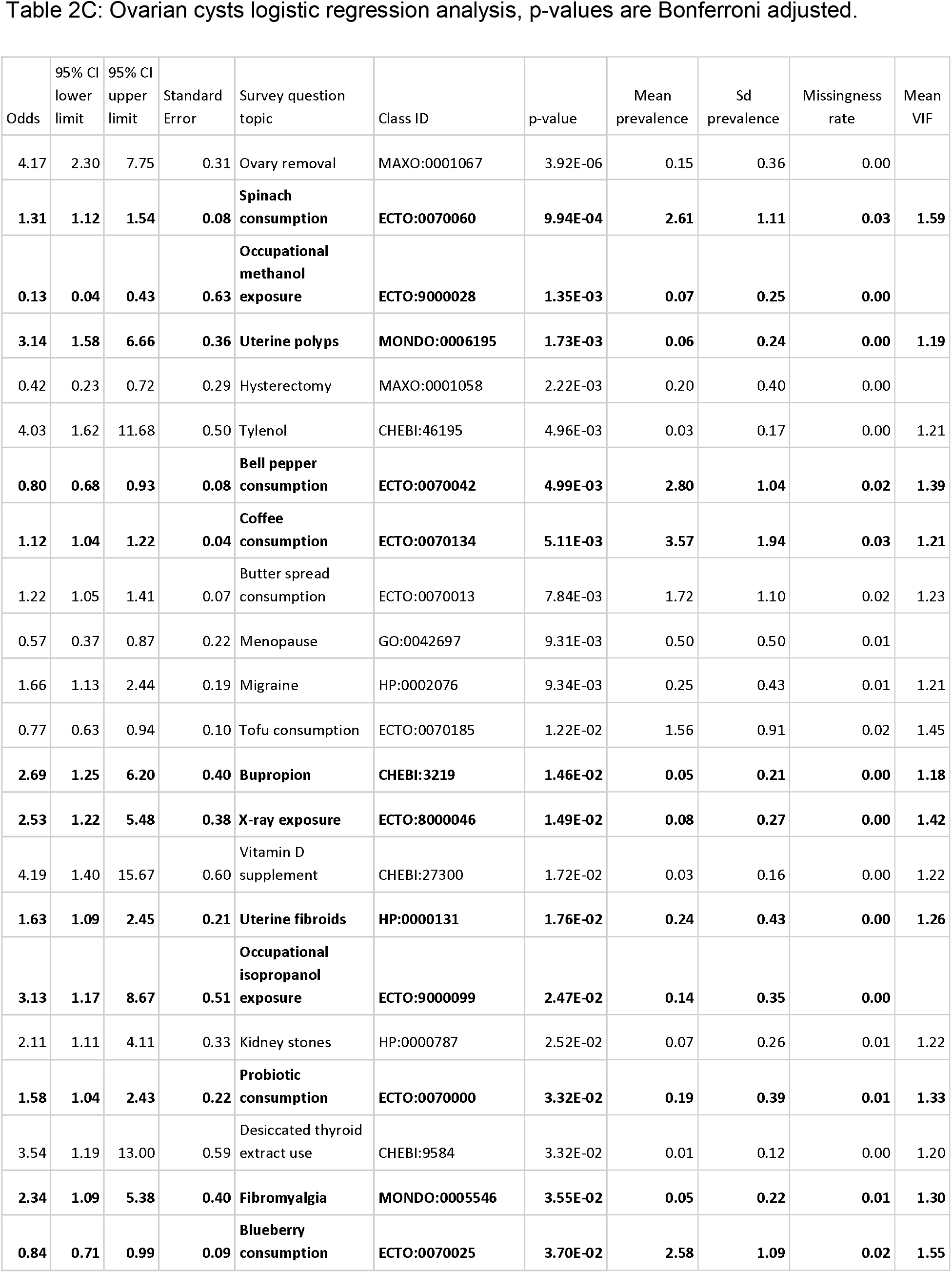
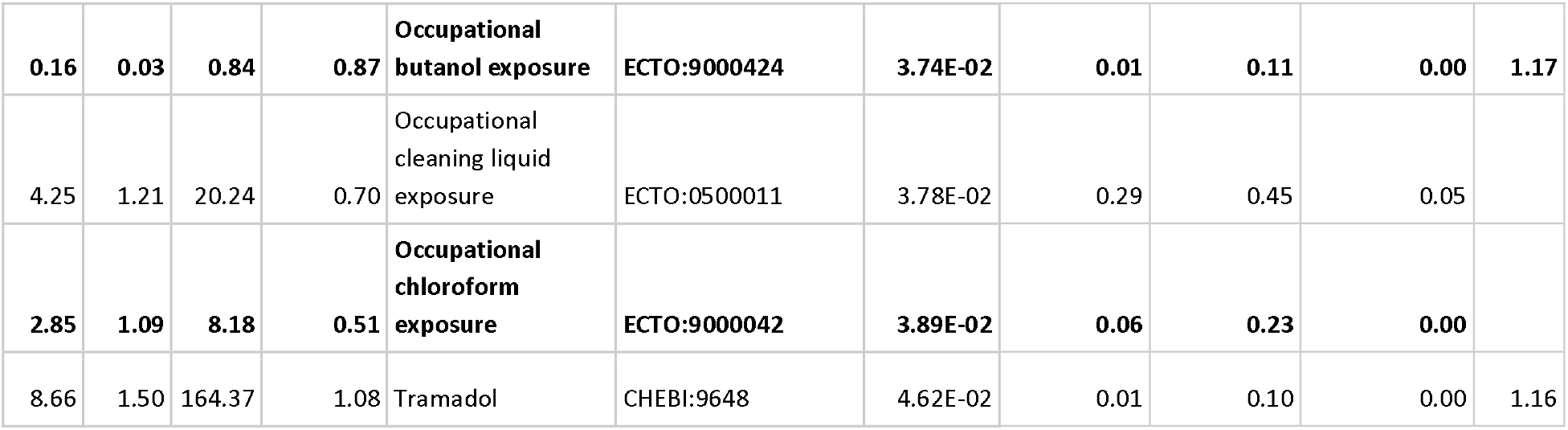
Significant features identified via logistic regression. Variables that are direct matches in the significant KG results are displayed in bold. Unreported Mean Variance Inflation Factor (VIF) scores indicate inadequate information available to calculate the score.

## DISCUSSION

Our work developing a KG with survey-based data and conducting machine learning to predict variables associated with FRDs is the first of its kind. The logistic regression model we developed for comparison supports our findings using this novel approach. Comparing the logistic regression and KG models resulted in numerous exact matches for medical conditions and procedures, environmental exposures, medications, and dietary exposures for the considered FRDs. Endometriosis and ovarian cysts were significantly associated with other gynecological conditions and procedures. Positive responses to questions regarding hysterectomy, ovary removal, and ovarian cysts were all associated with endometriosis. A possible explanation for the procedure associations is that ovary removal and hysterectomy are offered as endometriosis treatment options when other therapies have been unsuccessful.[51,52] However the timing of disease onset and medical procedures in this dataset was unavailable. Endometriosis can present as an ovarian endometrioma, an endometriotic cyst in the ovary,[53] which may be related to the significant endometriosis and ovarian cyst association identified. It is important to note that screening for any of these gynecological conditions may contribute to the identification of another gynecological comorbidity due to increased potential for detection.

Use of duloxetine was significantly associated with uterine fibroids in this study. Duloxetine is a medication primarily used for treatment of major depressive disorder, generalized anxiety disorder, chronic musculoskeletal pain, and fibromyalgia.[54] While duloxetine does not have a documented relationship with FRDs in current literature, there is a strong association between depression and mental health concerns in individuals with FRDs. Individuals with uterine fibroids have been documented to experience higher rates of depression and anxiety compared to controls, particularly amongst individuals who experience pain symptoms or who have undergone a hysterectomy.[55] Given the increased prevalence of mental health conditions amongst individuals with FRDs, individuals with these conditions may be more likely to take antidepressants or similar medications which may be related to this finding.

Omeprazole use was associated with increased odds of uterine fibroids. Omeprazole is a proton pump inhibitor, used to treat gastroesophageal reflux disease (GERD), ulcers, and other conditions characterized by excessive stomach acid.[56] Omeprazole has no reported side effects related to uterine fibroid development, but bulk-related symptoms may present due to uterine fibroids as the enlarged fibroids can distort the abdominal anatomy and cause abdominal bloating and pressure.[57] Uterine fibroids have been denoted as an associated disorder for individuals with Barrett’s esophagus, a gastrointestinal complication of GERD.[58,59]

We identified multiple associations between diet and FRDs. Tofu consumption was associated with significantly decreased odds of endometriosis. Tofu, a processed soybean curd, is often studied for its health benefits related to its high isoflavone content.[60] Isoflavones are of interest given their known antioxidant properties.[61] It is hypothesized that excessive inflammation observed with endometriosis may be mitigated through isoflavone exposure.[61,62] Supporting the findings of the present study, prior work has reported an inverse relationship between urinary isoflavone concentration and severe endometriosis.[63] However, a set of case studies investigating excessive soy consumption found high soy intake to be related to dysmenorrhea, endometriosis, and uterine fibroids.[64] Because of the higher rates of soy consumption among Asian individuals compared to other groups,[65] it is notable that prevalence of endometriosis is higher in Asian populations than in other racial groups.[66,67] However, data on race were unavailable for analysis. Notably, soy isoflavones are also phytoestrogens, given their ability to bind to estrogen receptors and contribute to estrogenic activity in humans[61]. Isoflavones have been denoted as potential endocrine disruptors, however these long term mechanistic effects are not fully elucidated.[60] While our results are inconclusive, further research evaluating soy consumption and endometriosis may be helpful for guidance on prevention and management.

Carrot consumption was also significantly associated with decreased odds of endometriosis. Consumption of fruits and vegetables has been identified as protective against endometriosis, potentially due to the anti-inflammatory properties of dietary components, including vitamins C and E.[68,69] Carrots contain high levels of antioxidant carotenoids, which may reduce the inflammatory responses that occur in individuals with endometriosis.[70] The effects of carrot consumption are inconsistent in the literature, with multiple investigations reporting no significant associations between carrots and endometriosis.[71,72] Further exploratory work is needed.

By utilizing a novel KG methodology and comparing the results with those from a traditional logistic regression model, we generated and corroborated multiple hypotheses of the effects of modifiable lifestyle factors on FRDs. The KG method presented here is an effective hypothesis-generation strategy, but the results should not be construed as causal as in other survey-based methodologies. The logistic regression approach indicated directionality for survey variables, which cannot be calculated using existing KG methods. The KG model identified an unranked list of predicted significant factors that require further assessment to identify variables of interest. Given the novelty of applying the KG method in survey-based data, its successful application in the present work showcases the potential of computational survey investigations using biomedical ontologies. Collecting data with ontology alignment in mind or retroactively performing ontology alignment for secondary data analysis will provide opportunities to apply KG study designs for hypothesis generation.

### Limitations

This work has limitations due to the nature of the PEGS dataset, namely the North Carolina-specific population and the lower percentage of individuals with FRDs compared to national prevalence estimates. Additionally, the dataset lacks information on temporality. PEGS participants are asked to describe their current eating habits, past and current exposures, and whether they have been diagnosed with an FRD. Given the lack of context for when onset of a condition occurred, it is difficult to identify the true impact of diet or environmental exposures, as they may have occurred before or after symptom presentation and disease diagnosis. Use of a survey design that includes temporality questions and collects information on gynecological history, demographics, and other potential confounders may improve the interpretation of findings.

Of note, our investigation used a binary variable of food consumption for individuals to indicate that they either do or do not consume a particular food. This approach was consistent for all food exposures, with no distinctions made between low and high consumption. Given the potentially wide range of consumption levels, this binary approach reduces the ability to decipher the impacts of dietary factors using the KG model. Binning data into “low”, “medium”, or “high” consumption levels should be considered for future work. Further, our named entity recognition approach to mapping string responses to survey questions can be improved by grouping similar medications (e.g., regular versus extended-release formats). Additionally, machine learning approaches that consider specific values for dietary intake (e.g., the number of apples consumed per week) when creating link predictions in a KG model would greatly benefit future nutrition investigations of this variety.

The performance of our KG model resulted in a substantial list of findings, many with similarly high prediction scores. While edge prediction provides prediction values between 0 and 1, equally ranked results make prioritization for hypothesis generation challenging. As such, efforts should be made to improve the prioritization of KG findings to enable hypothesis development. While areas for improvement exist in this study design, we identified multiple predicted variables, including modifiable lifestyle factors, for FRD. Additional results, including those resulting exclusively from KG analysis, may result in meaningful hypotheses in future investigations of FRDs.

## CONCLUSION

FRDs are highly impactful conditions for women globally, and there is a need to identify modifiable factors associated with these disorders. Limited investigations using ontologies or KG structures for investigations of FRDs have been conducted, and most existing studies have not accounted for modifiable lifestyle factors such as diet and environmental exposures. Using KG and logistic regression approaches, we identified a variety of potential intervention points for FRDs that can be pursued in future work. Because they are based on open-source, biomedical ontologies and computational resources, the novel methodologies used in this study can be repurposed for additional investigations.

## Author Contributions

Manuscript drafting: LEC, EC, QEH, MAH

Critical revision of the manuscript for important intellectual content: LEC, EC, JR, QEH, KS, HH, GV, CS, AMR, JEH, MAH

Biological subject matter expertise: LEC, QEH

Clinical subject matter expertise: LEC, QEH, JEH, PNR

Data analysis: LEC, EC, JR, KS, GV

Data curation: LEC, EC, HH, AMR

Data visualization: LEC, EC, TP, JR

Funding acquisition: LEC, JEH, MAH

Statistical analysis: LEC, EC, JR, CJM, MAH

## Data Availability

All data produced in the present work are contained in the manuscript. Data from the Personalized Environments and Genes Study used for the present work is available upon request from the National Institute of Environmental Health Sciences.

## Acknowledgements

We greatly appreciate the contributions of Hannah Blau, Bryan Laraway, Katherina Cortes, Harry Caufield, Anne Thessen, Sierra Moxon, Blessy Antony, and Karamarie Fecho to the design of this study.

## SUPPLEMENTAL METHODS

### OntoRunNER

OntoRunNER is a named entity recognition (NER) tool that reads strings of written content, which is then compared to a designated term list to identify what content is an exact or close match to the term list provided. For this project, we used OntoRunNER to coordinate strings of data input primarily regarding chemicals (medications or agricultural chemicals) to Chemical Entities of Biological Interest (ChEBI) Ontology terms. OntoRunNER allowed for both exact matches of the primary ChEBI label as well as synonyms of ChEBI content. It also allowed the string and ontology term to be considered a “match” if there were four or fewer different characters between them. Following use of OntoRunNER, we hand-reviewed terminology mappings for accuracy, which included additional manual mapping creation for common misspellings (e.g., ‘aderall’ versus ‘adderall’). We excluded from the dataset any string that could not be confidently mapped using OntoRunNER or manual means in an effort to avoid introducing inaccuracies.

### Knowledge graph preparation for embedding

To prepare the graph for embedding using GRAPE, we removed all disconnected (singleton) nodes that shared no edges with other nodes as they provided little to no information for our link prediction model. We then selected the largest component of the graph, or the subgraph of the knowledge graph with the most nodes. Components were connected subgraphs of the primary graph, with the largest component the one with the highest number of nodes and thus offering the greatest capacity to develop link predictions. Using only the largest component removed 7.1% of respondents (n = 691) from the final analysis. As these respondents were not included in the largest component, it is likely the data available from their survey responses was insufficient for informing significant link predictions.

We used the DeepWalkSkipGram approach to learn the latent representations of the nodes within this graph network. This approach uses the Deep Walk deep learning approach, where nodes within the graph are treated like words, and random walks between nodes can be taken to create sentences [36]. The Skip-gram component includes inputting a single node and then contextualizing and classifying the word based on other words from the same sentence, allowing for projections of words coming both before and after the single word (node) of interest [42]. This approach to embedding allows for the creation of sentence structure using nodes and then the assessment of which nodes should be located near each other in the low-dimensional embedding visualization.

### Logistic regression analysis

In more detail, the first two steps of supervised feature selection and random forest (RF) training were applied on 50 external stratified holdouts (train:test ratio 0.9:0.1). The variables that, on the average of all the holdouts, had an RF importance score greater than zero were chosen as the most important features.

For the first step of supervised feature selection on the training set, we ran preliminary experiments to choose among univariate feature selection techniques (where variables showing significant correlation with the label were selected), Boruta feature selection [49], permutation-based RF importance, and elastic nets (where the value of the elastic-net regularization parameter λ is set via internal five-fold cross-validation and the α parameter balancing the amount of lasso and ridge constraints is set to 0.5) [50]. Given the comparable preliminary results, we opted for elastic nets due to their higher regularization capability, which results in a lower number of selected features. To avoid overfitting, we trained the RF by balancing the samples used to choose each split (sampsize) and chose the RF parameters (number of trees for the RF and the number “mtry” of variables considered to define each split) by a grid search on 100 internal rebalanced holdouts (train:test ratio = 0.9:0.1) to maximize the area under the precision-recall curve, which is appropriate in the case of imbalanced classes.

Next, for the directionality of scores, the important variables were used to train *logistic regression classifiers.* Considering how rare events resulting in highly imbalanced datasets may cause sharp logistic regression underestimates [48], we ran logistic regression on 100 holdouts rebalanced by undersampling. We averaged the results of the 100 iterations (odds and *P* values) to get the final estimates. We also calculated the variance inflation factor (VIF) and mean prevalence for each variable. These values are reported with the full logistic regression results in Supplemental Tables 2A-C. Of note, variables with a large VIF (>4) or a low mean prevalence score (0.001) may not be reliable regression outcomes due to colinearity or lack of sufficient data to determine the influence of a variable on the outcomes of interest.

