## Supplemental Tables for "Predicting nutrition and environmental factors associated with female reproductive disorders using a knowledge graph and random forests"

Supplemental Table 1A: Health Conditions Key

| HE_B007_hypertension | HP:0000822 |
| --- | --- |
| HE_B007a_hypertension_preg | MONDO:0024664 |
| HE_B008_high_cholesterol | HP_0003124 |
| HE_B009_atherosclerosis | HP:0002621 |
| HE_B010_cardiac_arrhythmia | HP:0011675 |
| HE_B011_angina | HP:0001681 |
| HE_B012_heart_attack | HP:0001658 |
| HE_B013_coronary_artery | MONDO:0005010 |
| HE_B014_congestive_heart_failure | HP:0001635 |
| HE_B016_raynauds | MONDO:0008364 |
| HE_B017_blood_clots | HP:0004936 |
| HE_B019_stroke_mini | HP:0002326 |
| HE_B020_stroke | HP:0001297 |
| HE_B020a_stroke_type | HP:0002140 |
| HE_C021_pre_diabetes | MONDO:0006920 |
| HE_C022_diabetes | MONDO:0005015 |
| HE_C022a_diabetes_preg | MONDO:0005406 |
| HE_C022b_diabetes_insulin | MAXO:0000259 |
| HE_C022d_diabetes_pills | MAXO:0000257 |
| HE_C023_thyroid_disease | MONDO:0003240 |
| HE_C023a_hyperthyroidism | MONDO:0004425 |
| HE_C023b_hypothyroidism | MONDO:0005420 |
| HE_C023c_thyroid_enlarged | HP:0000853 |
| HE_C023d_nodule_benign | MONDO:0006107 |
| HE_D025_copd | MONDO:0005002 |
| HE_D026_ipf | MONDO:0008345 |
| HE_D027_tb | MONDO:0018076 |
| HE_D030_asthma | MONDO:0004979 |
| HE_E031_epilepsy | MONDO:0005027 |
| HE_E032_migraine | HP:0002076 |
| HE_F037_celiac | HP:0002608 |
| HE_F038_lactose_intolerance | HP:0004789 |
| HE_F039_crohns | HP:0100280 |
| HE_F040_ulcerative_colitis | HP:0100279 |
| HE_F041_polyps | HP:0200063 |
| HE_F042_gallbladder_disease | MONDO:0005281 |
| HE_F043_stomach_ulcer | HP:0002592 |
| HE_F044_cirrhosis | HP:0001394 |
| HE_F045_fatty_liver | MONDO:0004790 |
| HE_F046_hepatitis | MONDO:0002251 |
| HE_G047_ckd | HP:0012622 |
| HE_G048_esrd | MONDO:0004375 |
| HE_G049_kidney_stones | HP:0000787 |
| HE_G050_kidney_infection | HP:0012330 |
| HE_G051_pkd | MONDO:0020642 |
| HE_H053_scleroderma | HP:0100324 |
| HE_H055_fibromyalgia | MONDO:0005546 |
| HE_H056_lupus | MONDO:0007915 |
| HE_H057_sjogrens | MONDO:0010030 |
| HE_I058_hemochromatosis | MONDO:0006507 |
| HE_I059_iron_anemia | HP:0001891 |
| HE_I060_pernicious_anemia | MONDO:0008228 |
| HE_I061_sickle_cell | MONDO:0011382 |
| HE_J062_bone_loss | HP:0000938 |
| HE_J063_osteoporosis | HP:0000939 |
| HE_J064_gout | MONDO:0005393 |
| HE_J065_myositis | HP:0100614 |
| HE_J066_rheu_arthritis | HP:0001370 |
| HE_J067_osteoarthritis | HP:0002758 |
| HE_K069_psoriasis | MONDO:0005083 |
| HE_K070_eczema | HP:0000964 |
| HE_K071_urticaria | HP:0001025 |
| he_m083_hysterectomy | MAXO:0001058 |
| he_m084_ovaries | MAXO:0001067 |
| he_m085_menopause | GO:0042697 |
| he_m087_birth_control | ECTO:9001831 |
| he_m088_hormone_replacement | ECTO:2000005 |
| he_m089_endometriosis | MONDO:0005133 |
| he_m090_uterine_polyps | MONDO:0006195 |
| he_m091_uterine_tumors | HP:0000131 |
| he_m092_ovarian_cysts | HP:0000138 |
| he_o108a_cancer_breast_a | MONDO:0007254 |
| he_o109_cancer_cervix | MONDO:0002974 |
| he_o123_cancer_ovary | MONDO:0008170 |
| he_o134_cancer_uterus | MONDO:0002715 |

Supplemental Table 1B: External Exposures Key

| EA_A060_pest_regularly | ECTO:0000530 |
| --- | --- |
| ea_b092_solvent_d | ECTO:9000107 |
| ea_b092a_benzene | ECTO:9000034 |
| ea_b092b_chloroform | ECTO:9000042 |
| ea_b092c_chloroprene | ECTO:9000043 |
| ea_b092d_dichlorobenzene | ECTO:9000058 |
| ea_b092e_ethyl_benzene | ECTO:9000059 |
| ea_b092f_ethyl_dichloride | ECTO:9000039 |
| ea_b092g_perchloroethylene | ECTO:9000074 |
| ea_b092h_toluene | ECTO:9000036 |
| ea_b092i_trichloroethylene | ECTO:9000530 |
| ea_b092j_xylenes | ECTO:9000035 |
| ea_b098a_brake_fluid | ECTO:0500001 |
| ea_b098b_transmission_fluid | ECTO:0500003 |
| ea_b098c_hydraulic_fluid | ECTO:0500002 |
| ea_b098d_motor_oil | ECTO:0500004 |
| ea_b104_cleaning_d | ECTO:0500011 |
| ea_b104a_bleach | ECTO:9000076 |
| ea_b104b_ammonia | ECTO:9000031 |
| ea_b104c_carbon_tetrachloride | ECTO:9000048 |
| ea_b110_metal_d | ECTO:9002163 |
| ea_b110a_arsenic | ECTO_9000032 |
| ea_b110b_beryllium | ECTO:9000941 |
| ea_b110c_cadmium | ECTO_0001566 |
| ea_b110d_chromates | ECTO:9000038 |
| ea_b110e_lead | ECTO_9000945 |
| ea_b110f_mercury | ECTO_0001571 |
| ea_b110g_nickel | ECTO:9000063 |
| ea_b116_alcohol_d | ECTO:9000026 |
| ea_b116a_isopropanol | ECTO:9000099 |
| ea_b116b_methanol | ECTO:9000028 |
| ea_b116c_ethanol | ECTO:9000027 |
| ea_b116d_butanol | ECTO:9000424 |
| ea_b122_pest_d | ECTO:0000530 |
| ea_b122a_ethyl_dibromide | ECTO:9000060 |
| ea_b122b_chlorinated_naph | ECTO:9002147 |
| ea_b122c_insecticides | ECTO:9000089 |
| ea_b122d_fungicides | ECTO:9000092 |
| ea_b122e_herbicides | ECTO:0000515 |
| ea_b122f_ethyl_fumigants | ECTO:9002113 |
| ea_b122g_rodenticides | ECTO:9000090 |
| ea_b128_plastic_d | ECTO:7000147 |
| ea_b128a_bpa | ECTO:9000057 |
| ea_b128b_vinyl_chloride | ECTO:9001302 |
| ea_b128c_styrene | ECTO:9000069 |
| ea_b128d_phosgene | ECTO:9000070 |
| ea_b128e_phenol | ECTO:9000071 |
| ea_b128f_tdi | ECTO:9000103 |
| ea_b128g_methylene_bis | ECTO:9000104 |
| ea_b134_dust_d | ECTO:7000001 |
| ea_b134a_coal_dust | ECTO:7000005 |
| ea_b134b_fiberglass | ECTO:7000017 |
| ea_b134c_rock_dust | ECTO:7000144 |
| ea_b134d_silica | ECTO:7000030 |
| ea_b134e_talc | ECTO:7000028 |
| ea_b140_emissions_d | ECTO:7000139 |
| ea_b140a_nitrous_oxide | ECTO:9000051 |
| ea_b140b_carbon_dioxide | ECTO:9000049 |
| ea_b140c_carbon_monoxide | ECTO:0000207 |
| ea_b140d_ozone | ECTO:9000052 |
| ea_b146a_pbbs | ECTO:9001538 |
| ea_b146b_pcbs | ECTO:9000068 |
| ea_b146c_radiation | ECTO:0000001 |
| ea_b146d_xrays | ECTO:8000046 |
| ea_b146e_welding | ECTO:7000129 |
| ea_b152_acid_d | ECTO:9002146 |
| ea_b152a_hydrochloric_acid | ECTO:9000016 |
| ea_b152b_sulfuric_acid | ECTO:9000017 |
| ea_b152c_phosphoric_acid | ECTO:9000018 |
| ea_b152d_acetic_acid | ECTO:9000019 |
| ea_b152e_nitric_acid | ECTO:9000020 |
| ea_b158_alkalis_d | ECTO:9000021 |
| ea_b158a_sodium_oh | ECTO:9000022 |
| ea_b158b_calcium_oh | ECTO:9000024 |
| ea_b158c_potassium_oh | ECTO:9000025 |
| ea_b158d_magnesium_oh | ECTO:9000023 |
| ea_b164_stains_d | ECTO:0500015 |
| ea_b164a_shellac | ECTO:7000132 |
| ea_b164b_wood | ECTO:0500016 |
| ea_b164c_resin | ECTO:7000131 |
| ea_b164d_polyurethane | ECTO:9000082 |
| ea_b164e_lacquer | ECTO:0500017 |
| ea_b164f_acrylic | ECTO:7000142 |
| ea_b170_paint_d | ECTO:0500024 |
| ea_b170a_primer | ECTO:7000140 |
| ea_b170b_enamel | ECTO:0500021 |
| ea_b170c_oil_based | ECTO:7000141 |
| ea_b170d_acrylic | ECTO:7000142 |
| ea_b170e_luminescent | ECTO:7000143 |
| ea_b170f_acetone | ECTO:9000085 |
| ea_b170g_turpentine | ECTO:0500022 |
| ea_b170h_naphtha | ECTO:0500023 |
| ea_b170i_ethyl_ket | ECTO:9000083 |
| ea_b176a_desflurane | ECTO:9002164 |
| ea_b176b_enflurane | ECTO:9001437 |
| ea_b176c_halothane | ECTO:9001450 |
| ea_b176d_isoflurane | ECTO:9002166 |
| ea_b176e_methoxyflurane | ECTO:9001465 |
| ea_b176f_nitrous | ECTO:9000051 |
| ea_b176g_sevoflurane | ECTO:9002165 |
| ea_b182_adhesives_d | ECTO:7000130 |
| ea_b182a_white_glue | ECTO:9000079 |
| ea_b182b_rubber_cement | ECTO:7000149 |
| ea_b182c_neoprene | ECTO:9002141 |
| ea_b182d_vinyl_a | ECTO:9002150 |
| ea_b182e_epoxy | ECTO:9002142 |
| ea_b182f_urethane | ECTO:9000081 |
| ea_b182g_polyimides | ECTO:9002143 |
| ea_b182h_cyanoacrylates | ECTO:7000146 |
| ea_b182i_wallpaper_paste | ECTO:7000150 |
| ea_b194_ink_d | ECTO:7000157 |
| ea_b194a_hair_dye | ECTO:7000151 |
| ea_b194b_leather_dye | ECTO:0500006 |
| ea_b194c_textile_dye | ECTO:0500008 |
| ea_b194d_paper_dye | ECTO:0500007 |
| ea_b194e_india_ink | ECTO:0500009 |
| ea_b194f_inkjet | ECTO:7000155 |
| ea_b194g_gel_ink | ECTO:7000153 |
| ea_b194h_pen_ink | ECTO:0500010 |
| ea_b194j_soy_ink | ECTO:7000154 |
| ea_b194k_pharma_ink | ECTO:7000152 |

Supplemental Table 1C: Internal Exposures Key

| eb_a028_probiotic_PARQ | ECTO:0070000 |
| --- | --- |
| eb_i172_milk_skim | ECTO:0070001 |
| eb_i173_milk_lowfat | ECTO:0070002 |
| eb_i174_milk_whole | ECTO:0070003 |
| eb_i175_milk_nondairy | ECTO:0070004 |
| eb_i176_cream | ECTO:0070007 |
| eb_i177_cream_nondairy | ECTO:0070008 |
| eb_i178_frozen_yogurt | ECTO:0070009 |
| eb_i179_ice_cream | ECTO:0070010 |
| eb_i180_butter | ECTO:0070011 |
| eb_i181_margarine | ECTO:0070012 |
| eb_i182_spread_butter | ECTO:0070013 |
| eb_i183_yogurt_nonfat | ECTO:0070014 |
| eb_i184_yogurt_lowfat | ECTO:0070015 |
| eb_i185_yogurt_regular | ECTO:0070016 |
| eb_i186_cottage_cheese | ECTO:0070017 |
| eb_i187_cream_cheese | ECTO:0070018 |
| EB_I189_cheese_lowfat | ECTO:0070020 |
| EB_I190_cheese_regular | ECTO:0070021 |
| EB_I191_apple | ECTO:0070022 |
| EB_I192_avocado | ECTO:0070023 |
| EB_I193_banana | ECTO:0070024 |
| EB_I194_blueberries | ECTO:0070025 |
| EB_I195_cantaloupe | ECTO:0070026 |
| EB_I196_grapes | ECTO:0070027 |
| EB_I197_grapefruit | ECTO:0070028 |
| EB_I198_orange | ECTO:0070029 |
| EB_I199_peach_plum | ECTO:0070182 |
| EB_I200_pear | ECTO:0070033 |
| EB_I201_raspberries | ECTO:0070034 |
| EB_I202_strawberries | ECTO:0070035 |
| EB_I203_tomatoes | ECTO:0070036 |
| EB_I204_tomato_juice | ECTO:0070037 |
| EB_I205_tomato_sauce | ECTO:0070038 |
| EB_I206_watermelon | ECTO:0070039 |
| EB_I207_beans_lentils | ECTO:0070183 |
| EB_I208_bell_pepper | ECTO:0070042 |
| EB_I209_broccoli | ECTO:0070043 |
| EB_I210_brussels_sprouts | ECTO:0070044 |
| EB_I211_cabbage | ECTO:0070045 |
| EB_I212_carrots | ECTO:0070046 |
| EB_I213_cauliflower | ECTO:0070047 |
| EB_I214_corn | ECTO:0070048 |
| EB_I216_greens | ECTO:0070184 |
| EB_I217_onion | ECTO:0070056 |
| EB_I218_peas_lima_beans | ECTO:0070183 |
| EB_I220_spinach_raw | ECTO:0070060 |
| EB_I286_saccharin | ECTO:0070061 |
| EB_I221_green_beans | ECTO:0070062 |
| EB_I222_tofu | ECTO:0070185 |
| EB_I223_winter_squash | ECTO:0070067 |
| EB_I224_yams | ECTO:0070069 |
| EB_I225_eggs | ECTO:0070070 |
| EB_I226_hotdog_beef | ECTO:0070071 |
| EB_I227_hotdog_poultry | ECTO:0070186 |
| EB_I228_chicken_turkey | ECTO:0070075 |
| EB_I229_bacon | ECTO:0070185 |
| EB_I230_deli_meats | ECTO:0070078 |
| EB_I231_other_processed_meat | ECTO:0070084 |
| EB_I232_hamburger | ECTO:0070085 |
| EB_I233_beef_pork_lamb | ECTO:0070187 |
| EB_I234_ham | ECTO:0070090 |
| EB_I235_canned_tuna | ECTO:0070091 |
| EB_I236_crustacean | ECTO:0070188 |
| EB_I237_dark_fish | ECTO:0070209 |
| EB_I238_other_fish | ECTO:0070189 |
| EB_I239_cold_cereal | ECTO:0070104 |
| EB_I240_oatmeal | ECTO:0070107 |
| EB_I241_cooked_cereal | ECTO:0070107 |
| EB_I242_wheat_crackers | ECTO:0070108 |
| EB_I243_other_crackers | ECTO:0070109 |
| EB_I244_white_bread | ECTO:0070110 |
| EB_I245_rye | ECTO:0070112 |
| EB_I246_whole_wheat | ECTO:0070114 |
| EB_I247_bagel | ECTO:0070116 |
| EB_I248_muffin | ECTO:0070119 |
| EB_I249_pancakes | ECTO:0070121 |
| EB_I250_brown_rice | ECTO:0070123 |
| EB_I251_white_rice | ECTO:0070124 |
| EB_I252_pasta | ECTO:0070125 |
| EB_I253_tortillas | ECTO:0070128 |
| EB_I254_fries | ECTO:0070129 |
| EB_I255_potatoes | ECTO:0070130 |
| EB_I256_pizza | ECTO:0070131 |
| EB_I261_tea_decaf | ECTO:0070132 |
| EB_I262_tea | ECTO:0070133 |
| EB_I264_coffee | ECTO:0070134 |
| EB_I266_water | ECTO:0070135 |
| EB_I267_chocolate_milk | ECTO:0070137 |
| EB_I268_chocolate_dark | ECTO:0070138 |
| EB_I269_other_candy | ECTO:0070139 |
| EB_I270_cookies | ECTO:0070189 |
| EB_I271_cakes | ECTO:0070189 |
| EB_I272_jelly_honey | ECTO:0070148 |
| EB_I273_peanut_butter | ECTO:0070151 |
| EB_I274_potato_chips | ECTO:0070152 |
| EB_I275_pretzels | ECTO:0070153 |
| EB_I276_peanuts | ECTO:0070154 |
| EB_I277_walnuts | ECTO:0070155 |
| EB_I278_other_nuts | ECTO:0070156 |
| EB_I279_olive_oil | ECTO:0070157 |
| EB_I280_mayonnaise | ECTO:0070158 |
| EB_I281_salad_dressing | ECTO:0070159 |
| EB_I287_sugar | ECTO:0070161 |
| eb_a001_multivitamin_PARQ | ECTO:0070210 |
| eb_a002_vitamin_a_PARQ | ECTO:9000125 |
| eb_a003_vitamin_b3_PARQ | ECTO:9002151 |
| eb_a004_vitamin_b6_PARQ | ECTO:9000243 |
| eb_a005_vitamin_b12_PARQ | ECTO:9000229 |
| eb_a006_vitamin_b_comp_PARQ | ECTO:9002169 |
| eb_a007_vitamin_c_PARQ | ECTO:9000143 |
| eb_a008_vitamin_d_PARQ | ECTO:9000133 |
| eb_a009_vitamin_e_PARQ | ECTO:9000224 |
| eb_a010_calcium_PARQ | ECTO:9000299 |
| eb_a011_chromium_PARQ | ECTO:9000037 |
| eb_a012_iron_PARQ | ECTO:9000087 |
| eb_a013_magnesium_PARQ | ECTO:9000210 |
| eb_a014_potassium_PARQ | ECTO:9000253 |
| eb_a015_selenium_PARQ | ECTO:9000192 |
| eb_a016_zinc_PARQ | ECTO:9000954 |
| eb_a017_blk_cohosh_PARQ | ECTO:0070162 |
| eb_a018_coq10_PARQ | ECTO:0070163 |
| eb_a019_fish_oil_PARQ | ECTO:0070164 |
| eb_a020_flaxseed_oil_PARQ | ECTO:0070165 |
| eb_a021_folic_acid_PARQ | ECTO:9000123 |
| eb_a022_gingko_biloba_PARQ | ECTO:0070166 |
| eb_a023_ginseng_PARQ | ECTO:0070167 |
| eb_a024_glucosamine_PARQ | ECTO:0070168 |
| eb_a025_melatonin_PARQ | ECTO:0070169 |
| eb_a026_milk_thistle_PARQ | ECTO:0070170 |
| eb_a027_omega3_PARQ | ECTO:0070171 |
| eb_a028_probiotic_PARQ | ECTO:0070000 |
| eb_a029_red_yeast_rice_PARQ | ECTO:0070172 |
| eb_a030_resveratrol_PARQ | ECTO:0070173 |
| eb_a031_st_johns_wort_PARQ | ECTO:0070174 |

Supplemental Table 1D: Medications Key

| survey_question | survey_free_response | ontology_mapping |
| --- | --- | --- |
| eb_b034a_med3_name_CHILDQ | 5-htp | CHEBI:28171 |
| eb_b032a_med1_name_CHILDQ | 6-mercaptopurine | CHEBI:50667 |
| eb_b032a_med1_name_CHILDQ | abilify | CHEBI:31236 |
| eb_b033a_med2_name_CHILDQ | abilify | CHEBI:31236 |
| eb_b034a_med3_name_CHILDQ | abilify | CHEBI:31236 |
| eb_b036a_med5_name_CHILDQ | abilify | CHEBI:31236 |
| eb_b041a_med10_name_CHILDQ | abilify | CHEBI:31236 |
| eb_b033a_med2_name_CHILDQ | acebutolol | CHEBI:2379 |
| eb_b032a_med1_name_CHILDQ | acetaminophen | CHEBI:46195 |
| eb_b033a_med2_name_CHILDQ | acetaminophen | CHEBI:46195 |
| eb_b034a_med3_name_CHILDQ | acetaminophen | CHEBI:46195 |
| eb_b035a_med4_name_CHILDQ | acetaminophen | CHEBI:46195 |
| eb_b037a_med6_name_CHILDQ | acetaminophen | CHEBI:46195 |
| eb_b038a_med7_name_CHILDQ | acetaminophen | CHEBI:46195 |
| eb_b039a_med8_name_CHILDQ | acetaminophen | CHEBI:46195 |
| eb_b040a_med9_name_CHILDQ | acetaminophen | CHEBI:46195 |
| eb_b034a_med3_name_CHILDQ | acetazolamide | CHEBI:27690 |
| eb_b039a_med8_name_CHILDQ | acetyl-l-carnitine | CHEBI:57589 |
| eb_b032a_med1_name_CHILDQ | aciphex | CHEBI:8769 |
| eb_b034a_med3_name_CHILDQ | aciphex | CHEBI:8769 |
| eb_b033a_med2_name_CHILDQ | actonel | CHEBI:8869 |
| eb_b032a_med1_name_CHILDQ | acular | CHEBI:6130 |
| eb_b032a_med1_name_CHILDQ | acyclovir | CHEBI:2453 |
| eb_b033a_med2_name_CHILDQ | acyclovir | CHEBI:2453 |
| eb_b034a_med3_name_CHILDQ | acyclovir | CHEBI:2453 |
| eb_b035a_med4_name_CHILDQ | acyclovir | CHEBI:2453 |
| eb_b036a_med5_name_CHILDQ | acyclovir | CHEBI:2453 |
| eb_b037a_med6_name_CHILDQ | acyclovir | CHEBI:2453 |
| eb_b032a_med1_name_CHILDQ | advil | CHEBI:5855 |
| eb_b033a_med2_name_CHILDQ | advil | CHEBI:5855 |
| eb_b034a_med3_name_CHILDQ | advil | CHEBI:5855 |
| eb_b035a_med4_name_CHILDQ | advil | CHEBI:5855 |
| eb_b036a_med5_name_CHILDQ | advil | CHEBI:5855 |
| eb_b037a_med6_name_CHILDQ | advil | CHEBI:5855 |
| eb_b039a_med8_name_CHILDQ | advil | CHEBI:5855 |
| eb_b033a_med2_name_CHILDQ | alavert | CHEBI:6538 |
| eb_b034a_med3_name_CHILDQ | alavert | CHEBI:6538 |
| eb_b035a_med4_name_CHILDQ | alavert | CHEBI:6538 |
| eb_b032a_med1_name_CHILDQ | albuterol | CHEBI:2549 |
| eb_b033a_med2_name_CHILDQ | albuterol | CHEBI:2549 |
| eb_b034a_med3_name_CHILDQ | albuterol | CHEBI:2549 |
| eb_b035a_med4_name_CHILDQ | albuterol | CHEBI:2549 |
| eb_b036a_med5_name_CHILDQ | albuterol | CHEBI:2549 |
| eb_b037a_med6_name_CHILDQ | albuterol | CHEBI:2549 |
| eb_b038a_med7_name_CHILDQ | albuterol | CHEBI:2549 |
| eb_b040a_med9_name_CHILDQ | albuterol | CHEBI:2549 |
| eb_b034a_med3_name_CHILDQ | albuterol sulfate | CHEBI:2550 |
| eb_b036a_med5_name_CHILDQ | albuterol sulfate | CHEBI:2550 |
| eb_b032a_med1_name_CHILDQ | alendronate | CHEBI:2567 |
| eb_b033a_med2_name_CHILDQ | alendronate | CHEBI:2567 |
| eb_b034a_med3_name_CHILDQ | alendronate | CHEBI:2567 |
| eb_b035a_med4_name_CHILDQ | alendronate | CHEBI:2567 |
| eb_b036a_med5_name_CHILDQ | alendronate | CHEBI:2567 |
| eb_b038a_med7_name_CHILDQ | alendronate | CHEBI:2567 |
| eb_b032a_med1_name_CHILDQ | alfuzosin | CHEBI:51141 |
| eb_b033a_med2_name_CHILDQ | alfuzosin | CHEBI:51141 |
| eb_b033a_med2_name_CHILDQ | allerclear | CHEBI:6538 |
| eb_b036a_med5_name_CHILDQ | allerclear | CHEBI:6538 |
| eb_b032a_med1_name_CHILDQ | allopurinol | CHEBI:40279 |
| eb_b033a_med2_name_CHILDQ | allopurinol | CHEBI:40279 |
| eb_b034a_med3_name_CHILDQ | allopurinol | CHEBI:40279 |
| eb_b035a_med4_name_CHILDQ | allopurinol | CHEBI:40279 |
| eb_b036a_med5_name_CHILDQ | allopurinol | CHEBI:40279 |
| eb_b032a_med1_name_CHILDQ | alopurinol | CHEBI:40279 |
| eb_b035a_med4_name_CHILDQ | alopurinol | CHEBI:40279 |
| eb_b033a_med2_name_CHILDQ | alphagan p | CHEBI:51157 |
| eb_b032a_med1_name_CHILDQ | alprazolam | CHEBI:2611 |
| eb_b033a_med2_name_CHILDQ | alprazolam | CHEBI:2611 |
| eb_b034a_med3_name_CHILDQ | alprazolam | CHEBI:2611 |
| eb_b035a_med4_name_CHILDQ | alprazolam | CHEBI:2611 |
| eb_b036a_med5_name_CHILDQ | alprazolam | CHEBI:2611 |
| eb_b037a_med6_name_CHILDQ | alprazolam | CHEBI:2611 |
| eb_b033a_med2_name_CHILDQ | amaryl | CHEBI:5383 |
| eb_b036a_med5_name_CHILDQ | amaryl | CHEBI:5383 |
| eb_b032a_med1_name_CHILDQ | amiloride | CHEBI:2639 |
| eb_b034a_med3_name_CHILDQ | amiloride | CHEBI:2639 |
| eb_b033a_med2_name_CHILDQ | amiodarone | CHEBI:2663 |
| eb_b035a_med4_name_CHILDQ | amitiza | CHEBI:34945 |
| eb_b033a_med2_name_CHILDQ | amitriptylin | CHEBI:2666 |
| eb_b033a_med2_name_CHILDQ | amitriptyline | CHEBI:2666 |
| eb_b034a_med3_name_CHILDQ | amitriptyline | CHEBI:2666 |
| eb_b036a_med5_name_CHILDQ | amitriptyline | CHEBI:2666 |
| eb_b037a_med6_name_CHILDQ | amitriptyline | CHEBI:2666 |
| eb_b032a_med1_name_CHILDQ | amlodipine | CHEBI:2668 |
| eb_b033a_med2_name_CHILDQ | amlodipine | CHEBI:2668 |
| eb_b034a_med3_name_CHILDQ | amlodipine | CHEBI:2668 |
| eb_b035a_med4_name_CHILDQ | amlodipine | CHEBI:2668 |
| eb_b036a_med5_name_CHILDQ | amlodipine | CHEBI:2668 |
| eb_b037a_med6_name_CHILDQ | amlodipine | CHEBI:2668 |
| eb_b038a_med7_name_CHILDQ | amlodipine | CHEBI:2668 |
| eb_b039a_med8_name_CHILDQ | amlodipine | CHEBI:2668 |
| eb_b040a_med9_name_CHILDQ | amlodipine | CHEBI:2668 |
| eb_b032a_med1_name_CHILDQ | amlodipine besylate | CHEBI:2669 |
| eb_b033a_med2_name_CHILDQ | amlodipine besylate | CHEBI:2669 |
| eb_b034a_med3_name_CHILDQ | amlodipine besylate | CHEBI:2669 |
| eb_b035a_med4_name_CHILDQ | amlodipine besylate | CHEBI:2669 |
| eb_b036a_med5_name_CHILDQ | amlodipine besylate | CHEBI:2669 |
| eb_b037a_med6_name_CHILDQ | amlodipine besylate | CHEBI:2669 |
| eb_b039a_med8_name_CHILDQ | amlodipine besylate | CHEBI:2669 |
| eb_b033a_med2_name_CHILDQ | ammonium | CHEBI:28938 |
| eb_b032a_med1_name_CHILDQ | amoxicillin | CHEBI:2676 |
| eb_b035a_med4_name_CHILDQ | amoxicillin | CHEBI:2676 |
| eb_b040a_med9_name_CHILDQ | amoxicillin | CHEBI:2676 |
| eb_b035a_med4_name_CHILDQ | amphetamin | CHEBI:2679 |
| eb_b032a_med1_name_CHILDQ | amphetamine | CHEBI:2679 |
| eb_b032a_med1_name_CHILDQ | amphetamine | CHEBI:2679 |
| eb_b033a_med2_name_CHILDQ | amphetamine | CHEBI:2679 |
| eb_b033a_med2_name_CHILDQ | amphetamine | CHEBI:2679 |
| eb_b037a_med6_name_CHILDQ | amphetamine | CHEBI:2679 |
| eb_b037a_med6_name_CHILDQ | amphetamine | CHEBI:2679 |
| eb_b032a_med1_name_CHILDQ | ampyra | CHEBI:34385 |
| eb_b032a_med1_name_CHILDQ | anastrozole | CHEBI:2704 |
| eb_b033a_med2_name_CHILDQ | anastrozole | CHEBI:2704 |
| eb_b034a_med3_name_CHILDQ | anastrozole | CHEBI:2704 |
| eb_b035a_med4_name_CHILDQ | anastrozole | CHEBI:2704 |
| eb_b040a_med9_name_CHILDQ | anastrozole | CHEBI:2704 |
| eb_b035a_med4_name_CHILDQ | antacid | CHEBI:65265 |
| eb_b035a_med4_name_CHILDQ | antihistamine | CHEBI:37956 |
| eb_b035a_med4_name_CHILDQ | apap | CHEBI:46195 |
| eb_b036a_med5_name_CHILDQ | apap | CHEBI:46195 |
| eb_b032a_med1_name_CHILDQ | apixaban | CHEBI:72296 |
| eb_b040a_med9_name_CHILDQ | armodafinil | CHEBI:77590 |
| eb_b032a_med1_name_CHILDQ | armour thyroid | CHEBI:9584 |
| eb_b033a_med2_name_CHILDQ | armour thyroid | CHEBI:9584 |
| eb_b034a_med3_name_CHILDQ | armour thyroid | CHEBI:9584 |
| eb_b035a_med4_name_CHILDQ | armour thyroid | CHEBI:9584 |
| eb_b036a_med5_name_CHILDQ | armour thyroid | CHEBI:9584 |
| eb_b032a_med1_name_CHILDQ | arthrotec | CHEBI:63610 |
| eb_b032a_med1_name_CHILDQ | asa | CHEBI:15365 |
| eb_b033a_med2_name_CHILDQ | asa | CHEBI:15365 |
| eb_b034a_med3_name_CHILDQ | asa | CHEBI:15365 |
| eb_b035a_med4_name_CHILDQ | asa | CHEBI:15365 |
| eb_b036a_med5_name_CHILDQ | asa | CHEBI:15365 |
| eb_b037a_med6_name_CHILDQ | asa | CHEBI:15365 |
| eb_b038a_med7_name_CHILDQ | asa | CHEBI:15365 |
| eb_b032a_med1_name_CHILDQ | asacol | CHEBI:6775 |
| eb_b036a_med5_name_CHILDQ | asacol | CHEBI:6775 |
| eb_b037a_med6_name_CHILDQ | ascorbic acid | CHEBI:22652 |
| eb_b037a_med6_name_CHILDQ | ascorbic acid | CHEBI:22652 |
| eb_b032a_med1_name_CHILDQ | aspirin | CHEBI:15365 |
| eb_b033a_med2_name_CHILDQ | aspirin | CHEBI:15365 |
| eb_b034a_med3_name_CHILDQ | aspirin | CHEBI:15365 |
| eb_b035a_med4_name_CHILDQ | aspirin | CHEBI:15365 |
| eb_b036a_med5_name_CHILDQ | aspirin | CHEBI:15365 |
| eb_b037a_med6_name_CHILDQ | aspirin | CHEBI:15365 |
| eb_b038a_med7_name_CHILDQ | aspirin | CHEBI:15365 |
| eb_b039a_med8_name_CHILDQ | aspirin | CHEBI:15365 |
| eb_b040a_med9_name_CHILDQ | aspirin | CHEBI:15365 |
| eb_b041a_med10_name_CHILDQ | aspirin | CHEBI:15365 |
| eb_b041a_med10_name_CHILDQ | atarax | CHEBI:5819 |
| eb_b032a_med1_name_CHILDQ | atenolol | CHEBI:2904 |
| eb_b033a_med2_name_CHILDQ | atenolol | CHEBI:2904 |
| eb_b034a_med3_name_CHILDQ | atenolol | CHEBI:2904 |
| eb_b035a_med4_name_CHILDQ | atenolol | CHEBI:2904 |
| eb_b036a_med5_name_CHILDQ | atenolol | CHEBI:2904 |
| eb_b032a_med1_name_CHILDQ | ativan | CHEBI:6539 |
| eb_b033a_med2_name_CHILDQ | ativan | CHEBI:6539 |
| eb_b036a_med5_name_CHILDQ | ativan | CHEBI:6539 |
| eb_b037a_med6_name_CHILDQ | atomoxetine | CHEBI:127342 |
| eb_b032a_med1_name_CHILDQ | atorvastatin | CHEBI:39548 |
| eb_b033a_med2_name_CHILDQ | atorvastatin | CHEBI:39548 |
| eb_b034a_med3_name_CHILDQ | atorvastatin | CHEBI:39548 |
| eb_b035a_med4_name_CHILDQ | atorvastatin | CHEBI:39548 |
| eb_b036a_med5_name_CHILDQ | atorvastatin | CHEBI:39548 |
| eb_b037a_med6_name_CHILDQ | atorvastatin | CHEBI:39548 |
| eb_b038a_med7_name_CHILDQ | atorvastatin | CHEBI:39548 |
| eb_b039a_med8_name_CHILDQ | atorvastatin | CHEBI:39548 |
| eb_b040a_med9_name_CHILDQ | atorvastatin | CHEBI:39548 |
| eb_b041a_med10_name_CHILDQ | atorvastatin | CHEBI:39548 |
| eb_b032a_med1_name_CHILDQ | atorvastatin calcium | CHEBI:50686 |
| eb_b033a_med2_name_CHILDQ | atorvastatin calcium | CHEBI:50686 |
| eb_b034a_med3_name_CHILDQ | atorvastatin calcium | CHEBI:50686 |
| eb_b035a_med4_name_CHILDQ | atorvastatin calcium | CHEBI:50686 |
| eb_b036a_med5_name_CHILDQ | atorvastatin calcium | CHEBI:50686 |
| eb_b037a_med6_name_CHILDQ | atorvastatin calcium | CHEBI:50686 |
| eb_b038a_med7_name_CHILDQ | atorvastatin calcium | CHEBI:50686 |
| eb_b039a_med8_name_CHILDQ | atorvastatin calcium | CHEBI:50686 |
| eb_b032a_med1_name_CHILDQ | avapro | CHEBI:5959 |
| eb_b032a_med1_name_CHILDQ | azathioprine | CHEBI:2948 |
| eb_b033a_med2_name_CHILDQ | azathioprine | CHEBI:2948 |
| eb_b032a_med1_name_CHILDQ | azelastine | CHEBI:2950 |
| eb_b033a_med2_name_CHILDQ | azelastine | CHEBI:2950 |
| eb_b034a_med3_name_CHILDQ | azelastine | CHEBI:2950 |
| eb_b035a_med4_name_CHILDQ | azelastine | CHEBI:2950 |
| eb_b036a_med5_name_CHILDQ | azelastine | CHEBI:2950 |
| eb_b037a_med6_name_CHILDQ | azelastine | CHEBI:2950 |
| eb_b038a_med7_name_CHILDQ | azelastine | CHEBI:2950 |
| eb_b040a_med9_name_CHILDQ | azelastine | CHEBI:2950 |
| eb_b034a_med3_name_CHILDQ | azelastine hcl | CHEBI:2951 |
| eb_b035a_med4_name_CHILDQ | azelastine hcl | CHEBI:2951 |
| eb_b036a_med5_name_CHILDQ | azelastine hcl | CHEBI:2951 |
| eb_b037a_med6_name_CHILDQ | azelastine hcl | CHEBI:2951 |
| eb_b033a_med2_name_CHILDQ | azithromycin | CHEBI:2955 |
| eb_b036a_med5_name_CHILDQ | azopt | CHEBI:3176 |
| eb_b037a_med6_name_CHILDQ | azythromycin | CHEBI:176304 |
| eb_b032a_med1_name_CHILDQ | b vitamin | CHEBI:75769 |
| eb_b032a_med1_name_CHILDQ | baclofen | CHEBI:2972 |
| eb_b033a_med2_name_CHILDQ | baclofen | CHEBI:2972 |
| eb_b035a_med4_name_CHILDQ | baclofen | CHEBI:2972 |
| eb_b036a_med5_name_CHILDQ | baclofen | CHEBI:2972 |
| eb_b039a_med8_name_CHILDQ | baclofen | CHEBI:2972 |
| eb_b041a_med10_name_CHILDQ | baclofen | CHEBI:2972 |
| eb_b033a_med2_name_CHILDQ | beclometasone | CHEBI:3001 |
| eb_b032a_med1_name_CHILDQ | belviq | CHEBI:66852 |
| eb_b032a_med1_name_CHILDQ | benazepril | CHEBI:3011 |
| eb_b033a_med2_name_CHILDQ | benazepril | CHEBI:3011 |
| eb_b034a_med3_name_CHILDQ | benazepril | CHEBI:3011 |
| eb_b035a_med4_name_CHILDQ | benazepril | CHEBI:3011 |
| eb_b036a_med5_name_CHILDQ | benazepril | CHEBI:3011 |
| eb_b037a_med6_name_CHILDQ | benazepril | CHEBI:3011 |
| eb_b040a_med9_name_CHILDQ | benazepril | CHEBI:3011 |
| eb_b037a_med6_name_CHILDQ | benazepril hcl | CHEBI:3012 |
| eb_b034a_med3_name_CHILDQ | benzalkonium | CHEBI:188978 |
| eb_b034a_med3_name_CHILDQ | benzonatate | CHEBI:3032 |
| eb_b038a_med7_name_CHILDQ | benzonatate | CHEBI:3032 |
| eb_b035a_med4_name_CHILDQ | benzoyl | CHEBI:22733 |
| eb_b035a_med4_name_CHILDQ | benzoyl peroxide | CHEBI:82405 |
| eb_b035a_med4_name_CHILDQ | bepreve | CHEBI:31281 |
| eb_b032a_med1_name_CHILDQ | besylate | CHEBI:64457 |
| eb_b033a_med2_name_CHILDQ | besylate | CHEBI:64457 |
| eb_b034a_med3_name_CHILDQ | besylate | CHEBI:64457 |
| eb_b035a_med4_name_CHILDQ | besylate | CHEBI:64457 |
| eb_b036a_med5_name_CHILDQ | besylate | CHEBI:64457 |
| eb_b037a_med6_name_CHILDQ | besylate | CHEBI:64457 |
| eb_b039a_med8_name_CHILDQ | besylate | CHEBI:64457 |
| eb_b034a_med3_name_CHILDQ | beta | CHEBI:10545 |
| eb_b033a_med2_name_CHILDQ | betamethasone | CHEBI:3077 |
| eb_b033a_med2_name_CHILDQ | betamethasone dipropionate | CHEBI:31276 |
| eb_b036a_med5_name_CHILDQ | bicarbonate | CHEBI:17544 |
| eb_b032a_med1_name_CHILDQ | biotin | CHEBI:15956 |
| eb_b033a_med2_name_CHILDQ | biotin | CHEBI:15956 |
| eb_b034a_med3_name_CHILDQ | biotin | CHEBI:15956 |
| eb_b035a_med4_name_CHILDQ | biotin | CHEBI:15956 |
| eb_b036a_med5_name_CHILDQ | biotin | CHEBI:15956 |
| eb_b037a_med6_name_CHILDQ | biotin | CHEBI:15956 |
| eb_b038a_med7_name_CHILDQ | biotin | CHEBI:15956 |
| eb_b039a_med8_name_CHILDQ | biotin | CHEBI:15956 |
| eb_b040a_med9_name_CHILDQ | biotin | CHEBI:15956 |
| eb_b041a_med10_name_CHILDQ | biotin | CHEBI:15956 |
| eb_b032a_med1_name_CHILDQ | bisoprolol | CHEBI:3127 |
| eb_b033a_med2_name_CHILDQ | bisoprolol | CHEBI:3127 |
| eb_b035a_med4_name_CHILDQ | bisoprolol | CHEBI:3127 |
| eb_b033a_med2_name_CHILDQ | bisoprolol fumarate | CHEBI:3128 |
| eb_b032a_med1_name_CHILDQ | breo ellipta | CHEBI:75043 |
| eb_b035a_med4_name_CHILDQ | breo ellipta | CHEBI:75043 |
| eb_b032a_med1_name_CHILDQ | brilinta | CHEBI:68558 |
| eb_b037a_med6_name_CHILDQ | brilinta | CHEBI:68558 |
| eb_b033a_med2_name_CHILDQ | brimonidine | CHEBI:3175 |
| eb_b036a_med5_name_CHILDQ | brimonidine | CHEBI:3175 |
| eb_b036a_med5_name_CHILDQ | bromide | CHEBI:15858 |
| eb_b039a_med8_name_CHILDQ | bromide | CHEBI:15858 |
| eb_b032a_med1_name_CHILDQ | bromocriptine | CHEBI:3181 |
| eb_b032a_med1_name_CHILDQ | budesonide | CHEBI:3207 |
| eb_b035a_med4_name_CHILDQ | budesonide | CHEBI:3207 |
| eb_b033a_med2_name_CHILDQ | bumetanide | CHEBI:3213 |
| eb_b035a_med4_name_CHILDQ | bumetanide | CHEBI:3213 |
| eb_b032a_med1_name_CHILDQ | bupropion | CHEBI:3219 |
| eb_b033a_med2_name_CHILDQ | bupropion | CHEBI:3219 |
| eb_b034a_med3_name_CHILDQ | bupropion | CHEBI:3219 |
| eb_b035a_med4_name_CHILDQ | bupropion | CHEBI:3219 |
| eb_b036a_med5_name_CHILDQ | bupropion | CHEBI:3219 |
| eb_b037a_med6_name_CHILDQ | bupropion | CHEBI:3219 |
| eb_b038a_med7_name_CHILDQ | bupropion | CHEBI:3219 |
| eb_b039a_med8_name_CHILDQ | bupropion | CHEBI:3219 |
| eb_b033a_med2_name_CHILDQ | buspar | CHEBI:3224 |
| eb_b034a_med3_name_CHILDQ | buspar | CHEBI:3224 |
| eb_b032a_med1_name_CHILDQ | buspirone | CHEBI:3223 |
| eb_b033a_med2_name_CHILDQ | buspirone | CHEBI:3223 |
| eb_b035a_med4_name_CHILDQ | buspirone | CHEBI:3223 |
| eb_b036a_med5_name_CHILDQ | buspirone | CHEBI:3223 |
| eb_b041a_med10_name_CHILDQ | buspirone | CHEBI:3223 |
| eb_b032a_med1_name_CHILDQ | cabergoline | CHEBI:3286 |
| eb_b034a_med3_name_CHILDQ | cabergoline | CHEBI:3286 |
| eb_b033a_med2_name_CHILDQ | caffeine | CHEBI:27732 |
| eb_b036a_med5_name_CHILDQ | calciferol | CHEBI:28934 |
| eb_b032a_med1_name_CHILDQ | calcitonin | CHEBI:3306 |
| eb_b037a_med6_name_CHILDQ | calcitonin | CHEBI:3306 |
| eb_b032a_med1_name_CHILDQ | calcitriol | CHEBI:17823 |
| eb_b032a_med1_name_CHILDQ | calcium | CHEBI:22984 |
| eb_b033a_med2_name_CHILDQ | calcium | CHEBI:22984 |
| eb_b034a_med3_name_CHILDQ | calcium | CHEBI:22984 |
| eb_b035a_med4_name_CHILDQ | calcium | CHEBI:22984 |
| eb_b036a_med5_name_CHILDQ | calcium | CHEBI:22984 |
| eb_b037a_med6_name_CHILDQ | calcium | CHEBI:22984 |
| eb_b038a_med7_name_CHILDQ | calcium | CHEBI:22984 |
| eb_b039a_med8_name_CHILDQ | calcium | CHEBI:22984 |
| eb_b041a_med10_name_CHILDQ | calcium | CHEBI:22984 |
| eb_b034a_med3_name_CHILDQ | calcium carbonate | CHEBI:3311 |
| eb_b036a_med5_name_CHILDQ | calcium carbonate | CHEBI:3311 |
| eb_b035a_med4_name_CHILDQ | calcium citrate | CHEBI:190513 |
| eb_b035a_med4_name_CHILDQ | canagliflozin | CHEBI:73274 |
| eb_b035a_med4_name_CHILDQ | canagliflozin | CHEBI:73272 |
| eb_b033a_med2_name_CHILDQ | canasa | CHEBI:6775 |
| eb_b033a_med2_name_CHILDQ | carbamazepine | CHEBI:3387 |
| eb_b033a_med2_name_CHILDQ | carbidopa | CHEBI:3395 |
| eb_b033a_med2_name_CHILDQ | carbidopa-levodopa | CHEBI:3396 |
| eb_b032a_med1_name_CHILDQ | carboxymethylcellulose | CHEBI:85146 |
| eb_b038a_med7_name_CHILDQ | carboxymethylcellulose | CHEBI:85146 |
| eb_b034a_med3_name_CHILDQ | carfilzomib | CHEBI:65347 |
| eb_b032a_med1_name_CHILDQ | carisoprodol | CHEBI:3419 |
| eb_b039a_med8_name_CHILDQ | carnitine | CHEBI:17126 |
| eb_b039a_med8_name_CHILDQ | carnitine | CHEBI:3424 |
| eb_b032a_med1_name_CHILDQ | carvedilol | CHEBI:3441 |
| eb_b033a_med2_name_CHILDQ | carvedilol | CHEBI:3441 |
| eb_b034a_med3_name_CHILDQ | carvedilol | CHEBI:3441 |
| eb_b035a_med4_name_CHILDQ | carvedilol | CHEBI:3441 |
| eb_b037a_med6_name_CHILDQ | carvedilol | CHEBI:3441 |
| eb_b034a_med3_name_CHILDQ | cefuroxime | CHEBI:3515 |
| eb_b032a_med1_name_CHILDQ | celebrex | CHEBI:41423 |
| eb_b033a_med2_name_CHILDQ | celebrex | CHEBI:41423 |
| eb_b034a_med3_name_CHILDQ | celebrex | CHEBI:41423 |
| eb_b035a_med4_name_CHILDQ | celebrex | CHEBI:41423 |
| eb_b036a_med5_name_CHILDQ | celebrex | CHEBI:41423 |
| eb_b040a_med9_name_CHILDQ | celebrex | CHEBI:41423 |
| eb_b032a_med1_name_CHILDQ | celecoxib | CHEBI:41423 |
| eb_b033a_med2_name_CHILDQ | celecoxib | CHEBI:41423 |
| eb_b036a_med5_name_CHILDQ | celecoxib | CHEBI:41423 |
| eb_b038a_med7_name_CHILDQ | celecoxib | CHEBI:41423 |
| eb_b039a_med8_name_CHILDQ | celecoxib | CHEBI:41423 |
| eb_b032a_med1_name_CHILDQ | celexa | CHEBI:3724 |
| eb_b033a_med2_name_CHILDQ | celexa | CHEBI:3724 |
| eb_b034a_med3_name_CHILDQ | celexa | CHEBI:3724 |
| eb_b035a_med4_name_CHILDQ | celexa | CHEBI:3724 |
| eb_b036a_med5_name_CHILDQ | celexa | CHEBI:3724 |
| eb_b040a_med9_name_CHILDQ | celexa | CHEBI:3724 |
| eb_b033a_med2_name_CHILDQ | cellcept | CHEBI:8764 |
| eb_b034a_med3_name_CHILDQ | cellcept | CHEBI:8764 |
| eb_b032a_med1_name_CHILDQ | cephalexin | CHEBI:3534 |
| eb_b041a_med10_name_CHILDQ | cephalexin | CHEBI:3534 |
| eb_b032a_med1_name_CHILDQ | cetirizine | CHEBI:3561 |
| eb_b033a_med2_name_CHILDQ | cetirizine | CHEBI:3561 |
| eb_b034a_med3_name_CHILDQ | cetirizine | CHEBI:3561 |
| eb_b035a_med4_name_CHILDQ | cetirizine | CHEBI:3561 |
| eb_b036a_med5_name_CHILDQ | cetirizine | CHEBI:3561 |
| eb_b037a_med6_name_CHILDQ | cetirizine | CHEBI:3561 |
| eb_b038a_med7_name_CHILDQ | cetirizine | CHEBI:3561 |
| eb_b040a_med9_name_CHILDQ | cetirizine | CHEBI:3561 |
| eb_b032a_med1_name_CHILDQ | cetirizine hydrochloride | CHEBI:3562 |
| eb_b033a_med2_name_CHILDQ | cetirizine hydrochloride | CHEBI:3562 |
| eb_b037a_med6_name_CHILDQ | cetirizine hydrochloride | CHEBI:3562 |
| eb_b033a_med2_name_CHILDQ | cevimeline | CHEBI:3568 |
| eb_b034a_med3_name_CHILDQ | chlorpheniramine | CHEBI:52010 |
| eb_b037a_med6_name_CHILDQ | chlorpheniramine | CHEBI:52010 |
| eb_b038a_med7_name_CHILDQ | chlorpheniramine | CHEBI:52010 |
| eb_b038a_med7_name_CHILDQ | chlorpheniramine maleate | CHEBI:3645 |
| eb_b033a_med2_name_CHILDQ | chlorthalidone | CHEBI:3654 |
| eb_b035a_med4_name_CHILDQ | chlorthalidone | CHEBI:3654 |
| eb_b037a_med6_name_CHILDQ | chlorthalidone | CHEBI:3654 |
| eb_b039a_med8_name_CHILDQ | chlorthalidone | CHEBI:3654 |
| eb_b036a_med5_name_CHILDQ | cholecalciferol | CHEBI:28940 |
| eb_b032a_med1_name_CHILDQ | cholesterol | CHEBI:16113 |
| eb_b032a_med1_name_CHILDQ | chondroitin | CHEBI:16137 |
| eb_b035a_med4_name_CHILDQ | chondroitin | CHEBI:16137 |
| eb_b036a_med5_name_CHILDQ | chondroitin | CHEBI:16137 |
| eb_b037a_med6_name_CHILDQ | chondroitin | CHEBI:16137 |
| eb_b038a_med7_name_CHILDQ | chondroitin | CHEBI:16137 |
| eb_b037a_med6_name_CHILDQ | chondroitin sulfate | CHEBI:37397 |
| eb_b032a_med1_name_CHILDQ | cialis | CHEBI:71940 |
| eb_b033a_med2_name_CHILDQ | cialis | CHEBI:71940 |
| eb_b034a_med3_name_CHILDQ | cialis | CHEBI:71940 |
| eb_b037a_med6_name_CHILDQ | cialis | CHEBI:71940 |
| eb_b040a_med9_name_CHILDQ | cialis | CHEBI:71940 |
| eb_b035a_med4_name_CHILDQ | cilostazol | CHEBI:31401 |
| eb_b032a_med1_name_CHILDQ | citalopram | CHEBI:3723 |
| eb_b033a_med2_name_CHILDQ | citalopram | CHEBI:3723 |
| eb_b034a_med3_name_CHILDQ | citalopram | CHEBI:3723 |
| eb_b035a_med4_name_CHILDQ | citalopram | CHEBI:3723 |
| eb_b036a_med5_name_CHILDQ | citalopram | CHEBI:3723 |
| eb_b037a_med6_name_CHILDQ | citalopram | CHEBI:3723 |
| eb_b038a_med7_name_CHILDQ | citalopram | CHEBI:3723 |
| eb_b039a_med8_name_CHILDQ | citalopram | CHEBI:3723 |
| eb_b033a_med2_name_CHILDQ | citalopram hydrobromide | CHEBI:3724 |
| eb_b034a_med3_name_CHILDQ | citalopram hydrobromide | CHEBI:3724 |
| eb_b036a_med5_name_CHILDQ | citalopram hydrobromide | CHEBI:3724 |
| eb_b032a_med1_name_CHILDQ | claritin | CHEBI:6538 |
| eb_b033a_med2_name_CHILDQ | claritin | CHEBI:6538 |
| eb_b034a_med3_name_CHILDQ | claritin | CHEBI:6538 |
| eb_b035a_med4_name_CHILDQ | claritin | CHEBI:6538 |
| eb_b036a_med5_name_CHILDQ | claritin | CHEBI:6538 |
| eb_b037a_med6_name_CHILDQ | claritin | CHEBI:6538 |
| eb_b040a_med9_name_CHILDQ | claritin | CHEBI:6538 |
| eb_b041a_med10_name_CHILDQ | claritin | CHEBI:6538 |
| eb_b032a_med1_name_CHILDQ | clindamycin | CHEBI:3745 |
| eb_b035a_med4_name_CHILDQ | clindamycin | CHEBI:3745 |
| eb_b036a_med5_name_CHILDQ | clindamycin | CHEBI:3745 |
| eb_b037a_med6_name_CHILDQ | clindamycin | CHEBI:3745 |
| eb_b036a_med5_name_CHILDQ | clindamycin phosphate | CHEBI:3746 |
| eb_b032a_med1_name_CHILDQ | clobetasol | CHEBI:205919 |
| eb_b033a_med2_name_CHILDQ | clobetasol | CHEBI:205919 |
| eb_b035a_med4_name_CHILDQ | clobetasol | CHEBI:205919 |
| eb_b036a_med5_name_CHILDQ | clobetasol | CHEBI:205919 |
| eb_b032a_med1_name_CHILDQ | clobetasol propionate | CHEBI:31414 |
| eb_b033a_med2_name_CHILDQ | clobetasol propionate | CHEBI:31414 |
| eb_b035a_med4_name_CHILDQ | clobetasol propionate | CHEBI:31414 |
| eb_b036a_med5_name_CHILDQ | clobetasol propionate | CHEBI:31414 |
| eb_b032a_med1_name_CHILDQ | clonazepam | CHEBI:3756 |
| eb_b033a_med2_name_CHILDQ | clonazepam | CHEBI:3756 |
| eb_b034a_med3_name_CHILDQ | clonazepam | CHEBI:3756 |
| eb_b035a_med4_name_CHILDQ | clonazepam | CHEBI:3756 |
| eb_b036a_med5_name_CHILDQ | clonazepam | CHEBI:3756 |
| eb_b037a_med6_name_CHILDQ | clonazepam | CHEBI:3756 |
| eb_b038a_med7_name_CHILDQ | clonazepam | CHEBI:3756 |
| eb_b039a_med8_name_CHILDQ | clonazepam | CHEBI:3756 |
| eb_b033a_med2_name_CHILDQ | clonidine | CHEBI:46631 |
| eb_b035a_med4_name_CHILDQ | clonidine | CHEBI:46631 |
| eb_b036a_med5_name_CHILDQ | clonidine | CHEBI:46631 |
| eb_b037a_med6_name_CHILDQ | clonidine | CHEBI:46631 |
| eb_b032a_med1_name_CHILDQ | clopidogrel | CHEBI:37941 |
| eb_b033a_med2_name_CHILDQ | clopidogrel | CHEBI:37941 |
| eb_b034a_med3_name_CHILDQ | clopidogrel | CHEBI:37941 |
| eb_b035a_med4_name_CHILDQ | clopidogrel | CHEBI:37941 |
| eb_b036a_med5_name_CHILDQ | clopidogrel | CHEBI:37941 |
| eb_b037a_med6_name_CHILDQ | clopidogrel | CHEBI:37941 |
| eb_b036a_med5_name_CHILDQ | clopidogrel bisulfate | CHEBI:3759 |
| eb_b033a_med2_name_CHILDQ | clotrimazole | CHEBI:3764 |
| eb_b038a_med7_name_CHILDQ | clotrimazole | CHEBI:3764 |
| eb_b032a_med1_name_CHILDQ | codeine | CHEBI:16714 |
| eb_b032a_med1_name_CHILDQ | codeine | CHEBI:57871 |
| eb_b038a_med7_name_CHILDQ | codeine | CHEBI:16714 |
| eb_b038a_med7_name_CHILDQ | codeine | CHEBI:57871 |
| eb_b033a_med2_name_CHILDQ | colchicine | CHEBI:23359 |
| eb_b034a_med3_name_CHILDQ | colchicine | CHEBI:23359 |
| eb_b039a_med8_name_CHILDQ | colchicine | CHEBI:23359 |
| eb_b035a_med4_name_CHILDQ | colestipol | CHEBI:3814 |
| eb_b035a_med4_name_CHILDQ | colistimethate | CHEBI:59662 |
| eb_b035a_med4_name_CHILDQ | colistimethate | CHEBI:34650 |
| eb_b032a_med1_name_CHILDQ | concerta | CHEBI:31836 |
| eb_b033a_med2_name_CHILDQ | concerta | CHEBI:31836 |
| eb_b034a_med3_name_CHILDQ | concerta | CHEBI:31836 |
| eb_b038a_med7_name_CHILDQ | concerta | CHEBI:31836 |
| eb_b034a_med3_name_CHILDQ | coq10 | CHEBI:46245 |
| eb_b035a_med4_name_CHILDQ | coq10 | CHEBI:46245 |
| eb_b038a_med7_name_CHILDQ | coq10 | CHEBI:46245 |
| eb_b033a_med2_name_CHILDQ | coumadin | CHEBI:10034 |
| eb_b035a_med4_name_CHILDQ | coumadin | CHEBI:10034 |
| eb_b036a_med5_name_CHILDQ | coumadin | CHEBI:10034 |
| eb_b034a_med3_name_CHILDQ | creatinine | CHEBI:16737 |
| eb_b032a_med1_name_CHILDQ | crestor | CHEBI:77249 |
| eb_b033a_med2_name_CHILDQ | crestor | CHEBI:77249 |
| eb_b034a_med3_name_CHILDQ | crestor | CHEBI:77249 |
| eb_b035a_med4_name_CHILDQ | crestor | CHEBI:77249 |
| eb_b036a_med5_name_CHILDQ | crestor | CHEBI:77249 |
| eb_b037a_med6_name_CHILDQ | crestor | CHEBI:77249 |
| eb_b038a_med7_name_CHILDQ | crestor | CHEBI:77249 |
| eb_b039a_med8_name_CHILDQ | crestor | CHEBI:77249 |
| eb_b040a_med9_name_CHILDQ | crestor | CHEBI:77249 |
| eb_b033a_med2_name_CHILDQ | curcumin | CHEBI:3962 |
| eb_b035a_med4_name_CHILDQ | curcumin | CHEBI:3962 |
| eb_b039a_med8_name_CHILDQ | curcumin | CHEBI:3962 |
| eb_b034a_med3_name_CHILDQ | cyanocobalamin | CHEBI:17439 |
| eb_b035a_med4_name_CHILDQ | cyanocobalamin | CHEBI:17439 |
| eb_b036a_med5_name_CHILDQ | cyanocobalamin | CHEBI:17439 |
| eb_b041a_med10_name_CHILDQ | cyanocobalamin | CHEBI:17439 |
| eb_b032a_med1_name_CHILDQ | cyclen | CHEBI:37391 |
| eb_b033a_med2_name_CHILDQ | cyclobenzaprine | CHEBI:3996 |
| eb_b034a_med3_name_CHILDQ | cyclobenzaprine | CHEBI:3996 |
| eb_b035a_med4_name_CHILDQ | cyclobenzaprine | CHEBI:3996 |
| eb_b036a_med5_name_CHILDQ | cyclobenzaprine | CHEBI:3996 |
| eb_b037a_med6_name_CHILDQ | cyclobenzaprine | CHEBI:3996 |
| eb_b038a_med7_name_CHILDQ | cyclobenzaprine | CHEBI:3996 |
| eb_b041a_med10_name_CHILDQ | cyclobenzaprine | CHEBI:3996 |
| eb_b039a_med8_name_CHILDQ | cyclosporine | CHEBI:4031 |
| eb_b032a_med1_name_CHILDQ | cymbalta | CHEBI:31526 |
| eb_b033a_med2_name_CHILDQ | cymbalta | CHEBI:31526 |
| eb_b034a_med3_name_CHILDQ | cymbalta | CHEBI:31526 |
| eb_b035a_med4_name_CHILDQ | cymbalta | CHEBI:31526 |
| eb_b036a_med5_name_CHILDQ | cymbalta | CHEBI:31526 |
| eb_b037a_med6_name_CHILDQ | cymbalta | CHEBI:31526 |
| eb_b038a_med7_name_CHILDQ | cysteine | CHEBI:15356 |
| eb_b033a_med2_name_CHILDQ | cytomel | CHEBI:6484 |
| eb_b034a_med3_name_CHILDQ | cytomel | CHEBI:6484 |
| eb_b034a_med3_name_CHILDQ | ddavp | CHEBI:4450 |
| eb_b032a_med1_name_CHILDQ | decongestant | CHEBI:77715 |
| eb_b035a_med4_name_CHILDQ | dehydroepiandrosterone | CHEBI:28689 |
| eb_b032a_med1_name_CHILDQ | depo-provera | CHEBI:6716 |
| eb_b032a_med1_name_CHILDQ | descovy | CHEBI:133007 |
| eb_b033a_med2_name_CHILDQ | desloratadine | CHEBI:291342 |
| eb_b032a_med1_name_CHILDQ | desmopressin | CHEBI:4450 |
| eb_b032a_med1_name_CHILDQ | desonide | CHEBI:204734 |
| eb_b033a_med2_name_CHILDQ | dexamethasone | CHEBI:41879 |
| eb_b032a_med1_name_CHILDQ | dexilant | CHEBI:135931 |
| eb_b033a_med2_name_CHILDQ | dexilant | CHEBI:135931 |
| eb_b035a_med4_name_CHILDQ | dexilant | CHEBI:135931 |
| eb_b036a_med5_name_CHILDQ | dexilant | CHEBI:135931 |
| eb_b037a_med6_name_CHILDQ | dexilant | CHEBI:135931 |
| eb_b038a_med7_name_CHILDQ | dexilant | CHEBI:135931 |
| eb_b041a_med10_name_CHILDQ | dexilant | CHEBI:135931 |
| eb_b033a_med2_name_CHILDQ | dexmethylphenidate | CHEBI:51860 |
| eb_b032a_med1_name_CHILDQ | diazepam | CHEBI:49575 |
| eb_b033a_med2_name_CHILDQ | diazepam | CHEBI:49575 |
| eb_b034a_med3_name_CHILDQ | diazepam | CHEBI:49575 |
| eb_b036a_med5_name_CHILDQ | diazepam | CHEBI:49575 |
| eb_b032a_med1_name_CHILDQ | diclofenac | CHEBI:47381 |
| eb_b033a_med2_name_CHILDQ | diclofenac | CHEBI:47381 |
| eb_b034a_med3_name_CHILDQ | diclofenac | CHEBI:47381 |
| eb_b036a_med5_name_CHILDQ | diclofenac | CHEBI:47381 |
| eb_b040a_med9_name_CHILDQ | diclofenac | CHEBI:47381 |
| eb_b032a_med1_name_CHILDQ | diclofenac sodium | CHEBI:4509 |
| eb_b034a_med3_name_CHILDQ | diclofenac sodium | CHEBI:4509 |
| eb_b040a_med9_name_CHILDQ | diclofenac sodium | CHEBI:4509 |
| eb_b032a_med1_name_CHILDQ | dicyclomine | CHEBI:4514 |
| eb_b034a_med3_name_CHILDQ | dicyclomine | CHEBI:4514 |
| eb_b036a_med5_name_CHILDQ | dicyclomine | CHEBI:4514 |
| eb_b039a_med8_name_CHILDQ | dicyclomine | CHEBI:4514 |
| eb_b036a_med5_name_CHILDQ | diflucan | CHEBI:46081 |
| eb_b032a_med1_name_CHILDQ | digoxin | CHEBI:4551 |
| eb_b034a_med3_name_CHILDQ | digoxin | CHEBI:4551 |
| eb_b037a_med6_name_CHILDQ | digoxin | CHEBI:4551 |
| eb_b032a_med1_name_CHILDQ | dilantin | CHEBI:8107 |
| eb_b032a_med1_name_CHILDQ | diltiazem | CHEBI:101278 |
| eb_b033a_med2_name_CHILDQ | diltiazem | CHEBI:101278 |
| eb_b034a_med3_name_CHILDQ | diltiazem | CHEBI:101278 |
| eb_b035a_med4_name_CHILDQ | diltiazem | CHEBI:101278 |
| eb_b036a_med5_name_CHILDQ | diltiazem | CHEBI:101278 |
| eb_b037a_med6_name_CHILDQ | diltiazem | CHEBI:101278 |
| eb_b038a_med7_name_CHILDQ | diltiazem | CHEBI:101278 |
| eb_b039a_med8_name_CHILDQ | diltiazem | CHEBI:101278 |
| eb_b038a_med7_name_CHILDQ | diltiazem hydrochloride | CHEBI:645509 |
| eb_b032a_med1_name_CHILDQ | diovan | CHEBI:9927 |
| eb_b033a_med2_name_CHILDQ | diovan | CHEBI:9927 |
| eb_b034a_med3_name_CHILDQ | diovan | CHEBI:9927 |
| eb_b035a_med4_name_CHILDQ | diovan | CHEBI:9927 |
| eb_b032a_med1_name_CHILDQ | diphenhydramine | CHEBI:4636 |
| eb_b035a_med4_name_CHILDQ | diphenhydramine | CHEBI:4636 |
| eb_b036a_med5_name_CHILDQ | diphenhydramine | CHEBI:4636 |
| eb_b037a_med6_name_CHILDQ | diphenhydramine | CHEBI:4636 |
| eb_b035a_med4_name_CHILDQ | diphenhydramine hcl | CHEBI:4637 |
| eb_b036a_med5_name_CHILDQ | diphenhydramine hcl | CHEBI:4637 |
| eb_b035a_med4_name_CHILDQ | disulfiram | CHEBI:4659 |
| eb_b032a_med1_name_CHILDQ | docusate | CHEBI:4674 |
| eb_b033a_med2_name_CHILDQ | docusate | CHEBI:4674 |
| eb_b036a_med5_name_CHILDQ | docusate | CHEBI:4674 |
| eb_b037a_med6_name_CHILDQ | docusate | CHEBI:4674 |
| eb_b038a_med7_name_CHILDQ | docusate | CHEBI:4674 |
| eb_b041a_med10_name_CHILDQ | docusate | CHEBI:4674 |
| eb_b032a_med1_name_CHILDQ | docusate sodium | CHEBI:4674 |
| eb_b033a_med2_name_CHILDQ | docusate sodium | CHEBI:4674 |
| eb_b036a_med5_name_CHILDQ | docusate sodium | CHEBI:4674 |
| eb_b037a_med6_name_CHILDQ | docusate sodium | CHEBI:4674 |
| eb_b038a_med7_name_CHILDQ | docusate sodium | CHEBI:4674 |
| eb_b034a_med3_name_CHILDQ | donepezil | CHEBI:53289 |
| eb_b032a_med1_name_CHILDQ | dopa | CHEBI:49168 |
| eb_b032a_med1_name_CHILDQ | dorzolamide | CHEBI:4702 |
| eb_b034a_med3_name_CHILDQ | dorzolamide | CHEBI:4702 |
| eb_b032a_med1_name_CHILDQ | doxazosin | CHEBI:4708 |
| eb_b034a_med3_name_CHILDQ | doxazosin | CHEBI:4708 |
| eb_b035a_med4_name_CHILDQ | doxazosin | CHEBI:4708 |
| eb_b036a_med5_name_CHILDQ | doxazosin | CHEBI:4708 |
| eb_b032a_med1_name_CHILDQ | doxepin | CHEBI:4710 |
| eb_b035a_med4_name_CHILDQ | doxepin | CHEBI:4710 |
| eb_b037a_med6_name_CHILDQ | doxepin | CHEBI:4710 |
| eb_b032a_med1_name_CHILDQ | doxycycline | CHEBI:50845 |
| eb_b033a_med2_name_CHILDQ | doxycycline | CHEBI:50845 |
| eb_b035a_med4_name_CHILDQ | doxycycline | CHEBI:50845 |
| eb_b040a_med9_name_CHILDQ | doxycycline | CHEBI:50845 |
| eb_b032a_med1_name_CHILDQ | doxycycline hyclate | CHEBI:34730 |
| eb_b035a_med4_name_CHILDQ | drospirenone | CHEBI:50838 |
| eb_b038a_med7_name_CHILDQ | drug | CHEBI:23888 |
| eb_b032a_med1_name_CHILDQ | duloxetine | CHEBI:36796 |
| eb_b033a_med2_name_CHILDQ | duloxetine | CHEBI:36796 |
| eb_b034a_med3_name_CHILDQ | duloxetine | CHEBI:36796 |
| eb_b035a_med4_name_CHILDQ | duloxetine | CHEBI:36796 |
| eb_b036a_med5_name_CHILDQ | duloxetine | CHEBI:36796 |
| eb_b037a_med6_name_CHILDQ | duloxetine | CHEBI:36796 |
| eb_b038a_med7_name_CHILDQ | duloxetine | CHEBI:36796 |
| eb_b039a_med8_name_CHILDQ | duragesic | CHEBI:119915 |
| eb_b032a_med1_name_CHILDQ | dutasteride | CHEBI:521033 |
| eb_b034a_med3_name_CHILDQ | dutasteride | CHEBI:521033 |
| eb_b035a_med4_name_CHILDQ | dutasteride | CHEBI:521033 |
| eb_b036a_med5_name_CHILDQ | dutasteride | CHEBI:521033 |
| eb_b032a_med1_name_CHILDQ | effient | CHEBI:87697 |
| eb_b033a_med2_name_CHILDQ | effient | CHEBI:87697 |
| eb_b034a_med3_name_CHILDQ | effient | CHEBI:87697 |
| eb_b039a_med8_name_CHILDQ | effient | CHEBI:87697 |
| eb_b040a_med9_name_CHILDQ | effient | CHEBI:87697 |
| eb_b038a_med7_name_CHILDQ | elavil | CHEBI:2667 |
| eb_b032a_med1_name_CHILDQ | eliquis | CHEBI:72296 |
| eb_b033a_med2_name_CHILDQ | eliquis | CHEBI:72296 |
| eb_b034a_med3_name_CHILDQ | eliquis | CHEBI:72296 |
| eb_b035a_med4_name_CHILDQ | eliquis | CHEBI:72296 |
| eb_b038a_med7_name_CHILDQ | eliquis | CHEBI:72296 |
| eb_b032a_med1_name_CHILDQ | enalapril | CHEBI:4784 |
| eb_b033a_med2_name_CHILDQ | enalapril | CHEBI:4784 |
| eb_b034a_med3_name_CHILDQ | enalapril | CHEBI:4784 |
| eb_b035a_med4_name_CHILDQ | enalapril | CHEBI:4784 |
| eb_b036a_med5_name_CHILDQ | enalapril | CHEBI:4784 |
| eb_b038a_med7_name_CHILDQ | enalapril | CHEBI:4784 |
| eb_b033a_med2_name_CHILDQ | enalapril maleate | CHEBI:4785 |
| eb_b034a_med3_name_CHILDQ | enalapril maleate | CHEBI:4785 |
| eb_b038a_med7_name_CHILDQ | enalapril maleate | CHEBI:4785 |
| eb_b037a_med6_name_CHILDQ | epa | CHEBI:28364 |
| eb_b035a_med4_name_CHILDQ | epipen | CHEBI:28918 |
| eb_b032a_med1_name_CHILDQ | eprosartan | CHEBI:4814 |
| eb_b032a_med1_name_CHILDQ | eprosartan mesylate | CHEBI:48409 |
| eb_b032a_med1_name_CHILDQ | ergocalciferol | CHEBI:28934 |
| eb_b041a_med10_name_CHILDQ | ergocalciferol | CHEBI:28934 |
| eb_b032a_med1_name_CHILDQ | escitalopram | CHEBI:36791 |
| eb_b033a_med2_name_CHILDQ | escitalopram | CHEBI:36791 |
| eb_b034a_med3_name_CHILDQ | escitalopram | CHEBI:36791 |
| eb_b035a_med4_name_CHILDQ | escitalopram | CHEBI:36791 |
| eb_b036a_med5_name_CHILDQ | escitalopram | CHEBI:36791 |
| eb_b037a_med6_name_CHILDQ | escitalopram | CHEBI:36791 |
| eb_b032a_med1_name_CHILDQ | esomeprazol | CHEBI:50275 |
| eb_b032a_med1_name_CHILDQ | esomeprazole | CHEBI:50275 |
| eb_b033a_med2_name_CHILDQ | esomeprazole | CHEBI:50275 |
| eb_b034a_med3_name_CHILDQ | esomeprazole | CHEBI:50275 |
| eb_b035a_med4_name_CHILDQ | esomeprazole | CHEBI:50275 |
| eb_b032a_med1_name_CHILDQ | esomeprazole magnesium | CHEBI:50309 |
| eb_b032a_med1_name_CHILDQ | estradiol | CHEBI:23965 |
| eb_b033a_med2_name_CHILDQ | estradiol | CHEBI:23965 |
| eb_b034a_med3_name_CHILDQ | estradiol | CHEBI:23965 |
| eb_b035a_med4_name_CHILDQ | estradiol | CHEBI:23965 |
| eb_b036a_med5_name_CHILDQ | estradiol | CHEBI:23965 |
| eb_b037a_med6_name_CHILDQ | estradiol | CHEBI:23965 |
| eb_b038a_med7_name_CHILDQ | estradiol | CHEBI:23965 |
| eb_b040a_med9_name_CHILDQ | estradiol | CHEBI:23965 |
| eb_b033a_med2_name_CHILDQ | estriol | CHEBI:27974 |
| eb_b035a_med4_name_CHILDQ | estriol | CHEBI:27974 |
| eb_b032a_med1_name_CHILDQ | estrogen | CHEBI:50114 |
| eb_b033a_med2_name_CHILDQ | estrogen | CHEBI:50114 |
| eb_b034a_med3_name_CHILDQ | estrogen | CHEBI:50114 |
| eb_b035a_med4_name_CHILDQ | estrogen | CHEBI:50114 |
| eb_b032a_med1_name_CHILDQ | estropipate | CHEBI:4873 |
| eb_b034a_med3_name_CHILDQ | ethambutol | CHEBI:4877 |
| eb_b032a_med1_name_CHILDQ | ethinyl estradiol | CHEBI:4903 |
| eb_b035a_med4_name_CHILDQ | ethinyl estradiol | CHEBI:4903 |
| eb_b034a_med3_name_CHILDQ | etodolac | CHEBI:4909 |
| eb_b036a_med5_name_CHILDQ | etodolac | CHEBI:4909 |
| eb_b032a_med1_name_CHILDQ | evista | CHEBI:50740 |
| eb_b033a_med2_name_CHILDQ | evista | CHEBI:50740 |
| eb_b032a_med1_name_CHILDQ | exemestane | CHEBI:4953 |
| eb_b033a_med2_name_CHILDQ | exemestane | CHEBI:4953 |
| eb_b036a_med5_name_CHILDQ | exemestane | CHEBI:4953 |
| eb_b033a_med2_name_CHILDQ | ezetimibe | CHEBI:49040 |
| eb_b036a_med5_name_CHILDQ | ezetimibe | CHEBI:49040 |
| eb_b032a_med1_name_CHILDQ | famciclovir | CHEBI:4974 |
| eb_b032a_med1_name_CHILDQ | famotidine | CHEBI:4975 |
| eb_b033a_med2_name_CHILDQ | famotidine | CHEBI:4975 |
| eb_b034a_med3_name_CHILDQ | famotidine | CHEBI:4975 |
| eb_b035a_med4_name_CHILDQ | famotidine | CHEBI:4975 |
| eb_b036a_med5_name_CHILDQ | famotidine | CHEBI:4975 |
| eb_b041a_med10_name_CHILDQ | famotidine | CHEBI:4975 |
| eb_b032a_med1_name_CHILDQ | fanapt | CHEBI:65173 |
| eb_b033a_med2_name_CHILDQ | farxiga | CHEBI:85079 |
| eb_b035a_med4_name_CHILDQ | farxiga | CHEBI:85079 |
| eb_b036a_med5_name_CHILDQ | farxiga | CHEBI:85079 |
| eb_b037a_med6_name_CHILDQ | farxiga | CHEBI:85079 |
| eb_b039a_med8_name_CHILDQ | fatty acids | CHEBI:35366 |
| eb_b032a_med1_name_CHILDQ | felodipine | CHEBI:585948 |
| eb_b033a_med2_name_CHILDQ | felodipine | CHEBI:585948 |
| eb_b032a_med1_name_CHILDQ | femara | CHEBI:6413 |
| eb_b032a_med1_name_CHILDQ | fenofibrate | CHEBI:5001 |
| eb_b033a_med2_name_CHILDQ | fenofibrate | CHEBI:5001 |
| eb_b034a_med3_name_CHILDQ | fenofibrate | CHEBI:5001 |
| eb_b035a_med4_name_CHILDQ | fenofibrate | CHEBI:5001 |
| eb_b036a_med5_name_CHILDQ | fenofibrate | CHEBI:5001 |
| eb_b037a_med6_name_CHILDQ | fenofibrate | CHEBI:5001 |
| eb_b035a_med4_name_CHILDQ | fenofibric acid | CHEBI:83469 |
| eb_b035a_med4_name_CHILDQ | ferrous gluconate | CHEBI:31608 |
| eb_b038a_med7_name_CHILDQ | ferrous sulfate | CHEBI:75832 |
| eb_b040a_med9_name_CHILDQ | ferrous sulfate | CHEBI:75832 |
| eb_b035a_med4_name_CHILDQ | fetzima | CHEBI:136040 |
| eb_b032a_med1_name_CHILDQ | fexofenadine | CHEBI:5050 |
| eb_b033a_med2_name_CHILDQ | fexofenadine | CHEBI:5050 |
| eb_b034a_med3_name_CHILDQ | fexofenadine | CHEBI:5050 |
| eb_b036a_med5_name_CHILDQ | fexofenadine | CHEBI:5050 |
| eb_b037a_med6_name_CHILDQ | fexofenadine | CHEBI:5050 |
| eb_b039a_med8_name_CHILDQ | fexofenadine | CHEBI:5050 |
| eb_b041a_med10_name_CHILDQ | fexofenadine | CHEBI:5050 |
| eb_b034a_med3_name_CHILDQ | fexofenadine hydrochloride | CHEBI:5051 |
| eb_b039a_med8_name_CHILDQ | fexofenadine hydrochloride | CHEBI:5051 |
| eb_b032a_med1_name_CHILDQ | finasteride | CHEBI:5062 |
| eb_b033a_med2_name_CHILDQ | finasteride | CHEBI:5062 |
| eb_b034a_med3_name_CHILDQ | finasteride | CHEBI:5062 |
| eb_b035a_med4_name_CHILDQ | finasteride | CHEBI:5062 |
| eb_b039a_med8_name_CHILDQ | finasteride | CHEBI:5062 |
| eb_b032a_med1_name_CHILDQ | flecainide | CHEBI:75984 |
| eb_b033a_med2_name_CHILDQ | flecainide | CHEBI:75984 |
| eb_b034a_med3_name_CHILDQ | flecainide | CHEBI:75984 |
| eb_b032a_med1_name_CHILDQ | flomax | CHEBI:9399 |
| eb_b033a_med2_name_CHILDQ | flomax | CHEBI:9399 |
| eb_b035a_med4_name_CHILDQ | flomax | CHEBI:9399 |
| eb_b036a_med5_name_CHILDQ | flomax | CHEBI:9399 |
| eb_b037a_med6_name_CHILDQ | flomax | CHEBI:9399 |
| eb_b039a_med8_name_CHILDQ | flomax | CHEBI:9399 |
| eb_b033a_med2_name_CHILDQ | florinef | CHEBI:5102 |
| eb_b039a_med8_name_CHILDQ | fluconazole | CHEBI:46081 |
| eb_b041a_med10_name_CHILDQ | fluconazole | CHEBI:46081 |
| eb_b032a_med1_name_CHILDQ | fluocinonide | CHEBI:5109 |
| eb_b035a_med4_name_CHILDQ | fluocinonide | CHEBI:5109 |
| eb_b036a_med5_name_CHILDQ | fluocinonide | CHEBI:5109 |
| eb_b034a_med3_name_CHILDQ | fluorometholone | CHEBI:31625 |
| eb_b032a_med1_name_CHILDQ | fluoxetine | CHEBI:5118 |
| eb_b033a_med2_name_CHILDQ | fluoxetine | CHEBI:5118 |
| eb_b034a_med3_name_CHILDQ | fluoxetine | CHEBI:5118 |
| eb_b035a_med4_name_CHILDQ | fluoxetine | CHEBI:5118 |
| eb_b037a_med6_name_CHILDQ | fluoxetine | CHEBI:5118 |
| eb_b038a_med7_name_CHILDQ | fluoxetine | CHEBI:5118 |
| eb_b032a_med1_name_CHILDQ | fluoxetine hcl | CHEBI:5119 |
| eb_b033a_med2_name_CHILDQ | fluoxetine hcl | CHEBI:5119 |
| eb_b034a_med3_name_CHILDQ | fluoxetine hcl | CHEBI:5119 |
| eb_b038a_med7_name_CHILDQ | fluoxetine hcl | CHEBI:5119 |
| eb_b032a_med1_name_CHILDQ | fluticasone | CHEBI:5134 |
| eb_b033a_med2_name_CHILDQ | fluticasone | CHEBI:5134 |
| eb_b034a_med3_name_CHILDQ | fluticasone | CHEBI:5134 |
| eb_b035a_med4_name_CHILDQ | fluticasone | CHEBI:5134 |
| eb_b036a_med5_name_CHILDQ | fluticasone | CHEBI:5134 |
| eb_b037a_med6_name_CHILDQ | fluticasone | CHEBI:5134 |
| eb_b038a_med7_name_CHILDQ | fluticasone | CHEBI:5134 |
| eb_b039a_med8_name_CHILDQ | fluticasone | CHEBI:5134 |
| eb_b040a_med9_name_CHILDQ | fluticasone | CHEBI:5134 |
| eb_b041a_med10_name_CHILDQ | fluticasone | CHEBI:5134 |
| eb_b032a_med1_name_CHILDQ | fluticasone propionate | CHEBI:31441 |
| eb_b033a_med2_name_CHILDQ | fluticasone propionate | CHEBI:31441 |
| eb_b034a_med3_name_CHILDQ | fluticasone propionate | CHEBI:31441 |
| eb_b035a_med4_name_CHILDQ | fluticasone propionate | CHEBI:31441 |
| eb_b036a_med5_name_CHILDQ | fluticasone propionate | CHEBI:31441 |
| eb_b037a_med6_name_CHILDQ | fluticasone propionate | CHEBI:31441 |
| eb_b038a_med7_name_CHILDQ | fluticasone propionate | CHEBI:31441 |
| eb_b040a_med9_name_CHILDQ | fluticasone propionate | CHEBI:31441 |
| eb_b041a_med10_name_CHILDQ | fluticasone propionate | CHEBI:31441 |
| eb_b032a_med1_name_CHILDQ | folic acid | CHEBI:27470 |
| eb_b033a_med2_name_CHILDQ | folic acid | CHEBI:27470 |
| eb_b034a_med3_name_CHILDQ | folic acid | CHEBI:27470 |
| eb_b035a_med4_name_CHILDQ | folic acid | CHEBI:27470 |
| eb_b036a_med5_name_CHILDQ | folic acid | CHEBI:27470 |
| eb_b037a_med6_name_CHILDQ | folic acid | CHEBI:27470 |
| eb_b033a_med2_name_CHILDQ | fosamax | CHEBI:2566 |
| eb_b039a_med8_name_CHILDQ | fosamax | CHEBI:2566 |
| eb_b035a_med4_name_CHILDQ | fosinopril | CHEBI:5163 |
| eb_b033a_med2_name_CHILDQ | fumarate | CHEBI:29806 |
| eb_b034a_med3_name_CHILDQ | fumarate | CHEBI:29806 |
| eb_b039a_med8_name_CHILDQ | fumarate | CHEBI:29806 |
| eb_b032a_med1_name_CHILDQ | furosemide | CHEBI:47426 |
| eb_b033a_med2_name_CHILDQ | furosemide | CHEBI:47426 |
| eb_b034a_med3_name_CHILDQ | furosemide | CHEBI:47426 |
| eb_b035a_med4_name_CHILDQ | furosemide | CHEBI:47426 |
| eb_b036a_med5_name_CHILDQ | furosemide | CHEBI:47426 |
| eb_b037a_med6_name_CHILDQ | furosemide | CHEBI:47426 |
| eb_b038a_med7_name_CHILDQ | furosemide | CHEBI:47426 |
| eb_b039a_med8_name_CHILDQ | furosemide | CHEBI:47426 |
| eb_b040a_med9_name_CHILDQ | furosemide | CHEBI:47426 |
| eb_b032a_med1_name_CHILDQ | gabapentin | CHEBI:42797 |
| eb_b033a_med2_name_CHILDQ | gabapentin | CHEBI:42797 |
| eb_b034a_med3_name_CHILDQ | gabapentin | CHEBI:42797 |
| eb_b035a_med4_name_CHILDQ | gabapentin | CHEBI:42797 |
| eb_b036a_med5_name_CHILDQ | gabapentin | CHEBI:42797 |
| eb_b037a_med6_name_CHILDQ | gabapentin | CHEBI:42797 |
| eb_b038a_med7_name_CHILDQ | gabapentin | CHEBI:42797 |
| eb_b039a_med8_name_CHILDQ | gabapentin | CHEBI:42797 |
| eb_b040a_med9_name_CHILDQ | gabapentin | CHEBI:42797 |
| eb_b032a_med1_name_CHILDQ | gabapentine | CHEBI:42797 |
| eb_b032a_med1_name_CHILDQ | gemfibrozil | CHEBI:5296 |
| eb_b034a_med3_name_CHILDQ | gemfibrozil | CHEBI:5296 |
| eb_b036a_med5_name_CHILDQ | gemfibrozil | CHEBI:5296 |
| eb_b033a_med2_name_CHILDQ | genvoya | CHEBI:90922 |
| eb_b032a_med1_name_CHILDQ | geodon | CHEBI:32314 |
| eb_b033a_med2_name_CHILDQ | geodon | CHEBI:32314 |
| eb_b033a_med2_name_CHILDQ | gilenya | CHEBI:63112 |
| eb_b032a_med1_name_CHILDQ | glimepiride | CHEBI:5383 |
| eb_b033a_med2_name_CHILDQ | glimepiride | CHEBI:5383 |
| eb_b034a_med3_name_CHILDQ | glimepiride | CHEBI:5383 |
| eb_b035a_med4_name_CHILDQ | glimepiride | CHEBI:5383 |
| eb_b036a_med5_name_CHILDQ | glimepiride | CHEBI:5383 |
| eb_b038a_med7_name_CHILDQ | glimepiride | CHEBI:5383 |
| eb_b032a_med1_name_CHILDQ | glipizide | CHEBI:5384 |
| eb_b033a_med2_name_CHILDQ | glipizide | CHEBI:5384 |
| eb_b034a_med3_name_CHILDQ | glipizide | CHEBI:5384 |
| eb_b035a_med4_name_CHILDQ | glipizide | CHEBI:5384 |
| eb_b036a_med5_name_CHILDQ | glipizide | CHEBI:5384 |
| eb_b037a_med6_name_CHILDQ | glipizide | CHEBI:5384 |
| eb_b041a_med10_name_CHILDQ | glipizide | CHEBI:5384 |
| eb_b032a_med1_name_CHILDQ | gluconate | CHEBI:24265 |
| eb_b035a_med4_name_CHILDQ | gluconate | CHEBI:24265 |
| eb_b033a_med2_name_CHILDQ | glucophage | CHEBI:6802 |
| eb_b032a_med1_name_CHILDQ | glucosamine | CHEBI:5417 |
| eb_b033a_med2_name_CHILDQ | glucosamine | CHEBI:5417 |
| eb_b034a_med3_name_CHILDQ | glucosamine | CHEBI:5417 |
| eb_b035a_med4_name_CHILDQ | glucosamine | CHEBI:5417 |
| eb_b036a_med5_name_CHILDQ | glucosamine | CHEBI:5417 |
| eb_b037a_med6_name_CHILDQ | glucosamine | CHEBI:5417 |
| eb_b038a_med7_name_CHILDQ | glucosamine | CHEBI:5417 |
| eb_b037a_med6_name_CHILDQ | glutamine | CHEBI:28300 |
| eb_b032a_med1_name_CHILDQ | glycol | CHEBI:13643 |
| eb_b032a_med1_name_CHILDQ | guaifenesin | CHEBI:5551 |
| eb_b033a_med2_name_CHILDQ | guaifenesin | CHEBI:5551 |
| eb_b036a_med5_name_CHILDQ | guaifenesin | CHEBI:5551 |
| eb_b039a_med8_name_CHILDQ | guaifenesin | CHEBI:5551 |
| eb_b036a_med5_name_CHILDQ | guanfacine | CHEBI:5558 |
| eb_b036a_med5_name_CHILDQ | halcion | CHEBI:9674 |
| eb_b033a_med2_name_CHILDQ | hippurate | CHEBI:132966 |
| eb_b033a_med2_name_CHILDQ | horizant | CHEBI:68840 |
| eb_b032a_med1_name_CHILDQ | human insulin | CHEBI:5931 |
| eb_b038a_med7_name_CHILDQ | hyaluronic acid | CHEBI:16336 |
| eb_b033a_med2_name_CHILDQ | hydralazine | CHEBI:5775 |
| eb_b032a_med1_name_CHILDQ | hydrochlorothiazide | CHEBI:5778 |
| eb_b033a_med2_name_CHILDQ | hydrochlorothiazide | CHEBI:5778 |
| eb_b034a_med3_name_CHILDQ | hydrochlorothiazide | CHEBI:5778 |
| eb_b035a_med4_name_CHILDQ | hydrochlorothiazide | CHEBI:5778 |
| eb_b036a_med5_name_CHILDQ | hydrochlorothiazide | CHEBI:5778 |
| eb_b037a_med6_name_CHILDQ | hydrochlorothiazide | CHEBI:5778 |
| eb_b038a_med7_name_CHILDQ | hydrochlorothiazide | CHEBI:5778 |
| eb_b039a_med8_name_CHILDQ | hydrochlorothiazide | CHEBI:5778 |
| eb_b041a_med10_name_CHILDQ | hydrochlorothiazide | CHEBI:5778 |
| eb_b034a_med3_name_CHILDQ | hydrocodon | CHEBI:5779 |
| eb_b037a_med6_name_CHILDQ | hydrocodon | CHEBI:5779 |
| eb_b032a_med1_name_CHILDQ | hydrocodone | CHEBI:5779 |
| eb_b033a_med2_name_CHILDQ | hydrocodone | CHEBI:5779 |
| eb_b035a_med4_name_CHILDQ | hydrocodone | CHEBI:5779 |
| eb_b036a_med5_name_CHILDQ | hydrocodone | CHEBI:5779 |
| eb_b040a_med9_name_CHILDQ | hydrocodone | CHEBI:5779 |
| eb_b032a_med1_name_CHILDQ | hydrocortisone | CHEBI:17650 |
| eb_b035a_med4_name_CHILDQ | hydrocortisone | CHEBI:17650 |
| eb_b039a_med8_name_CHILDQ | hydrocortisone | CHEBI:17650 |
| eb_b039a_med8_name_CHILDQ | hydromorphone | CHEBI:5790 |
| eb_b034a_med3_name_CHILDQ | hydroquinone | CHEBI:17594 |
| eb_b032a_med1_name_CHILDQ | hydroxychloroquine | CHEBI:5801 |
| eb_b033a_med2_name_CHILDQ | hydroxychloroquine | CHEBI:5801 |
| eb_b034a_med3_name_CHILDQ | hydroxychloroquine | CHEBI:5801 |
| eb_b036a_med5_name_CHILDQ | hydroxychloroquine | CHEBI:5801 |
| eb_b039a_med8_name_CHILDQ | hydroxychloroquine | CHEBI:5801 |
| eb_b040a_med9_name_CHILDQ | hydroxychloroquine | CHEBI:5801 |
| eb_b034a_med3_name_CHILDQ | hydroxycobalamin | CHEBI:27786 |
| eb_b032a_med1_name_CHILDQ | hydroxyzine | CHEBI:5818 |
| eb_b039a_med8_name_CHILDQ | hydroxyzine | CHEBI:5818 |
| eb_b036a_med5_name_CHILDQ | hyoscyamine | CHEBI:17486 |
| eb_b033a_med2_name_CHILDQ | ibandronate | CHEBI:41060 |
| eb_b034a_med3_name_CHILDQ | ibandronate | CHEBI:41060 |
| eb_b037a_med6_name_CHILDQ | ibandronate | CHEBI:41060 |
| eb_b032a_med1_name_CHILDQ | ibuprofen | CHEBI:5855 |
| eb_b033a_med2_name_CHILDQ | ibuprofen | CHEBI:5855 |
| eb_b034a_med3_name_CHILDQ | ibuprofen | CHEBI:5855 |
| eb_b035a_med4_name_CHILDQ | ibuprofen | CHEBI:5855 |
| eb_b036a_med5_name_CHILDQ | ibuprofen | CHEBI:5855 |
| eb_b037a_med6_name_CHILDQ | ibuprofen | CHEBI:5855 |
| eb_b038a_med7_name_CHILDQ | ibuprofen | CHEBI:5855 |
| eb_b039a_med8_name_CHILDQ | ibuprofen | CHEBI:5855 |
| eb_b041a_med10_name_CHILDQ | ibuprofen | CHEBI:5855 |
| eb_b033a_med2_name_CHILDQ | imdur | CHEBI:6062 |
| eb_b034a_med3_name_CHILDQ | imdur | CHEBI:6062 |
| eb_b036a_med5_name_CHILDQ | imdur | CHEBI:6062 |
| eb_b037a_med6_name_CHILDQ | imdur | CHEBI:6062 |
| eb_b032a_med1_name_CHILDQ | imitrex | CHEBI:10650 |
| eb_b033a_med2_name_CHILDQ | imitrex | CHEBI:10650 |
| eb_b034a_med3_name_CHILDQ | imitrex | CHEBI:10650 |
| eb_b035a_med4_name_CHILDQ | imitrex | CHEBI:10650 |
| eb_b037a_med6_name_CHILDQ | imitrex | CHEBI:10650 |
| eb_b038a_med7_name_CHILDQ | imitrex | CHEBI:10650 |
| eb_b034a_med3_name_CHILDQ | imodium | CHEBI:6533 |
| eb_b035a_med4_name_CHILDQ | imodium | CHEBI:6533 |
| eb_b039a_med8_name_CHILDQ | imodium | CHEBI:6533 |
| eb_b032a_med1_name_CHILDQ | indapamide | CHEBI:5893 |
| eb_b033a_med2_name_CHILDQ | indapamide | CHEBI:5893 |
| eb_b036a_med5_name_CHILDQ | indapamide | CHEBI:5893 |
| eb_b034a_med3_name_CHILDQ | indomethacin | CHEBI:49662 |
| eb_b040a_med9_name_CHILDQ | indomethacin | CHEBI:49662 |
| eb_b032a_med1_name_CHILDQ | insulin | CHEBI:145810 |
| eb_b033a_med2_name_CHILDQ | insulin | CHEBI:145810 |
| eb_b034a_med3_name_CHILDQ | insulin | CHEBI:145810 |
| eb_b036a_med5_name_CHILDQ | insulin | CHEBI:145810 |
| eb_b037a_med6_name_CHILDQ | insulin | CHEBI:145810 |
| eb_b038a_med7_name_CHILDQ | insulin | CHEBI:145810 |
| eb_b039a_med8_name_CHILDQ | insulin | CHEBI:145810 |
| eb_b040a_med9_name_CHILDQ | insulin | CHEBI:145810 |
| eb_b041a_med10_name_CHILDQ | insulin | CHEBI:145810 |
| eb_b035a_med4_name_CHILDQ | invokana | CHEBI:73272 |
| eb_b036a_med5_name_CHILDQ | invokana | CHEBI:73272 |
| eb_b037a_med6_name_CHILDQ | invokana | CHEBI:73272 |
| eb_b034a_med3_name_CHILDQ | iodine | CHEBI:24859 |
| eb_b032a_med1_name_CHILDQ | ipratropium | CHEBI:5956 |
| eb_b036a_med5_name_CHILDQ | ipratropium | CHEBI:5956 |
| eb_b039a_med8_name_CHILDQ | ipratropium | CHEBI:5956 |
| eb_b036a_med5_name_CHILDQ | ipratropium bromide | CHEBI:46659 |
| eb_b039a_med8_name_CHILDQ | ipratropium bromide | CHEBI:46659 |
| eb_b032a_med1_name_CHILDQ | irbesartan | CHEBI:5959 |
| eb_b033a_med2_name_CHILDQ | irbesartan | CHEBI:5959 |
| eb_b034a_med3_name_CHILDQ | irbesartan | CHEBI:5959 |
| eb_b032a_med1_name_CHILDQ | iron | CHEBI:18248 |
| eb_b033a_med2_name_CHILDQ | iron | CHEBI:18248 |
| eb_b034a_med3_name_CHILDQ | iron | CHEBI:18248 |
| eb_b038a_med7_name_CHILDQ | isometheptene | CHEBI:134765 |
| eb_b033a_med2_name_CHILDQ | isosorbide | CHEBI:6060 |
| eb_b035a_med4_name_CHILDQ | isosorbide | CHEBI:6060 |
| eb_b038a_med7_name_CHILDQ | isosorbide | CHEBI:6060 |
| eb_b040a_med9_name_CHILDQ | isosorbide | CHEBI:6060 |
| eb_b035a_med4_name_CHILDQ | isosorbide mononitrate | CHEBI:6062 |
| eb_b040a_med9_name_CHILDQ | isosorbide mononitrate | CHEBI:6062 |
| eb_b032a_med1_name_CHILDQ | jardiance | CHEBI:82720 |
| eb_b038a_med7_name_CHILDQ | jardiance | CHEBI:82720 |
| eb_b035a_med4_name_CHILDQ | jublia | CHEBI:82718 |
| eb_b037a_med6_name_CHILDQ | kcl | CHEBI:32588 |
| eb_b032a_med1_name_CHILDQ | keppra | CHEBI:6437 |
| eb_b038a_med7_name_CHILDQ | ketoconazole | CHEBI:47519 |
| eb_b039a_med8_name_CHILDQ | ketoconazole | CHEBI:47519 |
| eb_b034a_med3_name_CHILDQ | klor-con | CHEBI:32588 |
| eb_b035a_med4_name_CHILDQ | klor-con | CHEBI:32588 |
| eb_b036a_med5_name_CHILDQ | klor-con | CHEBI:32588 |
| eb_b039a_med8_name_CHILDQ | l-carnitine | CHEBI:16347 |
| eb_b037a_med6_name_CHILDQ | l-glutamine | CHEBI:18050 |
| eb_b033a_med2_name_CHILDQ | l-thyroxine | CHEBI:18332 |
| eb_b034a_med3_name_CHILDQ | l-thyroxine | CHEBI:18332 |
| eb_b035a_med4_name_CHILDQ | l-thyroxine | CHEBI:18332 |
| eb_b032a_med1_name_CHILDQ | labetalol | CHEBI:6343 |
| eb_b033a_med2_name_CHILDQ | lactate | CHEBI:24996 |
| eb_b032a_med1_name_CHILDQ | lamictal | CHEBI:6367 |
| eb_b033a_med2_name_CHILDQ | lamictal | CHEBI:6367 |
| eb_b034a_med3_name_CHILDQ | lamictal | CHEBI:6367 |
| eb_b035a_med4_name_CHILDQ | lamictal | CHEBI:6367 |
| eb_b032a_med1_name_CHILDQ | lamotrigine | CHEBI:6367 |
| eb_b033a_med2_name_CHILDQ | lamotrigine | CHEBI:6367 |
| eb_b034a_med3_name_CHILDQ | lamotrigine | CHEBI:6367 |
| eb_b037a_med6_name_CHILDQ | lamotrigine | CHEBI:6367 |
| eb_b032a_med1_name_CHILDQ | lansoprazole | CHEBI:6375 |
| eb_b033a_med2_name_CHILDQ | lansoprazole | CHEBI:6375 |
| eb_b034a_med3_name_CHILDQ | lansoprazole | CHEBI:6375 |
| eb_b035a_med4_name_CHILDQ | lansoprazole | CHEBI:6375 |
| eb_b039a_med8_name_CHILDQ | lansoprazole | CHEBI:6375 |
| eb_b040a_med9_name_CHILDQ | lansoprazole | CHEBI:6375 |
| eb_b032a_med1_name_CHILDQ | latanoprost | CHEBI:6384 |
| eb_b033a_med2_name_CHILDQ | latanoprost | CHEBI:6384 |
| eb_b034a_med3_name_CHILDQ | latanoprost | CHEBI:6384 |
| eb_b036a_med5_name_CHILDQ | latanoprost | CHEBI:6384 |
| eb_b037a_med6_name_CHILDQ | latanoprost | CHEBI:6384 |
| eb_b039a_med8_name_CHILDQ | latanoprost | CHEBI:6384 |
| eb_b040a_med9_name_CHILDQ | latanoprost | CHEBI:6384 |
| eb_b032a_med1_name_CHILDQ | latuda | CHEBI:70732 |
| eb_b033a_med2_name_CHILDQ | latuda | CHEBI:70732 |
| eb_b035a_med4_name_CHILDQ | latuda | CHEBI:70732 |
| eb_b041a_med10_name_CHILDQ | laxative | CHEBI:50503 |
| eb_b033a_med2_name_CHILDQ | leflunomide | CHEBI:6402 |
| eb_b035a_med4_name_CHILDQ | leflunomide | CHEBI:6402 |
| eb_b033a_med2_name_CHILDQ | letrozol | CHEBI:6413 |
| eb_b032a_med1_name_CHILDQ | letrozole | CHEBI:6413 |
| eb_b035a_med4_name_CHILDQ | letrozole | CHEBI:6413 |
| eb_b034a_med3_name_CHILDQ | leucovorin | CHEBI:15640 |
| eb_b033a_med2_name_CHILDQ | levetiracetam | CHEBI:6437 |
| eb_b032a_med1_name_CHILDQ | levitra | CHEBI:46295 |
| eb_b033a_med2_name_CHILDQ | levodopa | CHEBI:15765 |
| eb_b032a_med1_name_CHILDQ | levofloxacin | CHEBI:63598 |
| eb_b032a_med1_name_CHILDQ | levonorgestrel | CHEBI:6443 |
| eb_b034a_med3_name_CHILDQ | levosalbutamol | CHEBI:8746 |
| eb_b032a_med1_name_CHILDQ | levothroid | CHEBI:6446 |
| eb_b032a_med1_name_CHILDQ | levothyroxin | CHEBI:18332 |
| eb_b033a_med2_name_CHILDQ | levothyroxin | CHEBI:18332 |
| eb_b034a_med3_name_CHILDQ | levothyroxin | CHEBI:18332 |
| eb_b035a_med4_name_CHILDQ | levothyroxin | CHEBI:18332 |
| eb_b036a_med5_name_CHILDQ | levothyroxin | CHEBI:18332 |
| eb_b038a_med7_name_CHILDQ | levothyroxin | CHEBI:18332 |
| eb_b032a_med1_name_CHILDQ | levothyroxine | CHEBI:18332 |
| eb_b033a_med2_name_CHILDQ | levothyroxine | CHEBI:18332 |
| eb_b034a_med3_name_CHILDQ | levothyroxine | CHEBI:18332 |
| eb_b035a_med4_name_CHILDQ | levothyroxine | CHEBI:18332 |
| eb_b036a_med5_name_CHILDQ | levothyroxine | CHEBI:18332 |
| eb_b037a_med6_name_CHILDQ | levothyroxine | CHEBI:18332 |
| eb_b038a_med7_name_CHILDQ | levothyroxine | CHEBI:18332 |
| eb_b039a_med8_name_CHILDQ | levothyroxine | CHEBI:18332 |
| eb_b040a_med9_name_CHILDQ | levothyroxine | CHEBI:18332 |
| eb_b032a_med1_name_CHILDQ | levothyroxine sodium | CHEBI:6446 |
| eb_b035a_med4_name_CHILDQ | levothyroxine sodium | CHEBI:6446 |
| eb_b036a_med5_name_CHILDQ | levothyroxine sodium | CHEBI:6446 |
| eb_b032a_med1_name_CHILDQ | levoxyl | CHEBI:6446 |
| eb_b033a_med2_name_CHILDQ | levoxyl | CHEBI:6446 |
| eb_b034a_med3_name_CHILDQ | levoxyl | CHEBI:6446 |
| eb_b041a_med10_name_CHILDQ | lidocaine | CHEBI:6456 |
| eb_b032a_med1_name_CHILDQ | linzess | CHEBI:68551 |
| eb_b032a_med1_name_CHILDQ | liothyronine | CHEBI:18258 |
| eb_b033a_med2_name_CHILDQ | liothyronine | CHEBI:18258 |
| eb_b034a_med3_name_CHILDQ | liothyronine | CHEBI:18258 |
| eb_b032a_med1_name_CHILDQ | lipitor | CHEBI:50686 |
| eb_b033a_med2_name_CHILDQ | lipitor | CHEBI:50686 |
| eb_b034a_med3_name_CHILDQ | lipitor | CHEBI:50686 |
| eb_b035a_med4_name_CHILDQ | lipitor | CHEBI:50686 |
| eb_b036a_med5_name_CHILDQ | lipitor | CHEBI:50686 |
| eb_b037a_med6_name_CHILDQ | lipitor | CHEBI:50686 |
| eb_b039a_med8_name_CHILDQ | lipitor | CHEBI:50686 |
| eb_b040a_med9_name_CHILDQ | lipitor | CHEBI:50686 |
| eb_b041a_med10_name_CHILDQ | lipitor | CHEBI:50686 |
| eb_b034a_med3_name_CHILDQ | lipofen | CHEBI:5001 |
| eb_b034a_med3_name_CHILDQ | lipoic acid | CHEBI:16494 |
| eb_b039a_med8_name_CHILDQ | lipoic acid | CHEBI:16494 |
| eb_b032a_med1_name_CHILDQ | lisinopril | CHEBI:43755 |
| eb_b033a_med2_name_CHILDQ | lisinopril | CHEBI:43755 |
| eb_b034a_med3_name_CHILDQ | lisinopril | CHEBI:43755 |
| eb_b035a_med4_name_CHILDQ | lisinopril | CHEBI:43755 |
| eb_b036a_med5_name_CHILDQ | lisinopril | CHEBI:43755 |
| eb_b037a_med6_name_CHILDQ | lisinopril | CHEBI:43755 |
| eb_b038a_med7_name_CHILDQ | lisinopril | CHEBI:43755 |
| eb_b040a_med9_name_CHILDQ | lisinopril | CHEBI:43755 |
| eb_b041a_med10_name_CHILDQ | lisinopril | CHEBI:43755 |
| eb_b032a_med1_name_CHILDQ | lithium | CHEBI:30145 |
| eb_b033a_med2_name_CHILDQ | lithium | CHEBI:30145 |
| eb_b036a_med5_name_CHILDQ | lithium | CHEBI:30145 |
| eb_b037a_med6_name_CHILDQ | lithium | CHEBI:30145 |
| eb_b041a_med10_name_CHILDQ | lithium | CHEBI:30145 |
| eb_b032a_med1_name_CHILDQ | lithium carbonate | CHEBI:6504 |
| eb_b036a_med5_name_CHILDQ | lithium carbonate | CHEBI:6504 |
| eb_b037a_med6_name_CHILDQ | lithium carbonate | CHEBI:6504 |
| eb_b037a_med6_name_CHILDQ | lomotil | CHEBI:6519 |
| eb_b040a_med9_name_CHILDQ | lomotil | CHEBI:6519 |
| eb_b037a_med6_name_CHILDQ | loperamide | CHEBI:6532 |
| eb_b039a_med8_name_CHILDQ | loperamide | CHEBI:6532 |
| eb_b041a_med10_name_CHILDQ | loperamide | CHEBI:6532 |
| eb_b039a_med8_name_CHILDQ | loperamide hcl | CHEBI:6533 |
| eb_b041a_med10_name_CHILDQ | loperamide hydrochloride | CHEBI:6533 |
| eb_b032a_med1_name_CHILDQ | loratadine | CHEBI:6538 |
| eb_b033a_med2_name_CHILDQ | loratadine | CHEBI:6538 |
| eb_b034a_med3_name_CHILDQ | loratadine | CHEBI:6538 |
| eb_b035a_med4_name_CHILDQ | loratadine | CHEBI:6538 |
| eb_b036a_med5_name_CHILDQ | loratadine | CHEBI:6538 |
| eb_b037a_med6_name_CHILDQ | loratadine | CHEBI:6538 |
| eb_b038a_med7_name_CHILDQ | loratadine | CHEBI:6538 |
| eb_b039a_med8_name_CHILDQ | loratadine | CHEBI:6538 |
| eb_b041a_med10_name_CHILDQ | loratadine | CHEBI:6538 |
| eb_b033a_med2_name_CHILDQ | lorazepam | CHEBI:6539 |
| eb_b035a_med4_name_CHILDQ | lorazepam | CHEBI:6539 |
| eb_b040a_med9_name_CHILDQ | lorazepam | CHEBI:6539 |
| eb_b041a_med10_name_CHILDQ | lorazepam | CHEBI:6539 |
| eb_b032a_med1_name_CHILDQ | losartan | CHEBI:6541 |
| eb_b033a_med2_name_CHILDQ | losartan | CHEBI:6541 |
| eb_b034a_med3_name_CHILDQ | losartan | CHEBI:6541 |
| eb_b035a_med4_name_CHILDQ | losartan | CHEBI:6541 |
| eb_b036a_med5_name_CHILDQ | losartan | CHEBI:6541 |
| eb_b037a_med6_name_CHILDQ | losartan | CHEBI:6541 |
| eb_b038a_med7_name_CHILDQ | losartan | CHEBI:6541 |
| eb_b039a_med8_name_CHILDQ | losartan | CHEBI:6541 |
| eb_b040a_med9_name_CHILDQ | losartan | CHEBI:6541 |
| eb_b041a_med10_name_CHILDQ | losartan | CHEBI:6541 |
| eb_b032a_med1_name_CHILDQ | lotensin | CHEBI:3012 |
| eb_b033a_med2_name_CHILDQ | lotensin | CHEBI:3012 |
| eb_b032a_med1_name_CHILDQ | lovastatin | CHEBI:40303 |
| eb_b033a_med2_name_CHILDQ | lovastatin | CHEBI:40303 |
| eb_b034a_med3_name_CHILDQ | lovastatin | CHEBI:40303 |
| eb_b035a_med4_name_CHILDQ | lovastatin | CHEBI:40303 |
| eb_b036a_med5_name_CHILDQ | lovastatin | CHEBI:40303 |
| eb_b037a_med6_name_CHILDQ | lovastatin | CHEBI:40303 |
| eb_b033a_med2_name_CHILDQ | lupron | CHEBI:63597 |
| eb_b039a_med8_name_CHILDQ | lupron | CHEBI:63597 |
| eb_b035a_med4_name_CHILDQ | lutein | CHEBI:28838 |
| eb_b036a_med5_name_CHILDQ | lutein | CHEBI:28838 |
| eb_b037a_med6_name_CHILDQ | lutein | CHEBI:28838 |
| eb_b040a_med9_name_CHILDQ | lutein | CHEBI:28838 |
| eb_b034a_med3_name_CHILDQ | lycopene | CHEBI:15948 |
| eb_b033a_med2_name_CHILDQ | lynparza | CHEBI:83766 |
| eb_b032a_med1_name_CHILDQ | lyrica | CHEBI:64356 |
| eb_b034a_med3_name_CHILDQ | lyrica | CHEBI:64356 |
| eb_b039a_med8_name_CHILDQ | lyrica | CHEBI:64356 |
| eb_b041a_med10_name_CHILDQ | lyrica | CHEBI:64356 |
| eb_b033a_med2_name_CHILDQ | lysine | CHEBI:25094 |
| eb_b039a_med8_name_CHILDQ | magnesia | CHEBI:31794 |
| eb_b032a_med1_name_CHILDQ | magnesium | CHEBI:25107 |
| eb_b033a_med2_name_CHILDQ | magnesium | CHEBI:25107 |
| eb_b035a_med4_name_CHILDQ | magnesium | CHEBI:25107 |
| eb_b036a_med5_name_CHILDQ | magnesium | CHEBI:25107 |
| eb_b038a_med7_name_CHILDQ | magnesium | CHEBI:25107 |
| eb_b039a_med8_name_CHILDQ | magnesium | CHEBI:25107 |
| eb_b040a_med9_name_CHILDQ | magnesium | CHEBI:25107 |
| eb_b038a_med7_name_CHILDQ | magnesium citrate | CHEBI:131389 |
| eb_b039a_med8_name_CHILDQ | magnesium citrate | CHEBI:131389 |
| eb_b032a_med1_name_CHILDQ | magnesium oxide | CHEBI:31794 |
| eb_b038a_med7_name_CHILDQ | magnesium oxide | CHEBI:31794 |
| eb_b040a_med9_name_CHILDQ | magnesium oxide | CHEBI:31794 |
| eb_b032a_med1_name_CHILDQ | maleate | CHEBI:132951 |
| eb_b033a_med2_name_CHILDQ | maleate | CHEBI:132951 |
| eb_b034a_med3_name_CHILDQ | maleate | CHEBI:132951 |
| eb_b035a_med4_name_CHILDQ | maleate | CHEBI:132951 |
| eb_b038a_med7_name_CHILDQ | maleate | CHEBI:132951 |
| eb_b033a_med2_name_CHILDQ | meclizine | CHEBI:6709 |
| eb_b037a_med6_name_CHILDQ | meclizine | CHEBI:6709 |
| eb_b032a_med1_name_CHILDQ | medroxyprogesterone | CHEBI:6715 |
| eb_b033a_med2_name_CHILDQ | medroxyprogesterone | CHEBI:6715 |
| eb_b034a_med3_name_CHILDQ | medroxyprogesterone | CHEBI:6715 |
| eb_b035a_med4_name_CHILDQ | medroxyprogesterone | CHEBI:6715 |
| eb_b032a_med1_name_CHILDQ | melatonin | CHEBI:16796 |
| eb_b033a_med2_name_CHILDQ | melatonin | CHEBI:16796 |
| eb_b035a_med4_name_CHILDQ | melatonin | CHEBI:16796 |
| eb_b036a_med5_name_CHILDQ | melatonin | CHEBI:16796 |
| eb_b038a_med7_name_CHILDQ | melatonin | CHEBI:16796 |
| eb_b032a_med1_name_CHILDQ | meloxicam | CHEBI:6741 |
| eb_b033a_med2_name_CHILDQ | meloxicam | CHEBI:6741 |
| eb_b034a_med3_name_CHILDQ | meloxicam | CHEBI:6741 |
| eb_b035a_med4_name_CHILDQ | meloxicam | CHEBI:6741 |
| eb_b036a_med5_name_CHILDQ | meloxicam | CHEBI:6741 |
| eb_b037a_med6_name_CHILDQ | meloxicam | CHEBI:6741 |
| eb_b038a_med7_name_CHILDQ | meloxicam | CHEBI:6741 |
| eb_b041a_med10_name_CHILDQ | meloxicam | CHEBI:6741 |
| eb_b035a_med4_name_CHILDQ | memantine | CHEBI:64312 |
| eb_b041a_med10_name_CHILDQ | memantine | CHEBI:64312 |
| eb_b032a_med1_name_CHILDQ | mercaptopurine | CHEBI:50667 |
| eb_b033a_med2_name_CHILDQ | mercaptopurine | CHEBI:50667 |
| eb_b032a_med1_name_CHILDQ | mesalamine | CHEBI:6775 |
| eb_b037a_med6_name_CHILDQ | metaxalone | CHEBI:6797 |
| eb_b038a_med7_name_CHILDQ | metaxalone | CHEBI:6797 |
| eb_b041a_med10_name_CHILDQ | metaxalone | CHEBI:6797 |
| eb_b032a_med1_name_CHILDQ | metformin | CHEBI:6801 |
| eb_b033a_med2_name_CHILDQ | metformin | CHEBI:6801 |
| eb_b034a_med3_name_CHILDQ | metformin | CHEBI:6801 |
| eb_b035a_med4_name_CHILDQ | metformin | CHEBI:6801 |
| eb_b036a_med5_name_CHILDQ | metformin | CHEBI:6801 |
| eb_b037a_med6_name_CHILDQ | metformin | CHEBI:6801 |
| eb_b038a_med7_name_CHILDQ | metformin | CHEBI:6801 |
| eb_b039a_med8_name_CHILDQ | metformin | CHEBI:6801 |
| eb_b040a_med9_name_CHILDQ | metformin | CHEBI:6801 |
| eb_b041a_med10_name_CHILDQ | metformin | CHEBI:6801 |
| eb_b032a_med1_name_CHILDQ | metformin hcl | CHEBI:6802 |
| eb_b033a_med2_name_CHILDQ | metformin hcl | CHEBI:6802 |
| eb_b034a_med3_name_CHILDQ | metformin hcl | CHEBI:6802 |
| eb_b037a_med6_name_CHILDQ | metformin hcl | CHEBI:6802 |
| eb_b041a_med10_name_CHILDQ | metformin hcl | CHEBI:6802 |
| eb_b034a_med3_name_CHILDQ | metformina | CHEBI:6801 |
| eb_b032a_med1_name_CHILDQ | methadone | CHEBI:6807 |
| eb_b033a_med2_name_CHILDQ | methenamine | CHEBI:6824 |
| eb_b033a_med2_name_CHILDQ | methenamine hippurate | CHEBI:6825 |
| eb_b032a_med1_name_CHILDQ | methimazole | CHEBI:50673 |
| eb_b033a_med2_name_CHILDQ | methimazole | CHEBI:50673 |
| eb_b035a_med4_name_CHILDQ | methimazole | CHEBI:50673 |
| eb_b032a_med1_name_CHILDQ | methocarbamol | CHEBI:6832 |
| eb_b035a_med4_name_CHILDQ | methocarbamol | CHEBI:6832 |
| eb_b032a_med1_name_CHILDQ | methotrexate | CHEBI:44185 |
| eb_b033a_med2_name_CHILDQ | methotrexate | CHEBI:44185 |
| eb_b035a_med4_name_CHILDQ | methotrexate | CHEBI:44185 |
| eb_b036a_med5_name_CHILDQ | methotrexate | CHEBI:44185 |
| eb_b037a_med6_name_CHILDQ | methotrexate | CHEBI:44185 |
| eb_b038a_med7_name_CHILDQ | methotrexate | CHEBI:44185 |
| eb_b039a_med8_name_CHILDQ | methotrexate | CHEBI:44185 |
| eb_b035a_med4_name_CHILDQ | methylcobalamin | CHEBI:28115 |
| eb_b032a_med1_name_CHILDQ | methyldopa | CHEBI:61058 |
| eb_b032a_med1_name_CHILDQ | methylphenidate | CHEBI:6887 |
| eb_b033a_med2_name_CHILDQ | methylphenidate | CHEBI:6887 |
| eb_b034a_med3_name_CHILDQ | methylphenidate | CHEBI:6887 |
| eb_b040a_med9_name_CHILDQ | methylphenidate | CHEBI:6887 |
| eb_b034a_med3_name_CHILDQ | methylphenidate hcl | CHEBI:31836 |
| eb_b033a_med2_name_CHILDQ | methyltestosterone | CHEBI:27436 |
| eb_b032a_med1_name_CHILDQ | metoprolol | CHEBI:6904 |
| eb_b033a_med2_name_CHILDQ | metoprolol | CHEBI:6904 |
| eb_b034a_med3_name_CHILDQ | metoprolol | CHEBI:6904 |
| eb_b035a_med4_name_CHILDQ | metoprolol | CHEBI:6904 |
| eb_b036a_med5_name_CHILDQ | metoprolol | CHEBI:6904 |
| eb_b037a_med6_name_CHILDQ | metoprolol | CHEBI:6904 |
| eb_b038a_med7_name_CHILDQ | metoprolol | CHEBI:6904 |
| eb_b040a_med9_name_CHILDQ | metoprolol | CHEBI:6904 |
| eb_b041a_med10_name_CHILDQ | metoprolol | CHEBI:6904 |
| eb_b032a_med1_name_CHILDQ | metoprolol succinate | CHEBI:6905 |
| eb_b033a_med2_name_CHILDQ | metoprolol succinate | CHEBI:6905 |
| eb_b035a_med4_name_CHILDQ | metoprolol succinate | CHEBI:6905 |
| eb_b036a_med5_name_CHILDQ | metoprolol succinate | CHEBI:6905 |
| eb_b040a_med9_name_CHILDQ | metoprolol succinate | CHEBI:6905 |
| eb_b032a_med1_name_CHILDQ | metoprolol tartrate | CHEBI:6906 |
| eb_b033a_med2_name_CHILDQ | metoprolol tartrate | CHEBI:6906 |
| eb_b035a_med4_name_CHILDQ | metoprolol tartrate | CHEBI:6906 |
| eb_b036a_med5_name_CHILDQ | metoprolol tartrate | CHEBI:6906 |
| eb_b034a_med3_name_CHILDQ | metronidazole | CHEBI:6909 |
| eb_b035a_med4_name_CHILDQ | metronidazole | CHEBI:6909 |
| eb_b032a_med1_name_CHILDQ | micardis | CHEBI:9434 |
| eb_b033a_med2_name_CHILDQ | micardis | CHEBI:9434 |
| eb_b036a_med5_name_CHILDQ | micardis | CHEBI:9434 |
| eb_b034a_med3_name_CHILDQ | micronor | CHEBI:7627 |
| eb_b039a_med8_name_CHILDQ | milk of magnesia | CHEBI:6637 |
| eb_b032a_med1_name_CHILDQ | minocycline | CHEBI:50694 |
| eb_b033a_med2_name_CHILDQ | minocycline | CHEBI:50694 |
| eb_b039a_med8_name_CHILDQ | minocycline | CHEBI:50694 |
| eb_b032a_med1_name_CHILDQ | minoxidil | CHEBI:6942 |
| eb_b034a_med3_name_CHILDQ | minoxidil | CHEBI:6942 |
| eb_b035a_med4_name_CHILDQ | mirabegron | CHEBI:65349 |
| eb_b032a_med1_name_CHILDQ | mirena | CHEBI:6443 |
| eb_b033a_med2_name_CHILDQ | mirena | CHEBI:6443 |
| eb_b036a_med5_name_CHILDQ | mirena | CHEBI:6443 |
| eb_b038a_med7_name_CHILDQ | mirena | CHEBI:6443 |
| eb_b032a_med1_name_CHILDQ | mirtazapine | CHEBI:6950 |
| eb_b034a_med3_name_CHILDQ | mirtazapine | CHEBI:6950 |
| eb_b037a_med6_name_CHILDQ | mirtazapine | CHEBI:6950 |
| eb_b032a_med1_name_CHILDQ | misoprostol | CHEBI:63610 |
| eb_b034a_med3_name_CHILDQ | mixture | CHEBI:60004 |
| eb_b032a_med1_name_CHILDQ | mobic | CHEBI:6741 |
| eb_b033a_med2_name_CHILDQ | mobic | CHEBI:6741 |
| eb_b034a_med3_name_CHILDQ | mobic | CHEBI:6741 |
| eb_b036a_med5_name_CHILDQ | mobic | CHEBI:6741 |
| eb_b033a_med2_name_CHILDQ | modafinil | CHEBI:31859 |
| eb_b034a_med3_name_CHILDQ | modafinil | CHEBI:31859 |
| eb_b035a_med4_name_CHILDQ | modafinil | CHEBI:31859 |
| eb_b036a_med5_name_CHILDQ | modafinil | CHEBI:31859 |
| eb_b032a_med1_name_CHILDQ | mometasone | CHEBI:6970 |
| eb_b033a_med2_name_CHILDQ | mometasone | CHEBI:6970 |
| eb_b032a_med1_name_CHILDQ | montelukast | CHEBI:50730 |
| eb_b033a_med2_name_CHILDQ | montelukast | CHEBI:50730 |
| eb_b034a_med3_name_CHILDQ | montelukast | CHEBI:50730 |
| eb_b035a_med4_name_CHILDQ | montelukast | CHEBI:50730 |
| eb_b036a_med5_name_CHILDQ | montelukast | CHEBI:50730 |
| eb_b037a_med6_name_CHILDQ | montelukast | CHEBI:50730 |
| eb_b038a_med7_name_CHILDQ | montelukast | CHEBI:50730 |
| eb_b039a_med8_name_CHILDQ | montelukast | CHEBI:50730 |
| eb_b040a_med9_name_CHILDQ | montelukast | CHEBI:50730 |
| eb_b041a_med10_name_CHILDQ | montelukast | CHEBI:50730 |
| eb_b033a_med2_name_CHILDQ | montelukast sodium | CHEBI:6993 |
| eb_b034a_med3_name_CHILDQ | montelukast sodium | CHEBI:6993 |
| eb_b035a_med4_name_CHILDQ | montelukast sodium | CHEBI:6993 |
| eb_b036a_med5_name_CHILDQ | montelukast sodium | CHEBI:6993 |
| eb_b039a_med8_name_CHILDQ | montelukast sodium | CHEBI:6993 |
| eb_b041a_med10_name_CHILDQ | montelukast sodium | CHEBI:6993 |
| eb_b040a_med9_name_CHILDQ | morphine | CHEBI:17303 |
| eb_b040a_med9_name_CHILDQ | morphine | CHEBI:58097 |
| eb_b032a_med1_name_CHILDQ | motrin | CHEBI:5855 |
| eb_b033a_med2_name_CHILDQ | motrin | CHEBI:5855 |
| eb_b036a_med5_name_CHILDQ | motrin | CHEBI:5855 |
| eb_b037a_med6_name_CHILDQ | motrin | CHEBI:5855 |
| eb_b041a_med10_name_CHILDQ | motrin | CHEBI:5855 |
| eb_b032a_med1_name_CHILDQ | multaq | CHEBI:50659 |
| eb_b038a_med7_name_CHILDQ | multaq | CHEBI:50659 |
| eb_b034a_med3_name_CHILDQ | mycophenolate | CHEBI:62932 |
| eb_b034a_med3_name_CHILDQ | mycophenolate | CHEBI:168396 |
| eb_b032a_med1_name_CHILDQ | myfortic | CHEBI:67155 |
| eb_b033a_med2_name_CHILDQ | myfortic | CHEBI:67155 |
| eb_b032a_med1_name_CHILDQ | myrbetriq | CHEBI:65349 |
| eb_b035a_med4_name_CHILDQ | myrbetriq | CHEBI:65349 |
| eb_b038a_med7_name_CHILDQ | myrbetriq | CHEBI:65349 |
| eb_b032a_med1_name_CHILDQ | nabumetone | CHEBI:7443 |
| eb_b033a_med2_name_CHILDQ | nabumetone | CHEBI:7443 |
| eb_b035a_med4_name_CHILDQ | nabumetone | CHEBI:7443 |
| eb_b035a_med4_name_CHILDQ | nadolol | CHEBI:7444 |
| eb_b034a_med3_name_CHILDQ | naltrexone | CHEBI:7465 |
| eb_b032a_med1_name_CHILDQ | naproxen | CHEBI:7476 |
| eb_b033a_med2_name_CHILDQ | naproxen | CHEBI:7476 |
| eb_b034a_med3_name_CHILDQ | naproxen | CHEBI:7476 |
| eb_b035a_med4_name_CHILDQ | naproxen | CHEBI:7476 |
| eb_b036a_med5_name_CHILDQ | naproxen | CHEBI:7476 |
| eb_b037a_med6_name_CHILDQ | naproxen | CHEBI:7476 |
| eb_b038a_med7_name_CHILDQ | naproxen | CHEBI:7476 |
| eb_b040a_med9_name_CHILDQ | naproxen | CHEBI:7476 |
| eb_b032a_med1_name_CHILDQ | naproxen sodium | CHEBI:7477 |
| eb_b033a_med2_name_CHILDQ | naproxen sodium | CHEBI:7477 |
| eb_b034a_med3_name_CHILDQ | naproxen sodium | CHEBI:7477 |
| eb_b035a_med4_name_CHILDQ | naproxen sodium | CHEBI:7477 |
| eb_b037a_med6_name_CHILDQ | naproxen sodium | CHEBI:7477 |
| eb_b038a_med7_name_CHILDQ | naproxen sodium | CHEBI:7477 |
| eb_b040a_med9_name_CHILDQ | naproxen sodium | CHEBI:7477 |
| eb_b032a_med1_name_CHILDQ | nasal decongestant | CHEBI:77715 |
| eb_b032a_med1_name_CHILDQ | nature-throid | CHEBI:9584 |
| eb_b033a_med2_name_CHILDQ | nature-throid | CHEBI:9584 |
| eb_b034a_med3_name_CHILDQ | nature-throid | CHEBI:9584 |
| eb_b035a_med4_name_CHILDQ | nature-throid | CHEBI:9584 |
| eb_b039a_med8_name_CHILDQ | neomycin | CHEBI:7507 |
| eb_b032a_med1_name_CHILDQ | neurontin | CHEBI:42797 |
| eb_b033a_med2_name_CHILDQ | neurontin | CHEBI:42797 |
| eb_b034a_med3_name_CHILDQ | neurontin | CHEBI:42797 |
| eb_b037a_med6_name_CHILDQ | neurontin | CHEBI:42797 |
| eb_b033a_med2_name_CHILDQ | nevirapine | CHEBI:63613 |
| eb_b032a_med1_name_CHILDQ | niacin | CHEBI:15940 |
| eb_b034a_med3_name_CHILDQ | niacin | CHEBI:15940 |
| eb_b039a_med8_name_CHILDQ | niacin | CHEBI:15940 |
| eb_b040a_med9_name_CHILDQ | niacin | CHEBI:15940 |
| eb_b033a_med2_name_CHILDQ | niaspan | CHEBI:15940 |
| eb_b035a_med4_name_CHILDQ | niaspan | CHEBI:15940 |
| eb_b032a_med1_name_CHILDQ | nifedipine | CHEBI:7565 |
| eb_b036a_med5_name_CHILDQ | nifedipine | CHEBI:7565 |
| eb_b040a_med9_name_CHILDQ | nifedipine | CHEBI:7565 |
| eb_b036a_med5_name_CHILDQ | nitrofurantoin | CHEBI:71415 |
| eb_b032a_med1_name_CHILDQ | nitrolingual | CHEBI:28787 |
| eb_b036a_med5_name_CHILDQ | nitrostat | CHEBI:28787 |
| eb_b039a_med8_name_CHILDQ | nitrostat | CHEBI:28787 |
| eb_b032a_med1_name_CHILDQ | norethindrone | CHEBI:7627 |
| eb_b036a_med5_name_CHILDQ | norethindrone | CHEBI:7627 |
| eb_b037a_med6_name_CHILDQ | norethindrone | CHEBI:7627 |
| eb_b038a_med7_name_CHILDQ | norethindrone | CHEBI:7627 |
| eb_b032a_med1_name_CHILDQ | nortriptyline | CHEBI:7640 |
| eb_b033a_med2_name_CHILDQ | nortriptyline | CHEBI:7640 |
| eb_b034a_med3_name_CHILDQ | nortriptyline | CHEBI:7640 |
| eb_b038a_med7_name_CHILDQ | nucynta | CHEBI:135935 |
| eb_b035a_med4_name_CHILDQ | nuvigil | CHEBI:77590 |
| eb_b037a_med6_name_CHILDQ | nystatin | CHEBI:7660 |
| eb_b037a_med6_name_CHILDQ | nystatin | CHEBI:59676 |
| eb_b032a_med1_name_CHILDQ | odefsey | CHEBI:133010 |
| eb_b032a_med1_name_CHILDQ | olmesartan | CHEBI:48416 |
| eb_b033a_med2_name_CHILDQ | olmesartan | CHEBI:48416 |
| eb_b034a_med3_name_CHILDQ | olmesartan | CHEBI:48416 |
| eb_b033a_med2_name_CHILDQ | olmesartan medoxomil | CHEBI:31932 |
| eb_b034a_med3_name_CHILDQ | olmesartan medoxomil | CHEBI:31932 |
| eb_b032a_med1_name_CHILDQ | olopatadine | CHEBI:7769 |
| eb_b035a_med4_name_CHILDQ | olopatadine | CHEBI:7769 |
| eb_b039a_med8_name_CHILDQ | olopatadine | CHEBI:7769 |
| eb_b039a_med8_name_CHILDQ | olopatadine hcl | CHEBI:7769 |
| eb_b039a_med8_name_CHILDQ | omega-3 fatty acids | CHEBI:25681 |
| eb_b033a_med2_name_CHILDQ | omeprazol | CHEBI:7772 |
| eb_b032a_med1_name_CHILDQ | omeprazole | CHEBI:7772 |
| eb_b033a_med2_name_CHILDQ | omeprazole | CHEBI:7772 |
| eb_b034a_med3_name_CHILDQ | omeprazole | CHEBI:7772 |
| eb_b035a_med4_name_CHILDQ | omeprazole | CHEBI:7772 |
| eb_b036a_med5_name_CHILDQ | omeprazole | CHEBI:7772 |
| eb_b037a_med6_name_CHILDQ | omeprazole | CHEBI:7772 |
| eb_b038a_med7_name_CHILDQ | omeprazole | CHEBI:7772 |
| eb_b040a_med9_name_CHILDQ | omeprazole | CHEBI:7772 |
| eb_b041a_med10_name_CHILDQ | omeprazole | CHEBI:7772 |
| eb_b035a_med4_name_CHILDQ | ondansetron | CHEBI:7773 |
| eb_b037a_med6_name_CHILDQ | onglyza | CHEBI:71271 |
| eb_b032a_med1_name_CHILDQ | ophthalmic | CHEBI:66981 |
| eb_b033a_med2_name_CHILDQ | ophthalmic | CHEBI:66981 |
| eb_b034a_med3_name_CHILDQ | ophthalmic | CHEBI:66981 |
| eb_b035a_med4_name_CHILDQ | ophthalmic | CHEBI:66981 |
| eb_b039a_med8_name_CHILDQ | ophthalmic | CHEBI:66981 |
| eb_b032a_med1_name_CHILDQ | oral contraceptive | CHEBI:49325 |
| eb_b033a_med2_name_CHILDQ | osphena | CHEBI:73275 |
| eb_b034a_med3_name_CHILDQ | osphena | CHEBI:73275 |
| eb_b034a_med3_name_CHILDQ | oxalate | CHEBI:132952 |
| eb_b034a_med3_name_CHILDQ | oxalate | CHEBI:30623 |
| eb_b035a_med4_name_CHILDQ | oxcarbazepine | CHEBI:7824 |
| eb_b032a_med1_name_CHILDQ | oxybutynin | CHEBI:7856 |
| eb_b033a_med2_name_CHILDQ | oxybutynin | CHEBI:7856 |
| eb_b034a_med3_name_CHILDQ | oxybutynin | CHEBI:7856 |
| eb_b035a_med4_name_CHILDQ | oxybutynin | CHEBI:7856 |
| eb_b032a_med1_name_CHILDQ | oxybutynin chloride | CHEBI:7857 |
| eb_b035a_med4_name_CHILDQ | oxybutynin chloride | CHEBI:7857 |
| eb_b032a_med1_name_CHILDQ | oxycodone | CHEBI:7852 |
| eb_b033a_med2_name_CHILDQ | oxycodone | CHEBI:7852 |
| eb_b034a_med3_name_CHILDQ | oxycodone | CHEBI:7852 |
| eb_b035a_med4_name_CHILDQ | oxycodone | CHEBI:7852 |
| eb_b036a_med5_name_CHILDQ | oxycodone | CHEBI:7852 |
| eb_b037a_med6_name_CHILDQ | oxycodone | CHEBI:7852 |
| eb_b039a_med8_name_CHILDQ | oxycodone | CHEBI:7852 |
| eb_b040a_med9_name_CHILDQ | oxycodone | CHEBI:7852 |
| eb_b041a_med10_name_CHILDQ | oxycodone | CHEBI:7852 |
| eb_b036a_med5_name_CHILDQ | oxycodone hcl | CHEBI:7859 |
| eb_b039a_med8_name_CHILDQ | oxycodone hcl | CHEBI:7859 |
| eb_b041a_med10_name_CHILDQ | oxycodone hcl | CHEBI:7859 |
| eb_b032a_med1_name_CHILDQ | pamelor | CHEBI:7641 |
| eb_b032a_med1_name_CHILDQ | pantoprazole | CHEBI:7915 |
| eb_b033a_med2_name_CHILDQ | pantoprazole | CHEBI:7915 |
| eb_b034a_med3_name_CHILDQ | pantoprazole | CHEBI:7915 |
| eb_b035a_med4_name_CHILDQ | pantoprazole | CHEBI:7915 |
| eb_b036a_med5_name_CHILDQ | pantoprazole | CHEBI:7915 |
| eb_b037a_med6_name_CHILDQ | pantoprazole | CHEBI:7915 |
| eb_b038a_med7_name_CHILDQ | pantoprazole | CHEBI:7915 |
| eb_b039a_med8_name_CHILDQ | pantoprazole | CHEBI:7915 |
| eb_b041a_med10_name_CHILDQ | pantoprazole | CHEBI:7915 |
| eb_b033a_med2_name_CHILDQ | pantoprazole sodium | CHEBI:50270 |
| eb_b041a_med10_name_CHILDQ | pantoprazole sodium | CHEBI:50270 |
| eb_b032a_med1_name_CHILDQ | paroxetine | CHEBI:7936 |
| eb_b033a_med2_name_CHILDQ | paroxetine | CHEBI:7936 |
| eb_b034a_med3_name_CHILDQ | paroxetine | CHEBI:7936 |
| eb_b033a_med2_name_CHILDQ | paroxetine hcl | CHEBI:7944 |
| eb_b035a_med4_name_CHILDQ | patanol | CHEBI:7769 |
| eb_b032a_med1_name_CHILDQ | paxil | CHEBI:7944 |
| eb_b033a_med2_name_CHILDQ | paxil | CHEBI:7944 |
| eb_b036a_med5_name_CHILDQ | paxil | CHEBI:7944 |
| eb_b033a_med2_name_CHILDQ | pentasa | CHEBI:6775 |
| eb_b034a_med3_name_CHILDQ | pentasa | CHEBI:6775 |
| eb_b034a_med3_name_CHILDQ | pentoxifylline | CHEBI:7986 |
| eb_b035a_med4_name_CHILDQ | pentoxifylline | CHEBI:7986 |
| eb_b032a_med1_name_CHILDQ | pepcid | CHEBI:4975 |
| eb_b033a_med2_name_CHILDQ | pepcid | CHEBI:4975 |
| eb_b034a_med3_name_CHILDQ | pepcid | CHEBI:4975 |
| eb_b035a_med4_name_CHILDQ | pepcid | CHEBI:4975 |
| eb_b036a_med5_name_CHILDQ | pepcid | CHEBI:4975 |
| eb_b037a_med6_name_CHILDQ | pepcid | CHEBI:4975 |
| eb_b038a_med7_name_CHILDQ | pepcid | CHEBI:4975 |
| eb_b039a_med8_name_CHILDQ | pepcid | CHEBI:4975 |
| eb_b040a_med9_name_CHILDQ | pepcid | CHEBI:4975 |
| eb_b037a_med6_name_CHILDQ | phenazopyridine | CHEBI:71416 |
| eb_b032a_med1_name_CHILDQ | phendimetrazine | CHEBI:8059 |
| eb_b032a_med1_name_CHILDQ | phenelzine | CHEBI:8060 |
| eb_b032a_med1_name_CHILDQ | phentermine | CHEBI:8080 |
| eb_b033a_med2_name_CHILDQ | phentermine | CHEBI:8080 |
| eb_b034a_med3_name_CHILDQ | phentermine | CHEBI:8080 |
| eb_b041a_med10_name_CHILDQ | phentermine | CHEBI:8080 |
| eb_b033a_med2_name_CHILDQ | phenylephrine | CHEBI:8093 |
| eb_b040a_med9_name_CHILDQ | phenylephrine | CHEBI:8093 |
| eb_b034a_med3_name_CHILDQ | pilocarpine | CHEBI:39462 |
| eb_b036a_med5_name_CHILDQ | pilocarpine | CHEBI:39462 |
| eb_b038a_med7_name_CHILDQ | pilocarpine | CHEBI:39462 |
| eb_b034a_med3_name_CHILDQ | pilocarpine hcl | CHEBI:141029 |
| eb_b035a_med4_name_CHILDQ | pindolol | CHEBI:8214 |
| eb_b033a_med2_name_CHILDQ | pioglitazone | CHEBI:8228 |
| eb_b035a_med4_name_CHILDQ | pioglitazone | CHEBI:8228 |
| eb_b032a_med1_name_CHILDQ | plavix | CHEBI:3759 |
| eb_b033a_med2_name_CHILDQ | plavix | CHEBI:3759 |
| eb_b034a_med3_name_CHILDQ | plavix | CHEBI:3759 |
| eb_b035a_med4_name_CHILDQ | plavix | CHEBI:3759 |
| eb_b036a_med5_name_CHILDQ | plavix | CHEBI:3759 |
| eb_b038a_med7_name_CHILDQ | plavix | CHEBI:3759 |
| eb_b039a_med8_name_CHILDQ | plavix | CHEBI:3759 |
| eb_b032a_med1_name_CHILDQ | potassium | CHEBI:26216 |
| eb_b033a_med2_name_CHILDQ | potassium | CHEBI:26216 |
| eb_b034a_med3_name_CHILDQ | potassium | CHEBI:26216 |
| eb_b035a_med4_name_CHILDQ | potassium | CHEBI:26216 |
| eb_b036a_med5_name_CHILDQ | potassium | CHEBI:26216 |
| eb_b037a_med6_name_CHILDQ | potassium | CHEBI:26216 |
| eb_b038a_med7_name_CHILDQ | potassium | CHEBI:26216 |
| eb_b039a_med8_name_CHILDQ | potassium | CHEBI:26216 |
| eb_b033a_med2_name_CHILDQ | potassium chloride | CHEBI:32588 |
| eb_b034a_med3_name_CHILDQ | potassium chloride | CHEBI:32588 |
| eb_b035a_med4_name_CHILDQ | potassium chloride | CHEBI:32588 |
| eb_b032a_med1_name_CHILDQ | potassium gluconate | CHEBI:32032 |
| eb_b033a_med2_name_CHILDQ | pradaxa | CHEBI:70743 |
| eb_b035a_med4_name_CHILDQ | pradaxa | CHEBI:70743 |
| eb_b037a_med6_name_CHILDQ | pradaxa | CHEBI:70743 |
| eb_b033a_med2_name_CHILDQ | pramipexole | CHEBI:8356 |
| eb_b034a_med3_name_CHILDQ | pramipexole | CHEBI:8356 |
| eb_b035a_med4_name_CHILDQ | pramipexole | CHEBI:8356 |
| eb_b037a_med6_name_CHILDQ | pramipexole | CHEBI:8356 |
| eb_b038a_med7_name_CHILDQ | pramipexole | CHEBI:8356 |
| eb_b039a_med8_name_CHILDQ | pramipexole | CHEBI:8356 |
| eb_b033a_med2_name_CHILDQ | pramipexole dihydrochloride | CHEBI:51148 |
| eb_b035a_med4_name_CHILDQ | pramipexole dihydrochloride | CHEBI:51148 |
| eb_b032a_med1_name_CHILDQ | pravastatin | CHEBI:63618 |
| eb_b033a_med2_name_CHILDQ | pravastatin | CHEBI:63618 |
| eb_b034a_med3_name_CHILDQ | pravastatin | CHEBI:63618 |
| eb_b035a_med4_name_CHILDQ | pravastatin | CHEBI:63618 |
| eb_b036a_med5_name_CHILDQ | pravastatin | CHEBI:63618 |
| eb_b037a_med6_name_CHILDQ | pravastatin | CHEBI:63618 |
| eb_b038a_med7_name_CHILDQ | pravastatin | CHEBI:63618 |
| eb_b039a_med8_name_CHILDQ | pravastatin | CHEBI:63618 |
| eb_b041a_med10_name_CHILDQ | pravastatin | CHEBI:63618 |
| eb_b033a_med2_name_CHILDQ | pravastatin sodium | CHEBI:8361 |
| eb_b034a_med3_name_CHILDQ | pravastatin sodium | CHEBI:8361 |
| eb_b035a_med4_name_CHILDQ | pravastatin sodium | CHEBI:8361 |
| eb_b038a_med7_name_CHILDQ | pravastatin sodium | CHEBI:8361 |
| eb_b040a_med9_name_CHILDQ | prazosin | CHEBI:8364 |
| eb_b032a_med1_name_CHILDQ | prednisolone | CHEBI:8378 |
| eb_b036a_med5_name_CHILDQ | prednisolone | CHEBI:8378 |
| eb_b032a_med1_name_CHILDQ | prednisolone acetate | CHEBI:8380 |
| eb_b032a_med1_name_CHILDQ | prednisone | CHEBI:8382 |
| eb_b033a_med2_name_CHILDQ | prednisone | CHEBI:8382 |
| eb_b034a_med3_name_CHILDQ | prednisone | CHEBI:8382 |
| eb_b036a_med5_name_CHILDQ | prednisone | CHEBI:8382 |
| eb_b037a_med6_name_CHILDQ | prednisone | CHEBI:8382 |
| eb_b038a_med7_name_CHILDQ | prednisone | CHEBI:8382 |
| eb_b039a_med8_name_CHILDQ | prednisone | CHEBI:8382 |
| eb_b032a_med1_name_CHILDQ | premarin | CHEBI:8389 |
| eb_b033a_med2_name_CHILDQ | premarin | CHEBI:8389 |
| eb_b037a_med6_name_CHILDQ | premarin | CHEBI:8389 |
| eb_b038a_med7_name_CHILDQ | premarin | CHEBI:8389 |
| eb_b041a_med10_name_CHILDQ | premarin | CHEBI:8389 |
| eb_b032a_med1_name_CHILDQ | prevacid | CHEBI:6375 |
| eb_b033a_med2_name_CHILDQ | prevacid | CHEBI:6375 |
| eb_b034a_med3_name_CHILDQ | prevacid | CHEBI:6375 |
| eb_b036a_med5_name_CHILDQ | prevacid | CHEBI:6375 |
| eb_b032a_med1_name_CHILDQ | primidone | CHEBI:8412 |
| eb_b038a_med7_name_CHILDQ | probenecid | CHEBI:8426 |
| eb_b032a_med1_name_CHILDQ | progesterone | CHEBI:17026 |
| eb_b033a_med2_name_CHILDQ | progesterone | CHEBI:17026 |
| eb_b034a_med3_name_CHILDQ | progesterone | CHEBI:17026 |
| eb_b037a_med6_name_CHILDQ | progesterone | CHEBI:17026 |
| eb_b038a_med7_name_CHILDQ | progesterone | CHEBI:17026 |
| eb_b032a_med1_name_CHILDQ | progestin | CHEBI:59826 |
| eb_b032a_med1_name_CHILDQ | prograf | CHEBI:61049 |
| eb_b036a_med5_name_CHILDQ | prograf | CHEBI:61049 |
| eb_b033a_med2_name_CHILDQ | promethazine | CHEBI:8461 |
| eb_b038a_med7_name_CHILDQ | promethazine | CHEBI:8461 |
| eb_b033a_med2_name_CHILDQ | promethazine hcl | CHEBI:8462 |
| eb_b033a_med2_name_CHILDQ | propafenone | CHEBI:63619 |
| eb_b032a_med1_name_CHILDQ | propanolol | CHEBI:8499 |
| eb_b032a_med1_name_CHILDQ | propionate | CHEBI:17272 |
| eb_b033a_med2_name_CHILDQ | propionate | CHEBI:17272 |
| eb_b034a_med3_name_CHILDQ | propionate | CHEBI:17272 |
| eb_b035a_med4_name_CHILDQ | propionate | CHEBI:17272 |
| eb_b036a_med5_name_CHILDQ | propionate | CHEBI:17272 |
| eb_b037a_med6_name_CHILDQ | propionate | CHEBI:17272 |
| eb_b038a_med7_name_CHILDQ | propionate | CHEBI:17272 |
| eb_b040a_med9_name_CHILDQ | propionate | CHEBI:17272 |
| eb_b041a_med10_name_CHILDQ | propionate | CHEBI:17272 |
| eb_b032a_med1_name_CHILDQ | propranolol | CHEBI:8499 |
| eb_b033a_med2_name_CHILDQ | propranolol | CHEBI:8499 |
| eb_b034a_med3_name_CHILDQ | propranolol | CHEBI:8499 |
| eb_b035a_med4_name_CHILDQ | propranolol | CHEBI:8499 |
| eb_b036a_med5_name_CHILDQ | propranolol | CHEBI:8499 |
| eb_b041a_med10_name_CHILDQ | propranolol | CHEBI:8499 |
| eb_b032a_med1_name_CHILDQ | protonix | CHEBI:50270 |
| eb_b033a_med2_name_CHILDQ | protonix | CHEBI:50270 |
| eb_b034a_med3_name_CHILDQ | protonix | CHEBI:50270 |
| eb_b035a_med4_name_CHILDQ | protonix | CHEBI:50270 |
| eb_b037a_med6_name_CHILDQ | protonix | CHEBI:50270 |
| eb_b032a_med1_name_CHILDQ | proventil | CHEBI:2549 |
| eb_b033a_med2_name_CHILDQ | proventil | CHEBI:2549 |
| eb_b037a_med6_name_CHILDQ | proventil | CHEBI:2549 |
| eb_b032a_med1_name_CHILDQ | provigil | CHEBI:31859 |
| eb_b041a_med10_name_CHILDQ | provigil | CHEBI:31859 |
| eb_b032a_med1_name_CHILDQ | prozac | CHEBI:5118 |
| eb_b033a_med2_name_CHILDQ | prozac | CHEBI:5118 |
| eb_b034a_med3_name_CHILDQ | prozac | CHEBI:5118 |
| eb_b035a_med4_name_CHILDQ | prozac | CHEBI:5118 |
| eb_b036a_med5_name_CHILDQ | prozac | CHEBI:5118 |
| eb_b034a_med3_name_CHILDQ | quetiapine | CHEBI:8707 |
| eb_b036a_med5_name_CHILDQ | quetiapine | CHEBI:8707 |
| eb_b039a_med8_name_CHILDQ | quetiapine | CHEBI:8707 |
| eb_b034a_med3_name_CHILDQ | quetiapine fumarate | CHEBI:8708 |
| eb_b039a_med8_name_CHILDQ | quetiapine fumarate | CHEBI:8708 |
| eb_b032a_med1_name_CHILDQ | quinapril | CHEBI:8713 |
| eb_b034a_med3_name_CHILDQ | quinapril | CHEBI:8713 |
| eb_b033a_med2_name_CHILDQ | rabeprazole | CHEBI:8768 |
| eb_b033a_med2_name_CHILDQ | raloxifene | CHEBI:8772 |
| eb_b035a_med4_name_CHILDQ | raloxifene | CHEBI:8772 |
| eb_b036a_med5_name_CHILDQ | raloxifene | CHEBI:8772 |
| eb_b032a_med1_name_CHILDQ | ramipril | CHEBI:8774 |
| eb_b033a_med2_name_CHILDQ | ramipril | CHEBI:8774 |
| eb_b034a_med3_name_CHILDQ | ramipril | CHEBI:8774 |
| eb_b035a_med4_name_CHILDQ | ramipril | CHEBI:8774 |
| eb_b038a_med7_name_CHILDQ | ramipril | CHEBI:8774 |
| eb_b032a_med1_name_CHILDQ | ranexa | CHEBI:87681 |
| eb_b033a_med2_name_CHILDQ | ranexa | CHEBI:87681 |
| eb_b032a_med1_name_CHILDQ | ranitidine | CHEBI:8776 |
| eb_b033a_med2_name_CHILDQ | ranitidine | CHEBI:8776 |
| eb_b034a_med3_name_CHILDQ | ranitidine | CHEBI:8776 |
| eb_b035a_med4_name_CHILDQ | ranitidine | CHEBI:8776 |
| eb_b036a_med5_name_CHILDQ | ranitidine | CHEBI:8776 |
| eb_b037a_med6_name_CHILDQ | ranitidine | CHEBI:8776 |
| eb_b038a_med7_name_CHILDQ | ranitidine | CHEBI:8776 |
| eb_b039a_med8_name_CHILDQ | ranitidine | CHEBI:8776 |
| eb_b040a_med9_name_CHILDQ | ranitidine | CHEBI:8776 |
| eb_b035a_med4_name_CHILDQ | ranitidine hcl | CHEBI:8777 |
| eb_b038a_med7_name_CHILDQ | ranitidine hcl | CHEBI:8777 |
| eb_b032a_med1_name_CHILDQ | ranolazine | CHEBI:87681 |
| eb_b033a_med2_name_CHILDQ | rapaflo | CHEBI:135929 |
| eb_b038a_med7_name_CHILDQ | rapaflo | CHEBI:135929 |
| eb_b032a_med1_name_CHILDQ | relafen | CHEBI:7443 |
| eb_b032a_med1_name_CHILDQ | relpax | CHEBI:61176 |
| eb_b033a_med2_name_CHILDQ | relpax | CHEBI:61176 |
| eb_b034a_med3_name_CHILDQ | relpax | CHEBI:61176 |
| eb_b038a_med7_name_CHILDQ | relpax | CHEBI:61176 |
| eb_b041a_med10_name_CHILDQ | relpax | CHEBI:61176 |
| eb_b032a_med1_name_CHILDQ | revlimid | CHEBI:63791 |
| eb_b039a_med8_name_CHILDQ | revlimid | CHEBI:63791 |
| eb_b032a_med1_name_CHILDQ | rexulti | CHEBI:134716 |
| eb_b033a_med2_name_CHILDQ | reyataz | CHEBI:31243 |
| eb_b037a_med6_name_CHILDQ | rifampin | CHEBI:28077 |
| eb_b032a_med1_name_CHILDQ | risedronate | CHEBI:8869 |
| eb_b036a_med5_name_CHILDQ | risedronate | CHEBI:8869 |
| eb_b033a_med2_name_CHILDQ | risperdal | CHEBI:8871 |
| eb_b032a_med1_name_CHILDQ | ritalin | CHEBI:31836 |
| eb_b034a_med3_name_CHILDQ | ritalin | CHEBI:31836 |
| eb_b035a_med4_name_CHILDQ | ritalin | CHEBI:31836 |
| eb_b037a_med6_name_CHILDQ | ritalin | CHEBI:31836 |
| eb_b033a_med2_name_CHILDQ | rivaroxaban | CHEBI:68579 |
| eb_b035a_med4_name_CHILDQ | rivastigmine | CHEBI:8874 |
| eb_b032a_med1_name_CHILDQ | rizatriptan | CHEBI:48273 |
| eb_b034a_med3_name_CHILDQ | rizatriptan | CHEBI:48273 |
| eb_b035a_med4_name_CHILDQ | rizatriptan | CHEBI:48273 |
| eb_b032a_med1_name_CHILDQ | ropinirole | CHEBI:8888 |
| eb_b034a_med3_name_CHILDQ | ropinirole | CHEBI:8888 |
| eb_b037a_med6_name_CHILDQ | ropinirole | CHEBI:8888 |
| eb_b032a_med1_name_CHILDQ | rosuvastatin | CHEBI:38545 |
| eb_b033a_med2_name_CHILDQ | rosuvastatin | CHEBI:38545 |
| eb_b034a_med3_name_CHILDQ | rosuvastatin | CHEBI:38545 |
| eb_b035a_med4_name_CHILDQ | rosuvastatin | CHEBI:38545 |
| eb_b036a_med5_name_CHILDQ | rosuvastatin | CHEBI:38545 |
| eb_b032a_med1_name_CHILDQ | rosuvastatin calcium | CHEBI:77249 |
| eb_b033a_med2_name_CHILDQ | rosuvastatin calcium | CHEBI:77249 |
| eb_b034a_med3_name_CHILDQ | rosuvastatin calcium | CHEBI:77249 |
| eb_b035a_med4_name_CHILDQ | rosuvastatin calcium | CHEBI:77249 |
| eb_b037a_med6_name_CHILDQ | s-adenosylmethionine | CHEBI:15414 |
| eb_b034a_med3_name_CHILDQ | sectral | CHEBI:2380 |
| eb_b036a_med5_name_CHILDQ | serevent | CHEBI:9012 |
| eb_b032a_med1_name_CHILDQ | seroquel | CHEBI:8708 |
| eb_b034a_med3_name_CHILDQ | seroquel | CHEBI:8708 |
| eb_b032a_med1_name_CHILDQ | sertraline | CHEBI:9123 |
| eb_b033a_med2_name_CHILDQ | sertraline | CHEBI:9123 |
| eb_b034a_med3_name_CHILDQ | sertraline | CHEBI:9123 |
| eb_b035a_med4_name_CHILDQ | sertraline | CHEBI:9123 |
| eb_b036a_med5_name_CHILDQ | sertraline | CHEBI:9123 |
| eb_b037a_med6_name_CHILDQ | sertraline | CHEBI:9123 |
| eb_b038a_med7_name_CHILDQ | sertraline | CHEBI:9123 |
| eb_b039a_med8_name_CHILDQ | sertraline | CHEBI:9123 |
| eb_b032a_med1_name_CHILDQ | sertraline hcl | CHEBI:9124 |
| eb_b033a_med2_name_CHILDQ | sertraline hcl | CHEBI:9124 |
| eb_b034a_med3_name_CHILDQ | sertraline hcl | CHEBI:9124 |
| eb_b035a_med4_name_CHILDQ | sertraline hcl | CHEBI:9124 |
| eb_b034a_med3_name_CHILDQ | sildenafil | CHEBI:9139 |
| eb_b036a_med5_name_CHILDQ | sildenafil | CHEBI:9139 |
| eb_b038a_med7_name_CHILDQ | sildenafil | CHEBI:9139 |
| eb_b038a_med7_name_CHILDQ | sildenafil citrate | CHEBI:58987 |
| eb_b037a_med6_name_CHILDQ | silver sulfadiazine | CHEBI:9142 |
| eb_b032a_med1_name_CHILDQ | simvastatin | CHEBI:9150 |
| eb_b033a_med2_name_CHILDQ | simvastatin | CHEBI:9150 |
| eb_b034a_med3_name_CHILDQ | simvastatin | CHEBI:9150 |
| eb_b035a_med4_name_CHILDQ | simvastatin | CHEBI:9150 |
| eb_b036a_med5_name_CHILDQ | simvastatin | CHEBI:9150 |
| eb_b037a_med6_name_CHILDQ | simvastatin | CHEBI:9150 |
| eb_b038a_med7_name_CHILDQ | simvastatin | CHEBI:9150 |
| eb_b041a_med10_name_CHILDQ | simvastatin | CHEBI:9150 |
| eb_b032a_med1_name_CHILDQ | simvastatine | CHEBI:9150 |
| eb_b032a_med1_name_CHILDQ | singulair | CHEBI:6993 |
| eb_b033a_med2_name_CHILDQ | singulair | CHEBI:6993 |
| eb_b034a_med3_name_CHILDQ | singulair | CHEBI:6993 |
| eb_b037a_med6_name_CHILDQ | singulair | CHEBI:6993 |
| eb_b032a_med1_name_CHILDQ | singular | CHEBI:6993 |
| eb_b033a_med2_name_CHILDQ | singular | CHEBI:6993 |
| eb_b034a_med3_name_CHILDQ | singular | CHEBI:6993 |
| eb_b037a_med6_name_CHILDQ | singular | CHEBI:6993 |
| eb_b038a_med7_name_CHILDQ | singular | CHEBI:6993 |
| eb_b032a_med1_name_CHILDQ | sitosterol | CHEBI:27693 |
| eb_b036a_med5_name_CHILDQ | sodium bicarbonate | CHEBI:32139 |
| eb_b033a_med2_name_CHILDQ | sodium chloride | CHEBI:26710 |
| eb_b038a_med7_name_CHILDQ | sodium chloride | CHEBI:26710 |
| eb_b036a_med5_name_CHILDQ | sodium sulfacetamide | CHEBI:63858 |
| eb_b038a_med7_name_CHILDQ | solifenacin | CHEBI:135530 |
| eb_b032a_med1_name_CHILDQ | sotalol | CHEBI:63622 |
| eb_b033a_med2_name_CHILDQ | sotalol | CHEBI:63622 |
| eb_b035a_med4_name_CHILDQ | sotalol | CHEBI:63622 |
| eb_b036a_med5_name_CHILDQ | sotalol | CHEBI:63622 |
| eb_b033a_med2_name_CHILDQ | sotalol hcl | CHEBI:9207 |
| eb_b035a_med4_name_CHILDQ | sotalol hcl | CHEBI:9207 |
| eb_b032a_med1_name_CHILDQ | spironolactone | CHEBI:9241 |
| eb_b033a_med2_name_CHILDQ | spironolactone | CHEBI:9241 |
| eb_b034a_med3_name_CHILDQ | spironolactone | CHEBI:9241 |
| eb_b035a_med4_name_CHILDQ | spironolactone | CHEBI:9241 |
| eb_b037a_med6_name_CHILDQ | spironolactone | CHEBI:9241 |
| eb_b038a_med7_name_CHILDQ | spironolactone | CHEBI:9241 |
| eb_b039a_med8_name_CHILDQ | spironolactone | CHEBI:9241 |
| eb_b040a_med9_name_CHILDQ | spironolactone | CHEBI:9241 |
| eb_b032a_med1_name_CHILDQ | statin | CHEBI:87631 |
| eb_b033a_med2_name_CHILDQ | statin | CHEBI:87631 |
| eb_b034a_med3_name_CHILDQ | statin | CHEBI:87631 |
| eb_b034a_med3_name_CHILDQ | sucralfate | CHEBI:9313 |
| eb_b035a_med4_name_CHILDQ | sucralfate | CHEBI:9313 |
| eb_b036a_med5_name_CHILDQ | sulfacetamide | CHEBI:63845 |
| eb_b037a_med6_name_CHILDQ | sulfadiazine | CHEBI:9328 |
| eb_b032a_med1_name_CHILDQ | sulfasalazine | CHEBI:9334 |
| eb_b032a_med1_name_CHILDQ | sumatriptan | CHEBI:10650 |
| eb_b033a_med2_name_CHILDQ | sumatriptan | CHEBI:10650 |
| eb_b034a_med3_name_CHILDQ | sumatriptan | CHEBI:10650 |
| eb_b035a_med4_name_CHILDQ | sumatriptan | CHEBI:10650 |
| eb_b037a_med6_name_CHILDQ | sumatriptan | CHEBI:10650 |
| eb_b039a_med8_name_CHILDQ | symlin | CHEBI:135922 |
| eb_b032a_med1_name_CHILDQ | synthroid | CHEBI:6446 |
| eb_b033a_med2_name_CHILDQ | synthroid | CHEBI:6446 |
| eb_b034a_med3_name_CHILDQ | synthroid | CHEBI:6446 |
| eb_b035a_med4_name_CHILDQ | synthroid | CHEBI:6446 |
| eb_b036a_med5_name_CHILDQ | synthroid | CHEBI:6446 |
| eb_b037a_med6_name_CHILDQ | synthroid | CHEBI:6446 |
| eb_b039a_med8_name_CHILDQ | synthroid | CHEBI:6446 |
| eb_b040a_med9_name_CHILDQ | synthroid | CHEBI:6446 |
| eb_b033a_med2_name_CHILDQ | tacrolimus | CHEBI:61049 |
| eb_b034a_med3_name_CHILDQ | tadalafil | CHEBI:71940 |
| eb_b037a_med6_name_CHILDQ | tadalafil | CHEBI:71940 |
| eb_b039a_med8_name_CHILDQ | tadalafil | CHEBI:71940 |
| eb_b035a_med4_name_CHILDQ | tagamet | CHEBI:50362 |
| eb_b032a_med1_name_CHILDQ | tamoxifen | CHEBI:41774 |
| eb_b033a_med2_name_CHILDQ | tamoxifen | CHEBI:41774 |
| eb_b034a_med3_name_CHILDQ | tamoxifen | CHEBI:41774 |
| eb_b032a_med1_name_CHILDQ | tamsulosin | CHEBI:9398 |
| eb_b033a_med2_name_CHILDQ | tamsulosin | CHEBI:9398 |
| eb_b034a_med3_name_CHILDQ | tamsulosin | CHEBI:9398 |
| eb_b035a_med4_name_CHILDQ | tamsulosin | CHEBI:9398 |
| eb_b037a_med6_name_CHILDQ | tamsulosin | CHEBI:9398 |
| eb_b038a_med7_name_CHILDQ | tamsulosin | CHEBI:9398 |
| eb_b041a_med10_name_CHILDQ | tamsulosin | CHEBI:9398 |
| eb_b034a_med3_name_CHILDQ | tanzeum | CHEBI:78425 |
| eb_b035a_med4_name_CHILDQ | tanzeum | CHEBI:78425 |
| eb_b032a_med1_name_CHILDQ | tartrate | CHEBI:132950 |
| eb_b033a_med2_name_CHILDQ | tartrate | CHEBI:132950 |
| eb_b034a_med3_name_CHILDQ | tartrate | CHEBI:132950 |
| eb_b035a_med4_name_CHILDQ | tartrate | CHEBI:132950 |
| eb_b036a_med5_name_CHILDQ | tartrate | CHEBI:132950 |
| eb_b038a_med7_name_CHILDQ | tartrate | CHEBI:132950 |
| eb_b032a_med1_name_CHILDQ | tecfidera | CHEBI:76004 |
| eb_b032a_med1_name_CHILDQ | telmisartan | CHEBI:9434 |
| eb_b034a_med3_name_CHILDQ | telmisartan | CHEBI:9434 |
| eb_b035a_med4_name_CHILDQ | telmisartan | CHEBI:9434 |
| eb_b034a_med3_name_CHILDQ | temazepam | CHEBI:9435 |
| eb_b032a_med1_name_CHILDQ | terazosin | CHEBI:9445 |
| eb_b032a_med1_name_CHILDQ | terbinafine | CHEBI:9448 |
| eb_b032a_med1_name_CHILDQ | terbinafine hcl | CHEBI:77614 |
| eb_b032a_med1_name_CHILDQ | testosteron | CHEBI:17347 |
| eb_b032a_med1_name_CHILDQ | testosterone | CHEBI:17347 |
| eb_b033a_med2_name_CHILDQ | testosterone | CHEBI:17347 |
| eb_b035a_med4_name_CHILDQ | testosterone | CHEBI:17347 |
| eb_b036a_med5_name_CHILDQ | testosterone | CHEBI:17347 |
| eb_b037a_med6_name_CHILDQ | testosterone | CHEBI:17347 |
| eb_b038a_med7_name_CHILDQ | testosterone | CHEBI:17347 |
| eb_b039a_med8_name_CHILDQ | testosterone | CHEBI:17347 |
| eb_b035a_med4_name_CHILDQ | testosterone cypionate | CHEBI:9463 |
| eb_b038a_med7_name_CHILDQ | testosterone cypionate | CHEBI:9463 |
| eb_b032a_med1_name_CHILDQ | theanine | CHEBI:58128 |
| eb_b032a_med1_name_CHILDQ | theophylline | CHEBI:28177 |
| eb_b032a_med1_name_CHILDQ | thyroid hormone | CHEBI:60311 |
| eb_b032a_med1_name_CHILDQ | thyroxine | CHEBI:30660 |
| eb_b033a_med2_name_CHILDQ | thyroxine | CHEBI:30660 |
| eb_b034a_med3_name_CHILDQ | thyroxine | CHEBI:30660 |
| eb_b035a_med4_name_CHILDQ | thyroxine | CHEBI:30660 |
| eb_b036a_med5_name_CHILDQ | thyroxine | CHEBI:30660 |
| eb_b036a_med5_name_CHILDQ | ticagrelor | CHEBI:68558 |
| eb_b032a_med1_name_CHILDQ | tikosyn | CHEBI:4681 |
| eb_b033a_med2_name_CHILDQ | timolol | CHEBI:39465 |
| eb_b035a_med4_name_CHILDQ | timolol | CHEBI:39465 |
| eb_b036a_med5_name_CHILDQ | timolol | CHEBI:39465 |
| eb_b033a_med2_name_CHILDQ | timolol maleate | CHEBI:9600 |
| eb_b035a_med4_name_CHILDQ | timolol maleate | CHEBI:9600 |
| eb_b033a_med2_name_CHILDQ | tizanidine | CHEBI:63629 |
| eb_b035a_med4_name_CHILDQ | tizanidine | CHEBI:63629 |
| eb_b036a_med5_name_CHILDQ | tizanidine | CHEBI:63629 |
| eb_b037a_med6_name_CHILDQ | tizanidine | CHEBI:63629 |
| eb_b034a_med3_name_CHILDQ | tolterodine | CHEBI:9622 |
| eb_b036a_med5_name_CHILDQ | tolterodine | CHEBI:9622 |
| eb_b032a_med1_name_CHILDQ | topamax | CHEBI:63631 |
| eb_b033a_med2_name_CHILDQ | topamax | CHEBI:63631 |
| eb_b037a_med6_name_CHILDQ | topamax | CHEBI:63631 |
| eb_b032a_med1_name_CHILDQ | topiramate | CHEBI:63631 |
| eb_b033a_med2_name_CHILDQ | topiramate | CHEBI:63631 |
| eb_b035a_med4_name_CHILDQ | topiramate | CHEBI:63631 |
| eb_b036a_med5_name_CHILDQ | topiramate | CHEBI:63631 |
| eb_b037a_med6_name_CHILDQ | topiramate | CHEBI:63631 |
| eb_b038a_med7_name_CHILDQ | topiramate | CHEBI:63631 |
| eb_b032a_med1_name_CHILDQ | torsemide | CHEBI:9637 |
| eb_b037a_med6_name_CHILDQ | torsemide | CHEBI:9637 |
| eb_b036a_med5_name_CHILDQ | toviaz | CHEBI:135920 |
| eb_b040a_med9_name_CHILDQ | toviaz | CHEBI:135920 |
| eb_b032a_med1_name_CHILDQ | tradjenta | CHEBI:68610 |
| eb_b032a_med1_name_CHILDQ | tramadol | CHEBI:9648 |
| eb_b033a_med2_name_CHILDQ | tramadol | CHEBI:9648 |
| eb_b034a_med3_name_CHILDQ | tramadol | CHEBI:9648 |
| eb_b035a_med4_name_CHILDQ | tramadol | CHEBI:9648 |
| eb_b036a_med5_name_CHILDQ | tramadol | CHEBI:9648 |
| eb_b037a_med6_name_CHILDQ | tramadol | CHEBI:9648 |
| eb_b038a_med7_name_CHILDQ | tramadol | CHEBI:9648 |
| eb_b039a_med8_name_CHILDQ | tramadol | CHEBI:9648 |
| eb_b040a_med9_name_CHILDQ | tramadol | CHEBI:9648 |
| eb_b032a_med1_name_CHILDQ | tramadol hcl | CHEBI:32250 |
| eb_b033a_med2_name_CHILDQ | tramadol hcl | CHEBI:32250 |
| eb_b034a_med3_name_CHILDQ | tramadol hcl | CHEBI:32250 |
| eb_b036a_med5_name_CHILDQ | tramadol hcl | CHEBI:32250 |
| eb_b038a_med7_name_CHILDQ | tramadol hcl | CHEBI:32250 |
| eb_b035a_med4_name_CHILDQ | tranexamic acid | CHEBI:48669 |
| eb_b032a_med1_name_CHILDQ | travatan | CHEBI:746859 |
| eb_b041a_med10_name_CHILDQ | travatan | CHEBI:746859 |
| eb_b032a_med1_name_CHILDQ | travatan z | CHEBI:746859 |
| eb_b034a_med3_name_CHILDQ | travoprost | CHEBI:746859 |
| eb_b032a_med1_name_CHILDQ | trazodone | CHEBI:9654 |
| eb_b033a_med2_name_CHILDQ | trazodone | CHEBI:9654 |
| eb_b034a_med3_name_CHILDQ | trazodone | CHEBI:9654 |
| eb_b035a_med4_name_CHILDQ | trazodone | CHEBI:9654 |
| eb_b036a_med5_name_CHILDQ | trazodone | CHEBI:9654 |
| eb_b037a_med6_name_CHILDQ | trazodone | CHEBI:9654 |
| eb_b038a_med7_name_CHILDQ | trazodone | CHEBI:9654 |
| eb_b039a_med8_name_CHILDQ | trazodone | CHEBI:9654 |
| eb_b040a_med9_name_CHILDQ | trazodone | CHEBI:9654 |
| eb_b041a_med10_name_CHILDQ | trazodone | CHEBI:9654 |
| eb_b032a_med1_name_CHILDQ | tretinoin | CHEBI:15367 |
| eb_b033a_med2_name_CHILDQ | tretinoin | CHEBI:15367 |
| eb_b037a_med6_name_CHILDQ | tretinoin | CHEBI:15367 |
| eb_b039a_med8_name_CHILDQ | tretinoin | CHEBI:15367 |
| eb_b033a_med2_name_CHILDQ | triamcinolone | CHEBI:9667 |
| eb_b036a_med5_name_CHILDQ | triamcinolone | CHEBI:9667 |
| eb_b039a_med8_name_CHILDQ | triamcinolone | CHEBI:9667 |
| eb_b040a_med9_name_CHILDQ | triamcinolone | CHEBI:9667 |
| eb_b033a_med2_name_CHILDQ | triamcinolone acetonide | CHEBI:71418 |
| eb_b036a_med5_name_CHILDQ | triamcinolone acetonide | CHEBI:71418 |
| eb_b032a_med1_name_CHILDQ | triamterene | CHEBI:9671 |
| eb_b033a_med2_name_CHILDQ | triamterene | CHEBI:9671 |
| eb_b034a_med3_name_CHILDQ | triamterene | CHEBI:9671 |
| eb_b035a_med4_name_CHILDQ | triamterene | CHEBI:9671 |
| eb_b038a_med7_name_CHILDQ | triamterene | CHEBI:9671 |
| eb_b041a_med10_name_CHILDQ | triamterene | CHEBI:9671 |
| eb_b032a_med1_name_CHILDQ | tricor | CHEBI:5001 |
| eb_b035a_med4_name_CHILDQ | tricor | CHEBI:5001 |
| eb_b036a_med5_name_CHILDQ | tricor | CHEBI:5001 |
| eb_b032a_med1_name_CHILDQ | trospium | CHEBI:145791 |
| eb_b032a_med1_name_CHILDQ | tylenol | CHEBI:46195 |
| eb_b033a_med2_name_CHILDQ | tylenol | CHEBI:46195 |
| eb_b034a_med3_name_CHILDQ | tylenol | CHEBI:46195 |
| eb_b035a_med4_name_CHILDQ | tylenol | CHEBI:46195 |
| eb_b036a_med5_name_CHILDQ | tylenol | CHEBI:46195 |
| eb_b037a_med6_name_CHILDQ | tylenol | CHEBI:46195 |
| eb_b038a_med7_name_CHILDQ | tylenol | CHEBI:46195 |
| eb_b039a_med8_name_CHILDQ | tylenol | CHEBI:46195 |
| eb_b040a_med9_name_CHILDQ | tylenol | CHEBI:46195 |
| eb_b041a_med10_name_CHILDQ | tylenol | CHEBI:46195 |
| eb_b032a_med1_name_CHILDQ | ubiquinol | CHEBI:17976 |
| eb_b032a_med1_name_CHILDQ | uloric | CHEBI:31596 |
| eb_b033a_med2_name_CHILDQ | uloric | CHEBI:31596 |
| eb_b034a_med3_name_CHILDQ | uloric | CHEBI:31596 |
| eb_b035a_med4_name_CHILDQ | uloric | CHEBI:31596 |
| eb_b032a_med1_name_CHILDQ | valacyclovir | CHEBI:35854 |
| eb_b033a_med2_name_CHILDQ | valacyclovir | CHEBI:35854 |
| eb_b034a_med3_name_CHILDQ | valacyclovir | CHEBI:35854 |
| eb_b035a_med4_name_CHILDQ | valacyclovir | CHEBI:35854 |
| eb_b037a_med6_name_CHILDQ | valacyclovir | CHEBI:35854 |
| eb_b041a_med10_name_CHILDQ | valacyclovir | CHEBI:35854 |
| eb_b038a_med7_name_CHILDQ | valganciclovir | CHEBI:63635 |
| eb_b032a_med1_name_CHILDQ | valium | CHEBI:49575 |
| eb_b035a_med4_name_CHILDQ | valium | CHEBI:49575 |
| eb_b032a_med1_name_CHILDQ | valproic acid | CHEBI:39867 |
| eb_b033a_med2_name_CHILDQ | valproic acid | CHEBI:39867 |
| eb_b032a_med1_name_CHILDQ | valsartan | CHEBI:9927 |
| eb_b033a_med2_name_CHILDQ | valsartan | CHEBI:9927 |
| eb_b034a_med3_name_CHILDQ | valsartan | CHEBI:9927 |
| eb_b035a_med4_name_CHILDQ | valsartan | CHEBI:9927 |
| eb_b036a_med5_name_CHILDQ | valsartan | CHEBI:9927 |
| eb_b038a_med7_name_CHILDQ | valsartan | CHEBI:9927 |
| eb_b040a_med9_name_CHILDQ | valsartan | CHEBI:9927 |
| eb_b035a_med4_name_CHILDQ | vancomycin | CHEBI:28001 |
| eb_b037a_med6_name_CHILDQ | vascepa | CHEBI:84883 |
| eb_b032a_med1_name_CHILDQ | venlafaxine | CHEBI:9943 |
| eb_b033a_med2_name_CHILDQ | venlafaxine | CHEBI:9943 |
| eb_b034a_med3_name_CHILDQ | venlafaxine | CHEBI:9943 |
| eb_b035a_med4_name_CHILDQ | venlafaxine | CHEBI:9943 |
| eb_b038a_med7_name_CHILDQ | venlafaxine | CHEBI:9943 |
| eb_b032a_med1_name_CHILDQ | venlafaxine hcl | CHEBI:9944 |
| eb_b032a_med1_name_CHILDQ | ventolin | CHEBI:2550 |
| eb_b034a_med3_name_CHILDQ | ventolin | CHEBI:2550 |
| eb_b035a_med4_name_CHILDQ | ventolin | CHEBI:2550 |
| eb_b036a_med5_name_CHILDQ | ventolin | CHEBI:2550 |
| eb_b037a_med6_name_CHILDQ | ventolin | CHEBI:2550 |
| eb_b038a_med7_name_CHILDQ | ventolin | CHEBI:2550 |
| eb_b039a_med8_name_CHILDQ | ventolin | CHEBI:2550 |
| eb_b040a_med9_name_CHILDQ | ventolin | CHEBI:2550 |
| eb_b041a_med10_name_CHILDQ | ventolin | CHEBI:2550 |
| eb_b035a_med4_name_CHILDQ | veramyst | CHEBI:74899 |
| eb_b032a_med1_name_CHILDQ | verapamil | CHEBI:9948 |
| eb_b033a_med2_name_CHILDQ | verapamil | CHEBI:9948 |
| eb_b035a_med4_name_CHILDQ | verapamil | CHEBI:9948 |
| eb_b037a_med6_name_CHILDQ | verapamil | CHEBI:9948 |
| eb_b033a_med2_name_CHILDQ | vesicare | CHEBI:135530 |
| eb_b034a_med3_name_CHILDQ | vesicare | CHEBI:135530 |
| eb_b035a_med4_name_CHILDQ | vesicare | CHEBI:135530 |
| eb_b032a_med1_name_CHILDQ | viagra | CHEBI:58987 |
| eb_b038a_med7_name_CHILDQ | viagra | CHEBI:58987 |
| eb_b032a_med1_name_CHILDQ | victoza | CHEBI:71193 |
| eb_b033a_med2_name_CHILDQ | victoza | CHEBI:71193 |
| eb_b034a_med3_name_CHILDQ | victoza | CHEBI:71193 |
| eb_b037a_med6_name_CHILDQ | victoza | CHEBI:71193 |
| eb_b038a_med7_name_CHILDQ | victoza | CHEBI:71193 |
| eb_b040a_med9_name_CHILDQ | victoza | CHEBI:71193 |
| eb_b032a_med1_name_CHILDQ | viibryd | CHEBI:70705 |
| eb_b033a_med2_name_CHILDQ | viibryd | CHEBI:70705 |
| eb_b036a_med5_name_CHILDQ | vilanterol | CHEBI:75037 |
| eb_b032a_med1_name_CHILDQ | vimpat | CHEBI:135939 |
| eb_b034a_med3_name_CHILDQ | vistaril | CHEBI:31680 |
| eb_b032a_med1_name_CHILDQ | vitamin b | CHEBI:75769 |
| eb_b035a_med4_name_CHILDQ | vitamin b | CHEBI:75769 |
| eb_b036a_med5_name_CHILDQ | vitamin b | CHEBI:75769 |
| eb_b040a_med9_name_CHILDQ | vitamin b | CHEBI:75769 |
| eb_b041a_med10_name_CHILDQ | vitamin b | CHEBI:75769 |
| eb_b032a_med1_name_CHILDQ | vitamin b12 | CHEBI:176843 |
| eb_b035a_med4_name_CHILDQ | vitamin b12 | CHEBI:176843 |
| eb_b036a_med5_name_CHILDQ | vitamin b12 | CHEBI:176843 |
| eb_b041a_med10_name_CHILDQ | vitamin b12 | CHEBI:176843 |
| eb_b034a_med3_name_CHILDQ | vitamin c | CHEBI:176783 |
| eb_b035a_med4_name_CHILDQ | vitamin c | CHEBI:176783 |
| eb_b037a_med6_name_CHILDQ | vitamin c | CHEBI:176783 |
| eb_b038a_med7_name_CHILDQ | vitamin c | CHEBI:176783 |
| eb_b039a_med8_name_CHILDQ | vitamin c | CHEBI:176783 |
| eb_b040a_med9_name_CHILDQ | vitamin c | CHEBI:176783 |
| eb_b032a_med1_name_CHILDQ | vitamin d | CHEBI:27300 |
| eb_b033a_med2_name_CHILDQ | vitamin d | CHEBI:27300 |
| eb_b034a_med3_name_CHILDQ | vitamin d | CHEBI:27300 |
| eb_b035a_med4_name_CHILDQ | vitamin d | CHEBI:27300 |
| eb_b036a_med5_name_CHILDQ | vitamin d | CHEBI:27300 |
| eb_b037a_med6_name_CHILDQ | vitamin d | CHEBI:27300 |
| eb_b038a_med7_name_CHILDQ | vitamin d | CHEBI:27300 |
| eb_b039a_med8_name_CHILDQ | vitamin d | CHEBI:27300 |
| eb_b040a_med9_name_CHILDQ | vitamin d | CHEBI:27300 |
| eb_b041a_med10_name_CHILDQ | vitamin d | CHEBI:27300 |
| eb_b032a_med1_name_CHILDQ | vitamin d2 | CHEBI:28934 |
| eb_b035a_med4_name_CHILDQ | vitamin d2 | CHEBI:28934 |
| eb_b037a_med6_name_CHILDQ | vitamin d2 | CHEBI:28934 |
| eb_b039a_med8_name_CHILDQ | vitamin d2 | CHEBI:28934 |
| eb_b040a_med9_name_CHILDQ | vitamin d2 | CHEBI:28934 |
| eb_b032a_med1_name_CHILDQ | vitamin d3 | CHEBI:28940 |
| eb_b033a_med2_name_CHILDQ | vitamin d3 | CHEBI:28940 |
| eb_b034a_med3_name_CHILDQ | vitamin d3 | CHEBI:28940 |
| eb_b035a_med4_name_CHILDQ | vitamin d3 | CHEBI:28940 |
| eb_b036a_med5_name_CHILDQ | vitamin d3 | CHEBI:28940 |
| eb_b037a_med6_name_CHILDQ | vitamin d3 | CHEBI:28940 |
| eb_b038a_med7_name_CHILDQ | vitamin d3 | CHEBI:28940 |
| eb_b041a_med10_name_CHILDQ | vitamin d3 | CHEBI:28940 |
| eb_b032a_med1_name_CHILDQ | vitamin e | CHEBI:33234 |
| eb_b035a_med4_name_CHILDQ | vitamin e | CHEBI:33234 |
| eb_b041a_med10_name_CHILDQ | vitamin e | CHEBI:33234 |
| eb_b032a_med1_name_CHILDQ | vyvanse | CHEBI:135925 |
| eb_b033a_med2_name_CHILDQ | vyvanse | CHEBI:135925 |
| eb_b036a_med5_name_CHILDQ | vyvanse | CHEBI:135925 |
| eb_b041a_med10_name_CHILDQ | vyvanse | CHEBI:135925 |
| eb_b032a_med1_name_CHILDQ | warfarin | CHEBI:10033 |
| eb_b033a_med2_name_CHILDQ | warfarin | CHEBI:10033 |
| eb_b032a_med1_name_CHILDQ | wellbutrin | CHEBI:3220 |
| eb_b033a_med2_name_CHILDQ | wellbutrin | CHEBI:3220 |
| eb_b034a_med3_name_CHILDQ | wellbutrin | CHEBI:3220 |
| eb_b035a_med4_name_CHILDQ | wellbutrin | CHEBI:3220 |
| eb_b036a_med5_name_CHILDQ | wellbutrin | CHEBI:3220 |
| eb_b037a_med6_name_CHILDQ | wellbutrin | CHEBI:3220 |
| eb_b038a_med7_name_CHILDQ | wellbutrin | CHEBI:3220 |
| eb_b032a_med1_name_CHILDQ | xalatan | CHEBI:6384 |
| eb_b032a_med1_name_CHILDQ | xanax | CHEBI:2611 |
| eb_b033a_med2_name_CHILDQ | xanax | CHEBI:2611 |
| eb_b034a_med3_name_CHILDQ | xanax | CHEBI:2611 |
| eb_b037a_med6_name_CHILDQ | xanax | CHEBI:2611 |
| eb_b032a_med1_name_CHILDQ | xarelto | CHEBI:68579 |
| eb_b033a_med2_name_CHILDQ | xarelto | CHEBI:68579 |
| eb_b034a_med3_name_CHILDQ | xarelto | CHEBI:68579 |
| eb_b032a_med1_name_CHILDQ | xeljanz | CHEBI:71197 |
| eb_b037a_med6_name_CHILDQ | xeljanz | CHEBI:71197 |
| eb_b039a_med8_name_CHILDQ | xeljanz | CHEBI:71197 |
| eb_b034a_med3_name_CHILDQ | xeloda | CHEBI:31348 |
| eb_b033a_med2_name_CHILDQ | xiidra | CHEBI:133023 |
| eb_b040a_med9_name_CHILDQ | xyrem | CHEBI:30830 |
| eb_b033a_med2_name_CHILDQ | xyzal | CHEBI:94559 |
| eb_b036a_med5_name_CHILDQ | zafirlukast | CHEBI:10100 |
| eb_b034a_med3_name_CHILDQ | zaleplon | CHEBI:10102 |
| eb_b036a_med5_name_CHILDQ | zaleplon | CHEBI:10102 |
| eb_b032a_med1_name_CHILDQ | zantac | CHEBI:8777 |
| eb_b033a_med2_name_CHILDQ | zantac | CHEBI:8777 |
| eb_b034a_med3_name_CHILDQ | zantac | CHEBI:8777 |
| eb_b035a_med4_name_CHILDQ | zantac | CHEBI:8777 |
| eb_b036a_med5_name_CHILDQ | zantac | CHEBI:8777 |
| eb_b037a_med6_name_CHILDQ | zantac | CHEBI:8777 |
| eb_b038a_med7_name_CHILDQ | zantac | CHEBI:8777 |
| eb_b039a_med8_name_CHILDQ | zantac | CHEBI:8777 |
| eb_b036a_med5_name_CHILDQ | zeaxanthin | CHEBI:27547 |
| eb_b037a_med6_name_CHILDQ | zeaxanthin | CHEBI:27547 |
| eb_b032a_med1_name_CHILDQ | zetia | CHEBI:49040 |
| eb_b033a_med2_name_CHILDQ | zetia | CHEBI:49040 |
| eb_b036a_med5_name_CHILDQ | zetia | CHEBI:49040 |
| eb_b037a_med6_name_CHILDQ | zetia | CHEBI:49040 |
| eb_b038a_med7_name_CHILDQ | zetia | CHEBI:49040 |
| eb_b033a_med2_name_CHILDQ | zinc | CHEBI:27363 |
| eb_b032a_med1_name_CHILDQ | zocor | CHEBI:9150 |
| eb_b033a_med2_name_CHILDQ | zocor | CHEBI:9150 |
| eb_b034a_med3_name_CHILDQ | zocor | CHEBI:9150 |
| eb_b035a_med4_name_CHILDQ | zocor | CHEBI:9150 |
| eb_b036a_med5_name_CHILDQ | zocor | CHEBI:9150 |
| eb_b037a_med6_name_CHILDQ | zocor | CHEBI:9150 |
| eb_b033a_med2_name_CHILDQ | zolmitriptan | CHEBI:10124 |
| eb_b036a_med5_name_CHILDQ | zolmitriptan | CHEBI:10124 |
| eb_b032a_med1_name_CHILDQ | zoloft | CHEBI:9124 |
| eb_b033a_med2_name_CHILDQ | zoloft | CHEBI:9124 |
| eb_b035a_med4_name_CHILDQ | zoloft | CHEBI:9124 |
| eb_b038a_med7_name_CHILDQ | zoloft | CHEBI:9124 |
| eb_b032a_med1_name_CHILDQ | zolpidem | CHEBI:10125 |
| eb_b033a_med2_name_CHILDQ | zolpidem | CHEBI:10125 |
| eb_b034a_med3_name_CHILDQ | zolpidem | CHEBI:10125 |
| eb_b035a_med4_name_CHILDQ | zolpidem | CHEBI:10125 |
| eb_b036a_med5_name_CHILDQ | zolpidem | CHEBI:10125 |
| eb_b038a_med7_name_CHILDQ | zolpidem | CHEBI:10125 |
| eb_b032a_med1_name_CHILDQ | zolpidem tartrate | CHEBI:10126 |
| eb_b033a_med2_name_CHILDQ | zolpidem tartrate | CHEBI:10126 |
| eb_b034a_med3_name_CHILDQ | zolpidem tartrate | CHEBI:10126 |
| eb_b036a_med5_name_CHILDQ | zolpidem tartrate | CHEBI:10126 |
| eb_b038a_med7_name_CHILDQ | zolpidem tartrate | CHEBI:10126 |
| eb_b034a_med3_name_CHILDQ | zomig | CHEBI:10124 |
| eb_b036a_med5_name_CHILDQ | zonisamide | CHEBI:10127 |
| eb_b032a_med1_name_CHILDQ | zyprexa | CHEBI:7735 |

Supplemental Table 2A: Endometriosis Logistic Regression

| **Log.odds** | **Log.odds.min** | **Log.odds.max** | **Odds** | **Odds.min** | **Odds.max** | **Standard.Error** | **Survey.question.label** | **Ontology.ID** | **p.value** | **mean_prevalence** | **sd_prevalence** | **miss_rate** | **mean_vif** |
| --- | --- | --- | --- | --- | --- | --- | --- | --- | --- | --- | --- | --- | --- |
| 1 | 0.44 | 1.57 | 2.71 | 1.55 | 4.80 | 0.29 | he_e032_migraine | HP:0002076 | 5.12E-04 | 0.25 | 0.43 | 0.01 | 1.47 |
| 1.92 | 0.86 | 3.16 | 6.84 | 2.37 | 23.46 | 0.58 | he_m090_uterine_polyps | MONDO:0006195 | 8.44E-04 | 0.06 | 0.24 | 0.00 | 1.54 |
| -0.35 | -0.61 | -0.10 | 0.70 | 0.55 | 0.90 | 0.13 | eb_i212_carrots | ECTO:0070046 | 6.33E-03 | 2.83 | 1.02 | 0.02 | 1.79 |
| 1.07 | 0.28 | 1.87 | 2.91 | 1.33 | 6.47 | 0.40 | he_m084_ovaries_parq | MAXO:0001067 | 7.99E-03 | 0.15 | 0.36 | 0.00 | 2.75 |
| 0.79 | 0.19 | 1.39 | 2.19 | 1.21 | 4.00 | 0.30 | he_m092_ovarian_cysts | HP:0000138 | 9.75E-03 | 0.22 | 0.41 | 0.00 | 1.63 |
| 0.89 | 0.20 | 1.60 | 2.44 | 1.22 | 4.94 | 0.36 | he_m083_hysterectomy | MAXO:0001058 | 1.20E-02 | 0.20 | 0.40 | 0.00 | 2.32 |
| 3.1 | 0.90 | 5.87 | 22.18 | 2.47 | >50 | 1.24 | CHEBI_6905 | CHEBI:6905 | 1.23E-02 | 0.04 | 0.19 | 0.00 |  |
| -1.54 | -2.77 | -0.31 | 0.22 | 0.06 | 0.73 | 0.62 | he_j063_osteoporosis | HP:0000939 | 1.35E-02 | 0.04 | 0.19 | 0.01 | 1.67 |
| 0.52 | 0.08 | 0.98 | 1.69 | 1.08 | 2.66 | 0.23 | ea_a060_pest_regularly_parq | ECTO:0000530 | 2.21E-02 | 0.44 | 0.50 | 0.05 | 1.36 |
| -0.25 | -0.46 | -0.04 | 0.78 | 0.63 | 0.96 | 0.11 | eb_i268_chocolate_dark | ECTO:0070138 | 2.22E-02 | 2.39 | 1.12 | 0.02 | 1.53 |
| -2.37 | -4.91 | -0.50 | 0.09 | 0.01 | 0.61 | 1.09 | CHEBI_6904 | CHEBI:6904 | 3.04E-02 | 0.05 | 0.22 | 0.00 |  |
| 1.78 | 0.30 | 3.74 | 5.95 | 1.36 | 42.05 | 0.84 | CHEBI_9584 | CHEBI:9584 | 3.29E-02 | 0.01 | 0.12 | 0.00 | 1.36 |
| -1.46 | -2.92 | -0.19 | 0.23 | 0.05 | 0.83 | 0.69 | ea_b116_alcohol_derived | ECTO:9000026 | 3.36E-02 | 0.19 | 0.39 | 0.11 |  |
| 1.48 | 0.13 | 3.00 | 4.41 | 1.13 | 20.13 | 0.72 | ea_b116a_isopropanol_parq | ECTO:9000099 | 4.02E-02 | 0.14 | 0.35 | 0.00 |  |
| -0.29 | -0.57 | -0.01 | 0.75 | 0.57 | 0.99 | 0.14 | eb_i222_tofu | ECTO:0070185 | 4.20E-02 | 1.56 | 0.91 | 0.02 | 1.70 |
| 0.65 | 0.01 | 1.31 | 1.92 | 1.01 | 3.69 | 0.33 | he_c023_thyroid_disease_parq | MONDO:0003240 | 4.71E-02 | 0.17 | 0.38 | 0.01 | 1.57 |
| 0.9 | -0.03 | 1.91 | 2.46 | 0.97 | 6.76 | 0.49 | CHEBI_6446 | CHEBI_6446 | 6.54E-02 | 0.14 | 0.34 | 0.00 |  |
| 2.29 | 0.22 | 5.57 | 9.91 | 1.24 | >50 | 1.27 | CHEBI_77249 | CHEBI_77249 | 7.06E-02 | 0.02 | 0.13 | 0.00 | 1.43 |
| -0.98 | -2.15 | 0.12 | 0.37 | 0.12 | 1.13 | 0.57 | eb_a021_folic_acid_parq | ECTO:9000123 | 8.69E-02 | 0.07 | 0.25 | 0.01 | 1.93 |
| 0.54 | -0.09 | 1.16 | 1.71 | 0.92 | 3.20 | 0.32 | he_m085_menopause_parq | GO:0042697 | 9.18E-02 | 0.50 | 0.50 | 0.01 | 2.09 |
| 0.77 | -0.12 | 1.71 | 2.16 | 0.89 | 5.52 | 0.46 | CHEBI_87631 | CHEBI_87631 | 9.44E-02 | 0.11 | 0.31 | 0.00 | 2.48 |
| 1.45 | -0.15 | 3.48 | 4.28 | 0.86 | 32.42 | 0.89 | CHEBI_50275 | CHEBI_50275 | 1.01E-01 | 0.02 | 0.16 | 0.00 | 1.66 |
| 0.18 | -0.04 | 0.39 | 1.19 | 0.96 | 1.48 | 0.11 | eb_i203_tomatoes | ECTO:0070036 | 1.05E-01 | 2.97 | 1.12 | 0.02 | 1.70 |
| 1.05 | -0.21 | 2.43 | 2.85 | 0.81 | 11.34 | 0.66 | ea_b092a_benzene_parq | ECTO:9000034 | 1.14E-01 | 0.04 | 0.20 | 0.00 | 1.89 |
| -0.71 | -1.63 | 0.15 | 0.49 | 0.20 | 1.16 | 0.45 | CHEBI_5855 | CHEBI_5855 | 1.15E-01 | 0.06 | 0.24 | 0.00 | 1.52 |
| 0.8 | -0.19 | 1.84 | 2.22 | 0.83 | 6.32 | 0.51 | eb_a004_vitamin_b6_parq | ECTO:9000243 | 1.21E-01 | 0.09 | 0.28 | 0.02 | 2.37 |
| 0.95 | -0.23 | 2.22 | 2.58 | 0.80 | 9.24 | 0.61 | CHEBI_46195 | CHEBI_46195 | 1.22E-01 | 0.03 | 0.17 | 0.00 | 1.54 |
| 1.17 | -0.25 | 2.82 | 3.21 | 0.78 | 16.75 | 0.76 | ea_b194a_hair_dye_parq | ECTO:7000151 | 1.26E-01 | 0.06 | 0.23 | 0.00 | 1.64 |
| 0.2 | -0.06 | 0.46 | 1.22 | 0.94 | 1.58 | 0.13 | eb_i206_watermelon | ECTO:0070039 | 1.35E-01 | 2.11 | 0.89 | 0.02 | 1.85 |
| -0.51 | -1.19 | 0.16 | 0.60 | 0.30 | 1.17 | 0.34 | eb_a007_vitamin_c_parq | ECTO:9000143 | 1.38E-01 | 0.20 | 0.40 | 0.01 | 1.74 |
| 0.44 | -0.16 | 1.04 | 1.56 | 0.86 | 2.83 | 0.30 | he_m091_uterine_tumors | HP:0000131 | 1.47E-01 | 0.24 | 0.43 | 0.00 | 1.66 |
| 0.3 | -0.09 | 0.76 | 1.35 | 0.91 | 2.15 | 0.21 | eb_i286_saccharin | ECTO:0070061 | 1.52E-01 | 1.11 | 0.52 | 0.03 | 1.43 |
| 0.79 | -0.27 | 1.93 | 2.21 | 0.76 | 6.89 | 0.56 | he_g049_kidney_stones | HP:0000787 | 1.54E-01 | 0.07 | 0.26 | 0.01 | 1.61 |
| 0.18 | -0.07 | 0.42 | 1.19 | 0.94 | 1.52 | 0.12 | eb_i243_other_crackers | ECTO:0070109 | 1.54E-01 | 2.27 | 0.98 | 0.03 | 1.68 |
| 0.75 | -0.34 | 1.87 | 2.12 | 0.71 | 6.51 | 0.56 | he_h055_fibromyalgia | MONDO:0005546 | 1.79E-01 | 0.05 | 0.22 | 0.01 | 1.70 |
| 0.18 | -0.09 | 0.46 | 1.20 | 0.91 | 1.59 | 0.14 | eb_i221_green_beans | ECTO:0070062 | 1.88E-01 | 2.71 | 0.91 | 0.04 | 1.67 |
| 0.36 | -0.18 | 0.91 | 1.44 | 0.83 | 2.49 | 0.28 | eb_a019_fish_oil_parq | ECTO:0070164 | 1.92E-01 | 0.23 | 0.42 | 0.01 | 1.73 |
| -0.56 | -1.42 | 0.29 | 0.57 | 0.24 | 1.34 | 0.43 | he_f038_lactose_intolerance | HP:0004789 | 2.00E-01 | 0.09 | 0.28 | 0.01 | 1.45 |
| 0.96 | -0.46 | 2.56 | 2.60 | 0.63 | 12.88 | 0.75 | CHEBI_9150 | CHEBI_9150 | 2.03E-01 | 0.04 | 0.19 | 0.00 | 1.83 |
| 0.51 | -0.30 | 1.34 | 1.67 | 0.74 | 3.83 | 0.42 | eb_a006_vitamin_b_comp_parq | ECTO:9002169 | 2.18E-01 | 0.12 | 0.33 | 0.01 | 2.13 |
| 1.36 | -0.67 | 4.41 | 3.90 | 0.51 | 82.06 | 1.18 | ea_b140a_nitrous_oxide_parq | ECTO:9000051 | 2.48E-01 | 0.02 | 0.14 | 0.00 | 1.47 |
| 0.89 | -0.61 | 2.57 | 2.43 | 0.54 | 13.08 | 0.78 | ea_b104_cleaning_derived | ECTO:0500011 | 2.57E-01 | 0.29 | 0.45 | 0.05 |  |
| 0.65 | -0.48 | 1.84 | 1.91 | 0.62 | 6.28 | 0.58 | ea_b134_dust_derived | ECTO:7000001 | 2.65E-01 | 0.08 | 0.27 | 0.10 | 1.86 |
| 0.84 | -0.59 | 2.49 | 2.32 | 0.55 | 12.10 | 0.76 | CHEBI_50730 | CHEBI_50730 | 2.71E-01 | 0.02 | 0.13 | 0.00 | 1.56 |
| 0.47 | -0.37 | 1.35 | 1.60 | 0.69 | 3.85 | 0.44 | eb_a018_coq10_parq | ECTO:0070163 | 2.81E-01 | 0.08 | 0.26 | 0.01 | 1.58 |
| -0.44 | -1.26 | 0.37 | 0.65 | 0.28 | 1.45 | 0.41 | he_f042_gallbladder_disease | MONDO:0005281 | 2.92E-01 | 0.10 | 0.31 | 0.01 | 1.55 |
| 0.52 | -0.44 | 1.50 | 1.67 | 0.64 | 4.47 | 0.49 | ea_b194g_gel_ink_parq | ECTO:7000153 | 2.95E-01 | 0.08 | 0.27 | 0.00 | 1.60 |
| -1.1 | -3.23 | 1.04 | 0.33 | 0.04 | 2.83 | 1.07 | ea_b128e_phenol_parq | ECTO:9000071 | 3.05E-01 | 0.03 | 0.17 | 0.00 | 1.88 |
| 1 | -0.83 | 3.17 | 2.71 | 0.44 | 23.81 | 0.98 | CHEBI_8776 | CHEBI_8776 | 3.08E-01 | 0.02 | 0.14 | 0.00 | 1.46 |
| 0.36 | -0.34 | 1.07 | 1.43 | 0.71 | 2.92 | 0.36 | he_j062_bone_loss | HP:0000938 | 3.16E-01 | 0.20 | 0.40 | 0.01 | 1.94 |
| 0.28 | -0.27 | 0.84 | 1.33 | 0.76 | 2.32 | 0.28 | he_b008_high_cholesterol | HP_0003124 | 3.18E-01 | 0.31 | 0.46 | 0.00 | 1.73 |
| -0.93 | -2.90 | 0.97 | 0.39 | 0.05 | 2.63 | 0.96 | CHEBI_30660 | CHEBI_30660 | 3.33E-01 | 0.07 | 0.25 | 0.00 |  |
| -0.38 | -1.19 | 0.41 | 0.68 | 0.30 | 1.50 | 0.41 | he_c021_pre_diabetes_parq | MONDO:0006920 | 3.44E-01 | 0.12 | 0.32 | 0.01 | 1.91 |
| -0.52 | -1.61 | 0.57 | 0.60 | 0.20 | 1.77 | 0.55 | he_f043_stomach_ulcer | HP:0002592 | 3.47E-01 | 0.05 | 0.22 | 0.01 | 1.71 |
| 0.96 | -1.01 | 3.19 | 2.62 | 0.36 | 24.18 | 1.03 | ea_b194c_textile_dye_parq | ECTO:0500008 | 3.48E-01 | 0.01 | 0.11 | 0.00 | 1.57 |
| 0.52 | -0.56 | 1.63 | 1.68 | 0.57 | 5.11 | 0.55 | ea_b146d_xrays_parq | ECTO:8000046 | 3.48E-01 | 0.08 | 0.27 | 0.00 | 1.92 |
| 0.09 | -0.10 | 0.28 | 1.09 | 0.90 | 1.32 | 0.10 | eb_i184_yogurt_lowfat | ECTO:0070015 | 3.62E-01 | 2.02 | 1.23 | 0.03 | 1.53 |
| -1.03 | -3.44 | 1.22 | 0.36 | 0.03 | 3.39 | 1.17 | CHEBI_5163 | CHEBI_5163 | 3.77E-01 | 0.04 | 0.20 | 0.00 | 1.50 |
| 1.26 | -1.50 | 4.64 | 3.51 | 0.22 | >50 | 1.45 | ea_b152_acid_derived | ECTO:9002146 | 3.87E-01 | 0.08 | 0.27 | 0.10 |  |
| 0.26 | -0.37 | 0.90 | 1.30 | 0.69 | 2.46 | 0.32 | eb_a013_magnesium_parq | ECTO:9000210 | 4.15E-01 | 0.16 | 0.37 | 0.01 | 1.74 |
| -0.53 | -1.90 | 0.75 | 0.59 | 0.15 | 2.12 | 0.67 | CHEBI_3219 | CHEBI_3219 | 4.29E-01 | 0.05 | 0.21 | 0.00 | 1.56 |
| 1.2 | -1.81 | 4.43 | 3.32 | 0.16 | 84.35 | 1.55 | ea_b164d_polyurethane_parq | ECTO:9000082 | 4.38E-01 | 0.03 | 0.17 | 0.00 |  |
| 0.19 | -0.30 | 0.67 | 1.21 | 0.74 | 1.96 | 0.25 | eb_a001_multivitamin_parq | ECTO:0070210 | 4.47E-01 | 0.48 | 0.50 | 0.00 | 1.60 |
| -0.68 | -2.50 | 1.10 | 0.51 | 0.08 | 3.01 | 0.90 | ea_b158_alkalis_derived | ECTO:9000021 | 4.49E-01 | 0.06 | 0.23 | 0.14 | 2.42 |
| 0.52 | -0.86 | 1.96 | 1.68 | 0.42 | 7.09 | 0.70 | CHEBI_9124 | CHEBI_9124 | 4.62E-01 | 0.04 | 0.20 | 0.00 | 2.12 |
| 0.24 | -0.41 | 0.88 | 1.27 | 0.67 | 2.42 | 0.33 | eb_a005_vitamin_b12_parq | ECTO:9000229 | 4.68E-01 | 0.21 | 0.40 | 0.01 | 1.92 |
| -0.08 | -0.28 | 0.13 | 0.93 | 0.75 | 1.14 | 0.11 | eb_i183_yogurt_nonfat | ECTO:0070014 | 4.70E-01 | 1.90 | 1.21 | 0.03 | 1.54 |
| 0.67 | -1.18 | 2.78 | 1.95 | 0.31 | 16.10 | 0.96 | CHEBI_3723 | CHEBI_3723 | 4.87E-01 | 0.02 | 0.14 | 0.00 | 1.52 |
| 0.09 | -0.16 | 0.34 | 1.09 | 0.85 | 1.41 | 0.13 | eb_i196_grapes | ECTO:0070027 | 4.87E-01 | 2.50 | 0.95 | 0.02 | 1.80 |
| 0.08 | -0.14 | 0.29 | 1.08 | 0.87 | 1.34 | 0.11 | eb_i242_wheat_crackers | ECTO:0070108 | 4.91E-01 | 2.35 | 1.09 | 0.02 | 1.78 |
| 0.1 | -0.18 | 0.38 | 1.10 | 0.84 | 1.46 | 0.14 | eb_i223_winter_squash | ECTO:0070067 | 4.92E-01 | 1.69 | 0.84 | 0.02 | 1.83 |
| -0.23 | -0.91 | 0.44 | 0.79 | 0.40 | 1.55 | 0.34 | he_j067_osteoarthritis_parq | HP:0002758 | 5.01E-01 | 0.19 | 0.39 | 0.02 | 1.67 |
| 0.2 | -0.40 | 0.81 | 1.23 | 0.67 | 2.24 | 0.31 | he_b007_hypertension_parq | HP:0000822 | 5.06E-01 | 0.29 | 0.45 | 0.00 | 1.99 |
| -0.71 | -2.87 | 1.54 | 0.49 | 0.06 | 4.67 | 1.07 | he_i060_pernicious_anemia | MONDO:0008228 | 5.11E-01 | 0.01 | 0.09 | 0.00 | 1.50 |
| 0.39 | -0.77 | 1.59 | 1.47 | 0.46 | 4.92 | 0.59 | CHEBI_42797 | CHEBI_42797 | 5.13E-01 | 0.03 | 0.18 | 0.00 | 1.42 |
| -0.89 | -4.18 | 1.77 | 0.41 | 0.02 | 5.89 | 1.39 | ea_b152a_hydrochloric_acid_parq | ECTO:9000016 | 5.23E-01 | 0.06 | 0.24 | 0.00 |  |
| 0.36 | -0.80 | 1.62 | 1.44 | 0.45 | 5.07 | 0.61 | he_g050_kidney_infection | HP:0012330 | 5.54E-01 | 0.05 | 0.22 | 0.01 | 1.54 |
| -0.55 | -2.59 | 1.30 | 0.57 | 0.08 | 3.67 | 0.96 | ea_b182_adhesives_derived | ECTO:7000130 | 5.64E-01 | 0.09 | 0.29 | 0.10 |  |
| 0.07 | -0.17 | 0.31 | 1.07 | 0.84 | 1.37 | 0.12 | eb_i186_cottage_cheese | ECTO:0070017 | 5.73E-01 | 1.69 | 0.94 | 0.02 | 1.52 |
| -0.08 | -0.36 | 0.20 | 0.92 | 0.70 | 1.22 | 0.14 | eb_i200_pear | ECTO:0070033 | 5.79E-01 | 1.93 | 0.84 | 0.02 | 1.62 |
| -0.37 | -1.70 | 0.95 | 0.69 | 0.18 | 2.60 | 0.67 | ea_b092b_chloroform_parq | ECTO:9000042 | 5.81E-01 | 0.06 | 0.23 | 0.00 | 2.06 |
| 0.43 | -1.12 | 2.14 | 1.54 | 0.33 | 8.53 | 0.81 | CHEBI_31441 | CHEBI_31441 | 5.95E-01 | 0.02 | 0.16 | 0.00 | 1.58 |
| 0.25 | -0.67 | 1.20 | 1.29 | 0.51 | 3.32 | 0.47 | ea_b104b_ammonia_parq | ECTO:9000031 | 5.95E-01 | 0.08 | 0.28 | 0.00 | 1.78 |
| -0.18 | -0.85 | 0.49 | 0.83 | 0.43 | 1.63 | 0.34 | he_f041_polyps | HP:0200063 | 5.98E-01 | 0.17 | 0.38 | 0.01 | 1.70 |
| -0.38 | -1.86 | 1.06 | 0.68 | 0.16 | 2.88 | 0.74 | he_b016_raynauds | MONDO:0008364 | 6.06E-01 | 0.05 | 0.21 | 0.01 | 1.47 |
| 0.3 | -0.82 | 1.47 | 1.34 | 0.44 | 4.33 | 0.57 | CHEBI_6801 | CHEBI_6801 | 6.07E-01 | 0.05 | 0.22 | 0.00 | 1.97 |
| 0.41 | -1.18 | 2.10 | 1.50 | 0.31 | 8.14 | 0.81 | CHEBI_27300 | CHEBI_27300 | 6.18E-01 | 0.03 | 0.16 | 0.00 | 1.48 |
| 0.24 | -0.77 | 1.26 | 1.27 | 0.46 | 3.52 | 0.51 | CHEBI_6538 | CHEBI_6538 | 6.41E-01 | 0.03 | 0.16 | 0.00 | 1.44 |
| -0.36 | -2.05 | 1.15 | 0.70 | 0.13 | 3.16 | 0.79 | ea_b104a_bleach_parq | ECTO:9000076 | 6.51E-01 | 0.27 | 0.45 | 0.00 |  |
| 0.13 | -0.48 | 0.74 | 1.14 | 0.62 | 2.10 | 0.31 | eb_a028_probiotic_parq | ECTO:0070000 | 6.77E-01 | 0.19 | 0.39 | 0.01 | 1.71 |
| -0.06 | -0.37 | 0.24 | 0.94 | 0.69 | 1.28 | 0.16 | eb_i254_fries | ECTO:0070129 | 6.81E-01 | 2.16 | 0.83 | 0.03 | 1.63 |
| -0.25 | -1.49 | 0.96 | 0.78 | 0.22 | 2.61 | 0.62 | he_k069_psoriasis | MONDO:0005083 | 6.82E-01 | 0.04 | 0.20 | 0.01 | 1.52 |
| -0.24 | -1.43 | 0.95 | 0.79 | 0.24 | 2.58 | 0.60 | CHEBI_3562 | CHEBI_3562 | 6.93E-01 | 0.04 | 0.21 | 0.00 | 1.38 |
| 0.54 | -2.21 | 3.95 | 1.72 | 0.11 | 51.96 | 1.48 | he_b012_heart_attack | HP:0001658 | 7.14E-01 | 0.01 | 0.10 | 0.01 | 1.49 |
| -0.34 | -2.23 | 1.55 | 0.71 | 0.11 | 4.73 | 0.94 | CHEBI_18332 | CHEBI_18332 | 7.15E-01 | 0.08 | 0.28 | 0.00 |  |
| 0.09 | -0.41 | 0.58 | 1.09 | 0.66 | 1.78 | 0.25 | eb_a008_vitamin_d_parq | ECTO:9000133 | 7.36E-01 | 0.51 | 0.50 | 0.01 | 1.85 |
| 0.04 | -0.21 | 0.29 | 1.04 | 0.81 | 1.33 | 0.13 | eb_i207_beans_lentils | ECTO:0070183 | 7.67E-01 | 2.94 | 0.97 | 0.02 | 1.65 |
| -0.17 | -1.37 | 1.04 | 0.84 | 0.25 | 2.84 | 0.61 | he_j066_rheu_arthritis_parq | HP:0001370 | 7.74E-01 | 0.04 | 0.20 | 0.04 | 1.79 |
| 0.11 | -0.65 | 0.87 | 1.12 | 0.52 | 2.38 | 0.39 | he_d030_asthma_parq | MONDO:0004979 | 7.76E-01 | 0.13 | 0.34 | 0.01 | 1.57 |
| 0.22 | -1.56 | 1.98 | 1.25 | 0.21 | 7.27 | 0.86 | CHEBI_9927 | CHEBI_9927 | 7.99E-01 | 0.01 | 0.12 | 0.00 | 1.39 |
| -0.2 | -1.79 | 1.39 | 0.82 | 0.17 | 4.02 | 0.80 | ea_b134e_talc_parq | ECTO:7000028 | 8.06E-01 | 0.03 | 0.16 | 0.00 | 1.83 |
| -0.19 | -1.73 | 1.40 | 0.83 | 0.18 | 4.05 | 0.79 | he_f045_fatty_liver | MONDO:0004790 | 8.13E-01 | 0.02 | 0.14 | 0.00 | 1.42 |
| 0.17 | -1.43 | 1.88 | 1.18 | 0.24 | 6.55 | 0.82 | CHEBI_43755 | CHEBI_43755 | 8.38E-01 | 0.06 | 0.24 | 0.00 | 1.70 |
| -0.26 | -3.04 | 2.38 | 0.77 | 0.05 | 10.86 | 1.32 | ea_b164_stains_derived | ECTO:0500015 | 8.46E-01 | 0.05 | 0.22 | 0.08 |  |
| -0.06 | -0.85 | 0.71 | 0.94 | 0.43 | 2.04 | 0.40 | CHEBI_15365 | CHEBI_15365 | 8.77E-01 | 0.12 | 0.32 | 0.00 | 1.71 |
| 0.02 | -0.24 | 0.29 | 1.02 | 0.78 | 1.33 | 0.14 | eb_i218_peas_lima_beans | ECTO:0070183 | 8.77E-01 | 2.33 | 0.94 | 0.03 | 1.83 |
| 0.08 | -0.98 | 1.14 | 1.08 | 0.38 | 3.11 | 0.53 | he_b010_cardiac_arrhythmia | HP:0011675 | 8.88E-01 | 0.05 | 0.22 | 0.00 | 1.42 |
| 0.01 | -0.20 | 0.23 | 1.01 | 0.82 | 1.26 | 0.11 | eb_i273_peanut_butter | ECTO:0070151 | 8.93E-01 | 2.76 | 1.15 | 0.02 | 1.55 |
| 0.08 | -1.23 | 1.43 | 1.09 | 0.29 | 4.19 | 0.67 | ea_b146c_radiation_parq | ECTO:0000001 | 9.02E-01 | 0.06 | 0.24 | 0.00 | 2.09 |
| 0.01 | -0.25 | 0.28 | 1.01 | 0.78 | 1.32 | 0.13 | eb_i178_frozen_yogurt | ECTO:0070009 | 9.14E-01 | 1.74 | 0.88 | 0.02 | 1.56 |
| -0.04 | -0.98 | 0.89 | 0.96 | 0.38 | 2.43 | 0.47 | CHEBI_7772 | CHEBI_7772 | 9.34E-01 | 0.05 | 0.22 | 0.00 | 1.93 |
| -0.07 | -2.37 | 2.37 | 0.93 | 0.09 | 10.68 | 1.17 | CHEBI_9123 | CHEBI_9123 | 9.49E-01 | 0.02 | 0.15 | 0.00 | 2.05 |
| 0.09 | -2.98 | 3.83 | 1.10 | 0.05 | 46.20 | 1.67 | he_b013_coronary_artery | MONDO:0005010 | 9.56E-01 | 0.01 | 0.09 | 0.01 | 1.58 |
| 0.05 | -2.04 | 2.28 | 1.05 | 0.13 | 9.81 | 1.08 | ea_b182a_white_glue_parq | ECTO:9000079 | 9.61E-01 | 0.06 | 0.24 | 0.00 |  |
| -0.02 | -1.10 | 1.05 | 0.98 | 0.33 | 2.85 | 0.54 | ea_b194_ink_derived | ECTO:7000157 | 9.74E-01 | 0.32 | 0.47 | 0.08 |  |
| 16.6 | -44.17 |  | >50 | 0.00 | >50 | >50 | CHEBI_50686 | CHEBI_50686 | 9.82E-01 | 0.02 | 0.13 | 0.00 | 1.58 |
| -15.21 | -337.19 | 2.63 | 0.00 | 0.00 | 13.81 | >50 | ea_b110f_mercury_parq | ECTO_0001571 | 9.85E-01 | 0.03 | 0.16 | 0.00 | 2.03 |
| 15.73 | -11.84 | 326.37 | >50 | 0.00 | >50 | >50 | ea_b110_metal_derived | ECTO:9002163 | 9.85E-01 | 0.05 | 0.22 | 0.14 | 2.23 |
| 0.01 | -1.01 | 1.02 | 1.01 | 0.36 | 2.78 | 0.51 | CHEBI_5778 | CHEBI_5778 | 9.85E-01 | 0.08 | 0.28 | 0.00 | 1.72 |
| 16.58 | -115.55 |  | >50 | 0.00 | >50 | >50 | ea_b134b_fiberglass_parq | ECTO:7000017 | 9.89E-01 | 0.01 | 0.08 | 0.00 | 1.36 |
| 0.01 | -1.06 | 1.10 | 1.01 | 0.35 | 3.01 | 0.55 | ea_b194f_inkjet_parq | ECTO:7000155 | 9.91E-01 | 0.23 | 0.42 | 0.00 |  |
| 17.37 | -246.91 |  | >50 | 0.00 | >50 | >50 | ea_b122g_rodenticides_parq | ECTO:9000090 | 9.92E-01 | 0.01 | 0.07 | 0.00 | 1.83 |
| 0 | -0.63 | 0.62 | 1.00 | 0.53 | 1.87 | 0.32 | he_m088_hormone_replacement | ECTO:2000005 | 9.98E-01 | 0.27 | 0.45 | 0.01 | 1.92 |

Supplemental Table 2B: Uterine Fibroid Logistic Regression

| **Log.odds** | **Log.odds.min** | **Log.odds.max** | **Odds** | **Odds.min** | **Odds.max** | **Standard.Error** | **Survey.question.label** | **Ontology.ID** | **p.value** | **mean_prevalence** | **sd_prevalence** | **miss_rate** | **mean_vif** |
| --- | --- | --- | --- | --- | --- | --- | --- | --- | --- | --- | --- | --- | --- |
| 1.43 | 0.95 | 1.94 | 4.20 | 2.58 | 6.94 | 0.25 | he_m083_hysterectomy | MAXO:0001058 | 1.20E-08 | 0.20 | 0.40 | 0.00 | 2.04 |
| 1.61 | 0.90 | 2.39 | 5.01 | 2.47 | 10.94 | 0.38 | he_m090_uterine_polyps | MONDO:0006195 | 1.90E-05 | 0.06 | 0.24 | 0.00 | 1.27 |
| 1.31 | 0.57 | 2.12 | 3.71 | 1.77 | 8.35 | 0.39 | CHEBI_7772 | CHEBI:7772 | 8.29E-04 | 0.05 | 0.22 | 0.00 | 1.62 |
| 0.53 | 0.14 | 0.91 | 1.69 | 1.16 | 2.49 | 0.20 | he_i059_iron_anemia | HP:0001891 | 7.04E-03 | 0.24 | 0.43 | 0.00 | 1.26 |
| 0.54 | 0.15 | 0.93 | 1.71 | 1.16 | 2.54 | 0.20 | he_b007_hypertension_parq | HP:0000822 | 7.27E-03 | 0.29 | 0.45 | 0.00 | 1.92 |
| 0.53 | 0.12 | 0.94 | 1.70 | 1.13 | 2.56 | 0.21 | he_m085_menopause_parq | GO:0042697 | 1.04E-02 | 0.50 | 0.50 | 0.01 | 2.10 |
| 0.52 | 0.12 | 0.93 | 1.69 | 1.12 | 2.54 | 0.21 | he_m092_ovarian_cysts | HP:0000138 | 1.19E-02 | 0.22 | 0.41 | 0.00 | 1.41 |
| 0.20 | 0.04 | 0.36 | 1.22 | 1.04 | 1.43 | 0.08 | eb_i198_orange | ECTO:0070029 | 1.37E-02 | 2.53 | 1.05 | 0.02 | 1.63 |
| 0.52 | 0.07 | 0.98 | 1.69 | 1.07 | 2.68 | 0.23 | eb_a013_magnesium_parq | ECTO:9000210 | 2.46E-02 | 0.16 | 0.37 | 0.01 | 1.43 |
| 0.95 | 0.10 | 1.85 | 2.58 | 1.11 | 6.34 | 0.44 | he_g050_kidney_infection | HP:0012330 | 3.25E-02 | 0.05 | 0.22 | 0.01 | 1.30 |
| 0.35 | 0.02 | 0.68 | 1.42 | 1.02 | 1.96 | 0.17 | eb_a008_vitamin_d_parq | ECTO:9000133 | 3.53E-02 | 0.51 | 0.50 | 0.01 | 1.55 |
| 1.38 | 0.11 | 2.93 | 3.97 | 1.12 | 18.82 | 0.70 | CHEBI_36796 | CHEBI:36796 | 4.82E-02 | 0.01 | 0.12 | 0.00 | 1.40 |
| -0.57 | -1.13 | 0.00 | 0.57 | 0.32 | 1.00 | 0.29 | he_f042_gallbladder_disease | MONDO:0005281 | 4.91E-02 | 0.10 | 0.31 | 0.01 | 1.42 |
| 2.09 | 0.30 | 5.06 | 8.12 | 1.35 | 156.98 | 1.10 | ea_b134b_fiberglass_parq | ECTO:7000017 | 5.65E-02 | 0.01 | 0.08 | 0.00 | 1.20 |
| 0.55 | -0.02 | 1.14 | 1.73 | 0.98 | 3.11 | 0.29 | CHEBI_5778 | CHEBI:5778 | 6.08E-02 | 0.08 | 0.28 | 0.00 | 1.53 |
| -0.17 | -0.36 | 0.01 | 0.84 | 0.70 | 1.01 | 0.09 | eb_i205_tomato_sauce | ECTO:0070038 | 6.72E-02 | 2.48 | 0.91 | 0.02 | 1.52 |
| 0.13 | -0.01 | 0.28 | 1.14 | 0.99 | 1.32 | 0.07 | eb_i277_walnuts | ECTO:0070155 | 7.51E-02 | 2.18 | 1.07 | 0.03 | 1.56 |
| 0.89 | -0.08 | 1.90 | 2.43 | 0.93 | 6.71 | 0.50 | ea_b116b_methanol_parq | ECTO:9000028 | 7.73E-02 | 0.07 | 0.25 | 0.00 | 3.05 |
| -0.14 | -0.30 | 0.02 | 0.87 | 0.74 | 1.02 | 0.08 | eb_i192_avocado | ECTO:0070023 | 7.76E-02 | 2.23 | 1.04 | 0.02 | 1.45 |
| 1.07 | -0.08 | 2.40 | 2.90 | 0.92 | 11.07 | 0.62 | CHEBI_9927 | CHEBI:9927 | 8.48E-02 | 0.01 | 0.12 | 0.00 | 1.24 |
| 0.54 | -0.07 | 1.18 | 1.72 | 0.93 | 3.25 | 0.32 | eb_a018_coq10_parq | ECTO:0070163 | 8.88E-02 | 0.08 | 0.26 | 0.01 | 1.45 |
| 1.13 | -0.13 | 2.66 | 3.09 | 0.88 | 14.34 | 0.69 | CHEBI_2904 | CHEBI:2904 | 1.02E-01 | 0.01 | 0.11 | 0.00 | 1.20 |
| 0.31 | -0.07 | 0.69 | 1.37 | 0.94 | 2.00 | 0.19 | eb_a019_fish_oil_parq | ECTO:0070164 | 1.07E-01 | 0.23 | 0.42 | 0.01 | 1.53 |
| 0.14 | -0.03 | 0.32 | 1.16 | 0.97 | 1.38 | 0.09 | eb_i229_bacon | ECTO:0070185 | 1.07E-01 | 2.28 | 0.93 | 0.02 | 1.47 |
| 0.94 | -0.17 | 2.18 | 2.56 | 0.84 | 8.86 | 0.59 | ea_b140c_carbon_monoxide_parq | ECTO:0000207 | 1.11E-01 | 0.02 | 0.15 | 0.00 | 1.40 |
| -0.40 | -0.91 | 0.11 | 0.67 | 0.40 | 1.12 | 0.26 | he_m087_birth_control | ECTO:9001831 | 1.26E-01 | 0.88 | 0.33 | 0.01 | 1.25 |
| 0.60 | -0.16 | 1.40 | 1.82 | 0.85 | 4.04 | 0.39 | he_k069_psoriasis | MONDO:0005083 | 1.27E-01 | 0.04 | 0.20 | 0.01 | 1.22 |
| 0.57 | -0.18 | 1.35 | 1.76 | 0.84 | 3.87 | 0.39 | CHEBI_6801 | CHEBI:6801 | 1.45E-01 | 0.05 | 0.22 | 0.00 | 1.96 |
| 0.66 | -0.24 | 1.60 | 1.94 | 0.79 | 4.94 | 0.47 | ea_b092b_chloroform_parq | ECTO:9000042 | 1.56E-01 | 0.06 | 0.23 | 0.00 | 2.34 |
| -0.13 | -0.32 | 0.05 | 0.87 | 0.72 | 1.05 | 0.10 | eb_i251_white_rice | ECTO:0070124 | 1.61E-01 | 2.28 | 0.92 | 0.02 | 1.48 |
| -0.58 | -1.43 | 0.22 | 0.56 | 0.24 | 1.25 | 0.42 | ea_b116c_ethanol_parq | ECTO:9000027 | 1.62E-01 | 0.10 | 0.31 | 0.00 | 2.84 |
| 1.01 | -0.35 | 2.61 | 2.75 | 0.71 | 13.63 | 0.73 | CHEBI_2453 | CHEBI:2453 | 1.67E-01 | 0.01 | 0.11 | 0.00 | 1.24 |
| 1.57 | -0.40 | 4.59 | 4.82 | 0.67 | 98.72 | 1.16 | ea_b194e_india_ink_parq | ECTO:0500009 | 1.74E-01 | 0.01 | 0.10 | 0.00 | 1.46 |
| 0.79 | -0.30 | 2.02 | 2.19 | 0.74 | 7.52 | 0.58 | CHEBI_5134 | CHEBI:5134 | 1.76E-01 | 0.02 | 0.15 | 0.00 | 1.29 |
| -0.64 | -1.61 | 0.28 | 0.53 | 0.20 | 1.32 | 0.48 | ea_b092j_xylenes_parq | ECTO:9000035 | 1.79E-01 | 0.05 | 0.23 | 0.00 | 2.74 |
| -0.42 | -1.04 | 0.20 | 0.66 | 0.35 | 1.22 | 0.32 | he_g049_kidney_stones | HP:0000787 | 1.81E-01 | 0.07 | 0.26 | 0.01 | 1.36 |
| 0.53 | -0.26 | 1.35 | 1.70 | 0.77 | 3.87 | 0.41 | ea_b170f_acetone_parq | ECTO:9000085 | 1.92E-01 | 0.05 | 0.21 | 0.00 | 1.55 |
| 0.63 | -0.32 | 1.67 | 1.88 | 0.73 | 5.29 | 0.50 | CHEBI_2668 | CHEBI:2668 | 2.06E-01 | 0.03 | 0.18 | 0.00 | 1.35 |
| -1.34 | -3.64 | 0.64 | 0.26 | 0.03 | 1.90 | 1.06 | ea_b158_alkalis_derived | ECTO:9000021 | 2.09E-01 | 0.06 | 0.23 | 0.14 |  |
| 0.75 | -0.40 | 1.97 | 2.12 | 0.67 | 7.14 | 0.60 | CHEBI_9150 | CHEBI:9150 | 2.10E-01 | 0.04 | 0.19 | 0.00 | 1.42 |
| -0.56 | -1.44 | 0.34 | 0.57 | 0.24 | 1.40 | 0.45 | he_j063_osteoporosis | HP:0000939 | 2.15E-01 | 0.04 | 0.19 | 0.01 | 1.56 |
| 0.11 | -0.07 | 0.29 | 1.12 | 0.93 | 1.34 | 0.09 | eb_i221_green_beans | ECTO:0070062 | 2.25E-01 | 2.71 | 0.91 | 0.04 | 1.51 |
| 0.65 | -0.38 | 1.77 | 1.92 | 0.68 | 5.90 | 0.54 | CHEBI_6741 | CHEBI:6741 | 2.26E-01 | 0.02 | 0.14 | 0.00 | 1.24 |
| 0.12 | -0.07 | 0.31 | 1.12 | 0.93 | 1.36 | 0.10 | eb_i199_peach_plum | ECTO:0070182 | 2.34E-01 | 2.10 | 0.88 | 0.02 | 1.72 |
| 0.84 | -0.49 | 2.41 | 2.31 | 0.61 | 11.15 | 0.71 | ea_b194c_textile_dye_parq | ECTO:0500008 | 2.41E-01 | 0.01 | 0.11 | 0.00 | 1.35 |
| 0.37 | -0.25 | 1.01 | 1.45 | 0.78 | 2.73 | 0.32 | ea_b146d_xrays_parq | ECTO:8000046 | 2.42E-01 | 0.08 | 0.27 | 0.00 | 1.69 |
| 0.68 | -0.45 | 1.93 | 1.98 | 0.64 | 6.90 | 0.59 | ea_b194d_paper_dye_parq | ECTO:0500007 | 2.51E-01 | 0.02 | 0.13 | 0.00 | 1.26 |
| 0.24 | -0.18 | 0.67 | 1.27 | 0.83 | 1.95 | 0.22 | he_f041_polyps | HP:0200063 | 2.64E-01 | 0.17 | 0.38 | 0.01 | 1.44 |
| -0.30 | -0.84 | 0.23 | 0.74 | 0.43 | 1.26 | 0.27 | ea_b116a_isopropanol_parq | ECTO:9000099 | 2.68E-01 | 0.14 | 0.35 | 0.00 | 1.89 |
| 0.07 | -0.05 | 0.19 | 1.07 | 0.95 | 1.21 | 0.06 | eb_i175_milk_nondairy | ECTO:0070004 | 2.69E-01 | 1.74 | 1.28 | 0.02 | 1.33 |
| 0.31 | -0.24 | 0.88 | 1.37 | 0.78 | 2.42 | 0.29 | ea_b104b_ammonia_parq | ECTO:9000031 | 2.72E-01 | 0.08 | 0.28 | 0.00 | 1.55 |
| 0.46 | -0.36 | 1.31 | 1.59 | 0.70 | 3.70 | 0.42 | CHEBI_46195 | CHEBI:46195 | 2.74E-01 | 0.03 | 0.17 | 0.00 | 1.31 |
| 0.29 | -0.23 | 0.82 | 1.34 | 0.79 | 2.27 | 0.27 | he_m089_endometriosis | MONDO:0005133 | 2.79E-01 | 0.12 | 0.32 | 0.00 | 1.34 |
| 0.10 | -0.09 | 0.30 | 1.11 | 0.92 | 1.34 | 0.10 | eb_i235_canned_tuna | ECTO:0070091 | 2.81E-01 | 1.98 | 0.85 | 0.02 | 1.36 |
| 0.39 | -0.32 | 1.14 | 1.48 | 0.72 | 3.11 | 0.37 | he_b010_cardiac_arrhythmia | HP:0011675 | 2.88E-01 | 0.05 | 0.22 | 0.00 | 1.30 |
| 0.24 | -0.21 | 0.69 | 1.27 | 0.81 | 2.00 | 0.23 | he_j067_osteoarthritis_parq | HP:0002758 | 2.93E-01 | 0.19 | 0.39 | 0.02 | 1.50 |
| 0.80 | -0.69 | 2.56 | 2.23 | 0.50 | 12.88 | 0.80 | ea_b140a_nitrous_oxide_parq | ECTO:9000051 | 3.17E-01 | 0.02 | 0.14 | 0.00 | 1.77 |
| -0.10 | -0.31 | 0.10 | 0.90 | 0.73 | 1.11 | 0.11 | eb_i195_cantaloupe | ECTO:0070026 | 3.24E-01 | 2.05 | 0.81 | 0.02 | 1.60 |
| -0.09 | -0.27 | 0.09 | 0.91 | 0.76 | 1.09 | 0.09 | eb_i201_raspberries | ECTO:0070034 | 3.24E-01 | 1.94 | 0.92 | 0.02 | 1.56 |
| -0.60 | -1.87 | 0.57 | 0.55 | 0.15 | 1.78 | 0.61 | CHEBI_9123 | CHEBI:9123 | 3.27E-01 | 0.02 | 0.15 | 0.00 | 1.27 |
| 0.47 | -0.46 | 1.45 | 1.60 | 0.63 | 4.28 | 0.48 | CHEBI_27300 | CHEBI:27300 | 3.31E-01 | 0.03 | 0.16 | 0.00 | 1.25 |
| 0.45 | -0.47 | 1.41 | 1.57 | 0.63 | 4.11 | 0.47 | ea_b182_adhesives_derived | ECTO:7000130 | 3.43E-01 | 0.09 | 0.29 | 0.10 | 3.07 |
| -0.49 | -1.55 | 0.55 | 0.61 | 0.21 | 1.74 | 0.53 | CHEBI_50275 | CHEBI:50275 | 3.54E-01 | 0.02 | 0.16 | 0.00 | 1.63 |
| 0.48 | -0.53 | 1.59 | 1.62 | 0.59 | 4.89 | 0.53 | CHEBI_8776 | CHEBI:8776 | 3.61E-01 | 0.02 | 0.14 | 0.00 | 1.31 |
| 0.21 | -0.25 | 0.67 | 1.24 | 0.78 | 1.96 | 0.23 | he_j062_bone_loss | HP:0000938 | 3.65E-01 | 0.20 | 0.40 | 0.01 | 1.70 |
| -0.52 | -1.67 | 0.60 | 0.59 | 0.19 | 1.82 | 0.58 | ea_b182a_white_glue_parq | ECTO:9000079 | 3.65E-01 | 0.06 | 0.24 | 0.00 | 3.31 |
| 0.40 | -0.47 | 1.30 | 1.50 | 0.63 | 3.68 | 0.45 | ea_b110_metal_derived | ECTO:9002163 | 3.67E-01 | 0.05 | 0.22 | 0.14 | 1.61 |
| 0.50 | -0.62 | 1.65 | 1.65 | 0.54 | 5.21 | 0.57 | CHEBI_39548 | CHEBI:39548 | 3.81E-01 | 0.05 | 0.22 | 0.00 | 1.42 |
| 0.09 | -0.11 | 0.29 | 1.09 | 0.90 | 1.33 | 0.10 | eb_i237_dark_fish | ECTO:0070209 | 3.82E-01 | 1.97 | 0.89 | 0.02 | 1.67 |
| -0.49 | -1.60 | 0.64 | 0.62 | 0.20 | 1.89 | 0.57 | CHEBI_30660 | CHEBI:30660 | 3.93E-01 | 0.07 | 0.25 | 0.00 | 2.59 |
| -0.08 | -0.26 | 0.11 | 0.93 | 0.77 | 1.11 | 0.09 | eb_i222_tofu | ECTO:0070185 | 4.08E-01 | 1.56 | 0.91 | 0.02 | 1.41 |
| 0.23 | -0.32 | 0.78 | 1.26 | 0.73 | 2.18 | 0.28 | he_f038_lactose_intolerance | HP:0004789 | 4.16E-01 | 0.09 | 0.28 | 0.01 | 1.27 |
| -0.31 | -1.08 | 0.45 | 0.73 | 0.34 | 1.56 | 0.39 | ea_b146c_radiation_parq | ECTO:0000001 | 4.23E-01 | 0.06 | 0.24 | 0.00 | 1.75 |
| -0.08 | -0.29 | 0.12 | 0.92 | 0.75 | 1.13 | 0.10 | eb_i253_tortillas | ECTO:0070128 | 4.23E-01 | 2.23 | 0.86 | 0.03 | 1.57 |
| 0.20 | -0.30 | 0.70 | 1.22 | 0.74 | 2.01 | 0.25 | ea_b194_ink_derived | ECTO:7000157 | 4.26E-01 | 0.32 | 0.47 | 0.08 | 1.54 |
| -0.21 | -0.73 | 0.31 | 0.81 | 0.48 | 1.36 | 0.26 | he_d030_asthma_parq | MONDO:0004979 | 4.28E-01 | 0.13 | 0.34 | 0.01 | 1.34 |
| -0.05 | -0.19 | 0.08 | 0.95 | 0.83 | 1.09 | 0.07 | eb_i240_oatmeal | ECTO:0070107 | 4.54E-01 | 2.38 | 1.14 | 0.02 | 1.29 |
| 0.39 | -0.62 | 1.47 | 1.47 | 0.54 | 4.35 | 0.53 | ea_b170d_acrylic_parq | ECTO:7000142 | 4.61E-01 | 0.03 | 0.17 | 0.00 | 1.55 |
| 0.15 | -0.26 | 0.56 | 1.16 | 0.77 | 1.75 | 0.21 | he_m088_hormone_replacement | ECTO:2000005 | 4.75E-01 | 0.27 | 0.45 | 0.01 | 1.74 |
| 0.43 | -0.77 | 1.64 | 1.54 | 0.46 | 5.15 | 0.61 | CHEBI_5163 | CHEBI:5163 | 4.78E-01 | 0.04 | 0.20 | 0.00 | 2.88 |
| -0.22 | -0.84 | 0.41 | 0.80 | 0.43 | 1.51 | 0.32 | eb_a014_potassium_parq | ECTO:9000253 | 4.96E-01 | 0.06 | 0.24 | 0.01 | 1.39 |
| 0.34 | -0.64 | 1.32 | 1.40 | 0.53 | 3.75 | 0.50 | ea_b092h_toluene_parq | ECTO:9000036 | 5.00E-01 | 0.05 | 0.23 | 0.00 | 2.79 |
| 0.55 | -1.06 | 2.32 | 1.73 | 0.35 | 10.14 | 0.84 | he_h057_sjogrens | MONDO:0010030 | 5.14E-01 | 0.01 | 0.10 | 0.01 | 1.43 |
| -0.37 | -1.54 | 0.80 | 0.69 | 0.21 | 2.21 | 0.59 | CHEBI_9654 | CHEBI:9654 | 5.28E-01 | 0.02 | 0.15 | 0.00 | 1.31 |
| 0.23 | -0.53 | 1.02 | 1.26 | 0.59 | 2.78 | 0.39 | ea_b194a_hair_dye_parq | ECTO:7000151 | 5.56E-01 | 0.06 | 0.23 | 0.00 | 1.54 |
| 0.57 | -1.45 | 2.89 | 1.77 | 0.23 | 18.01 | 1.08 | ea_b158a_sodium_oh_parq | ECTO:9000022 | 5.98E-01 | 0.05 | 0.21 | 0.00 |  |
| 0.42 | -1.20 | 2.02 | 1.52 | 0.30 | 7.53 | 0.80 | he_f039_crohns | HP:0100280 | 5.99E-01 | 0.01 | 0.10 | 0.01 | 1.19 |
| 0.28 | -0.76 | 1.36 | 1.32 | 0.47 | 3.88 | 0.53 | CHEBI_77249 | CHEBI:77249 | 6.01E-01 | 0.02 | 0.13 | 0.00 | 1.29 |
| 0.10 | -0.28 | 0.47 | 1.10 | 0.76 | 1.60 | 0.19 | he_b008_high_cholesterol | HP:0003124 | 6.14E-01 | 0.31 | 0.46 | 0.00 | 1.62 |
| -0.23 | -1.18 | 0.70 | 0.80 | 0.31 | 2.01 | 0.47 | CHEBI_18332 | CHEBI:18332 | 6.28E-01 | 0.08 | 0.28 | 0.00 | 3.67 |
| 0.22 | -0.82 | 1.27 | 1.24 | 0.44 | 3.57 | 0.53 | ea_b176d_isoflurane_parq | ECTO:9002166 | 6.84E-01 | 0.02 | 0.15 | 0.00 | 1.47 |
| 0.20 | -0.75 | 1.16 | 1.22 | 0.47 | 3.19 | 0.48 | ea_b092a_benzene_parq | ECTO:9000034 | 6.85E-01 | 0.04 | 0.20 | 0.00 | 2.12 |
| -0.19 | -1.14 | 0.77 | 0.82 | 0.32 | 2.15 | 0.48 | CHEBI_43755 | CHEBI:43755 | 6.87E-01 | 0.06 | 0.24 | 0.00 | 3.66 |
| 0.12 | -0.48 | 0.72 | 1.12 | 0.62 | 2.05 | 0.31 | he_m084_ovaries_parq | MAXO:0001067 | 7.02E-01 | 0.15 | 0.36 | 0.00 | 2.15 |
| 0.23 | -1.01 | 1.50 | 1.26 | 0.36 | 4.47 | 0.63 | CHEBI_6905 | CHEBI:6905 | 7.14E-01 | 0.04 | 0.19 | 0.00 | 3.27 |
| -0.15 | -0.96 | 0.66 | 0.86 | 0.38 | 1.93 | 0.41 | ea_b098d_motor_oil_parq | ECTO:0500004 | 7.18E-01 | 0.04 | 0.19 | 0.00 | 1.35 |
| -0.19 | -1.26 | 0.89 | 0.83 | 0.28 | 2.42 | 0.54 | ea_b182b_rubber_cement_parq | ECTO:7000149 | 7.33E-01 | 0.04 | 0.19 | 0.00 | 1.89 |
| 0.04 | -0.18 | 0.25 | 1.04 | 0.84 | 1.29 | 0.11 | eb_i238_other_fish | ECTO:0070189 | 7.36E-01 | 1.86 | 0.80 | 0.02 | 1.57 |
| 0.19 | -0.93 | 1.41 | 1.21 | 0.40 | 4.09 | 0.59 | he_f045_fatty_liver | MONDO:0004790 | 7.44E-01 | 0.02 | 0.14 | 0.00 | 1.33 |
| 0.10 | -0.62 | 0.84 | 1.11 | 0.54 | 2.31 | 0.37 | CHEBI_6541 | CHEBI:6541 | 7.80E-01 | 0.05 | 0.21 | 0.00 | 1.49 |
| 0.10 | -0.63 | 0.84 | 1.10 | 0.53 | 2.31 | 0.37 | he_h055_fibromyalgia | MONDO:0005546 | 7.99E-01 | 0.05 | 0.22 | 0.01 | 1.52 |
| 0.02 | -0.14 | 0.18 | 1.02 | 0.87 | 1.20 | 0.08 | eb_i243_other_crackers | ECTO:0070109 | 8.02E-01 | 2.27 | 0.98 | 0.03 | 1.39 |
| -0.13 | -1.22 | 0.96 | 0.88 | 0.29 | 2.61 | 0.55 | CHEBI_6904 | CHEBI:6904 | 8.15E-01 | 0.05 | 0.22 | 0.00 | 3.34 |
| 0.05 | -0.40 | 0.50 | 1.05 | 0.67 | 1.65 | 0.23 | he_c023_thyroid_disease_parq | MONDO:0003240 | 8.21E-01 | 0.17 | 0.38 | 0.01 | 1.55 |
| 0.02 | -0.16 | 0.20 | 1.02 | 0.85 | 1.23 | 0.09 | eb_i211_cabbage | ECTO:0070045 | 8.21E-01 | 2.41 | 0.92 | 0.02 | 1.59 |
| 0.09 | -0.74 | 0.96 | 1.10 | 0.47 | 2.61 | 0.43 | CHEBI_42797 | CHEBI:42797 | 8.26E-01 | 0.03 | 0.18 | 0.00 | 1.34 |
| -0.03 | -0.33 | 0.26 | 0.97 | 0.72 | 1.30 | 0.15 | ea_a060_pest_regularly_parq | ECTO:0000530 | 8.26E-01 | 0.44 | 0.50 | 0.05 | 1.21 |
| 0.09 | -0.71 | 0.90 | 1.09 | 0.49 | 2.45 | 0.41 | eb_a020_flaxseed_oil_parq | ECTO:0070165 | 8.28E-01 | 0.04 | 0.18 | 0.01 | 1.30 |
| 0.06 | -0.52 | 0.64 | 1.06 | 0.59 | 1.90 | 0.30 | he_c021_pre_diabetes_parq | MONDO:0006920 | 8.47E-01 | 0.12 | 0.32 | 0.01 | 1.77 |
| 0.02 | -0.16 | 0.19 | 1.02 | 0.85 | 1.21 | 0.09 | eb_i196_grapes | ECTO:0070027 | 8.65E-01 | 2.50 | 0.95 | 0.02 | 1.67 |
| -0.06 | -0.82 | 0.70 | 0.94 | 0.44 | 2.01 | 0.39 | he_b016_raynauds | MONDO:0008364 | 8.81E-01 | 0.05 | 0.21 | 0.01 | 1.31 |
| 0.04 | -0.46 | 0.53 | 1.04 | 0.63 | 1.70 | 0.25 | he_k071_urticaria | HP:0001025 | 8.86E-01 | 0.14 | 0.34 | 0.01 | 1.29 |
| 0.01 | -0.21 | 0.24 | 1.02 | 0.81 | 1.27 | 0.12 | eb_i256_pizza | ECTO:0070131 | 8.98E-01 | 2.28 | 0.74 | 0.02 | 1.58 |
| -0.01 | -0.24 | 0.21 | 0.99 | 0.79 | 1.23 | 0.11 | eb_i252_pasta | ECTO:0070125 | 9.05E-01 | 2.65 | 0.87 | 0.02 | 1.99 |
| 0.06 | -1.04 | 1.17 | 1.06 | 0.35 | 3.21 | 0.55 | CHEBI_50686 | CHEBI:50686 | 9.15E-01 | 0.02 | 0.13 | 0.00 | 1.35 |
| 0.03 | -0.50 | 0.56 | 1.03 | 0.60 | 1.75 | 0.27 | eb_a027_omega3_parq | ECTO:0070171 | 9.26E-01 | 0.10 | 0.29 | 0.01 | 1.51 |
| -0.07 | -1.65 | 1.51 | 0.94 | 0.19 | 4.54 | 0.78 | ea_b176f_nitrous_parq | ECTO:9000051 | 9.33E-01 | 0.02 | 0.14 | 0.00 | 1.87 |
| 0.06 | -1.44 | 1.57 | 1.06 | 0.24 | 4.80 | 0.76 | CHEBI_63618 | CHEBI:63618 | 9.36E-01 | 0.02 | 0.12 | 0.00 | 1.32 |
| 0.01 | -0.45 | 0.48 | 1.01 | 0.64 | 1.61 | 0.23 | CHEBI_15365 | CHEBI:15365 | 9.52E-01 | 0.12 | 0.32 | 0.00 | 1.42 |
| -0.02 | -0.71 | 0.68 | 0.98 | 0.49 | 1.97 | 0.35 | he_c022_diabetes_parq | MONDO:0005015 | 9.64E-01 | 0.08 | 0.27 | 0.03 | 2.03 |
| 0.00 | -0.17 | 0.18 | 1.00 | 0.84 | 1.20 | 0.09 | eb_i218_peas_lima_beans | ECTO:0070183 | 9.68E-01 | 2.33 | 0.94 | 0.03 | 1.54 |
| -0.01 | -0.41 | 0.39 | 0.99 | 0.66 | 1.48 | 0.20 | eb_a028_probiotic_parq | ECTO:0070000 | 9.72E-01 | 0.19 | 0.39 | 0.01 | 1.46 |
| 0.01 | -1.01 | 1.01 | 1.01 | 0.37 | 2.74 | 0.51 | CHEBI_87631 | CHEBI:87631 | 9.85E-01 | 0.11 | 0.31 | 0.00 | 3.79 |
| 0.01 | -1.01 | 1.01 | 1.01 | 0.37 | 2.74 | 0.51 | CHEBI_87631 | CHEBI:87631 | 9.85E-01 |  |  |  |  |

Supplemental Table 2C: Ovarian Cysts Logistic Regression

| **Log.odds** | **Log.odds.min** | **Log.odds.max** | **Odds** | **Odds.min** | **Odds.max** | **Standard.Error** | **Survey.question.label** | **Ontology.ID** | **p.value** | **mean_prevalence** | **sd_prevalence** | **miss_rate** | **mean_vif** |
| --- | --- | --- | --- | --- | --- | --- | --- | --- | --- | --- | --- | --- | --- |
| 1.43 | 0.83 | 2.05 | 4.17 | 2.30 | 7.75 | 0.31 | he_m084_ovaries_parq | MAXO:0001067 | 3.92E-06 | 0.15 | 0.36 | 0.00 |  |
| 0.27 | 0.11 | 0.43 | 1.31 | 1.12 | 1.54 | 0.08 | eb_i220_spinach_raw | ECTO:0070060 | 9.94E-04 | 2.61 | 1.11 | 0.03 | 1.59 |
| -2.02 | -3.33 | -0.85 | 0.13 | 0.04 | 0.43 | 0.63 | ea_b116b_methanol_parq | ECTO:9000028 | 1.35E-03 | 0.07 | 0.25 | 0.00 |  |
| 1.14 | 0.46 | 1.90 | 3.14 | 1.58 | 6.66 | 0.36 | he_m090_uterine_polyps | MONDO:0006195 | 1.73E-03 | 0.06 | 0.24 | 0.00 | 1.19 |
| -0.88 | -1.45 | -0.33 | 0.42 | 0.23 | 0.72 | 0.29 | he_m083_hysterectomy | MAXO:0001058 | 2.22E-03 | 0.20 | 0.40 | 0.00 |  |
| 1.39 | 0.48 | 2.46 | 4.03 | 1.62 | 11.68 | 0.50 | CHEBI_46195 | CHEBI:46195 | 4.96E-03 | 0.03 | 0.17 | 0.00 | 1.21 |
| -0.23 | -0.39 | -0.07 | 0.80 | 0.68 | 0.93 | 0.08 | eb_i208_bell_pepper | ECTO:0070042 | 4.99E-03 | 2.80 | 1.04 | 0.02 | 1.39 |
| 0.12 | 0.04 | 0.20 | 1.12 | 1.04 | 1.22 | 0.04 | eb_i264_coffee | ECTO:0070134 | 5.11E-03 | 3.57 | 1.94 | 0.03 | 1.21 |
| 0.2 | 0.05 | 0.34 | 1.22 | 1.05 | 1.41 | 0.07 | eb_i182_spread_butter | ECTO:0070013 | 7.84E-03 | 1.72 | 1.10 | 0.02 | 1.23 |
| -0.56 | -0.99 | -0.14 | 0.57 | 0.37 | 0.87 | 0.22 | he_m085_menopause_parq | GO:0042697 | 9.31E-03 | 0.50 | 0.50 | 0.01 |  |
| 0.51 | 0.13 | 0.89 | 1.66 | 1.13 | 2.44 | 0.19 | he_e032_migraine | HP:0002076 | 9.34E-03 | 0.25 | 0.43 | 0.01 | 1.21 |
| -0.26 | -0.46 | -0.06 | 0.77 | 0.63 | 0.94 | 0.10 | eb_i222_tofu | ECTO:0070185 | 1.22E-02 | 1.56 | 0.91 | 0.02 | 1.45 |
| 0.99 | 0.22 | 1.83 | 2.69 | 1.25 | 6.20 | 0.40 | CHEBI_3219 | CHEBI:3219 | 1.46E-02 | 0.05 | 0.21 | 0.00 | 1.18 |
| 0.93 | 0.20 | 1.70 | 2.53 | 1.22 | 5.48 | 0.38 | ea_b146d_xrays_parq | ECTO:8000046 | 1.49E-02 | 0.08 | 0.27 | 0.00 | 1.42 |
| 1.43 | 0.34 | 2.75 | 4.19 | 1.40 | 15.67 | 0.60 | CHEBI_27300 | CHEBI:27300 | 1.72E-02 | 0.03 | 0.16 | 0.00 | 1.22 |
| 0.49 | 0.09 | 0.89 | 1.63 | 1.09 | 2.45 | 0.21 | he_m091_uterine_tumors | HP:0000131 | 1.76E-02 | 0.24 | 0.43 | 0.00 | 1.26 |
| 1.14 | 0.16 | 2.16 | 3.13 | 1.17 | 8.67 | 0.51 | ea_b116a_isopropanol_parq | ECTO:9000099 | 2.47E-02 | 0.14 | 0.35 | 0.00 |  |
| 0.74 | 0.10 | 1.41 | 2.11 | 1.11 | 4.11 | 0.33 | he_g049_kidney_stones | HP:0000787 | 2.52E-02 | 0.07 | 0.26 | 0.01 | 1.22 |
| 0.46 | 0.04 | 0.89 | 1.58 | 1.04 | 2.43 | 0.22 | eb_a028_probiotic_parq | ECTO:0070000 | 3.32E-02 | 0.19 | 0.39 | 0.01 | 1.33 |
| 1.27 | 0.17 | 2.56 | 3.54 | 1.19 | 13.00 | 0.59 | CHEBI_9584 | CHEBI:9584 | 3.32E-02 | 0.01 | 0.12 | 0.00 | 1.20 |
| 0.85 | 0.08 | 1.68 | 2.34 | 1.09 | 5.38 | 0.40 | he_h055_fibromyalgia | MONDO:0005546 | 3.55E-02 | 0.05 | 0.22 | 0.01 | 1.30 |
| -0.18 | -0.35 | -0.01 | 0.84 | 0.71 | 0.99 | 0.09 | eb_i194_blueberries | ECTO:0070025 | 3.70E-02 | 2.58 | 1.09 | 0.02 | 1.55 |
| -1.82 | -3.67 | -0.18 | 0.16 | 0.03 | 0.84 | 0.87 | ea_b116d_butanol_parq | ECTO:9000424 | 3.74E-02 | 0.01 | 0.11 | 0.00 | 1.17 |
| 1.45 | 0.19 | 3.01 | 4.25 | 1.21 | 20.24 | 0.70 | ea_b104_cleaning_derived | ECTO:0500011 | 3.78E-02 | 0.29 | 0.45 | 0.05 |  |
| 1.05 | 0.09 | 2.10 | 2.85 | 1.09 | 8.18 | 0.51 | ea_b092b_chloroform_parq | ECTO:9000042 | 3.89E-02 | 0.06 | 0.23 | 0.00 |  |
| 2.16 | 0.41 | 5.10 | 8.66 | 1.50 | >50 | 1.08 | CHEBI_9648 | CHEBI:9648 | 4.62E-02 | 0.01 | 0.10 | 0.00 | 1.16 |
| 0.72 | 0.01 | 1.47 | 2.05 | 1.01 | 4.34 | 0.37 | eb_a014_potassium_parq | ECTO:9000253 | 5.12E-02 | 0.06 | 0.24 | 0.01 | 1.43 |
| 1.14 | 0.00 | 2.47 | 3.13 | 1.00 | 11.87 | 0.62 | CHEBI_10126 | CHEBI:10126 | 6.35E-02 | 0.02 | 0.12 | 0.00 | 1.19 |
| -0.99 | -2.11 | 0.03 | 0.37 | 0.12 | 1.03 | 0.54 | ea_b182_adhesives_derived | ECTO:7000130 | 6.55E-02 | 0.09 | 0.29 | 0.10 |  |
| -0.95 | -2.05 | 0.02 | 0.39 | 0.13 | 1.02 | 0.52 | CHEBI_9654 | CHEBI:9654 | 6.82E-02 | 0.02 | 0.15 | 0.00 | 1.28 |
| 1.59 | -0.06 | 3.46 | 4.88 | 0.94 | 31.79 | 0.88 | ea_b164f_acrylic_parq | ECTO:7000142 | 7.20E-02 | 0.02 | 0.15 | 0.00 |  |
| 1.31 | -0.13 | 2.84 | 3.70 | 0.88 | 17.18 | 0.75 | CHEBI_6905 | CHEBI:6905 | 8.15E-02 | 0.04 | 0.19 | 0.00 |  |
| 0.44 | -0.05 | 0.95 | 1.56 | 0.95 | 2.58 | 0.26 | he_m089_endometriosis | MONDO:0005133 | 8.16E-02 | 0.12 | 0.32 | 0.00 | 1.29 |
| 0.48 | -0.06 | 1.03 | 1.62 | 0.94 | 2.80 | 0.28 | he_f042_gallbladder_disease | MONDO:0005281 | 8.28E-02 | 0.10 | 0.31 | 0.01 | 1.26 |
| -0.9 | -1.96 | 0.12 | 0.41 | 0.14 | 1.12 | 0.53 | ea_b116_alcohol_derived | ECTO:9000026 | 8.60E-02 | 0.19 | 0.39 | 0.11 |  |
| 1.2 | -0.14 | 2.64 | 3.32 | 0.87 | 14.03 | 0.71 | ea_b182e_epoxy_parq | ECTO:9002142 | 8.86E-02 | 0.02 | 0.15 | 0.00 |  |
| 0.87 | -0.09 | 1.94 | 2.38 | 0.91 | 6.93 | 0.51 | CHEBI_7772 | CHEBI:7772 | 9.00E-02 | 0.05 | 0.22 | 0.00 | 1.56 |
| 0.38 | -0.07 | 0.83 | 1.46 | 0.94 | 2.29 | 0.23 | eb_a007_vitamin_c_parq | ECTO:9000143 | 9.60E-02 | 0.20 | 0.40 | 0.01 | 1.48 |
| -0.78 | -1.72 | 0.13 | 0.46 | 0.18 | 1.14 | 0.47 | eb_a002_vitamin_a_parq | ECTO:9000125 | 9.65E-02 | 0.06 | 0.23 | 0.01 | 1.60 |
| 0.63 | -0.12 | 1.42 | 1.89 | 0.89 | 4.14 | 0.39 | CHEBI_39548 | CHEBI:39548 | 1.04E-01 | 0.05 | 0.22 | 0.00 | 1.36 |
| 0.37 | -0.10 | 0.84 | 1.45 | 0.91 | 2.32 | 0.24 | he_m088_hormone_replacement | ECTO:2000005 | 1.21E-01 | 0.27 | 0.45 | 0.01 |  |
| 1.12 | -0.27 | 2.60 | 3.06 | 0.77 | 13.43 | 0.72 | ea_b152e_nitric_acid_parq | ECTO:9000020 | 1.23E-01 | 0.03 | 0.17 | 0.00 |  |
| 0.27 | -0.08 | 0.62 | 1.30 | 0.92 | 1.85 | 0.18 | eb_a008_vitamin_d_parq | ECTO:9000133 | 1.35E-01 | 0.51 | 0.50 | 0.01 | 1.56 |
| 0.82 | -0.24 | 1.97 | 2.26 | 0.79 | 7.16 | 0.55 | CHEBI_43755 | CHEBI:43755 | 1.42E-01 | 0.06 | 0.24 | 0.00 |  |
| 0.1 | -0.04 | 0.25 | 1.11 | 0.96 | 1.28 | 0.07 | eb_i189_cheese_lowfat | ECTO:0070020 | 1.48E-01 | 1.88 | 1.13 | 0.03 | 1.19 |
| 0.3 | -0.11 | 0.72 | 1.35 | 0.90 | 2.05 | 0.21 | eb_a019_fish_oil_parq | ECTO:0070164 | 1.50E-01 | 0.23 | 0.42 | 0.01 | 1.36 |
| 0.87 | -0.31 | 2.15 | 2.40 | 0.73 | 8.55 | 0.62 | ea_b182a_white_glue_parq | ECTO:9000079 | 1.60E-01 | 0.06 | 0.24 | 0.00 |  |
| 0.7 | -0.27 | 1.71 | 2.02 | 0.76 | 5.54 | 0.50 | ea_b116c_ethanol_parq | ECTO:9000027 | 1.63E-01 | 0.10 | 0.31 | 0.00 |  |
| -0.76 | -1.86 | 0.30 | 0.47 | 0.16 | 1.35 | 0.55 | ea_b170_paint_derived | ECTO:0500024 | 1.65E-01 | 0.09 | 0.28 | 0.09 |  |
| -0.58 | -1.43 | 0.24 | 0.56 | 0.24 | 1.27 | 0.42 | eb_a020_flaxseed_oil_parq | ECTO:0070165 | 1.71E-01 | 0.04 | 0.18 | 0.01 | 1.24 |
| -0.92 | -2.48 | 0.35 | 0.40 | 0.08 | 1.42 | 0.70 | ea_b104a_bleach_parq | ECTO:9000076 | 1.90E-01 | 0.27 | 0.45 | 0.00 |  |
| 0.1 | -0.05 | 0.25 | 1.11 | 0.95 | 1.29 | 0.08 | eb_i267_chocolate_milk | ECTO:0070137 | 1.93E-01 | 2.34 | 1.02 | 0.03 | 1.17 |
| 0.77 | -0.35 | 2.01 | 2.16 | 0.70 | 7.47 | 0.59 | ea_b194d_paper_dye_parq | ECTO:0500007 | 1.94E-01 | 0.02 | 0.13 | 0.00 | 1.23 |
| -0.91 | -2.32 | 0.44 | 0.40 | 0.10 | 1.55 | 0.70 | CHEBI_5163 | CHEBI:5163 | 1.94E-01 | 0.04 | 0.20 | 0.00 |  |
| 0.68 | -0.38 | 1.82 | 1.97 | 0.68 | 6.17 | 0.55 | CHEBI_50686 | CHEBI:50686 | 2.21E-01 | 0.02 | 0.13 | 0.00 | 1.30 |
| 1.04 | -0.48 | 3.02 | 2.83 | 0.62 | 20.46 | 0.85 | he_j064_gout | MONDO:0005393 | 2.23E-01 | 0.01 | 0.11 | 0.01 | 1.20 |
| 0.36 | -0.22 | 0.95 | 1.43 | 0.80 | 2.58 | 0.30 | eb_a006_vitamin_b_comp_parq | ECTO:9002169 | 2.30E-01 | 0.12 | 0.33 | 0.01 | 1.63 |
| -0.47 | -1.25 | 0.30 | 0.62 | 0.29 | 1.35 | 0.39 | eb_a003_vitamin_b3_parq | ECTO:9002151 | 2.31E-01 | 0.08 | 0.28 | 0.01 | 1.86 |
| -0.57 | -1.55 | 0.40 | 0.57 | 0.21 | 1.49 | 0.49 | he_d025_copd | MONDO:0005002 | 2.48E-01 | 0.03 | 0.18 | 0.01 | 1.30 |
| 0.28 | -0.20 | 0.77 | 1.33 | 0.82 | 2.16 | 0.25 | he_k071_urticaria | HP:0001025 | 2.48E-01 | 0.14 | 0.34 | 0.01 | 1.25 |
| 0.28 | -0.24 | 0.80 | 1.32 | 0.79 | 2.22 | 0.26 | he_d030_asthma_parq | MONDO:0004979 | 2.90E-01 | 0.13 | 0.34 | 0.01 | 1.27 |
| 0.7 | -0.56 | 2.08 | 2.02 | 0.57 | 7.99 | 0.66 | eb_a015_selenium_parq | ECTO:9000192 | 2.91E-01 | 0.02 | 0.14 | 0.01 | 1.48 |
| 0.35 | -0.33 | 1.04 | 1.42 | 0.72 | 2.83 | 0.35 | ea_b194g_gel_ink_parq | ECTO:7000153 | 3.14E-01 | 0.08 | 0.27 | 0.00 | 1.38 |
| 0.37 | -0.37 | 1.13 | 1.45 | 0.69 | 3.09 | 0.38 | CHEBI_6541 | CHEBI:6541 | 3.27E-01 | 0.05 | 0.21 | 0.00 | 1.34 |
| 0.37 | -0.38 | 1.15 | 1.44 | 0.68 | 3.15 | 0.39 | he_g050_kidney_infection | HP:0012330 | 3.44E-01 | 0.05 | 0.22 | 0.01 | 1.22 |
| -0.57 | -1.83 | 0.59 | 0.57 | 0.16 | 1.80 | 0.61 | CHEBI_6904 | CHEBI:6904 | 3.49E-01 | 0.05 | 0.22 | 0.00 |  |
| 0.18 | -0.21 | 0.56 | 1.19 | 0.81 | 1.75 | 0.20 | he_b008_high_cholesterol | HP:0003124 | 3.67E-01 | 0.31 | 0.46 | 0.00 | 1.39 |
| 0.09 | -0.10 | 0.28 | 1.09 | 0.90 | 1.32 | 0.10 | eb_i235_canned_tuna | ECTO:0070091 | 3.68E-01 | 1.98 | 0.85 | 0.02 | 1.23 |
| -0.2 | -0.65 | 0.24 | 0.82 | 0.52 | 1.28 | 0.23 | he_c023_thyroid_disease_parq | MONDO:0003240 | 3.74E-01 | 0.17 | 0.38 | 0.01 |  |
| 0.21 | -0.26 | 0.69 | 1.24 | 0.77 | 2.00 | 0.24 | he_f041_polyps | HP:0200063 | 3.81E-01 | 0.17 | 0.38 | 0.01 | 1.31 |
| 0.44 | -0.56 | 1.50 | 1.56 | 0.57 | 4.48 | 0.52 | CHEBI_5051 | CHEBI:5051 | 3.90E-01 | 0.02 | 0.13 | 0.00 | 1.14 |
| -0.3 | -0.98 | 0.38 | 0.74 | 0.38 | 1.46 | 0.34 | eb_a021_folic_acid_parq | ECTO:9000123 | 3.91E-01 | 0.07 | 0.25 | 0.01 | 1.30 |
| -0.08 | -0.26 | 0.10 | 0.93 | 0.77 | 1.11 | 0.09 | eb_i229_bacon | ECTO:0070185 | 4.07E-01 | 2.28 | 0.93 | 0.02 | 1.30 |
| 0.28 | -0.40 | 0.98 | 1.32 | 0.67 | 2.65 | 0.35 | CHEBI_3562 | CHEBI:3562 | 4.22E-01 | 0.04 | 0.21 | 0.00 | 1.17 |
| -0.26 | -0.91 | 0.38 | 0.77 | 0.40 | 1.46 | 0.33 | eb_a018_coq10_parq | ECTO:0070163 | 4.25E-01 | 0.08 | 0.26 | 0.01 | 1.39 |
| -0.05 | -0.17 | 0.07 | 0.95 | 0.84 | 1.08 | 0.06 | eb_i261_tea_decaf | ECTO:0070132 | 4.34E-01 | 2.08 | 1.34 | 0.03 | 1.21 |
| -0.22 | -0.79 | 0.34 | 0.80 | 0.46 | 1.41 | 0.29 | eb_a012_iron_parq | ECTO:9000087 | 4.42E-01 | 0.10 | 0.30 | 0.01 | 1.28 |
| 0.54 | -0.86 | 2.05 | 1.72 | 0.42 | 7.74 | 0.73 | ea_b164_stains_derived | ECTO:0500015 | 4.55E-01 | 0.05 | 0.22 | 0.08 |  |
| -0.7 | -2.74 | 1.16 | 0.50 | 0.06 | 3.19 | 0.97 | CHEBI_3756 | CHEBI:3756 | 4.71E-01 | 0.01 | 0.11 | 0.00 |  |
| -0.24 | -0.91 | 0.42 | 0.78 | 0.40 | 1.52 | 0.34 | he_b010_cardiac_arrhythmia | HP:0011675 | 4.74E-01 | 0.05 | 0.22 | 0.00 | 1.18 |
| -0.4 | -1.63 | 0.77 | 0.67 | 0.20 | 2.16 | 0.61 | ea_b182b_rubber_cement_parq | ECTO:7000149 | 5.16E-01 | 0.04 | 0.19 | 0.00 | 1.21 |
| -0.24 | -0.99 | 0.51 | 0.79 | 0.37 | 1.66 | 0.38 | he_b016_raynauds | MONDO:0008364 | 5.27E-01 | 0.05 | 0.21 | 0.01 | 1.20 |
| -0.38 | -1.65 | 0.81 | 0.68 | 0.19 | 2.26 | 0.62 | CHEBI_18332 | CHEBI:18332 | 5.35E-01 | 0.08 | 0.28 | 0.00 |  |
| 0.37 | -0.78 | 1.57 | 1.44 | 0.46 | 4.81 | 0.60 | ea_b134e_talc_parq | ECTO:7000028 | 5.40E-01 | 0.03 | 0.16 | 0.00 | 1.18 |
| 0.15 | -0.33 | 0.63 | 1.16 | 0.72 | 1.87 | 0.24 | he_j062_bone_loss | HP:0000938 | 5.41E-01 | 0.20 | 0.40 | 0.01 | 1.51 |
| 0.14 | -0.32 | 0.60 | 1.15 | 0.73 | 1.83 | 0.23 | ea_b194_ink_derived | ECTO:7000157 | 5.42E-01 | 0.32 | 0.47 | 0.08 |  |
| 0.39 | -0.85 | 1.65 | 1.47 | 0.43 | 5.22 | 0.64 | ea_b170f_acetone_parq | ECTO:9000085 | 5.45E-01 | 0.05 | 0.21 | 0.00 |  |
| 0.22 | -0.50 | 0.97 | 1.25 | 0.60 | 2.64 | 0.37 | eb_a009_vitamin_e_parq | ECTO:9000224 | 5.50E-01 | 0.07 | 0.26 | 0.01 | 1.52 |
| -0.27 | -1.20 | 0.67 | 0.76 | 0.30 | 1.96 | 0.47 | ea_b092a_benzene_parq | ECTO:9000034 | 5.68E-01 | 0.04 | 0.20 | 0.00 |  |
| 0.13 | -0.32 | 0.58 | 1.14 | 0.73 | 1.78 | 0.23 | he_j067_osteoarthritis_parq | HP:0002758 | 5.73E-01 | 0.19 | 0.39 | 0.02 | 1.39 |
| 0.28 | -0.72 | 1.31 | 1.32 | 0.48 | 3.71 | 0.52 | ea_b128_plastic_derived | ECTO:7000147 | 5.91E-01 | 0.05 | 0.21 | 0.16 |  |
| -0.23 | -1.09 | 0.63 | 0.80 | 0.34 | 1.87 | 0.44 | ea_b146c_radiation_parq | ECTO:0000001 | 5.98E-01 | 0.06 | 0.24 | 0.00 | 1.41 |
| -0.14 | -0.65 | 0.37 | 0.87 | 0.52 | 1.45 | 0.26 | CHEBI_15365 | CHEBI:15365 | 5.98E-01 | 0.12 | 0.32 | 0.00 | 1.31 |
| 0.33 | -0.88 | 1.59 | 1.39 | 0.42 | 4.89 | 0.62 | CHEBI_30660 | CHEBI:30660 | 5.99E-01 | 0.07 | 0.25 | 0.00 |  |
| 0.11 | -0.35 | 0.58 | 1.12 | 0.70 | 1.79 | 0.24 | eb_a005_vitamin_b12_parq | ECTO:9000229 | 6.33E-01 | 0.21 | 0.40 | 0.01 | 1.50 |
| -0.14 | -0.73 | 0.45 | 0.87 | 0.48 | 1.56 | 0.30 | ea_b194h_pen_ink_parq | ECTO:0500010 | 6.42E-01 | 0.13 | 0.34 | 0.00 | 1.40 |
| 0.09 | -0.29 | 0.47 | 1.09 | 0.75 | 1.59 | 0.19 | he_i059_iron_anemia | HP:0001891 | 6.46E-01 | 0.24 | 0.43 | 0.00 | 1.21 |
| -0.17 | -0.89 | 0.55 | 0.85 | 0.41 | 1.73 | 0.37 | he_c022_diabetes_parq | MONDO:0005015 | 6.49E-01 | 0.08 | 0.27 | 0.03 | 1.59 |
| -0.27 | -1.72 | 1.06 | 0.76 | 0.18 | 2.88 | 0.69 | CHEBI_36796 | CHEBI:36796 | 6.98E-01 | 0.01 | 0.12 | 0.00 | 1.31 |
| 0.15 | -0.64 | 0.97 | 1.17 | 0.53 | 2.63 | 0.41 | eb_a016_zinc_parq | ECTO:9000954 | 7.05E-01 | 0.06 | 0.23 | 0.01 | 1.69 |
| -0.27 | -1.77 | 1.13 | 0.76 | 0.17 | 3.10 | 0.72 | he_b019_stroke_mini | HP:0002326 | 7.06E-01 | 0.01 | 0.12 | 0.00 | 1.25 |
| -0.12 | -0.78 | 0.52 | 0.88 | 0.46 | 1.69 | 0.33 | CHEBI_6446 | CHEBI:6446 | 7.06E-01 | 0.14 | 0.34 | 0.00 |  |
| -0.11 | -0.75 | 0.51 | 0.89 | 0.47 | 1.67 | 0.32 | CHEBI_5778 | CHEBI:5778 | 7.26E-01 | 0.08 | 0.28 | 0.00 | 1.24 |
| 0.1 | -0.51 | 0.71 | 1.10 | 0.60 | 2.04 | 0.31 | eb_a024_glucosamine_parq | ECTO:0070168 | 7.52E-01 | 0.08 | 0.27 | 0.01 | 1.23 |
| 0.24 | -1.26 | 1.91 | 1.27 | 0.28 | 6.77 | 0.78 | CHEBI_28364 | CHEBI:28364 | 7.61E-01 | 0.02 | 0.14 | 0.00 |  |
| 0.12 | -0.73 | 0.97 | 1.13 | 0.48 | 2.64 | 0.43 | CHEBI_7476 | CHEBI:7476 | 7.80E-01 | 0.03 | 0.18 | 0.00 | 1.17 |
| -0.15 | -1.29 | 1.02 | 0.86 | 0.27 | 2.77 | 0.58 | CHEBI_50275 | CHEBI:50275 | 7.97E-01 | 0.02 | 0.16 | 0.00 | 1.50 |
| 0.1 | -0.64 | 0.84 | 1.10 | 0.53 | 2.31 | 0.37 | ea_b134_dust_derived | ECTO:7000001 | 7.98E-01 | 0.08 | 0.27 | 0.10 |  |
| -0.03 | -0.25 | 0.19 | 0.97 | 0.78 | 1.21 | 0.11 | eb_i223_winter_squash | ECTO:0070067 | 8.06E-01 | 1.69 | 0.84 | 0.02 | 1.61 |
| 0.1 | -0.74 | 0.95 | 1.11 | 0.48 | 2.57 | 0.43 | CHEBI_6538 | CHEBI:6538 | 8.08E-01 | 0.03 | 0.16 | 0.00 | 1.22 |
| -0.13 | -1.18 | 0.94 | 0.88 | 0.31 | 2.57 | 0.54 | he_j063_osteoporosis | HP:0000939 | 8.09E-01 | 0.04 | 0.19 | 0.01 | 1.40 |
| 0.07 | -0.49 | 0.63 | 1.07 | 0.61 | 1.87 | 0.28 | he_f038_lactose_intolerance | HP:0004789 | 8.16E-01 | 0.09 | 0.28 | 0.01 | 1.21 |
| -0.11 | -1.05 | 0.83 | 0.90 | 0.35 | 2.30 | 0.47 | CHEBI_2668 | CHEBI:2668 | 8.23E-01 | 0.03 | 0.18 | 0.00 | 1.26 |
| 0.08 | -0.96 | 1.14 | 1.08 | 0.38 | 3.12 | 0.53 | CHEBI_6741 | CHEBI:6741 | 8.79E-01 | 0.02 | 0.14 | 0.00 | 1.19 |
| -0.03 | -0.52 | 0.45 | 0.97 | 0.60 | 1.57 | 0.25 | eb_a013_magnesium_parq | ECTO:9000210 | 8.91E-01 | 0.16 | 0.37 | 0.01 | 1.54 |
| -0.01 | -0.19 | 0.17 | 0.99 | 0.83 | 1.19 | 0.09 | eb_i224_yams | ECTO:0070069 | 9.24E-01 | 2.40 | 0.94 | 0.02 | 1.40 |
| 0.07 | -1.52 | 1.64 | 1.08 | 0.22 | 5.15 | 0.80 | ea_b164d_polyurethane_parq | ECTO:9000082 | 9.25E-01 | 0.03 | 0.17 | 0.00 |  |
| 0.02 | -0.73 | 0.78 | 1.02 | 0.48 | 2.18 | 0.38 | ea_b194a_hair_dye_parq | ECTO:7000151 | 9.62E-01 | 0.06 | 0.23 | 0.00 | 1.27 |
| 0.02 | -0.82 | 0.87 | 1.02 | 0.44 | 2.38 | 0.43 | CHEBI_42797 | CHEBI:42797 | 9.67E-01 | 0.03 | 0.18 | 0.00 | 1.42 |
| 0.01 | -1.01 | 1.05 | 1.01 | 0.37 | 2.85 | 0.52 | ea_b152a_hydrochloric_acid_parq | ECTO:9000016 | 9.79E-01 | 0.06 | 0.24 | 0.00 |  |
| -0.01 | -0.59 | 0.58 | 0.99 | 0.56 | 1.78 | 0.30 | he_c021_pre_diabetes_parq | MONDO:0006920 | 9.84E-01 | 0.12 | 0.32 | 0.01 | 1.70 |
| 0.01 | -0.66 | 0.69 | 1.01 | 0.52 | 1.99 | 0.34 | ea_b104b_ammonia_parq | ECTO:9000031 | 9.84E-01 | 0.08 | 0.28 | 0.00 | 1.33 |
| 0 | -0.68 | 0.68 | 1.00 | 0.51 | 1.98 | 0.35 | he_f043_stomach_ulcer | HP:0002592 | 9.97E-01 | 0.05 | 0.22 | 0.01 | 1.26 |

Supplemental Table 3: KG predicted links

| prediction_score | FRD_of_interest | predicted_link_variable |
| --- | --- | --- |
| 1 | MONDO:0005133 | ECTO:9000087 |
| 1 | MONDO:0005133 | ECTO:9002151 |
| 1 | MONDO:0005133 | ECTO:0070001 |
| 1 | MONDO:0005133 | ECTO:0070014 |
| 1 | MONDO:0005133 | ECTO:0070186 |
| 1 | MONDO:0005133 | ECTO:0070091 |
| 1 | MONDO:0005133 | MONDO:0002251 |
| 1 | MONDO:0005133 | MONDO:0002974 |
| 1 | MONDO:0005133 | MONDO:0003240 |
| 1 | MONDO:0005133 | ECTO:0070043 |
| 1 | MONDO:0005133 | ECTO:0070069 |
| 1 | MONDO:0005133 | ECTO:0070166 |
| 1 | MONDO:0005133 | ECTO:0070135 |
| 1 | MONDO:0005133 | ECTO:0070137 |
| 1 | MONDO:0005133 | ECTO:0070033 |
| 1 | MONDO:0005133 | ECTO:0070184 |
| 1 | MONDO:0005133 | ECTO:0070034 |
| 1 | MONDO:0005133 | ECTO:0070163 |
| 1 | MONDO:0005133 | ECTO:0070129 |
| 1 | MONDO:0005133 | ECTO:7000144 |
| 1 | MONDO:0005133 | ECTO:9000038 |
| 1 | MONDO:0005133 | ECTO:7000143 |
| 1 | MONDO:0005133 | ECTO:9000052 |
| 1 | MONDO:0005133 | ECTO:0500008 |
| 1 | MONDO:0005133 | ECTO:0500017 |
| 1 | MONDO:0005133 | ECTO:0500016 |
| 1 | MONDO:0005133 | ECTO:9002164 |
| 1 | MONDO:0005133 | ECTO:9000031 |
| 1 | MONDO:0005133 | ECTO:9000018 |
| 1 | MONDO:0005133 | ECTO:9000074 |
| 1 | MONDO:0005133 | ECTO:9000063 |
| 1 | MONDO:0005133 | ECTO:7000030 |
| 1 | MONDO:0005133 | ECTO:9000103 |
| 1 | MONDO:0005133 | ECTO:0500023 |
| 1 | MONDO:0005133 | HP:0012887 |
| 1 | MONDO:0005133 | ECTO:0000207 |
| 1 | MONDO:0005133 | CHEBI:10125 |
| 1 | MONDO:0005133 | ECTO:0070013 |
| 1 | MONDO:0005133 | MONDO:0004425 |
| 1 | MONDO:0005133 | ECTO:0070029 |
| 1 | MONDO:0005133 | ECTO:0070170 |
| 1 | MONDO:0005133 | ECTO:0070152 |
| 1 | MONDO:0005133 | ECTO:0070038 |
| 1 | MONDO:0005133 | ECTO:9002169 |
| 1 | MONDO:0005133 | ECTO:0070161 |
| 1 | MONDO:0005133 | ECTO:0070016 |
| 1 | MONDO:0005133 | ECTO:0070028 |
| 1 | MONDO:0005133 | ECTO:0070061 |
| 1 | MONDO:0005133 | ECTO:0070123 |
| 1 | MONDO:0005133 | ECTO:0070090 |
| 1 | MONDO:0005133 | ECTO:0070071 |
| 1 | MONDO:0005133 | ECTO:0070168 |
| 1 | MONDO:0005133 | ECTO:9000037 |
| 1 | MONDO:0005133 | ECTO:9000125 |
| 1 | MONDO:0005133 | MONDO:0006195 |
| 1 | MONDO:0005133 | MONDO:0005393 |
| 1 | MONDO:0005133 | MONDO:0002715 |
| 1 | MONDO:0005133 | MONDO:0004375 |
| 1 | MONDO:0005133 | MONDO:0010030 |
| 1 | MONDO:0005133 | ECTO:9000083 |
| 1 | MONDO:0005133 | CHEBI:6993 |
| 1 | MONDO:0005133 | ECTO:0001566 |
| 1 | MONDO:0005133 | ECTO:9000104 |
| 1 | MONDO:0005133 | ECTO:0000515 |
| 1 | MONDO:0005133 | ECTO:7000129 |
| 1 | MONDO:0005133 | ECTO:7000132 |
| 1 | MONDO:0005133 | ECTO:0500004 |
| 1 | MONDO:0005133 | ECTO:7000155 |
| 1 | MONDO:0005133 | ECTO:9000036 |
| 1 | MONDO:0005133 | ECTO:9000941 |
| 1 | MONDO:0005133 | CHEBI:10650 |
| 1 | MONDO:0005133 | ECTO:9000085 |
| 1 | MONDO:0005133 | ECTO:9000039 |
| 1 | MONDO:0005133 | ECTO:9000016 |
| 1 | MONDO:0005133 | CHEBI:7769 |
| 1 | MONDO:0005133 | CHEBI:6904 |
| 1 | MONDO:0005133 | CHEBI:6801 |
| 1 | MONDO:0005133 | CHEBI:3756 |
| 1 | MONDO:0005133 | CHEBI:6538 |
| 1 | MONDO:0005133 | CHEBI:8777 |
| 1 | MONDO:0005133 | CHEBI:50270 |
| 1 | MONDO:0005133 | CHEBI:6367 |
| 1 | MONDO:0005133 | CHEBI:43755 |
| 1 | MONDO:0005133 | GO:0042697 |
| 1 | MONDO:0005133 | HP:0100279 |
| 1 | MONDO:0005133 | HP:0002140 |
| 1 | MONDO:0005133 | HP:0002076 |
| 1 | MONDO:0005133 | HP:0200063 |
| 1 | MONDO:0005133 | HP:0002592 |
| 1 | MONDO:0005133 | HP:0000964 |
| 1 | MONDO:0005133 | HP:0000938 |
| 1 | MONDO:0005133 | CHEBI:5778 |
| 1 | MONDO:0005133 | HP:0033167 |
| 1 | MONDO:0005133 | HP:0008251 |
| 1 | MONDO:0005133 | ECTO:0070131 |
| 1 | MONDO:0005133 | ECTO:0070172 |
| 1 | MONDO:0005133 | ECTO:0070153 |
| 1 | MONDO:0005133 | ECTO:0070104 |
| 1 | MONDO:0005133 | ECTO:0070084 |
| 1 | MONDO:0005133 | ECTO:0070020 |
| 1 | MONDO:0005133 | ECTO:0070010 |
| 1 | MONDO:0005133 | ECTO:9000954 |
| 1 | MONDO:0005133 | ECTO:9000133 |
| 1 | MONDO:0005133 | ECTO:0070124 |
| 1 | MONDO:0005133 | ECTO:0070044 |
| 1 | MONDO:0005133 | ECTO:0070027 |
| 1 | MONDO:0005133 | ECTO:0070114 |
| 1 | MONDO:0005133 | MONDO:0008170 |
| 1 | MONDO:0005133 | MONDO:0005010 |
| 1 | MONDO:0005133 | MONDO:0024664 |
| 1 | MONDO:0005133 | ECTO:0070130 |
| 1 | MONDO:0005133 | ECTO:0070060 |
| 1 | MONDO:0005133 | ECTO:0070070 |
| 1 | MONDO:0005133 | ECTO:9002147 |
| 1 | MONDO:0005133 | ECTO:9000945 |
| 1 | MONDO:0005133 | ECTO:7000146 |
| 1 | MONDO:0005133 | ECTO:9000082 |
| 1 | MONDO:0005133 | ECTO:9001538 |
| 1 | MONDO:0005133 | CHEBI:36796 |
| 1 | MONDO:0005133 | ECTO:2000005 |
| 1 | MONDO:0005133 | HP:0005994 |
| 1 | MONDO:0005133 | CHEBI:3562 |
| 1 | MONDO:0005133 | HP:0000147 |
| 1 | MONDO:0005133 | HP:0012886 |
| 1 | MONDO:0005133 | CHEBI:6541 |
| 1 | MONDO:0005133 | CHEBI:5118 |
| 1 | MONDO:0005133 | CHEBI:101278 |
| 1 | MONDO:0005133 | HP:0001891 |
| 1 | MONDO:0005133 | HP:0001635 |
| 1 | MONDO:0005133 | HP:0000131 |
| 1 | MONDO:0005133 | MAXO:0001058 |
| 1 | MONDO:0005133 | ECTO:9000092 |
| 1 | MONDO:0005133 | ECTO:7000142 |
| 1 | MONDO:0005133 | HP:0001370 |
| 1 | MONDO:0005133 | CHEBI:7772 |
| 1 | MONDO:0005133 | ECTO:0070037 |
| 1 | MONDO:0005133 | ECTO:0070174 |
| 1 | MONDO:0005133 | ECTO:0070022 |
| 1 | MONDO:0005133 | ECTO:0070159 |
| 1 | MONDO:0005133 | ECTO:0070138 |
| 1 | MONDO:0005133 | ECTO:0070036 |
| 1 | MONDO:0005133 | ECTO:0070128 |
| 1 | MONDO:0005133 | ECTO:0070155 |
| 1 | MONDO:0005133 | ECTO:0070003 |
| 1 | MONDO:0005133 | ECTO:0070009 |
| 1 | MONDO:0005133 | ECTO:0070158 |
| 1 | MONDO:0005133 | ECTO:0070121 |
| 1 | MONDO:0005133 | ECTO:0070187 |
| 1 | MONDO:0005133 | ECTO:0070012 |
| 1 | MONDO:0005133 | ECTO:9000123 |
| 1 | MONDO:0005133 | ECTO:9000299 |
| 1 | MONDO:0005133 | ECTO:0070210 |
| 1 | MONDO:0005133 | MONDO:0011382 |
| 1 | MONDO:0005133 | MONDO:0008364 |
| 1 | MONDO:0005133 | MONDO:0020642 |
| 1 | MONDO:0005133 | MONDO:0018076 |
| 1 | MONDO:0005133 | HP:0004936 |
| 1 | MONDO:0005133 | ECTO:9000424 |
| 1 | MONDO:0005133 | ECTO:9000051 |
| 1 | MONDO:0005133 | ECTO:0000001 |
| 1 | MONDO:0005133 | ECTO:9000025 |
| 1 | MONDO:0005133 | ECTO:9000028 |
| 1 | MONDO:0005133 | CHEBI:3219 |
| 1 | MONDO:0005133 | CHEBI:5051 |
| 1 | MONDO:0005133 | CHEBI:36791 |
| 1 | MONDO:0005133 | HP:0100280 |
| 1 | MONDO:0005133 | HP:0004789 |
| 1 | MONDO:0005133 | HP:0001297 |
| 1 | MONDO:0005133 | HP:0000822 |
| 1 | MONDO:0005133 | ECTO:0000530 |
| 1 | MONDO:0005133 | ECTO:9000017 |
| 1 | MONDO:0005133 | CHEBI:5855 |
| 1 | MONDO:0005133 | MAXO:0001067 |
| 1 | MONDO:0005133 | HP:0000787 |
| 1 | MONDO:0005133 | HP:0001025 |
| 1 | MONDO:0005133 | HP:0000939 |
| 1 | MONDO:0005133 | HP:0000853 |
| 1 | MONDO:0005133 | HP:0001681 |
| 1 | MONDO:0005133 | ECTO:9000068 |
| 1 | MONDO:0005133 | ECTO:0070188 |
| 1 | MONDO:0005133 | ECTO:0070133 |
| 1 | MONDO:0005133 | ECTO:0070165 |
| 1 | MONDO:0005133 | ECTO:0070035 |
| 1 | MONDO:0005133 | ECTO:0070183 |
| 1 | MONDO:0005133 | ECTO:0070047 |
| 1 | MONDO:0005133 | ECTO:0070110 |
| 1 | MONDO:0005133 | ECTO:0070021 |
| 1 | MONDO:0005133 | ECTO:0070171 |
| 1 | MONDO:0005133 | ECTO:0070024 |
| 1 | MONDO:0005133 | ECTO:0070167 |
| 1 | MONDO:0005133 | MONDO:0005420 |
| 1 | MONDO:0005133 | MONDO:0005015 |
| 1 | MONDO:0005133 | MONDO:0004979 |
| 1 | MONDO:0005133 | MONDO:0005027 |
| 1 | MONDO:0005133 | MONDO:0008345 |
| 1 | MONDO:0005133 | ECTO:0070075 |
| 1 | MONDO:0005133 | ECTO:0070015 |
| 1 | MONDO:0005133 | ECTO:0070002 |
| 1 | MONDO:0005133 | ECTO:9000210 |
| 1 | MONDO:0005133 | ECTO:9000243 |
| 1 | MONDO:0005133 | CHEBI:6539 |
| 1 | MONDO:0005133 | CHEBI:6741 |
| 1 | MONDO:0005133 | MAXO:0000257 |
| 1 | MONDO:0005133 | HP:0012330 |
| 1 | MONDO:0005133 | HP:0002621 |
| 1 | MONDO:0005133 | HP:0012622 |
| 1 | MONDO:0005133 | HP:0002608 |
| 1 | MONDO:0005133 | CHEBI:6446 |
| 1 | MONDO:0005133 | CHEBI:6484 |
| 1 | MONDO:0005133 | ECTO:7000154 |
| 1 | MONDO:0005133 | ECTO:9000060 |
| 1 | MONDO:0005133 | ECTO:9002143 |
| 1 | MONDO:0005133 | CHEBI:63631 |
| 1 | MONDO:0005133 | ECTO:9000099 |
| 1 | MONDO:0005133 | ECTO:9000022 |
| 1 | MONDO:0005133 | ECTO:0500010 |
| 1 | MONDO:0005133 | ECTO:9000071 |
| 1 | MONDO:0005133 | ECTO:9002142 |
| 1 | MONDO:0005133 | ECTO:9000059 |
| 1 | MONDO:0005133 | ECTO:7000141 |
| 1 | MONDO:0005133 | ECTO:7000131 |
| 1 | MONDO:0005133 | ECTO:9000090 |
| 1 | MONDO:0005133 | MONDO:0006920 |
| 1 | MONDO:0005133 | MONDO:0004790 |
| 1 | MONDO:0005133 | MONDO:0005002 |
| 1 | MONDO:0005133 | CHEBI:42797 |
| 1 | MONDO:0005133 | MONDO:0005546 |
| 1 | MONDO:0005133 | ECTO:0070046 |
| 1 | MONDO:0005133 | ECTO:0070042 |
| 1 | MONDO:0005133 | ECTO:0070056 |
| 1 | MONDO:0005133 | ECTO:9000229 |
| 1 | MONDO:0005133 | ECTO:9000253 |
| 1 | MONDO:0005133 | ECTO:0070004 |
| 1 | MONDO:0005133 | ECTO:0070017 |
| 1 | MONDO:0005133 | ECTO:0070185 |
| 1 | MONDO:0005133 | ECTO:0070209 |
| 1 | MONDO:0005133 | ECTO:0070139 |
| 1 | MONDO:0005133 | ECTO:0070169 |
| 1 | MONDO:0005133 | ECTO:0070039 |
| 1 | MONDO:0005133 | ECTO:0070112 |
| 1 | MONDO:0005133 | ECTO:0070067 |
| 1 | MONDO:0005133 | ECTO:0070025 |
| 1 | MONDO:0005133 | ECTO:0070108 |
| 1 | MONDO:0005133 | CHEBI:7944 |
| 1 | MONDO:0005133 | HP:0000138 |
| 1 | MONDO:0005133 | HP:0002758 |
| 1 | MONDO:0005133 | HP:0002326 |
| 1 | MONDO:0005133 | ECTO:9001831 |
| 1 | MONDO:0005133 | CHEBI:50275 |
| 1 | MONDO:0005133 | HP:0001394 |
| 1 | MONDO:0005133 | HP:0100614 |
| 1 | MONDO:0005133 | HP:0011675 |
| 1 | MONDO:0005133 | HP:0001658 |
| 1 | MONDO:0005133 | CHEBI:50309 |
| 1 | MONDO:0005133 | HP:0100324 |
| 1 | MONDO:0005133 | MAXO:0000259 |
| 1 | MONDO:0005133 | CHEBI:10126 |
| 1 | MONDO:0005133 | CHEBI:39548 |
| 1 | MONDO:0005133 | CHEBI:7476 |
| 1 | MONDO:0005133 | ECTO:0070132 |
| 1 | MONDO:0005133 | ECTO:0070119 |
| 1 | MONDO:0005133 | ECTO:0070048 |
| 1 | MONDO:0005133 | ECTO:0070000 |
| 1 | MONDO:0005133 | ECTO:0070151 |
| 1 | MONDO:0005133 | ECTO:0070189 |
| 1 | MONDO:0005133 | ECTO:0070078 |
| 1 | MONDO:0005133 | ECTO:0070018 |
| 1 | MONDO:0005133 | ECTO:0070007 |
| 1 | MONDO:0005133 | ECTO:9000192 |
| 1 | MONDO:0005133 | ECTO:9000143 |
| 1 | MONDO:0005133 | ECTO:0070026 |
| 1 | MONDO:0005133 | ECTO:0070109 |
| 1 | MONDO:0005133 | ECTO:0070062 |
| 1 | MONDO:0005133 | ECTO:0070157 |
| 1 | MONDO:0005133 | CHEBI:77249 |
| 1 | MONDO:0005133 | MONDO:0005406 |
| 1 | MONDO:0005133 | MONDO:0008228 |
| 1 | MONDO:0005133 | MONDO:0007915 |
| 1 | MONDO:0005133 | ECTO:0070125 |
| 1 | MONDO:0005133 | ECTO:9000069 |
| 1 | MONDO:0005133 | ECTO:9000043 |
| 1 | MONDO:0005133 | ECTO:9000058 |
| 1 | MONDO:0005133 | ECTO:7000140 |
| 1 | MONDO:0005133 | ECTO:7000005 |
| 1 | MONDO:0005133 | ECTO:9000081 |
| 1 | MONDO:0005133 | ECTO:9001302 |
| 1 | MONDO:0005133 | ECTO:7000017 |
| 1 | MONDO:0005133 | ECTO:0500006 |
| 1 | MONDO:0005133 | ECTO:9001437 |
| 1 | MONDO:0005133 | ECTO:9002166 |
| 1 | MONDO:0005133 | ECTO:9000048 |
| 1 | MONDO:0005133 | ECTO:9000076 |
| 1 | MONDO:0005133 | ECTO:9000020 |
| 1 | MONDO:0005133 | ECTO:7000153 |
| 1 | MONDO:0005133 | ECTO:0001571 |
| 1 | MONDO:0005133 | ECTO:9000079 |
| 1 | MONDO:0005133 | ECTO:7000151 |
| 1 | MONDO:0005133 | ECTO:9002165 |
| 1 | MONDO:0005133 | ECTO:9000032 |
| 1 | MONDO:0005133 | ECTO:7000152 |
| 1 | MONDO:0005133 | ECTO:9000042 |
| 1 | MONDO:0005133 | ECTO:9000019 |
| 1 | MONDO:0005133 | ECTO:0500009 |
| 1 | MONDO:0005133 | ECTO:9000024 |
| 1 | MONDO:0005133 | ECTO:0500021 |
| 1 | MONDO:0005133 | ECTO:9000057 |
| 1 | MONDO:0005133 | ECTO:7000150 |
| 1 | MONDO:0005133 | ECTO:9001465 |
| 1 | MONDO:0005133 | ECTO:9000027 |
| 1 | MONDO:0005133 | ECTO:9001450 |
| 1 | MONDO:0005133 | ECTO:9000034 |
| 1 | MONDO:0005133 | MONDO:0006507 |
| 1 | MONDO:0005133 | ECTO:0070164 |
| 1 | MONDO:0005133 | ECTO:9000224 |
| 1 | MONDO:0005133 | ECTO:0070011 |
| 1 | MONDO:0005133 | ECTO:0070023 |
| 1 | MONDO:0005133 | MONDO:0006107 |
| 1 | MONDO:0005133 | MONDO:0005281 |
| 1 | MONDO:0005133 | MONDO:0005083 |
| 1 | MONDO:0005133 | ECTO:0070173 |
| 1 | MONDO:0005133 | ECTO:0070156 |
| 1 | MONDO:0005133 | ECTO:0070148 |
| 1 | MONDO:0005133 | ECTO:0070116 |
| 1 | MONDO:0005133 | ECTO:0070182 |
| 1 | MONDO:0005133 | ECTO:0070045 |
| 1 | MONDO:0005133 | ECTO:0070134 |
| 1 | MONDO:0005133 | ECTO:0070162 |
| 1 | MONDO:0005133 | ECTO:0070008 |
| 1 | MONDO:0005133 | ECTO:0070154 |
| 1 | MONDO:0005133 | ECTO:0070107 |
| 1 | MONDO:0005133 | ECTO:0070085 |
| 1 | MONDO:0005133 | ECTO:8000046 |
| 1 | MONDO:0005133 | ECTO:9000070 |
| 1 | MONDO:0005133 | ECTO:9002113 |
| 1 | MONDO:0005133 | ECTO:9002141 |
| 1 | MONDO:0005133 | CHEBI:50686 |
| 1 | MONDO:0005133 | ECTO:0500007 |
| 1 | MONDO:0005133 | ECTO:0500022 |
| 1 | MONDO:0005133 | ECTO:9000089 |
| 1 | MONDO:0005133 | ECTO:7000028 |
| 1 | MONDO:0005133 | ECTO:9000023 |
| 1 | MONDO:0005133 | ECTO:7000149 |
| 1 | MONDO:0005133 | ECTO:9000049 |
| 1 | MONDO:0005133 | CHEBI:135931 |
| 1 | MONDO:0005133 | CHEBI:9124 |
| 1 | MONDO:0005133 | CHEBI:135925 |
| 1 | MONDO:0005133 | HP:0025081 |
| 1 | MONDO:0005133 | HP:0009798 |
| 1 | MONDO:0005133 | HP:0002077 |
| 1 | MONDO:0005133 | HP:0002640 |
| 1 | MONDO:0005133 | HP:0008843 |
| 1 | MONDO:0005133 | CHEBI:9648 |
| 1 | MONDO:0005133 | HP:0032282 |
| 1 | MONDO:0005133 | HP:0030425 |
| 1 | MONDO:0005133 | CHEBI:3654 |
| 1 | MONDO:0005133 | CHEBI:3723 |
| 1 | MONDO:0005133 | HP:0012623 |
| 1 | MONDO:0005133 | CHEBI:2668 |
| 1 | MONDO:0005133 | HP:0002083 |
| 1 | MONDO:0005133 | CHEBI:9584 |
| 1 | MONDO:0005133 | HP:0033761 |
| 1 | MONDO:0005133 | CHEBI:31836 |
| 1 | MONDO:0005133 | CHEBI:2611 |
| 1 | MONDO:0005133 | CHEBI:9123 |
| 1 | MONDO:0005133 | CHEBI:2550 |
| 1 | MONDO:0005133 | HP:0001051 |
| 1 | MONDO:0005133 | HP:0001962 |
| 1 | MONDO:0005133 | CHEBI:9150 |
| 1 | MONDO:0005133 | CHEBI:9654 |
| 1 | MONDO:0005133 | CHEBI:3724 |
| 1 | MONDO:0005133 | CHEBI:9435 |
| 1 | MONDO:0005133 | CHEBI:31526 |
| 1 | MONDO:0005133 | CHEBI:47426 |
| 1 | MONDO:0005133 | CHEBI:15365 |
| 1 | HP:0000138 | CHEBI:115822 |
| 1 | HP:0000138 | CHEBI:115264 |
| 1 | HP:0000138 | ECTO:0500023 |
| 1 | HP:0000138 | ECTO:0000207 |
| 1 | HP:0000138 | ECTO:9000085 |
| 1 | HP:0000138 | ECTO:9000039 |
| 1 | HP:0000138 | ECTO:9000016 |
| 1 | HP:0000138 | CHEBI:116071 |
| 1 | HP:0000138 | CHEBI:97038 |
| 1 | HP:0000138 | CHEBI:103162 |
| 1 | HP:0000138 | CHEBI:117714 |
| 1 | HP:0000138 | CHEBI:95556 |
| 1 | HP:0000138 | CHEBI:103445 |
| 1 | HP:0000138 | CHEBI:7769 |
| 1 | HP:0000138 | CHEBI:115800 |
| 1 | HP:0000138 | CHEBI:98727 |
| 1 | HP:0000138 | CHEBI:95428 |
| 1 | HP:0000138 | CHEBI:98794 |
| 1 | HP:0000138 | CHEBI:98735 |
| 1 | HP:0000138 | CHEBI:115942 |
| 1 | HP:0000138 | ECTO:9000103 |
| 1 | HP:0000138 | ECTO:7000030 |
| 1 | HP:0000138 | CHEBI:96992 |
| 1 | HP:0000138 | CHEBI:117800 |
| 1 | HP:0000138 | ECTO:9000018 |
| 1 | HP:0000138 | ECTO:9000031 |
| 1 | HP:0000138 | ECTO:9000074 |
| 1 | HP:0000138 | ECTO:7000144 |
| 1 | HP:0000138 | ECTO:9000038 |
| 1 | HP:0000138 | ECTO:7000143 |
| 1 | HP:0000138 | ECTO:9000052 |
| 1 | HP:0000138 | ECTO:0500008 |
| 1 | HP:0000138 | ECTO:0500017 |
| 1 | HP:0000138 | ECTO:0500016 |
| 1 | HP:0000138 | ECTO:9002164 |
| 1 | HP:0000138 | ECTO:9000063 |
| 1 | HP:0000138 | MONDO:0002251 |
| 1 | HP:0000138 | MONDO:0002974 |
| 1 | HP:0000138 | ECTO:0070043 |
| 1 | HP:0000138 | ECTO:0070069 |
| 1 | HP:0000138 | ECTO:0070166 |
| 1 | HP:0000138 | ECTO:0070135 |
| 1 | HP:0000138 | ECTO:0070137 |
| 1 | HP:0000138 | ECTO:0070184 |
| 1 | HP:0000138 | ECTO:0070163 |
| 1 | HP:0000138 | ECTO:0070129 |
| 1 | HP:0000138 | ECTO:9000087 |
| 1 | HP:0000138 | ECTO:9002151 |
| 1 | HP:0000138 | CHEBI:100647 |
| 1 | HP:0000138 | CHEBI:96310 |
| 1 | HP:0000138 | CHEBI:129605 |
| 1 | HP:0000138 | CHEBI:6904 |
| 1 | HP:0000138 | MONDO:0004375 |
| 1 | HP:0000138 | MONDO:0010030 |
| 1 | HP:0000138 | MONDO:0005393 |
| 1 | HP:0000138 | MONDO:0006195 |
| 1 | HP:0000138 | MONDO:0004425 |
| 1 | HP:0000138 | ECTO:0070029 |
| 1 | HP:0000138 | ECTO:0070170 |
| 1 | HP:0000138 | ECTO:0070152 |
| 1 | HP:0000138 | ECTO:0070038 |
| 1 | HP:0000138 | ECTO:9002169 |
| 1 | HP:0000138 | ECTO:0070123 |
| 1 | HP:0000138 | ECTO:0070090 |
| 1 | HP:0000138 | ECTO:0070071 |
| 1 | HP:0000138 | CHEBI:97933 |
| 1 | HP:0000138 | ECTO:0070168 |
| 1 | HP:0000138 | ECTO:9000037 |
| 1 | HP:0000138 | ECTO:9000125 |
| 1 | HP:0000138 | CHEBI:10125 |
| 1 | HP:0000138 | MONDO:0002715 |
| 1 | HP:0000138 | CHEBI:104743 |
| 1 | HP:0000138 | ECTO:9000941 |
| 1 | HP:0000138 | CHEBI:97612 |
| 1 | HP:0000138 | CHEBI:104332 |
| 1 | HP:0000138 | CHEBI:3804 |
| 1 | HP:0000138 | HP:0100279 |
| 1 | HP:0000138 | HP:0002140 |
| 1 | HP:0000138 | CHEBI:100696 |
| 1 | HP:0000138 | CHEBI:122563 |
| 1 | HP:0000138 | CHEBI:104776 |
| 1 | HP:0000138 | CHEBI:98553 |
| 1 | HP:0000138 | CHEBI:96565 |
| 1 | HP:0000138 | CHEBI:96920 |
| 1 | HP:0000138 | CHEBI:104330 |
| 1 | HP:0000138 | CHEBI:104505 |
| 1 | HP:0000138 | CHEBI:97886 |
| 1 | HP:0000138 | CHEBI:43755 |
| 1 | HP:0000138 | CHEBI:104612 |
| 1 | HP:0000138 | CHEBI:115801 |
| 1 | HP:0000138 | CHEBI:97358 |
| 1 | HP:0000138 | CHEBI:104684 |
| 1 | HP:0000138 | CHEBI:101210 |
| 1 | HP:0000138 | CHEBI:104674 |
| 1 | HP:0000138 | CHEBI:98362 |
| 1 | HP:0000138 | CHEBI:101472 |
| 1 | HP:0000138 | CHEBI:95933 |
| 1 | HP:0000138 | CHEBI:115900 |
| 1 | HP:0000138 | CHEBI:129595 |
| 1 | HP:0000138 | CHEBI:104735 |
| 1 | HP:0000138 | CHEBI:115939 |
| 1 | HP:0000138 | CHEBI:122807 |
| 1 | HP:0000138 | CHEBI:8777 |
| 1 | HP:0000138 | CHEBI:117642 |
| 1 | HP:0000138 | CHEBI:117646 |
| 1 | HP:0000138 | CHEBI:6801 |
| 1 | HP:0000138 | CHEBI:123776 |
| 1 | HP:0000138 | CHEBI:97776 |
| 1 | HP:0000138 | CHEBI:115759 |
| 1 | HP:0000138 | CHEBI:6538 |
| 1 | HP:0000138 | CHEBI:97541 |
| 1 | HP:0000138 | CHEBI:96831 |
| 1 | HP:0000138 | CHEBI:96778 |
| 1 | HP:0000138 | CHEBI:115445 |
| 1 | HP:0000138 | CHEBI:115435 |
| 1 | HP:0000138 | CHEBI:123849 |
| 1 | HP:0000138 | CHEBI:122605 |
| 1 | HP:0000138 | CHEBI:97361 |
| 1 | HP:0000138 | CHEBI:103578 |
| 1 | HP:0000138 | CHEBI:129871 |
| 1 | HP:0000138 | CHEBI:115214 |
| 1 | HP:0000138 | CHEBI:122537 |
| 1 | HP:0000138 | CHEBI:124084 |
| 1 | HP:0000138 | CHEBI:104579 |
| 1 | HP:0000138 | CHEBI:104364 |
| 1 | HP:0000138 | CHEBI:96046 |
| 1 | HP:0000138 | CHEBI:50270 |
| 1 | HP:0000138 | CHEBI:115362 |
| 1 | HP:0000138 | CHEBI:115304 |
| 1 | HP:0000138 | CHEBI:115876 |
| 1 | HP:0000138 | CHEBI:124067 |
| 1 | HP:0000138 | CHEBI:104499 |
| 1 | HP:0000138 | CHEBI:124023 |
| 1 | HP:0000138 | CHEBI:104633 |
| 1 | HP:0000138 | CHEBI:104638 |
| 1 | HP:0000138 | CHEBI:10650 |
| 1 | HP:0000138 | ECTO:0070060 |
| 1 | HP:0000138 | ECTO:0070070 |
| 1 | HP:0000138 | ECTO:0070114 |
| 1 | HP:0000138 | ECTO:0070027 |
| 1 | HP:0000138 | ECTO:0070124 |
| 1 | HP:0000138 | ECTO:0070131 |
| 1 | HP:0000138 | ECTO:0070172 |
| 1 | HP:0000138 | ECTO:0070104 |
| 1 | HP:0000138 | ECTO:0070010 |
| 1 | HP:0000138 | ECTO:9000954 |
| 1 | HP:0000138 | CHEBI:102532 |
| 1 | HP:0000138 | CHEBI:5778 |
| 1 | HP:0000138 | ECTO:0070130 |
| 1 | HP:0000138 | MONDO:0024664 |
| 1 | HP:0000138 | MONDO:0005010 |
| 1 | HP:0000138 | HP:0008251 |
| 1 | HP:0000138 | CHEBI:96289 |
| 1 | HP:0000138 | MONDO:0008170 |
| 1 | HP:0000138 | CHEBI:96396 |
| 1 | HP:0000138 | ECTO:9002147 |
| 1 | HP:0000138 | ECTO:9001538 |
| 1 | HP:0000138 | CHEBI:6375 |
| 1 | HP:0000138 | CHEBI:100512 |
| 1 | HP:0000138 | CHEBI:36796 |
| 1 | HP:0000138 | HP:0001635 |
| 1 | HP:0000138 | HP:0000791 |
| 1 | HP:0000138 | CHEBI:127722 |
| 1 | HP:0000138 | CHEBI:95591 |
| 1 | HP:0000138 | CHEBI:128568 |
| 1 | HP:0000138 | HP:0008675 |
| 1 | HP:0000138 | CHEBI:5118 |
| 1 | HP:0000138 | CHEBI:113461 |
| 1 | HP:0000138 | CHEBI:100572 |
| 1 | HP:0000138 | HP:0003940 |
| 1 | HP:0000138 | HP:0001047 |
| 1 | HP:0000138 | HP:0001931 |
| 1 | HP:0000138 | CHEBI:113275 |
| 1 | HP:0000138 | HP:0001413 |
| 1 | HP:0000138 | CHEBI:102270 |
| 1 | HP:0000138 | HP:0004699 |
| 1 | HP:0000138 | CHEBI:113281 |
| 1 | HP:0000138 | HP:0030035 |
| 1 | HP:0000138 | CHEBI:127882 |
| 1 | HP:0000138 | HP:0004398 |
| 1 | HP:0000138 | CHEBI:116002 |
| 1 | HP:0000138 | HP:0033762 |
| 1 | HP:0000138 | HP:0100735 |
| 1 | HP:0000138 | CHEBI:95405 |
| 1 | HP:0000138 | CHEBI:3562 |
| 1 | HP:0000138 | CHEBI:95802 |
| 1 | HP:0000138 | HP:0000804 |
| 1 | HP:0000138 | HP:0005086 |
| 1 | HP:0000138 | CHEBI:127127 |
| 1 | HP:0000138 | CHEBI:116030 |
| 1 | HP:0000138 | CHEBI:95531 |
| 1 | HP:0000138 | HP:0030255 |
| 1 | HP:0000138 | CHEBI:102168 |
| 1 | HP:0000138 | CHEBI:94851 |
| 1 | HP:0000138 | HP:0031011 |
| 1 | HP:0000138 | HP:0031012 |
| 1 | HP:0000138 | HP:0011127 |
| 1 | HP:0000138 | HP:0003774 |
| 1 | HP:0000138 | CHEBI:112616 |
| 1 | HP:0000138 | CHEBI:107011 |
| 1 | HP:0000138 | HP:0003088 |
| 1 | HP:0000138 | HP:0011005 |
| 1 | HP:0000138 | HP:0005994 |
| 1 | HP:0000138 | HP:0033167 |
| 1 | HP:0000138 | HP:0033564 |
| 1 | HP:0000138 | CHEBI:101353 |
| 1 | HP:0000138 | CHEBI:115346 |
| 1 | HP:0000138 | CHEBI:7772 |
| 1 | HP:0000138 | CHEBI:96816 |
| 1 | HP:0000138 | ECTO:9000299 |
| 1 | HP:0000138 | MONDO:0020642 |
| 1 | HP:0000138 | MONDO:0018076 |
| 1 | HP:0000138 | MONDO:0008364 |
| 1 | HP:0000138 | MONDO:0011382 |
| 1 | HP:0000138 | ECTO:0070174 |
| 1 | HP:0000138 | ECTO:0070022 |
| 1 | HP:0000138 | ECTO:0070159 |
| 1 | HP:0000138 | ECTO:0070138 |
| 1 | HP:0000138 | ECTO:0070036 |
| 1 | HP:0000138 | ECTO:0070128 |
| 1 | HP:0000138 | ECTO:0070158 |
| 1 | HP:0000138 | ECTO:0070121 |
| 1 | HP:0000138 | ECTO:0070187 |
| 1 | HP:0000138 | ECTO:9000123 |
| 1 | HP:0000138 | HP:0004936 |
| 1 | HP:0000138 | HP:0001370 |
| 1 | HP:0000138 | HP:0002592 |
| 1 | HP:0000138 | CHEBI:102802 |
| 1 | HP:0000138 | CHEBI:123856 |
| 1 | HP:0000138 | CHEBI:123827 |
| 1 | HP:0000138 | CHEBI:101728 |
| 1 | HP:0000138 | CHEBI:5551 |
| 1 | HP:0000138 | HP:0001297 |
| 1 | HP:0000138 | HP:0004789 |
| 1 | HP:0000138 | HP:0100280 |
| 1 | HP:0000138 | CHEBI:96754 |
| 1 | HP:0000138 | CHEBI:96699 |
| 1 | HP:0000138 | CHEBI:116050 |
| 1 | HP:0000138 | CHEBI:94772 |
| 1 | HP:0000138 | CHEBI:97223 |
| 1 | HP:0000138 | CHEBI:96955 |
| 1 | HP:0000138 | CHEBI:97203 |
| 1 | HP:0000138 | CHEBI:5051 |
| 1 | HP:0000138 | CHEBI:36791 |
| 1 | HP:0000138 | CHEBI:101206 |
| 1 | HP:0000138 | ECTO:9000051 |
| 1 | HP:0000138 | ECTO:9000945 |
| 1 | HP:0000138 | ECTO:7000146 |
| 1 | HP:0000138 | ECTO:9000082 |
| 1 | HP:0000138 | ECTO:9000092 |
| 1 | HP:0000138 | ECTO:7000142 |
| 1 | HP:0000138 | ECTO:9000424 |
| 1 | HP:0000138 | CHEBI:115816 |
| 1 | HP:0000138 | ECTO:0000001 |
| 1 | HP:0000138 | ECTO:9000025 |
| 1 | HP:0000138 | ECTO:9000028 |
| 1 | HP:0000138 | CHEBI:96568 |
| 1 | HP:0000138 | CHEBI:115262 |
| 1 | HP:0000138 | CHEBI:129371 |
| 1 | HP:0000138 | CHEBI:115770 |
| 1 | HP:0000138 | CHEBI:101157 |
| 1 | HP:0000138 | CHEBI:117762 |
| 1 | HP:0000138 | CHEBI:102857 |
| 1 | HP:0000138 | CHEBI:102849 |
| 1 | HP:0000138 | CHEBI:3219 |
| 1 | HP:0000138 | CHEBI:101535 |
| 1 | HP:0000138 | HP:0002401 |
| 1 | HP:0000138 | ECTO:9000068 |
| 1 | HP:0000138 | CHEBI:104812 |
| 1 | HP:0000138 | CHEBI:115919 |
| 1 | HP:0000138 | HP:0001681 |
| 1 | HP:0000138 | HP:0001025 |
| 1 | HP:0000138 | HP:0000939 |
| 1 | HP:0000138 | HP:0000853 |
| 1 | HP:0000138 | MONDO:0008345 |
| 1 | HP:0000138 | ECTO:0070021 |
| 1 | HP:0000138 | ECTO:9000243 |
| 1 | HP:0000138 | ECTO:9000210 |
| 1 | HP:0000138 | ECTO:0070075 |
| 1 | HP:0000138 | ECTO:0070133 |
| 1 | HP:0000138 | ECTO:0070165 |
| 1 | HP:0000138 | ECTO:0070035 |
| 1 | HP:0000138 | ECTO:0070183 |
| 1 | HP:0000138 | ECTO:0070047 |
| 1 | HP:0000138 | ECTO:0070171 |
| 1 | HP:0000138 | ECTO:0070024 |
| 1 | HP:0000138 | ECTO:0070167 |
| 1 | HP:0000138 | MONDO:0005420 |
| 1 | HP:0000138 | MONDO:0005027 |
| 1 | HP:0000138 | CHEBI:6539 |
| 1 | HP:0000138 | CHEBI:6741 |
| 1 | HP:0000138 | MAXO:0000257 |
| 1 | HP:0000138 | HP:0012330 |
| 1 | HP:0000138 | HP:0002621 |
| 1 | HP:0000138 | HP:0012622 |
| 1 | HP:0000138 | HP:0002608 |
| 1 | HP:0000138 | CHEBI:6446 |
| 1 | HP:0000138 | ECTO:9002143 |
| 1 | HP:0000138 | ECTO:0500010 |
| 1 | HP:0000138 | ECTO:9000071 |
| 1 | HP:0000138 | ECTO:9002142 |
| 1 | HP:0000138 | ECTO:9000059 |
| 1 | HP:0000138 | ECTO:7000141 |
| 1 | HP:0000138 | ECTO:7000131 |
| 1 | HP:0000138 | ECTO:9000090 |
| 1 | HP:0000138 | CHEBI:123758 |
| 1 | HP:0000138 | ECTO:9000099 |
| 1 | HP:0000138 | ECTO:7000154 |
| 1 | HP:0000138 | CHEBI:117602 |
| 1 | HP:0000138 | ECTO:9000060 |
| 1 | HP:0000138 | CHEBI:97043 |
| 1 | HP:0000138 | ECTO:9000022 |
| 1 | HP:0000138 | CHEBI:129107 |
| 1 | HP:0000138 | ECTO:9000229 |
| 1 | HP:0000138 | ECTO:9000253 |
| 1 | HP:0000138 | CHEBI:42797 |
| 1 | HP:0000138 | ECTO:0070139 |
| 1 | HP:0000138 | ECTO:0070169 |
| 1 | HP:0000138 | ECTO:0070039 |
| 1 | HP:0000138 | ECTO:0070025 |
| 1 | HP:0000138 | ECTO:0070108 |
| 1 | HP:0000138 | ECTO:0070056 |
| 1 | HP:0000138 | ECTO:0070042 |
| 1 | HP:0000138 | ECTO:0070046 |
| 1 | HP:0000138 | MONDO:0005546 |
| 1 | HP:0000138 | MONDO:0004790 |
| 1 | HP:0000138 | MONDO:0005002 |
| 1 | HP:0000138 | CHEBI:104367 |
| 1 | HP:0000138 | CHEBI:115818 |
| 1 | HP:0000138 | HP:0002326 |
| 1 | HP:0000138 | CHEBI:103645 |
| 1 | HP:0000138 | CHEBI:122562 |
| 1 | HP:0000138 | MAXO:0000259 |
| 1 | HP:0000138 | HP:0100324 |
| 1 | HP:0000138 | HP:0001394 |
| 1 | HP:0000138 | HP:0100614 |
| 1 | HP:0000138 | HP:0011675 |
| 1 | HP:0000138 | HP:0001658 |
| 1 | HP:0000138 | CHEBI:97360 |
| 1 | HP:0000138 | CHEBI:117690 |
| 1 | HP:0000138 | ECTO:7000155 |
| 1 | HP:0000138 | ECTO:9000036 |
| 1 | HP:0000138 | ECTO:9000017 |
| 1 | HP:0000138 | CHEBI:104498 |
| 1 | HP:0000138 | CHEBI:6993 |
| 1 | HP:0000138 | CHEBI:8869 |
| 1 | HP:0000138 | CHEBI:96308 |
| 1 | HP:0000138 | CHEBI:104417 |
| 1 | HP:0000138 | CHEBI:104507 |
| 1 | HP:0000138 | CHEBI:117851 |
| 1 | HP:0000138 | CHEBI:104592 |
| 1 | HP:0000138 | CHEBI:96985 |
| 1 | HP:0000138 | CHEBI:95652 |
| 1 | HP:0000138 | CHEBI:5855 |
| 1 | HP:0000138 | CHEBI:103497 |
| 1 | HP:0000138 | CHEBI:103412 |
| 1 | HP:0000138 | CHEBI:97472 |
| 1 | HP:0000138 | CHEBI:50275 |
| 1 | HP:0000138 | CHEBI:123752 |
| 1 | HP:0000138 | CHEBI:96788 |
| 1 | HP:0000138 | CHEBI:96730 |
| 1 | HP:0000138 | CHEBI:96758 |
| 1 | HP:0000138 | CHEBI:123640 |
| 1 | HP:0000138 | CHEBI:104525 |
| 1 | HP:0000138 | CHEBI:123921 |
| 1 | HP:0000138 | CHEBI:104516 |
| 1 | HP:0000138 | CHEBI:103627 |
| 1 | HP:0000138 | CHEBI:123835 |
| 1 | HP:0000138 | CHEBI:104454 |
| 1 | HP:0000138 | ECTO:0500004 |
| 1 | HP:0000138 | ECTO:9000083 |
| 1 | HP:0000138 | ECTO:0001566 |
| 1 | HP:0000138 | ECTO:9000104 |
| 1 | HP:0000138 | ECTO:0000515 |
| 1 | HP:0000138 | ECTO:7000129 |
| 1 | HP:0000138 | ECTO:7000132 |
| 1 | HP:0000138 | CHEBI:104839 |
| 1 | HP:0000138 | CHEBI:98306 |
| 1 | HP:0000138 | CHEBI:104677 |
| 1 | HP:0000138 | CHEBI:129336 |
| 1 | HP:0000138 | CHEBI:50309 |
| 1 | HP:0000138 | CHEBI:122545 |
| 1 | HP:0000138 | CHEBI:97734 |
| 1 | HP:0000138 | CHEBI:103634 |
| 1 | HP:0000138 | CHEBI:97317 |
| 1 | HP:0000138 | CHEBI:10126 |
| 1 | HP:0000138 | CHEBI:97056 |
| 1 | HP:0000138 | CHEBI:103464 |
| 1 | HP:0000138 | CHEBI:103493 |
| 1 | HP:0000138 | MONDO:0007915 |
| 1 | HP:0000138 | MONDO:0005133 |
| 1 | HP:0000138 | ECTO:0070125 |
| 1 | HP:0000138 | ECTO:0070157 |
| 1 | HP:0000138 | ECTO:0070062 |
| 1 | HP:0000138 | ECTO:0070109 |
| 1 | HP:0000138 | ECTO:0070026 |
| 1 | HP:0000138 | ECTO:0070119 |
| 1 | HP:0000138 | ECTO:0070048 |
| 1 | HP:0000138 | ECTO:0070000 |
| 1 | HP:0000138 | ECTO:0070151 |
| 1 | HP:0000138 | ECTO:9000192 |
| 1 | HP:0000138 | ECTO:9000143 |
| 1 | HP:0000138 | CHEBI:123810 |
| 1 | HP:0000138 | CHEBI:103519 |
| 1 | HP:0000138 | CHEBI:117767 |
| 1 | HP:0000138 | CHEBI:103416 |
| 1 | HP:0000138 | CHEBI:103405 |
| 1 | HP:0000138 | CHEBI:103491 |
| 1 | HP:0000138 | MONDO:0008228 |
| 1 | HP:0000138 | CHEBI:98784 |
| 1 | HP:0000138 | MONDO:0005406 |
| 1 | HP:0000138 | CHEBI:117793 |
| 1 | HP:0000138 | CHEBI:97990 |
| 1 | HP:0000138 | ECTO:9000042 |
| 1 | HP:0000138 | ECTO:9000019 |
| 1 | HP:0000138 | ECTO:9000048 |
| 1 | HP:0000138 | ECTO:9002166 |
| 1 | HP:0000138 | ECTO:9001437 |
| 1 | HP:0000138 | ECTO:9000058 |
| 1 | HP:0000138 | ECTO:9000057 |
| 1 | HP:0000138 | ECTO:7000150 |
| 1 | HP:0000138 | ECTO:9001465 |
| 1 | HP:0000138 | ECTO:9000032 |
| 1 | HP:0000138 | ECTO:7000152 |
| 1 | HP:0000138 | ECTO:7000140 |
| 1 | HP:0000138 | ECTO:7000005 |
| 1 | HP:0000138 | ECTO:9000081 |
| 1 | HP:0000138 | ECTO:9000069 |
| 1 | HP:0000138 | ECTO:9001302 |
| 1 | HP:0000138 | ECTO:7000017 |
| 1 | HP:0000138 | ECTO:0500006 |
| 1 | HP:0000138 | ECTO:9000043 |
| 1 | HP:0000138 | ECTO:0500021 |
| 1 | HP:0000138 | CHEBI:117784 |
| 1 | HP:0000138 | ECTO:9000024 |
| 1 | HP:0000138 | CHEBI:117887 |
| 1 | HP:0000138 | CHEBI:117879 |
| 1 | HP:0000138 | ECTO:9000076 |
| 1 | HP:0000138 | ECTO:9000020 |
| 1 | HP:0000138 | ECTO:7000153 |
| 1 | HP:0000138 | ECTO:0001571 |
| 1 | HP:0000138 | ECTO:9000079 |
| 1 | HP:0000138 | ECTO:7000151 |
| 1 | HP:0000138 | ECTO:9002165 |
| 1 | HP:0000138 | ECTO:0500009 |
| 1 | HP:0000138 | CHEBI:97251 |
| 1 | HP:0000138 | CHEBI:117834 |
| 1 | HP:0000138 | CHEBI:96934 |
| 1 | HP:0000138 | CHEBI:96235 |
| 1 | HP:0000138 | MONDO:0006507 |
| 1 | HP:0000138 | ECTO:0070156 |
| 1 | HP:0000138 | MONDO:0006107 |
| 1 | HP:0000138 | CHEBI:103641 |
| 1 | HP:0000138 | MONDO:0005083 |
| 1 | HP:0000138 | ECTO:0070173 |
| 1 | HP:0000138 | ECTO:0070148 |
| 1 | HP:0000138 | ECTO:0070116 |
| 1 | HP:0000138 | ECTO:0070045 |
| 1 | HP:0000138 | ECTO:0070134 |
| 1 | HP:0000138 | ECTO:0070162 |
| 1 | HP:0000138 | ECTO:0070154 |
| 1 | HP:0000138 | ECTO:0070085 |
| 1 | HP:0000138 | ECTO:0070011 |
| 1 | HP:0000138 | ECTO:0070164 |
| 1 | HP:0000138 | ECTO:9000224 |
| 1 | HP:0000138 | ECTO:9002113 |
| 1 | HP:0000138 | CHEBI:124083 |
| 1 | HP:0000138 | CHEBI:98726 |
| 1 | HP:0000138 | ECTO:9000089 |
| 1 | HP:0000138 | ECTO:9002141 |
| 1 | HP:0000138 | ECTO:0500007 |
| 1 | HP:0000138 | ECTO:0500022 |
| 1 | HP:0000138 | CHEBI:98861 |
| 1 | HP:0000138 | ECTO:7000028 |
| 1 | HP:0000138 | ECTO:9000023 |
| 1 | HP:0000138 | ECTO:7000149 |
| 1 | HP:0000138 | CHEBI:50686 |
| 1 | HP:0000138 | ECTO:9000049 |
| 1 | HP:0000138 | ECTO:8000046 |
| 1 | HP:0000138 | ECTO:9000034 |
| 1 | HP:0000138 | ECTO:9001450 |
| 1 | HP:0000138 | ECTO:9000027 |
| 1 | HP:0000138 | ECTO:9000070 |
| 1 | HP:0000138 | CHEBI:117641 |
| 1 | HP:0000138 | CHEBI:117621 |
| 1 | HP:0000138 | CHEBI:135530 |
| 1 | HP:0000138 | CHEBI:110697 |
| 1 | HP:0000138 | CHEBI:126342 |
| 1 | HP:0000138 | CHEBI:95851 |
| 1 | HP:0000138 | HP:0009798 |
| 1 | HP:0000138 | CHEBI:106809 |
| 1 | HP:0000138 | CHEBI:110685 |
| 1 | HP:0000138 | HP:0009805 |
| 1 | HP:0000138 | CHEBI:130372 |
| 1 | HP:0000138 | CHEBI:99718 |
| 1 | HP:0000138 | CHEBI:110900 |
| 1 | HP:0000138 | CHEBI:9435 |
| 1 | HP:0000138 | HP:0001722 |
| 1 | HP:0000138 | CHEBI:111130 |
| 1 | HP:0000138 | HP:0004724 |
| 1 | HP:0000138 | CHEBI:94889 |
| 1 | HP:0000138 | CHEBI:99284 |
| 1 | HP:0000138 | CHEBI:106465 |
| 1 | HP:0000138 | HP:0033120 |
| 1 | HP:0000138 | HP:0012626 |
| 1 | HP:0000138 | CHEBI:99296 |
| 1 | HP:0000138 | CHEBI:128223 |
| 1 | HP:0000138 | CHEBI:111076 |
| 1 | HP:0000138 | HP:0004349 |
| 1 | HP:0000138 | HP:0025081 |
| 1 | HP:0000138 | CHEBI:106860 |
| 1 | HP:0000138 | CHEBI:110748 |
| 1 | HP:0000138 | CHEBI:111057 |
| 1 | HP:0000138 | CHEBI:110567 |
| 1 | HP:0000138 | CHEBI:113321 |
| 1 | HP:0000138 | HP:0003574 |
| 1 | HP:0000138 | CHEBI:106459 |
| 1 | HP:0000138 | HP:0000976 |
| 1 | HP:0000138 | HP:0003964 |
| 1 | HP:0000138 | CHEBI:106672 |
| 1 | HP:0000138 | CHEBI:127381 |
| 1 | HP:0000138 | CHEBI:110580 |
| 1 | HP:0000138 | CHEBI:3654 |
| 1 | HP:0000138 | HP:0001662 |
| 1 | HP:0000138 | HP:0001962 |
| 1 | HP:0000138 | CHEBI:126080 |
| 1 | HP:0000138 | CHEBI:110888 |
| 1 | HP:0000138 | CHEBI:112065 |
| 1 | HP:0000138 | CHEBI:99884 |
| 1 | HP:0000138 | HP:0033591 |
| 1 | HP:0000138 | CHEBI:110762 |
| 1 | HP:0000138 | CHEBI:128499 |
| 1 | HP:0000138 | HP:0002077 |
| 1 | HP:0000138 | HP:0012317 |
| 1 | HP:0000138 | HP:0003876 |
| 1 | HP:0000138 | CHEBI:112518 |
| 1 | HP:0000138 | CHEBI:113502 |
| 1 | HP:0000138 | CHEBI:99928 |
| 1 | HP:0000138 | CHEBI:130963 |
| 1 | HP:0000138 | CHEBI:106446 |
| 1 | HP:0000138 | CHEBI:111971 |
| 1 | HP:0000138 | CHEBI:112049 |
| 1 | HP:0000138 | CHEBI:110809 |
| 1 | HP:0000138 | CHEBI:110629 |
| 1 | HP:0000138 | CHEBI:111897 |
| 1 | HP:0000138 | CHEBI:9124 |
| 1 | HP:0000138 | CHEBI:130077 |
| 1 | HP:0000138 | CHEBI:106592 |
| 1 | HP:0000138 | CHEBI:106598 |
| 1 | HP:0000138 | CHEBI:106753 |
| 1 | HP:0000138 | CHEBI:106405 |
| 1 | HP:0000138 | CHEBI:112008 |
| 1 | HP:0000138 | CHEBI:99209 |
| 1 | HP:0000138 | CHEBI:106412 |
| 1 | HP:0000138 | HP:0010784 |
| 1 | HP:0000138 | CHEBI:110905 |
| 1 | HP:0000138 | CHEBI:94559 |
| 1 | HP:0000138 | CHEBI:125302 |
| 1 | HP:0000138 | CHEBI:127737 |
| 1 | HP:0000138 | HP:0000875 |
| 1 | HP:0000138 | CHEBI:111069 |
| 1 | HP:0000138 | CHEBI:110890 |
| 1 | HP:0000138 | CHEBI:2668 |
| 1 | HP:0000138 | CHEBI:106975 |
| 1 | HP:0000138 | CHEBI:111958 |
| 1 | HP:0000138 | CHEBI:15365 |
| 1 | HP:0000138 | CHEBI:131265 |
| 1 | HP:0000138 | CHEBI:110639 |
| 1 | HP:0000138 | HP:0033761 |
| 1 | HP:0000138 | CHEBI:110618 |
| 1 | HP:0000138 | CHEBI:111956 |
| 1 | HP:0000138 | CHEBI:53796 |
| 1 | HP:0000138 | CHEBI:2550 |
| 1 | HP:0000138 | HP:0040160 |
| 1 | HP:0000138 | HP:0008843 |
| 1 | HP:0000138 | CHEBI:128977 |
| 1 | HP:0000138 | CHEBI:111117 |
| 1 | HP:0000138 | HP:0001051 |
| 1 | HP:0000138 | CHEBI:3724 |
| 1 | HP:0000138 | CHEBI:106933 |
| 1 | HP:0000138 | HP:0002640 |
| 1 | HP:0000138 | CHEBI:127494 |
| 1 | HP:0000138 | CHEBI:130005 |
| 1 | HP:0000138 | CHEBI:111919 |
| 1 | HP:0000138 | CHEBI:111849 |
| 1 | HP:0000138 | CHEBI:111089 |
| 1 | HP:0000138 | CHEBI:100275 |
| 1 | HP:0000138 | CHEBI:99565 |
| 1 | HP:0000138 | CHEBI:106915 |
| 1 | HP:0000138 | CHEBI:100253 |
| 1 | HP:0000138 | CHEBI:110698 |
| 1 | HP:0000138 | HP:0008076 |
| 1 | HP:0000138 | CHEBI:106619 |
| 1 | HP:0000138 | HP:0032282 |
| 1 | HP:0000138 | CHEBI:106644 |
| 1 | HP:0000138 | HP:0012625 |
| 1 | HP:0000138 | HP:0012623 |
| 1 | HP:0000138 | CHEBI:106624 |
| 1 | HP:0000138 | CHEBI:130649 |
| 1 | HP:0000138 | CHEBI:111872 |
| 1 | HP:0000138 | CHEBI:106989 |
| 1 | HP:0000138 | CHEBI:107022 |
| 1 | HP:0000138 | CHEBI:106924 |
| 1 | HP:0000138 | HP:0550003 |
| 1 | HP:0000138 | CHEBI:135925 |
| 1 | HP:0000138 | CHEBI:111859 |
| 1 | HP:0000138 | CHEBI:95721 |
| 1 | HP:0000138 | CHEBI:131237 |
| 1 | HP:0000138 | CHEBI:106510 |
| 1 | HP:0000138 | CHEBI:130855 |
| 1 | HP:0000138 | CHEBI:32068 |
| 1 | HP:0000138 | CHEBI:126158 |
| 1 | HP:0000138 | HP:0005305 |
| 1 | HP:0000138 | CHEBI:106731 |
| 1 | HP:0000138 | CHEBI:99467 |
| 1 | HP:0000138 | HP:0012624 |
| 1 | HP:0000138 | CHEBI:111055 |
| 1 | HP:0000138 | CHEBI:99599 |
| 1 | HP:0000138 | CHEBI:106506 |
| 1 | HP:0000138 | HP:0004943 |
| 1 | HP:0000138 | CHEBI:106573 |
| 1 | HP:0000138 | CHEBI:9123 |
| 1 | HP:0000138 | CHEBI:106529 |
| 1 | HP:0000138 | HP:0002083 |
| 1 | HP:0000138 | CHEBI:31526 |
| 1 | HP:0000138 | CHEBI:126583 |
| 1 | HP:0000138 | HP:0006577 |
| 1 | HP:0000138 | CHEBI:127953 |
| 1 | HP:0000138 | HP:0100817 |
| 1 | HP:0000138 | CHEBI:29516 |
| 1 | HP:0000138 | HP:0100281 |
| 1 | HP:0000138 | CHEBI:131032 |
| 1 | HP:0000138 | CHEBI:128200 |
| 1 | HP:0000138 | CHEBI:95611 |
| 1 | HP:0000138 | CHEBI:110596 |
| 1 | HP:0000138 | CHEBI:31836 |
| 1 | HP:0000138 | CHEBI:110956 |
| 1 | HP:0000138 | CHEBI:126051 |
| 1 | HP:0000138 | CHEBI:99088 |
| 1 | HP:0000138 | HP:0005681 |
| 1 | HP:0000138 | CHEBI:127466 |
| 1 | HP:0000138 | CHEBI:131276 |
| 1 | HP:0000138 | CHEBI:112463 |
| 1 | HP:0000138 | CHEBI:126308 |
| 1 | MONDO:0006195 | ECTO:9002151 |
| 1 | MONDO:0006195 | ECTO:0070001 |
| 1 | MONDO:0006195 | ECTO:9000087 |
| 1 | MONDO:0006195 | ECTO:0070014 |
| 1 | MONDO:0006195 | MONDO:0002251 |
| 1 | MONDO:0006195 | MONDO:0002974 |
| 1 | MONDO:0006195 | ECTO:0070186 |
| 1 | MONDO:0006195 | MONDO:0003240 |
| 1 | MONDO:0006195 | ECTO:0070043 |
| 1 | MONDO:0006195 | ECTO:0070069 |
| 1 | MONDO:0006195 | ECTO:0070166 |
| 1 | MONDO:0006195 | ECTO:0070135 |
| 1 | MONDO:0006195 | ECTO:0070137 |
| 1 | MONDO:0006195 | ECTO:0070033 |
| 1 | MONDO:0006195 | ECTO:0070184 |
| 1 | MONDO:0006195 | ECTO:0070034 |
| 1 | MONDO:0006195 | ECTO:0070163 |
| 1 | MONDO:0006195 | ECTO:0070129 |
| 1 | MONDO:0006195 | ECTO:0070091 |
| 1 | MONDO:0006195 | ECTO:9000074 |
| 1 | MONDO:0006195 | ECTO:7000144 |
| 1 | MONDO:0006195 | ECTO:9000038 |
| 1 | MONDO:0006195 | ECTO:7000143 |
| 1 | MONDO:0006195 | ECTO:9000052 |
| 1 | MONDO:0006195 | ECTO:0500008 |
| 1 | MONDO:0006195 | ECTO:0500017 |
| 1 | MONDO:0006195 | ECTO:0500016 |
| 1 | MONDO:0006195 | ECTO:9000063 |
| 1 | MONDO:0006195 | ECTO:9002164 |
| 1 | MONDO:0006195 | ECTO:7000030 |
| 1 | MONDO:0006195 | ECTO:9000103 |
| 1 | MONDO:0006195 | ECTO:0000207 |
| 1 | MONDO:0006195 | ECTO:9000085 |
| 1 | MONDO:0006195 | ECTO:9000039 |
| 1 | MONDO:0006195 | ECTO:9000016 |
| 1 | MONDO:0006195 | CHEBI:6904 |
| 1 | MONDO:0006195 | ECTO:9000031 |
| 1 | MONDO:0006195 | ECTO:9000018 |
| 1 | MONDO:0006195 | ECTO:0500023 |
| 1 | MONDO:0006195 | CHEBI:6801 |
| 1 | MONDO:0006195 | CHEBI:3756 |
| 1 | MONDO:0006195 | CHEBI:6538 |
| 1 | MONDO:0006195 | ECTO:0070038 |
| 1 | MONDO:0006195 | ECTO:9002169 |
| 1 | MONDO:0006195 | ECTO:0070161 |
| 1 | MONDO:0006195 | ECTO:0070016 |
| 1 | MONDO:0006195 | ECTO:0070028 |
| 1 | MONDO:0006195 | ECTO:0070061 |
| 1 | MONDO:0006195 | ECTO:0070123 |
| 1 | MONDO:0006195 | ECTO:0070090 |
| 1 | MONDO:0006195 | ECTO:0070071 |
| 1 | MONDO:0006195 | ECTO:0070013 |
| 1 | MONDO:0006195 | ECTO:0070168 |
| 1 | MONDO:0006195 | ECTO:9000037 |
| 1 | MONDO:0006195 | ECTO:9000125 |
| 1 | MONDO:0006195 | ECTO:0070170 |
| 1 | MONDO:0006195 | ECTO:0070152 |
| 1 | MONDO:0006195 | ECTO:0070029 |
| 1 | MONDO:0006195 | MONDO:0002715 |
| 1 | MONDO:0006195 | MONDO:0004375 |
| 1 | MONDO:0006195 | MONDO:0010030 |
| 1 | MONDO:0006195 | MONDO:0005393 |
| 1 | MONDO:0006195 | MONDO:0004425 |
| 1 | MONDO:0006195 | CHEBI:8777 |
| 1 | MONDO:0006195 | CHEBI:6993 |
| 1 | MONDO:0006195 | ECTO:9000083 |
| 1 | MONDO:0006195 | ECTO:0001566 |
| 1 | MONDO:0006195 | ECTO:9000104 |
| 1 | MONDO:0006195 | ECTO:0000515 |
| 1 | MONDO:0006195 | ECTO:7000129 |
| 1 | MONDO:0006195 | ECTO:7000132 |
| 1 | MONDO:0006195 | ECTO:9000941 |
| 1 | MONDO:0006195 | CHEBI:10650 |
| 1 | MONDO:0006195 | CHEBI:50270 |
| 1 | MONDO:0006195 | CHEBI:6367 |
| 1 | MONDO:0006195 | CHEBI:43755 |
| 1 | MONDO:0006195 | GO:0042697 |
| 1 | MONDO:0006195 | HP:0100279 |
| 1 | MONDO:0006195 | HP:0002140 |
| 1 | MONDO:0006195 | HP:0002076 |
| 1 | MONDO:0006195 | HP:0200063 |
| 1 | MONDO:0006195 | CHEBI:10125 |
| 1 | MONDO:0006195 | CHEBI:5778 |
| 1 | MONDO:0006195 | ECTO:0070084 |
| 1 | MONDO:0006195 | ECTO:0070020 |
| 1 | MONDO:0006195 | ECTO:0070010 |
| 1 | MONDO:0006195 | ECTO:9000954 |
| 1 | MONDO:0006195 | ECTO:9000133 |
| 1 | MONDO:0006195 | ECTO:0070153 |
| 1 | MONDO:0006195 | HP:0033167 |
| 1 | MONDO:0006195 | ECTO:0070104 |
| 1 | MONDO:0006195 | ECTO:0070172 |
| 1 | MONDO:0006195 | ECTO:0070131 |
| 1 | MONDO:0006195 | MONDO:0008170 |
| 1 | MONDO:0006195 | MONDO:0005010 |
| 1 | MONDO:0006195 | MONDO:0024664 |
| 1 | MONDO:0006195 | ECTO:0070130 |
| 1 | MONDO:0006195 | ECTO:0070060 |
| 1 | MONDO:0006195 | ECTO:0070070 |
| 1 | MONDO:0006195 | ECTO:0070114 |
| 1 | MONDO:0006195 | ECTO:0070027 |
| 1 | MONDO:0006195 | ECTO:0070044 |
| 1 | MONDO:0006195 | ECTO:0070124 |
| 1 | MONDO:0006195 | ECTO:9002147 |
| 1 | MONDO:0006195 | ECTO:9000945 |
| 1 | MONDO:0006195 | ECTO:7000146 |
| 1 | MONDO:0006195 | ECTO:9000082 |
| 1 | MONDO:0006195 | ECTO:9000092 |
| 1 | MONDO:0006195 | ECTO:7000142 |
| 1 | MONDO:0006195 | ECTO:9001538 |
| 1 | MONDO:0006195 | CHEBI:36796 |
| 1 | MONDO:0006195 | ECTO:2000005 |
| 1 | MONDO:0006195 | HP:0000147 |
| 1 | MONDO:0006195 | HP:0012886 |
| 1 | MONDO:0006195 | CHEBI:6541 |
| 1 | MONDO:0006195 | CHEBI:5118 |
| 1 | MONDO:0006195 | MAXO:0001058 |
| 1 | MONDO:0006195 | HP:0001891 |
| 1 | MONDO:0006195 | HP:0001635 |
| 1 | MONDO:0006195 | HP:0000131 |
| 1 | MONDO:0006195 | HP:0005994 |
| 1 | MONDO:0006195 | ECTO:9000424 |
| 1 | MONDO:0006195 | ECTO:9000051 |
| 1 | MONDO:0006195 | CHEBI:7772 |
| 1 | MONDO:0006195 | ECTO:0070128 |
| 1 | MONDO:0006195 | ECTO:0070155 |
| 1 | MONDO:0006195 | ECTO:0070003 |
| 1 | MONDO:0006195 | ECTO:0070009 |
| 1 | MONDO:0006195 | ECTO:0070158 |
| 1 | MONDO:0006195 | ECTO:0070121 |
| 1 | MONDO:0006195 | ECTO:0070187 |
| 1 | MONDO:0006195 | ECTO:0070037 |
| 1 | MONDO:0006195 | ECTO:0070012 |
| 1 | MONDO:0006195 | ECTO:9000123 |
| 1 | MONDO:0006195 | ECTO:9000299 |
| 1 | MONDO:0006195 | ECTO:0070210 |
| 1 | MONDO:0006195 | ECTO:0070138 |
| 1 | MONDO:0006195 | ECTO:0070036 |
| 1 | MONDO:0006195 | ECTO:0070159 |
| 1 | MONDO:0006195 | MONDO:0020642 |
| 1 | MONDO:0006195 | ECTO:0070022 |
| 1 | MONDO:0006195 | MONDO:0018076 |
| 1 | MONDO:0006195 | MONDO:0008364 |
| 1 | MONDO:0006195 | MONDO:0011382 |
| 1 | MONDO:0006195 | ECTO:0070174 |
| 1 | MONDO:0006195 | HP:0004936 |
| 1 | MONDO:0006195 | HP:0001370 |
| 1 | MONDO:0006195 | HP:0000938 |
| 1 | MONDO:0006195 | HP:0000964 |
| 1 | MONDO:0006195 | HP:0002592 |
| 1 | MONDO:0006195 | ECTO:0000001 |
| 1 | MONDO:0006195 | ECTO:9000025 |
| 1 | MONDO:0006195 | ECTO:9000028 |
| 1 | MONDO:0006195 | CHEBI:3219 |
| 1 | MONDO:0006195 | CHEBI:36791 |
| 1 | MONDO:0006195 | CHEBI:5051 |
| 1 | MONDO:0006195 | HP:0000822 |
| 1 | MONDO:0006195 | HP:0100280 |
| 1 | MONDO:0006195 | HP:0004789 |
| 1 | MONDO:0006195 | HP:0001297 |
| 1 | MONDO:0006195 | HP:0012887 |
| 1 | MONDO:0006195 | MAXO:0001067 |
| 1 | MONDO:0006195 | HP:0000787 |
| 1 | MONDO:0006195 | HP:0001025 |
| 1 | MONDO:0006195 | ECTO:9000068 |
| 1 | MONDO:0006195 | HP:0000939 |
| 1 | MONDO:0006195 | HP:0000853 |
| 1 | MONDO:0006195 | HP:0001681 |
| 1 | MONDO:0006195 | ECTO:0500004 |
| 1 | MONDO:0006195 | MONDO:0008345 |
| 1 | MONDO:0006195 | ECTO:0070133 |
| 1 | MONDO:0006195 | ECTO:9000243 |
| 1 | MONDO:0006195 | ECTO:9000210 |
| 1 | MONDO:0006195 | ECTO:0070002 |
| 1 | MONDO:0006195 | ECTO:0070015 |
| 1 | MONDO:0006195 | ECTO:0070075 |
| 1 | MONDO:0006195 | ECTO:0070188 |
| 1 | MONDO:0006195 | ECTO:0070165 |
| 1 | MONDO:0006195 | MONDO:0005027 |
| 1 | MONDO:0006195 | ECTO:0070035 |
| 1 | MONDO:0006195 | ECTO:0070183 |
| 1 | MONDO:0006195 | ECTO:0070047 |
| 1 | MONDO:0006195 | ECTO:0070110 |
| 1 | MONDO:0006195 | ECTO:0070021 |
| 1 | MONDO:0006195 | ECTO:0070171 |
| 1 | MONDO:0006195 | ECTO:0070024 |
| 1 | MONDO:0006195 | ECTO:0070167 |
| 1 | MONDO:0006195 | MONDO:0005420 |
| 1 | MONDO:0006195 | MONDO:0005015 |
| 1 | MONDO:0006195 | MONDO:0004979 |
| 1 | MONDO:0006195 | MAXO:0000257 |
| 1 | MONDO:0006195 | HP:0012330 |
| 1 | MONDO:0006195 | HP:0002621 |
| 1 | MONDO:0006195 | HP:0012622 |
| 1 | MONDO:0006195 | HP:0002608 |
| 1 | MONDO:0006195 | ECTO:9000099 |
| 1 | MONDO:0006195 | ECTO:9000022 |
| 1 | MONDO:0006195 | ECTO:0500010 |
| 1 | MONDO:0006195 | ECTO:9000071 |
| 1 | MONDO:0006195 | ECTO:9002142 |
| 1 | MONDO:0006195 | ECTO:9000059 |
| 1 | MONDO:0006195 | ECTO:7000141 |
| 1 | MONDO:0006195 | ECTO:7000131 |
| 1 | MONDO:0006195 | ECTO:9000090 |
| 1 | MONDO:0006195 | CHEBI:6539 |
| 1 | MONDO:0006195 | CHEBI:6741 |
| 1 | MONDO:0006195 | ECTO:7000154 |
| 1 | MONDO:0006195 | ECTO:9000060 |
| 1 | MONDO:0006195 | ECTO:9002143 |
| 1 | MONDO:0006195 | ECTO:0070112 |
| 1 | MONDO:0006195 | ECTO:0070067 |
| 1 | MONDO:0006195 | ECTO:0070025 |
| 1 | MONDO:0006195 | ECTO:0070108 |
| 1 | MONDO:0006195 | ECTO:0070056 |
| 1 | MONDO:0006195 | ECTO:0070042 |
| 1 | MONDO:0006195 | ECTO:0070046 |
| 1 | MONDO:0006195 | MONDO:0005546 |
| 1 | MONDO:0006195 | MONDO:0006920 |
| 1 | MONDO:0006195 | MONDO:0004790 |
| 1 | MONDO:0006195 | MONDO:0005002 |
| 1 | MONDO:0006195 | ECTO:0070169 |
| 1 | MONDO:0006195 | CHEBI:42797 |
| 1 | MONDO:0006195 | ECTO:0070039 |
| 1 | MONDO:0006195 | ECTO:0070139 |
| 1 | MONDO:0006195 | ECTO:0070209 |
| 1 | MONDO:0006195 | ECTO:9000229 |
| 1 | MONDO:0006195 | ECTO:9000253 |
| 1 | MONDO:0006195 | ECTO:0070004 |
| 1 | MONDO:0006195 | ECTO:0070017 |
| 1 | MONDO:0006195 | ECTO:0070185 |
| 1 | MONDO:0006195 | CHEBI:6484 |
| 1 | MONDO:0006195 | CHEBI:6446 |
| 1 | MONDO:0006195 | HP:0002758 |
| 1 | MONDO:0006195 | HP:0002326 |
| 1 | MONDO:0006195 | HP:0000138 |
| 1 | MONDO:0006195 | ECTO:9001831 |
| 1 | MONDO:0006195 | CHEBI:50309 |
| 1 | MONDO:0006195 | ECTO:7000155 |
| 1 | MONDO:0006195 | ECTO:9000036 |
| 1 | MONDO:0006195 | ECTO:0000530 |
| 1 | MONDO:0006195 | ECTO:9000017 |
| 1 | MONDO:0006195 | CHEBI:50275 |
| 1 | MONDO:0006195 | CHEBI:5855 |
| 1 | MONDO:0006195 | MAXO:0000259 |
| 1 | MONDO:0006195 | HP:0001394 |
| 1 | MONDO:0006195 | HP:0100614 |
| 1 | MONDO:0006195 | HP:0011675 |
| 1 | MONDO:0006195 | HP:0001658 |
| 1 | MONDO:0006195 | HP:0100324 |
| 1 | MONDO:0006195 | CHEBI:10126 |
| 1 | MONDO:0006195 | ECTO:9000076 |
| 1 | MONDO:0006195 | ECTO:9000020 |
| 1 | MONDO:0006195 | ECTO:7000153 |
| 1 | MONDO:0006195 | ECTO:0001571 |
| 1 | MONDO:0006195 | ECTO:9000079 |
| 1 | MONDO:0006195 | ECTO:7000151 |
| 1 | MONDO:0006195 | ECTO:7000152 |
| 1 | MONDO:0006195 | ECTO:9000042 |
| 1 | MONDO:0006195 | ECTO:9000019 |
| 1 | MONDO:0006195 | ECTO:9000048 |
| 1 | MONDO:0006195 | ECTO:9002166 |
| 1 | MONDO:0006195 | ECTO:9001437 |
| 1 | MONDO:0006195 | ECTO:9000058 |
| 1 | MONDO:0006195 | ECTO:7000140 |
| 1 | MONDO:0006195 | ECTO:7000005 |
| 1 | MONDO:0006195 | ECTO:9000081 |
| 1 | MONDO:0006195 | ECTO:9001302 |
| 1 | MONDO:0006195 | ECTO:7000017 |
| 1 | MONDO:0006195 | ECTO:0500006 |
| 1 | MONDO:0006195 | ECTO:9000032 |
| 1 | MONDO:0006195 | ECTO:9002165 |
| 1 | MONDO:0006195 | ECTO:0500009 |
| 1 | MONDO:0006195 | ECTO:9000024 |
| 1 | MONDO:0006195 | ECTO:0500021 |
| 1 | MONDO:0006195 | ECTO:9000057 |
| 1 | MONDO:0006195 | ECTO:7000150 |
| 1 | MONDO:0006195 | ECTO:9001465 |
| 1 | MONDO:0006195 | CHEBI:39548 |
| 1 | MONDO:0006195 | MONDO:0005406 |
| 1 | MONDO:0006195 | ECTO:0070026 |
| 1 | MONDO:0006195 | MONDO:0008228 |
| 1 | MONDO:0006195 | MONDO:0007915 |
| 1 | MONDO:0006195 | MONDO:0005133 |
| 1 | MONDO:0006195 | ECTO:0070125 |
| 1 | MONDO:0006195 | ECTO:0070157 |
| 1 | MONDO:0006195 | ECTO:0070062 |
| 1 | MONDO:0006195 | ECTO:0070109 |
| 1 | MONDO:0006195 | ECTO:0070132 |
| 1 | MONDO:0006195 | ECTO:0070119 |
| 1 | MONDO:0006195 | ECTO:0070048 |
| 1 | MONDO:0006195 | ECTO:0070000 |
| 1 | MONDO:0006195 | ECTO:0070151 |
| 1 | MONDO:0006195 | ECTO:0070189 |
| 1 | MONDO:0006195 | ECTO:0070078 |
| 1 | MONDO:0006195 | ECTO:0070018 |
| 1 | MONDO:0006195 | ECTO:0070007 |
| 1 | MONDO:0006195 | ECTO:9000192 |
| 1 | MONDO:0006195 | ECTO:9000143 |
| 1 | MONDO:0006195 | ECTO:9000043 |
| 1 | MONDO:0006195 | ECTO:9000069 |
| 1 | MONDO:0006195 | ECTO:9000027 |
| 1 | MONDO:0006195 | ECTO:9001450 |
| 1 | MONDO:0006195 | MONDO:0006507 |
| 1 | MONDO:0006195 | ECTO:0070023 |
| 1 | MONDO:0006195 | ECTO:0070011 |
| 1 | MONDO:0006195 | ECTO:0070164 |
| 1 | MONDO:0006195 | ECTO:9000224 |
| 1 | MONDO:0006195 | ECTO:0070107 |
| 1 | MONDO:0006195 | ECTO:0070085 |
| 1 | MONDO:0006195 | ECTO:0070154 |
| 1 | MONDO:0006195 | MONDO:0006107 |
| 1 | MONDO:0006195 | ECTO:0070008 |
| 1 | MONDO:0006195 | MONDO:0005281 |
| 1 | MONDO:0006195 | MONDO:0005083 |
| 1 | MONDO:0006195 | ECTO:0070173 |
| 1 | MONDO:0006195 | ECTO:0070156 |
| 1 | MONDO:0006195 | ECTO:0070148 |
| 1 | MONDO:0006195 | ECTO:0070116 |
| 1 | MONDO:0006195 | ECTO:0070182 |
| 1 | MONDO:0006195 | ECTO:0070045 |
| 1 | MONDO:0006195 | ECTO:0070134 |
| 1 | MONDO:0006195 | ECTO:0070162 |
| 1 | MONDO:0006195 | ECTO:9000070 |
| 1 | MONDO:0006195 | ECTO:9002113 |
| 1 | MONDO:0006195 | ECTO:9002141 |
| 1 | MONDO:0006195 | ECTO:0500007 |
| 1 | MONDO:0006195 | ECTO:0500022 |
| 1 | MONDO:0006195 | ECTO:9000089 |
| 1 | MONDO:0006195 | ECTO:7000028 |
| 1 | MONDO:0006195 | ECTO:9000023 |
| 1 | MONDO:0006195 | ECTO:7000149 |
| 1 | MONDO:0006195 | ECTO:9000049 |
| 1 | MONDO:0006195 | ECTO:8000046 |
| 1 | MONDO:0006195 | ECTO:9000034 |
| 1 | MONDO:0006195 | CHEBI:50686 |
| 1 | MONDO:0006195 | CHEBI:3562 |
| 1 | MONDO:0006195 | CHEBI:2668 |
| 1 | MONDO:0006195 | HP:0025081 |
| 1 | MONDO:0006195 | CHEBI:9584 |
| 1 | MONDO:0006195 | CHEBI:3723 |
| 1 | MONDO:0006195 | CHEBI:135925 |
| 1 | MONDO:0006195 | CHEBI:3724 |
| 1 | MONDO:0006195 | HP:0030425 |
| 1 | MONDO:0006195 | CHEBI:9123 |
| 1 | MONDO:0006195 | CHEBI:9124 |
| 1 | MONDO:0006195 | CHEBI:135931 |
| 1 | MONDO:0006195 | HP:0002083 |
| 1 | MONDO:0006195 | CHEBI:3654 |
| 1 | MONDO:0006195 | HP:0002077 |
| 1 | MONDO:0006195 | CHEBI:9654 |
| 1 | MONDO:0006195 | CHEBI:2611 |
| 1 | MONDO:0006195 | CHEBI:2550 |
| 1 | MONDO:0006195 | CHEBI:31836 |
| 1 | MONDO:0006195 | CHEBI:9648 |
| 1 | MONDO:0006195 | HP:0009798 |
| 1 | MONDO:0006195 | CHEBI:15365 |
| 1 | MONDO:0006195 | CHEBI:31526 |
| 1 | MONDO:0006195 | CHEBI:9150 |
| 1 | MONDO:0006195 | CHEBI:9435 |
| 1 | MONDO:0006195 | HP:0002640 |
| 1 | HP:0000131 | CHEBI:103596 |
| 1 | HP:0000131 | CHEBI:122544 |
| 1 | HP:0000131 | CHEBI:103567 |
| 1 | HP:0000131 | CHEBI:122573 |
| 1 | HP:0000131 | CHEBI:103552 |
| 1 | HP:0000131 | CHEBI:103585 |
| 1 | HP:0000131 | CHEBI:97111 |
| 1 | HP:0000131 | CHEBI:97120 |
| 1 | HP:0000131 | CHEBI:103578 |
| 1 | HP:0000131 | CHEBI:103572 |
| 1 | HP:0000131 | CHEBI:122561 |
| 1 | HP:0000131 | CHEBI:122527 |
| 1 | HP:0000131 | CHEBI:95897 |
| 1 | HP:0000131 | CHEBI:122532 |
| 1 | HP:0000131 | CHEBI:122515 |
| 1 | HP:0000131 | CHEBI:122511 |
| 1 | HP:0000131 | CHEBI:122505 |
| 1 | HP:0000131 | CHEBI:122534 |
| 1 | HP:0000131 | CHEBI:98024 |
| 1 | HP:0000131 | CHEBI:103633 |
| 1 | HP:0000131 | CHEBI:97175 |
| 1 | HP:0000131 | CHEBI:8869 |
| 1 | HP:0000131 | CHEBI:103646 |
| 1 | HP:0000131 | CHEBI:102500 |
| 1 | HP:0000131 | CHEBI:102510 |
| 1 | HP:0000131 | CHEBI:103619 |
| 1 | HP:0000131 | CHEBI:103665 |
| 1 | HP:0000131 | CHEBI:103653 |
| 1 | HP:0000131 | CHEBI:103605 |
| 1 | HP:0000131 | CHEBI:102573 |
| 1 | HP:0000131 | CHEBI:103622 |
| 1 | HP:0000131 | CHEBI:103628 |
| 1 | HP:0000131 | CHEBI:102614 |
| 1 | HP:0000131 | CHEBI:122681 |
| 1 | HP:0000131 | CHEBI:122687 |
| 1 | HP:0000131 | CHEBI:122686 |
| 1 | HP:0000131 | CHEBI:97044 |
| 1 | HP:0000131 | CHEBI:97012 |
| 1 | HP:0000131 | CHEBI:97020 |
| 1 | HP:0000131 | CHEBI:97007 |
| 1 | HP:0000131 | CHEBI:97039 |
| 1 | HP:0000131 | CHEBI:122692 |
| 1 | HP:0000131 | CHEBI:122648 |
| 1 | HP:0000131 | CHEBI:122627 |
| 1 | HP:0000131 | CHEBI:122622 |
| 1 | HP:0000131 | CHEBI:122653 |
| 1 | HP:0000131 | CHEBI:122661 |
| 1 | HP:0000131 | CHEBI:122677 |
| 1 | HP:0000131 | CHEBI:122673 |
| 1 | HP:0000131 | CHEBI:122669 |
| 1 | HP:0000131 | CHEBI:122666 |
| 1 | HP:0000131 | CHEBI:102233 |
| 1 | HP:0000131 | CHEBI:122616 |
| 1 | HP:0000131 | CHEBI:102662 |
| 1 | HP:0000131 | CHEBI:122717 |
| 1 | HP:0000131 | CHEBI:122716 |
| 1 | HP:0000131 | CHEBI:122706 |
| 1 | HP:0000131 | CHEBI:122705 |
| 1 | HP:0000131 | CHEBI:122700 |
| 1 | HP:0000131 | CHEBI:102694 |
| 1 | HP:0000131 | CHEBI:95824 |
| 1 | HP:0000131 | CHEBI:122584 |
| 1 | HP:0000131 | CHEBI:122720 |
| 1 | HP:0000131 | CHEBI:122611 |
| 1 | HP:0000131 | CHEBI:8270 |
| 1 | HP:0000131 | CHEBI:122779 |
| 1 | HP:0000131 | CHEBI:122600 |
| 1 | HP:0000131 | CHEBI:122771 |
| 1 | HP:0000131 | CHEBI:122766 |
| 1 | HP:0000131 | CHEBI:122798 |
| 1 | HP:0000131 | CHEBI:122781 |
| 1 | HP:0000131 | CHEBI:102682 |
| 1 | HP:0000131 | CHEBI:104621 |
| 1 | HP:0000131 | CHEBI:104629 |
| 1 | HP:0000131 | CHEBI:104625 |
| 1 | HP:0000131 | HP:0005994 |
| 1 | HP:0000131 | CHEBI:104546 |
| 1 | HP:0000131 | CHEBI:104550 |
| 1 | HP:0000131 | CHEBI:104558 |
| 1 | HP:0000131 | CHEBI:104526 |
| 1 | HP:0000131 | CHEBI:104608 |
| 1 | HP:0000131 | CHEBI:103709 |
| 1 | HP:0000131 | CHEBI:98120 |
| 1 | HP:0000131 | CHEBI:103690 |
| 1 | HP:0000131 | CHEBI:95990 |
| 1 | HP:0000131 | CHEBI:97238 |
| 1 | HP:0000131 | CHEBI:95952 |
| 1 | HP:0000131 | CHEBI:98150 |
| 1 | HP:0000131 | CHEBI:103732 |
| 1 | HP:0000131 | CHEBI:98187 |
| 1 | HP:0000131 | CHEBI:97306 |
| 1 | HP:0000131 | CHEBI:103722 |
| 1 | HP:0000131 | CHEBI:122803 |
| 1 | HP:0000131 | CHEBI:122811 |
| 1 | HP:0000131 | CHEBI:104534 |
| 1 | HP:0000131 | CHEBI:98259 |
| 1 | HP:0000131 | CHEBI:123672 |
| 1 | HP:0000131 | CHEBI:103627 |
| 1 | HP:0000131 | CHEBI:103609 |
| 1 | HP:0000131 | CHEBI:123691 |
| 1 | HP:0000131 | CHEBI:103651 |
| 1 | HP:0000131 | CHEBI:103658 |
| 1 | HP:0000131 | CHEBI:103386 |
| 1 | HP:0000131 | CHEBI:103643 |
| 1 | HP:0000131 | CHEBI:102745 |
| 1 | HP:0000131 | CHEBI:103372 |
| 1 | HP:0000131 | CHEBI:123647 |
| 1 | HP:0000131 | CHEBI:123637 |
| 1 | HP:0000131 | CHEBI:123631 |
| 1 | HP:0000131 | CHEBI:104717 |
| 1 | HP:0000131 | CHEBI:104710 |
| 1 | HP:0000131 | CHEBI:102729 |
| 1 | HP:0000131 | CHEBI:104714 |
| 1 | HP:0000131 | CHEBI:103347 |
| 1 | HP:0000131 | CHEBI:103564 |
| 1 | HP:0000131 | CHEBI:103614 |
| 1 | HP:0000131 | CHEBI:103357 |
| 1 | HP:0000131 | CHEBI:103362 |
| 1 | HP:0000131 | CHEBI:116021 |
| 1 | HP:0000131 | CHEBI:102870 |
| 1 | HP:0000131 | CHEBI:102878 |
| 1 | HP:0000131 | CHEBI:116013 |
| 1 | HP:0000131 | CHEBI:104850 |
| 1 | HP:0000131 | CHEBI:104854 |
| 1 | HP:0000131 | CHEBI:102802 |
| 1 | HP:0000131 | CHEBI:116047 |
| 1 | HP:0000131 | CHEBI:123728 |
| 1 | HP:0000131 | CHEBI:116024 |
| 1 | HP:0000131 | CHEBI:104776 |
| 1 | HP:0000131 | HP:0005086 |
| 1 | HP:0000131 | CHEBI:123760 |
| 1 | HP:0000131 | CHEBI:123762 |
| 1 | HP:0000131 | CHEBI:123772 |
| 1 | HP:0000131 | CHEBI:104786 |
| 1 | HP:0000131 | CHEBI:103675 |
| 1 | HP:0000131 | CHEBI:104767 |
| 1 | HP:0000131 | CHEBI:104770 |
| 1 | HP:0000131 | CHEBI:104744 |
| 1 | HP:0000131 | CHEBI:103593 |
| 1 | HP:0000131 | CHEBI:122519 |
| 1 | HP:0000131 | CHEBI:104748 |
| 1 | HP:0000131 | CHEBI:103478 |
| 1 | HP:0000131 | CHEBI:103471 |
| 1 | HP:0000131 | CHEBI:104656 |
| 1 | HP:0000131 | CHEBI:122569 |
| 1 | HP:0000131 | CHEBI:122576 |
| 1 | HP:0000131 | CHEBI:98950 |
| 1 | HP:0000131 | CHEBI:122571 |
| 1 | HP:0000131 | CHEBI:122547 |
| 1 | HP:0000131 | CHEBI:122548 |
| 1 | HP:0000131 | CHEBI:122558 |
| 1 | HP:0000131 | CHEBI:122525 |
| 1 | HP:0000131 | CHEBI:104688 |
| 1 | HP:0000131 | CHEBI:103445 |
| 1 | HP:0000131 | CHEBI:123854 |
| 1 | HP:0000131 | CHEBI:104691 |
| 1 | HP:0000131 | CHEBI:122539 |
| 1 | HP:0000131 | CHEBI:122503 |
| 1 | HP:0000131 | CHEBI:122507 |
| 1 | HP:0000131 | CHEBI:122513 |
| 1 | HP:0000131 | CHEBI:103482 |
| 1 | HP:0000131 | CHEBI:103487 |
| 1 | HP:0000131 | CHEBI:104643 |
| 1 | HP:0000131 | CHEBI:104724 |
| 1 | HP:0000131 | CHEBI:98238 |
| 1 | HP:0000131 | CHEBI:104750 |
| 1 | HP:0000131 | CHEBI:103334 |
| 1 | HP:0000131 | CHEBI:104758 |
| 1 | HP:0000131 | CHEBI:123981 |
| 1 | HP:0000131 | CHEBI:123973 |
| 1 | HP:0000131 | CHEBI:123996 |
| 1 | HP:0000131 | HP:0100735 |
| 1 | HP:0000131 | CHEBI:104735 |
| 1 | HP:0000131 | CHEBI:50309 |
| 1 | HP:0000131 | CHEBI:123920 |
| 1 | HP:0000131 | CHEBI:123910 |
| 1 | HP:0000131 | CHEBI:103422 |
| 1 | HP:0000131 | CHEBI:100443 |
| 1 | HP:0000131 | CHEBI:104662 |
| 1 | HP:0000131 | CHEBI:104667 |
| 1 | HP:0000131 | CHEBI:98271 |
| 1 | HP:0000131 | CHEBI:104678 |
| 1 | HP:0000131 | CHEBI:116000 |
| 1 | HP:0000131 | CHEBI:104841 |
| 1 | HP:0000131 | CHEBI:103706 |
| 1 | HP:0000131 | CHEBI:100171 |
| 1 | HP:0000131 | CHEBI:95572 |
| 1 | HP:0000131 | CHEBI:104612 |
| 1 | HP:0000131 | CHEBI:128305 |
| 1 | HP:0000131 | CHEBI:117704 |
| 1 | HP:0000131 | CHEBI:102021 |
| 1 | HP:0000131 | CHEBI:104543 |
| 1 | HP:0000131 | CHEBI:104555 |
| 1 | HP:0000131 | CHEBI:98826 |
| 1 | HP:0000131 | CHEBI:104521 |
| 1 | HP:0000131 | CHEBI:104528 |
| 1 | HP:0000131 | CHEBI:100988 |
| 1 | HP:0000131 | CHEBI:128216 |
| 1 | HP:0000131 | CHEBI:104532 |
| 1 | HP:0000131 | CHEBI:98873 |
| 1 | HP:0000131 | CHEBI:104581 |
| 1 | HP:0000131 | CHEBI:98214 |
| 1 | HP:0000131 | CHEBI:97871 |
| 1 | HP:0000131 | CHEBI:97886 |
| 1 | HP:0000131 | CHEBI:128256 |
| 1 | HP:0000131 | CHEBI:128312 |
| 1 | HP:0000131 | CHEBI:104605 |
| 1 | HP:0000131 | CHEBI:104635 |
| 1 | HP:0000131 | CHEBI:122512 |
| 1 | HP:0000131 | CHEBI:116040 |
| 1 | HP:0000131 | CHEBI:95511 |
| 1 | HP:0000131 | CHEBI:98308 |
| 1 | HP:0000131 | CHEBI:98333 |
| 1 | HP:0000131 | CHEBI:117691 |
| 1 | HP:0000131 | CHEBI:102075 |
| 1 | HP:0000131 | CHEBI:117683 |
| 1 | HP:0000131 | CHEBI:102079 |
| 1 | HP:0000131 | CHEBI:122509 |
| 1 | HP:0000131 | CHEBI:104630 |
| 1 | HP:0000131 | CHEBI:122506 |
| 1 | HP:0000131 | CHEBI:122528 |
| 1 | HP:0000131 | CHEBI:122524 |
| 1 | HP:0000131 | CHEBI:122556 |
| 1 | HP:0000131 | CHEBI:122545 |
| 1 | HP:0000131 | CHEBI:117670 |
| 1 | HP:0000131 | CHEBI:122577 |
| 1 | HP:0000131 | CHEBI:122574 |
| 1 | HP:0000131 | CHEBI:128251 |
| 1 | HP:0000131 | CHEBI:97853 |
| 1 | HP:0000131 | CHEBI:104561 |
| 1 | HP:0000131 | CHEBI:104836 |
| 1 | HP:0000131 | CHEBI:104477 |
| 1 | HP:0000131 | CHEBI:104443 |
| 1 | HP:0000131 | CHEBI:116065 |
| 1 | HP:0000131 | CHEBI:116069 |
| 1 | HP:0000131 | CHEBI:103772 |
| 1 | HP:0000131 | CHEBI:98735 |
| 1 | HP:0000131 | CHEBI:104455 |
| 1 | HP:0000131 | CHEBI:104821 |
| 1 | HP:0000131 | CHEBI:104818 |
| 1 | HP:0000131 | CHEBI:97992 |
| 1 | HP:0000131 | CHEBI:103753 |
| 1 | HP:0000131 | CHEBI:103759 |
| 1 | HP:0000131 | CHEBI:103768 |
| 1 | HP:0000131 | CHEBI:104860 |
| 1 | HP:0000131 | CHEBI:104864 |
| 1 | HP:0000131 | CHEBI:104869 |
| 1 | HP:0000131 | CHEBI:104849 |
| 1 | HP:0000131 | CHEBI:98760 |
| 1 | HP:0000131 | CHEBI:103738 |
| 1 | HP:0000131 | CHEBI:104464 |
| 1 | HP:0000131 | CHEBI:104566 |
| 1 | HP:0000131 | CHEBI:104488 |
| 1 | HP:0000131 | CHEBI:104577 |
| 1 | HP:0000131 | CHEBI:104508 |
| 1 | HP:0000131 | CHEBI:104505 |
| 1 | HP:0000131 | CHEBI:104512 |
| 1 | HP:0000131 | CHEBI:97957 |
| 1 | HP:0000131 | CHEBI:98803 |
| 1 | HP:0000131 | CHEBI:104490 |
| 1 | HP:0000131 | CHEBI:104484 |
| 1 | HP:0000131 | CHEBI:104494 |
| 1 | HP:0000131 | CHEBI:104416 |
| 1 | HP:0000131 | CHEBI:104422 |
| 1 | HP:0000131 | CHEBI:104427 |
| 1 | HP:0000131 | CHEBI:100914 |
| 1 | HP:0000131 | CHEBI:104432 |
| 1 | HP:0000131 | CHEBI:98787 |
| 1 | HP:0000131 | CHEBI:104409 |
| 1 | HP:0000131 | CHEBI:104401 |
| 1 | HP:0000131 | CHEBI:104407 |
| 1 | HP:0000131 | CHEBI:104696 |
| 1 | HP:0000131 | CHEBI:123872 |
| 1 | HP:0000131 | ECTO:9000016 |
| 1 | HP:0000131 | ECTO:9000039 |
| 1 | HP:0000131 | ECTO:9000085 |
| 1 | HP:0000131 | ECTO:0000207 |
| 1 | HP:0000131 | CHEBI:98626 |
| 1 | HP:0000131 | ECTO:9000103 |
| 1 | HP:0000131 | ECTO:7000030 |
| 1 | HP:0000131 | CHEBI:95683 |
| 1 | HP:0000131 | CHEBI:100528 |
| 1 | HP:0000131 | CHEBI:104379 |
| 1 | HP:0000131 | CHEBI:97607 |
| 1 | HP:0000131 | CHEBI:95628 |
| 1 | HP:0000131 | CHEBI:117894 |
| 1 | HP:0000131 | CHEBI:117843 |
| 1 | HP:0000131 | CHEBI:103229 |
| 1 | HP:0000131 | CHEBI:117867 |
| 1 | HP:0000131 | CHEBI:117856 |
| 1 | HP:0000131 | CHEBI:102792 |
| 1 | HP:0000131 | CHEBI:97601 |
| 1 | HP:0000131 | CHEBI:104395 |
| 1 | HP:0000131 | CHEBI:104375 |
| 1 | HP:0000131 | CHEBI:100541 |
| 1 | HP:0000131 | CHEBI:104370 |
| 1 | HP:0000131 | CHEBI:104364 |
| 1 | HP:0000131 | CHEBI:104369 |
| 1 | HP:0000131 | CHEBI:100552 |
| 1 | HP:0000131 | CHEBI:104371 |
| 1 | HP:0000131 | CHEBI:102711 |
| 1 | HP:0000131 | CHEBI:102389 |
| 1 | HP:0000131 | CHEBI:117644 |
| 1 | HP:0000131 | ECTO:0500023 |
| 1 | HP:0000131 | CHEBI:95732 |
| 1 | HP:0000131 | CHEBI:95747 |
| 1 | HP:0000131 | CHEBI:95714 |
| 1 | HP:0000131 | CHEBI:122819 |
| 1 | HP:0000131 | CHEBI:122815 |
| 1 | HP:0000131 | CHEBI:43755 |
| 1 | HP:0000131 | CHEBI:95648 |
| 1 | HP:0000131 | CHEBI:97525 |
| 1 | HP:0000131 | CHEBI:100585 |
| 1 | HP:0000131 | CHEBI:122664 |
| 1 | HP:0000131 | CHEBI:122667 |
| 1 | HP:0000131 | CHEBI:122675 |
| 1 | HP:0000131 | CHEBI:117769 |
| 1 | HP:0000131 | CHEBI:117765 |
| 1 | HP:0000131 | CHEBI:122640 |
| 1 | HP:0000131 | CHEBI:102929 |
| 1 | HP:0000131 | CHEBI:122657 |
| 1 | HP:0000131 | CHEBI:122620 |
| 1 | HP:0000131 | CHEBI:122628 |
| 1 | HP:0000131 | CHEBI:122634 |
| 1 | HP:0000131 | CHEBI:100638 |
| 1 | HP:0000131 | CHEBI:122639 |
| 1 | HP:0000131 | CHEBI:102936 |
| 1 | HP:0000131 | CHEBI:97445 |
| 1 | HP:0000131 | CHEBI:117617 |
| 1 | HP:0000131 | CHEBI:117613 |
| 1 | HP:0000131 | HP:0033762 |
| 1 | HP:0000131 | CHEBI:100342 |
| 1 | HP:0000131 | CHEBI:98984 |
| 1 | HP:0000131 | CHEBI:102995 |
| 1 | HP:0000131 | CHEBI:103538 |
| 1 | HP:0000131 | CHEBI:103819 |
| 1 | HP:0000131 | CHEBI:117640 |
| 1 | HP:0000131 | CHEBI:103467 |
| 1 | HP:0000131 | CHEBI:103462 |
| 1 | HP:0000131 | CHEBI:100412 |
| 1 | HP:0000131 | CHEBI:97361 |
| 1 | HP:0000131 | CHEBI:123802 |
| 1 | HP:0000131 | CHEBI:122587 |
| 1 | HP:0000131 | CHEBI:122581 |
| 1 | HP:0000131 | CHEBI:122592 |
| 1 | HP:0000131 | CHEBI:117603 |
| 1 | HP:0000131 | CHEBI:98306 |
| 1 | HP:0000131 | CHEBI:117668 |
| 1 | HP:0000131 | CHEBI:98313 |
| 1 | HP:0000131 | CHEBI:117661 |
| 1 | HP:0000131 | CHEBI:103521 |
| 1 | HP:0000131 | CHEBI:122617 |
| 1 | HP:0000131 | CHEBI:117658 |
| 1 | HP:0000131 | CHEBI:103544 |
| 1 | HP:0000131 | CHEBI:102919 |
| 1 | HP:0000131 | CHEBI:117787 |
| 1 | HP:0000131 | CHEBI:117782 |
| 1 | HP:0000131 | CHEBI:117822 |
| 1 | HP:0000131 | CHEBI:103184 |
| 1 | HP:0000131 | CHEBI:103191 |
| 1 | HP:0000131 | CHEBI:100309 |
| 1 | HP:0000131 | CHEBI:102826 |
| 1 | HP:0000131 | CHEBI:102419 |
| 1 | HP:0000131 | CHEBI:117801 |
| 1 | HP:0000131 | CHEBI:117708 |
| 1 | HP:0000131 | CHEBI:117829 |
| 1 | HP:0000131 | CHEBI:98533 |
| 1 | HP:0000131 | CHEBI:117792 |
| 1 | HP:0000131 | CHEBI:117702 |
| 1 | HP:0000131 | CHEBI:117811 |
| 1 | HP:0000131 | CHEBI:98514 |
| 1 | HP:0000131 | CHEBI:117889 |
| 1 | HP:0000131 | CHEBI:102439 |
| 1 | HP:0000131 | CHEBI:98585 |
| 1 | HP:0000131 | CHEBI:117878 |
| 1 | HP:0000131 | CHEBI:117873 |
| 1 | HP:0000131 | CHEBI:117695 |
| 1 | HP:0000131 | CHEBI:117699 |
| 1 | HP:0000131 | CHEBI:102966 |
| 1 | HP:0000131 | CHEBI:117722 |
| 1 | HP:0000131 | CHEBI:103133 |
| 1 | HP:0000131 | CHEBI:100329 |
| 1 | HP:0000131 | CHEBI:95117 |
| 1 | HP:0000131 | CHEBI:36796 |
| 1 | HP:0000131 | CHEBI:117775 |
| 1 | HP:0000131 | CHEBI:98442 |
| 1 | HP:0000131 | CHEBI:102957 |
| 1 | HP:0000131 | CHEBI:117728 |
| 1 | HP:0000131 | CHEBI:117719 |
| 1 | HP:0000131 | CHEBI:100653 |
| 1 | HP:0000131 | CHEBI:117714 |
| 1 | HP:0000131 | CHEBI:117688 |
| 1 | HP:0000131 | CHEBI:117744 |
| 1 | HP:0000131 | CHEBI:98912 |
| 1 | HP:0000131 | CHEBI:117733 |
| 1 | HP:0000131 | CHEBI:117678 |
| 1 | HP:0000131 | CHEBI:117671 |
| 1 | HP:0000131 | CHEBI:103162 |
| 1 | HP:0000131 | CHEBI:104539 |
| 1 | HP:0000131 | CHEBI:115313 |
| 1 | HP:0000131 | CHEBI:115378 |
| 1 | HP:0000131 | CHEBI:104397 |
| 1 | HP:0000131 | CHEBI:115381 |
| 1 | HP:0000131 | CHEBI:115387 |
| 1 | HP:0000131 | CHEBI:115388 |
| 1 | HP:0000131 | CHEBI:96821 |
| 1 | HP:0000131 | CHEBI:96827 |
| 1 | HP:0000131 | CHEBI:96831 |
| 1 | HP:0000131 | CHEBI:96839 |
| 1 | HP:0000131 | CHEBI:96803 |
| 1 | HP:0000131 | CHEBI:104373 |
| 1 | HP:0000131 | CHEBI:96811 |
| 1 | HP:0000131 | CHEBI:96819 |
| 1 | HP:0000131 | CHEBI:115376 |
| 1 | HP:0000131 | CHEBI:96841 |
| 1 | HP:0000131 | CHEBI:115394 |
| 1 | HP:0000131 | CHEBI:98598 |
| 1 | HP:0000131 | CHEBI:98802 |
| 1 | HP:0000131 | CHEBI:98560 |
| 1 | HP:0000131 | CHEBI:115395 |
| 1 | HP:0000131 | CHEBI:98553 |
| 1 | HP:0000131 | CHEBI:104389 |
| 1 | HP:0000131 | CHEBI:96875 |
| 1 | HP:0000131 | CHEBI:98794 |
| 1 | HP:0000131 | CHEBI:104377 |
| 1 | HP:0000131 | CHEBI:96886 |
| 1 | HP:0000131 | CHEBI:98727 |
| 1 | HP:0000131 | CHEBI:115336 |
| 1 | HP:0000131 | CHEBI:97666 |
| 1 | HP:0000131 | CHEBI:96884 |
| 1 | HP:0000131 | CHEBI:115341 |
| 1 | HP:0000131 | CHEBI:104314 |
| 1 | HP:0000131 | CHEBI:104310 |
| 1 | HP:0000131 | CHEBI:115327 |
| 1 | HP:0000131 | CHEBI:129933 |
| 1 | HP:0000131 | CHEBI:115306 |
| 1 | HP:0000131 | CHEBI:104300 |
| 1 | HP:0000131 | CHEBI:104304 |
| 1 | HP:0000131 | CHEBI:104307 |
| 1 | HP:0000131 | CHEBI:115351 |
| 1 | HP:0000131 | CHEBI:115358 |
| 1 | HP:0000131 | CHEBI:115408 |
| 1 | HP:0000131 | CHEBI:97590 |
| 1 | HP:0000131 | CHEBI:96743 |
| 1 | HP:0000131 | CHEBI:96756 |
| 1 | HP:0000131 | CHEBI:96727 |
| 1 | HP:0000131 | CHEBI:96732 |
| 1 | HP:0000131 | CHEBI:96739 |
| 1 | HP:0000131 | CHEBI:96650 |
| 1 | HP:0000131 | CHEBI:97478 |
| 1 | HP:0000131 | CHEBI:97486 |
| 1 | HP:0000131 | CHEBI:96778 |
| 1 | HP:0000131 | CHEBI:97522 |
| 1 | HP:0000131 | CHEBI:96761 |
| 1 | HP:0000131 | CHEBI:96785 |
| 1 | HP:0000131 | CHEBI:96772 |
| 1 | HP:0000131 | CHEBI:97514 |
| 1 | HP:0000131 | CHEBI:5051 |
| 1 | HP:0000131 | CHEBI:115413 |
| 1 | HP:0000131 | CHEBI:115435 |
| 1 | HP:0000131 | CHEBI:115440 |
| 1 | HP:0000131 | CHEBI:115445 |
| 1 | HP:0000131 | CHEBI:115428 |
| 1 | HP:0000131 | CHEBI:98506 |
| 1 | HP:0000131 | CHEBI:96789 |
| 1 | HP:0000131 | CHEBI:96891 |
| 1 | HP:0000131 | CHEBI:103397 |
| 1 | HP:0000131 | CHEBI:97732 |
| 1 | HP:0000131 | CHEBI:129348 |
| 1 | HP:0000131 | CHEBI:122807 |
| 1 | HP:0000131 | CHEBI:122802 |
| 1 | HP:0000131 | CHEBI:103403 |
| 1 | HP:0000131 | CHEBI:103345 |
| 1 | HP:0000131 | CHEBI:103332 |
| 1 | HP:0000131 | CHEBI:97751 |
| 1 | HP:0000131 | CHEBI:103427 |
| 1 | HP:0000131 | CHEBI:103421 |
| 1 | HP:0000131 | CHEBI:103496 |
| 1 | HP:0000131 | CHEBI:103461 |
| 1 | HP:0000131 | CHEBI:103476 |
| 1 | HP:0000131 | CHEBI:96208 |
| 1 | HP:0000131 | CHEBI:103417 |
| 1 | HP:0000131 | CHEBI:103411 |
| 1 | HP:0000131 | CHEBI:103485 |
| 1 | HP:0000131 | CHEBI:96136 |
| 1 | HP:0000131 | HP:0008675 |
| 1 | HP:0000131 | CHEBI:96002 |
| 1 | HP:0000131 | CHEBI:98764 |
| 1 | HP:0000131 | CHEBI:96050 |
| 1 | HP:0000131 | CHEBI:96058 |
| 1 | HP:0000131 | CHEBI:104346 |
| 1 | HP:0000131 | CHEBI:104351 |
| 1 | HP:0000131 | CHEBI:104359 |
| 1 | HP:0000131 | CHEBI:104338 |
| 1 | HP:0000131 | CHEBI:104324 |
| 1 | HP:0000131 | CHEBI:104330 |
| 1 | HP:0000131 | CHEBI:104270 |
| 1 | HP:0000131 | CHEBI:104284 |
| 1 | HP:0000131 | CHEBI:103389 |
| 1 | HP:0000131 | CHEBI:103384 |
| 1 | HP:0000131 | CHEBI:96168 |
| 1 | HP:0000131 | CHEBI:103371 |
| 1 | HP:0000131 | CHEBI:104288 |
| 1 | HP:0000131 | CHEBI:104296 |
| 1 | HP:0000131 | CHEBI:96154 |
| 1 | HP:0000131 | CHEBI:104257 |
| 1 | HP:0000131 | CHEBI:104266 |
| 1 | HP:0000131 | CHEBI:104281 |
| 1 | HP:0000131 | CHEBI:104276 |
| 1 | HP:0000131 | CHEBI:103351 |
| 1 | HP:0000131 | CHEBI:103365 |
| 1 | HP:0000131 | CHEBI:103360 |
| 1 | HP:0000131 | CHEBI:103359 |
| 1 | HP:0000131 | CHEBI:103355 |
| 1 | HP:0000131 | CHEBI:129738 |
| 1 | HP:0000131 | MONDO:0008364 |
| 1 | HP:0000131 | CHEBI:98206 |
| 1 | HP:0000131 | MONDO:0018076 |
| 1 | HP:0000131 | MONDO:0020642 |
| 1 | HP:0000131 | MONDO:0011382 |
| 1 | HP:0000131 | CHEBI:104802 |
| 1 | HP:0000131 | CHEBI:104446 |
| 1 | HP:0000131 | ECTO:0070158 |
| 1 | HP:0000131 | CHEBI:104309 |
| 1 | HP:0000131 | CHEBI:104458 |
| 1 | HP:0000131 | CHEBI:104828 |
| 1 | HP:0000131 | CHEBI:104824 |
| 1 | HP:0000131 | CHEBI:104831 |
| 1 | HP:0000131 | CHEBI:104806 |
| 1 | HP:0000131 | ECTO:0070128 |
| 1 | HP:0000131 | ECTO:0070174 |
| 1 | HP:0000131 | CHEBI:104810 |
| 1 | HP:0000131 | CHEBI:104815 |
| 1 | HP:0000131 | ECTO:0070036 |
| 1 | HP:0000131 | ECTO:0070138 |
| 1 | HP:0000131 | ECTO:0070159 |
| 1 | HP:0000131 | ECTO:0070022 |
| 1 | HP:0000131 | CHEBI:104842 |
| 1 | HP:0000131 | CHEBI:104846 |
| 1 | HP:0000131 | CHEBI:104701 |
| 1 | HP:0000131 | CHEBI:104711 |
| 1 | HP:0000131 | CHEBI:104793 |
| 1 | HP:0000131 | CHEBI:104798 |
| 1 | HP:0000131 | CHEBI:104773 |
| 1 | HP:0000131 | CHEBI:104440 |
| 1 | HP:0000131 | ECTO:0070121 |
| 1 | HP:0000131 | ECTO:0070187 |
| 1 | HP:0000131 | ECTO:9000123 |
| 1 | HP:0000131 | CHEBI:104506 |
| 1 | HP:0000131 | CHEBI:104582 |
| 1 | HP:0000131 | CHEBI:104591 |
| 1 | HP:0000131 | CHEBI:97882 |
| 1 | HP:0000131 | CHEBI:104569 |
| 1 | HP:0000131 | CHEBI:104570 |
| 1 | HP:0000131 | CHEBI:104404 |
| 1 | HP:0000131 | CHEBI:104408 |
| 1 | HP:0000131 | CHEBI:104411 |
| 1 | HP:0000131 | CHEBI:104419 |
| 1 | HP:0000131 | CHEBI:104461 |
| 1 | HP:0000131 | CHEBI:104467 |
| 1 | HP:0000131 | ECTO:9000299 |
| 1 | HP:0000131 | CHEBI:104429 |
| 1 | HP:0000131 | CHEBI:104481 |
| 1 | HP:0000131 | CHEBI:97947 |
| 1 | HP:0000131 | CHEBI:104497 |
| 1 | HP:0000131 | CHEBI:104745 |
| 1 | HP:0000131 | CHEBI:98917 |
| 1 | HP:0000131 | CHEBI:117682 |
| 1 | HP:0000131 | CHEBI:115153 |
| 1 | HP:0000131 | CHEBI:96635 |
| 1 | HP:0000131 | CHEBI:129779 |
| 1 | HP:0000131 | CHEBI:97346 |
| 1 | HP:0000131 | CHEBI:103096 |
| 1 | HP:0000131 | CHEBI:117619 |
| 1 | HP:0000131 | CHEBI:115210 |
| 1 | HP:0000131 | CHEBI:115214 |
| 1 | HP:0000131 | CHEBI:97377 |
| 1 | HP:0000131 | CHEBI:117627 |
| 1 | HP:0000131 | CHEBI:117623 |
| 1 | HP:0000131 | CHEBI:115223 |
| 1 | HP:0000131 | CHEBI:129871 |
| 1 | HP:0000131 | CHEBI:96531 |
| 1 | HP:0000131 | CHEBI:117610 |
| 1 | HP:0000131 | CHEBI:97358 |
| 1 | HP:0000131 | CHEBI:98405 |
| 1 | HP:0000131 | CHEBI:98410 |
| 1 | HP:0000131 | CHEBI:129703 |
| 1 | HP:0000131 | CHEBI:96688 |
| 1 | HP:0000131 | CHEBI:96680 |
| 1 | HP:0000131 | CHEBI:117706 |
| 1 | HP:0000131 | CHEBI:117703 |
| 1 | HP:0000131 | CHEBI:129719 |
| 1 | HP:0000131 | CHEBI:115129 |
| 1 | HP:0000131 | CHEBI:97414 |
| 1 | HP:0000131 | CHEBI:115140 |
| 1 | HP:0000131 | CHEBI:115145 |
| 1 | HP:0000131 | CHEBI:115195 |
| 1 | HP:0000131 | CHEBI:101938 |
| 1 | HP:0000131 | CHEBI:115179 |
| 1 | HP:0000131 | CHEBI:115183 |
| 1 | HP:0000131 | CHEBI:115186 |
| 1 | HP:0000131 | CHEBI:104699 |
| 1 | HP:0000131 | CHEBI:115249 |
| 1 | HP:0000131 | CHEBI:104663 |
| 1 | HP:0000131 | CHEBI:104752 |
| 1 | HP:0000131 | CHEBI:117596 |
| 1 | HP:0000131 | CHEBI:104738 |
| 1 | HP:0000131 | CHEBI:115279 |
| 1 | HP:0000131 | CHEBI:117652 |
| 1 | HP:0000131 | CHEBI:117662 |
| 1 | HP:0000131 | CHEBI:115202 |
| 1 | HP:0000131 | CHEBI:117655 |
| 1 | HP:0000131 | CHEBI:117650 |
| 1 | HP:0000131 | CHEBI:115238 |
| 1 | HP:0000131 | CHEBI:117607 |
| 1 | HP:0000131 | CHEBI:104684 |
| 1 | HP:0000131 | CHEBI:115244 |
| 1 | HP:0000131 | CHEBI:117665 |
| 1 | HP:0000131 | CHEBI:104651 |
| 1 | HP:0000131 | CHEBI:115282 |
| 1 | HP:0000131 | CHEBI:103002 |
| 1 | HP:0000131 | CHEBI:104674 |
| 1 | HP:0000131 | CHEBI:117646 |
| 1 | HP:0000131 | CHEBI:117642 |
| 1 | HP:0000131 | CHEBI:115252 |
| 1 | HP:0000131 | CHEBI:117639 |
| 1 | HP:0000131 | CHEBI:115263 |
| 1 | HP:0000131 | CHEBI:115267 |
| 1 | HP:0000131 | CHEBI:104640 |
| 1 | HP:0000131 | CHEBI:115271 |
| 1 | HP:0000131 | CHEBI:117633 |
| 1 | HP:0000131 | CHEBI:117630 |
| 1 | HP:0000131 | CHEBI:104646 |
| 1 | HP:0000131 | CHEBI:116037 |
| 1 | HP:0000131 | CHEBI:128394 |
| 1 | HP:0000131 | CHEBI:116061 |
| 1 | HP:0000131 | ECTO:0070026 |
| 1 | HP:0000131 | CHEBI:103525 |
| 1 | HP:0000131 | ECTO:0070119 |
| 1 | HP:0000131 | ECTO:0070048 |
| 1 | HP:0000131 | ECTO:0070000 |
| 1 | HP:0000131 | ECTO:0070151 |
| 1 | HP:0000131 | ECTO:9000192 |
| 1 | HP:0000131 | ECTO:9000143 |
| 1 | HP:0000131 | CHEBI:103455 |
| 1 | HP:0000131 | CHEBI:103460 |
| 1 | HP:0000131 | ECTO:0070109 |
| 1 | HP:0000131 | ECTO:0070062 |
| 1 | HP:0000131 | CHEBI:103515 |
| 1 | HP:0000131 | MONDO:0005406 |
| 1 | HP:0000131 | CHEBI:117767 |
| 1 | HP:0000131 | MONDO:0008228 |
| 1 | HP:0000131 | CHEBI:103507 |
| 1 | HP:0000131 | CHEBI:103503 |
| 1 | HP:0000131 | ECTO:0070157 |
| 1 | HP:0000131 | MONDO:0007915 |
| 1 | HP:0000131 | CHEBI:103530 |
| 1 | HP:0000131 | MONDO:0005133 |
| 1 | HP:0000131 | CHEBI:103541 |
| 1 | HP:0000131 | CHEBI:103546 |
| 1 | HP:0000131 | ECTO:0070125 |
| 1 | HP:0000131 | CHEBI:103464 |
| 1 | HP:0000131 | CHEBI:103401 |
| 1 | HP:0000131 | CHEBI:100716 |
| 1 | HP:0000131 | CHEBI:103410 |
| 1 | HP:0000131 | CHEBI:103415 |
| 1 | HP:0000131 | CHEBI:103425 |
| 1 | HP:0000131 | CHEBI:98343 |
| 1 | HP:0000131 | CHEBI:98323 |
| 1 | HP:0000131 | CHEBI:103409 |
| 1 | HP:0000131 | CHEBI:103484 |
| 1 | HP:0000131 | CHEBI:103480 |
| 1 | HP:0000131 | CHEBI:103431 |
| 1 | HP:0000131 | CHEBI:100765 |
| 1 | HP:0000131 | CHEBI:103442 |
| 1 | HP:0000131 | CHEBI:98213 |
| 1 | HP:0000131 | CHEBI:117796 |
| 1 | HP:0000131 | CHEBI:117803 |
| 1 | HP:0000131 | CHEBI:100840 |
| 1 | HP:0000131 | CHEBI:117813 |
| 1 | HP:0000131 | CHEBI:117820 |
| 1 | HP:0000131 | CHEBI:117825 |
| 1 | HP:0000131 | CHEBI:117772 |
| 1 | HP:0000131 | CHEBI:117778 |
| 1 | HP:0000131 | CHEBI:103137 |
| 1 | HP:0000131 | CHEBI:117781 |
| 1 | HP:0000131 | CHEBI:117789 |
| 1 | HP:0000131 | CHEBI:117726 |
| 1 | HP:0000131 | CHEBI:103199 |
| 1 | HP:0000131 | CHEBI:117731 |
| 1 | HP:0000131 | CHEBI:117737 |
| 1 | HP:0000131 | CHEBI:117742 |
| 1 | HP:0000131 | CHEBI:117748 |
| 1 | HP:0000131 | CHEBI:117710 |
| 1 | HP:0000131 | CHEBI:117717 |
| 1 | HP:0000131 | CHEBI:117881 |
| 1 | HP:0000131 | CHEBI:98536 |
| 1 | HP:0000131 | CHEBI:98762 |
| 1 | HP:0000131 | CHEBI:98770 |
| 1 | HP:0000131 | CHEBI:98747 |
| 1 | HP:0000131 | CHEBI:129008 |
| 1 | HP:0000131 | CHEBI:98726 |
| 1 | HP:0000131 | CHEBI:98729 |
| 1 | HP:0000131 | CHEBI:98564 |
| 1 | HP:0000131 | CHEBI:98577 |
| 1 | HP:0000131 | CHEBI:98732 |
| 1 | HP:0000131 | CHEBI:98702 |
| 1 | HP:0000131 | CHEBI:103338 |
| 1 | HP:0000131 | CHEBI:98396 |
| 1 | HP:0000131 | CHEBI:6993 |
| 1 | HP:0000131 | CHEBI:103392 |
| 1 | HP:0000131 | CHEBI:129023 |
| 1 | HP:0000131 | CHEBI:124024 |
| 1 | HP:0000131 | CHEBI:124015 |
| 1 | HP:0000131 | CHEBI:98698 |
| 1 | HP:0000131 | CHEBI:124089 |
| 1 | HP:0000131 | CHEBI:103369 |
| 1 | HP:0000131 | CHEBI:103375 |
| 1 | HP:0000131 | CHEBI:103382 |
| 1 | HP:0000131 | CHEBI:129056 |
| 1 | HP:0000131 | CHEBI:124068 |
| 1 | HP:0000131 | CHEBI:103354 |
| 1 | HP:0000131 | CHEBI:122585 |
| 1 | HP:0000131 | CHEBI:122580 |
| 1 | HP:0000131 | CHEBI:122586 |
| 1 | HP:0000131 | CHEBI:123674 |
| 1 | HP:0000131 | CHEBI:123686 |
| 1 | HP:0000131 | CHEBI:122613 |
| 1 | HP:0000131 | CHEBI:122609 |
| 1 | HP:0000131 | CHEBI:122606 |
| 1 | HP:0000131 | CHEBI:122602 |
| 1 | HP:0000131 | CHEBI:123668 |
| 1 | HP:0000131 | CHEBI:123640 |
| 1 | HP:0000131 | CHEBI:122516 |
| 1 | HP:0000131 | CHEBI:123788 |
| 1 | HP:0000131 | CHEBI:123752 |
| 1 | HP:0000131 | CHEBI:123745 |
| 1 | HP:0000131 | CHEBI:122546 |
| 1 | HP:0000131 | CHEBI:122538 |
| 1 | HP:0000131 | CHEBI:122535 |
| 1 | HP:0000131 | CHEBI:122557 |
| 1 | HP:0000131 | CHEBI:122570 |
| 1 | HP:0000131 | CHEBI:122575 |
| 1 | HP:0000131 | CHEBI:123701 |
| 1 | HP:0000131 | CHEBI:123931 |
| 1 | HP:0000131 | CHEBI:123934 |
| 1 | HP:0000131 | CHEBI:123921 |
| 1 | HP:0000131 | CHEBI:123907 |
| 1 | HP:0000131 | CHEBI:122693 |
| 1 | HP:0000131 | CHEBI:123952 |
| 1 | HP:0000131 | CHEBI:122678 |
| 1 | HP:0000131 | CHEBI:122663 |
| 1 | HP:0000131 | CHEBI:122649 |
| 1 | HP:0000131 | CHEBI:122689 |
| 1 | HP:0000131 | CHEBI:122646 |
| 1 | HP:0000131 | CHEBI:123997 |
| 1 | HP:0000131 | CHEBI:122638 |
| 1 | HP:0000131 | CHEBI:122633 |
| 1 | HP:0000131 | CHEBI:123975 |
| 1 | HP:0000131 | CHEBI:123982 |
| 1 | HP:0000131 | CHEBI:122624 |
| 1 | HP:0000131 | CHEBI:122659 |
| 1 | HP:0000131 | ECTO:9002165 |
| 1 | HP:0000131 | ECTO:7000150 |
| 1 | HP:0000131 | ECTO:9000057 |
| 1 | HP:0000131 | ECTO:0500021 |
| 1 | HP:0000131 | ECTO:9000024 |
| 1 | HP:0000131 | ECTO:0500009 |
| 1 | HP:0000131 | ECTO:7000151 |
| 1 | HP:0000131 | ECTO:9000079 |
| 1 | HP:0000131 | ECTO:0001571 |
| 1 | HP:0000131 | ECTO:7000153 |
| 1 | HP:0000131 | ECTO:9000020 |
| 1 | HP:0000131 | ECTO:9000076 |
| 1 | HP:0000131 | CHEBI:117847 |
| 1 | HP:0000131 | CHEBI:117870 |
| 1 | HP:0000131 | CHEBI:117876 |
| 1 | HP:0000131 | CHEBI:117844 |
| 1 | HP:0000131 | CHEBI:117840 |
| 1 | HP:0000131 | CHEBI:117834 |
| 1 | HP:0000131 | ECTO:9001465 |
| 1 | HP:0000131 | ECTO:9000068 |
| 1 | HP:0000131 | CHEBI:123721 |
| 1 | HP:0000131 | CHEBI:123795 |
| 1 | HP:0000131 | ECTO:9000032 |
| 1 | HP:0000131 | ECTO:7000152 |
| 1 | HP:0000131 | CHEBI:98189 |
| 1 | HP:0000131 | CHEBI:103729 |
| 1 | HP:0000131 | CHEBI:104626 |
| 1 | HP:0000131 | CHEBI:96951 |
| 1 | HP:0000131 | CHEBI:98539 |
| 1 | HP:0000131 | CHEBI:97332 |
| 1 | HP:0000131 | CHEBI:98519 |
| 1 | HP:0000131 | CHEBI:96923 |
| 1 | HP:0000131 | CHEBI:96929 |
| 1 | HP:0000131 | CHEBI:103363 |
| 1 | HP:0000131 | CHEBI:101037 |
| 1 | HP:0000131 | CHEBI:98692 |
| 1 | HP:0000131 | CHEBI:98548 |
| 1 | HP:0000131 | CHEBI:97415 |
| 1 | HP:0000131 | CHEBI:103752 |
| 1 | HP:0000131 | CHEBI:103696 |
| 1 | HP:0000131 | CHEBI:98152 |
| 1 | HP:0000131 | CHEBI:103676 |
| 1 | HP:0000131 | CHEBI:98767 |
| 1 | HP:0000131 | CHEBI:98774 |
| 1 | HP:0000131 | CHEBI:104633 |
| 1 | HP:0000131 | CHEBI:104693 |
| 1 | HP:0000131 | CHEBI:104638 |
| 1 | HP:0000131 | CHEBI:97835 |
| 1 | HP:0000131 | CHEBI:104664 |
| 1 | HP:0000131 | CHEBI:103688 |
| 1 | HP:0000131 | CHEBI:104670 |
| 1 | HP:0000131 | CHEBI:103683 |
| 1 | HP:0000131 | CHEBI:128512 |
| 1 | HP:0000131 | CHEBI:96960 |
| 1 | HP:0000131 | CHEBI:103679 |
| 1 | HP:0000131 | CHEBI:104647 |
| 1 | HP:0000131 | CHEBI:104652 |
| 1 | HP:0000131 | CHEBI:103379 |
| 1 | HP:0000131 | CHEBI:104614 |
| 1 | HP:0000131 | CHEBI:104610 |
| 1 | HP:0000131 | CHEBI:103739 |
| 1 | HP:0000131 | CHEBI:103731 |
| 1 | HP:0000131 | CHEBI:96941 |
| 1 | HP:0000131 | CHEBI:103374 |
| 1 | HP:0000131 | CHEBI:104680 |
| 1 | HP:0000131 | CHEBI:103717 |
| 1 | HP:0000131 | CHEBI:34349 |
| 1 | HP:0000131 | CHEBI:98183 |
| 1 | HP:0000131 | CHEBI:122814 |
| 1 | HP:0000131 | CHEBI:103617 |
| 1 | HP:0000131 | CHEBI:104380 |
| 1 | HP:0000131 | CHEBI:128738 |
| 1 | HP:0000131 | CHEBI:104301 |
| 1 | HP:0000131 | CHEBI:128727 |
| 1 | HP:0000131 | CHEBI:104311 |
| 1 | HP:0000131 | CHEBI:104315 |
| 1 | HP:0000131 | CHEBI:104317 |
| 1 | HP:0000131 | CHEBI:95307 |
| 1 | HP:0000131 | CHEBI:128744 |
| 1 | HP:0000131 | CHEBI:98062 |
| 1 | HP:0000131 | CHEBI:115952 |
| 1 | HP:0000131 | CHEBI:115960 |
| 1 | HP:0000131 | CHEBI:97639 |
| 1 | HP:0000131 | CHEBI:115933 |
| 1 | HP:0000131 | CHEBI:104347 |
| 1 | HP:0000131 | CHEBI:115942 |
| 1 | HP:0000131 | CHEBI:115946 |
| 1 | HP:0000131 | CHEBI:97614 |
| 1 | HP:0000131 | CHEBI:104325 |
| 1 | HP:0000131 | CHEBI:104366 |
| 1 | HP:0000131 | CHEBI:98795 |
| 1 | HP:0000131 | CHEBI:98784 |
| 1 | HP:0000131 | CHEBI:97519 |
| 1 | HP:0000131 | CHEBI:103620 |
| 1 | HP:0000131 | CHEBI:103659 |
| 1 | HP:0000131 | CHEBI:103663 |
| 1 | HP:0000131 | CHEBI:103631 |
| 1 | HP:0000131 | CHEBI:103638 |
| 1 | HP:0000131 | CHEBI:95242 |
| 1 | HP:0000131 | CHEBI:98100 |
| 1 | HP:0000131 | CHEBI:97533 |
| 1 | HP:0000131 | CHEBI:103570 |
| 1 | HP:0000131 | CHEBI:104385 |
| 1 | HP:0000131 | CHEBI:103576 |
| 1 | HP:0000131 | CHEBI:128807 |
| 1 | HP:0000131 | CHEBI:103583 |
| 1 | HP:0000131 | CHEBI:103550 |
| 1 | HP:0000131 | CHEBI:103557 |
| 1 | HP:0000131 | CHEBI:103565 |
| 1 | HP:0000131 | CHEBI:103594 |
| 1 | HP:0000131 | CHEBI:104390 |
| 1 | HP:0000131 | CHEBI:104660 |
| 1 | HP:0000131 | CHEBI:96966 |
| 1 | HP:0000131 | CHEBI:103349 |
| 1 | HP:0000131 | CHEBI:104540 |
| 1 | HP:0000131 | CHEBI:104405 |
| 1 | HP:0000131 | CHEBI:104436 |
| 1 | HP:0000131 | CHEBI:115759 |
| 1 | HP:0000131 | CHEBI:103473 |
| 1 | HP:0000131 | CHEBI:104425 |
| 1 | HP:0000131 | CHEBI:103490 |
| 1 | HP:0000131 | CHEBI:97908 |
| 1 | HP:0000131 | CHEBI:101120 |
| 1 | HP:0000131 | CHEBI:104499 |
| 1 | HP:0000131 | CHEBI:104784 |
| 1 | HP:0000131 | CHEBI:104789 |
| 1 | HP:0000131 | CHEBI:101126 |
| 1 | HP:0000131 | CHEBI:104790 |
| 1 | HP:0000131 | CHEBI:104799 |
| 1 | HP:0000131 | CHEBI:103598 |
| 1 | HP:0000131 | CHEBI:103590 |
| 1 | HP:0000131 | CHEBI:104763 |
| 1 | HP:0000131 | CHEBI:104769 |
| 1 | HP:0000131 | CHEBI:103561 |
| 1 | HP:0000131 | CHEBI:103494 |
| 1 | HP:0000131 | CHEBI:104856 |
| 1 | HP:0000131 | CHEBI:104852 |
| 1 | HP:0000131 | CHEBI:104829 |
| 1 | HP:0000131 | CHEBI:104833 |
| 1 | HP:0000131 | CHEBI:104826 |
| 1 | HP:0000131 | CHEBI:104470 |
| 1 | HP:0000131 | CHEBI:104474 |
| 1 | HP:0000131 | CHEBI:98418 |
| 1 | HP:0000131 | CHEBI:104441 |
| 1 | HP:0000131 | CHEBI:104447 |
| 1 | HP:0000131 | CHEBI:104452 |
| 1 | HP:0000131 | CHEBI:104448 |
| 1 | HP:0000131 | CHEBI:104413 |
| 1 | HP:0000131 | CHEBI:104825 |
| 1 | HP:0000131 | CHEBI:104807 |
| 1 | HP:0000131 | CHEBI:104803 |
| 1 | HP:0000131 | CHEBI:104862 |
| 1 | HP:0000131 | CHEBI:104866 |
| 1 | HP:0000131 | CHEBI:103435 |
| 1 | HP:0000131 | CHEBI:104469 |
| 1 | HP:0000131 | CHEBI:103441 |
| 1 | HP:0000131 | CHEBI:103414 |
| 1 | HP:0000131 | CHEBI:103554 |
| 1 | HP:0000131 | CHEBI:104779 |
| 1 | HP:0000131 | CHEBI:104732 |
| 1 | HP:0000131 | CHEBI:103667 |
| 1 | HP:0000131 | CHEBI:103655 |
| 1 | HP:0000131 | CHEBI:104722 |
| 1 | HP:0000131 | CHEBI:104739 |
| 1 | HP:0000131 | CHEBI:103624 |
| 1 | HP:0000131 | CHEBI:104589 |
| 1 | HP:0000131 | CHEBI:104583 |
| 1 | HP:0000131 | CHEBI:104590 |
| 1 | HP:0000131 | CHEBI:103611 |
| 1 | HP:0000131 | CHEBI:104564 |
| 1 | HP:0000131 | CHEBI:104536 |
| 1 | HP:0000131 | CHEBI:103337 |
| 1 | HP:0000131 | CHEBI:104527 |
| 1 | HP:0000131 | CHEBI:104524 |
| 1 | HP:0000131 | HP:0012886 |
| 1 | HP:0000131 | CHEBI:104552 |
| 1 | HP:0000131 | CHEBI:103342 |
| 1 | HP:0000131 | CHEBI:104548 |
| 1 | HP:0000131 | CHEBI:97856 |
| 1 | HP:0000131 | CHEBI:97867 |
| 1 | HP:0000131 | CHEBI:103587 |
| 1 | HP:0000131 | CHEBI:104702 |
| 1 | HP:0000131 | CHEBI:103424 |
| 1 | HP:0000131 | CHEBI:103581 |
| 1 | HP:0000131 | CHEBI:103573 |
| 1 | HP:0000131 | CHEBI:104515 |
| 1 | HP:0000131 | CHEBI:104503 |
| 1 | HP:0000131 | CHEBI:103400 |
| 1 | HP:0000131 | CHEBI:103407 |
| 1 | HP:0000131 | CHEBI:104706 |
| 1 | HP:0000131 | CHEBI:128568 |
| 1 | HP:0000131 | CHEBI:103635 |
| 1 | HP:0000131 | CHEBI:104712 |
| 1 | HP:0000131 | CHEBI:104716 |
| 1 | HP:0000131 | CHEBI:96903 |
| 1 | HP:0000131 | CHEBI:103647 |
| 1 | HP:0000131 | CHEBI:103640 |
| 1 | HP:0000131 | CHEBI:103396 |
| 1 | HP:0000131 | CHEBI:96907 |
| 1 | HP:0000131 | CHEBI:104571 |
| 1 | HP:0000131 | CHEBI:122818 |
| 1 | HP:0000131 | CHEBI:97627 |
| 1 | HP:0000131 | CHEBI:95491 |
| 1 | HP:0000131 | CHEBI:117593 |
| 1 | HP:0000131 | CHEBI:124023 |
| 1 | HP:0000131 | CHEBI:124046 |
| 1 | HP:0000131 | CHEBI:124082 |
| 1 | HP:0000131 | CHEBI:103775 |
| 1 | HP:0000131 | CHEBI:103771 |
| 1 | HP:0000131 | CHEBI:103728 |
| 1 | HP:0000131 | CHEBI:103710 |
| 1 | HP:0000131 | CHEBI:115868 |
| 1 | HP:0000131 | CHEBI:98558 |
| 1 | HP:0000131 | CHEBI:102208 |
| 1 | HP:0000131 | CHEBI:115800 |
| 1 | HP:0000131 | CHEBI:115807 |
| 1 | HP:0000131 | CHEBI:98125 |
| 1 | HP:0000131 | CHEBI:95412 |
| 1 | HP:0000131 | CHEBI:95427 |
| 1 | HP:0000131 | CHEBI:95403 |
| 1 | HP:0000131 | HP:0002625 |
| 1 | HP:0000131 | CHEBI:102194 |
| 1 | HP:0000131 | CHEBI:115882 |
| 1 | HP:0000131 | CHEBI:124002 |
| 1 | HP:0000131 | CHEBI:101236 |
| 1 | HP:0000131 | CHEBI:95597 |
| 1 | HP:0000131 | CHEBI:98658 |
| 1 | HP:0000131 | CHEBI:117632 |
| 1 | HP:0000131 | CHEBI:98645 |
| 1 | HP:0000131 | CHEBI:115834 |
| 1 | HP:0000131 | CHEBI:117648 |
| 1 | HP:0000131 | CHEBI:128124 |
| 1 | HP:0000131 | CHEBI:115846 |
| 1 | HP:0000131 | CHEBI:128144 |
| 1 | HP:0000131 | CHEBI:115812 |
| 1 | HP:0000131 | CHEBI:124014 |
| 1 | HP:0000131 | CHEBI:98047 |
| 1 | HP:0000131 | CHEBI:115824 |
| 1 | HP:0000131 | CHEBI:115875 |
| 1 | HP:0000131 | CHEBI:95583 |
| 1 | HP:0000131 | CHEBI:95556 |
| 1 | HP:0000131 | CHEBI:103705 |
| 1 | HP:0000131 | CHEBI:102023 |
| 1 | HP:0000131 | CHEBI:103767 |
| 1 | HP:0000131 | CHEBI:95490 |
| 1 | HP:0000131 | CHEBI:95496 |
| 1 | HP:0000131 | CHEBI:122806 |
| 1 | HP:0000131 | CHEBI:128008 |
| 1 | HP:0000131 | CHEBI:128335 |
| 1 | HP:0000131 | CHEBI:116071 |
| 1 | HP:0000131 | CHEBI:117656 |
| 1 | HP:0000131 | CHEBI:122619 |
| 1 | HP:0000131 | CHEBI:122612 |
| 1 | HP:0000131 | CHEBI:122601 |
| 1 | HP:0000131 | CHEBI:116005 |
| 1 | HP:0000131 | CHEBI:116008 |
| 1 | HP:0000131 | CHEBI:10650 |
| 1 | HP:0000131 | CHEBI:116010 |
| 1 | HP:0000131 | CHEBI:116018 |
| 1 | HP:0000131 | CHEBI:117624 |
| 1 | HP:0000131 | CHEBI:95532 |
| 1 | HP:0000131 | CHEBI:117612 |
| 1 | HP:0000131 | CHEBI:128395 |
| 1 | HP:0000131 | CHEBI:116048 |
| 1 | HP:0000131 | CHEBI:122599 |
| 1 | HP:0000131 | CHEBI:116054 |
| 1 | HP:0000131 | CHEBI:116057 |
| 1 | HP:0000131 | CHEBI:122699 |
| 1 | HP:0000131 | CHEBI:122662 |
| 1 | HP:0000131 | CHEBI:116068 |
| 1 | HP:0000131 | CHEBI:117647 |
| 1 | HP:0000131 | CHEBI:115799 |
| 1 | HP:0000131 | CHEBI:117598 |
| 1 | HP:0000131 | CHEBI:115771 |
| 1 | HP:0000131 | CHEBI:101139 |
| 1 | HP:0000131 | CHEBI:98155 |
| 1 | HP:0000131 | CHEBI:101192 |
| 1 | HP:0000131 | CHEBI:95435 |
| 1 | HP:0000131 | CHEBI:95430 |
| 1 | HP:0000131 | CHEBI:98940 |
| 1 | HP:0000131 | CHEBI:117666 |
| 1 | HP:0000131 | CHEBI:122632 |
| 1 | HP:0000131 | CHEBI:122629 |
| 1 | HP:0000131 | CHEBI:122623 |
| 1 | HP:0000131 | CHEBI:122658 |
| 1 | HP:0000131 | CHEBI:98443 |
| 1 | HP:0000131 | CHEBI:103026 |
| 1 | HP:0000131 | CHEBI:98473 |
| 1 | HP:0000131 | CHEBI:122674 |
| 1 | HP:0000131 | CHEBI:98654 |
| 1 | HP:0000131 | CHEBI:102081 |
| 1 | HP:0000131 | CHEBI:98099 |
| 1 | HP:0000131 | CHEBI:117669 |
| 1 | HP:0000131 | CHEBI:104246 |
| 1 | HP:0000131 | CHEBI:122786 |
| 1 | HP:0000131 | CHEBI:103589 |
| 1 | HP:0000131 | CHEBI:103582 |
| 1 | HP:0000131 | CHEBI:122760 |
| 1 | HP:0000131 | CHEBI:104250 |
| 1 | HP:0000131 | CHEBI:104255 |
| 1 | HP:0000131 | CHEBI:104258 |
| 1 | HP:0000131 | CHEBI:122773 |
| 1 | HP:0000131 | CHEBI:122742 |
| 1 | HP:0000131 | CHEBI:122754 |
| 1 | HP:0000131 | CHEBI:104297 |
| 1 | HP:0000131 | CHEBI:95428 |
| 1 | HP:0000131 | CHEBI:122750 |
| 1 | HP:0000131 | CHEBI:122770 |
| 1 | HP:0000131 | CHEBI:122776 |
| 1 | HP:0000131 | CHEBI:122769 |
| 1 | HP:0000131 | CHEBI:122763 |
| 1 | HP:0000131 | CHEBI:97770 |
| 1 | HP:0000131 | CHEBI:122783 |
| 1 | HP:0000131 | CHEBI:103827 |
| 1 | HP:0000131 | CHEBI:95399 |
| 1 | HP:0000131 | CHEBI:115920 |
| 1 | HP:0000131 | CHEBI:122821 |
| 1 | HP:0000131 | CHEBI:115973 |
| 1 | HP:0000131 | CHEBI:10125 |
| 1 | HP:0000131 | CHEBI:115980 |
| 1 | HP:0000131 | CHEBI:122725 |
| 1 | HP:0000131 | CHEBI:97697 |
| 1 | HP:0000131 | CHEBI:122710 |
| 1 | HP:0000131 | CHEBI:115914 |
| 1 | HP:0000131 | CHEBI:102203 |
| 1 | HP:0000131 | CHEBI:103556 |
| 1 | HP:0000131 | CHEBI:115928 |
| 1 | HP:0000131 | CHEBI:98840 |
| 1 | HP:0000131 | CHEBI:97711 |
| 1 | HP:0000131 | CHEBI:115906 |
| 1 | HP:0000131 | CHEBI:104271 |
| 1 | HP:0000131 | CHEBI:103599 |
| 1 | HP:0000131 | CHEBI:103592 |
| 1 | HP:0000131 | CHEBI:103563 |
| 1 | HP:0000131 | CHEBI:122791 |
| 1 | HP:0000131 | CHEBI:122793 |
| 1 | HP:0000131 | CHEBI:128633 |
| 1 | HP:0000131 | CHEBI:128181 |
| 1 | HP:0000131 | CHEBI:95434 |
| 1 | HP:0000131 | CHEBI:101275 |
| 1 | HP:0000131 | CHEBI:117680 |
| 1 | HP:0000131 | CHEBI:117684 |
| 1 | HP:0000131 | CHEBI:98916 |
| 1 | HP:0000131 | CHEBI:101287 |
| 1 | HP:0000131 | CHEBI:102155 |
| 1 | HP:0000131 | HP:0011127 |
| 1 | HP:0000131 | CHEBI:103094 |
| 1 | HP:0000131 | CHEBI:117675 |
| 1 | HP:0000131 | CHEBI:128173 |
| 1 | HP:0000131 | CHEBI:117620 |
| 1 | HP:0000131 | CHEBI:117606 |
| 1 | HP:0000131 | CHEBI:117609 |
| 1 | HP:0000131 | CHEBI:117653 |
| 1 | HP:0000131 | CHEBI:117659 |
| 1 | HP:0000131 | CHEBI:101293 |
| 1 | HP:0000131 | CHEBI:101297 |
| 1 | HP:0000131 | CHEBI:98906 |
| 1 | HP:0000131 | CHEBI:103613 |
| 1 | HP:0000131 | CHEBI:117705 |
| 1 | HP:0000131 | CHEBI:103669 |
| 1 | HP:0000131 | CHEBI:122788 |
| 1 | HP:0000131 | CHEBI:103642 |
| 1 | HP:0000131 | CHEBI:128674 |
| 1 | HP:0000131 | CHEBI:103637 |
| 1 | HP:0000131 | CHEBI:122713 |
| 1 | HP:0000131 | CHEBI:95476 |
| 1 | HP:0000131 | CHEBI:117693 |
| 1 | HP:0000131 | CHEBI:98924 |
| 1 | HP:0000131 | CHEBI:122702 |
| 1 | HP:0000131 | CHEBI:122812 |
| 1 | HP:0000131 | CHEBI:102118 |
| 1 | HP:0000131 | CHEBI:103662 |
| 1 | HP:0000131 | CHEBI:122735 |
| 1 | HP:0000131 | CHEBI:103657 |
| 1 | HP:0000131 | CHEBI:103601 |
| 1 | HP:0000131 | CHEBI:122732 |
| 1 | HP:0000131 | CHEBI:122804 |
| 1 | HP:0000131 | CHEBI:103608 |
| 1 | HP:0000131 | CHEBI:104468 |
| 1 | HP:0000131 | CHEBI:104808 |
| 1 | HP:0000131 | CHEBI:104804 |
| 1 | HP:0000131 | CHEBI:104312 |
| 1 | HP:0000131 | CHEBI:104342 |
| 1 | HP:0000131 | CHEBI:129150 |
| 1 | HP:0000131 | CHEBI:117696 |
| 1 | HP:0000131 | CHEBI:98928 |
| 1 | HP:0000131 | CHEBI:129175 |
| 1 | HP:0000131 | CHEBI:103051 |
| 1 | HP:0000131 | CHEBI:98933 |
| 1 | HP:0000131 | CHEBI:98939 |
| 1 | HP:0000131 | CHEBI:104348 |
| 1 | HP:0000131 | CHEBI:104332 |
| 1 | HP:0000131 | CHEBI:97623 |
| 1 | HP:0000131 | CHEBI:104326 |
| 1 | HP:0000131 | CHEBI:97612 |
| 1 | HP:0000131 | CHEBI:97641 |
| 1 | HP:0000131 | CHEBI:104353 |
| 1 | HP:0000131 | CHEBI:104412 |
| 1 | HP:0000131 | CHEBI:98900 |
| 1 | HP:0000131 | CHEBI:122718 |
| 1 | HP:0000131 | CHEBI:122707 |
| 1 | HP:0000131 | CHEBI:104398 |
| 1 | HP:0000131 | CHEBI:103088 |
| 1 | HP:0000131 | CHEBI:122738 |
| 1 | HP:0000131 | CHEBI:117618 |
| 1 | HP:0000131 | CHEBI:122734 |
| 1 | HP:0000131 | CHEBI:122728 |
| 1 | HP:0000131 | CHEBI:122724 |
| 1 | HP:0000131 | CHEBI:104391 |
| 1 | HP:0000131 | CHEBI:122721 |
| 1 | HP:0000131 | CHEBI:104374 |
| 1 | HP:0000131 | CHEBI:103031 |
| 1 | HP:0000131 | CHEBI:122753 |
| 1 | HP:0000131 | CHEBI:122746 |
| 1 | HP:0000131 | CHEBI:104305 |
| 1 | HP:0000131 | CHEBI:122772 |
| 1 | HP:0000131 | CHEBI:122767 |
| 1 | HP:0000131 | CHEBI:122796 |
| 1 | HP:0000131 | CHEBI:122782 |
| 1 | HP:0000131 | CHEBI:117681 |
| 1 | HP:0000131 | CHEBI:117689 |
| 1 | HP:0000131 | CHEBI:122816 |
| 1 | HP:0000131 | CHEBI:129253 |
| 1 | HP:0000131 | CHEBI:129297 |
| 1 | HP:0000131 | CHEBI:104298 |
| 1 | HP:0000131 | CHEBI:97731 |
| 1 | HP:0000131 | CHEBI:98808 |
| 1 | HP:0000131 | CHEBI:104290 |
| 1 | HP:0000131 | CHEBI:104259 |
| 1 | HP:0000131 | CHEBI:104251 |
| 1 | HP:0000131 | CHEBI:97700 |
| 1 | HP:0000131 | CHEBI:104277 |
| 1 | HP:0000131 | CHEBI:104282 |
| 1 | HP:0000131 | CHEBI:129211 |
| 1 | HP:0000131 | CHEBI:117621 |
| 1 | HP:0000131 | CHEBI:117626 |
| 1 | HP:0000131 | CHEBI:122685 |
| 1 | HP:0000131 | CHEBI:104641 |
| 1 | HP:0000131 | CHEBI:104675 |
| 1 | HP:0000131 | CHEBI:98270 |
| 1 | HP:0000131 | CHEBI:117816 |
| 1 | HP:0000131 | CHEBI:117807 |
| 1 | HP:0000131 | CHEBI:103172 |
| 1 | HP:0000131 | CHEBI:104733 |
| 1 | HP:0000131 | CHEBI:117795 |
| 1 | HP:0000131 | CHEBI:103801 |
| 1 | HP:0000131 | CHEBI:103806 |
| 1 | HP:0000131 | CHEBI:104637 |
| 1 | HP:0000131 | CHEBI:104603 |
| 1 | HP:0000131 | CHEBI:104618 |
| 1 | HP:0000131 | CHEBI:104753 |
| 1 | HP:0000131 | CHEBI:104743 |
| 1 | HP:0000131 | CHEBI:117736 |
| 1 | HP:0000131 | CHEBI:117747 |
| 1 | HP:0000131 | CHEBI:104703 |
| 1 | HP:0000131 | CHEBI:104707 |
| 1 | HP:0000131 | HP:0002092 |
| 1 | HP:0000131 | CHEBI:104653 |
| 1 | HP:0000131 | CHEBI:104659 |
| 1 | HP:0000131 | CHEBI:117896 |
| 1 | HP:0000131 | CHEBI:122697 |
| 1 | HP:0000131 | CHEBI:122698 |
| 1 | HP:0000131 | CHEBI:122668 |
| 1 | HP:0000131 | CHEBI:50275 |
| 1 | HP:0000131 | CHEBI:122660 |
| 1 | HP:0000131 | CHEBI:122672 |
| 1 | HP:0000131 | CHEBI:122645 |
| 1 | HP:0000131 | CHEBI:117892 |
| 1 | HP:0000131 | CHEBI:122652 |
| 1 | HP:0000131 | CHEBI:117887 |
| 1 | HP:0000131 | CHEBI:122621 |
| 1 | HP:0000131 | CHEBI:98159 |
| 1 | HP:0000131 | CHEBI:122631 |
| 1 | HP:0000131 | CHEBI:122637 |
| 1 | HP:0000131 | CHEBI:117879 |
| 1 | HP:0000131 | CHEBI:104694 |
| 1 | HP:0000131 | CHEBI:104686 |
| 1 | HP:0000131 | CHEBI:117880 |
| 1 | HP:0000131 | CHEBI:104547 |
| 1 | HP:0000131 | CHEBI:104551 |
| 1 | HP:0000131 | CHEBI:104559 |
| 1 | HP:0000131 | CHEBI:104482 |
| 1 | HP:0000131 | CHEBI:104502 |
| 1 | HP:0000131 | CHEBI:104507 |
| 1 | HP:0000131 | CHEBI:104514 |
| 1 | HP:0000131 | CHEBI:104518 |
| 1 | HP:0000131 | CHEBI:103502 |
| 1 | HP:0000131 | CHEBI:101615 |
| 1 | HP:0000131 | CHEBI:103540 |
| 1 | HP:0000131 | CHEBI:98394 |
| 1 | HP:0000131 | CHEBI:104486 |
| 1 | HP:0000131 | CHEBI:103117 |
| 1 | HP:0000131 | CHEBI:103519 |
| 1 | HP:0000131 | CHEBI:104498 |
| 1 | HP:0000131 | CHEBI:103514 |
| 1 | HP:0000131 | CHEBI:104857 |
| 1 | HP:0000131 | CHEBI:103524 |
| 1 | HP:0000131 | CHEBI:104848 |
| 1 | HP:0000131 | CHEBI:104435 |
| 1 | HP:0000131 | CHEBI:104811 |
| 1 | HP:0000131 | CHEBI:104780 |
| 1 | HP:0000131 | CHEBI:103112 |
| 1 | HP:0000131 | CHEBI:117725 |
| 1 | HP:0000131 | CHEBI:104588 |
| 1 | HP:0000131 | CHEBI:98371 |
| 1 | HP:0000131 | CHEBI:117770 |
| 1 | HP:0000131 | CHEBI:101668 |
| 1 | HP:0000131 | CHEBI:104535 |
| 1 | HP:0000131 | CHEBI:98338 |
| 1 | HP:0000131 | CHEBI:117783 |
| 1 | HP:0000131 | CHEBI:98301 |
| 1 | HP:0000131 | CHEBI:98310 |
| 1 | HP:0000131 | CHEBI:103145 |
| 1 | HP:0000131 | CHEBI:104794 |
| 1 | HP:0000131 | CHEBI:104592 |
| 1 | HP:0000131 | CHEBI:104596 |
| 1 | HP:0000131 | CHEBI:117751 |
| 1 | HP:0000131 | CHEBI:104764 |
| 1 | HP:0000131 | CHEBI:104563 |
| 1 | HP:0000131 | CHEBI:97850 |
| 1 | HP:0000131 | CHEBI:117756 |
| 1 | HP:0000131 | CHEBI:104575 |
| 1 | HP:0000131 | CHEBI:117860 |
| 1 | HP:0000131 | CHEBI:117859 |
| 1 | HP:0000131 | CHEBI:117853 |
| 1 | HP:0000131 | CHEBI:98973 |
| 1 | HP:0000131 | CHEBI:98082 |
| 1 | HP:0000131 | CHEBI:98086 |
| 1 | HP:0000131 | CHEBI:117645 |
| 1 | HP:0000131 | CHEBI:117664 |
| 1 | HP:0000131 | ECTO:7000154 |
| 1 | HP:0000131 | ECTO:9000060 |
| 1 | HP:0000131 | CHEBI:117600 |
| 1 | HP:0000131 | CHEBI:117604 |
| 1 | HP:0000131 | CHEBI:117654 |
| 1 | HP:0000131 | ECTO:9002143 |
| 1 | HP:0000131 | ECTO:9000099 |
| 1 | HP:0000131 | ECTO:9000059 |
| 1 | HP:0000131 | ECTO:9002142 |
| 1 | HP:0000131 | ECTO:9000071 |
| 1 | HP:0000131 | CHEBI:97488 |
| 1 | HP:0000131 | ECTO:0500010 |
| 1 | HP:0000131 | ECTO:9000022 |
| 1 | HP:0000131 | CHEBI:122583 |
| 1 | HP:0000131 | ECTO:7000141 |
| 1 | HP:0000131 | CHEBI:122598 |
| 1 | HP:0000131 | CHEBI:122590 |
| 1 | HP:0000131 | CHEBI:122605 |
| 1 | HP:0000131 | CHEBI:97428 |
| 1 | HP:0000131 | CHEBI:122610 |
| 1 | HP:0000131 | CHEBI:122618 |
| 1 | HP:0000131 | CHEBI:8777 |
| 1 | HP:0000131 | CHEBI:97439 |
| 1 | HP:0000131 | ECTO:7000131 |
| 1 | HP:0000131 | ECTO:9000090 |
| 1 | HP:0000131 | CHEBI:103457 |
| 1 | HP:0000131 | CHEBI:123826 |
| 1 | HP:0000131 | CHEBI:123813 |
| 1 | HP:0000131 | CHEBI:127091 |
| 1 | HP:0000131 | CHEBI:127079 |
| 1 | HP:0000131 | CHEBI:101990 |
| 1 | HP:0000131 | CHEBI:123804 |
| 1 | HP:0000131 | CHEBI:95290 |
| 1 | HP:0000131 | ECTO:9000954 |
| 1 | HP:0000131 | ECTO:0070010 |
| 1 | HP:0000131 | ECTO:0070104 |
| 1 | HP:0000131 | ECTO:0070172 |
| 1 | HP:0000131 | ECTO:0070131 |
| 1 | HP:0000131 | ECTO:0070124 |
| 1 | HP:0000131 | ECTO:0070027 |
| 1 | HP:0000131 | ECTO:0070114 |
| 1 | HP:0000131 | ECTO:0070070 |
| 1 | HP:0000131 | ECTO:0070060 |
| 1 | HP:0000131 | ECTO:0070130 |
| 1 | HP:0000131 | MONDO:0024664 |
| 1 | HP:0000131 | MONDO:0005010 |
| 1 | HP:0000131 | CHEBI:127062 |
| 1 | HP:0000131 | CHEBI:123885 |
| 1 | HP:0000131 | CHEBI:123912 |
| 1 | HP:0000131 | CHEBI:123949 |
| 1 | HP:0000131 | CHEBI:123938 |
| 1 | HP:0000131 | CHEBI:123936 |
| 1 | HP:0000131 | CHEBI:123902 |
| 1 | HP:0000131 | CHEBI:123955 |
| 1 | HP:0000131 | CHEBI:123978 |
| 1 | HP:0000131 | CHEBI:97138 |
| 1 | HP:0000131 | CHEBI:123977 |
| 1 | HP:0000131 | CHEBI:123991 |
| 1 | HP:0000131 | CHEBI:123992 |
| 1 | HP:0000131 | CHEBI:97286 |
| 1 | HP:0000131 | CHEBI:123843 |
| 1 | HP:0000131 | CHEBI:123832 |
| 1 | HP:0000131 | CHEBI:123863 |
| 1 | HP:0000131 | CHEBI:123855 |
| 1 | HP:0000131 | MONDO:0008170 |
| 1 | HP:0000131 | CHEBI:95517 |
| 1 | HP:0000131 | CHEBI:113478 |
| 1 | HP:0000131 | CHEBI:5118 |
| 1 | HP:0000131 | CHEBI:113414 |
| 1 | HP:0000131 | CHEBI:113421 |
| 1 | HP:0000131 | CHEBI:113426 |
| 1 | HP:0000131 | CHEBI:113496 |
| 1 | HP:0000131 | CHEBI:7769 |
| 1 | HP:0000131 | CHEBI:95542 |
| 1 | HP:0000131 | CHEBI:102049 |
| 1 | HP:0000131 | CHEBI:102052 |
| 1 | HP:0000131 | CHEBI:95580 |
| 1 | HP:0000131 | CHEBI:113443 |
| 1 | HP:0000131 | CHEBI:95522 |
| 1 | HP:0000131 | CHEBI:102072 |
| 1 | HP:0000131 | CHEBI:113457 |
| 1 | HP:0000131 | CHEBI:113461 |
| 1 | HP:0000131 | CHEBI:113431 |
| 1 | HP:0000131 | CHEBI:113437 |
| 1 | HP:0000131 | CHEBI:102018 |
| 1 | HP:0000131 | CHEBI:113506 |
| 1 | HP:0000131 | CHEBI:101836 |
| 1 | HP:0000131 | CHEBI:95418 |
| 1 | HP:0000131 | CHEBI:95425 |
| 1 | HP:0000131 | CHEBI:95421 |
| 1 | HP:0000131 | CHEBI:95312 |
| 1 | HP:0000131 | CHEBI:101896 |
| 1 | HP:0000131 | CHEBI:95401 |
| 1 | HP:0000131 | CHEBI:95487 |
| 1 | HP:0000131 | CHEBI:113504 |
| 1 | HP:0000131 | CHEBI:95436 |
| 1 | HP:0000131 | HP:0000804 |
| 1 | HP:0000131 | CHEBI:102199 |
| 1 | HP:0000131 | CHEBI:101815 |
| 1 | HP:0000131 | CHEBI:102129 |
| 1 | HP:0000131 | CHEBI:113527 |
| 1 | HP:0000131 | CHEBI:101823 |
| 1 | HP:0000131 | CHEBI:123958 |
| 1 | HP:0000131 | CHEBI:117780 |
| 1 | HP:0000131 | CHEBI:97776 |
| 1 | HP:0000131 | CHEBI:104260 |
| 1 | HP:0000131 | CHEBI:104254 |
| 1 | HP:0000131 | CHEBI:104283 |
| 1 | HP:0000131 | CHEBI:104295 |
| 1 | HP:0000131 | CHEBI:127420 |
| 1 | HP:0000131 | CHEBI:6801 |
| 1 | HP:0000131 | ECTO:9001538 |
| 1 | HP:0000131 | ECTO:9002147 |
| 1 | HP:0000131 | CHEBI:104245 |
| 1 | HP:0000131 | CHEBI:123805 |
| 1 | HP:0000131 | CHEBI:102613 |
| 1 | HP:0000131 | CHEBI:117824 |
| 1 | HP:0000131 | CHEBI:117812 |
| 1 | HP:0000131 | CHEBI:95873 |
| 1 | HP:0000131 | CHEBI:123814 |
| 1 | HP:0000131 | CHEBI:117886 |
| 1 | HP:0000131 | CHEBI:127288 |
| 1 | HP:0000131 | CHEBI:129142 |
| 1 | HP:0000131 | CHEBI:117875 |
| 1 | HP:0000131 | CHEBI:104383 |
| 1 | HP:0000131 | CHEBI:104388 |
| 1 | HP:0000131 | CHEBI:104396 |
| 1 | HP:0000131 | CHEBI:104365 |
| 1 | HP:0000131 | CHEBI:104372 |
| 1 | HP:0000131 | CHEBI:117891 |
| 1 | HP:0000131 | CHEBI:127209 |
| 1 | HP:0000131 | CHEBI:117838 |
| 1 | HP:0000131 | CHEBI:127228 |
| 1 | HP:0000131 | CHEBI:97526 |
| 1 | HP:0000131 | CHEBI:97541 |
| 1 | HP:0000131 | CHEBI:117802 |
| 1 | HP:0000131 | CHEBI:117735 |
| 1 | HP:0000131 | CHEBI:117777 |
| 1 | HP:0000131 | CHEBI:129003 |
| 1 | HP:0000131 | CHEBI:97493 |
| 1 | HP:0000131 | CHEBI:117724 |
| 1 | HP:0000131 | CHEBI:97473 |
| 1 | HP:0000131 | CHEBI:129264 |
| 1 | HP:0000131 | CHEBI:117716 |
| 1 | HP:0000131 | CHEBI:117741 |
| 1 | HP:0000131 | CHEBI:117730 |
| 1 | HP:0000131 | CHEBI:123894 |
| 1 | HP:0000131 | CHEBI:123849 |
| 1 | HP:0000131 | CHEBI:123838 |
| 1 | HP:0000131 | CHEBI:123830 |
| 1 | HP:0000131 | CHEBI:117794 |
| 1 | HP:0000131 | CHEBI:123853 |
| 1 | HP:0000131 | CHEBI:123882 |
| 1 | HP:0000131 | CHEBI:97566 |
| 1 | HP:0000131 | CHEBI:117806 |
| 1 | HP:0000131 | CHEBI:117831 |
| 1 | HP:0000131 | CHEBI:129123 |
| 1 | HP:0000131 | CHEBI:102572 |
| 1 | HP:0000131 | ECTO:9000082 |
| 1 | HP:0000131 | ECTO:9000424 |
| 1 | HP:0000131 | ECTO:7000142 |
| 1 | HP:0000131 | ECTO:9000092 |
| 1 | HP:0000131 | ECTO:7000146 |
| 1 | HP:0000131 | ECTO:9000945 |
| 1 | HP:0000131 | ECTO:9000051 |
| 1 | HP:0000131 | ECTO:0000001 |
| 1 | HP:0000131 | CHEBI:102579 |
| 1 | HP:0000131 | HP:0003774 |
| 1 | HP:0000131 | CHEBI:117869 |
| 1 | HP:0000131 | CHEBI:117863 |
| 1 | HP:0000131 | CHEBI:102551 |
| 1 | HP:0000131 | CHEBI:117858 |
| 1 | HP:0000131 | CHEBI:117852 |
| 1 | HP:0000131 | CHEBI:95923 |
| 1 | HP:0000131 | CHEBI:104341 |
| 1 | HP:0000131 | CHEBI:104345 |
| 1 | HP:0000131 | CHEBI:104350 |
| 1 | HP:0000131 | CHEBI:104358 |
| 1 | HP:0000131 | CHEBI:129269 |
| 1 | HP:0000131 | CHEBI:104323 |
| 1 | HP:0000131 | CHEBI:129296 |
| 1 | HP:0000131 | CHEBI:104337 |
| 1 | HP:0000131 | CHEBI:127169 |
| 1 | HP:0000131 | ECTO:9000028 |
| 1 | HP:0000131 | ECTO:9000025 |
| 1 | HP:0000131 | CHEBI:123681 |
| 1 | HP:0000131 | CHEBI:123693 |
| 1 | HP:0000131 | CHEBI:97016 |
| 1 | HP:0000131 | CHEBI:123776 |
| 1 | HP:0000131 | CHEBI:123774 |
| 1 | HP:0000131 | CHEBI:97023 |
| 1 | HP:0000131 | CHEBI:97006 |
| 1 | HP:0000131 | CHEBI:97003 |
| 1 | HP:0000131 | CHEBI:123639 |
| 1 | HP:0000131 | CHEBI:123654 |
| 1 | HP:0000131 | CHEBI:123634 |
| 1 | HP:0000131 | CHEBI:123655 |
| 1 | HP:0000131 | CHEBI:97031 |
| 1 | HP:0000131 | CHEBI:123757 |
| 1 | HP:0000131 | CHEBI:124018 |
| 1 | HP:0000131 | CHEBI:123724 |
| 1 | HP:0000131 | CHEBI:123716 |
| 1 | HP:0000131 | CHEBI:124005 |
| 1 | HP:0000131 | CHEBI:123741 |
| 1 | HP:0000131 | CHEBI:123766 |
| 1 | HP:0000131 | CHEBI:124041 |
| 1 | HP:0000131 | CHEBI:124080 |
| 1 | HP:0000131 | CHEBI:6904 |
| 1 | HP:0000131 | CHEBI:127355 |
| 1 | HP:0000131 | CHEBI:124092 |
| 1 | HP:0000131 | CHEBI:123797 |
| 1 | HP:0000131 | CHEBI:96014 |
| 1 | HP:0000131 | CHEBI:113304 |
| 1 | HP:0000131 | CHEBI:127933 |
| 1 | HP:0000131 | CHEBI:113330 |
| 1 | HP:0000131 | HP:0001635 |
| 1 | HP:0000131 | CHEBI:113343 |
| 1 | HP:0000131 | CHEBI:129441 |
| 1 | HP:0000131 | CHEBI:129434 |
| 1 | HP:0000131 | CHEBI:96881 |
| 1 | HP:0000131 | CHEBI:96899 |
| 1 | HP:0000131 | CHEBI:115345 |
| 1 | HP:0000131 | CHEBI:115346 |
| 1 | HP:0000131 | CHEBI:129997 |
| 1 | HP:0000131 | CHEBI:96895 |
| 1 | HP:0000131 | CHEBI:7772 |
| 1 | HP:0000131 | CHEBI:96065 |
| 1 | HP:0000131 | CHEBI:96075 |
| 1 | HP:0000131 | CHEBI:96042 |
| 1 | HP:0000131 | CHEBI:127927 |
| 1 | HP:0000131 | CHEBI:96038 |
| 1 | HP:0000131 | CHEBI:113268 |
| 1 | HP:0000131 | CHEBI:113323 |
| 1 | HP:0000131 | CHEBI:113327 |
| 1 | HP:0000131 | CHEBI:115339 |
| 1 | HP:0000131 | CHEBI:102710 |
| 1 | HP:0000131 | CHEBI:96879 |
| 1 | HP:0000131 | CHEBI:97258 |
| 1 | HP:0000131 | CHEBI:96849 |
| 1 | HP:0000131 | CHEBI:96852 |
| 1 | HP:0000131 | CHEBI:97223 |
| 1 | HP:0000131 | CHEBI:115397 |
| 1 | HP:0000131 | CHEBI:115405 |
| 1 | HP:0000131 | CHEBI:102869 |
| 1 | HP:0000131 | CHEBI:102849 |
| 1 | HP:0000131 | CHEBI:102859 |
| 1 | HP:0000131 | CHEBI:102857 |
| 1 | HP:0000131 | CHEBI:127600 |
| 1 | HP:0000131 | CHEBI:96822 |
| 1 | HP:0000131 | CHEBI:129958 |
| 1 | HP:0000131 | CHEBI:115360 |
| 1 | HP:0000131 | CHEBI:115332 |
| 1 | HP:0000131 | CHEBI:102737 |
| 1 | HP:0000131 | CHEBI:96836 |
| 1 | HP:0000131 | CHEBI:96804 |
| 1 | HP:0000131 | CHEBI:96816 |
| 1 | HP:0000131 | CHEBI:115314 |
| 1 | HP:0000131 | CHEBI:115302 |
| 1 | HP:0000131 | CHEBI:115324 |
| 1 | HP:0000131 | CHEBI:115168 |
| 1 | HP:0000131 | CHEBI:102532 |
| 1 | HP:0000131 | CHEBI:115206 |
| 1 | HP:0000131 | CHEBI:96593 |
| 1 | HP:0000131 | CHEBI:96568 |
| 1 | HP:0000131 | CHEBI:115234 |
| 1 | HP:0000131 | CHEBI:95948 |
| 1 | HP:0000131 | CHEBI:115212 |
| 1 | HP:0000131 | CHEBI:95906 |
| 1 | HP:0000131 | CHEBI:130999 |
| 1 | HP:0000131 | CHEBI:101378 |
| 1 | HP:0000131 | CHEBI:115224 |
| 1 | HP:0000131 | CHEBI:95999 |
| 1 | HP:0000131 | CHEBI:115227 |
| 1 | HP:0000131 | CHEBI:5551 |
| 1 | HP:0000131 | CHEBI:95982 |
| 1 | HP:0000131 | HP:0031012 |
| 1 | HP:0000131 | HP:0031011 |
| 1 | HP:0000131 | CHEBI:130484 |
| 1 | HP:0000131 | CHEBI:115159 |
| 1 | HP:0000131 | CHEBI:115297 |
| 1 | HP:0000131 | CHEBI:115265 |
| 1 | HP:0000131 | CHEBI:115262 |
| 1 | HP:0000131 | CHEBI:129918 |
| 1 | HP:0000131 | CHEBI:102690 |
| 1 | HP:0000131 | CHEBI:107028 |
| 1 | HP:0000131 | CHEBI:107024 |
| 1 | HP:0000131 | CHEBI:97232 |
| 1 | HP:0000131 | CHEBI:101443 |
| 1 | HP:0000131 | CHEBI:101448 |
| 1 | HP:0000131 | CHEBI:107004 |
| 1 | HP:0000131 | CHEBI:97225 |
| 1 | HP:0000131 | CHEBI:101470 |
| 1 | HP:0000131 | CHEBI:101409 |
| 1 | HP:0000131 | CHEBI:129911 |
| 1 | HP:0000131 | CHEBI:96500 |
| 1 | HP:0000131 | CHEBI:101462 |
| 1 | HP:0000131 | CHEBI:101466 |
| 1 | HP:0000131 | CHEBI:115283 |
| 1 | HP:0000131 | CHEBI:115288 |
| 1 | HP:0000131 | CHEBI:115289 |
| 1 | HP:0000131 | CHEBI:115251 |
| 1 | HP:0000131 | CHEBI:96637 |
| 1 | HP:0000131 | CHEBI:127204 |
| 1 | HP:0000131 | HP:0000147 |
| 1 | HP:0000131 | CHEBI:112548 |
| 1 | HP:0000131 | CHEBI:112555 |
| 1 | HP:0000131 | CHEBI:96747 |
| 1 | HP:0000131 | CHEBI:112557 |
| 1 | HP:0000131 | CHEBI:112587 |
| 1 | HP:0000131 | CHEBI:96759 |
| 1 | HP:0000131 | CHEBI:112591 |
| 1 | HP:0000131 | CHEBI:96736 |
| 1 | HP:0000131 | CHEBI:96700 |
| 1 | HP:0000131 | CHEBI:112521 |
| 1 | HP:0000131 | CHEBI:112526 |
| 1 | HP:0000131 | CHEBI:112527 |
| 1 | HP:0000131 | CHEBI:101686 |
| 1 | HP:0000131 | CHEBI:112532 |
| 1 | HP:0000131 | CHEBI:112537 |
| 1 | HP:0000131 | CHEBI:115433 |
| 1 | HP:0000131 | CHEBI:115418 |
| 1 | HP:0000131 | CHEBI:96786 |
| 1 | HP:0000131 | CHEBI:96781 |
| 1 | HP:0000131 | CHEBI:112560 |
| 1 | HP:0000131 | CHEBI:127173 |
| 1 | HP:0000131 | CHEBI:96655 |
| 1 | HP:0000131 | CHEBI:112646 |
| 1 | HP:0000131 | CHEBI:115149 |
| 1 | HP:0000131 | CHEBI:130474 |
| 1 | HP:0000131 | CHEBI:115178 |
| 1 | HP:0000131 | CHEBI:101728 |
| 1 | HP:0000131 | CHEBI:115188 |
| 1 | HP:0000131 | CHEBI:115192 |
| 1 | HP:0000131 | CHEBI:112616 |
| 1 | HP:0000131 | HP:0006577 |
| 1 | HP:0000131 | CHEBI:129713 |
| 1 | HP:0000131 | CHEBI:129769 |
| 1 | HP:0000131 | CHEBI:112650 |
| 1 | HP:0000131 | CHEBI:112620 |
| 1 | HP:0000131 | CHEBI:115125 |
| 1 | HP:0000131 | CHEBI:112630 |
| 1 | HP:0000131 | CHEBI:112633 |
| 1 | HP:0000131 | CHEBI:101740 |
| 1 | HP:0000131 | CHEBI:101745 |
| 1 | HP:0000131 | HP:0030255 |
| 1 | HP:0000131 | CHEBI:97261 |
| 1 | HP:0000131 | CHEBI:107011 |
| 1 | HP:0000131 | CHEBI:107019 |
| 1 | HP:0000131 | CHEBI:101584 |
| 1 | HP:0000131 | CHEBI:127815 |
| 1 | HP:0000131 | HP:0004699 |
| 1 | HP:0000131 | CHEBI:127846 |
| 1 | HP:0000131 | CHEBI:113282 |
| 1 | HP:0000131 | CHEBI:96133 |
| 1 | HP:0000131 | CHEBI:113275 |
| 1 | HP:0000131 | CHEBI:129370 |
| 1 | HP:0000131 | CHEBI:129368 |
| 1 | HP:0000131 | CHEBI:102307 |
| 1 | HP:0000131 | CHEBI:129627 |
| 1 | HP:0000131 | CHEBI:97067 |
| 1 | HP:0000131 | CHEBI:130712 |
| 1 | HP:0000131 | CHEBI:129562 |
| 1 | HP:0000131 | CHEBI:129552 |
| 1 | HP:0000131 | CHEBI:129577 |
| 1 | HP:0000131 | CHEBI:130749 |
| 1 | HP:0000131 | CHEBI:96441 |
| 1 | HP:0000131 | CHEBI:96401 |
| 1 | HP:0000131 | HP:0002401 |
| 1 | HP:0000131 | CHEBI:95701 |
| 1 | HP:0000131 | CHEBI:97161 |
| 1 | HP:0000131 | CHEBI:97144 |
| 1 | HP:0000131 | CHEBI:101565 |
| 1 | HP:0000131 | CHEBI:101535 |
| 1 | HP:0000131 | CHEBI:95766 |
| 1 | HP:0000131 | CHEBI:97173 |
| 1 | HP:0000131 | CHEBI:97180 |
| 1 | HP:0000131 | CHEBI:96307 |
| 1 | HP:0000131 | CHEBI:29692 |
| 1 | HP:0000131 | CHEBI:96271 |
| 1 | HP:0000131 | CHEBI:129356 |
| 1 | HP:0000131 | CHEBI:102941 |
| 1 | HP:0000131 | CHEBI:96212 |
| 1 | HP:0000131 | CHEBI:102953 |
| 1 | HP:0000131 | CHEBI:130805 |
| 1 | HP:0000131 | CHEBI:96291 |
| 1 | HP:0000131 | CHEBI:96276 |
| 1 | HP:0000131 | CHEBI:97092 |
| 1 | HP:0000131 | CHEBI:96241 |
| 1 | HP:0000131 | CHEBI:96253 |
| 1 | HP:0000131 | CHEBI:130866 |
| 1 | HP:0000131 | CHEBI:129169 |
| 1 | HP:0000131 | CHEBI:102427 |
| 1 | HP:0000131 | CHEBI:97035 |
| 1 | HP:0000131 | CHEBI:97047 |
| 1 | HP:0000131 | CHEBI:97015 |
| 1 | HP:0000131 | CHEBI:102893 |
| 1 | HP:0000131 | CHEBI:96770 |
| 1 | HP:0000131 | CHEBI:123951 |
| 1 | HP:0000131 | CHEBI:116066 |
| 1 | HP:0000131 | CHEBI:116070 |
| 1 | HP:0000131 | CHEBI:96488 |
| 1 | HP:0000131 | CHEBI:123960 |
| 1 | HP:0000131 | CHEBI:123657 |
| 1 | HP:0000131 | CHEBI:115274 |
| 1 | HP:0000131 | CHEBI:115298 |
| 1 | HP:0000131 | CHEBI:123904 |
| 1 | HP:0000131 | CHEBI:97034 |
| 1 | HP:0000131 | CHEBI:97038 |
| 1 | HP:0000131 | CHEBI:115200 |
| 1 | HP:0000131 | CHEBI:115207 |
| 1 | HP:0000131 | CHEBI:123924 |
| 1 | HP:0000131 | CHEBI:97042 |
| 1 | HP:0000131 | CHEBI:97018 |
| 1 | HP:0000131 | CHEBI:115268 |
| 1 | HP:0000131 | CHEBI:115264 |
| 1 | HP:0000131 | CHEBI:123987 |
| 1 | HP:0000131 | CHEBI:115286 |
| 1 | HP:0000131 | CHEBI:116014 |
| 1 | HP:0000131 | CHEBI:116017 |
| 1 | HP:0000131 | CHEBI:115290 |
| 1 | HP:0000131 | CHEBI:116045 |
| 1 | HP:0000131 | CHEBI:116032 |
| 1 | HP:0000131 | CHEBI:116049 |
| 1 | HP:0000131 | CHEBI:116050 |
| 1 | HP:0000131 | CHEBI:123994 |
| 1 | HP:0000131 | CHEBI:116059 |
| 1 | HP:0000131 | CHEBI:116063 |
| 1 | HP:0000131 | CHEBI:116025 |
| 1 | HP:0000131 | CHEBI:116029 |
| 1 | HP:0000131 | CHEBI:113395 |
| 1 | HP:0000131 | CHEBI:123733 |
| 1 | HP:0000131 | CHEBI:123768 |
| 1 | HP:0000131 | CHEBI:123750 |
| 1 | HP:0000131 | CHEBI:123785 |
| 1 | HP:0000131 | CHEBI:123713 |
| 1 | HP:0000131 | CHEBI:113366 |
| 1 | HP:0000131 | CHEBI:94861 |
| 1 | HP:0000131 | CHEBI:123794 |
| 1 | HP:0000131 | CHEBI:94873 |
| 1 | HP:0000131 | CHEBI:113370 |
| 1 | HP:0000131 | CHEBI:113356 |
| 1 | HP:0000131 | CHEBI:123718 |
| 1 | HP:0000131 | CHEBI:123683 |
| 1 | HP:0000131 | CHEBI:123684 |
| 1 | HP:0000131 | CHEBI:94927 |
| 1 | HP:0000131 | CHEBI:123694 |
| 1 | HP:0000131 | CHEBI:94908 |
| 1 | HP:0000131 | CHEBI:94914 |
| 1 | HP:0000131 | CHEBI:94946 |
| 1 | HP:0000131 | HP:0008843 |
| 1 | HP:0000131 | CHEBI:96570 |
| 1 | HP:0000131 | CHEBI:97014 |
| 1 | HP:0000131 | CHEBI:97142 |
| 1 | HP:0000131 | CHEBI:96748 |
| 1 | HP:0000131 | CHEBI:96753 |
| 1 | HP:0000131 | CHEBI:123827 |
| 1 | HP:0000131 | CHEBI:96679 |
| 1 | HP:0000131 | CHEBI:115126 |
| 1 | HP:0000131 | CHEBI:123824 |
| 1 | HP:0000131 | CHEBI:123821 |
| 1 | HP:0000131 | CHEBI:96728 |
| 1 | HP:0000131 | HP:0004398 |
| 1 | HP:0000131 | CHEBI:36791 |
| 1 | HP:0000131 | CHEBI:97203 |
| 1 | HP:0000131 | CHEBI:96737 |
| 1 | HP:0000131 | CHEBI:127636 |
| 1 | HP:0000131 | CHEBI:115410 |
| 1 | HP:0000131 | CHEBI:115414 |
| 1 | HP:0000131 | CHEBI:115419 |
| 1 | HP:0000131 | CHEBI:115424 |
| 1 | HP:0000131 | CHEBI:123809 |
| 1 | HP:0000131 | CHEBI:115233 |
| 1 | HP:0000131 | CHEBI:97022 |
| 1 | HP:0000131 | CHEBI:97027 |
| 1 | HP:0000131 | CHEBI:115241 |
| 1 | HP:0000131 | CHEBI:115247 |
| 1 | HP:0000131 | CHEBI:115215 |
| 1 | HP:0000131 | CHEBI:129889 |
| 1 | HP:0000131 | CHEBI:115222 |
| 1 | HP:0000131 | CHEBI:96628 |
| 1 | HP:0000131 | CHEBI:115194 |
| 1 | HP:0000131 | CHEBI:123876 |
| 1 | HP:0000131 | CHEBI:115176 |
| 1 | HP:0000131 | CHEBI:123878 |
| 1 | HP:0000131 | CHEBI:115180 |
| 1 | HP:0000131 | CHEBI:127722 |
| 1 | HP:0000131 | CHEBI:123889 |
| 1 | HP:0000131 | CHEBI:123893 |
| 1 | HP:0000131 | CHEBI:115135 |
| 1 | HP:0000131 | CHEBI:115161 |
| 1 | HP:0000131 | CHEBI:115165 |
| 1 | HP:0000131 | CHEBI:123845 |
| 1 | HP:0000131 | CHEBI:115170 |
| 1 | HP:0000131 | CHEBI:115132 |
| 1 | HP:0000131 | CHEBI:115133 |
| 1 | HP:0000131 | CHEBI:123856 |
| 1 | HP:0000131 | CHEBI:123834 |
| 1 | HP:0000131 | CHEBI:123869 |
| 1 | HP:0000131 | CHEBI:123866 |
| 1 | HP:0000131 | CHEBI:115148 |
| 1 | HP:0000131 | CHEBI:103135 |
| 1 | HP:0000131 | CHEBI:123939 |
| 1 | HP:0000131 | CHEBI:96258 |
| 1 | HP:0000131 | CHEBI:101655 |
| 1 | HP:0000131 | CHEBI:101669 |
| 1 | HP:0000131 | CHEBI:115756 |
| 1 | HP:0000131 | CHEBI:115767 |
| 1 | HP:0000131 | CHEBI:101724 |
| 1 | HP:0000131 | MONDO:0002251 |
| 1 | HP:0000131 | MONDO:0002974 |
| 1 | HP:0000131 | ECTO:9002151 |
| 1 | HP:0000131 | ECTO:9000087 |
| 1 | HP:0000131 | ECTO:0070129 |
| 1 | HP:0000131 | ECTO:0070163 |
| 1 | HP:0000131 | ECTO:0070184 |
| 1 | HP:0000131 | ECTO:0070137 |
| 1 | HP:0000131 | ECTO:0070135 |
| 1 | HP:0000131 | ECTO:0070166 |
| 1 | HP:0000131 | ECTO:0070069 |
| 1 | HP:0000131 | ECTO:0070043 |
| 1 | HP:0000131 | CHEBI:115955 |
| 1 | HP:0000131 | CHEBI:115967 |
| 1 | HP:0000131 | CHEBI:101328 |
| 1 | HP:0000131 | CHEBI:115931 |
| 1 | HP:0000131 | CHEBI:115938 |
| 1 | HP:0000131 | CHEBI:96992 |
| 1 | HP:0000131 | CHEBI:96965 |
| 1 | HP:0000131 | CHEBI:96978 |
| 1 | HP:0000131 | CHEBI:96970 |
| 1 | HP:0000131 | CHEBI:96944 |
| 1 | HP:0000131 | CHEBI:96958 |
| 1 | HP:0000131 | CHEBI:96997 |
| 1 | HP:0000131 | CHEBI:96913 |
| 1 | HP:0000131 | CHEBI:101105 |
| 1 | HP:0000131 | CHEBI:96901 |
| 1 | HP:0000131 | CHEBI:96917 |
| 1 | HP:0000131 | CHEBI:96986 |
| 1 | HP:0000131 | CHEBI:96950 |
| 1 | HP:0000131 | CHEBI:101074 |
| 1 | HP:0000131 | CHEBI:96931 |
| 1 | HP:0000131 | CHEBI:123943 |
| 1 | HP:0000131 | CHEBI:117810 |
| 1 | HP:0000131 | CHEBI:124067 |
| 1 | HP:0000131 | CHEBI:117800 |
| 1 | HP:0000131 | CHEBI:96471 |
| 1 | HP:0000131 | CHEBI:96453 |
| 1 | HP:0000131 | CHEBI:117827 |
| 1 | HP:0000131 | CHEBI:96422 |
| 1 | HP:0000131 | CHEBI:124074 |
| 1 | HP:0000131 | CHEBI:124006 |
| 1 | HP:0000131 | CHEBI:124031 |
| 1 | HP:0000131 | CHEBI:103169 |
| 1 | HP:0000131 | CHEBI:124022 |
| 1 | HP:0000131 | CHEBI:98092 |
| 1 | HP:0000131 | CHEBI:124057 |
| 1 | HP:0000131 | CHEBI:117798 |
| 1 | HP:0000131 | CHEBI:129541 |
| 1 | HP:0000131 | CHEBI:124045 |
| 1 | HP:0000131 | CHEBI:124042 |
| 1 | HP:0000131 | HP:0001297 |
| 1 | HP:0000131 | HP:0004789 |
| 1 | HP:0000131 | HP:0100280 |
| 1 | HP:0000131 | CHEBI:117890 |
| 1 | HP:0000131 | CHEBI:103202 |
| 1 | HP:0000131 | CHEBI:117845 |
| 1 | HP:0000131 | CHEBI:117835 |
| 1 | HP:0000131 | CHEBI:117832 |
| 1 | HP:0000131 | CHEBI:101546 |
| 1 | HP:0000131 | CHEBI:101553 |
| 1 | HP:0000131 | CHEBI:101557 |
| 1 | HP:0000131 | CHEBI:98023 |
| 1 | HP:0000131 | CHEBI:117888 |
| 1 | HP:0000131 | CHEBI:117882 |
| 1 | HP:0000131 | CHEBI:124019 |
| 1 | HP:0000131 | CHEBI:117732 |
| 1 | HP:0000131 | CHEBI:96310 |
| 1 | HP:0000131 | CHEBI:117739 |
| 1 | HP:0000131 | CHEBI:103517 |
| 1 | HP:0000131 | CHEBI:103548 |
| 1 | HP:0000131 | CHEBI:122715 |
| 1 | HP:0000131 | CHEBI:103536 |
| 1 | HP:0000131 | CHEBI:122704 |
| 1 | HP:0000131 | CHEBI:103509 |
| 1 | HP:0000131 | CHEBI:103125 |
| 1 | HP:0000131 | CHEBI:122737 |
| 1 | HP:0000131 | CHEBI:117768 |
| 1 | HP:0000131 | CHEBI:103520 |
| 1 | HP:0000131 | CHEBI:122797 |
| 1 | HP:0000131 | CHEBI:122752 |
| 1 | HP:0000131 | CHEBI:122749 |
| 1 | HP:0000131 | CHEBI:122741 |
| 1 | HP:0000131 | CHEBI:122778 |
| 1 | HP:0000131 | CHEBI:122765 |
| 1 | HP:0000131 | CHEBI:122795 |
| 1 | HP:0000131 | CHEBI:103527 |
| 1 | HP:0000131 | CHEBI:122789 |
| 1 | HP:0000131 | CHEBI:122730 |
| 1 | HP:0000131 | CHEBI:129676 |
| 1 | HP:0000131 | CHEBI:117727 |
| 1 | HP:0000131 | CHEBI:117720 |
| 1 | HP:0000131 | CHEBI:96355 |
| 1 | HP:0000131 | CHEBI:117718 |
| 1 | HP:0000131 | CHEBI:129605 |
| 1 | HP:0000131 | CHEBI:117712 |
| 1 | HP:0000131 | CHEBI:129626 |
| 1 | HP:0000131 | CHEBI:96317 |
| 1 | HP:0000131 | CHEBI:117773 |
| 1 | HP:0000131 | CHEBI:117785 |
| 1 | HP:0000131 | CHEBI:103228 |
| 1 | HP:0000131 | CHEBI:101513 |
| 1 | HP:0000131 | CHEBI:101397 |
| 1 | HP:0000131 | ECTO:9000063 |
| 1 | HP:0000131 | CHEBI:117865 |
| 1 | HP:0000131 | CHEBI:101798 |
| 1 | HP:0000131 | CHEBI:117854 |
| 1 | HP:0000131 | CHEBI:98134 |
| 1 | HP:0000131 | ECTO:9000018 |
| 1 | HP:0000131 | ECTO:9000074 |
| 1 | HP:0000131 | ECTO:9000038 |
| 1 | HP:0000131 | ECTO:7000144 |
| 1 | HP:0000131 | ECTO:9000031 |
| 1 | HP:0000131 | ECTO:0500017 |
| 1 | HP:0000131 | ECTO:9002164 |
| 1 | HP:0000131 | CHEBI:101438 |
| 1 | HP:0000131 | ECTO:0500016 |
| 1 | HP:0000131 | ECTO:0500008 |
| 1 | HP:0000131 | ECTO:9000052 |
| 1 | HP:0000131 | ECTO:7000143 |
| 1 | HP:0000131 | CHEBI:115940 |
| 1 | HP:0000131 | CHEBI:115947 |
| 1 | HP:0000131 | CHEBI:100687 |
| 1 | HP:0000131 | HP:0004936 |
| 1 | HP:0000131 | HP:0001370 |
| 1 | HP:0000131 | HP:0002592 |
| 1 | HP:0000131 | CHEBI:100647 |
| 1 | HP:0000131 | CHEBI:123786 |
| 1 | HP:0000131 | CHEBI:124012 |
| 1 | HP:0000131 | CHEBI:104742 |
| 1 | HP:0000131 | CHEBI:124034 |
| 1 | HP:0000131 | CHEBI:104751 |
| 1 | HP:0000131 | CHEBI:104755 |
| 1 | HP:0000131 | CHEBI:104728 |
| 1 | HP:0000131 | CHEBI:123719 |
| 1 | HP:0000131 | CHEBI:124025 |
| 1 | HP:0000131 | CHEBI:103488 |
| 1 | HP:0000131 | CHEBI:104726 |
| 1 | HP:0000131 | CHEBI:124058 |
| 1 | HP:0000131 | CHEBI:103483 |
| 1 | HP:0000131 | CHEBI:123739 |
| 1 | HP:0000131 | CHEBI:104731 |
| 1 | HP:0000131 | CHEBI:104737 |
| 1 | HP:0000131 | CHEBI:123761 |
| 1 | HP:0000131 | CHEBI:124040 |
| 1 | HP:0000131 | CHEBI:123754 |
| 1 | HP:0000131 | CHEBI:123751 |
| 1 | HP:0000131 | CHEBI:123793 |
| 1 | HP:0000131 | CHEBI:103419 |
| 1 | HP:0000131 | CHEBI:104719 |
| 1 | HP:0000131 | CHEBI:104787 |
| 1 | HP:0000131 | CHEBI:100758 |
| 1 | HP:0000131 | CHEBI:104851 |
| 1 | HP:0000131 | CHEBI:104855 |
| 1 | HP:0000131 | CHEBI:100003 |
| 1 | HP:0000131 | CHEBI:103336 |
| 1 | HP:0000131 | CHEBI:102440 |
| 1 | HP:0000131 | CHEBI:102453 |
| 1 | HP:0000131 | CHEBI:104783 |
| 1 | HP:0000131 | CHEBI:103395 |
| 1 | HP:0000131 | CHEBI:104700 |
| 1 | HP:0000131 | CHEBI:104797 |
| 1 | HP:0000131 | CHEBI:100762 |
| 1 | HP:0000131 | CHEBI:104762 |
| 1 | HP:0000131 | HP:0012887 |
| 1 | HP:0000131 | CHEBI:104772 |
| 1 | HP:0000131 | CHEBI:104778 |
| 1 | HP:0000131 | CHEBI:103406 |
| 1 | HP:0000131 | CHEBI:124084 |
| 1 | HP:0000131 | CHEBI:124048 |
| 1 | HP:0000131 | CHEBI:129020 |
| 1 | HP:0000131 | CHEBI:123771 |
| 1 | HP:0000131 | CHEBI:123666 |
| 1 | HP:0000131 | CHEBI:123650 |
| 1 | HP:0000131 | CHEBI:123658 |
| 1 | HP:0000131 | CHEBI:97363 |
| 1 | HP:0000131 | CHEBI:103523 |
| 1 | HP:0000131 | CHEBI:103513 |
| 1 | HP:0000131 | CHEBI:103545 |
| 1 | HP:0000131 | CHEBI:102318 |
| 1 | HP:0000131 | CHEBI:123969 |
| 1 | HP:0000131 | CHEBI:95779 |
| 1 | HP:0000131 | CHEBI:123989 |
| 1 | HP:0000131 | CHEBI:123988 |
| 1 | HP:0000131 | CHEBI:103501 |
| 1 | HP:0000131 | CHEBI:123995 |
| 1 | HP:0000131 | HP:0003088 |
| 1 | HP:0000131 | CHEBI:117766 |
| 1 | HP:0000131 | CHEBI:117762 |
| 1 | HP:0000131 | CHEBI:123914 |
| 1 | HP:0000131 | CHEBI:123945 |
| 1 | HP:0000131 | CHEBI:97396 |
| 1 | HP:0000131 | CHEBI:104698 |
| 1 | HP:0000131 | CHEBI:104692 |
| 1 | HP:0000131 | CHEBI:104669 |
| 1 | HP:0000131 | CHEBI:123779 |
| 1 | HP:0000131 | CHEBI:124066 |
| 1 | HP:0000131 | CHEBI:95695 |
| 1 | HP:0000131 | CHEBI:123703 |
| 1 | HP:0000131 | CHEBI:103449 |
| 1 | HP:0000131 | CHEBI:103446 |
| 1 | HP:0000131 | CHEBI:103430 |
| 1 | HP:0000131 | CHEBI:95686 |
| 1 | HP:0000131 | CHEBI:103469 |
| 1 | HP:0000131 | CHEBI:104689 |
| 1 | HP:0000131 | CHEBI:123679 |
| 1 | HP:0000131 | CHEBI:103453 |
| 1 | HP:0000131 | CHEBI:129006 |
| 1 | HP:0000131 | CHEBI:104645 |
| 1 | HP:0000131 | CHEBI:104650 |
| 1 | HP:0000131 | CHEBI:104658 |
| 1 | HP:0000131 | CHEBI:104683 |
| 1 | HP:0000131 | CHEBI:123646 |
| 1 | HP:0000131 | CHEBI:104845 |
| 1 | HP:0000131 | CHEBI:104865 |
| 1 | HP:0000131 | CHEBI:104801 |
| 1 | HP:0000131 | CHEBI:104805 |
| 1 | HP:0000131 | CHEBI:100536 |
| 1 | HP:0000131 | CHEBI:104632 |
| 1 | HP:0000131 | CHEBI:104636 |
| 1 | HP:0000131 | CHEBI:104607 |
| 1 | HP:0000131 | CHEBI:104274 |
| 1 | HP:0000131 | CHEBI:104602 |
| 1 | HP:0000131 | CHEBI:104606 |
| 1 | HP:0000131 | CHEBI:104613 |
| 1 | HP:0000131 | CHEBI:104617 |
| 1 | HP:0000131 | CHEBI:104248 |
| 1 | HP:0000131 | HP:0001931 |
| 1 | HP:0000131 | CHEBI:97817 |
| 1 | HP:0000131 | CHEBI:97824 |
| 1 | HP:0000131 | CHEBI:100859 |
| 1 | HP:0000131 | CHEBI:98773 |
| 1 | HP:0000131 | CHEBI:104545 |
| 1 | HP:0000131 | CHEBI:104557 |
| 1 | HP:0000131 | CHEBI:104293 |
| 1 | HP:0000131 | CHEBI:98766 |
| 1 | HP:0000131 | CHEBI:104263 |
| 1 | HP:0000131 | CHEBI:100516 |
| 1 | HP:0000131 | CHEBI:104335 |
| 1 | HP:0000131 | CHEBI:100576 |
| 1 | HP:0000131 | CHEBI:104319 |
| 1 | HP:0000131 | CHEBI:98811 |
| 1 | HP:0000131 | CHEBI:104340 |
| 1 | HP:0000131 | CHEBI:104356 |
| 1 | HP:0000131 | CHEBI:100590 |
| 1 | HP:0000131 | CHEBI:98798 |
| 1 | HP:0000131 | CHEBI:104321 |
| 1 | HP:0000131 | CHEBI:104329 |
| 1 | HP:0000131 | CHEBI:104529 |
| 1 | HP:0000131 | CHEBI:97879 |
| 1 | HP:0000131 | CHEBI:102993 |
| 1 | HP:0000131 | CHEBI:103381 |
| 1 | HP:0000131 | CHEBI:103358 |
| 1 | HP:0000131 | CHEBI:104403 |
| 1 | HP:0000131 | CHEBI:104410 |
| 1 | HP:0000131 | CHEBI:104418 |
| 1 | HP:0000131 | HP:0011005 |
| 1 | HP:0000131 | CHEBI:104460 |
| 1 | HP:0000131 | CHEBI:104466 |
| 1 | HP:0000131 | CHEBI:103387 |
| 1 | HP:0000131 | CHEBI:104473 |
| 1 | HP:0000131 | CHEBI:104434 |
| 1 | HP:0000131 | CHEBI:104479 |
| 1 | HP:0000131 | CHEBI:103373 |
| 1 | HP:0000131 | CHEBI:104445 |
| 1 | HP:0000131 | CHEBI:103380 |
| 1 | HP:0000131 | CHEBI:104457 |
| 1 | HP:0000131 | CHEBI:104827 |
| 1 | HP:0000131 | CHEBI:104823 |
| 1 | HP:0000131 | CHEBI:104830 |
| 1 | HP:0000131 | CHEBI:100816 |
| 1 | HP:0000131 | CHEBI:103367 |
| 1 | HP:0000131 | CHEBI:104587 |
| 1 | HP:0000131 | CHEBI:100855 |
| 1 | HP:0000131 | CHEBI:97774 |
| 1 | HP:0000131 | CHEBI:104595 |
| 1 | HP:0000131 | CHEBI:104562 |
| 1 | HP:0000131 | CHEBI:104568 |
| 1 | HP:0000131 | CHEBI:104574 |
| 1 | HP:0000131 | CHEBI:104579 |
| 1 | HP:0000131 | CHEBI:100865 |
| 1 | HP:0000131 | CHEBI:104509 |
| 1 | HP:0000131 | CHEBI:104501 |
| 1 | HP:0000131 | CHEBI:100835 |
| 1 | HP:0000131 | CHEBI:104513 |
| 1 | HP:0000131 | CHEBI:97952 |
| 1 | HP:0000131 | CHEBI:97748 |
| 1 | HP:0000131 | CHEBI:128862 |
| 1 | HP:0000131 | CHEBI:104485 |
| 1 | HP:0000131 | CHEBI:104489 |
| 1 | HP:0000131 | CHEBI:104496 |
| 1 | HP:0000131 | CHEBI:97905 |
| 1 | HP:0000131 | CHEBI:115864 |
| 1 | HP:0000131 | CHEBI:115884 |
| 1 | HP:0000131 | CHEBI:115885 |
| 1 | HP:0000131 | CHEBI:115889 |
| 1 | HP:0000131 | CHEBI:115833 |
| 1 | HP:0000131 | CHEBI:115841 |
| 1 | HP:0000131 | CHEBI:115811 |
| 1 | HP:0000131 | CHEBI:115819 |
| 1 | HP:0000131 | CHEBI:115822 |
| 1 | HP:0000131 | CHEBI:115806 |
| 1 | HP:0000131 | CHEBI:115910 |
| 1 | HP:0000131 | CHEBI:115917 |
| 1 | HP:0000131 | CHEBI:115927 |
| 1 | HP:0000131 | CHEBI:115905 |
| 1 | HP:0000131 | CHEBI:115981 |
| 1 | HP:0000131 | CHEBI:115890 |
| 1 | HP:0000131 | CHEBI:100957 |
| 1 | HP:0000131 | CHEBI:101177 |
| 1 | HP:0000131 | CHEBI:115777 |
| 1 | HP:0000131 | CHEBI:115783 |
| 1 | HP:0000131 | CHEBI:96765 |
| 1 | HP:0000131 | CHEBI:122751 |
| 1 | HP:0000131 | CHEBI:122703 |
| 1 | HP:0000131 | CHEBI:107015 |
| 1 | HP:0000131 | CHEBI:129336 |
| 1 | HP:0000131 | CHEBI:115174 |
| 1 | HP:0000131 | CHEBI:100989 |
| 1 | HP:0000131 | CHEBI:129354 |
| 1 | HP:0000131 | CHEBI:115185 |
| 1 | HP:0000131 | CHEBI:96229 |
| 1 | HP:0000131 | CHEBI:100984 |
| 1 | HP:0000131 | CHEBI:115190 |
| 1 | HP:0000131 | CHEBI:98600 |
| 1 | HP:0000131 | CHEBI:107000 |
| 1 | HP:0000131 | CHEBI:122794 |
| 1 | HP:0000131 | CHEBI:122792 |
| 1 | HP:0000131 | CHEBI:129708 |
| 1 | HP:0000131 | CHEBI:122764 |
| 1 | HP:0000131 | CHEBI:122777 |
| 1 | HP:0000131 | CHEBI:122740 |
| 1 | HP:0000131 | CHEBI:98266 |
| 1 | HP:0000131 | CHEBI:103757 |
| 1 | HP:0000131 | CHEBI:115144 |
| 1 | HP:0000131 | CHEBI:96613 |
| 1 | HP:0000131 | CHEBI:122800 |
| 1 | HP:0000131 | CHEBI:98069 |
| 1 | HP:0000131 | CHEBI:115221 |
| 1 | HP:0000131 | CHEBI:115228 |
| 1 | HP:0000131 | CHEBI:96184 |
| 1 | HP:0000131 | CHEBI:96194 |
| 1 | HP:0000131 | CHEBI:122820 |
| 1 | HP:0000131 | CHEBI:98002 |
| 1 | HP:0000131 | CHEBI:96144 |
| 1 | HP:0000131 | CHEBI:122801 |
| 1 | HP:0000131 | CHEBI:115137 |
| 1 | HP:0000131 | CHEBI:122813 |
| 1 | HP:0000131 | CHEBI:115151 |
| 1 | HP:0000131 | CHEBI:98309 |
| 1 | HP:0000131 | CHEBI:129783 |
| 1 | HP:0000131 | CHEBI:115163 |
| 1 | HP:0000131 | CHEBI:98344 |
| 1 | HP:0000131 | CHEBI:103778 |
| 1 | HP:0000131 | CHEBI:115171 |
| 1 | HP:0000131 | CHEBI:98261 |
| 1 | HP:0000131 | CHEBI:122759 |
| 1 | HP:0000131 | CHEBI:96383 |
| 1 | HP:0000131 | CHEBI:98643 |
| 1 | HP:0000131 | CHEBI:103636 |
| 1 | HP:0000131 | CHEBI:103641 |
| 1 | HP:0000131 | CHEBI:103648 |
| 1 | HP:0000131 | CHEBI:122810 |
| 1 | HP:0000131 | CHEBI:96757 |
| 1 | HP:0000131 | CHEBI:103588 |
| 1 | HP:0000131 | CHEBI:96751 |
| 1 | HP:0000131 | CHEBI:103555 |
| 1 | HP:0000131 | CHEBI:103559 |
| 1 | HP:0000131 | CHEBI:103562 |
| 1 | HP:0000131 | CHEBI:103591 |
| 1 | HP:0000131 | CHEBI:96733 |
| 1 | HP:0000131 | CHEBI:98518 |
| 1 | HP:0000131 | CHEBI:115430 |
| 1 | HP:0000131 | CHEBI:115444 |
| 1 | HP:0000131 | CHEBI:115412 |
| 1 | HP:0000131 | CHEBI:115426 |
| 1 | HP:0000131 | CHEBI:96787 |
| 1 | HP:0000131 | CHEBI:96312 |
| 1 | HP:0000131 | CHEBI:103668 |
| 1 | HP:0000131 | CHEBI:100933 |
| 1 | HP:0000131 | CHEBI:103661 |
| 1 | HP:0000131 | CHEBI:96245 |
| 1 | HP:0000131 | CHEBI:129726 |
| 1 | HP:0000131 | CHEBI:96669 |
| 1 | HP:0000131 | CHEBI:115122 |
| 1 | HP:0000131 | CHEBI:103670 |
| 1 | HP:0000131 | CHEBI:96674 |
| 1 | HP:0000131 | CHEBI:129766 |
| 1 | HP:0000131 | CHEBI:96249 |
| 1 | HP:0000131 | CHEBI:96242 |
| 1 | HP:0000131 | CHEBI:100909 |
| 1 | HP:0000131 | CHEBI:103656 |
| 1 | HP:0000131 | CHEBI:96653 |
| 1 | HP:0000131 | CHEBI:98236 |
| 1 | HP:0000131 | CHEBI:122822 |
| 1 | HP:0000131 | CHEBI:100952 |
| 1 | HP:0000131 | CHEBI:103625 |
| 1 | HP:0000131 | CHEBI:103607 |
| 1 | HP:0000131 | CHEBI:103600 |
| 1 | HP:0000131 | CHEBI:103650 |
| 1 | HP:0000131 | CHEBI:115219 |
| 1 | HP:0000131 | CHEBI:129460 |
| 1 | HP:0000131 | CHEBI:122757 |
| 1 | HP:0000131 | CHEBI:122756 |
| 1 | HP:0000131 | CHEBI:96876 |
| 1 | HP:0000131 | CHEBI:115372 |
| 1 | HP:0000131 | CHEBI:96843 |
| 1 | HP:0000131 | CHEBI:115808 |
| 1 | HP:0000131 | CHEBI:115379 |
| 1 | HP:0000131 | CHEBI:98111 |
| 1 | HP:0000131 | CHEBI:115383 |
| 1 | HP:0000131 | CHEBI:115384 |
| 1 | HP:0000131 | CHEBI:96859 |
| 1 | HP:0000131 | CHEBI:98556 |
| 1 | HP:0000131 | CHEBI:96850 |
| 1 | HP:0000131 | CHEBI:98133 |
| 1 | HP:0000131 | CHEBI:98540 |
| 1 | HP:0000131 | CHEBI:96834 |
| 1 | HP:0000131 | CHEBI:96802 |
| 1 | HP:0000131 | CHEBI:96809 |
| 1 | HP:0000131 | CHEBI:96814 |
| 1 | HP:0000131 | CHEBI:115312 |
| 1 | HP:0000131 | CHEBI:98582 |
| 1 | HP:0000131 | CHEBI:96867 |
| 1 | HP:0000131 | CHEBI:115396 |
| 1 | HP:0000131 | CHEBI:115865 |
| 1 | HP:0000131 | CHEBI:96780 |
| 1 | HP:0000131 | CHEBI:115415 |
| 1 | HP:0000131 | CHEBI:98156 |
| 1 | HP:0000131 | CHEBI:127774 |
| 1 | HP:0000131 | CHEBI:115422 |
| 1 | HP:0000131 | CHEBI:115425 |
| 1 | HP:0000131 | CHEBI:115813 |
| 1 | HP:0000131 | CHEBI:115821 |
| 1 | HP:0000131 | CHEBI:115829 |
| 1 | HP:0000131 | CHEBI:96797 |
| 1 | HP:0000131 | CHEBI:98185 |
| 1 | HP:0000131 | CHEBI:98178 |
| 1 | HP:0000131 | CHEBI:96791 |
| 1 | HP:0000131 | CHEBI:96768 |
| 1 | HP:0000131 | CHEBI:115876 |
| 1 | HP:0000131 | CHEBI:96779 |
| 1 | HP:0000131 | CHEBI:50686 |
| 1 | HP:0000131 | CHEBI:115850 |
| 1 | HP:0000131 | CHEBI:115855 |
| 1 | HP:0000131 | CHEBI:115322 |
| 1 | HP:0000131 | CHEBI:115328 |
| 1 | HP:0000131 | CHEBI:115272 |
| 1 | HP:0000131 | CHEBI:98431 |
| 1 | HP:0000131 | CHEBI:96034 |
| 1 | HP:0000131 | CHEBI:98032 |
| 1 | HP:0000131 | CHEBI:115203 |
| 1 | HP:0000131 | CHEBI:129498 |
| 1 | HP:0000131 | CHEBI:96565 |
| 1 | HP:0000131 | HP:0003940 |
| 1 | HP:0000131 | CHEBI:96573 |
| 1 | HP:0000131 | CHEBI:122785 |
| 1 | HP:0000131 | CHEBI:115231 |
| 1 | HP:0000131 | CHEBI:122709 |
| 1 | HP:0000131 | CHEBI:115235 |
| 1 | HP:0000131 | CHEBI:122790 |
| 1 | HP:0000131 | CHEBI:122762 |
| 1 | HP:0000131 | CHEBI:122768 |
| 1 | HP:0000131 | CHEBI:122775 |
| 1 | HP:0000131 | CHEBI:129861 |
| 1 | HP:0000131 | CHEBI:129439 |
| 1 | HP:0000131 | CHEBI:122744 |
| 1 | HP:0000131 | CHEBI:122712 |
| 1 | HP:0000131 | CHEBI:96057 |
| 1 | HP:0000131 | CHEBI:115795 |
| 1 | HP:0000131 | CHEBI:115333 |
| 1 | HP:0000131 | CHEBI:129959 |
| 1 | HP:0000131 | CHEBI:115352 |
| 1 | HP:0000131 | CHEBI:115359 |
| 1 | HP:0000131 | CHEBI:115284 |
| 1 | HP:0000131 | CHEBI:115361 |
| 1 | HP:0000131 | CHEBI:115292 |
| 1 | HP:0000131 | CHEBI:115255 |
| 1 | HP:0000131 | CHEBI:115370 |
| 1 | HP:0000131 | CHEBI:115338 |
| 1 | HP:0000131 | CHEBI:115294 |
| 1 | HP:0000131 | CHEBI:115269 |
| 1 | HP:0000131 | CHEBI:122723 |
| 1 | HP:0000131 | CHEBI:115342 |
| 1 | HP:0000131 | CHEBI:115347 |
| 1 | HP:0000131 | CHEBI:122727 |
| 1 | HP:0000131 | CHEBI:101133 |
| 1 | HP:0000131 | CHEBI:115785 |
| 1 | HP:0000131 | CHEBI:115789 |
| 1 | HP:0000131 | CHEBI:129633 |
| 1 | HP:0000131 | CHEBI:96798 |
| 1 | HP:0000131 | CHEBI:115843 |
| 1 | HP:0000131 | CHEBI:122604 |
| 1 | HP:0000131 | CHEBI:6539 |
| 1 | HP:0000131 | CHEBI:101675 |
| 1 | HP:0000131 | CHEBI:122630 |
| 1 | HP:0000131 | CHEBI:122626 |
| 1 | HP:0000131 | CHEBI:122654 |
| 1 | HP:0000131 | CHEBI:122651 |
| 1 | HP:0000131 | CHEBI:122647 |
| 1 | HP:0000131 | CHEBI:122644 |
| 1 | HP:0000131 | CHEBI:122671 |
| 1 | HP:0000131 | CHEBI:100832 |
| 1 | HP:0000131 | CHEBI:122691 |
| 1 | HP:0000131 | CHEBI:122696 |
| 1 | HP:0000131 | CHEBI:122680 |
| 1 | HP:0000131 | CHEBI:122684 |
| 1 | HP:0000131 | CHEBI:98222 |
| 1 | HP:0000131 | CHEBI:129677 |
| 1 | HP:0000131 | CHEBI:101644 |
| 1 | HP:0000131 | CHEBI:101618 |
| 1 | HP:0000131 | CHEBI:122615 |
| 1 | HP:0000131 | CHEBI:122636 |
| 1 | HP:0000131 | CHEBI:50270 |
| 1 | HP:0000131 | CHEBI:6538 |
| 1 | HP:0000131 | CHEBI:122694 |
| 1 | HP:0000131 | CHEBI:101796 |
| 1 | HP:0000131 | CHEBI:129070 |
| 1 | HP:0000131 | CHEBI:122595 |
| 1 | HP:0000131 | CHEBI:129012 |
| 1 | HP:0000131 | CHEBI:122607 |
| 1 | HP:0000131 | CHEBI:129329 |
| 1 | HP:0000131 | CHEBI:129334 |
| 1 | HP:0000131 | CHEBI:129352 |
| 1 | HP:0000131 | CHEBI:98363 |
| 1 | HP:0000131 | CHEBI:6741 |
| 1 | HP:0000131 | CHEBI:122641 |
| 1 | HP:0000131 | CHEBI:96230 |
| 1 | HP:0000131 | CHEBI:122635 |
| 1 | HP:0000131 | CHEBI:96210 |
| 1 | HP:0000131 | CHEBI:98273 |
| 1 | HP:0000131 | CHEBI:100726 |
| 1 | HP:0000131 | CHEBI:96247 |
| 1 | HP:0000131 | HP:0001047 |
| 1 | HP:0000131 | CHEBI:122608 |
| 1 | HP:0000131 | CHEBI:100774 |
| 1 | HP:0000131 | CHEBI:96364 |
| 1 | HP:0000131 | CHEBI:96350 |
| 1 | HP:0000131 | CHEBI:122559 |
| 1 | HP:0000131 | CHEBI:122553 |
| 1 | HP:0000131 | CHEBI:122542 |
| 1 | HP:0000131 | CHEBI:96996 |
| 1 | HP:0000131 | CHEBI:122579 |
| 1 | HP:0000131 | CHEBI:122560 |
| 1 | HP:0000131 | CHEBI:122564 |
| 1 | HP:0000131 | CHEBI:96993 |
| 1 | HP:0000131 | CHEBI:96968 |
| 1 | HP:0000131 | CHEBI:6446 |
| 1 | HP:0000131 | CHEBI:96973 |
| 1 | HP:0000131 | CHEBI:129546 |
| 1 | HP:0000131 | CHEBI:96943 |
| 1 | HP:0000131 | CHEBI:96953 |
| 1 | HP:0000131 | CHEBI:129560 |
| 1 | HP:0000131 | CHEBI:129556 |
| 1 | HP:0000131 | CHEBI:96925 |
| 1 | HP:0000131 | CHEBI:96928 |
| 1 | HP:0000131 | CHEBI:96938 |
| 1 | HP:0000131 | CHEBI:96985 |
| 1 | HP:0000131 | CHEBI:122523 |
| 1 | HP:0000131 | CHEBI:122526 |
| 1 | HP:0000131 | CHEBI:96303 |
| 1 | HP:0000131 | CHEBI:122597 |
| 1 | HP:0000131 | CHEBI:122582 |
| 1 | HP:0000131 | CHEBI:122589 |
| 1 | HP:0000131 | CHEBI:115753 |
| 1 | HP:0000131 | CHEBI:115763 |
| 1 | HP:0000131 | CHEBI:115764 |
| 1 | HP:0000131 | CHEBI:115769 |
| 1 | HP:0000131 | CHEBI:101122 |
| 1 | HP:0000131 | CHEBI:129643 |
| 1 | HP:0000131 | CHEBI:96918 |
| 1 | HP:0000131 | CHEBI:96380 |
| 1 | HP:0000131 | CHEBI:96384 |
| 1 | HP:0000131 | CHEBI:122518 |
| 1 | HP:0000131 | CHEBI:122510 |
| 1 | HP:0000131 | CHEBI:96361 |
| 1 | HP:0000131 | CHEBI:122530 |
| 1 | HP:0000131 | CHEBI:96902 |
| 1 | HP:0000131 | CHEBI:96912 |
| 1 | HP:0000131 | CHEBI:129059 |
| 1 | HP:0000131 | CHEBI:129095 |
| 1 | HP:0000131 | CHEBI:129081 |
| 1 | HP:0000131 | CHEBI:96145 |
| 1 | HP:0000131 | CHEBI:98449 |
| 1 | HP:0000131 | CHEBI:101566 |
| 1 | HP:0000131 | CHEBI:96869 |
| 1 | HP:0000131 | CHEBI:96877 |
| 1 | HP:0000131 | CHEBI:96873 |
| 1 | HP:0000131 | CHEBI:112561 |
| 1 | HP:0000131 | CHEBI:96844 |
| 1 | HP:0000131 | CHEBI:112569 |
| 1 | HP:0000131 | CHEBI:112572 |
| 1 | HP:0000131 | CHEBI:112576 |
| 1 | HP:0000131 | CHEBI:101572 |
| 1 | HP:0000131 | CHEBI:96853 |
| 1 | HP:0000131 | CHEBI:96855 |
| 1 | HP:0000131 | CHEBI:112559 |
| 1 | HP:0000131 | CHEBI:115392 |
| 1 | HP:0000131 | CHEBI:96826 |
| 1 | HP:0000131 | CHEBI:112585 |
| 1 | HP:0000131 | CHEBI:112592 |
| 1 | HP:0000131 | CHEBI:96832 |
| 1 | HP:0000131 | CHEBI:115398 |
| 1 | HP:0000131 | CHEBI:101555 |
| 1 | HP:0000131 | CHEBI:96411 |
| 1 | HP:0000131 | CHEBI:112640 |
| 1 | HP:0000131 | CHEBI:96790 |
| 1 | HP:0000131 | CHEBI:112607 |
| 1 | HP:0000131 | CHEBI:112609 |
| 1 | HP:0000131 | CHEBI:96769 |
| 1 | HP:0000131 | CHEBI:96762 |
| 1 | HP:0000131 | CHEBI:98651 |
| 1 | HP:0000131 | CHEBI:129550 |
| 1 | HP:0000131 | CHEBI:129107 |
| 1 | HP:0000131 | CHEBI:96773 |
| 1 | HP:0000131 | CHEBI:96414 |
| 1 | HP:0000131 | CHEBI:96461 |
| 1 | HP:0000131 | CHEBI:112622 |
| 1 | HP:0000131 | CHEBI:112628 |
| 1 | HP:0000131 | CHEBI:96451 |
| 1 | HP:0000131 | CHEBI:115401 |
| 1 | HP:0000131 | CHEBI:112632 |
| 1 | HP:0000131 | CHEBI:129158 |
| 1 | HP:0000131 | CHEBI:129525 |
| 1 | HP:0000131 | CHEBI:96808 |
| 1 | HP:0000131 | CHEBI:101524 |
| 1 | HP:0000131 | CHEBI:96812 |
| 1 | HP:0000131 | CHEBI:122578 |
| 1 | HP:0000131 | CHEBI:96079 |
| 1 | HP:0000131 | CHEBI:96046 |
| 1 | HP:0000131 | CHEBI:96055 |
| 1 | HP:0000131 | CHEBI:96010 |
| 1 | HP:0000131 | CHEBI:100581 |
| 1 | HP:0000131 | CHEBI:129438 |
| 1 | HP:0000131 | CHEBI:100569 |
| 1 | HP:0000131 | CHEBI:122562 |
| 1 | HP:0000131 | CHEBI:122572 |
| 1 | HP:0000131 | CHEBI:100679 |
| 1 | HP:0000131 | CHEBI:122540 |
| 1 | HP:0000131 | CHEBI:122549 |
| 1 | HP:0000131 | CHEBI:122551 |
| 1 | HP:0000131 | CHEBI:122520 |
| 1 | HP:0000131 | CHEBI:122529 |
| 1 | HP:0000131 | CHEBI:129001 |
| 1 | HP:0000131 | CHEBI:96185 |
| 1 | HP:0000131 | CHEBI:122517 |
| 1 | HP:0000131 | CHEBI:96893 |
| 1 | HP:0000131 | CHEBI:115343 |
| 1 | HP:0000131 | CHEBI:115320 |
| 1 | HP:0000131 | CHEBI:101492 |
| 1 | HP:0000131 | CHEBI:112524 |
| 1 | HP:0000131 | CHEBI:103813 |
| 1 | HP:0000131 | CHEBI:100698 |
| 1 | HP:0000131 | CHEBI:112531 |
| 1 | HP:0000131 | CHEBI:115329 |
| 1 | HP:0000131 | CHEBI:103807 |
| 1 | HP:0000131 | CHEBI:103802 |
| 1 | HP:0000131 | CHEBI:112510 |
| 1 | HP:0000131 | CHEBI:115301 |
| 1 | HP:0000131 | CHEBI:115334 |
| 1 | HP:0000131 | CHEBI:115304 |
| 1 | HP:0000131 | CHEBI:115308 |
| 1 | HP:0000131 | CHEBI:115350 |
| 1 | HP:0000131 | CHEBI:115362 |
| 1 | HP:0000131 | CHEBI:115368 |
| 1 | HP:0000131 | CHEBI:98362 |
| 1 | HP:0000131 | CHEBI:129971 |
| 1 | HP:0000131 | CHEBI:115330 |
| 1 | HP:0000131 | CHEBI:115443 |
| 1 | HP:0000131 | CHEBI:115438 |
| 1 | HP:0000131 | CHEBI:96465 |
| 1 | HP:0000131 | CHEBI:104814 |
| 1 | HP:0000131 | CHEBI:96916 |
| 1 | HP:0000131 | CHEBI:98983 |
| 1 | HP:0000131 | CHEBI:127662 |
| 1 | HP:0000131 | CHEBI:96988 |
| 1 | HP:0000131 | CHEBI:104576 |
| 1 | HP:0000131 | CHEBI:104572 |
| 1 | HP:0000131 | CHEBI:117602 |
| 1 | HP:0000131 | CHEBI:127652 |
| 1 | HP:0000131 | CHEBI:104775 |
| 1 | HP:0000131 | CHEBI:117608 |
| 1 | HP:0000131 | CHEBI:104560 |
| 1 | HP:0000131 | CHEBI:104875 |
| 1 | HP:0000131 | CHEBI:104871 |
| 1 | HP:0000131 | CHEBI:104597 |
| 1 | HP:0000131 | CHEBI:104766 |
| 1 | HP:0000131 | CHEBI:104761 |
| 1 | HP:0000131 | CHEBI:96991 |
| 1 | HP:0000131 | CHEBI:117657 |
| 1 | HP:0000131 | CHEBI:104580 |
| 1 | HP:0000131 | CHEBI:96908 |
| 1 | HP:0000131 | CHEBI:96904 |
| 1 | HP:0000131 | CHEBI:104844 |
| 1 | HP:0000131 | CHEBI:104747 |
| 1 | HP:0000131 | CHEBI:116028 |
| 1 | HP:0000131 | CHEBI:104420 |
| 1 | HP:0000131 | CHEBI:116062 |
| 1 | HP:0000131 | CHEBI:116055 |
| 1 | HP:0000131 | CHEBI:97901 |
| 1 | HP:0000131 | CHEBI:97925 |
| 1 | HP:0000131 | CHEBI:116044 |
| 1 | HP:0000131 | CHEBI:115765 |
| 1 | HP:0000131 | CHEBI:104492 |
| 1 | HP:0000131 | CHEBI:117629 |
| 1 | HP:0000131 | CHEBI:104487 |
| 1 | HP:0000131 | CHEBI:116006 |
| 1 | HP:0000131 | CHEBI:116001 |
| 1 | HP:0000131 | CHEBI:97930 |
| 1 | HP:0000131 | CHEBI:103092 |
| 1 | HP:0000131 | HP:0033564 |
| 1 | HP:0000131 | CHEBI:117616 |
| 1 | HP:0000131 | CHEBI:117625 |
| 1 | HP:0000131 | CHEBI:104795 |
| 1 | HP:0000131 | CHEBI:104530 |
| 1 | HP:0000131 | CHEBI:104450 |
| 1 | HP:0000131 | CHEBI:96961 |
| 1 | HP:0000131 | CHEBI:104853 |
| 1 | HP:0000131 | CHEBI:104444 |
| 1 | HP:0000131 | CHEBI:104840 |
| 1 | HP:0000131 | CHEBI:103009 |
| 1 | HP:0000131 | CHEBI:104619 |
| 1 | HP:0000131 | CHEBI:104604 |
| 1 | HP:0000131 | CHEBI:104478 |
| 1 | HP:0000131 | CHEBI:104472 |
| 1 | HP:0000131 | CHEBI:104480 |
| 1 | HP:0000131 | CHEBI:104609 |
| 1 | HP:0000131 | CHEBI:104868 |
| 1 | HP:0000131 | CHEBI:104634 |
| 1 | HP:0000131 | CHEBI:104863 |
| 1 | HP:0000131 | CHEBI:104465 |
| 1 | HP:0000131 | CHEBI:104813 |
| 1 | HP:0000131 | CHEBI:104817 |
| 1 | HP:0000131 | CHEBI:104835 |
| 1 | HP:0000131 | CHEBI:104839 |
| 1 | HP:0000131 | CHEBI:104820 |
| 1 | HP:0000131 | CHEBI:104859 |
| 1 | HP:0000131 | CHEBI:101031 |
| 1 | HP:0000131 | CHEBI:117643 |
| 1 | HP:0000131 | CHEBI:104822 |
| 1 | HP:0000131 | CHEBI:104819 |
| 1 | HP:0000131 | CHEBI:104800 |
| 1 | HP:0000131 | CHEBI:117663 |
| 1 | HP:0000131 | CHEBI:96971 |
| 1 | HP:0000131 | CHEBI:96946 |
| 1 | HP:0000131 | CHEBI:117667 |
| 1 | HP:0000131 | CHEBI:96952 |
| 1 | HP:0000131 | CHEBI:104553 |
| 1 | HP:0000131 | CHEBI:96920 |
| 1 | HP:0000131 | CHEBI:97814 |
| 1 | HP:0000131 | CHEBI:97984 |
| 1 | HP:0000131 | CHEBI:104541 |
| 1 | HP:0000131 | CHEBI:104456 |
| 1 | HP:0000131 | CHEBI:96933 |
| 1 | HP:0000131 | CHEBI:117635 |
| 1 | HP:0000131 | CHEBI:117638 |
| 1 | HP:0000131 | CHEBI:104451 |
| 1 | HP:0000131 | CHEBI:97988 |
| 1 | HP:0000131 | CHEBI:116031 |
| 1 | HP:0000131 | CHEBI:116035 |
| 1 | HP:0000131 | CHEBI:104426 |
| 1 | HP:0000131 | CHEBI:104757 |
| 1 | HP:0000131 | CHEBI:104740 |
| 1 | HP:0000131 | CHEBI:98866 |
| 1 | HP:0000131 | CHEBI:104725 |
| 1 | HP:0000131 | CHEBI:104713 |
| 1 | HP:0000131 | CHEBI:104720 |
| 1 | HP:0000131 | CHEBI:104704 |
| 1 | HP:0000131 | CHEBI:104708 |
| 1 | HP:0000131 | CHEBI:104759 |
| 1 | HP:0000131 | CHEBI:104754 |
| 1 | HP:0000131 | CHEBI:104749 |
| 1 | HP:0000131 | CHEBI:101650 |
| 1 | HP:0000131 | CHEBI:104741 |
| 1 | HP:0000131 | CHEBI:97368 |
| 1 | HP:0000131 | CHEBI:97360 |
| 1 | HP:0000131 | CHEBI:104715 |
| 1 | HP:0000131 | CHEBI:104774 |
| 1 | HP:0000131 | CHEBI:104718 |
| 1 | HP:0000131 | CHEBI:101695 |
| 1 | HP:0000131 | CHEBI:104765 |
| 1 | HP:0000131 | CHEBI:104746 |
| 1 | HP:0000131 | CHEBI:104736 |
| 1 | HP:0000131 | CHEBI:104756 |
| 1 | HP:0000131 | CHEBI:104676 |
| 1 | HP:0000131 | CHEBI:104690 |
| 1 | HP:0000131 | CHEBI:104687 |
| 1 | HP:0000131 | CHEBI:104681 |
| 1 | HP:0000131 | CHEBI:104682 |
| 1 | HP:0000131 | CHEBI:104657 |
| 1 | HP:0000131 | CHEBI:104648 |
| 1 | HP:0000131 | CHEBI:104649 |
| 1 | HP:0000131 | CHEBI:97325 |
| 1 | HP:0000131 | CHEBI:104671 |
| 1 | HP:0000131 | CHEBI:104729 |
| 1 | HP:0000131 | CHEBI:104644 |
| 1 | HP:0000131 | CHEBI:104665 |
| 1 | HP:0000131 | CHEBI:104679 |
| 1 | HP:0000131 | CHEBI:104673 |
| 1 | HP:0000131 | CHEBI:97309 |
| 1 | HP:0000131 | CHEBI:104668 |
| 1 | HP:0000131 | CHEBI:3219 |
| 1 | HP:0000131 | HP:0033167 |
| 1 | HP:0000131 | CHEBI:104760 |
| 1 | HP:0000131 | CHEBI:104791 |
| 1 | HP:0000131 | CHEBI:104785 |
| 1 | HP:0000131 | CHEBI:117674 |
| 1 | HP:0000131 | CHEBI:104809 |
| 1 | HP:0000131 | CHEBI:104834 |
| 1 | HP:0000131 | CHEBI:104838 |
| 1 | HP:0000131 | CHEBI:117692 |
| 1 | HP:0000131 | CHEBI:98923 |
| 1 | HP:0000131 | CHEBI:104453 |
| 1 | HP:0000131 | CHEBI:104782 |
| 1 | HP:0000131 | CHEBI:104475 |
| 1 | HP:0000131 | CHEBI:97990 |
| 1 | HP:0000131 | CHEBI:104792 |
| 1 | HP:0000131 | CHEBI:104462 |
| 1 | HP:0000131 | CHEBI:98908 |
| 1 | HP:0000131 | CHEBI:104414 |
| 1 | HP:0000131 | CHEBI:103038 |
| 1 | HP:0000131 | CHEBI:117687 |
| 1 | HP:0000131 | CHEBI:104400 |
| 1 | HP:0000131 | CHEBI:104437 |
| 1 | HP:0000131 | CHEBI:104430 |
| 1 | HP:0000131 | CHEBI:104816 |
| 1 | HP:0000131 | CHEBI:104812 |
| 1 | HP:0000131 | CHEBI:104781 |
| 1 | HP:0000131 | CHEBI:117707 |
| 1 | HP:0000131 | CHEBI:104655 |
| 1 | HP:0000131 | CHEBI:104642 |
| 1 | HP:0000131 | CHEBI:104677 |
| 1 | HP:0000131 | CHEBI:104672 |
| 1 | HP:0000131 | CHEBI:104777 |
| 1 | HP:0000131 | CHEBI:127720 |
| 1 | HP:0000131 | CHEBI:104666 |
| 1 | HP:0000131 | CHEBI:104661 |
| 1 | HP:0000131 | CHEBI:104771 |
| 1 | HP:0000131 | CHEBI:104796 |
| 1 | HP:0000131 | CHEBI:104858 |
| 1 | HP:0000131 | CHEBI:104768 |
| 1 | HP:0000131 | CHEBI:127711 |
| 1 | HP:0000131 | CHEBI:104843 |
| 1 | HP:0000131 | CHEBI:127745 |
| 1 | HP:0000131 | CHEBI:104874 |
| 1 | HP:0000131 | CHEBI:104870 |
| 1 | HP:0000131 | CHEBI:104867 |
| 1 | HP:0000131 | CHEBI:104454 |
| 1 | HP:0000131 | CHEBI:104417 |
| 1 | HP:0000131 | CHEBI:115437 |
| 1 | HP:0000131 | CHEBI:115911 |
| 1 | HP:0000131 | CHEBI:113294 |
| 1 | HP:0000131 | CHEBI:122722 |
| 1 | HP:0000131 | CHEBI:97832 |
| 1 | HP:0000131 | CHEBI:122726 |
| 1 | HP:0000131 | CHEBI:122729 |
| 1 | HP:0000131 | CHEBI:104616 |
| 1 | HP:0000131 | CHEBI:113273 |
| 1 | HP:0000131 | CHEBI:122708 |
| 1 | HP:0000131 | CHEBI:122711 |
| 1 | HP:0000131 | CHEBI:104601 |
| 1 | HP:0000131 | CHEBI:122719 |
| 1 | HP:0000131 | CHEBI:101335 |
| 1 | HP:0000131 | CHEBI:115985 |
| 1 | HP:0000131 | CHEBI:96625 |
| 1 | HP:0000131 | CHEBI:127807 |
| 1 | HP:0000131 | CHEBI:115988 |
| 1 | HP:0000131 | CHEBI:101341 |
| 1 | HP:0000131 | CHEBI:96638 |
| 1 | HP:0000131 | CHEBI:115130 |
| 1 | HP:0000131 | CHEBI:115995 |
| 1 | HP:0000131 | CHEBI:115943 |
| 1 | HP:0000131 | CHEBI:96530 |
| 1 | HP:0000131 | CHEBI:104623 |
| 1 | HP:0000131 | CHEBI:96560 |
| 1 | HP:0000131 | CHEBI:104544 |
| 1 | HP:0000131 | CHEBI:129835 |
| 1 | HP:0000131 | CHEBI:115957 |
| 1 | HP:0000131 | CHEBI:98083 |
| 1 | HP:0000131 | CHEBI:115237 |
| 1 | HP:0000131 | CHEBI:95397 |
| 1 | HP:0000131 | CHEBI:104627 |
| 1 | HP:0000131 | CHEBI:115242 |
| 1 | HP:0000131 | CHEBI:115939 |
| 1 | HP:0000131 | CHEBI:115245 |
| 1 | HP:0000131 | CHEBI:129854 |
| 1 | HP:0000131 | CHEBI:115969 |
| 1 | HP:0000131 | CHEBI:115211 |
| 1 | HP:0000131 | CHEBI:129886 |
| 1 | HP:0000131 | CHEBI:115220 |
| 1 | HP:0000131 | CHEBI:115934 |
| 1 | HP:0000131 | CHEBI:115229 |
| 1 | HP:0000131 | CHEBI:127819 |
| 1 | HP:0000131 | CHEBI:115921 |
| 1 | HP:0000131 | CHEBI:104449 |
| 1 | HP:0000131 | CHEBI:122784 |
| 1 | HP:0000131 | CHEBI:98148 |
| 1 | HP:0000131 | CHEBI:129720 |
| 1 | HP:0000131 | CHEBI:96695 |
| 1 | HP:0000131 | CHEBI:96676 |
| 1 | HP:0000131 | CHEBI:115124 |
| 1 | HP:0000131 | CHEBI:101276 |
| 1 | HP:0000131 | CHEBI:122808 |
| 1 | HP:0000131 | CHEBI:122805 |
| 1 | HP:0000131 | CHEBI:98191 |
| 1 | HP:0000131 | CHEBI:122817 |
| 1 | HP:0000131 | CHEBI:115893 |
| 1 | HP:0000131 | CHEBI:115897 |
| 1 | HP:0000131 | CHEBI:98106 |
| 1 | HP:0000131 | CHEBI:96746 |
| 1 | HP:0000131 | CHEBI:96740 |
| 1 | HP:0000131 | CHEBI:101292 |
| 1 | HP:0000131 | CHEBI:96729 |
| 1 | HP:0000131 | CHEBI:96735 |
| 1 | HP:0000131 | CHEBI:115835 |
| 1 | HP:0000131 | CHEBI:115191 |
| 1 | HP:0000131 | CHEBI:115184 |
| 1 | HP:0000131 | CHEBI:115177 |
| 1 | HP:0000131 | CHEBI:122743 |
| 1 | HP:0000131 | CHEBI:122787 |
| 1 | HP:0000131 | CHEBI:8253 |
| 1 | HP:0000131 | CHEBI:122799 |
| 1 | HP:0000131 | CHEBI:115929 |
| 1 | HP:0000131 | CHEBI:115907 |
| 1 | HP:0000131 | CHEBI:122774 |
| 1 | HP:0000131 | CHEBI:115142 |
| 1 | HP:0000131 | CHEBI:115147 |
| 1 | HP:0000131 | CHEBI:104631 |
| 1 | HP:0000131 | CHEBI:115173 |
| 1 | HP:0000131 | CHEBI:122747 |
| 1 | HP:0000131 | CHEBI:122755 |
| 1 | HP:0000131 | CHEBI:115150 |
| 1 | HP:0000131 | CHEBI:104628 |
| 1 | HP:0000131 | CHEBI:95277 |
| 1 | HP:0000131 | CHEBI:104624 |
| 1 | HP:0000131 | CHEBI:127832 |
| 1 | HP:0000131 | CHEBI:104620 |
| 1 | HP:0000131 | CHEBI:104639 |
| 1 | HP:0000131 | CHEBI:95424 |
| 1 | HP:0000131 | CHEBI:98097 |
| 1 | HP:0000131 | CHEBI:95420 |
| 1 | HP:0000131 | CHEBI:101062 |
| 1 | HP:0000131 | CHEBI:97958 |
| 1 | HP:0000131 | CHEBI:104516 |
| 1 | HP:0000131 | CHEBI:104511 |
| 1 | HP:0000131 | CHEBI:95599 |
| 1 | HP:0000131 | CHEBI:104519 |
| 1 | HP:0000131 | CHEBI:104504 |
| 1 | HP:0000131 | CHEBI:104500 |
| 1 | HP:0000131 | CHEBI:113306 |
| 1 | HP:0000131 | CHEBI:113309 |
| 1 | HP:0000131 | CHEBI:101004 |
| 1 | HP:0000131 | CHEBI:104517 |
| 1 | HP:0000131 | CHEBI:113336 |
| 1 | HP:0000131 | CHEBI:95543 |
| 1 | HP:0000131 | CHEBI:104565 |
| 1 | HP:0000131 | CHEBI:102092 |
| 1 | HP:0000131 | CHEBI:98101 |
| 1 | HP:0000131 | CHEBI:98138 |
| 1 | HP:0000131 | CHEBI:104598 |
| 1 | HP:0000131 | CHEBI:102046 |
| 1 | HP:0000131 | CHEBI:98153 |
| 1 | HP:0000131 | CHEBI:97933 |
| 1 | HP:0000131 | CHEBI:104428 |
| 1 | HP:0000131 | CHEBI:104442 |
| 1 | HP:0000131 | CHEBI:104476 |
| 1 | HP:0000131 | CHEBI:104471 |
| 1 | HP:0000131 | CHEBI:104402 |
| 1 | HP:0000131 | CHEBI:104463 |
| 1 | HP:0000131 | CHEBI:104415 |
| 1 | HP:0000131 | CHEBI:104433 |
| 1 | HP:0000131 | CHEBI:104406 |
| 1 | HP:0000131 | CHEBI:104423 |
| 1 | HP:0000131 | CHEBI:104491 |
| 1 | HP:0000131 | CHEBI:117599 |
| 1 | HP:0000131 | CHEBI:104438 |
| 1 | HP:0000131 | CHEBI:104431 |
| 1 | HP:0000131 | CHEBI:104421 |
| 1 | HP:0000131 | CHEBI:104495 |
| 1 | HP:0000131 | CHEBI:97918 |
| 1 | HP:0000131 | CHEBI:104493 |
| 1 | HP:0000131 | CHEBI:104483 |
| 1 | HP:0000131 | HP:0000791 |
| 1 | HP:0000131 | CHEBI:97870 |
| 1 | HP:0000131 | CHEBI:104584 |
| 1 | HP:0000131 | CHEBI:115296 |
| 1 | HP:0000131 | CHEBI:115250 |
| 1 | HP:0000131 | CHEBI:115254 |
| 1 | HP:0000131 | CHEBI:95489 |
| 1 | HP:0000131 | CHEBI:115258 |
| 1 | HP:0000131 | CHEBI:102105 |
| 1 | HP:0000131 | CHEBI:98058 |
| 1 | HP:0000131 | CHEBI:115261 |
| 1 | HP:0000131 | CHEBI:104522 |
| 1 | HP:0000131 | CHEBI:104615 |
| 1 | HP:0000131 | CHEBI:102116 |
| 1 | HP:0000131 | CHEBI:98074 |
| 1 | HP:0000131 | CHEBI:98071 |
| 1 | HP:0000131 | CHEBI:102124 |
| 1 | HP:0000131 | CHEBI:104556 |
| 1 | HP:0000131 | CHEBI:104600 |
| 1 | HP:0000131 | CHEBI:96599 |
| 1 | HP:0000131 | CHEBI:115205 |
| 1 | HP:0000131 | CHEBI:95408 |
| 1 | HP:0000131 | CHEBI:104533 |
| 1 | HP:0000131 | CHEBI:96517 |
| 1 | HP:0000131 | CHEBI:113319 |
| 1 | HP:0000131 | CHEBI:104520 |
| 1 | HP:0000131 | CHEBI:104578 |
| 1 | HP:0000131 | CHEBI:104573 |
| 1 | HP:0000131 | CHEBI:104537 |
| 1 | HP:0000131 | CHEBI:104531 |
| 1 | HP:0000131 | CHEBI:113329 |
| 1 | HP:0000131 | CHEBI:129902 |
| 1 | HP:0000131 | CHEBI:104567 |
| 1 | HP:0000131 | CHEBI:104525 |
| 1 | HP:0000131 | CHEBI:102150 |
| 1 | HP:0000131 | CHEBI:104538 |
| 1 | HP:0000131 | CHEBI:97880 |
| 1 | HP:0000131 | CHEBI:104554 |
| 1 | HP:0000131 | CHEBI:104542 |
| 1 | HP:0000131 | CHEBI:96503 |
| 1 | HP:0000131 | CHEBI:115276 |
| 1 | HP:0000131 | CHEBI:102171 |
| 1 | HP:0000131 | CHEBI:102181 |
| 1 | HP:0000131 | CHEBI:98034 |
| 1 | HP:0000131 | CHEBI:129573 |
| 1 | HP:0000131 | CHEBI:100544 |
| 1 | HP:0000131 | CHEBI:101024 |
| 1 | HP:0000131 | CHEBI:117615 |
| 1 | HP:0000131 | CHEBI:117611 |
| 1 | HP:0000131 | CHEBI:100604 |
| 1 | HP:0000131 | CHEBI:117601 |
| 1 | HP:0000131 | CHEBI:117605 |
| 1 | HP:0000131 | CHEBI:100692 |
| 1 | HP:0000131 | CHEBI:100696 |
| 1 | HP:0000131 | CHEBI:101054 |
| 1 | HP:0000131 | CHEBI:96999 |
| 1 | HP:0000131 | CHEBI:96921 |
| 1 | HP:0000131 | CHEBI:117698 |
| 1 | HP:0000131 | CHEBI:100644 |
| 1 | HP:0000131 | CHEBI:96957 |
| 1 | HP:0000131 | CHEBI:96947 |
| 1 | HP:0000131 | CHEBI:96972 |
| 1 | HP:0000131 | CHEBI:96977 |
| 1 | HP:0000131 | CHEBI:96964 |
| 1 | HP:0000131 | CHEBI:96934 |
| 1 | HP:0000131 | CHEBI:117690 |
| 1 | HP:0000131 | CHEBI:117686 |
| 1 | HP:0000131 | CHEBI:117677 |
| 1 | HP:0000131 | CHEBI:117673 |
| 1 | HP:0000131 | CHEBI:98935 |
| 1 | HP:0000131 | CHEBI:100674 |
| 1 | HP:0000131 | CHEBI:117631 |
| 1 | HP:0000131 | CHEBI:117634 |
| 1 | HP:0000131 | CHEBI:115937 |
| 1 | HP:0000131 | CHEBI:115966 |
| 1 | HP:0000131 | CHEBI:115961 |
| 1 | HP:0000131 | CHEBI:115974 |
| 1 | HP:0000131 | CHEBI:115975 |
| 1 | HP:0000131 | CHEBI:115992 |
| 1 | HP:0000131 | CHEBI:115999 |
| 1 | HP:0000131 | CHEBI:115996 |
| 1 | HP:0000131 | CHEBI:117597 |
| 1 | HP:0000131 | CHEBI:96980 |
| 1 | HP:0000131 | CHEBI:101688 |
| 1 | HP:0000131 | CHEBI:101747 |
| 1 | HP:0000131 | HP:0002608 |
| 1 | HP:0000131 | HP:0012622 |
| 1 | HP:0000131 | HP:0002621 |
| 1 | HP:0000131 | HP:0012330 |
| 1 | HP:0000131 | MAXO:0000257 |
| 1 | HP:0000131 | CHEBI:98776 |
| 1 | HP:0000131 | CHEBI:98703 |
| 1 | HP:0000131 | CHEBI:115760 |
| 1 | HP:0000131 | CHEBI:115754 |
| 1 | HP:0000131 | CHEBI:98846 |
| 1 | HP:0000131 | CHEBI:96989 |
| 1 | HP:0000131 | CHEBI:117701 |
| 1 | HP:0000131 | CHEBI:96919 |
| 1 | HP:0000131 | CHEBI:96900 |
| 1 | HP:0000131 | CHEBI:101620 |
| 1 | HP:0000131 | CHEBI:101606 |
| 1 | HP:0000131 | CHEBI:98897 |
| 1 | HP:0000131 | CHEBI:96433 |
| 1 | HP:0000131 | CHEBI:101175 |
| 1 | HP:0000131 | CHEBI:115780 |
| 1 | HP:0000131 | CHEBI:115788 |
| 1 | HP:0000131 | CHEBI:122531 |
| 1 | HP:0000131 | CHEBI:122537 |
| 1 | HP:0000131 | CHEBI:129170 |
| 1 | HP:0000131 | CHEBI:122508 |
| 1 | HP:0000131 | CHEBI:115792 |
| 1 | HP:0000131 | CHEBI:115798 |
| 1 | HP:0000131 | CHEBI:115774 |
| 1 | HP:0000131 | CHEBI:101185 |
| 1 | HP:0000131 | CHEBI:42797 |
| 1 | HP:0000131 | CHEBI:122550 |
| 1 | HP:0000131 | CHEBI:122554 |
| 1 | HP:0000131 | CHEBI:122543 |
| 1 | HP:0000131 | CHEBI:122683 |
| 1 | HP:0000131 | CHEBI:100699 |
| 1 | HP:0000131 | CHEBI:122695 |
| 1 | HP:0000131 | CHEBI:100685 |
| 1 | HP:0000131 | CHEBI:122665 |
| 1 | HP:0000131 | CHEBI:115954 |
| 1 | HP:0000131 | CHEBI:115962 |
| 1 | HP:0000131 | CHEBI:122676 |
| 1 | HP:0000131 | CHEBI:122670 |
| 1 | HP:0000131 | CHEBI:122642 |
| 1 | HP:0000131 | CHEBI:122643 |
| 1 | HP:0000131 | CHEBI:115936 |
| 1 | HP:0000131 | CHEBI:122650 |
| 1 | HP:0000131 | CHEBI:115945 |
| 1 | HP:0000131 | CHEBI:115948 |
| 1 | HP:0000131 | CHEBI:122625 |
| 1 | HP:0000131 | CHEBI:115970 |
| 1 | HP:0000131 | CHEBI:115971 |
| 1 | HP:0000131 | CHEBI:103597 |
| 1 | HP:0000131 | CHEBI:122614 |
| 1 | HP:0000131 | CHEBI:100620 |
| 1 | HP:0000131 | CHEBI:100605 |
| 1 | HP:0000131 | CHEBI:122536 |
| 1 | HP:0000131 | CHEBI:129034 |
| 1 | HP:0000131 | CHEBI:129098 |
| 1 | HP:0000131 | CHEBI:101563 |
| 1 | HP:0000131 | CHEBI:101092 |
| 1 | HP:0000131 | CHEBI:122563 |
| 1 | HP:0000131 | CHEBI:122566 |
| 1 | HP:0000131 | CHEBI:122552 |
| 1 | HP:0000131 | CHEBI:122522 |
| 1 | HP:0000131 | CHEBI:122504 |
| 1 | HP:0000131 | CHEBI:122593 |
| 1 | HP:0000131 | CHEBI:122514 |
| 1 | HP:0000131 | CHEBI:100654 |
| 1 | HP:0000131 | CHEBI:100650 |
| 1 | HP:0000131 | CHEBI:100675 |
| 1 | HP:0000131 | CHEBI:100665 |
| 1 | HP:0000131 | CHEBI:122588 |
| 1 | HP:0000131 | CHEBI:100612 |
| 1 | HP:0000131 | CHEBI:122596 |
| 1 | HP:0000131 | CHEBI:103568 |
| 1 | HP:0000131 | CHEBI:103553 |
| 1 | HP:0000131 | CHEBI:103580 |
| 1 | HP:0000131 | CHEBI:103782 |
| 1 | HP:0000131 | CHEBI:115826 |
| 1 | HP:0000131 | CHEBI:100930 |
| 1 | HP:0000131 | CHEBI:115873 |
| 1 | HP:0000131 | CHEBI:115874 |
| 1 | HP:0000131 | CHEBI:115880 |
| 1 | HP:0000131 | CHEBI:115881 |
| 1 | HP:0000131 | CHEBI:103751 |
| 1 | HP:0000131 | CHEBI:103789 |
| 1 | HP:0000131 | CHEBI:101242 |
| 1 | HP:0000131 | CHEBI:115848 |
| 1 | HP:0000131 | CHEBI:115853 |
| 1 | HP:0000131 | CHEBI:115859 |
| 1 | HP:0000131 | CHEBI:101210 |
| 1 | HP:0000131 | CHEBI:115862 |
| 1 | HP:0000131 | CHEBI:115866 |
| 1 | HP:0000131 | CHEBI:101226 |
| 1 | HP:0000131 | CHEBI:122565 |
| 1 | HP:0000131 | CHEBI:115801 |
| 1 | HP:0000131 | CHEBI:115814 |
| 1 | HP:0000131 | CHEBI:115836 |
| 1 | HP:0000131 | CHEBI:103579 |
| 1 | HP:0000131 | CHEBI:103654 |
| 1 | HP:0000131 | CHEBI:115916 |
| 1 | HP:0000131 | CHEBI:115922 |
| 1 | HP:0000131 | CHEBI:115900 |
| 1 | HP:0000131 | CHEBI:115908 |
| 1 | HP:0000131 | CHEBI:101468 |
| 1 | HP:0000131 | CHEBI:103634 |
| 1 | HP:0000131 | CHEBI:103666 |
| 1 | HP:0000131 | CHEBI:103660 |
| 1 | HP:0000131 | CHEBI:103623 |
| 1 | HP:0000131 | CHEBI:129225 |
| 1 | HP:0000131 | CHEBI:103629 |
| 1 | HP:0000131 | CHEBI:103610 |
| 1 | HP:0000131 | CHEBI:101272 |
| 1 | HP:0000131 | CHEBI:101280 |
| 1 | HP:0000131 | CHEBI:103687 |
| 1 | HP:0000131 | CHEBI:115892 |
| 1 | HP:0000131 | CHEBI:103678 |
| 1 | HP:0000131 | CHEBI:129263 |
| 1 | HP:0000131 | CHEBI:115912 |
| 1 | HP:0000131 | CHEBI:101267 |
| 1 | HP:0000131 | CHEBI:115818 |
| 1 | HP:0000131 | CHEBI:115872 |
| 1 | HP:0000131 | CHEBI:115870 |
| 1 | HP:0000131 | CHEBI:115827 |
| 1 | HP:0000131 | CHEBI:115810 |
| 1 | HP:0000131 | CHEBI:115832 |
| 1 | HP:0000131 | CHEBI:100919 |
| 1 | HP:0000131 | CHEBI:115844 |
| 1 | HP:0000131 | CHEBI:101262 |
| 1 | HP:0000131 | CHEBI:115926 |
| 1 | HP:0000131 | CHEBI:115919 |
| 1 | HP:0000131 | CHEBI:115772 |
| 1 | HP:0000131 | CHEBI:115796 |
| 1 | HP:0000131 | CHEBI:115791 |
| 1 | HP:0000131 | CHEBI:115790 |
| 1 | HP:0000131 | MAXO:0000259 |
| 1 | HP:0000131 | HP:0001394 |
| 1 | HP:0000131 | HP:0100614 |
| 1 | HP:0000131 | CHEBI:101147 |
| 1 | HP:0000131 | HP:0011675 |
| 1 | HP:0000131 | HP:0001658 |
| 1 | HP:0000131 | HP:0100324 |
| 1 | HP:0000131 | CHEBI:115786 |
| 1 | HP:0000131 | CHEBI:115782 |
| 1 | HP:0000131 | CHEBI:101137 |
| 1 | HP:0000131 | CHEBI:115860 |
| 1 | HP:0000131 | CHEBI:115856 |
| 1 | HP:0000131 | CHEBI:115888 |
| 1 | HP:0000131 | CHEBI:115883 |
| 1 | HP:0000131 | CHEBI:115805 |
| 1 | HP:0000131 | CHEBI:104697 |
| 1 | HP:0000131 | CHEBI:97375 |
| 1 | HP:0000131 | CHEBI:97369 |
| 1 | HP:0000131 | CHEBI:124085 |
| 1 | HP:0000131 | CHEBI:124013 |
| 1 | HP:0000131 | CHEBI:7902 |
| 1 | HP:0000131 | CHEBI:124035 |
| 1 | HP:0000131 | CHEBI:124026 |
| 1 | HP:0000131 | CHEBI:124087 |
| 1 | HP:0000131 | CHEBI:97005 |
| 1 | HP:0000131 | CHEBI:97009 |
| 1 | HP:0000131 | CHEBI:97025 |
| 1 | HP:0000131 | CHEBI:97029 |
| 1 | HP:0000131 | CHEBI:97010 |
| 1 | HP:0000131 | CHEBI:97045 |
| 1 | HP:0000131 | CHEBI:97037 |
| 1 | HP:0000131 | CHEBI:97085 |
| 1 | HP:0000131 | CHEBI:124091 |
| 1 | HP:0000131 | HP:0002326 |
| 1 | HP:0000131 | CHEBI:129927 |
| 1 | HP:0000131 | CHEBI:115277 |
| 1 | HP:0000131 | CHEBI:115295 |
| 1 | HP:0000131 | CHEBI:115204 |
| 1 | HP:0000131 | CHEBI:96566 |
| 1 | HP:0000131 | CHEBI:96544 |
| 1 | HP:0000131 | CHEBI:123896 |
| 1 | HP:0000131 | CHEBI:115232 |
| 1 | HP:0000131 | CHEBI:101925 |
| 1 | HP:0000131 | CHEBI:123801 |
| 1 | HP:0000131 | CHEBI:115278 |
| 1 | HP:0000131 | CHEBI:115293 |
| 1 | HP:0000131 | CHEBI:115260 |
| 1 | HP:0000131 | CHEBI:115266 |
| 1 | HP:0000131 | CHEBI:124049 |
| 1 | HP:0000131 | CHEBI:124077 |
| 1 | HP:0000131 | ECTO:0070116 |
| 1 | HP:0000131 | MONDO:0005083 |
| 1 | HP:0000131 | ECTO:0070173 |
| 1 | HP:0000131 | ECTO:0070156 |
| 1 | HP:0000131 | ECTO:0070148 |
| 1 | HP:0000131 | ECTO:0070045 |
| 1 | HP:0000131 | ECTO:0070134 |
| 1 | HP:0000131 | ECTO:0070162 |
| 1 | HP:0000131 | ECTO:0070154 |
| 1 | HP:0000131 | ECTO:0070085 |
| 1 | HP:0000131 | ECTO:0070011 |
| 1 | HP:0000131 | ECTO:0070164 |
| 1 | HP:0000131 | ECTO:9000224 |
| 1 | HP:0000131 | MONDO:0006107 |
| 1 | HP:0000131 | MONDO:0006507 |
| 1 | HP:0000131 | CHEBI:98788 |
| 1 | HP:0000131 | CHEBI:97292 |
| 1 | HP:0000131 | CHEBI:97317 |
| 1 | HP:0000131 | CHEBI:97214 |
| 1 | HP:0000131 | CHEBI:123870 |
| 1 | HP:0000131 | CHEBI:117846 |
| 1 | HP:0000131 | ECTO:8000046 |
| 1 | HP:0000131 | ECTO:9000034 |
| 1 | HP:0000131 | ECTO:9001450 |
| 1 | HP:0000131 | ECTO:9000027 |
| 1 | HP:0000131 | CHEBI:129324 |
| 1 | HP:0000131 | CHEBI:123897 |
| 1 | HP:0000131 | CHEBI:95845 |
| 1 | HP:0000131 | CHEBI:123828 |
| 1 | HP:0000131 | CHEBI:95883 |
| 1 | HP:0000131 | CHEBI:123820 |
| 1 | HP:0000131 | CHEBI:123818 |
| 1 | HP:0000131 | CHEBI:123816 |
| 1 | HP:0000131 | CHEBI:95874 |
| 1 | HP:0000131 | CHEBI:123810 |
| 1 | HP:0000131 | CHEBI:96207 |
| 1 | HP:0000131 | CHEBI:117857 |
| 1 | HP:0000131 | CHEBI:117868 |
| 1 | HP:0000131 | CHEBI:117830 |
| 1 | HP:0000131 | ECTO:9000049 |
| 1 | HP:0000131 | CHEBI:102577 |
| 1 | HP:0000131 | ECTO:7000149 |
| 1 | HP:0000131 | CHEBI:123844 |
| 1 | HP:0000131 | CHEBI:95905 |
| 1 | HP:0000131 | CHEBI:129390 |
| 1 | HP:0000131 | ECTO:0500007 |
| 1 | HP:0000131 | CHEBI:95915 |
| 1 | HP:0000131 | ECTO:0500022 |
| 1 | HP:0000131 | ECTO:9000023 |
| 1 | HP:0000131 | CHEBI:95944 |
| 1 | HP:0000131 | ECTO:9000089 |
| 1 | HP:0000131 | ECTO:7000028 |
| 1 | HP:0000131 | CHEBI:123833 |
| 1 | HP:0000131 | CHEBI:95933 |
| 1 | HP:0000131 | CHEBI:123868 |
| 1 | HP:0000131 | CHEBI:129396 |
| 1 | HP:0000131 | CHEBI:117836 |
| 1 | HP:0000131 | CHEBI:117895 |
| 1 | HP:0000131 | CHEBI:95690 |
| 1 | HP:0000131 | CHEBI:96262 |
| 1 | HP:0000131 | CHEBI:96348 |
| 1 | HP:0000131 | CHEBI:104294 |
| 1 | HP:0000131 | CHEBI:104287 |
| 1 | HP:0000131 | CHEBI:117723 |
| 1 | HP:0000131 | CHEBI:117776 |
| 1 | HP:0000131 | CHEBI:104253 |
| 1 | HP:0000131 | CHEBI:102397 |
| 1 | HP:0000131 | CHEBI:103134 |
| 1 | HP:0000131 | CHEBI:102391 |
| 1 | HP:0000131 | CHEBI:96308 |
| 1 | HP:0000131 | CHEBI:95672 |
| 1 | HP:0000131 | CHEBI:117788 |
| 1 | HP:0000131 | CHEBI:97702 |
| 1 | HP:0000131 | CHEBI:104249 |
| 1 | HP:0000131 | CHEBI:117790 |
| 1 | HP:0000131 | CHEBI:117750 |
| 1 | HP:0000131 | CHEBI:117755 |
| 1 | HP:0000131 | CHEBI:104279 |
| 1 | HP:0000131 | CHEBI:104275 |
| 1 | HP:0000131 | CHEBI:117715 |
| 1 | HP:0000131 | CHEBI:103106 |
| 1 | HP:0000131 | CHEBI:117745 |
| 1 | HP:0000131 | CHEBI:103157 |
| 1 | HP:0000131 | CHEBI:117874 |
| 1 | HP:0000131 | CHEBI:117885 |
| 1 | HP:0000131 | CHEBI:117815 |
| 1 | HP:0000131 | CHEBI:117823 |
| 1 | HP:0000131 | CHEBI:97735 |
| 1 | HP:0000131 | CHEBI:95728 |
| 1 | HP:0000131 | CHEBI:95761 |
| 1 | HP:0000131 | CHEBI:117793 |
| 1 | HP:0000131 | CHEBI:124061 |
| 1 | HP:0000131 | CHEBI:124016 |
| 1 | HP:0000131 | CHEBI:124065 |
| 1 | HP:0000131 | CHEBI:124070 |
| 1 | HP:0000131 | CHEBI:124079 |
| 1 | HP:0000131 | CHEBI:95704 |
| 1 | HP:0000131 | CHEBI:124020 |
| 1 | HP:0000131 | CHEBI:117734 |
| 1 | HP:0000131 | CHEBI:124038 |
| 1 | HP:0000131 | CHEBI:124008 |
| 1 | HP:0000131 | ECTO:9002141 |
| 1 | HP:0000131 | CHEBI:129435 |
| 1 | HP:0000131 | CHEBI:127320 |
| 1 | HP:0000131 | CHEBI:123923 |
| 1 | HP:0000131 | CHEBI:127310 |
| 1 | HP:0000131 | ECTO:9000070 |
| 1 | HP:0000131 | CHEBI:127397 |
| 1 | HP:0000131 | CHEBI:96089 |
| 1 | HP:0000131 | CHEBI:123966 |
| 1 | HP:0000131 | CHEBI:123959 |
| 1 | HP:0000131 | ECTO:9002113 |
| 1 | HP:0000131 | CHEBI:123913 |
| 1 | HP:0000131 | CHEBI:96190 |
| 1 | HP:0000131 | CHEBI:96179 |
| 1 | HP:0000131 | CHEBI:96142 |
| 1 | HP:0000131 | CHEBI:95978 |
| 1 | HP:0000131 | CHEBI:96156 |
| 1 | HP:0000131 | CHEBI:96102 |
| 1 | HP:0000131 | CHEBI:104280 |
| 1 | HP:0000131 | CHEBI:117761 |
| 1 | HP:0000131 | CHEBI:104386 |
| 1 | HP:0000131 | CHEBI:97500 |
| 1 | HP:0000131 | CHEBI:97150 |
| 1 | HP:0000131 | CHEBI:104278 |
| 1 | HP:0000131 | CHEBI:104273 |
| 1 | HP:0000131 | CHEBI:97521 |
| 1 | HP:0000131 | CHEBI:97132 |
| 1 | HP:0000131 | CHEBI:104308 |
| 1 | HP:0000131 | CHEBI:104268 |
| 1 | HP:0000131 | CHEBI:104262 |
| 1 | HP:0000131 | CHEBI:103378 |
| 1 | HP:0000131 | CHEBI:97088 |
| 1 | HP:0000131 | CHEBI:103391 |
| 1 | HP:0000131 | CHEBI:97107 |
| 1 | HP:0000131 | CHEBI:104378 |
| 1 | HP:0000131 | CHEBI:103353 |
| 1 | HP:0000131 | CHEBI:104367 |
| 1 | HP:0000131 | CHEBI:104363 |
| 1 | HP:0000131 | CHEBI:97699 |
| 1 | HP:0000131 | CHEBI:104393 |
| 1 | HP:0000131 | CHEBI:101480 |
| 1 | HP:0000131 | CHEBI:101478 |
| 1 | HP:0000131 | CHEBI:101472 |
| 1 | HP:0000131 | CHEBI:104354 |
| 1 | HP:0000131 | CHEBI:97030 |
| 1 | HP:0000131 | CHEBI:113428 |
| 1 | HP:0000131 | CHEBI:104327 |
| 1 | HP:0000131 | CHEBI:97043 |
| 1 | HP:0000131 | CHEBI:113405 |
| 1 | HP:0000131 | CHEBI:97618 |
| 1 | HP:0000131 | CHEBI:97019 |
| 1 | HP:0000131 | CHEBI:113453 |
| 1 | HP:0000131 | CHEBI:113459 |
| 1 | HP:0000131 | CHEBI:104256 |
| 1 | HP:0000131 | CHEBI:113465 |
| 1 | HP:0000131 | CHEBI:104286 |
| 1 | HP:0000131 | CHEBI:104349 |
| 1 | HP:0000131 | CHEBI:104292 |
| 1 | HP:0000131 | CHEBI:113433 |
| 1 | HP:0000131 | CHEBI:103341 |
| 1 | HP:0000131 | CHEBI:103348 |
| 1 | HP:0000131 | CHEBI:97002 |
| 1 | HP:0000131 | CHEBI:103366 |
| 1 | HP:0000131 | CHEBI:10126 |
| 1 | HP:0000131 | CHEBI:124071 |
| 1 | HP:0000131 | CHEBI:104334 |
| 1 | HP:0000131 | CHEBI:97475 |
| 1 | HP:0000131 | CHEBI:104387 |
| 1 | HP:0000131 | CHEBI:104392 |
| 1 | HP:0000131 | CHEBI:97244 |
| 1 | HP:0000131 | CHEBI:98744 |
| 1 | HP:0000131 | CHEBI:97219 |
| 1 | HP:0000131 | CHEBI:98724 |
| 1 | HP:0000131 | CHEBI:97417 |
| 1 | HP:0000131 | CHEBI:97293 |
| 1 | HP:0000131 | CHEBI:97284 |
| 1 | HP:0000131 | CHEBI:102856 |
| 1 | HP:0000131 | CHEBI:97251 |
| 1 | HP:0000131 | CHEBI:97260 |
| 1 | HP:0000131 | CHEBI:97268 |
| 1 | HP:0000131 | CHEBI:97537 |
| 1 | HP:0000131 | CHEBI:97246 |
| 1 | HP:0000131 | HP:0008251 |
| 1 | HP:0000131 | CHEBI:97224 |
| 1 | HP:0000131 | CHEBI:101723 |
| 1 | HP:0000131 | CHEBI:104394 |
| 1 | HP:0000131 | CHEBI:104399 |
| 1 | HP:0000131 | CHEBI:104381 |
| 1 | HP:0000131 | CHEBI:113361 |
| 1 | HP:0000131 | CHEBI:113382 |
| 1 | HP:0000131 | CHEBI:104328 |
| 1 | HP:0000131 | CHEBI:104320 |
| 1 | HP:0000131 | CHEBI:113384 |
| 1 | HP:0000131 | CHEBI:104355 |
| 1 | HP:0000131 | CHEBI:113352 |
| 1 | HP:0000131 | CHEBI:97528 |
| 1 | HP:0000131 | CHEBI:104343 |
| 1 | HP:0000131 | CHEBI:97545 |
| 1 | HP:0000131 | CHEBI:97472 |
| 1 | HP:0000131 | CHEBI:113363 |
| 1 | HP:0000131 | CHEBI:113391 |
| 1 | HP:0000131 | CHEBI:97160 |
| 1 | HP:0000131 | CHEBI:104318 |
| 1 | HP:0000131 | CHEBI:104316 |
| 1 | HP:0000131 | CHEBI:102785 |
| 1 | HP:0000131 | CHEBI:102741 |
| 1 | HP:0000131 | CHEBI:97460 |
| 1 | HP:0000131 | CHEBI:104333 |
| 1 | HP:0000131 | CHEBI:113423 |
| 1 | HP:0000131 | CHEBI:113418 |
| 1 | HP:0000131 | CHEBI:97578 |
| 1 | HP:0000131 | CHEBI:103518 |
| 1 | HP:0000131 | CHEBI:124090 |
| 1 | HP:0000131 | CHEBI:95639 |
| 1 | HP:0000131 | CHEBI:129595 |
| 1 | HP:0000131 | CHEBI:95652 |
| 1 | HP:0000131 | CHEBI:103512 |
| 1 | HP:0000131 | CHEBI:96460 |
| 1 | HP:0000131 | CHEBI:97046 |
| 1 | HP:0000131 | CHEBI:97017 |
| 1 | HP:0000131 | CHEBI:96448 |
| 1 | HP:0000131 | CHEBI:103522 |
| 1 | HP:0000131 | CHEBI:96452 |
| 1 | HP:0000131 | CHEBI:129509 |
| 1 | HP:0000131 | CHEBI:97026 |
| 1 | HP:0000131 | CHEBI:97001 |
| 1 | HP:0000131 | CHEBI:104336 |
| 1 | HP:0000131 | CHEBI:103452 |
| 1 | HP:0000131 | CHEBI:103459 |
| 1 | HP:0000131 | CHEBI:103463 |
| 1 | HP:0000131 | CHEBI:124083 |
| 1 | HP:0000131 | CHEBI:95610 |
| 1 | HP:0000131 | CHEBI:95618 |
| 1 | HP:0000131 | CHEBI:97714 |
| 1 | HP:0000131 | CHEBI:124076 |
| 1 | HP:0000131 | CHEBI:124047 |
| 1 | HP:0000131 | CHEBI:124043 |
| 1 | HP:0000131 | CHEBI:102350 |
| 1 | HP:0000131 | CHEBI:104269 |
| 1 | HP:0000131 | CHEBI:124053 |
| 1 | HP:0000131 | CHEBI:103500 |
| 1 | HP:0000131 | CHEBI:124029 |
| 1 | HP:0000131 | CHEBI:124039 |
| 1 | HP:0000131 | CHEBI:95602 |
| 1 | HP:0000131 | CHEBI:124033 |
| 1 | HP:0000131 | CHEBI:124009 |
| 1 | HP:0000131 | CHEBI:124017 |
| 1 | HP:0000131 | CHEBI:124011 |
| 1 | HP:0000131 | CHEBI:103533 |
| 1 | HP:0000131 | CHEBI:102364 |
| 1 | HP:0000131 | CHEBI:129557 |
| 1 | HP:0000131 | CHEBI:103539 |
| 1 | HP:0000131 | CHEBI:103468 |
| 1 | HP:0000131 | CHEBI:104322 |
| 1 | HP:0000131 | CHEBI:103437 |
| 1 | HP:0000131 | CHEBI:94772 |
| 1 | HP:0000131 | CHEBI:104303 |
| 1 | HP:0000131 | CHEBI:103405 |
| 1 | HP:0000131 | CHEBI:104252 |
| 1 | HP:0000131 | CHEBI:97679 |
| 1 | HP:0000131 | CHEBI:104376 |
| 1 | HP:0000131 | CHEBI:104267 |
| 1 | HP:0000131 | CHEBI:104261 |
| 1 | HP:0000131 | CHEBI:103399 |
| 1 | HP:0000131 | CHEBI:104360 |
| 1 | HP:0000131 | CHEBI:113508 |
| 1 | HP:0000131 | CHEBI:97734 |
| 1 | HP:0000131 | CHEBI:113492 |
| 1 | HP:0000131 | CHEBI:113498 |
| 1 | HP:0000131 | CHEBI:113474 |
| 1 | HP:0000131 | CHEBI:104382 |
| 1 | HP:0000131 | CHEBI:113484 |
| 1 | HP:0000131 | CHEBI:97773 |
| 1 | HP:0000131 | CHEBI:103335 |
| 1 | HP:0000131 | CHEBI:103408 |
| 1 | HP:0000131 | CHEBI:104291 |
| 1 | HP:0000131 | CHEBI:101594 |
| 1 | HP:0000131 | CHEBI:113515 |
| 1 | HP:0000131 | CHEBI:104357 |
| 1 | HP:0000131 | CHEBI:104344 |
| 1 | HP:0000131 | CHEBI:103493 |
| 1 | HP:0000131 | CHEBI:103499 |
| 1 | HP:0000131 | CHEBI:97638 |
| 1 | HP:0000131 | CHEBI:97665 |
| 1 | HP:0000131 | CHEBI:103472 |
| 1 | HP:0000131 | CHEBI:103479 |
| 1 | HP:0000131 | CHEBI:113523 |
| 1 | HP:0000131 | CHEBI:104285 |
| 1 | HP:0000131 | CHEBI:101527 |
| 1 | HP:0000131 | CHEBI:102956 |
| 1 | HP:0000131 | CHEBI:97083 |
| 1 | HP:0000131 | CHEBI:97772 |
| 1 | HP:0000131 | CHEBI:103413 |
| 1 | HP:0000131 | CHEBI:113500 |
| 1 | HP:0000131 | CHEBI:104299 |
| 1 | HP:0000131 | CHEBI:104289 |
| 1 | HP:0000131 | CHEBI:96024 |
| 1 | HP:0000131 | CHEBI:129714 |
| 1 | HP:0000131 | CHEBI:123652 |
| 1 | HP:0000131 | CHEBI:129718 |
| 1 | HP:0000131 | CHEBI:129746 |
| 1 | HP:0000131 | CHEBI:115123 |
| 1 | HP:0000131 | CHEBI:123659 |
| 1 | HP:0000131 | CHEBI:123661 |
| 1 | HP:0000131 | CHEBI:123638 |
| 1 | HP:0000131 | CHEBI:123642 |
| 1 | HP:0000131 | CHEBI:129723 |
| 1 | HP:0000131 | CHEBI:115189 |
| 1 | HP:0000131 | CHEBI:123953 |
| 1 | HP:0000131 | CHEBI:123954 |
| 1 | HP:0000131 | CHEBI:123957 |
| 1 | HP:0000131 | CHEBI:3804 |
| 1 | HP:0000131 | CHEBI:127128 |
| 1 | HP:0000131 | CHEBI:129763 |
| 1 | HP:0000131 | CHEBI:123789 |
| 1 | HP:0000131 | CHEBI:123780 |
| 1 | HP:0000131 | CHEBI:123702 |
| 1 | HP:0000131 | CHEBI:123753 |
| 1 | HP:0000131 | CHEBI:123705 |
| 1 | HP:0000131 | CHEBI:123758 |
| 1 | HP:0000131 | CHEBI:123709 |
| 1 | HP:0000131 | CHEBI:123708 |
| 1 | HP:0000131 | CHEBI:123698 |
| 1 | HP:0000131 | CHEBI:123697 |
| 1 | HP:0000131 | CHEBI:96641 |
| 1 | HP:0000131 | CHEBI:96658 |
| 1 | HP:0000131 | CHEBI:123692 |
| 1 | HP:0000131 | CHEBI:123688 |
| 1 | HP:0000131 | CHEBI:123983 |
| 1 | HP:0000131 | CHEBI:129873 |
| 1 | HP:0000131 | CHEBI:115225 |
| 1 | HP:0000131 | CHEBI:96537 |
| 1 | HP:0000131 | CHEBI:123831 |
| 1 | HP:0000131 | CHEBI:123881 |
| 1 | HP:0000131 | CHEBI:123852 |
| 1 | HP:0000131 | CHEBI:127286 |
| 1 | HP:0000131 | CHEBI:123858 |
| 1 | HP:0000131 | CHEBI:123862 |
| 1 | HP:0000131 | CHEBI:123864 |
| 1 | HP:0000131 | CHEBI:115243 |
| 1 | HP:0000131 | CHEBI:123839 |
| 1 | HP:0000131 | CHEBI:123861 |
| 1 | HP:0000131 | CHEBI:115216 |
| 1 | HP:0000131 | CHEBI:123926 |
| 1 | HP:0000131 | CHEBI:123900 |
| 1 | HP:0000131 | CHEBI:115131 |
| 1 | HP:0000131 | CHEBI:123908 |
| 1 | HP:0000131 | CHEBI:115136 |
| 1 | HP:0000131 | CHEBI:115138 |
| 1 | HP:0000131 | CHEBI:123901 |
| 1 | HP:0000131 | CHEBI:115146 |
| 1 | HP:0000131 | CHEBI:115198 |
| 1 | HP:0000131 | CHEBI:123999 |
| 1 | HP:0000131 | CHEBI:123929 |
| 1 | HP:0000131 | CHEBI:101956 |
| 1 | HP:0000131 | CHEBI:123935 |
| 1 | HP:0000131 | CHEBI:96627 |
| 1 | HP:0000131 | CHEBI:123941 |
| 1 | HP:0000131 | CHEBI:123922 |
| 1 | HP:0000131 | CHEBI:123940 |
| 1 | HP:0000131 | CHEBI:123947 |
| 1 | HP:0000131 | CHEBI:123915 |
| 1 | HP:0000131 | CHEBI:96758 |
| 1 | HP:0000131 | CHEBI:96847 |
| 1 | HP:0000131 | CHEBI:96805 |
| 1 | HP:0000131 | CHEBI:96813 |
| 1 | HP:0000131 | CHEBI:115315 |
| 1 | HP:0000131 | CHEBI:115321 |
| 1 | HP:0000131 | CHEBI:123706 |
| 1 | HP:0000131 | CHEBI:123777 |
| 1 | HP:0000131 | CHEBI:96833 |
| 1 | HP:0000131 | CHEBI:123732 |
| 1 | HP:0000131 | CHEBI:123720 |
| 1 | HP:0000131 | CHEBI:127418 |
| 1 | HP:0000131 | CHEBI:123712 |
| 1 | HP:0000131 | CHEBI:115380 |
| 1 | HP:0000131 | CHEBI:123742 |
| 1 | HP:0000131 | CHEBI:123738 |
| 1 | HP:0000131 | CHEBI:115386 |
| 1 | HP:0000131 | CHEBI:123767 |
| 1 | HP:0000131 | CHEBI:127433 |
| 1 | HP:0000131 | CHEBI:123759 |
| 1 | HP:0000131 | CHEBI:96851 |
| 1 | HP:0000131 | CHEBI:127356 |
| 1 | HP:0000131 | CHEBI:123644 |
| 1 | HP:0000131 | CHEBI:115340 |
| 1 | HP:0000131 | CHEBI:123664 |
| 1 | HP:0000131 | CHEBI:96076 |
| 1 | HP:0000131 | CHEBI:115335 |
| 1 | HP:0000131 | CHEBI:129950 |
| 1 | HP:0000131 | CHEBI:115353 |
| 1 | HP:0000131 | CHEBI:123682 |
| 1 | HP:0000131 | CHEBI:123677 |
| 1 | HP:0000131 | CHEBI:96889 |
| 1 | HP:0000131 | CHEBI:115369 |
| 1 | HP:0000131 | CHEBI:123695 |
| 1 | HP:0000131 | CHEBI:123725 |
| 1 | HP:0000131 | CHEBI:96752 |
| 1 | HP:0000131 | CHEBI:115374 |
| 1 | HP:0000131 | CHEBI:123723 |
| 1 | HP:0000131 | CHEBI:127507 |
| 1 | HP:0000131 | CHEBI:123796 |
| 1 | HP:0000131 | CHEBI:96788 |
| 1 | HP:0000131 | CHEBI:96784 |
| 1 | HP:0000131 | CHEBI:123715 |
| 1 | HP:0000131 | CHEBI:123710 |
| 1 | HP:0000131 | CHEBI:123747 |
| 1 | HP:0000131 | CHEBI:96726 |
| 1 | HP:0000131 | CHEBI:96734 |
| 1 | HP:0000131 | CHEBI:96730 |
| 1 | HP:0000131 | CHEBI:123783 |
| 1 | HP:0000131 | CHEBI:115431 |
| 1 | HP:0000131 | CHEBI:115434 |
| 1 | HP:0000131 | CHEBI:123737 |
| 1 | HP:0000131 | CHEBI:115441 |
| 1 | HP:0000131 | CHEBI:115393 |
| 1 | HP:0000131 | CHEBI:123799 |
| 1 | HP:0000131 | CHEBI:127428 |
| 1 | HP:0000131 | CHEBI:96870 |
| 1 | HP:0000131 | CHEBI:96774 |
| 1 | HP:0000131 | CHEBI:96763 |
| 1 | HP:0000131 | CHEBI:115402 |
| 1 | HP:0000131 | CHEBI:115281 |
| 1 | HP:0000131 | MONDO:0006195 |
| 1 | HP:0000131 | MONDO:0005393 |
| 1 | HP:0000131 | CHEBI:115273 |
| 1 | HP:0000131 | MONDO:0010030 |
| 1 | HP:0000131 | MONDO:0004375 |
| 1 | HP:0000131 | MONDO:0002715 |
| 1 | HP:0000131 | CHEBI:98819 |
| 1 | HP:0000131 | CHEBI:115285 |
| 1 | HP:0000131 | CHEBI:96518 |
| 1 | HP:0000131 | MONDO:0004425 |
| 1 | HP:0000131 | CHEBI:115270 |
| 1 | HP:0000131 | ECTO:9000125 |
| 1 | HP:0000131 | ECTO:9000037 |
| 1 | HP:0000131 | ECTO:0070168 |
| 1 | HP:0000131 | ECTO:0070071 |
| 1 | HP:0000131 | ECTO:0070090 |
| 1 | HP:0000131 | ECTO:0070123 |
| 1 | HP:0000131 | ECTO:0070029 |
| 1 | HP:0000131 | ECTO:9002169 |
| 1 | HP:0000131 | ECTO:0070038 |
| 1 | HP:0000131 | CHEBI:115256 |
| 1 | HP:0000131 | ECTO:0070152 |
| 1 | HP:0000131 | CHEBI:98796 |
| 1 | HP:0000131 | ECTO:0070170 |
| 1 | HP:0000131 | CHEBI:98861 |
| 1 | HP:0000131 | CHEBI:115201 |
| 1 | HP:0000131 | CHEBI:115172 |
| 1 | HP:0000131 | CHEBI:115169 |
| 1 | HP:0000131 | CHEBI:115164 |
| 1 | HP:0000131 | CHEBI:115160 |
| 1 | HP:0000131 | CHEBI:115152 |
| 1 | HP:0000131 | CHEBI:96608 |
| 1 | HP:0000131 | CHEBI:115139 |
| 1 | HP:0000131 | CHEBI:115182 |
| 1 | HP:0000131 | CHEBI:115175 |
| 1 | HP:0000131 | CHEBI:96615 |
| 1 | HP:0000131 | CHEBI:96545 |
| 1 | HP:0000131 | CHEBI:115213 |
| 1 | HP:0000131 | CHEBI:98710 |
| 1 | HP:0000131 | CHEBI:129845 |
| 1 | HP:0000131 | CHEBI:96567 |
| 1 | HP:0000131 | CHEBI:96523 |
| 1 | HP:0000131 | CHEBI:98771 |
| 1 | HP:0000131 | CHEBI:129874 |
| 1 | HP:0000131 | CHEBI:115226 |
| 1 | HP:0000131 | CHEBI:117637 |
| 1 | HP:0000131 | CHEBI:117641 |
| 1 | HP:0000131 | CHEBI:117649 |
| 1 | HP:0000131 | CHEBI:103005 |
| 1 | HP:0000131 | CHEBI:98965 |
| 1 | HP:0000131 | CHEBI:117651 |
| 1 | HP:0000131 | CHEBI:98970 |
| 1 | HP:0000131 | CHEBI:117595 |
| 1 | HP:0000131 | CHEBI:117700 |
| 1 | HP:0000131 | CHEBI:117709 |
| 1 | HP:0000131 | CHEBI:117694 |
| 1 | HP:0000131 | CHEBI:117697 |
| 1 | HP:0000131 | CHEBI:103084 |
| 1 | HP:0000131 | CHEBI:117614 |
| 1 | HP:0000131 | CHEBI:117676 |
| 1 | HP:0000131 | CHEBI:103064 |
| 1 | HP:0000131 | CHEBI:117672 |
| 1 | HP:0000131 | CHEBI:117685 |
| 1 | HP:0000131 | CHEBI:129709 |
| 1 | HP:0000131 | CHEBI:115344 |
| 1 | HP:0000131 | CHEBI:117804 |
| 1 | HP:0000131 | CHEBI:117809 |
| 1 | HP:0000131 | CHEBI:96053 |
| 1 | HP:0000131 | CHEBI:96890 |
| 1 | HP:0000131 | CHEBI:117797 |
| 1 | HP:0000131 | CHEBI:117826 |
| 1 | HP:0000131 | CHEBI:117877 |
| 1 | HP:0000131 | CHEBI:117817 |
| 1 | HP:0000131 | CHEBI:115349 |
| 1 | HP:0000131 | CHEBI:96818 |
| 1 | HP:0000131 | CHEBI:96883 |
| 1 | HP:0000131 | CHEBI:115309 |
| 1 | HP:0000131 | CHEBI:115305 |
| 1 | HP:0000131 | CHEBI:117784 |
| 1 | HP:0000131 | CHEBI:115319 |
| 1 | HP:0000131 | CHEBI:115316 |
| 1 | HP:0000131 | CHEBI:103146 |
| 1 | HP:0000131 | CHEBI:115354 |
| 1 | HP:0000131 | CHEBI:117711 |
| 1 | HP:0000131 | CHEBI:115357 |
| 1 | HP:0000131 | CHEBI:96880 |
| 1 | HP:0000131 | CHEBI:115337 |
| 1 | HP:0000131 | CHEBI:115331 |
| 1 | HP:0000131 | CHEBI:117738 |
| 1 | HP:0000131 | CHEBI:115366 |
| 1 | HP:0000131 | CHEBI:115363 |
| 1 | HP:0000131 | CHEBI:117749 |
| 1 | HP:0000131 | CHEBI:117871 |
| 1 | HP:0000131 | ECTO:9000019 |
| 1 | HP:0000131 | ECTO:9000042 |
| 1 | HP:0000131 | CHEBI:96126 |
| 1 | HP:0000131 | CHEBI:96150 |
| 1 | HP:0000131 | CHEBI:96143 |
| 1 | HP:0000131 | ECTO:9000048 |
| 1 | HP:0000131 | ECTO:9002166 |
| 1 | HP:0000131 | ECTO:0500006 |
| 1 | HP:0000131 | ECTO:7000017 |
| 1 | HP:0000131 | ECTO:9001302 |
| 1 | HP:0000131 | ECTO:9000081 |
| 1 | HP:0000131 | ECTO:7000005 |
| 1 | HP:0000131 | ECTO:7000140 |
| 1 | HP:0000131 | ECTO:9000058 |
| 1 | HP:0000131 | ECTO:9001437 |
| 1 | HP:0000131 | CHEBI:96195 |
| 1 | HP:0000131 | CHEBI:117837 |
| 1 | HP:0000131 | CHEBI:117841 |
| 1 | HP:0000131 | CHEBI:117849 |
| 1 | HP:0000131 | CHEBI:117893 |
| 1 | HP:0000131 | CHEBI:103221 |
| 1 | HP:0000131 | CHEBI:117861 |
| 1 | HP:0000131 | CHEBI:117752 |
| 1 | HP:0000131 | CHEBI:103393 |
| 1 | HP:0000131 | CHEBI:96750 |
| 1 | HP:0000131 | CHEBI:96755 |
| 1 | HP:0000131 | CHEBI:96742 |
| 1 | HP:0000131 | CHEBI:103416 |
| 1 | HP:0000131 | CHEBI:115417 |
| 1 | HP:0000131 | CHEBI:103420 |
| 1 | HP:0000131 | CHEBI:103426 |
| 1 | HP:0000131 | CHEBI:115442 |
| 1 | HP:0000131 | CHEBI:103402 |
| 1 | HP:0000131 | CHEBI:115439 |
| 1 | HP:0000131 | CHEBI:103331 |
| 1 | HP:0000131 | CHEBI:96810 |
| 1 | HP:0000131 | CHEBI:115128 |
| 1 | HP:0000131 | CHEBI:103383 |
| 1 | HP:0000131 | CHEBI:103388 |
| 1 | HP:0000131 | CHEBI:103364 |
| 1 | HP:0000131 | CHEBI:96683 |
| 1 | HP:0000131 | CHEBI:103370 |
| 1 | HP:0000131 | CHEBI:103344 |
| 1 | HP:0000131 | CHEBI:96659 |
| 1 | HP:0000131 | CHEBI:115423 |
| 1 | HP:0000131 | CHEBI:115427 |
| 1 | HP:0000131 | CHEBI:103547 |
| 1 | HP:0000131 | CHEBI:115377 |
| 1 | HP:0000131 | CHEBI:96846 |
| 1 | HP:0000131 | CHEBI:115371 |
| 1 | HP:0000131 | CHEBI:103510 |
| 1 | HP:0000131 | CHEBI:96871 |
| 1 | HP:0000131 | CHEBI:96878 |
| 1 | HP:0000131 | CHEBI:96874 |
| 1 | HP:0000131 | CHEBI:115399 |
| 1 | HP:0000131 | CHEBI:103542 |
| 1 | HP:0000131 | CHEBI:115390 |
| 1 | HP:0000131 | CHEBI:96806 |
| 1 | HP:0000131 | CHEBI:96830 |
| 1 | HP:0000131 | CHEBI:96829 |
| 1 | HP:0000131 | CHEBI:117763 |
| 1 | HP:0000131 | CHEBI:96824 |
| 1 | HP:0000131 | CHEBI:103535 |
| 1 | HP:0000131 | CHEBI:96858 |
| 1 | HP:0000131 | CHEBI:103508 |
| 1 | HP:0000131 | CHEBI:103504 |
| 1 | HP:0000131 | CHEBI:96840 |
| 1 | HP:0000131 | CHEBI:103531 |
| 1 | HP:0000131 | CHEBI:103475 |
| 1 | HP:0000131 | CHEBI:103495 |
| 1 | HP:0000131 | CHEBI:96760 |
| 1 | HP:0000131 | CHEBI:96796 |
| 1 | HP:0000131 | CHEBI:103470 |
| 1 | HP:0000131 | CHEBI:96777 |
| 1 | HP:0000131 | CHEBI:96799 |
| 1 | HP:0000131 | CHEBI:103491 |
| 1 | HP:0000131 | CHEBI:115407 |
| 1 | HP:0000131 | CHEBI:115400 |
| 1 | HP:0000131 | CHEBI:103450 |
| 1 | HP:0000131 | CHEBI:103465 |
| 1 | HP:0000131 | CHEBI:103432 |
| 1 | HP:0000131 | CHEBI:103436 |
| 1 | HP:0000131 | CHEBI:103438 |
| 1 | HP:0000131 | CHEBI:96771 |
| 1 | HP:0000131 | CHEBI:103443 |
| 1 | HP:0000131 | CHEBI:103447 |
| 1 | HP:0000131 | CHEBI:103618 |
| 1 | HP:0000131 | CHEBI:103616 |
| 1 | HP:0000131 | CHEBI:103621 |
| 1 | HP:0000131 | CHEBI:103604 |
| 1 | HP:0000131 | CHEBI:103652 |
| 1 | HP:0000131 | CHEBI:103664 |
| 1 | HP:0000131 | CHEBI:103639 |
| 1 | HP:0000131 | CHEBI:103632 |
| 1 | HP:0000131 | CHEBI:103645 |
| 1 | HP:0000131 | CHEBI:103677 |
| 1 | HP:0000131 | CHEBI:103685 |
| 1 | HP:0000131 | CHEBI:103584 |
| 1 | HP:0000131 | CHEBI:103571 |
| 1 | HP:0000131 | CHEBI:103577 |
| 1 | HP:0000131 | CHEBI:103551 |
| 1 | HP:0000131 | CHEBI:103558 |
| 1 | HP:0000131 | CHEBI:103595 |
| 1 | HP:0000131 | CHEBI:123812 |
| 1 | HP:0000131 | CHEBI:123819 |
| 1 | HP:0000131 | CHEBI:123829 |
| 1 | HP:0000131 | CHEBI:123879 |
| 1 | HP:0000131 | CHEBI:123860 |
| 1 | HP:0000131 | CHEBI:123835 |
| 1 | HP:0000131 | CHEBI:123841 |
| 1 | HP:0000131 | CHEBI:123873 |
| 1 | HP:0000131 | CHEBI:123807 |
| 1 | HP:0000131 | ECTO:9000229 |
| 1 | HP:0000131 | ECTO:9000253 |
| 1 | HP:0000131 | CHEBI:103796 |
| 1 | HP:0000131 | ECTO:0070056 |
| 1 | HP:0000131 | MONDO:0005002 |
| 1 | HP:0000131 | MONDO:0004790 |
| 1 | HP:0000131 | MONDO:0005546 |
| 1 | HP:0000131 | ECTO:0070046 |
| 1 | HP:0000131 | ECTO:0070042 |
| 1 | HP:0000131 | ECTO:0070108 |
| 1 | HP:0000131 | ECTO:0070025 |
| 1 | HP:0000131 | ECTO:0070039 |
| 1 | HP:0000131 | ECTO:0070169 |
| 1 | HP:0000131 | ECTO:0070139 |
| 1 | HP:0000131 | CHEBI:97239 |
| 1 | HP:0000131 | CHEBI:97264 |
| 1 | HP:0000131 | CHEBI:103781 |
| 1 | HP:0000131 | CHEBI:97090 |
| 1 | HP:0000131 | CHEBI:103754 |
| 1 | HP:0000131 | CHEBI:97063 |
| 1 | HP:0000131 | CHEBI:97056 |
| 1 | HP:0000131 | CHEBI:97004 |
| 1 | HP:0000131 | CHEBI:97000 |
| 1 | HP:0000131 | CHEBI:97008 |
| 1 | HP:0000131 | CHEBI:97028 |
| 1 | HP:0000131 | CHEBI:97032 |
| 1 | HP:0000131 | CHEBI:97013 |
| 1 | HP:0000131 | CHEBI:97049 |
| 1 | HP:0000131 | CHEBI:97041 |
| 1 | HP:0000131 | CHEBI:97036 |
| 1 | HP:0000131 | HP:0002140 |
| 1 | HP:0000131 | HP:0100279 |
| 1 | HP:0000131 | ECTO:7000132 |
| 1 | HP:0000131 | ECTO:7000129 |
| 1 | HP:0000131 | ECTO:0000515 |
| 1 | HP:0000131 | ECTO:9000104 |
| 1 | HP:0000131 | ECTO:0001566 |
| 1 | HP:0000131 | ECTO:9000083 |
| 1 | HP:0000131 | ECTO:0500004 |
| 1 | HP:0000131 | ECTO:7000155 |
| 1 | HP:0000131 | CHEBI:96244 |
| 1 | HP:0000131 | ECTO:9000036 |
| 1 | HP:0000131 | CHEBI:96266 |
| 1 | HP:0000131 | ECTO:9000017 |
| 1 | HP:0000131 | CHEBI:129373 |
| 1 | HP:0000131 | CHEBI:129347 |
| 1 | HP:0000131 | ECTO:9000941 |
| 1 | HP:0000131 | CHEBI:96235 |
| 1 | HP:0000131 | HP:0001681 |
| 1 | HP:0000131 | HP:0000853 |
| 1 | HP:0000131 | HP:0000939 |
| 1 | HP:0000131 | HP:0001025 |
| 1 | HP:0000131 | CHEBI:103428 |
| 1 | HP:0000131 | CHEBI:103418 |
| 1 | HP:0000131 | CHEBI:103412 |
| 1 | HP:0000131 | CHEBI:103486 |
| 1 | HP:0000131 | CHEBI:103481 |
| 1 | HP:0000131 | CHEBI:103394 |
| 1 | HP:0000131 | CHEBI:103404 |
| 1 | HP:0000131 | CHEBI:101404 |
| 1 | HP:0000131 | CHEBI:103477 |
| 1 | HP:0000131 | CHEBI:103458 |
| 1 | HP:0000131 | CHEBI:103466 |
| 1 | HP:0000131 | CHEBI:101415 |
| 1 | HP:0000131 | CHEBI:103497 |
| 1 | HP:0000131 | CHEBI:103492 |
| 1 | HP:0000131 | CHEBI:101437 |
| 1 | HP:0000131 | CHEBI:103439 |
| 1 | HP:0000131 | CHEBI:103433 |
| 1 | HP:0000131 | CHEBI:98677 |
| 1 | HP:0000131 | CHEBI:98667 |
| 1 | HP:0000131 | CHEBI:98683 |
| 1 | HP:0000131 | CHEBI:101727 |
| 1 | HP:0000131 | CHEBI:103361 |
| 1 | HP:0000131 | CHEBI:103333 |
| 1 | HP:0000131 | CHEBI:103346 |
| 1 | HP:0000131 | CHEBI:103339 |
| 1 | HP:0000131 | CHEBI:103356 |
| 1 | HP:0000131 | CHEBI:103377 |
| 1 | HP:0000131 | CHEBI:103352 |
| 1 | HP:0000131 | CHEBI:103385 |
| 1 | HP:0000131 | CHEBI:103390 |
| 1 | HP:0000131 | CHEBI:96330 |
| 1 | HP:0000131 | CHEBI:117883 |
| 1 | HP:0000131 | CHEBI:117872 |
| 1 | HP:0000131 | CHEBI:117805 |
| 1 | HP:0000131 | CHEBI:96385 |
| 1 | HP:0000131 | CHEBI:117828 |
| 1 | HP:0000131 | CHEBI:117821 |
| 1 | HP:0000131 | CHEBI:117819 |
| 1 | HP:0000131 | CHEBI:117855 |
| 1 | HP:0000131 | CHEBI:117851 |
| 1 | HP:0000131 | CHEBI:129654 |
| 1 | HP:0000131 | CHEBI:129695 |
| 1 | HP:0000131 | CHEBI:103203 |
| 1 | HP:0000131 | CHEBI:117848 |
| 1 | HP:0000131 | CHEBI:117842 |
| 1 | HP:0000131 | CHEBI:117839 |
| 1 | HP:0000131 | CHEBI:117862 |
| 1 | HP:0000131 | CHEBI:117833 |
| 1 | HP:0000131 | CHEBI:117866 |
| 1 | HP:0000131 | CHEBI:117764 |
| 1 | HP:0000131 | CHEBI:117760 |
| 1 | HP:0000131 | CHEBI:103119 |
| 1 | HP:0000131 | CHEBI:101580 |
| 1 | HP:0000131 | CHEBI:117759 |
| 1 | HP:0000131 | CHEBI:117757 |
| 1 | HP:0000131 | CHEBI:117754 |
| 1 | HP:0000131 | CHEBI:101545 |
| 1 | HP:0000131 | CHEBI:117786 |
| 1 | HP:0000131 | CHEBI:103528 |
| 1 | HP:0000131 | CHEBI:103529 |
| 1 | HP:0000131 | CHEBI:103511 |
| 1 | HP:0000131 | CHEBI:103549 |
| 1 | HP:0000131 | CHEBI:103543 |
| 1 | HP:0000131 | CHEBI:103537 |
| 1 | HP:0000131 | CHEBI:103532 |
| 1 | HP:0000131 | CHEBI:103506 |
| 1 | HP:0000131 | CHEBI:103194 |
| 1 | HP:0000131 | CHEBI:96445 |
| 1 | HP:0000131 | CHEBI:96475 |
| 1 | HP:0000131 | CHEBI:96426 |
| 1 | HP:0000131 | CHEBI:117799 |
| 1 | HP:0000131 | CHEBI:117791 |
| 1 | HP:0000131 | CHEBI:117779 |
| 1 | HP:0000131 | CHEBI:117774 |
| 1 | HP:0000131 | CHEBI:117771 |
| 1 | HP:0000131 | CHEBI:117721 |
| 1 | HP:0000131 | CHEBI:117713 |
| 1 | HP:0000131 | ECTO:9000043 |
| 1 | HP:0000131 | ECTO:9000069 |
| 1 | HP:0000131 | MONDO:0008345 |
| 1 | HP:0000131 | MONDO:0005027 |
| 1 | HP:0000131 | ECTO:0070047 |
| 1 | HP:0000131 | CHEBI:129379 |
| 1 | HP:0000131 | MONDO:0005420 |
| 1 | HP:0000131 | ECTO:0070167 |
| 1 | HP:0000131 | ECTO:0070024 |
| 1 | HP:0000131 | ECTO:0070171 |
| 1 | HP:0000131 | ECTO:0070021 |
| 1 | HP:0000131 | ECTO:0070183 |
| 1 | HP:0000131 | ECTO:9000243 |
| 1 | HP:0000131 | ECTO:0070035 |
| 1 | HP:0000131 | ECTO:0070165 |
| 1 | HP:0000131 | ECTO:0070133 |
| 1 | HP:0000131 | ECTO:0070075 |
| 1 | HP:0000131 | ECTO:9000210 |
| 1 | HP:0000131 | CHEBI:96792 |
| 1 | HP:0000131 | CHEBI:124027 |
| 1 | HP:0000131 | CHEBI:106422 |
| 1 | HP:0000131 | CHEBI:106859 |
| 1 | HP:0000131 | CHEBI:106911 |
| 1 | HP:0000131 | HP:0030425 |
| 1 | HP:0000131 | CHEBI:106560 |
| 1 | HP:0000131 | CHEBI:106576 |
| 1 | HP:0000131 | CHEBI:110887 |
| 1 | HP:0000131 | CHEBI:106843 |
| 1 | HP:0000131 | HP:0100281 |
| 1 | HP:0000131 | CHEBI:106549 |
| 1 | HP:0000131 | CHEBI:106547 |
| 1 | HP:0000131 | CHEBI:106900 |
| 1 | HP:0000131 | CHEBI:106852 |
| 1 | HP:0000131 | CHEBI:106831 |
| 1 | HP:0000131 | CHEBI:106582 |
| 1 | HP:0000131 | CHEBI:106595 |
| 1 | HP:0000131 | CHEBI:106899 |
| 1 | HP:0000131 | HP:0002083 |
| 1 | HP:0000131 | CHEBI:106892 |
| 1 | HP:0000131 | CHEBI:106860 |
| 1 | HP:0000131 | CHEBI:106914 |
| 1 | HP:0000131 | CHEBI:110824 |
| 1 | HP:0000131 | CHEBI:110621 |
| 1 | HP:0000131 | CHEBI:127240 |
| 1 | HP:0000131 | CHEBI:110782 |
| 1 | HP:0000131 | HP:0004943 |
| 1 | HP:0000131 | CHEBI:106987 |
| 1 | HP:0000131 | CHEBI:106983 |
| 1 | HP:0000131 | CHEBI:127254 |
| 1 | HP:0000131 | CHEBI:106992 |
| 1 | HP:0000131 | CHEBI:127264 |
| 1 | HP:0000131 | CHEBI:110857 |
| 1 | HP:0000131 | CHEBI:110847 |
| 1 | HP:0000131 | CHEBI:110839 |
| 1 | HP:0000131 | CHEBI:110873 |
| 1 | HP:0000131 | CHEBI:106927 |
| 1 | HP:0000131 | CHEBI:99534 |
| 1 | HP:0000131 | CHEBI:110819 |
| 1 | HP:0000131 | CHEBI:110815 |
| 1 | HP:0000131 | CHEBI:106933 |
| 1 | HP:0000131 | CHEBI:99549 |
| 1 | HP:0000131 | CHEBI:110805 |
| 1 | HP:0000131 | CHEBI:106877 |
| 1 | HP:0000131 | CHEBI:106528 |
| 1 | HP:0000131 | CHEBI:106520 |
| 1 | HP:0000131 | CHEBI:112118 |
| 1 | HP:0000131 | CHEBI:106496 |
| 1 | HP:0000131 | CHEBI:126735 |
| 1 | HP:0000131 | CHEBI:112124 |
| 1 | HP:0000131 | CHEBI:126751 |
| 1 | HP:0000131 | CHEBI:106461 |
| 1 | HP:0000131 | CHEBI:106474 |
| 1 | HP:0000131 | CHEBI:126749 |
| 1 | HP:0000131 | CHEBI:112109 |
| 1 | HP:0000131 | CHEBI:130205 |
| 1 | HP:0000131 | CHEBI:106530 |
| 1 | HP:0000131 | CHEBI:99717 |
| 1 | HP:0000131 | CHEBI:113291 |
| 1 | HP:0000131 | CHEBI:113287 |
| 1 | HP:0000131 | CHEBI:113278 |
| 1 | HP:0000131 | CHEBI:113272 |
| 1 | HP:0000131 | CHEBI:112049 |
| 1 | HP:0000131 | CHEBI:113296 |
| 1 | HP:0000131 | CHEBI:113293 |
| 1 | HP:0000131 | CHEBI:106493 |
| 1 | HP:0000131 | CHEBI:106488 |
| 1 | HP:0000131 | CHEBI:106423 |
| 1 | HP:0000131 | CHEBI:106427 |
| 1 | HP:0000131 | CHEBI:106515 |
| 1 | HP:0000131 | CHEBI:126842 |
| 1 | HP:0000131 | CHEBI:106511 |
| 1 | HP:0000131 | CHEBI:99667 |
| 1 | HP:0000131 | CHEBI:127757 |
| 1 | HP:0000131 | CHEBI:99870 |
| 1 | HP:0000131 | CHEBI:130183 |
| 1 | HP:0000131 | CHEBI:130167 |
| 1 | HP:0000131 | CHEBI:80023 |
| 1 | HP:0000131 | CHEBI:131262 |
| 1 | HP:0000131 | CHEBI:112141 |
| 1 | HP:0000131 | CHEBI:131268 |
| 1 | HP:0000131 | HP:0012317 |
| 1 | HP:0000131 | CHEBI:131264 |
| 1 | HP:0000131 | CHEBI:112150 |
| 1 | HP:0000131 | CHEBI:106458 |
| 1 | HP:0000131 | CHEBI:131279 |
| 1 | HP:0000131 | CHEBI:110789 |
| 1 | HP:0000131 | CHEBI:110795 |
| 1 | HP:0000131 | CHEBI:110798 |
| 1 | HP:0000131 | CHEBI:106726 |
| 1 | HP:0000131 | CHEBI:110946 |
| 1 | HP:0000131 | CHEBI:106785 |
| 1 | HP:0000131 | CHEBI:106781 |
| 1 | HP:0000131 | CHEBI:106798 |
| 1 | HP:0000131 | CHEBI:112631 |
| 1 | HP:0000131 | CHEBI:112626 |
| 1 | HP:0000131 | CHEBI:112625 |
| 1 | HP:0000131 | CHEBI:112621 |
| 1 | HP:0000131 | CHEBI:112652 |
| 1 | HP:0000131 | CHEBI:106749 |
| 1 | HP:0000131 | CHEBI:106730 |
| 1 | HP:0000131 | CHEBI:106703 |
| 1 | HP:0000131 | CHEBI:106713 |
| 1 | HP:0000131 | CHEBI:106766 |
| 1 | HP:0000131 | CHEBI:131313 |
| 1 | HP:0000131 | CHEBI:130789 |
| 1 | HP:0000131 | CHEBI:112611 |
| 1 | HP:0000131 | CHEBI:112610 |
| 1 | HP:0000131 | CHEBI:110953 |
| 1 | HP:0000131 | CHEBI:110955 |
| 1 | HP:0000131 | CHEBI:110922 |
| 1 | HP:0000131 | CHEBI:112575 |
| 1 | HP:0000131 | CHEBI:110977 |
| 1 | HP:0000131 | CHEBI:127441 |
| 1 | HP:0000131 | CHEBI:99999 |
| 1 | HP:0000131 | CHEBI:110967 |
| 1 | HP:0000131 | CHEBI:110999 |
| 1 | HP:0000131 | CHEBI:130733 |
| 1 | HP:0000131 | CHEBI:112589 |
| 1 | HP:0000131 | CHEBI:112588 |
| 1 | HP:0000131 | CHEBI:127429 |
| 1 | HP:0000131 | CHEBI:130738 |
| 1 | HP:0000131 | CHEBI:110989 |
| 1 | HP:0000131 | CHEBI:112558 |
| 1 | HP:0000131 | CHEBI:112550 |
| 1 | HP:0000131 | CHEBI:112546 |
| 1 | HP:0000131 | CHEBI:112544 |
| 1 | HP:0000131 | CHEBI:94175 |
| 1 | HP:0000131 | CHEBI:110933 |
| 1 | HP:0000131 | CHEBI:112601 |
| 1 | HP:0000131 | CHEBI:106744 |
| 1 | HP:0000131 | CHEBI:110760 |
| 1 | HP:0000131 | CHEBI:107002 |
| 1 | HP:0000131 | CHEBI:131236 |
| 1 | HP:0000131 | CHEBI:110728 |
| 1 | HP:0000131 | CHEBI:110752 |
| 1 | HP:0000131 | CHEBI:110719 |
| 1 | HP:0000131 | CHEBI:110711 |
| 1 | HP:0000131 | CHEBI:131270 |
| 1 | HP:0000131 | CHEBI:106965 |
| 1 | HP:0000131 | CHEBI:130906 |
| 1 | HP:0000131 | CHEBI:107022 |
| 1 | HP:0000131 | CHEBI:106741 |
| 1 | HP:0000131 | CHEBI:106948 |
| 1 | HP:0000131 | CHEBI:106945 |
| 1 | HP:0000131 | CHEBI:107020 |
| 1 | HP:0000131 | CHEBI:107017 |
| 1 | HP:0000131 | CHEBI:110777 |
| 1 | HP:0000131 | CHEBI:110771 |
| 1 | HP:0000131 | CHEBI:110767 |
| 1 | HP:0000131 | CHEBI:110764 |
| 1 | HP:0000131 | CHEBI:106631 |
| 1 | HP:0000131 | CHEBI:106636 |
| 1 | HP:0000131 | CHEBI:106622 |
| 1 | HP:0000131 | CHEBI:106628 |
| 1 | HP:0000131 | CHEBI:106758 |
| 1 | HP:0000131 | CHEBI:106752 |
| 1 | HP:0000131 | HP:0033761 |
| 1 | HP:0000131 | CHEBI:99698 |
| 1 | HP:0000131 | CHEBI:106689 |
| 1 | HP:0000131 | CHEBI:106684 |
| 1 | HP:0000131 | CHEBI:106697 |
| 1 | HP:0000131 | CHEBI:106693 |
| 1 | HP:0000131 | CHEBI:130863 |
| 1 | HP:0000131 | CHEBI:106662 |
| 1 | HP:0000131 | CHEBI:106679 |
| 1 | HP:0000131 | CHEBI:106674 |
| 1 | HP:0000131 | CHEBI:106615 |
| 1 | HP:0000131 | CHEBI:127167 |
| 1 | HP:0000131 | CHEBI:106648 |
| 1 | HP:0000131 | CHEBI:106654 |
| 1 | HP:0000131 | CHEBI:106650 |
| 1 | HP:0000131 | CHEBI:106412 |
| 1 | HP:0000131 | CHEBI:112054 |
| 1 | HP:0000131 | CHEBI:112027 |
| 1 | HP:0000131 | CHEBI:99336 |
| 1 | HP:0000131 | CHEBI:112005 |
| 1 | HP:0000131 | CHEBI:112009 |
| 1 | HP:0000131 | CHEBI:112010 |
| 1 | HP:0000131 | CHEBI:126626 |
| 1 | HP:0000131 | CHEBI:113362 |
| 1 | HP:0000131 | CHEBI:126646 |
| 1 | HP:0000131 | CHEBI:113351 |
| 1 | HP:0000131 | CHEBI:99339 |
| 1 | HP:0000131 | CHEBI:113387 |
| 1 | HP:0000131 | CHEBI:131030 |
| 1 | HP:0000131 | CHEBI:99370 |
| 1 | HP:0000131 | CHEBI:94862 |
| 1 | HP:0000131 | HP:0009798 |
| 1 | HP:0000131 | CHEBI:94881 |
| 1 | HP:0000131 | CHEBI:131015 |
| 1 | HP:0000131 | CHEBI:126540 |
| 1 | HP:0000131 | CHEBI:99368 |
| 1 | HP:0000131 | CHEBI:131045 |
| 1 | HP:0000131 | CHEBI:131102 |
| 1 | HP:0000131 | CHEBI:99329 |
| 1 | HP:0000131 | CHEBI:101818 |
| 1 | HP:0000131 | CHEBI:101811 |
| 1 | HP:0000131 | CHEBI:99428 |
| 1 | HP:0000131 | CHEBI:131076 |
| 1 | HP:0000131 | CHEBI:99447 |
| 1 | HP:0000131 | CHEBI:99460 |
| 1 | HP:0000131 | CHEBI:131069 |
| 1 | HP:0000131 | CHEBI:126346 |
| 1 | HP:0000131 | CHEBI:107006 |
| 1 | HP:0000131 | CHEBI:99289 |
| 1 | HP:0000131 | CHEBI:99388 |
| 1 | HP:0000131 | CHEBI:107013 |
| 1 | HP:0000131 | CHEBI:126274 |
| 1 | HP:0000131 | CHEBI:126266 |
| 1 | HP:0000131 | CHEBI:101860 |
| 1 | HP:0000131 | CHEBI:99389 |
| 1 | HP:0000131 | CHEBI:101885 |
| 1 | HP:0000131 | CHEBI:99334 |
| 1 | HP:0000131 | CHEBI:101829 |
| 1 | HP:0000131 | CHEBI:101902 |
| 1 | HP:0000131 | CHEBI:99249 |
| 1 | HP:0000131 | CHEBI:99484 |
| 1 | HP:0000131 | CHEBI:110547 |
| 1 | HP:0000131 | CHEBI:110583 |
| 1 | HP:0000131 | CHEBI:110530 |
| 1 | HP:0000131 | CHEBI:110529 |
| 1 | HP:0000131 | CHEBI:110558 |
| 1 | HP:0000131 | CHEBI:113525 |
| 1 | HP:0000131 | CHEBI:113518 |
| 1 | HP:0000131 | CHEBI:113512 |
| 1 | HP:0000131 | CHEBI:99493 |
| 1 | HP:0000131 | CHEBI:127779 |
| 1 | HP:0000131 | CHEBI:113467 |
| 1 | HP:0000131 | CHEBI:110541 |
| 1 | HP:0000131 | CHEBI:131012 |
| 1 | HP:0000131 | HP:0001722 |
| 1 | HP:0000131 | CHEBI:92797 |
| 1 | HP:0000131 | CHEBI:110619 |
| 1 | HP:0000131 | CHEBI:110608 |
| 1 | HP:0000131 | CHEBI:110636 |
| 1 | HP:0000131 | CHEBI:99142 |
| 1 | HP:0000131 | CHEBI:110591 |
| 1 | HP:0000131 | CHEBI:101951 |
| 1 | HP:0000131 | CHEBI:110599 |
| 1 | HP:0000131 | CHEBI:113502 |
| 1 | HP:0000131 | CHEBI:99230 |
| 1 | HP:0000131 | CHEBI:113408 |
| 1 | HP:0000131 | CHEBI:113402 |
| 1 | HP:0000131 | CHEBI:113422 |
| 1 | HP:0000131 | CHEBI:113420 |
| 1 | HP:0000131 | CHEBI:113415 |
| 1 | HP:0000131 | CHEBI:99264 |
| 1 | HP:0000131 | CHEBI:113486 |
| 1 | HP:0000131 | CHEBI:113476 |
| 1 | HP:0000131 | CHEBI:113470 |
| 1 | HP:0000131 | CHEBI:101992 |
| 1 | HP:0000131 | CHEBI:99416 |
| 1 | HP:0000131 | CHEBI:110574 |
| 1 | HP:0000131 | CHEBI:110571 |
| 1 | HP:0000131 | CHEBI:110569 |
| 1 | HP:0000131 | CHEBI:110563 |
| 1 | HP:0000131 | CHEBI:113507 |
| 1 | HP:0000131 | CHEBI:126394 |
| 1 | HP:0000131 | CHEBI:127707 |
| 1 | HP:0000131 | CHEBI:112028 |
| 1 | HP:0000131 | CHEBI:113335 |
| 1 | HP:0000131 | CHEBI:106541 |
| 1 | HP:0000131 | CHEBI:99174 |
| 1 | HP:0000131 | CHEBI:106559 |
| 1 | HP:0000131 | CHEBI:106556 |
| 1 | HP:0000131 | CHEBI:113341 |
| 1 | HP:0000131 | CHEBI:112538 |
| 1 | HP:0000131 | CHEBI:112536 |
| 1 | HP:0000131 | CHEBI:112529 |
| 1 | HP:0000131 | CHEBI:106585 |
| 1 | HP:0000131 | CHEBI:106534 |
| 1 | HP:0000131 | CHEBI:106591 |
| 1 | HP:0000131 | CHEBI:112525 |
| 1 | HP:0000131 | CHEBI:99156 |
| 1 | HP:0000131 | CHEBI:113305 |
| 1 | HP:0000131 | CHEBI:127580 |
| 1 | HP:0000131 | CHEBI:112586 |
| 1 | HP:0000131 | CHEBI:112581 |
| 1 | HP:0000131 | CHEBI:106538 |
| 1 | HP:0000131 | CHEBI:127953 |
| 1 | HP:0000131 | CHEBI:106546 |
| 1 | HP:0000131 | CHEBI:113310 |
| 1 | HP:0000131 | CHEBI:106548 |
| 1 | HP:0000131 | CHEBI:127878 |
| 1 | HP:0000131 | CHEBI:99104 |
| 1 | HP:0000131 | CHEBI:99786 |
| 1 | HP:0000131 | CHEBI:130274 |
| 1 | HP:0000131 | CHEBI:130241 |
| 1 | HP:0000131 | CHEBI:112069 |
| 1 | HP:0000131 | CHEBI:99761 |
| 1 | HP:0000131 | HP:0012626 |
| 1 | HP:0000131 | CHEBI:112077 |
| 1 | HP:0000131 | CHEBI:112457 |
| 1 | HP:0000131 | CHEBI:112517 |
| 1 | HP:0000131 | CHEBI:112480 |
| 1 | HP:0000131 | CHEBI:112499 |
| 1 | HP:0000131 | CHEBI:112466 |
| 1 | HP:0000131 | CHEBI:112472 |
| 1 | HP:0000131 | CHEBI:106565 |
| 1 | HP:0000131 | CHEBI:106573 |
| 1 | HP:0000131 | CHEBI:127554 |
| 1 | HP:0000131 | CHEBI:99140 |
| 1 | HP:0000131 | CHEBI:130091 |
| 1 | HP:0000131 | CHEBI:112618 |
| 1 | HP:0000131 | CHEBI:106468 |
| 1 | HP:0000131 | CHEBI:112654 |
| 1 | HP:0000131 | CHEBI:99212 |
| 1 | HP:0000131 | CHEBI:106472 |
| 1 | HP:0000131 | CHEBI:127682 |
| 1 | HP:0000131 | CHEBI:112649 |
| 1 | HP:0000131 | CHEBI:112645 |
| 1 | HP:0000131 | CHEBI:99980 |
| 1 | HP:0000131 | CHEBI:112613 |
| 1 | HP:0000131 | CHEBI:106517 |
| 1 | HP:0000131 | CHEBI:112604 |
| 1 | HP:0000131 | CHEBI:126913 |
| 1 | HP:0000131 | CHEBI:106419 |
| 1 | HP:0000131 | CHEBI:106414 |
| 1 | HP:0000131 | CHEBI:99295 |
| 1 | HP:0000131 | CHEBI:99284 |
| 1 | HP:0000131 | CHEBI:99254 |
| 1 | HP:0000131 | CHEBI:99506 |
| 1 | HP:0000131 | CHEBI:106499 |
| 1 | HP:0000131 | CHEBI:106490 |
| 1 | HP:0000131 | CHEBI:106486 |
| 1 | HP:0000131 | CHEBI:106430 |
| 1 | HP:0000131 | CHEBI:106514 |
| 1 | HP:0000131 | CHEBI:106510 |
| 1 | HP:0000131 | CHEBI:127518 |
| 1 | HP:0000131 | CHEBI:112554 |
| 1 | HP:0000131 | CHEBI:112540 |
| 1 | HP:0000131 | CHEBI:112562 |
| 1 | HP:0000131 | CHEBI:99884 |
| 1 | HP:0000131 | CHEBI:112638 |
| 1 | HP:0000131 | CHEBI:112636 |
| 1 | HP:0000131 | CHEBI:106446 |
| 1 | HP:0000131 | CHEBI:106440 |
| 1 | HP:0000131 | CHEBI:106457 |
| 1 | HP:0000131 | CHEBI:106452 |
| 1 | HP:0000131 | CHEBI:127692 |
| 1 | HP:0000131 | CHEBI:112655 |
| 1 | HP:0000131 | CHEBI:94348 |
| 1 | HP:0000131 | CHEBI:106436 |
| 1 | HP:0000131 | CHEBI:127432 |
| 1 | HP:0000131 | CHEBI:112522 |
| 1 | HP:0000131 | CHEBI:99994 |
| 1 | HP:0000131 | CHEBI:125738 |
| 1 | HP:0000131 | CHEBI:112097 |
| 1 | HP:0000131 | CHEBI:112095 |
| 1 | HP:0000131 | CHEBI:112088 |
| 1 | HP:0000131 | CHEBI:111152 |
| 1 | HP:0000131 | CHEBI:125758 |
| 1 | HP:0000131 | CHEBI:111102 |
| 1 | HP:0000131 | CHEBI:112031 |
| 1 | HP:0000131 | CHEBI:111128 |
| 1 | HP:0000131 | CHEBI:111111 |
| 1 | HP:0000131 | CHEBI:111888 |
| 1 | HP:0000131 | CHEBI:112026 |
| 1 | HP:0000131 | CHEBI:126272 |
| 1 | HP:0000131 | CHEBI:112053 |
| 1 | HP:0000131 | CHEBI:112048 |
| 1 | HP:0000131 | CHEBI:111871 |
| 1 | HP:0000131 | CHEBI:111876 |
| 1 | HP:0000131 | CHEBI:112117 |
| 1 | HP:0000131 | CHEBI:112111 |
| 1 | HP:0000131 | CHEBI:111133 |
| 1 | HP:0000131 | CHEBI:112067 |
| 1 | HP:0000131 | CHEBI:112068 |
| 1 | HP:0000131 | CHEBI:111143 |
| 1 | HP:0000131 | CHEBI:111929 |
| 1 | HP:0000131 | CHEBI:111901 |
| 1 | HP:0000131 | CHEBI:111956 |
| 1 | HP:0000131 | CHEBI:111962 |
| 1 | HP:0000131 | CHEBI:111932 |
| 1 | HP:0000131 | CHEBI:125713 |
| 1 | HP:0000131 | CHEBI:111933 |
| 1 | HP:0000131 | CHEBI:111937 |
| 1 | HP:0000131 | CHEBI:111946 |
| 1 | HP:0000131 | CHEBI:128476 |
| 1 | HP:0000131 | CHEBI:112471 |
| 1 | HP:0000131 | CHEBI:112465 |
| 1 | HP:0000131 | CHEBI:112498 |
| 1 | HP:0000131 | CHEBI:112492 |
| 1 | HP:0000131 | CHEBI:128449 |
| 1 | HP:0000131 | CHEBI:112073 |
| 1 | HP:0000131 | CHEBI:111144 |
| 1 | HP:0000131 | CHEBI:111882 |
| 1 | HP:0000131 | CHEBI:112100 |
| 1 | HP:0000131 | CHEBI:111910 |
| 1 | HP:0000131 | CHEBI:126547 |
| 1 | HP:0000131 | CHEBI:110812 |
| 1 | HP:0000131 | CHEBI:126888 |
| 1 | HP:0000131 | CHEBI:110863 |
| 1 | HP:0000131 | CHEBI:110866 |
| 1 | HP:0000131 | CHEBI:127383 |
| 1 | HP:0000131 | CHEBI:110874 |
| 1 | HP:0000131 | CHEBI:113396 |
| 1 | HP:0000131 | CHEBI:126874 |
| 1 | HP:0000131 | CHEBI:127360 |
| 1 | HP:0000131 | CHEBI:111885 |
| 1 | HP:0000131 | CHEBI:126570 |
| 1 | HP:0000131 | CHEBI:126579 |
| 1 | HP:0000131 | CHEBI:92101 |
| 1 | HP:0000131 | CHEBI:126562 |
| 1 | HP:0000131 | CHEBI:110858 |
| 1 | HP:0000131 | CHEBI:126580 |
| 1 | HP:0000131 | CHEBI:113367 |
| 1 | HP:0000131 | CHEBI:113359 |
| 1 | HP:0000131 | CHEBI:111892 |
| 1 | HP:0000131 | CHEBI:110802 |
| 1 | HP:0000131 | CHEBI:126814 |
| 1 | HP:0000131 | CHEBI:110827 |
| 1 | HP:0000131 | CHEBI:110882 |
| 1 | HP:0000131 | CHEBI:112139 |
| 1 | HP:0000131 | CHEBI:112133 |
| 1 | HP:0000131 | CHEBI:110884 |
| 1 | HP:0000131 | CHEBI:112123 |
| 1 | HP:0000131 | CHEBI:127307 |
| 1 | HP:0000131 | CHEBI:110894 |
| 1 | HP:0000131 | CHEBI:111854 |
| 1 | HP:0000131 | CHEBI:110896 |
| 1 | HP:0000131 | CHEBI:94904 |
| 1 | HP:0000131 | CHEBI:110899 |
| 1 | HP:0000131 | CHEBI:111859 |
| 1 | HP:0000131 | CHEBI:128502 |
| 1 | HP:0000131 | CHEBI:112148 |
| 1 | HP:0000131 | CHEBI:112140 |
| 1 | HP:0000131 | CHEBI:112019 |
| 1 | HP:0000131 | CHEBI:110825 |
| 1 | HP:0000131 | CHEBI:111915 |
| 1 | HP:0000131 | CHEBI:111117 |
| 1 | HP:0000131 | CHEBI:111835 |
| 1 | HP:0000131 | CHEBI:128690 |
| 1 | HP:0000131 | CHEBI:126342 |
| 1 | HP:0000131 | CHEBI:128662 |
| 1 | HP:0000131 | CHEBI:126306 |
| 1 | HP:0000131 | CHEBI:128649 |
| 1 | HP:0000131 | CHEBI:128631 |
| 1 | HP:0000131 | CHEBI:128839 |
| 1 | HP:0000131 | CHEBI:128617 |
| 1 | HP:0000131 | CHEBI:127677 |
| 1 | HP:0000131 | CHEBI:126949 |
| 1 | HP:0000131 | CHEBI:130298 |
| 1 | HP:0000131 | CHEBI:126946 |
| 1 | HP:0000131 | CHEBI:126393 |
| 1 | HP:0000131 | CHEBI:126402 |
| 1 | HP:0000131 | CHEBI:128469 |
| 1 | HP:0000131 | CHEBI:127641 |
| 1 | HP:0000131 | CHEBI:111029 |
| 1 | HP:0000131 | CHEBI:111019 |
| 1 | HP:0000131 | CHEBI:111041 |
| 1 | HP:0000131 | CHEBI:127042 |
| 1 | HP:0000131 | CHEBI:112478 |
| 1 | HP:0000131 | CHEBI:112464 |
| 1 | HP:0000131 | CHEBI:112497 |
| 1 | HP:0000131 | CHEBI:112025 |
| 1 | HP:0000131 | CHEBI:126693 |
| 1 | HP:0000131 | CHEBI:112030 |
| 1 | HP:0000131 | CHEBI:127533 |
| 1 | HP:0000131 | CHEBI:112085 |
| 1 | HP:0000131 | CHEBI:112087 |
| 1 | HP:0000131 | CHEBI:112092 |
| 1 | HP:0000131 | CHEBI:112094 |
| 1 | HP:0000131 | CHEBI:112063 |
| 1 | HP:0000131 | CHEBI:112066 |
| 1 | HP:0000131 | CHEBI:127053 |
| 1 | HP:0000131 | CHEBI:112076 |
| 1 | HP:0000131 | CHEBI:112081 |
| 1 | HP:0000131 | CHEBI:128766 |
| 1 | HP:0000131 | CHEBI:125306 |
| 1 | HP:0000131 | CHEBI:112484 |
| 1 | HP:0000131 | CHEBI:112491 |
| 1 | HP:0000131 | CHEBI:111031 |
| 1 | HP:0000131 | CHEBI:111099 |
| 1 | HP:0000131 | CHEBI:111073 |
| 1 | HP:0000131 | CHEBI:128417 |
| 1 | HP:0000131 | CHEBI:111989 |
| 1 | HP:0000131 | CHEBI:126950 |
| 1 | HP:0000131 | CHEBI:111148 |
| 1 | HP:0000131 | CHEBI:111142 |
| 1 | HP:0000131 | CHEBI:111096 |
| 1 | HP:0000131 | CHEBI:111092 |
| 1 | HP:0000131 | CHEBI:111138 |
| 1 | HP:0000131 | CHEBI:111132 |
| 1 | HP:0000131 | CHEBI:130170 |
| 1 | HP:0000131 | CHEBI:111067 |
| 1 | HP:0000131 | CHEBI:111068 |
| 1 | HP:0000131 | CHEBI:111158 |
| 1 | HP:0000131 | CHEBI:130192 |
| 1 | HP:0000131 | CHEBI:111051 |
| 1 | HP:0000131 | CHEBI:111100 |
| 1 | HP:0000131 | CHEBI:111126 |
| 1 | HP:0000131 | CHEBI:111120 |
| 1 | HP:0000131 | CHEBI:111075 |
| 1 | HP:0000131 | CHEBI:111008 |
| 1 | HP:0000131 | CHEBI:111033 |
| 1 | HP:0000131 | CHEBI:111039 |
| 1 | HP:0000131 | CHEBI:111043 |
| 1 | HP:0000131 | CHEBI:111061 |
| 1 | HP:0000131 | CHEBI:111059 |
| 1 | HP:0000131 | CHEBI:128529 |
| 1 | HP:0000131 | CHEBI:111089 |
| 1 | HP:0000131 | CHEBI:111081 |
| 1 | HP:0000131 | CHEBI:111080 |
| 1 | HP:0000131 | CHEBI:111079 |
| 1 | HP:0000131 | CHEBI:2668 |
| 1 | HP:0000131 | CHEBI:126984 |
| 1 | HP:0000131 | CHEBI:126414 |
| 1 | HP:0000131 | CHEBI:128561 |
| 1 | HP:0000131 | CHEBI:111024 |
| 1 | HP:0000131 | CHEBI:126964 |
| 1 | HP:0000131 | CHEBI:111013 |
| 1 | HP:0000131 | CHEBI:111970 |
| 1 | HP:0000131 | CHEBI:111975 |
| 1 | HP:0000131 | CHEBI:111979 |
| 1 | HP:0000131 | CHEBI:113354 |
| 1 | HP:0000131 | CHEBI:126836 |
| 1 | HP:0000131 | CHEBI:130904 |
| 1 | HP:0000131 | CHEBI:106816 |
| 1 | HP:0000131 | CHEBI:130308 |
| 1 | HP:0000131 | CHEBI:110978 |
| 1 | HP:0000131 | CHEBI:110904 |
| 1 | HP:0000131 | CHEBI:99998 |
| 1 | HP:0000131 | CHEBI:110915 |
| 1 | HP:0000131 | CHEBI:111912 |
| 1 | HP:0000131 | CHEBI:111920 |
| 1 | HP:0000131 | CHEBI:111926 |
| 1 | HP:0000131 | CHEBI:111906 |
| 1 | HP:0000131 | CHEBI:3724 |
| 1 | HP:0000131 | CHEBI:111961 |
| 1 | HP:0000131 | CHEBI:111967 |
| 1 | HP:0000131 | CHEBI:111935 |
| 1 | HP:0000131 | CHEBI:130395 |
| 1 | HP:0000131 | CHEBI:130929 |
| 1 | HP:0000131 | CHEBI:127294 |
| 1 | HP:0000131 | CHEBI:126051 |
| 1 | HP:0000131 | CHEBI:111881 |
| 1 | HP:0000131 | CHEBI:110994 |
| 1 | HP:0000131 | CHEBI:106780 |
| 1 | HP:0000131 | CHEBI:106794 |
| 1 | HP:0000131 | CHEBI:130324 |
| 1 | HP:0000131 | CHEBI:125812 |
| 1 | HP:0000131 | CHEBI:106754 |
| 1 | HP:0000131 | CHEBI:106748 |
| 1 | HP:0000131 | CHEBI:106774 |
| 1 | HP:0000131 | CHEBI:106761 |
| 1 | HP:0000131 | CHEBI:106765 |
| 1 | HP:0000131 | CHEBI:106712 |
| 1 | HP:0000131 | CHEBI:106718 |
| 1 | HP:0000131 | CHEBI:110940 |
| 1 | HP:0000131 | CHEBI:110947 |
| 1 | HP:0000131 | CHEBI:110956 |
| 1 | HP:0000131 | CHEBI:106736 |
| 1 | HP:0000131 | CHEBI:110923 |
| 1 | HP:0000131 | CHEBI:130424 |
| 1 | HP:0000131 | CHEBI:110934 |
| 1 | HP:0000131 | CHEBI:130438 |
| 1 | HP:0000131 | CHEBI:110983 |
| 1 | HP:0000131 | CHEBI:126034 |
| 1 | HP:0000131 | CHEBI:111853 |
| 1 | HP:0000131 | CHEBI:130390 |
| 1 | HP:0000131 | CHEBI:110914 |
| 1 | HP:0000131 | CHEBI:130781 |
| 1 | HP:0000131 | CHEBI:99531 |
| 1 | HP:0000131 | CHEBI:130855 |
| 1 | HP:0000131 | CHEBI:99513 |
| 1 | HP:0000131 | CHEBI:125893 |
| 1 | HP:0000131 | CHEBI:106805 |
| 1 | HP:0000131 | CHEBI:106801 |
| 1 | HP:0000131 | CHEBI:106813 |
| 1 | HP:0000131 | CHEBI:110911 |
| 1 | HP:0000131 | CHEBI:111858 |
| 1 | HP:0000131 | CHEBI:127466 |
| 1 | HP:0000131 | CHEBI:130881 |
| 1 | HP:0000131 | CHEBI:112503 |
| 1 | HP:0000131 | CHEBI:110900 |
| 1 | HP:0000131 | CHEBI:112534 |
| 1 | HP:0000131 | CHEBI:112530 |
| 1 | HP:0000131 | CHEBI:106818 |
| 1 | HP:0000131 | CHEBI:112528 |
| 1 | HP:0000131 | CHEBI:127328 |
| 1 | HP:0000131 | CHEBI:126132 |
| 1 | HP:0000131 | CHEBI:127332 |
| 1 | HP:0000131 | CHEBI:126188 |
| 1 | HP:0000131 | CHEBI:111861 |
| 1 | HP:0000131 | CHEBI:105484 |
| 1 | HP:0000131 | HP:0009805 |
| 1 | HP:0000131 | CHEBI:111896 |
| 1 | HP:0000131 | CHEBI:99001 |
| 1 | HP:0000131 | CHEBI:31836 |
| 1 | HP:0000131 | CHEBI:99092 |
| 1 | HP:0000131 | CHEBI:111843 |
| 1 | HP:0000131 | CHEBI:111847 |
| 1 | HP:0000131 | CHEBI:99089 |
| 1 | HP:0000131 | CHEBI:99080 |
| 1 | HP:0000131 | CHEBI:130978 |
| 1 | HP:0000131 | CHEBI:126176 |
| 1 | HP:0000131 | CHEBI:125980 |
| 1 | HP:0000131 | CHEBI:126158 |
| 1 | HP:0000131 | CHEBI:29631 |
| 1 | HP:0000131 | CHEBI:130938 |
| 1 | HP:0000131 | CHEBI:130369 |
| 1 | HP:0000131 | CHEBI:130314 |
| 1 | HP:0000131 | CHEBI:111839 |
| 1 | HP:0000131 | CHEBI:113401 |
| 1 | HP:0000131 | CHEBI:130581 |
| 1 | HP:0000131 | CHEBI:127459 |
| 1 | HP:0000131 | CHEBI:125975 |
| 1 | HP:0000131 | CHEBI:113444 |
| 1 | HP:0000131 | CHEBI:113434 |
| 1 | HP:0000131 | CHEBI:113462 |
| 1 | HP:0000131 | CHEBI:106917 |
| 1 | HP:0000131 | CHEBI:113407 |
| 1 | HP:0000131 | HP:0012624 |
| 1 | HP:0000131 | CHEBI:113497 |
| 1 | HP:0000131 | CHEBI:113413 |
| 1 | HP:0000131 | CHEBI:106902 |
| 1 | HP:0000131 | CHEBI:106935 |
| 1 | HP:0000131 | CHEBI:106923 |
| 1 | HP:0000131 | CHEBI:130627 |
| 1 | HP:0000131 | CHEBI:110706 |
| 1 | HP:0000131 | CHEBI:113485 |
| 1 | HP:0000131 | CHEBI:113479 |
| 1 | HP:0000131 | CHEBI:126449 |
| 1 | HP:0000131 | CHEBI:110768 |
| 1 | HP:0000131 | CHEBI:106851 |
| 1 | HP:0000131 | CHEBI:106825 |
| 1 | HP:0000131 | CHEBI:111840 |
| 1 | HP:0000131 | CHEBI:113386 |
| 1 | HP:0000131 | CHEBI:111848 |
| 1 | HP:0000131 | CHEBI:106872 |
| 1 | HP:0000131 | CHEBI:106876 |
| 1 | HP:0000131 | CHEBI:113375 |
| 1 | HP:0000131 | CHEBI:106869 |
| 1 | HP:0000131 | CHEBI:106898 |
| 1 | HP:0000131 | CHEBI:99081 |
| 1 | HP:0000131 | CHEBI:106882 |
| 1 | HP:0000131 | CHEBI:106888 |
| 1 | HP:0000131 | CHEBI:94889 |
| 1 | HP:0000131 | CHEBI:130507 |
| 1 | HP:0000131 | CHEBI:106834 |
| 1 | HP:0000131 | CHEBI:106838 |
| 1 | HP:0000131 | CHEBI:106829 |
| 1 | HP:0000131 | CHEBI:110783 |
| 1 | HP:0000131 | CHEBI:110712 |
| 1 | HP:0000131 | CHEBI:110742 |
| 1 | HP:0000131 | CHEBI:106688 |
| 1 | HP:0000131 | CHEBI:106647 |
| 1 | HP:0000131 | CHEBI:113517 |
| 1 | HP:0000131 | CHEBI:113511 |
| 1 | HP:0000131 | CHEBI:130668 |
| 1 | HP:0000131 | CHEBI:106630 |
| 1 | HP:0000131 | CHEBI:106635 |
| 1 | HP:0000131 | CHEBI:106627 |
| 1 | HP:0000131 | CHEBI:125860 |
| 1 | HP:0000131 | CHEBI:106641 |
| 1 | HP:0000131 | CHEBI:125851 |
| 1 | HP:0000131 | CHEBI:110746 |
| 1 | HP:0000131 | CHEBI:106611 |
| 1 | HP:0000131 | CHEBI:106604 |
| 1 | HP:0000131 | CHEBI:106673 |
| 1 | HP:0000131 | CHEBI:106678 |
| 1 | HP:0000131 | CHEBI:106661 |
| 1 | HP:0000131 | CHEBI:106665 |
| 1 | HP:0000131 | CHEBI:125877 |
| 1 | HP:0000131 | CHEBI:106683 |
| 1 | HP:0000131 | CHEBI:113524 |
| 1 | HP:0000131 | CHEBI:106961 |
| 1 | HP:0000131 | CHEBI:127477 |
| 1 | HP:0000131 | CHEBI:130658 |
| 1 | HP:0000131 | CHEBI:110753 |
| 1 | HP:0000131 | CHEBI:110756 |
| 1 | HP:0000131 | CHEBI:106980 |
| 1 | HP:0000131 | CHEBI:106986 |
| 1 | HP:0000131 | CHEBI:110722 |
| 1 | HP:0000131 | CHEBI:110726 |
| 1 | HP:0000131 | CHEBI:110729 |
| 1 | HP:0000131 | CHEBI:110733 |
| 1 | HP:0000131 | CHEBI:110737 |
| 1 | HP:0000131 | CHEBI:125913 |
| 1 | HP:0000131 | CHEBI:113505 |
| 1 | HP:0000131 | CHEBI:106956 |
| 1 | HP:0000131 | CHEBI:125935 |
| 1 | HP:0000131 | CHEBI:106942 |
| 1 | HP:0000131 | CHEBI:130664 |
| 1 | HP:0000131 | CHEBI:106973 |
| 1 | HP:0000131 | CHEBI:106979 |
| 1 | HP:0000131 | CHEBI:110626 |
| 1 | HP:0000131 | CHEBI:110696 |
| 1 | HP:0000131 | CHEBI:113399 |
| 1 | HP:0000131 | CHEBI:95881 |
| 1 | HP:0000131 | CHEBI:128743 |
| 1 | HP:0000131 | CHEBI:102545 |
| 1 | HP:0000131 | CHEBI:4975 |
| 1 | HP:0000131 | CHEBI:102568 |
| 1 | HP:0000131 | CHEBI:102511 |
| 1 | HP:0000131 | CHEBI:126719 |
| 1 | HP:0000131 | CHEBI:102520 |
| 1 | HP:0000131 | CHEBI:95831 |
| 1 | HP:0000131 | CHEBI:95878 |
| 1 | HP:0000131 | CHEBI:112120 |
| 1 | HP:0000131 | CHEBI:128834 |
| 1 | HP:0000131 | CHEBI:102676 |
| 1 | HP:0000131 | CHEBI:128407 |
| 1 | HP:0000131 | CHEBI:102634 |
| 1 | HP:0000131 | CHEBI:102625 |
| 1 | HP:0000131 | CHEBI:126847 |
| 1 | HP:0000131 | CHEBI:95793 |
| 1 | HP:0000131 | CHEBI:106870 |
| 1 | HP:0000131 | CHEBI:126737 |
| 1 | HP:0000131 | CHEBI:112137 |
| 1 | HP:0000131 | CHEBI:110686 |
| 1 | HP:0000131 | CHEBI:102470 |
| 1 | HP:0000131 | CHEBI:112075 |
| 1 | HP:0000131 | CHEBI:112071 |
| 1 | HP:0000131 | CHEBI:112065 |
| 1 | HP:0000131 | CHEBI:112082 |
| 1 | HP:0000131 | CHEBI:112035 |
| 1 | HP:0000131 | CHEBI:112033 |
| 1 | HP:0000131 | CHEBI:126691 |
| 1 | HP:0000131 | CHEBI:102478 |
| 1 | HP:0000131 | CHEBI:128715 |
| 1 | HP:0000131 | CHEBI:112101 |
| 1 | HP:0000131 | CHEBI:112056 |
| 1 | HP:0000131 | CHEBI:126676 |
| 1 | HP:0000131 | CHEBI:126671 |
| 1 | HP:0000131 | CHEBI:95954 |
| 1 | HP:0000131 | CHEBI:95980 |
| 1 | HP:0000131 | CHEBI:95992 |
| 1 | HP:0000131 | HP:0004268 |
| 1 | HP:0000131 | CHEBI:112113 |
| 1 | HP:0000131 | CHEBI:106862 |
| 1 | HP:0000131 | CHEBI:106865 |
| 1 | HP:0000131 | CHEBI:128466 |
| 1 | HP:0000131 | CHEBI:112596 |
| 1 | HP:0000131 | CHEBI:128579 |
| 1 | HP:0000131 | CHEBI:106952 |
| 1 | HP:0000131 | CHEBI:112513 |
| 1 | HP:0000131 | CHEBI:112514 |
| 1 | HP:0000131 | CHEBI:112535 |
| 1 | HP:0000131 | CHEBI:106971 |
| 1 | HP:0000131 | CHEBI:106963 |
| 1 | HP:0000131 | CHEBI:106966 |
| 1 | HP:0000131 | CHEBI:112595 |
| 1 | HP:0000131 | CHEBI:106894 |
| 1 | HP:0000131 | CHEBI:112580 |
| 1 | HP:0000131 | CHEBI:112551 |
| 1 | HP:0000131 | CHEBI:112541 |
| 1 | HP:0000131 | CHEBI:95645 |
| 1 | HP:0000131 | CHEBI:112566 |
| 1 | HP:0000131 | CHEBI:102428 |
| 1 | HP:0000131 | CHEBI:102451 |
| 1 | HP:0000131 | CHEBI:112634 |
| 1 | HP:0000131 | CHEBI:106989 |
| 1 | HP:0000131 | CHEBI:128595 |
| 1 | HP:0000131 | CHEBI:102388 |
| 1 | HP:0000131 | CHEBI:106997 |
| 1 | HP:0000131 | CHEBI:106885 |
| 1 | HP:0000131 | CHEBI:128481 |
| 1 | HP:0000131 | CHEBI:102220 |
| 1 | HP:0000131 | CHEBI:106833 |
| 1 | HP:0000131 | CHEBI:102250 |
| 1 | HP:0000131 | CHEBI:106820 |
| 1 | HP:0000131 | CHEBI:106857 |
| 1 | HP:0000131 | CHEBI:106841 |
| 1 | HP:0000131 | CHEBI:95734 |
| 1 | HP:0000131 | CHEBI:95756 |
| 1 | HP:0000131 | CHEBI:128526 |
| 1 | HP:0000131 | CHEBI:106915 |
| 1 | HP:0000131 | CHEBI:106908 |
| 1 | HP:0000131 | CHEBI:128510 |
| 1 | HP:0000131 | CHEBI:106929 |
| 1 | HP:0000131 | CHEBI:102322 |
| 1 | HP:0000131 | CHEBI:95685 |
| 1 | HP:0000131 | CHEBI:95253 |
| 1 | HP:0000131 | CHEBI:112489 |
| 1 | HP:0000131 | CHEBI:112495 |
| 1 | HP:0000131 | CHEBI:99445 |
| 1 | HP:0000131 | CHEBI:128296 |
| 1 | HP:0000131 | CHEBI:127274 |
| 1 | HP:0000131 | CHEBI:95584 |
| 1 | HP:0000131 | CHEBI:127258 |
| 1 | HP:0000131 | CHEBI:102040 |
| 1 | HP:0000131 | CHEBI:128309 |
| 1 | HP:0000131 | CHEBI:127234 |
| 1 | HP:0000131 | CHEBI:131081 |
| 1 | HP:0000131 | CHEBI:102014 |
| 1 | HP:0000131 | CHEBI:128345 |
| 1 | HP:0000131 | CHEBI:126287 |
| 1 | HP:0000131 | CHEBI:99405 |
| 1 | HP:0000131 | HP:0008076 |
| 1 | HP:0000131 | CHEBI:126269 |
| 1 | HP:0000131 | CHEBI:102068 |
| 1 | HP:0000131 | CHEBI:131006 |
| 1 | HP:0000131 | CHEBI:131000 |
| 1 | HP:0000131 | CHEBI:95564 |
| 1 | HP:0000131 | CHEBI:126244 |
| 1 | HP:0000131 | CHEBI:112003 |
| 1 | HP:0000131 | CHEBI:112007 |
| 1 | HP:0000131 | CHEBI:126629 |
| 1 | HP:0000131 | CHEBI:126507 |
| 1 | HP:0000131 | CHEBI:100275 |
| 1 | HP:0000131 | CHEBI:126514 |
| 1 | HP:0000131 | CHEBI:116030 |
| 1 | HP:0000131 | CHEBI:116053 |
| 1 | HP:0000131 | CHEBI:116046 |
| 1 | HP:0000131 | CHEBI:116043 |
| 1 | HP:0000131 | CHEBI:116012 |
| 1 | HP:0000131 | CHEBI:116004 |
| 1 | HP:0000131 | HP:0033120 |
| 1 | HP:0000131 | CHEBI:126575 |
| 1 | HP:0000131 | CHEBI:130890 |
| 1 | HP:0000131 | CHEBI:101918 |
| 1 | HP:0000131 | CHEBI:99371 |
| 1 | HP:0000131 | CHEBI:116067 |
| 1 | HP:0000131 | CHEBI:131137 |
| 1 | HP:0000131 | CHEBI:101988 |
| 1 | HP:0000131 | HP:0550003 |
| 1 | HP:0000131 | CHEBI:95586 |
| 1 | HP:0000131 | CHEBI:112470 |
| 1 | HP:0000131 | CHEBI:126924 |
| 1 | HP:0000131 | CHEBI:102211 |
| 1 | HP:0000131 | CHEBI:128047 |
| 1 | HP:0000131 | CHEBI:128050 |
| 1 | HP:0000131 | CHEBI:128056 |
| 1 | HP:0000131 | CHEBI:102126 |
| 1 | HP:0000131 | CHEBI:95407 |
| 1 | HP:0000131 | CHEBI:102191 |
| 1 | HP:0000131 | CHEBI:95423 |
| 1 | HP:0000131 | CHEBI:95326 |
| 1 | HP:0000131 | CHEBI:126366 |
| 1 | HP:0000131 | CHEBI:128145 |
| 1 | HP:0000131 | CHEBI:95279 |
| 1 | HP:0000131 | CHEBI:128195 |
| 1 | HP:0000131 | CHEBI:95295 |
| 1 | HP:0000131 | CHEBI:95291 |
| 1 | HP:0000131 | CHEBI:128647 |
| 1 | HP:0000131 | CHEBI:128610 |
| 1 | HP:0000131 | CHEBI:112475 |
| 1 | HP:0000131 | CHEBI:102113 |
| 1 | HP:0000131 | CHEBI:102168 |
| 1 | HP:0000131 | CHEBI:126411 |
| 1 | HP:0000131 | CHEBI:126416 |
| 1 | HP:0000131 | CHEBI:102060 |
| 1 | HP:0000131 | CHEBI:95571 |
| 1 | HP:0000131 | CHEBI:102041 |
| 1 | HP:0000131 | CHEBI:128336 |
| 1 | HP:0000131 | CHEBI:102016 |
| 1 | HP:0000131 | CHEBI:95506 |
| 1 | HP:0000131 | CHEBI:126308 |
| 1 | HP:0000131 | CHEBI:102097 |
| 1 | HP:0000131 | CHEBI:95531 |
| 1 | HP:0000131 | CHEBI:32054 |
| 1 | HP:0000131 | CHEBI:95494 |
| 1 | HP:0000131 | CHEBI:126383 |
| 1 | HP:0000131 | CHEBI:95405 |
| 1 | HP:0000131 | CHEBI:95429 |
| 1 | HP:0000131 | CHEBI:95449 |
| 1 | HP:0000131 | CHEBI:95419 |
| 1 | HP:0000131 | CHEBI:95414 |
| 1 | HP:0000131 | CHEBI:112635 |
| 1 | HP:0000131 | CHEBI:106639 |
| 1 | HP:0000131 | CHEBI:106623 |
| 1 | HP:0000131 | CHEBI:100757 |
| 1 | HP:0000131 | CHEBI:116011 |
| 1 | HP:0000131 | CHEBI:116007 |
| 1 | HP:0000131 | CHEBI:100770 |
| 1 | HP:0000131 | CHEBI:100830 |
| 1 | HP:0000131 | CHEBI:100821 |
| 1 | HP:0000131 | CHEBI:127614 |
| 1 | HP:0000131 | CHEBI:100789 |
| 1 | HP:0000131 | CHEBI:100730 |
| 1 | HP:0000131 | CHEBI:34949 |
| 1 | HP:0000131 | CHEBI:110761 |
| 1 | HP:0000131 | CHEBI:127651 |
| 1 | HP:0000131 | CHEBI:100866 |
| 1 | HP:0000131 | CHEBI:110735 |
| 1 | HP:0000131 | CHEBI:110723 |
| 1 | HP:0000131 | CHEBI:110750 |
| 1 | HP:0000131 | CHEBI:100828 |
| 1 | HP:0000131 | CHEBI:127731 |
| 1 | HP:0000131 | CHEBI:110770 |
| 1 | HP:0000131 | CHEBI:116042 |
| 1 | HP:0000131 | CHEBI:116056 |
| 1 | HP:0000131 | CHEBI:116060 |
| 1 | HP:0000131 | CHEBI:116022 |
| 1 | HP:0000131 | CHEBI:110957 |
| 1 | HP:0000131 | CHEBI:110943 |
| 1 | HP:0000131 | CHEBI:100797 |
| 1 | HP:0000131 | CHEBI:100788 |
| 1 | HP:0000131 | CHEBI:100742 |
| 1 | HP:0000131 | CHEBI:113324 |
| 1 | HP:0000131 | CHEBI:113318 |
| 1 | HP:0000131 | CHEBI:113313 |
| 1 | HP:0000131 | CHEBI:113348 |
| 1 | HP:0000131 | CHEBI:113346 |
| 1 | HP:0000131 | CHEBI:127967 |
| 1 | HP:0000131 | CHEBI:113308 |
| 1 | HP:0000131 | CHEBI:3562 |
| 1 | HP:0000131 | CHEBI:127591 |
| 1 | HP:0000131 | CHEBI:3361 |
| 1 | HP:0000131 | CHEBI:116036 |
| 1 | HP:0000131 | CHEBI:116026 |
| 1 | HP:0000131 | CHEBI:110766 |
| 1 | HP:0000131 | CHEBI:110790 |
| 1 | HP:0000131 | CHEBI:101998 |
| 1 | HP:0000131 | CHEBI:128940 |
| 1 | HP:0000131 | CHEBI:110837 |
| 1 | HP:0000131 | CHEBI:110835 |
| 1 | HP:0000131 | CHEBI:128871 |
| 1 | HP:0000131 | CHEBI:128851 |
| 1 | HP:0000131 | CHEBI:110897 |
| 1 | HP:0000131 | CHEBI:110889 |
| 1 | HP:0000131 | CHEBI:100603 |
| 1 | HP:0000131 | CHEBI:110883 |
| 1 | HP:0000131 | CHEBI:100621 |
| 1 | HP:0000131 | CHEBI:110788 |
| 1 | HP:0000131 | CHEBI:102132 |
| 1 | HP:0000131 | CHEBI:100518 |
| 1 | HP:0000131 | CHEBI:94905 |
| 1 | HP:0000131 | CHEBI:94939 |
| 1 | HP:0000131 | CHEBI:94931 |
| 1 | HP:0000131 | CHEBI:95406 |
| 1 | HP:0000131 | CHEBI:95422 |
| 1 | HP:0000131 | CHEBI:95415 |
| 1 | HP:0000131 | CHEBI:110803 |
| 1 | HP:0000131 | CHEBI:128866 |
| 1 | HP:0000131 | CHEBI:110807 |
| 1 | HP:0000131 | CHEBI:128896 |
| 1 | HP:0000131 | CHEBI:127700 |
| 1 | HP:0000131 | CHEBI:135931 |
| 1 | HP:0000131 | CHEBI:95559 |
| 1 | HP:0000131 | CHEBI:102044 |
| 1 | HP:0000131 | CHEBI:100572 |
| 1 | HP:0000131 | CHEBI:110855 |
| 1 | HP:0000131 | CHEBI:110852 |
| 1 | HP:0000131 | CHEBI:95591 |
| 1 | HP:0000131 | CHEBI:102013 |
| 1 | HP:0000131 | CHEBI:110841 |
| 1 | HP:0000131 | CHEBI:110878 |
| 1 | HP:0000131 | CHEBI:110870 |
| 1 | HP:0000131 | CHEBI:95505 |
| 1 | HP:0000131 | CHEBI:110865 |
| 1 | HP:0000131 | CHEBI:95527 |
| 1 | HP:0000131 | CHEBI:110817 |
| 1 | HP:0000131 | CHEBI:110808 |
| 1 | HP:0000131 | CHEBI:110959 |
| 1 | HP:0000131 | CHEBI:110920 |
| 1 | HP:0000131 | CHEBI:112657 |
| 1 | HP:0000131 | CHEBI:106796 |
| 1 | HP:0000131 | CHEBI:106746 |
| 1 | HP:0000131 | CHEBI:106772 |
| 1 | HP:0000131 | CHEBI:106777 |
| 1 | HP:0000131 | CHEBI:106763 |
| 1 | HP:0000131 | CHEBI:106769 |
| 1 | HP:0000131 | CHEBI:106721 |
| 1 | HP:0000131 | CHEBI:106728 |
| 1 | HP:0000131 | HP:0002077 |
| 1 | HP:0000131 | CHEBI:106783 |
| 1 | HP:0000131 | CHEBI:107018 |
| 1 | HP:0000131 | CHEBI:129108 |
| 1 | HP:0000131 | CHEBI:106810 |
| 1 | HP:0000131 | CHEBI:106814 |
| 1 | HP:0000131 | CHEBI:106808 |
| 1 | HP:0000131 | CHEBI:107003 |
| 1 | HP:0000131 | CHEBI:107008 |
| 1 | HP:0000131 | CHEBI:129178 |
| 1 | HP:0000131 | CHEBI:107023 |
| 1 | HP:0000131 | CHEBI:106742 |
| 1 | HP:0000131 | CHEBI:106753 |
| 1 | HP:0000131 | CHEBI:106750 |
| 1 | HP:0000131 | CHEBI:102901 |
| 1 | HP:0000131 | CHEBI:112653 |
| 1 | HP:0000131 | CHEBI:106657 |
| 1 | HP:0000131 | CHEBI:106643 |
| 1 | HP:0000131 | CHEBI:112643 |
| 1 | HP:0000131 | CHEBI:106613 |
| 1 | HP:0000131 | CHEBI:106618 |
| 1 | HP:0000131 | CHEBI:112612 |
| 1 | HP:0000131 | CHEBI:106600 |
| 1 | HP:0000131 | CHEBI:112608 |
| 1 | HP:0000131 | CHEBI:112606 |
| 1 | HP:0000131 | CHEBI:106603 |
| 1 | HP:0000131 | CHEBI:106607 |
| 1 | HP:0000131 | CHEBI:106680 |
| 1 | HP:0000131 | CHEBI:106676 |
| 1 | HP:0000131 | CHEBI:106663 |
| 1 | HP:0000131 | CHEBI:106668 |
| 1 | HP:0000131 | CHEBI:102947 |
| 1 | HP:0000131 | CHEBI:107021 |
| 1 | HP:0000131 | CHEBI:126013 |
| 1 | HP:0000131 | CHEBI:110926 |
| 1 | HP:0000131 | CHEBI:113276 |
| 1 | HP:0000131 | CHEBI:101916 |
| 1 | HP:0000131 | CHEBI:110919 |
| 1 | HP:0000131 | CHEBI:110913 |
| 1 | HP:0000131 | CHEBI:110909 |
| 1 | HP:0000131 | CHEBI:113288 |
| 1 | HP:0000131 | CHEBI:113284 |
| 1 | HP:0000131 | CHEBI:113280 |
| 1 | HP:0000131 | CHEBI:113279 |
| 1 | HP:0000131 | CHEBI:110973 |
| 1 | HP:0000131 | CHEBI:129112 |
| 1 | HP:0000131 | CHEBI:110971 |
| 1 | HP:0000131 | CHEBI:110962 |
| 1 | HP:0000131 | CHEBI:110997 |
| 1 | HP:0000131 | CHEBI:100706 |
| 1 | HP:0000131 | CHEBI:110986 |
| 1 | HP:0000131 | CHEBI:110937 |
| 1 | HP:0000131 | CHEBI:110931 |
| 1 | HP:0000131 | CHEBI:113266 |
| 1 | HP:0000131 | CHEBI:101900 |
| 1 | HP:0000131 | CHEBI:129038 |
| 1 | HP:0000131 | CHEBI:101891 |
| 1 | HP:0000131 | CHEBI:129061 |
| 1 | HP:0000131 | CHEBI:129109 |
| 1 | HP:0000131 | CHEBI:126028 |
| 1 | HP:0000131 | CHEBI:129248 |
| 1 | HP:0000131 | CHEBI:129148 |
| 1 | HP:0000131 | CHEBI:129256 |
| 1 | HP:0000131 | CHEBI:129273 |
| 1 | HP:0000131 | CHEBI:126110 |
| 1 | HP:0000131 | CHEBI:135530 |
| 1 | HP:0000131 | CHEBI:126194 |
| 1 | HP:0000131 | CHEBI:126171 |
| 1 | HP:0000131 | CHEBI:126143 |
| 1 | HP:0000131 | CHEBI:126153 |
| 1 | HP:0000131 | CHEBI:129278 |
| 1 | HP:0000131 | CHEBI:129295 |
| 1 | HP:0000131 | CHEBI:101873 |
| 1 | HP:0000131 | CHEBI:127767 |
| 1 | HP:0000131 | CHEBI:129089 |
| 1 | HP:0000131 | CHEBI:99322 |
| 1 | HP:0000131 | CHEBI:126530 |
| 1 | HP:0000131 | CHEBI:126529 |
| 1 | HP:0000131 | CHEBI:102680 |
| 1 | HP:0000131 | CHEBI:95888 |
| 1 | HP:0000131 | CHEBI:99527 |
| 1 | HP:0000131 | CHEBI:125855 |
| 1 | HP:0000131 | CHEBI:99537 |
| 1 | HP:0000131 | CHEBI:127168 |
| 1 | HP:0000131 | CHEBI:99568 |
| 1 | HP:0000131 | CHEBI:95806 |
| 1 | HP:0000131 | CHEBI:95818 |
| 1 | HP:0000131 | CHEBI:125801 |
| 1 | HP:0000131 | CHEBI:100422 |
| 1 | HP:0000131 | CHEBI:125811 |
| 1 | HP:0000131 | CHEBI:95798 |
| 1 | HP:0000131 | CHEBI:100345 |
| 1 | HP:0000131 | CHEBI:94912 |
| 1 | HP:0000131 | CHEBI:94936 |
| 1 | HP:0000131 | CHEBI:95721 |
| 1 | HP:0000131 | CHEBI:95755 |
| 1 | HP:0000131 | CHEBI:127292 |
| 1 | HP:0000131 | CHEBI:9040 |
| 1 | HP:0000131 | CHEBI:95851 |
| 1 | HP:0000131 | CHEBI:95830 |
| 1 | HP:0000131 | CHEBI:102592 |
| 1 | HP:0000131 | CHEBI:125982 |
| 1 | HP:0000131 | CHEBI:99673 |
| 1 | HP:0000131 | CHEBI:125990 |
| 1 | HP:0000131 | CHEBI:102477 |
| 1 | HP:0000131 | CHEBI:102469 |
| 1 | HP:0000131 | CHEBI:127494 |
| 1 | HP:0000131 | CHEBI:127495 |
| 1 | HP:0000131 | CHEBI:102487 |
| 1 | HP:0000131 | CHEBI:102480 |
| 1 | HP:0000131 | CHEBI:99620 |
| 1 | HP:0000131 | CHEBI:95953 |
| 1 | HP:0000131 | CHEBI:95975 |
| 1 | HP:0000131 | CHEBI:131324 |
| 1 | HP:0000131 | CHEBI:125295 |
| 1 | HP:0000131 | CHEBI:127505 |
| 1 | HP:0000131 | CHEBI:95929 |
| 1 | HP:0000131 | CHEBI:102536 |
| 1 | HP:0000131 | CHEBI:100436 |
| 1 | HP:0000131 | CHEBI:113397 |
| 1 | HP:0000131 | CHEBI:127467 |
| 1 | HP:0000131 | CHEBI:113416 |
| 1 | HP:0000131 | CHEBI:113469 |
| 1 | HP:0000131 | CHEBI:102345 |
| 1 | HP:0000131 | CHEBI:113463 |
| 1 | HP:0000131 | CHEBI:102594 |
| 1 | HP:0000131 | CHEBI:113403 |
| 1 | HP:0000131 | CHEBI:125710 |
| 1 | HP:0000131 | CHEBI:102378 |
| 1 | HP:0000131 | CHEBI:125723 |
| 1 | HP:0000131 | CHEBI:95836 |
| 1 | HP:0000131 | CHEBI:102479 |
| 1 | HP:0000131 | CHEBI:95868 |
| 1 | HP:0000131 | CHEBI:95611 |
| 1 | HP:0000131 | CHEBI:113487 |
| 1 | HP:0000131 | CHEBI:95649 |
| 1 | HP:0000131 | CHEBI:113481 |
| 1 | HP:0000131 | CHEBI:113472 |
| 1 | HP:0000131 | CHEBI:125795 |
| 1 | HP:0000131 | CHEBI:111147 |
| 1 | HP:0000131 | CHEBI:113439 |
| 1 | HP:0000131 | CHEBI:113445 |
| 1 | HP:0000131 | CHEBI:102539 |
| 1 | HP:0000131 | CHEBI:111077 |
| 1 | HP:0000131 | CHEBI:113368 |
| 1 | HP:0000131 | CHEBI:127998 |
| 1 | HP:0000131 | CHEBI:111020 |
| 1 | HP:0000131 | CHEBI:113388 |
| 1 | HP:0000131 | CHEBI:113380 |
| 1 | HP:0000131 | CHEBI:111016 |
| 1 | HP:0000131 | CHEBI:111010 |
| 1 | HP:0000131 | CHEBI:111046 |
| 1 | HP:0000131 | CHEBI:113379 |
| 1 | HP:0000131 | CHEBI:113371 |
| 1 | HP:0000131 | CHEBI:111036 |
| 1 | HP:0000131 | CHEBI:94868 |
| 1 | HP:0000131 | CHEBI:111003 |
| 1 | HP:0000131 | CHEBI:111063 |
| 1 | HP:0000131 | CHEBI:95677 |
| 1 | HP:0000131 | CHEBI:111056 |
| 1 | HP:0000131 | CHEBI:111054 |
| 1 | HP:0000131 | CHEBI:127713 |
| 1 | HP:0000131 | CHEBI:99643 |
| 1 | HP:0000131 | CHEBI:95664 |
| 1 | HP:0000131 | CHEBI:94559 |
| 1 | HP:0000131 | CHEBI:106479 |
| 1 | HP:0000131 | CHEBI:106498 |
| 1 | HP:0000131 | CHEBI:106484 |
| 1 | HP:0000131 | CHEBI:106489 |
| 1 | HP:0000131 | CHEBI:106433 |
| 1 | HP:0000131 | CHEBI:106420 |
| 1 | HP:0000131 | CHEBI:101817 |
| 1 | HP:0000131 | CHEBI:106455 |
| 1 | HP:0000131 | CHEBI:101838 |
| 1 | HP:0000131 | CHEBI:113331 |
| 1 | HP:0000131 | CHEBI:99872 |
| 1 | HP:0000131 | CHEBI:113326 |
| 1 | HP:0000131 | CHEBI:99896 |
| 1 | HP:0000131 | CHEBI:113311 |
| 1 | HP:0000131 | HP:0025274 |
| 1 | HP:0000131 | CHEBI:113349 |
| 1 | HP:0000131 | CHEBI:34950 |
| 1 | HP:0000131 | CHEBI:106503 |
| 1 | HP:0000131 | CHEBI:106475 |
| 1 | HP:0000131 | HP:0003964 |
| 1 | HP:0000131 | CHEBI:113269 |
| 1 | HP:0000131 | CHEBI:135925 |
| 1 | HP:0000131 | CHEBI:110680 |
| 1 | HP:0000131 | CHEBI:110650 |
| 1 | HP:0000131 | CHEBI:127034 |
| 1 | HP:0000131 | CHEBI:110647 |
| 1 | HP:0000131 | CHEBI:127848 |
| 1 | HP:0000131 | CHEBI:127016 |
| 1 | HP:0000131 | CHEBI:110676 |
| 1 | HP:0000131 | CHEBI:127844 |
| 1 | HP:0000131 | CHEBI:110670 |
| 1 | HP:0000131 | CHEBI:113285 |
| 1 | HP:0000131 | CHEBI:99924 |
| 1 | HP:0000131 | CHEBI:113270 |
| 1 | HP:0000131 | CHEBI:106413 |
| 1 | HP:0000131 | CHEBI:106408 |
| 1 | HP:0000131 | HP:0001962 |
| 1 | HP:0000131 | CHEBI:113298 |
| 1 | HP:0000131 | CHEBI:127897 |
| 1 | HP:0000131 | CHEBI:106507 |
| 1 | HP:0000131 | CHEBI:106532 |
| 1 | HP:0000131 | CHEBI:99076 |
| 1 | HP:0000131 | CHEBI:99014 |
| 1 | HP:0000131 | CHEBI:100002 |
| 1 | HP:0000131 | CHEBI:111941 |
| 1 | HP:0000131 | CHEBI:127628 |
| 1 | HP:0000131 | CHEBI:127603 |
| 1 | HP:0000131 | CHEBI:99709 |
| 1 | HP:0000131 | CHEBI:128895 |
| 1 | HP:0000131 | CHEBI:111873 |
| 1 | HP:0000131 | CHEBI:111889 |
| 1 | HP:0000131 | CHEBI:127369 |
| 1 | HP:0000131 | CHEBI:106521 |
| 1 | HP:0000131 | CHEBI:131266 |
| 1 | HP:0000131 | CHEBI:131261 |
| 1 | HP:0000131 | CHEBI:111897 |
| 1 | HP:0000131 | CHEBI:128956 |
| 1 | HP:0000131 | CHEBI:131237 |
| 1 | HP:0000131 | CHEBI:131246 |
| 1 | HP:0000131 | CHEBI:111834 |
| 1 | HP:0000131 | CHEBI:111844 |
| 1 | HP:0000131 | CHEBI:94452 |
| 1 | HP:0000131 | CHEBI:111969 |
| 1 | HP:0000131 | CHEBI:111955 |
| 1 | HP:0000131 | CHEBI:111950 |
| 1 | HP:0000131 | CHEBI:106525 |
| 1 | HP:0000131 | CHEBI:99832 |
| 1 | HP:0000131 | CHEBI:106583 |
| 1 | HP:0000131 | CHEBI:106588 |
| 1 | HP:0000131 | CHEBI:106551 |
| 1 | HP:0000131 | CHEBI:106558 |
| 1 | HP:0000131 | CHEBI:127558 |
| 1 | HP:0000131 | CHEBI:106543 |
| 1 | HP:0000131 | CHEBI:106581 |
| 1 | HP:0000131 | CHEBI:106578 |
| 1 | HP:0000131 | CHEBI:106562 |
| 1 | HP:0000131 | CHEBI:106568 |
| 1 | HP:0000131 | CHEBI:101941 |
| 1 | HP:0000131 | CHEBI:111914 |
| 1 | HP:0000131 | CHEBI:111919 |
| 1 | HP:0000131 | CHEBI:111928 |
| 1 | HP:0000131 | CHEBI:111900 |
| 1 | HP:0000131 | CHEBI:111136 |
| 1 | HP:0000131 | CHEBI:125776 |
| 1 | HP:0000131 | CHEBI:100253 |
| 1 | HP:0000131 | CHEBI:106495 |
| 1 | HP:0000131 | CHEBI:130498 |
| 1 | HP:0000131 | CHEBI:102780 |
| 1 | HP:0000131 | CHEBI:106442 |
| 1 | HP:0000131 | CHEBI:102774 |
| 1 | HP:0000131 | CHEBI:106454 |
| 1 | HP:0000131 | CHEBI:106437 |
| 1 | HP:0000131 | CHEBI:106432 |
| 1 | HP:0000131 | CHEBI:106487 |
| 1 | HP:0000131 | CHEBI:106469 |
| 1 | HP:0000131 | CHEBI:127381 |
| 1 | HP:0000131 | CHEBI:106465 |
| 1 | HP:0000131 | CHEBI:106471 |
| 1 | HP:0000131 | CHEBI:106478 |
| 1 | HP:0000131 | HP:0100743 |
| 1 | HP:0000131 | CHEBI:100413 |
| 1 | HP:0000131 | CHEBI:99986 |
| 1 | HP:0000131 | CHEBI:130959 |
| 1 | HP:0000131 | CHEBI:130924 |
| 1 | HP:0000131 | CHEBI:128689 |
| 1 | HP:0000131 | CHEBI:127314 |
| 1 | HP:0000131 | CHEBI:111993 |
| 1 | HP:0000131 | CHEBI:111997 |
| 1 | HP:0000131 | CHEBI:111964 |
| 1 | HP:0000131 | CHEBI:111958 |
| 1 | HP:0000131 | CHEBI:130360 |
| 1 | HP:0000131 | CHEBI:111909 |
| 1 | HP:0000131 | CHEBI:111903 |
| 1 | HP:0000131 | CHEBI:106599 |
| 1 | HP:0000131 | CHEBI:99853 |
| 1 | HP:0000131 | CHEBI:111923 |
| 1 | HP:0000131 | CHEBI:111917 |
| 1 | HP:0000131 | CHEBI:100362 |
| 1 | HP:0000131 | CHEBI:99816 |
| 1 | HP:0000131 | CHEBI:106535 |
| 1 | HP:0000131 | CHEBI:106505 |
| 1 | HP:0000131 | CHEBI:106501 |
| 1 | HP:0000131 | CHEBI:100320 |
| 1 | HP:0000131 | CHEBI:111986 |
| 1 | HP:0000131 | CHEBI:99895 |
| 1 | HP:0000131 | CHEBI:128731 |
| 1 | HP:0000131 | CHEBI:130932 |
| 1 | HP:0000131 | CHEBI:106557 |
| 1 | HP:0000131 | CHEBI:127103 |
| 1 | HP:0000131 | CHEBI:116033 |
| 1 | HP:0000131 | CHEBI:127508 |
| 1 | HP:0000131 | CHEBI:116027 |
| 1 | HP:0000131 | HP:0033591 |
| 1 | HP:0000131 | CHEBI:116023 |
| 1 | HP:0000131 | CHEBI:116051 |
| 1 | HP:0000131 | CHEBI:116015 |
| 1 | HP:0000131 | CHEBI:116002 |
| 1 | HP:0000131 | CHEBI:130793 |
| 1 | HP:0000131 | CHEBI:106406 |
| 1 | HP:0000131 | CHEBI:100163 |
| 1 | HP:0000131 | CHEBI:99246 |
| 1 | HP:0000131 | CHEBI:116064 |
| 1 | HP:0000131 | CHEBI:126484 |
| 1 | HP:0000131 | CHEBI:126451 |
| 1 | HP:0000131 | CHEBI:127187 |
| 1 | HP:0000131 | CHEBI:101842 |
| 1 | HP:0000131 | CHEBI:99345 |
| 1 | HP:0000131 | CHEBI:116041 |
| 1 | HP:0000131 | CHEBI:130700 |
| 1 | HP:0000131 | CHEBI:128511 |
| 1 | HP:0000131 | CHEBI:130750 |
| 1 | HP:0000131 | CHEBI:100428 |
| 1 | HP:0000131 | CHEBI:106416 |
| 1 | HP:0000131 | CHEBI:99945 |
| 1 | HP:0000131 | CHEBI:102855 |
| 1 | HP:0000131 | CHEBI:128823 |
| 1 | HP:0000131 | CHEBI:102808 |
| 1 | HP:0000131 | CHEBI:102881 |
| 1 | HP:0000131 | CHEBI:128404 |
| 1 | HP:0000131 | CHEBI:100091 |
| 1 | HP:0000131 | CHEBI:95147 |
| 1 | HP:0000131 | CHEBI:128483 |
| 1 | HP:0000131 | CHEBI:128468 |
| 1 | HP:0000131 | CHEBI:127415 |
| 1 | HP:0000131 | CHEBI:99123 |
| 1 | HP:0000131 | CHEBI:9439 |
| 1 | HP:0000131 | CHEBI:130745 |
| 1 | HP:0000131 | CHEBI:128530 |
| 1 | HP:0000131 | CHEBI:106554 |
| 1 | HP:0000131 | CHEBI:111934 |
| 1 | HP:0000131 | CHEBI:113490 |
| 1 | HP:0000131 | CHEBI:99583 |
| 1 | HP:0000131 | CHEBI:113521 |
| 1 | HP:0000131 | CHEBI:113513 |
| 1 | HP:0000131 | CHEBI:100014 |
| 1 | HP:0000131 | CHEBI:125823 |
| 1 | HP:0000131 | CHEBI:100277 |
| 1 | HP:0000131 | CHEBI:100036 |
| 1 | HP:0000131 | CHEBI:100237 |
| 1 | HP:0000131 | CHEBI:95749 |
| 1 | HP:0000131 | CHEBI:95788 |
| 1 | HP:0000131 | CHEBI:128235 |
| 1 | HP:0000131 | CHEBI:95775 |
| 1 | HP:0000131 | CHEBI:99565 |
| 1 | HP:0000131 | CHEBI:95707 |
| 1 | HP:0000131 | CHEBI:128269 |
| 1 | HP:0000131 | CHEBI:128272 |
| 1 | HP:0000131 | CHEBI:102922 |
| 1 | HP:0000131 | CHEBI:128881 |
| 1 | HP:0000131 | CHEBI:102339 |
| 1 | HP:0000131 | HP:0001662 |
| 1 | HP:0000131 | CHEBI:93431 |
| 1 | HP:0000131 | CHEBI:102270 |
| 1 | HP:0000131 | CHEBI:102407 |
| 1 | HP:0000131 | CHEBI:80108 |
| 1 | HP:0000131 | CHEBI:125785 |
| 1 | HP:0000131 | CHEBI:111155 |
| 1 | HP:0000131 | CHEBI:111150 |
| 1 | HP:0000131 | CHEBI:111105 |
| 1 | HP:0000131 | CHEBI:125731 |
| 1 | HP:0000131 | CHEBI:111123 |
| 1 | HP:0000131 | CHEBI:102423 |
| 1 | HP:0000131 | CHEBI:111119 |
| 1 | HP:0000131 | CHEBI:111114 |
| 1 | HP:0000131 | CHEBI:102429 |
| 1 | HP:0000131 | CHEBI:102677 |
| 1 | HP:0000131 | CHEBI:102661 |
| 1 | HP:0000131 | CHEBI:100162 |
| 1 | HP:0000131 | CHEBI:102626 |
| 1 | HP:0000131 | CHEBI:100191 |
| 1 | HP:0000131 | CHEBI:100184 |
| 1 | HP:0000131 | CHEBI:128223 |
| 1 | HP:0000131 | CHEBI:128202 |
| 1 | HP:0000131 | CHEBI:111938 |
| 1 | HP:0000131 | CHEBI:111862 |
| 1 | HP:0000131 | CHEBI:102713 |
| 1 | HP:0000131 | CHEBI:102715 |
| 1 | HP:0000131 | CHEBI:102731 |
| 1 | HP:0000131 | CHEBI:102786 |
| 1 | HP:0000131 | CHEBI:102757 |
| 1 | HP:0000131 | CHEBI:111899 |
| 1 | HP:0000131 | CHEBI:111866 |
| 1 | HP:0000131 | CHEBI:111860 |
| 1 | HP:0000131 | CHEBI:111856 |
| 1 | HP:0000131 | CHEBI:128847 |
| 1 | HP:0000131 | CHEBI:111883 |
| 1 | HP:0000131 | CHEBI:99068 |
| 1 | HP:0000131 | CHEBI:99050 |
| 1 | HP:0000131 | CHEBI:102835 |
| 1 | HP:0000131 | CHEBI:102844 |
| 1 | HP:0000131 | CHEBI:106566 |
| 1 | HP:0000131 | CHEBI:111943 |
| 1 | HP:0000131 | CHEBI:106575 |
| 1 | HP:0000131 | CHEBI:130649 |
| 1 | HP:0000131 | CHEBI:111836 |
| 1 | HP:0000131 | CHEBI:128010 |
| 1 | HP:0000131 | CHEBI:130665 |
| 1 | HP:0000131 | CHEBI:128840 |
| 1 | HP:0000131 | CHEBI:128294 |
| 1 | HP:0000131 | CHEBI:102384 |
| 1 | HP:0000131 | CHEBI:128300 |
| 1 | HP:0000131 | CHEBI:102347 |
| 1 | HP:0000131 | CHEBI:128958 |
| 1 | HP:0000131 | CHEBI:99605 |
| 1 | HP:0000131 | CHEBI:95601 |
| 1 | HP:0000131 | CHEBI:95635 |
| 1 | HP:0000131 | CHEBI:102447 |
| 1 | HP:0000131 | CHEBI:102441 |
| 1 | HP:0000131 | CHEBI:131213 |
| 1 | HP:0000131 | CHEBI:131245 |
| 1 | HP:0000131 | CHEBI:131238 |
| 1 | HP:0000131 | CHEBI:131265 |
| 1 | HP:0000131 | CHEBI:130695 |
| 1 | HP:0000131 | CHEBI:128026 |
| 1 | HP:0000131 | CHEBI:112023 |
| 1 | HP:0000131 | CHEBI:112059 |
| 1 | HP:0000131 | CHEBI:112052 |
| 1 | HP:0000131 | CHEBI:111035 |
| 1 | HP:0000131 | CHEBI:130850 |
| 1 | HP:0000131 | CHEBI:106928 |
| 1 | HP:0000131 | CHEBI:106921 |
| 1 | HP:0000131 | CHEBI:111093 |
| 1 | HP:0000131 | CHEBI:111097 |
| 1 | HP:0000131 | CHEBI:106996 |
| 1 | HP:0000131 | CHEBI:111002 |
| 1 | HP:0000131 | CHEBI:106981 |
| 1 | HP:0000131 | CHEBI:100150 |
| 1 | HP:0000131 | CHEBI:106903 |
| 1 | HP:0000131 | CHEBI:111040 |
| 1 | HP:0000131 | CHEBI:111045 |
| 1 | HP:0000131 | CHEBI:111015 |
| 1 | HP:0000131 | CHEBI:111026 |
| 1 | HP:0000131 | CHEBI:106976 |
| 1 | HP:0000131 | CHEBI:130894 |
| 1 | HP:0000131 | CHEBI:106949 |
| 1 | HP:0000131 | CHEBI:106946 |
| 1 | HP:0000131 | CHEBI:106936 |
| 1 | HP:0000131 | CHEBI:111087 |
| 1 | HP:0000131 | CHEBI:112047 |
| 1 | HP:0000131 | CHEBI:91572 |
| 1 | HP:0000131 | CHEBI:106846 |
| 1 | HP:0000131 | CHEBI:106849 |
| 1 | HP:0000131 | CHEBI:100067 |
| 1 | HP:0000131 | CHEBI:106687 |
| 1 | HP:0000131 | CHEBI:106681 |
| 1 | HP:0000131 | CHEBI:106692 |
| 1 | HP:0000131 | CHEBI:106677 |
| 1 | HP:0000131 | CHEBI:106671 |
| 1 | HP:0000131 | CHEBI:106919 |
| 1 | HP:0000131 | CHEBI:106621 |
| 1 | HP:0000131 | CHEBI:106614 |
| 1 | HP:0000131 | CHEBI:106912 |
| 1 | HP:0000131 | CHEBI:111076 |
| 1 | HP:0000131 | CHEBI:106645 |
| 1 | HP:0000131 | CHEBI:106659 |
| 1 | HP:0000131 | CHEBI:106652 |
| 1 | HP:0000131 | CHEBI:106907 |
| 1 | HP:0000131 | CHEBI:106625 |
| 1 | HP:0000131 | CHEBI:106944 |
| 1 | HP:0000131 | CHEBI:106941 |
| 1 | HP:0000131 | CHEBI:106951 |
| 1 | HP:0000131 | CHEBI:106837 |
| 1 | HP:0000131 | CHEBI:106848 |
| 1 | HP:0000131 | CHEBI:106606 |
| 1 | HP:0000131 | CHEBI:100279 |
| 1 | HP:0000131 | CHEBI:106858 |
| 1 | HP:0000131 | CHEBI:100263 |
| 1 | HP:0000131 | CHEBI:126592 |
| 1 | HP:0000131 | CHEBI:106821 |
| 1 | HP:0000131 | CHEBI:106667 |
| 1 | HP:0000131 | CHEBI:106694 |
| 1 | HP:0000131 | CHEBI:106954 |
| 1 | HP:0000131 | CHEBI:106887 |
| 1 | HP:0000131 | CHEBI:106881 |
| 1 | HP:0000131 | CHEBI:106686 |
| 1 | HP:0000131 | CHEBI:106896 |
| 1 | HP:0000131 | CHEBI:106891 |
| 1 | HP:0000131 | CHEBI:106867 |
| 1 | HP:0000131 | CHEBI:106875 |
| 1 | HP:0000131 | CHEBI:112018 |
| 1 | HP:0000131 | CHEBI:106617 |
| 1 | HP:0000131 | CHEBI:106656 |
| 1 | HP:0000131 | CHEBI:106651 |
| 1 | HP:0000131 | CHEBI:126537 |
| 1 | HP:0000131 | CHEBI:106957 |
| 1 | HP:0000131 | CHEBI:106953 |
| 1 | HP:0000131 | CHEBI:106940 |
| 1 | HP:0000131 | CHEBI:106943 |
| 1 | HP:0000131 | CHEBI:106970 |
| 1 | HP:0000131 | CHEBI:106985 |
| 1 | HP:0000131 | CHEBI:106962 |
| 1 | HP:0000131 | HP:0004349 |
| 1 | HP:0000131 | CHEBI:106999 |
| 1 | HP:0000131 | CHEBI:106994 |
| 1 | HP:0000131 | CHEBI:106968 |
| 1 | HP:0000131 | CHEBI:126499 |
| 1 | HP:0000131 | CHEBI:106934 |
| 1 | HP:0000131 | CHEBI:106930 |
| 1 | HP:0000131 | CHEBI:106904 |
| 1 | HP:0000131 | CHEBI:106632 |
| 1 | HP:0000131 | CHEBI:106638 |
| 1 | HP:0000131 | CHEBI:106844 |
| 1 | HP:0000131 | CHEBI:106826 |
| 1 | HP:0000131 | CHEBI:106823 |
| 1 | HP:0000131 | CHEBI:111851 |
| 1 | HP:0000131 | CHEBI:112070 |
| 1 | HP:0000131 | CHEBI:99505 |
| 1 | HP:0000131 | CHEBI:112061 |
| 1 | HP:0000131 | CHEBI:112099 |
| 1 | HP:0000131 | CHEBI:112089 |
| 1 | HP:0000131 | CHEBI:111868 |
| 1 | HP:0000131 | CHEBI:111864 |
| 1 | HP:0000131 | CHEBI:111855 |
| 1 | HP:0000131 | CHEBI:112029 |
| 1 | HP:0000131 | CHEBI:112136 |
| 1 | HP:0000131 | CHEBI:112021 |
| 1 | HP:0000131 | CHEBI:111886 |
| 1 | HP:0000131 | CHEBI:128593 |
| 1 | HP:0000131 | CHEBI:111878 |
| 1 | HP:0000131 | CHEBI:111872 |
| 1 | HP:0000131 | CHEBI:128575 |
| 1 | HP:0000131 | CHEBI:112043 |
| 1 | HP:0000131 | CHEBI:112112 |
| 1 | HP:0000131 | CHEBI:112078 |
| 1 | HP:0000131 | CHEBI:112079 |
| 1 | HP:0000131 | CHEBI:111891 |
| 1 | HP:0000131 | CHEBI:130513 |
| 1 | HP:0000131 | CHEBI:130425 |
| 1 | HP:0000131 | CHEBI:99479 |
| 1 | HP:0000131 | CHEBI:2550 |
| 1 | HP:0000131 | CHEBI:130559 |
| 1 | HP:0000131 | CHEBI:130571 |
| 1 | HP:0000131 | CHEBI:130982 |
| 1 | HP:0000131 | CHEBI:111846 |
| 1 | HP:0000131 | CHEBI:126146 |
| 1 | HP:0000131 | CHEBI:130963 |
| 1 | HP:0000131 | CHEBI:100287 |
| 1 | HP:0000131 | CHEBI:112479 |
| 1 | HP:0000131 | CHEBI:112474 |
| 1 | HP:0000131 | CHEBI:112460 |
| 1 | HP:0000131 | CHEBI:112494 |
| 1 | HP:0000131 | CHEBI:112488 |
| 1 | HP:0000131 | CHEBI:112458 |
| 1 | HP:0000131 | CHEBI:111893 |
| 1 | HP:0000131 | CHEBI:112108 |
| 1 | HP:0000131 | CHEBI:126736 |
| 1 | HP:0000131 | CHEBI:111146 |
| 1 | HP:0000131 | CHEBI:106764 |
| 1 | HP:0000131 | CHEBI:106729 |
| 1 | HP:0000131 | CHEBI:106725 |
| 1 | HP:0000131 | CHEBI:106864 |
| 1 | HP:0000131 | CHEBI:111122 |
| 1 | HP:0000131 | CHEBI:106702 |
| 1 | HP:0000131 | CHEBI:111104 |
| 1 | HP:0000131 | CHEBI:106716 |
| 1 | HP:0000131 | CHEBI:125750 |
| 1 | HP:0000131 | CHEBI:111154 |
| 1 | HP:0000131 | CHEBI:126739 |
| 1 | HP:0000131 | CHEBI:111159 |
| 1 | HP:0000131 | CHEBI:125781 |
| 1 | HP:0000131 | CHEBI:106773 |
| 1 | HP:0000131 | CHEBI:106747 |
| 1 | HP:0000131 | CHEBI:106743 |
| 1 | HP:0000131 | CHEBI:106880 |
| 1 | HP:0000131 | CHEBI:111135 |
| 1 | HP:0000131 | CHEBI:106832 |
| 1 | HP:0000131 | CHEBI:111113 |
| 1 | HP:0000131 | CHEBI:106861 |
| 1 | HP:0000131 | CHEBI:106879 |
| 1 | HP:0000131 | CHEBI:106873 |
| 1 | HP:0000131 | CHEBI:111944 |
| 1 | HP:0000131 | CHEBI:111947 |
| 1 | HP:0000131 | CHEBI:111965 |
| 1 | HP:0000131 | CHEBI:111951 |
| 1 | HP:0000131 | CHEBI:111904 |
| 1 | HP:0000131 | CHEBI:111924 |
| 1 | HP:0000131 | CHEBI:111918 |
| 1 | HP:0000131 | CHEBI:112149 |
| 1 | HP:0000131 | CHEBI:93665 |
| 1 | HP:0000131 | CHEBI:111987 |
| 1 | HP:0000131 | CHEBI:111976 |
| 1 | HP:0000131 | CHEBI:111994 |
| 1 | HP:0000131 | CHEBI:106811 |
| 1 | HP:0000131 | CHEBI:130741 |
| 1 | HP:0000131 | HP:0032282 |
| 1 | HP:0000131 | CHEBI:106788 |
| 1 | HP:0000131 | CHEBI:106792 |
| 1 | HP:0000131 | CHEBI:128247 |
| 1 | HP:0000131 | CHEBI:106755 |
| 1 | HP:0000131 | CHEBI:106759 |
| 1 | HP:0000131 | CHEBI:110905 |
| 1 | HP:0000131 | CHEBI:99469 |
| 1 | HP:0000131 | CHEBI:112145 |
| 1 | HP:0000131 | CHEBI:112143 |
| 1 | HP:0000131 | CHEBI:110784 |
| 1 | HP:0000131 | CHEBI:110787 |
| 1 | HP:0000131 | CHEBI:126781 |
| 1 | HP:0000131 | CHEBI:110918 |
| 1 | HP:0000131 | CHEBI:110912 |
| 1 | HP:0000131 | CHEBI:110902 |
| 1 | HP:0000131 | CHEBI:110985 |
| 1 | HP:0000131 | CHEBI:110765 |
| 1 | HP:0000131 | CHEBI:110972 |
| 1 | HP:0000131 | CHEBI:126289 |
| 1 | HP:0000131 | CHEBI:110961 |
| 1 | HP:0000131 | CHEBI:110960 |
| 1 | HP:0000131 | CHEBI:110996 |
| 1 | HP:0000131 | CHEBI:110774 |
| 1 | HP:0000131 | CHEBI:128761 |
| 1 | HP:0000131 | CHEBI:99435 |
| 1 | HP:0000131 | CHEBI:99439 |
| 1 | HP:0000131 | CHEBI:126363 |
| 1 | HP:0000131 | CHEBI:112121 |
| 1 | HP:0000131 | CHEBI:110833 |
| 1 | HP:0000131 | CHEBI:112057 |
| 1 | HP:0000131 | CHEBI:110806 |
| 1 | HP:0000131 | CHEBI:110813 |
| 1 | HP:0000131 | CHEBI:112045 |
| 1 | HP:0000131 | CHEBI:110816 |
| 1 | HP:0000131 | CHEBI:126337 |
| 1 | HP:0000131 | CHEBI:110864 |
| 1 | HP:0000131 | CHEBI:110867 |
| 1 | HP:0000131 | CHEBI:126329 |
| 1 | HP:0000131 | CHEBI:110875 |
| 1 | HP:0000131 | CHEBI:112114 |
| 1 | HP:0000131 | CHEBI:110840 |
| 1 | HP:0000131 | CHEBI:112102 |
| 1 | HP:0000131 | CHEBI:99417 |
| 1 | HP:0000131 | CHEBI:112130 |
| 1 | HP:0000131 | CHEBI:112127 |
| 1 | HP:0000131 | CHEBI:128779 |
| 1 | HP:0000131 | CHEBI:110936 |
| 1 | HP:0000131 | CHEBI:131020 |
| 1 | HP:0000131 | CHEBI:130366 |
| 1 | HP:0000131 | CHEBI:110646 |
| 1 | HP:0000131 | CHEBI:106847 |
| 1 | HP:0000131 | CHEBI:99396 |
| 1 | HP:0000131 | CHEBI:112015 |
| 1 | HP:0000131 | CHEBI:128828 |
| 1 | HP:0000131 | CHEBI:110754 |
| 1 | HP:0000131 | CHEBI:110684 |
| 1 | HP:0000131 | CHEBI:130372 |
| 1 | HP:0000131 | CHEBI:110690 |
| 1 | HP:0000131 | CHEBI:105483 |
| 1 | HP:0000131 | CHEBI:106910 |
| 1 | HP:0000131 | CHEBI:110693 |
| 1 | HP:0000131 | CHEBI:106909 |
| 1 | HP:0000131 | CHEBI:110699 |
| 1 | HP:0000131 | CHEBI:126551 |
| 1 | HP:0000131 | CHEBI:106901 |
| 1 | HP:0000131 | CHEBI:106932 |
| 1 | HP:0000131 | CHEBI:110631 |
| 1 | HP:0000131 | CHEBI:112008 |
| 1 | HP:0000131 | CHEBI:112004 |
| 1 | HP:0000131 | CHEBI:110642 |
| 1 | HP:0000131 | CHEBI:110641 |
| 1 | HP:0000131 | CHEBI:110930 |
| 1 | HP:0000131 | CHEBI:110925 |
| 1 | HP:0000131 | CHEBI:126821 |
| 1 | HP:0000131 | CHEBI:99387 |
| 1 | HP:0000131 | CHEBI:110949 |
| 1 | HP:0000131 | CHEBI:110942 |
| 1 | HP:0000131 | CHEBI:126229 |
| 1 | HP:0000131 | CHEBI:106866 |
| 1 | HP:0000131 | CHEBI:106890 |
| 1 | HP:0000131 | CHEBI:106895 |
| 1 | HP:0000131 | CHEBI:110664 |
| 1 | HP:0000131 | CHEBI:106886 |
| 1 | HP:0000131 | CHEBI:106836 |
| 1 | HP:0000131 | CHEBI:110671 |
| 1 | HP:0000131 | CHEBI:110716 |
| 1 | HP:0000131 | CHEBI:106824 |
| 1 | HP:0000131 | CHEBI:106827 |
| 1 | HP:0000131 | CHEBI:112022 |
| 1 | HP:0000131 | CHEBI:110820 |
| 1 | HP:0000131 | CHEBI:126234 |
| 1 | HP:0000131 | CHEBI:126348 |
| 1 | HP:0000131 | CHEBI:99991 |
| 1 | HP:0000131 | CHEBI:130278 |
| 1 | HP:0000131 | CHEBI:112040 |
| 1 | HP:0000131 | CHEBI:112046 |
| 1 | HP:0000131 | CHEBI:106807 |
| 1 | HP:0000131 | CHEBI:126672 |
| 1 | HP:0000131 | CHEBI:112051 |
| 1 | HP:0000131 | CHEBI:112058 |
| 1 | HP:0000131 | CHEBI:126344 |
| 1 | HP:0000131 | CHEBI:112093 |
| 1 | HP:0000131 | CHEBI:106802 |
| 1 | HP:0000131 | CHEBI:106806 |
| 1 | HP:0000131 | CHEBI:112024 |
| 1 | HP:0000131 | CHEBI:126696 |
| 1 | HP:0000131 | CHEBI:112037 |
| 1 | HP:0000131 | CHEBI:126336 |
| 1 | HP:0000131 | CHEBI:112084 |
| 1 | HP:0000131 | CHEBI:126300 |
| 1 | HP:0000131 | CHEBI:112115 |
| 1 | HP:0000131 | CHEBI:125294 |
| 1 | HP:0000131 | CHEBI:112103 |
| 1 | HP:0000131 | CHEBI:112131 |
| 1 | HP:0000131 | CHEBI:106745 |
| 1 | HP:0000131 | CHEBI:126812 |
| 1 | HP:0000131 | CHEBI:106776 |
| 1 | HP:0000131 | CHEBI:106768 |
| 1 | HP:0000131 | CHEBI:128290 |
| 1 | HP:0000131 | CHEBI:106714 |
| 1 | HP:0000131 | CHEBI:106709 |
| 1 | HP:0000131 | CHEBI:106700 |
| 1 | HP:0000131 | CHEBI:106704 |
| 1 | HP:0000131 | CHEBI:112146 |
| 1 | HP:0000131 | CHEBI:106731 |
| 1 | HP:0000131 | CHEBI:106737 |
| 1 | HP:0000131 | CHEBI:106724 |
| 1 | HP:0000131 | CHEBI:126270 |
| 1 | HP:0000131 | CHEBI:106782 |
| 1 | HP:0000131 | CHEBI:106786 |
| 1 | HP:0000131 | CHEBI:112128 |
| 1 | HP:0000131 | CHEBI:112091 |
| 1 | HP:0000131 | CHEBI:130858 |
| 1 | HP:0000131 | CHEBI:112034 |
| 1 | HP:0000131 | CHEBI:112476 |
| 1 | HP:0000131 | CHEBI:126975 |
| 1 | HP:0000131 | CHEBI:126943 |
| 1 | HP:0000131 | CHEBI:126952 |
| 1 | HP:0000131 | CHEBI:128121 |
| 1 | HP:0000131 | CHEBI:110885 |
| 1 | HP:0000131 | CHEBI:110891 |
| 1 | HP:0000131 | CHEBI:126391 |
| 1 | HP:0000131 | CHEBI:126388 |
| 1 | HP:0000131 | CHEBI:112462 |
| 1 | HP:0000131 | CHEBI:112080 |
| 1 | HP:0000131 | CHEBI:112496 |
| 1 | HP:0000131 | CHEBI:112482 |
| 1 | HP:0000131 | CHEBI:112459 |
| 1 | HP:0000131 | CHEBI:112072 |
| 1 | HP:0000131 | CHEBI:112062 |
| 1 | HP:0000131 | CHEBI:112090 |
| 1 | HP:0000131 | CHEBI:112086 |
| 1 | HP:0000131 | CHEBI:112083 |
| 1 | HP:0000131 | CHEBI:126152 |
| 1 | HP:0000131 | CHEBI:126155 |
| 1 | HP:0000131 | CHEBI:130104 |
| 1 | HP:0000131 | CHEBI:130116 |
| 1 | HP:0000131 | CHEBI:112456 |
| 1 | HP:0000131 | CHEBI:112490 |
| 1 | HP:0000131 | CHEBI:112463 |
| 1 | HP:0000131 | CHEBI:112469 |
| 1 | HP:0000131 | CHEBI:128339 |
| 1 | HP:0000131 | HP:0005305 |
| 1 | HP:0000131 | CHEBI:130136 |
| 1 | HP:0000131 | CHEBI:128038 |
| 1 | HP:0000131 | CHEBI:130124 |
| 1 | HP:0000131 | CHEBI:126120 |
| 1 | HP:0000131 | CHEBI:126976 |
| 1 | HP:0000131 | CHEBI:126115 |
| 1 | HP:0000131 | CHEBI:130701 |
| 1 | HP:0000131 | CHEBI:126937 |
| 1 | HP:0000131 | CHEBI:128052 |
| 1 | HP:0000131 | CHEBI:126166 |
| 1 | HP:0000131 | CHEBI:126933 |
| 1 | HP:0000131 | CHEBI:130471 |
| 1 | HP:0000131 | CHEBI:110575 |
| 1 | HP:0000131 | CHEBI:110566 |
| 1 | HP:0000131 | CHEBI:106485 |
| 1 | HP:0000131 | CHEBI:99957 |
| 1 | HP:0000131 | CHEBI:130082 |
| 1 | HP:0000131 | CHEBI:9123 |
| 1 | HP:0000131 | CHEBI:131276 |
| 1 | HP:0000131 | CHEBI:106476 |
| 1 | HP:0000131 | CHEBI:106463 |
| 1 | HP:0000131 | CHEBI:106467 |
| 1 | HP:0000131 | CHEBI:100179 |
| 1 | HP:0000131 | CHEBI:106434 |
| 1 | HP:0000131 | CHEBI:99882 |
| 1 | HP:0000131 | CHEBI:111981 |
| 1 | HP:0000131 | CHEBI:106425 |
| 1 | HP:0000131 | CHEBI:106450 |
| 1 | HP:0000131 | CHEBI:130005 |
| 1 | HP:0000131 | CHEBI:111971 |
| 1 | HP:0000131 | CHEBI:100180 |
| 1 | HP:0000131 | CHEBI:111996 |
| 1 | HP:0000131 | CHEBI:111992 |
| 1 | HP:0000131 | CHEBI:111916 |
| 1 | HP:0000131 | CHEBI:106403 |
| 1 | HP:0000131 | CHEBI:111922 |
| 1 | HP:0000131 | CHEBI:106418 |
| 1 | HP:0000131 | CHEBI:99161 |
| 1 | HP:0000131 | CHEBI:111022 |
| 1 | HP:0000131 | CHEBI:99664 |
| 1 | HP:0000131 | CHEBI:99185 |
| 1 | HP:0000131 | CHEBI:99633 |
| 1 | HP:0000131 | CHEBI:125318 |
| 1 | HP:0000131 | CHEBI:100136 |
| 1 | HP:0000131 | CHEBI:131243 |
| 1 | HP:0000131 | CHEBI:125307 |
| 1 | HP:0000131 | CHEBI:111942 |
| 1 | HP:0000131 | CHEBI:100143 |
| 1 | HP:0000131 | CHEBI:111963 |
| 1 | HP:0000131 | CHEBI:131260 |
| 1 | HP:0000131 | CHEBI:111957 |
| 1 | HP:0000131 | CHEBI:111908 |
| 1 | HP:0000131 | CHEBI:106410 |
| 1 | HP:0000131 | CHEBI:111902 |
| 1 | HP:0000131 | CHEBI:130029 |
| 1 | HP:0000131 | CHEBI:131206 |
| 1 | HP:0000131 | CHEBI:99139 |
| 1 | HP:0000131 | CHEBI:130718 |
| 1 | HP:0000131 | CHEBI:130759 |
| 1 | HP:0000131 | CHEBI:106550 |
| 1 | HP:0000131 | CHEBI:106587 |
| 1 | HP:0000131 | CHEBI:130899 |
| 1 | HP:0000131 | CHEBI:106596 |
| 1 | HP:0000131 | CHEBI:130709 |
| 1 | HP:0000131 | CHEBI:125303 |
| 1 | HP:0000131 | CHEBI:99794 |
| 1 | HP:0000131 | CHEBI:99842 |
| 1 | HP:0000131 | CHEBI:99733 |
| 1 | HP:0000131 | CHEBI:128200 |
| 1 | HP:0000131 | CHEBI:99849 |
| 1 | HP:0000131 | CHEBI:130812 |
| 1 | HP:0000131 | CHEBI:106529 |
| 1 | HP:0000131 | CHEBI:106524 |
| 1 | HP:0000131 | CHEBI:106536 |
| 1 | HP:0000131 | CHEBI:99716 |
| 1 | HP:0000131 | CHEBI:99747 |
| 1 | HP:0000131 | CHEBI:106580 |
| 1 | HP:0000131 | CHEBI:106577 |
| 1 | HP:0000131 | CHEBI:106561 |
| 1 | HP:0000131 | CHEBI:106567 |
| 1 | HP:0000131 | CHEBI:99736 |
| 1 | HP:0000131 | CHEBI:130077 |
| 1 | HP:0000131 | CHEBI:99742 |
| 1 | HP:0000131 | CHEBI:99705 |
| 1 | HP:0000131 | CHEBI:106513 |
| 1 | HP:0000131 | CHEBI:106516 |
| 1 | HP:0000131 | CHEBI:106508 |
| 1 | HP:0000131 | CHEBI:106533 |
| 1 | HP:0000131 | CHEBI:106537 |
| 1 | HP:0000131 | CHEBI:100215 |
| 1 | HP:0000131 | CHEBI:100202 |
| 1 | HP:0000131 | CHEBI:106552 |
| 1 | HP:0000131 | CHEBI:106555 |
| 1 | HP:0000131 | CHEBI:100208 |
| 1 | HP:0000131 | CHEBI:106571 |
| 1 | HP:0000131 | CHEBI:106563 |
| 1 | HP:0000131 | CHEBI:106569 |
| 1 | HP:0000131 | CHEBI:111018 |
| 1 | HP:0000131 | CHEBI:111048 |
| 1 | HP:0000131 | CHEBI:125298 |
| 1 | HP:0000131 | CHEBI:110573 |
| 1 | HP:0000131 | CHEBI:128857 |
| 1 | HP:0000131 | CHEBI:110532 |
| 1 | HP:0000131 | CHEBI:128845 |
| 1 | HP:0000131 | CHEBI:110588 |
| 1 | HP:0000131 | CHEBI:110593 |
| 1 | HP:0000131 | CHEBI:110598 |
| 1 | HP:0000131 | CHEBI:128870 |
| 1 | HP:0000131 | CHEBI:110567 |
| 1 | HP:0000131 | CHEBI:110578 |
| 1 | HP:0000131 | CHEBI:99578 |
| 1 | HP:0000131 | CHEBI:125881 |
| 1 | HP:0000131 | CHEBI:99376 |
| 1 | HP:0000131 | CHEBI:100360 |
| 1 | HP:0000131 | CHEBI:125867 |
| 1 | HP:0000131 | CHEBI:100385 |
| 1 | HP:0000131 | CHEBI:99398 |
| 1 | HP:0000131 | CHEBI:100037 |
| 1 | HP:0000131 | CHEBI:100038 |
| 1 | HP:0000131 | CHEBI:110556 |
| 1 | HP:0000131 | CHEBI:110545 |
| 1 | HP:0000131 | CHEBI:110540 |
| 1 | HP:0000131 | CHEBI:125815 |
| 1 | HP:0000131 | CHEBI:128977 |
| 1 | HP:0000131 | CHEBI:128974 |
| 1 | HP:0000131 | CHEBI:110663 |
| 1 | HP:0000131 | CHEBI:110668 |
| 1 | HP:0000131 | CHEBI:110659 |
| 1 | HP:0000131 | CHEBI:130586 |
| 1 | HP:0000131 | CHEBI:131032 |
| 1 | HP:0000131 | CHEBI:130611 |
| 1 | HP:0000131 | CHEBI:99474 |
| 1 | HP:0000131 | CHEBI:110695 |
| 1 | HP:0000131 | CHEBI:128908 |
| 1 | HP:0000131 | CHEBI:110620 |
| 1 | HP:0000131 | CHEBI:110630 |
| 1 | HP:0000131 | CHEBI:110607 |
| 1 | HP:0000131 | CHEBI:125842 |
| 1 | HP:0000131 | CHEBI:99461 |
| 1 | HP:0000131 | CHEBI:99467 |
| 1 | HP:0000131 | CHEBI:125953 |
| 1 | HP:0000131 | CHEBI:99588 |
| 1 | HP:0000131 | CHEBI:111030 |
| 1 | HP:0000131 | CHEBI:99028 |
| 1 | HP:0000131 | CHEBI:111070 |
| 1 | HP:0000131 | CHEBI:111078 |
| 1 | HP:0000131 | CHEBI:111898 |
| 1 | HP:0000131 | CHEBI:111088 |
| 1 | HP:0000131 | CHEBI:125699 |
| 1 | HP:0000131 | CHEBI:111091 |
| 1 | HP:0000131 | CHEBI:111055 |
| 1 | HP:0000131 | CHEBI:111058 |
| 1 | HP:0000131 | CHEBI:111060 |
| 1 | HP:0000131 | CHEBI:99584 |
| 1 | HP:0000131 | CHEBI:111064 |
| 1 | HP:0000131 | CHEBI:100053 |
| 1 | HP:0000131 | CHEBI:100049 |
| 1 | HP:0000131 | CHEBI:111863 |
| 1 | HP:0000131 | CHEBI:99041 |
| 1 | HP:0000131 | CHEBI:111095 |
| 1 | HP:0000131 | CHEBI:111852 |
| 1 | HP:0000131 | CHEBI:111000 |
| 1 | HP:0000131 | CHEBI:130495 |
| 1 | HP:0000131 | CHEBI:111833 |
| 1 | HP:0000131 | CHEBI:53796 |
| 1 | HP:0000131 | CHEBI:111841 |
| 1 | HP:0000131 | CHEBI:99547 |
| 1 | HP:0000131 | CHEBI:111116 |
| 1 | HP:0000131 | CHEBI:111125 |
| 1 | HP:0000131 | CHEBI:111107 |
| 1 | HP:0000131 | CHEBI:111108 |
| 1 | HP:0000131 | CHEBI:111157 |
| 1 | HP:0000131 | CHEBI:125995 |
| 1 | HP:0000131 | CHEBI:111131 |
| 1 | HP:0000131 | CHEBI:99242 |
| 1 | HP:0000131 | CHEBI:111137 |
| 1 | HP:0000131 | CHEBI:111141 |
| 1 | HP:0000131 | CHEBI:125989 |
| 1 | HP:0000131 | CHEBI:99253 |
| 1 | HP:0000131 | CHEBI:130455 |
| 1 | HP:0000131 | CHEBI:99085 |
| 1 | HP:0000131 | CHEBI:111849 |
| 1 | HP:0000131 | CHEBI:111845 |
| 1 | HP:0000131 | CHEBI:106531 |
| 1 | HP:0000131 | CHEBI:130247 |
| 1 | HP:0000131 | CHEBI:126972 |
| 1 | HP:0000131 | CHEBI:110677 |
| 1 | HP:0000131 | CHEBI:126340 |
| 1 | HP:0000131 | CHEBI:99326 |
| 1 | HP:0000131 | CHEBI:126320 |
| 1 | HP:0000131 | CHEBI:130391 |
| 1 | HP:0000131 | CHEBI:130388 |
| 1 | HP:0000131 | CHEBI:128713 |
| 1 | HP:0000131 | CHEBI:130648 |
| 1 | HP:0000131 | CHEBI:110673 |
| 1 | HP:0000131 | CHEBI:110643 |
| 1 | HP:0000131 | CHEBI:126410 |
| 1 | HP:0000131 | CHEBI:110648 |
| 1 | HP:0000131 | CHEBI:131116 |
| 1 | HP:0000131 | CHEBI:110657 |
| 1 | HP:0000131 | CHEBI:126398 |
| 1 | HP:0000131 | CHEBI:99378 |
| 1 | HP:0000131 | CHEBI:130337 |
| 1 | HP:0000131 | CHEBI:125838 |
| 1 | HP:0000131 | CHEBI:131134 |
| 1 | HP:0000131 | CHEBI:99332 |
| 1 | HP:0000131 | CHEBI:130636 |
| 1 | HP:0000131 | CHEBI:105482 |
| 1 | HP:0000131 | CHEBI:130418 |
| 1 | HP:0000131 | CHEBI:110791 |
| 1 | HP:0000131 | CHEBI:110793 |
| 1 | HP:0000131 | CHEBI:110762 |
| 1 | HP:0000131 | CHEBI:110763 |
| 1 | HP:0000131 | CHEBI:99296 |
| 1 | HP:0000131 | CHEBI:130949 |
| 1 | HP:0000131 | CHEBI:128177 |
| 1 | HP:0000131 | CHEBI:110700 |
| 1 | HP:0000131 | CHEBI:110709 |
| 1 | HP:0000131 | CHEBI:110715 |
| 1 | HP:0000131 | CHEBI:110718 |
| 1 | HP:0000131 | CHEBI:130582 |
| 1 | HP:0000131 | CHEBI:126297 |
| 1 | HP:0000131 | CHEBI:110759 |
| 1 | HP:0000131 | CHEBI:130545 |
| 1 | HP:0000131 | CHEBI:130538 |
| 1 | HP:0000131 | CHEBI:128673 |
| 1 | HP:0000131 | CHEBI:130349 |
| 1 | HP:0000131 | CHEBI:110685 |
| 1 | HP:0000131 | CHEBI:110781 |
| 1 | HP:0000131 | CHEBI:131070 |
| 1 | HP:0000131 | CHEBI:99466 |
| 1 | HP:0000131 | CHEBI:131059 |
| 1 | HP:0000131 | CHEBI:130642 |
| 1 | HP:0000131 | CHEBI:110546 |
| 1 | HP:0000131 | CHEBI:110543 |
| 1 | HP:0000131 | CHEBI:110548 |
| 1 | HP:0000131 | CHEBI:110554 |
| 1 | HP:0000131 | CHEBI:110559 |
| 1 | HP:0000131 | CHEBI:130694 |
| 1 | HP:0000131 | CHEBI:110944 |
| 1 | HP:0000131 | CHEBI:110531 |
| 1 | HP:0000131 | CHEBI:130682 |
| 1 | HP:0000131 | CHEBI:126988 |
| 1 | HP:0000131 | CHEBI:110582 |
| 1 | HP:0000131 | CHEBI:110585 |
| 1 | HP:0000131 | CHEBI:110597 |
| 1 | HP:0000131 | CHEBI:126965 |
| 1 | HP:0000131 | CHEBI:110560 |
| 1 | HP:0000131 | CHEBI:128436 |
| 1 | HP:0000131 | CHEBI:126923 |
| 1 | HP:0000131 | CHEBI:128812 |
| 1 | HP:0000131 | CHEBI:130304 |
| 1 | HP:0000131 | CHEBI:110697 |
| 1 | HP:0000131 | CHEBI:110928 |
| 1 | HP:0000131 | CHEBI:110929 |
| 1 | HP:0000131 | CHEBI:110939 |
| 1 | HP:0000131 | CHEBI:110622 |
| 1 | HP:0000131 | CHEBI:110981 |
| 1 | HP:0000131 | CHEBI:110987 |
| 1 | HP:0000131 | CHEBI:110991 |
| 1 | HP:0000131 | CHEBI:110964 |
| 1 | HP:0000131 | CHEBI:125895 |
| 1 | HP:0000131 | CHEBI:110966 |
| 1 | HP:0000131 | CHEBI:110632 |
| 1 | HP:0000131 | CHEBI:110975 |
| 1 | HP:0000131 | CHEBI:110609 |
| 1 | HP:0000131 | CHEBI:110611 |
| 1 | HP:0000131 | CHEBI:130356 |
| 1 | HP:0000131 | CHEBI:128830 |
| 1 | HP:0000131 | CHEBI:110785 |
| 1 | HP:0000131 | CHEBI:110780 |
| 1 | HP:0000131 | CHEBI:106506 |
| 1 | HP:0000131 | CHEBI:125722 |
| 1 | HP:0000131 | CHEBI:106459 |
| 1 | HP:0000131 | CHEBI:106428 |
| 1 | HP:0000131 | CHEBI:106424 |
| 1 | HP:0000131 | CHEBI:106438 |
| 1 | HP:0000131 | CHEBI:106483 |
| 1 | HP:0000131 | CHEBI:106497 |
| 1 | HP:0000131 | CHEBI:106466 |
| 1 | HP:0000131 | CHEBI:106462 |
| 1 | HP:0000131 | CHEBI:128364 |
| 1 | HP:0000131 | CHEBI:130843 |
| 1 | HP:0000131 | CHEBI:128377 |
| 1 | HP:0000131 | CHEBI:99976 |
| 1 | HP:0000131 | CHEBI:130190 |
| 1 | HP:0000131 | CHEBI:130182 |
| 1 | HP:0000131 | CHEBI:111134 |
| 1 | HP:0000131 | CHEBI:99601 |
| 1 | HP:0000131 | CHEBI:106407 |
| 1 | HP:0000131 | CHEBI:106402 |
| 1 | HP:0000131 | CHEBI:131215 |
| 1 | HP:0000131 | CHEBI:106443 |
| 1 | HP:0000131 | CHEBI:106448 |
| 1 | HP:0000131 | CHEBI:111084 |
| 1 | HP:0000131 | CHEBI:111025 |
| 1 | HP:0000131 | CHEBI:106502 |
| 1 | HP:0000131 | CHEBI:130825 |
| 1 | HP:0000131 | CHEBI:111014 |
| 1 | HP:0000131 | CHEBI:106512 |
| 1 | HP:0000131 | CHEBI:111049 |
| 1 | HP:0000131 | CHEBI:111044 |
| 1 | HP:0000131 | CHEBI:130288 |
| 1 | HP:0000131 | CHEBI:130284 |
| 1 | HP:0000131 | CHEBI:111034 |
| 1 | HP:0000131 | CHEBI:111001 |
| 1 | HP:0000131 | CHEBI:130876 |
| 1 | HP:0000131 | CHEBI:131277 |
| 1 | HP:0000131 | CHEBI:130212 |
| 1 | HP:0000131 | CHEBI:111069 |
| 1 | HP:0000131 | CHEBI:111062 |
| 1 | HP:0000131 | CHEBI:111086 |
| 1 | HP:0000131 | CHEBI:106417 |
| 1 | HP:0000131 | CHEBI:131318 |
| 1 | HP:0000131 | CHEBI:128102 |
| 1 | HP:0000131 | CHEBI:110879 |
| 1 | HP:0000131 | CHEBI:110822 |
| 1 | HP:0000131 | CHEBI:110830 |
| 1 | HP:0000131 | CHEBI:110800 |
| 1 | HP:0000131 | CHEBI:110804 |
| 1 | HP:0000131 | CHEBI:110818 |
| 1 | HP:0000131 | CHEBI:110861 |
| 1 | HP:0000131 | CHEBI:110869 |
| 1 | HP:0000131 | CHEBI:110872 |
| 1 | HP:0000131 | CHEBI:110842 |
| 1 | HP:0000131 | HP:0100817 |
| 1 | HP:0000131 | CHEBI:110854 |
| 1 | HP:0000131 | CHEBI:112014 |
| 1 | HP:0000131 | CHEBI:112012 |
| 1 | HP:0000131 | CHEBI:99218 |
| 1 | HP:0000131 | CHEBI:112001 |
| 1 | HP:0000131 | CHEBI:128154 |
| 1 | HP:0000131 | CHEBI:128129 |
| 1 | HP:0000131 | CHEBI:128132 |
| 1 | HP:0000131 | CHEBI:9435 |
| 1 | HP:0000131 | CHEBI:126560 |
| 1 | HP:0000131 | CHEBI:128051 |
| 1 | HP:0000131 | CHEBI:110895 |
| 1 | HP:0000131 | CHEBI:111103 |
| 1 | HP:0000131 | CHEBI:125737 |
| 1 | HP:0000131 | CHEBI:111129 |
| 1 | HP:0000131 | CHEBI:99911 |
| 1 | HP:0000131 | CHEBI:125744 |
| 1 | HP:0000131 | CHEBI:111112 |
| 1 | HP:0000131 | CHEBI:99928 |
| 1 | HP:0000131 | CHEBI:99105 |
| 1 | HP:0000131 | CHEBI:126473 |
| 1 | HP:0000131 | CHEBI:99598 |
| 1 | HP:0000131 | CHEBI:128022 |
| 1 | HP:0000131 | CHEBI:128006 |
| 1 | HP:0000131 | CHEBI:99153 |
| 1 | HP:0000131 | CHEBI:130892 |
| 1 | HP:0000131 | CHEBI:99138 |
| 1 | HP:0000131 | CHEBI:128091 |
| 1 | HP:0000131 | CHEBI:99533 |
| 1 | HP:0000131 | CHEBI:126569 |
| 1 | HP:0000131 | CHEBI:110635 |
| 1 | HP:0000131 | CHEBI:110639 |
| 1 | HP:0000131 | CHEBI:111875 |
| 1 | HP:0000131 | CHEBI:106522 |
| 1 | HP:0000131 | CHEBI:99030 |
| 1 | HP:0000131 | CHEBI:106509 |
| 1 | HP:0000131 | CHEBI:106504 |
| 1 | HP:0000131 | CHEBI:100127 |
| 1 | HP:0000131 | CHEBI:100121 |
| 1 | HP:0000131 | CHEBI:100445 |
| 1 | HP:0000131 | CHEBI:111879 |
| 1 | HP:0000131 | CHEBI:111870 |
| 1 | HP:0000131 | CHEBI:106449 |
| 1 | HP:0000131 | CHEBI:99054 |
| 1 | HP:0000131 | CHEBI:127737 |
| 1 | HP:0000131 | CHEBI:110660 |
| 1 | HP:0000131 | CHEBI:110669 |
| 1 | HP:0000131 | CHEBI:110672 |
| 1 | HP:0000131 | CHEBI:110645 |
| 1 | HP:0000131 | CHEBI:110651 |
| 1 | HP:0000131 | CHEBI:126186 |
| 1 | HP:0000131 | CHEBI:106526 |
| 1 | HP:0000131 | CHEBI:111865 |
| 1 | HP:0000131 | CHEBI:110661 |
| 1 | HP:0000131 | CHEBI:110666 |
| 1 | HP:0000131 | CHEBI:110683 |
| 1 | HP:0000131 | CHEBI:99768 |
| 1 | HP:0000131 | CHEBI:106564 |
| 1 | HP:0000131 | CHEBI:110653 |
| 1 | HP:0000131 | CHEBI:106579 |
| 1 | HP:0000131 | CHEBI:106572 |
| 1 | HP:0000131 | CHEBI:110652 |
| 1 | HP:0000131 | CHEBI:111890 |
| 1 | HP:0000131 | CHEBI:112619 |
| 1 | HP:0000131 | CHEBI:106540 |
| 1 | HP:0000131 | CHEBI:99027 |
| 1 | HP:0000131 | CHEBI:106589 |
| 1 | HP:0000131 | CHEBI:106584 |
| 1 | HP:0000131 | CHEBI:106590 |
| 1 | HP:0000131 | CHEBI:112600 |
| 1 | HP:0000131 | CHEBI:106593 |
| 1 | HP:0000131 | CHEBI:110674 |
| 1 | HP:0000131 | CHEBI:110655 |
| 1 | HP:0000131 | CHEBI:106445 |
| 1 | HP:0000131 | CHEBI:111838 |
| 1 | HP:0000131 | CHEBI:111982 |
| 1 | HP:0000131 | CHEBI:106411 |
| 1 | HP:0000131 | CHEBI:111921 |
| 1 | HP:0000131 | CHEBI:111913 |
| 1 | HP:0000131 | CHEBI:110600 |
| 1 | HP:0000131 | CHEBI:110604 |
| 1 | HP:0000131 | CHEBI:110610 |
| 1 | HP:0000131 | CHEBI:111985 |
| 1 | HP:0000131 | CHEBI:111980 |
| 1 | HP:0000131 | CHEBI:111977 |
| 1 | HP:0000131 | CHEBI:106456 |
| 1 | HP:0000131 | CHEBI:111974 |
| 1 | HP:0000131 | CHEBI:111972 |
| 1 | HP:0000131 | CHEBI:111995 |
| 1 | HP:0000131 | CHEBI:111991 |
| 1 | HP:0000131 | CHEBI:110542 |
| 1 | HP:0000131 | CHEBI:100250 |
| 1 | HP:0000131 | CHEBI:126007 |
| 1 | HP:0000131 | CHEBI:110553 |
| 1 | HP:0000131 | CHEBI:111927 |
| 1 | HP:0000131 | CHEBI:111907 |
| 1 | HP:0000131 | CHEBI:106404 |
| 1 | HP:0000131 | CHEBI:106409 |
| 1 | HP:0000131 | CHEBI:106451 |
| 1 | HP:0000131 | CHEBI:106429 |
| 1 | HP:0000131 | CHEBI:106421 |
| 1 | HP:0000131 | CHEBI:106439 |
| 1 | HP:0000131 | CHEBI:106435 |
| 1 | HP:0000131 | CHEBI:100063 |
| 1 | HP:0000131 | CHEBI:106482 |
| 1 | HP:0000131 | CHEBI:110682 |
| 1 | HP:0000131 | CHEBI:110687 |
| 1 | HP:0000131 | CHEBI:100174 |
| 1 | HP:0000131 | CHEBI:110691 |
| 1 | HP:0000131 | CHEBI:111931 |
| 1 | HP:0000131 | CHEBI:111968 |
| 1 | HP:0000131 | CHEBI:100168 |
| 1 | HP:0000131 | CHEBI:126124 |
| 1 | HP:0000131 | CHEBI:111954 |
| 1 | HP:0000131 | CHEBI:99967 |
| 1 | HP:0000131 | CHEBI:111837 |
| 1 | HP:0000131 | CHEBI:112637 |
| 1 | HP:0000131 | CHEBI:100201 |
| 1 | HP:0000131 | CHEBI:125966 |
| 1 | HP:0000131 | CHEBI:110586 |
| 1 | HP:0000131 | CHEBI:110539 |
| 1 | HP:0000131 | CHEBI:131221 |
| 1 | HP:0000131 | CHEBI:110527 |
| 1 | HP:0000131 | CHEBI:110945 |
| 1 | HP:0000131 | CHEBI:112042 |
| 1 | HP:0000131 | CHEBI:110952 |
| 1 | HP:0000131 | CHEBI:110954 |
| 1 | HP:0000131 | CHEBI:125962 |
| 1 | HP:0000131 | CHEBI:110992 |
| 1 | HP:0000131 | CHEBI:131247 |
| 1 | HP:0000131 | CHEBI:131244 |
| 1 | HP:0000131 | CHEBI:110921 |
| 1 | HP:0000131 | CHEBI:130008 |
| 1 | HP:0000131 | HP:0003876 |
| 1 | HP:0000131 | CHEBI:110932 |
| 1 | HP:0000131 | CHEBI:110982 |
| 1 | HP:0000131 | CHEBI:110988 |
| 1 | HP:0000131 | CHEBI:99630 |
| 1 | HP:0000131 | CHEBI:126768 |
| 1 | HP:0000131 | CHEBI:126765 |
| 1 | HP:0000131 | CHEBI:100292 |
| 1 | HP:0000131 | CHEBI:113332 |
| 1 | HP:0000131 | CHEBI:113300 |
| 1 | HP:0000131 | CHEBI:112142 |
| 1 | HP:0000131 | CHEBI:112151 |
| 1 | HP:0000131 | CHEBI:127570 |
| 1 | HP:0000131 | CHEBI:29516 |
| 1 | HP:0000131 | CHEBI:99621 |
| 1 | HP:0000131 | CHEBI:127564 |
| 1 | HP:0000131 | CHEBI:127523 |
| 1 | HP:0000131 | CHEBI:112125 |
| 1 | HP:0000131 | CHEBI:126758 |
| 1 | HP:0000131 | CHEBI:112135 |
| 1 | HP:0000131 | CHEBI:112107 |
| 1 | HP:0000131 | CHEBI:110577 |
| 1 | HP:0000131 | CHEBI:112119 |
| 1 | HP:0000131 | CHEBI:110561 |
| 1 | HP:0000131 | CHEBI:110564 |
| 1 | HP:0000131 | CHEBI:130015 |
| 1 | HP:0000131 | CHEBI:110998 |
| 1 | HP:0000131 | CHEBI:110694 |
| 1 | HP:0000131 | CHEBI:112571 |
| 1 | HP:0000131 | CHEBI:112487 |
| 1 | HP:0000131 | CHEBI:99744 |
| 1 | HP:0000131 | CHEBI:112493 |
| 1 | HP:0000131 | CHEBI:112473 |
| 1 | HP:0000131 | CHEBI:100057 |
| 1 | HP:0000131 | CHEBI:125988 |
| 1 | HP:0000131 | CHEBI:112545 |
| 1 | HP:0000131 | CHEBI:112579 |
| 1 | HP:0000131 | CHEBI:112568 |
| 1 | HP:0000131 | CHEBI:112055 |
| 1 | HP:0000131 | CHEBI:112565 |
| 1 | HP:0000131 | CHEBI:127686 |
| 1 | HP:0000131 | CHEBI:110601 |
| 1 | HP:0000131 | CHEBI:110637 |
| 1 | HP:0000131 | CHEBI:110633 |
| 1 | HP:0000131 | CHEBI:99799 |
| 1 | HP:0000131 | CHEBI:110628 |
| 1 | HP:0000131 | CHEBI:100075 |
| 1 | HP:0000131 | CHEBI:112481 |
| 1 | HP:0000131 | CHEBI:112599 |
| 1 | HP:0000131 | CHEBI:112074 |
| 1 | HP:0000131 | CHEBI:112064 |
| 1 | HP:0000131 | CHEBI:110969 |
| 1 | HP:0000131 | CHEBI:100454 |
| 1 | HP:0000131 | CHEBI:112020 |
| 1 | HP:0000131 | CHEBI:130081 |
| 1 | HP:0000131 | CHEBI:110903 |
| 1 | HP:0000131 | CHEBI:110907 |
| 1 | HP:0000131 | CHEBI:9124 |
| 1 | HP:0000131 | CHEBI:110910 |
| 1 | HP:0000131 | CHEBI:131278 |
| 1 | HP:0000131 | CHEBI:112039 |
| 1 | HP:0000131 | CHEBI:112512 |
| 1 | HP:0000131 | CHEBI:112509 |
| 1 | HP:0000131 | CHEBI:112539 |
| 1 | HP:0000131 | CHEBI:112096 |
| 1 | HP:0000131 | CHEBI:112533 |
| 1 | HP:0000131 | CHEBI:112098 |
| 1 | HP:0000131 | CHEBI:112060 |
| 1 | HP:0000131 | CHEBI:107005 |
| 1 | HP:0000131 | CHEBI:100207 |
| 1 | HP:0000131 | CHEBI:113338 |
| 1 | HP:0000131 | CHEBI:106845 |
| 1 | HP:0000131 | CHEBI:113473 |
| 1 | HP:0000131 | CHEBI:113491 |
| 1 | HP:0000131 | CHEBI:128093 |
| 1 | HP:0000131 | CHEBI:128032 |
| 1 | HP:0000131 | CHEBI:128017 |
| 1 | HP:0000131 | CHEBI:128069 |
| 1 | HP:0000131 | CHEBI:128059 |
| 1 | HP:0000131 | CHEBI:127860 |
| 1 | HP:0000131 | CHEBI:106842 |
| 1 | HP:0000131 | CHEBI:127814 |
| 1 | HP:0000131 | CHEBI:125299 |
| 1 | HP:0000131 | CHEBI:131047 |
| 1 | HP:0000131 | CHEBI:106855 |
| 1 | HP:0000131 | CHEBI:106828 |
| 1 | HP:0000131 | CHEBI:106822 |
| 1 | HP:0000131 | CHEBI:126563 |
| 1 | HP:0000131 | CHEBI:127845 |
| 1 | HP:0000131 | CHEBI:126571 |
| 1 | HP:0000131 | CHEBI:106916 |
| 1 | HP:0000131 | CHEBI:126583 |
| 1 | HP:0000131 | CHEBI:113482 |
| 1 | HP:0000131 | CHEBI:131098 |
| 1 | HP:0000131 | CHEBI:126488 |
| 1 | HP:0000131 | CHEBI:126463 |
| 1 | HP:0000131 | CHEBI:127762 |
| 1 | HP:0000131 | CHEBI:106995 |
| 1 | HP:0000131 | CHEBI:106991 |
| 1 | HP:0000131 | CHEBI:113446 |
| 1 | HP:0000131 | CHEBI:113436 |
| 1 | HP:0000131 | CHEBI:113430 |
| 1 | HP:0000131 | CHEBI:113456 |
| 1 | HP:0000131 | CHEBI:113404 |
| 1 | HP:0000131 | CHEBI:113417 |
| 1 | HP:0000131 | CHEBI:113412 |
| 1 | HP:0000131 | CHEBI:131052 |
| 1 | HP:0000131 | CHEBI:128013 |
| 1 | HP:0000131 | CHEBI:106922 |
| 1 | HP:0000131 | CHEBI:106937 |
| 1 | HP:0000131 | CHEBI:126565 |
| 1 | HP:0000131 | CHEBI:128094 |
| 1 | HP:0000131 | CHEBI:113509 |
| 1 | HP:0000131 | CHEBI:112017 |
| 1 | HP:0000131 | CHEBI:112132 |
| 1 | HP:0000131 | CHEBI:113317 |
| 1 | HP:0000131 | CHEBI:128679 |
| 1 | HP:0000131 | CHEBI:128122 |
| 1 | HP:0000131 | CHEBI:113340 |
| 1 | HP:0000131 | CHEBI:126702 |
| 1 | HP:0000131 | CHEBI:113339 |
| 1 | HP:0000131 | CHEBI:126724 |
| 1 | HP:0000131 | CHEBI:112129 |
| 1 | HP:0000131 | CHEBI:112138 |
| 1 | HP:0000131 | CHEBI:127827 |
| 1 | HP:0000131 | CHEBI:112104 |
| 1 | HP:0000131 | CHEBI:126778 |
| 1 | HP:0000131 | CHEBI:112110 |
| 1 | HP:0000131 | CHEBI:112116 |
| 1 | HP:0000131 | CHEBI:128681 |
| 1 | HP:0000131 | CHEBI:130059 |
| 1 | HP:0000131 | CHEBI:130056 |
| 1 | HP:0000131 | CHEBI:112041 |
| 1 | HP:0000131 | CHEBI:127983 |
| 1 | HP:0000131 | CHEBI:113321 |
| 1 | HP:0000131 | CHEBI:112147 |
| 1 | HP:0000131 | CHEBI:112144 |
| 1 | HP:0000131 | CHEBI:127829 |
| 1 | HP:0000131 | CHEBI:106897 |
| 1 | HP:0000131 | CHEBI:106868 |
| 1 | HP:0000131 | CHEBI:113290 |
| 1 | HP:0000131 | CHEBI:113289 |
| 1 | HP:0000131 | CHEBI:113283 |
| 1 | HP:0000131 | CHEBI:106871 |
| 1 | HP:0000131 | CHEBI:113271 |
| 1 | HP:0000131 | CHEBI:125304 |
| 1 | HP:0000131 | CHEBI:113299 |
| 1 | HP:0000131 | CHEBI:128135 |
| 1 | HP:0000131 | CHEBI:113292 |
| 1 | HP:0000131 | CHEBI:112016 |
| 1 | HP:0000131 | CHEBI:126472 |
| 1 | HP:0000131 | CHEBI:113526 |
| 1 | HP:0000131 | CHEBI:113514 |
| 1 | HP:0000131 | CHEBI:126607 |
| 1 | HP:0000131 | CHEBI:127788 |
| 1 | HP:0000131 | CHEBI:112013 |
| 1 | HP:0000131 | CHEBI:100238 |
| 1 | HP:0000131 | CHEBI:110605 |
| 1 | HP:0000131 | CHEBI:106797 |
| 1 | HP:0000131 | CHEBI:106793 |
| 1 | HP:0000131 | CHEBI:100264 |
| 1 | HP:0000131 | CHEBI:106733 |
| 1 | HP:0000131 | CHEBI:106717 |
| 1 | HP:0000131 | CHEBI:106711 |
| 1 | HP:0000131 | CHEBI:126307 |
| 1 | HP:0000131 | CHEBI:126333 |
| 1 | HP:0000131 | CHEBI:106770 |
| 1 | HP:0000131 | CHEBI:126368 |
| 1 | HP:0000131 | CHEBI:106740 |
| 1 | HP:0000131 | CHEBI:110634 |
| 1 | HP:0000131 | CHEBI:110629 |
| 1 | HP:0000131 | CHEBI:110625 |
| 1 | HP:0000131 | CHEBI:32068 |
| 1 | HP:0000131 | CHEBI:9584 |
| 1 | HP:0000131 | CHEBI:128215 |
| 1 | HP:0000131 | CHEBI:110689 |
| 1 | HP:0000131 | CHEBI:106790 |
| 1 | HP:0000131 | CHEBI:106784 |
| 1 | HP:0000131 | CHEBI:106789 |
| 1 | HP:0000131 | CHEBI:110576 |
| 1 | HP:0000131 | CHEBI:106800 |
| 1 | HP:0000131 | CHEBI:110572 |
| 1 | HP:0000131 | CHEBI:107025 |
| 1 | HP:0000131 | CHEBI:106812 |
| 1 | HP:0000131 | CHEBI:110584 |
| 1 | HP:0000131 | CHEBI:107012 |
| 1 | HP:0000131 | CHEBI:110594 |
| 1 | HP:0000131 | CHEBI:110590 |
| 1 | HP:0000131 | CHEBI:110587 |
| 1 | HP:0000131 | CHEBI:110580 |
| 1 | HP:0000131 | CHEBI:110536 |
| 1 | HP:0000131 | CHEBI:110596 |
| 1 | HP:0000131 | CHEBI:110535 |
| 1 | HP:0000131 | CHEBI:99195 |
| 1 | HP:0000131 | CHEBI:110568 |
| 1 | HP:0000131 | CHEBI:110565 |
| 1 | HP:0000131 | CHEBI:110579 |
| 1 | HP:0000131 | CHEBI:99235 |
| 1 | HP:0000131 | CHEBI:110658 |
| 1 | HP:0000131 | CHEBI:112011 |
| 1 | HP:0000131 | CHEBI:106967 |
| 1 | HP:0000131 | CHEBI:113365 |
| 1 | HP:0000131 | CHEBI:113355 |
| 1 | HP:0000131 | CHEBI:113353 |
| 1 | HP:0000131 | CHEBI:113389 |
| 1 | HP:0000131 | CHEBI:113381 |
| 1 | HP:0000131 | CHEBI:113377 |
| 1 | HP:0000131 | CHEBI:94898 |
| 1 | HP:0000131 | CHEBI:128341 |
| 1 | HP:0000131 | CHEBI:106964 |
| 1 | HP:0000131 | CHEBI:110654 |
| 1 | HP:0000131 | CHEBI:106960 |
| 1 | HP:0000131 | CHEBI:131136 |
| 1 | HP:0000131 | CHEBI:128350 |
| 1 | HP:0000131 | CHEBI:128355 |
| 1 | HP:0000131 | CHEBI:106959 |
| 1 | HP:0000131 | CHEBI:112000 |
| 1 | HP:0000131 | CHEBI:112006 |
| 1 | HP:0000131 | CHEBI:127773 |
| 1 | HP:0000131 | CHEBI:113398 |
| 1 | HP:0000131 | CHEBI:106634 |
| 1 | HP:0000131 | CHEBI:128303 |
| 1 | HP:0000131 | CHEBI:106626 |
| 1 | HP:0000131 | CHEBI:110649 |
| 1 | HP:0000131 | CHEBI:110644 |
| 1 | HP:0000131 | CHEBI:110679 |
| 1 | HP:0000131 | CHEBI:110678 |
| 1 | HP:0000131 | CHEBI:110667 |
| 1 | HP:0000131 | CHEBI:106682 |
| 1 | HP:0000131 | CHEBI:106696 |
| 1 | HP:0000131 | CHEBI:128285 |
| 1 | HP:0000131 | CHEBI:106660 |
| 1 | HP:0000131 | CHEBI:106672 |
| 1 | HP:0000131 | CHEBI:106601 |
| 1 | HP:0000131 | CHEBI:99337 |
| 1 | HP:0000131 | CHEBI:106610 |
| 1 | HP:0000131 | CHEBI:106646 |
| 1 | HP:0000131 | CHEBI:99313 |
| 1 | HP:0000131 | CHEBI:99317 |
| 1 | HP:0000131 | CHEBI:106653 |
| 1 | HP:0000131 | CHEBI:113334 |
| 1 | HP:0000131 | CHEBI:127962 |
| 1 | HP:0000131 | CHEBI:110603 |
| 1 | HP:0000131 | CHEBI:112615 |
| 1 | HP:0000131 | CHEBI:112656 |
| 1 | HP:0000131 | CHEBI:112651 |
| 1 | HP:0000131 | CHEBI:126139 |
| 1 | HP:0000131 | CHEBI:99878 |
| 1 | HP:0000131 | CHEBI:92112 |
| 1 | HP:0000131 | CHEBI:111990 |
| 1 | HP:0000131 | CHEBI:111998 |
| 1 | HP:0000131 | CHEBI:112641 |
| 1 | HP:0000131 | CHEBI:111973 |
| 1 | HP:0000131 | CHEBI:111911 |
| 1 | HP:0000131 | CHEBI:112605 |
| 1 | HP:0000131 | CHEBI:112602 |
| 1 | HP:0000131 | CHEBI:111984 |
| 1 | HP:0000131 | CHEBI:126190 |
| 1 | HP:0000131 | CHEBI:106518 |
| 1 | HP:0000131 | CHEBI:106500 |
| 1 | HP:0000131 | CHEBI:106539 |
| 1 | HP:0000131 | CHEBI:106523 |
| 1 | HP:0000131 | CHEBI:106447 |
| 1 | HP:0000131 | CHEBI:110880 |
| 1 | HP:0000131 | CHEBI:106453 |
| 1 | HP:0000131 | CHEBI:99906 |
| 1 | HP:0000131 | CHEBI:99944 |
| 1 | HP:0000131 | CHEBI:112553 |
| 1 | HP:0000131 | CHEBI:106415 |
| 1 | HP:0000131 | CHEBI:112547 |
| 1 | HP:0000131 | CHEBI:112543 |
| 1 | HP:0000131 | CHEBI:130515 |
| 1 | HP:0000131 | CHEBI:106405 |
| 1 | HP:0000131 | CHEBI:112578 |
| 1 | HP:0000131 | CHEBI:112574 |
| 1 | HP:0000131 | CHEBI:112570 |
| 1 | HP:0000131 | CHEBI:112567 |
| 1 | HP:0000131 | CHEBI:106470 |
| 1 | HP:0000131 | CHEBI:106494 |
| 1 | HP:0000131 | CHEBI:106491 |
| 1 | HP:0000131 | CHEBI:106431 |
| 1 | HP:0000131 | CHEBI:110888 |
| 1 | HP:0000131 | CHEBI:106426 |
| 1 | HP:0000131 | HP:0001051 |
| 1 | HP:0000131 | CHEBI:111925 |
| 1 | HP:0000131 | CHEBI:110828 |
| 1 | HP:0000131 | CHEBI:99718 |
| 1 | HP:0000131 | CHEBI:111887 |
| 1 | HP:0000131 | CHEBI:130158 |
| 1 | HP:0000131 | CHEBI:111857 |
| 1 | HP:0000131 | CHEBI:130130 |
| 1 | HP:0000131 | CHEBI:15365 |
| 1 | HP:0000131 | CHEBI:126112 |
| 1 | HP:0000131 | CHEBI:111115 |
| 1 | HP:0000131 | CHEBI:111124 |
| 1 | HP:0000131 | CHEBI:111106 |
| 1 | HP:0000131 | CHEBI:111905 |
| 1 | HP:0000131 | CHEBI:111156 |
| 1 | HP:0000131 | CHEBI:99009 |
| 1 | HP:0000131 | CHEBI:111160 |
| 1 | HP:0000131 | CHEBI:111130 |
| 1 | HP:0000131 | CHEBI:111140 |
| 1 | HP:0000131 | CHEBI:130236 |
| 1 | HP:0000131 | CHEBI:111842 |
| 1 | HP:0000131 | CHEBI:110976 |
| 1 | HP:0000131 | CHEBI:111880 |
| 1 | HP:0000131 | CHEBI:111874 |
| 1 | HP:0000131 | CHEBI:111877 |
| 1 | HP:0000131 | CHEBI:99762 |
| 1 | HP:0000131 | CHEBI:111952 |
| 1 | HP:0000131 | CHEBI:111959 |
| 1 | HP:0000131 | CHEBI:111960 |
| 1 | HP:0000131 | CHEBI:111966 |
| 1 | HP:0000131 | CHEBI:111930 |
| 1 | HP:0000131 | CHEBI:130114 |
| 1 | HP:0000131 | CHEBI:111939 |
| 1 | HP:0000131 | CHEBI:111940 |
| 1 | HP:0000131 | CHEBI:111945 |
| 1 | HP:0000131 | CHEBI:111948 |
| 1 | HP:0000131 | CHEBI:106598 |
| 1 | HP:0000131 | CHEBI:106592 |
| 1 | HP:0000131 | CHEBI:106586 |
| 1 | HP:0000131 | CHEBI:106542 |
| 1 | HP:0000131 | CHEBI:106544 |
| 1 | HP:0000131 | CHEBI:106574 |
| 1 | HP:0000131 | CHEBI:106570 |
| 1 | HP:0000131 | CHEBI:110821 |
| 1 | HP:0000131 | CHEBI:112556 |
| 1 | HP:0000131 | CHEBI:99088 |
| 1 | HP:0000131 | CHEBI:106670 |
| 1 | HP:0000131 | CHEBI:106620 |
| 1 | HP:0000131 | CHEBI:106624 |
| 1 | HP:0000131 | CHEBI:106658 |
| 1 | HP:0000131 | CHEBI:106640 |
| 1 | HP:0000131 | CHEBI:106644 |
| 1 | HP:0000131 | CHEBI:106619 |
| 1 | HP:0000131 | CHEBI:110734 |
| 1 | HP:0000131 | CHEBI:110730 |
| 1 | HP:0000131 | CHEBI:99209 |
| 1 | HP:0000131 | CHEBI:110747 |
| 1 | HP:0000131 | CHEBI:106664 |
| 1 | HP:0000131 | CHEBI:106669 |
| 1 | HP:0000131 | CHEBI:106695 |
| 1 | HP:0000131 | CHEBI:106699 |
| 1 | HP:0000131 | CHEBI:106691 |
| 1 | HP:0000131 | CHEBI:110757 |
| 1 | HP:0000131 | CHEBI:110755 |
| 1 | HP:0000131 | CHEBI:110749 |
| 1 | HP:0000131 | CHEBI:106633 |
| 1 | HP:0000131 | CHEBI:128508 |
| 1 | HP:0000131 | CHEBI:99282 |
| 1 | HP:0000131 | CHEBI:110570 |
| 1 | HP:0000131 | CHEBI:110614 |
| 1 | HP:0000131 | CHEBI:110618 |
| 1 | HP:0000131 | CHEBI:106990 |
| 1 | HP:0000131 | CHEBI:106998 |
| 1 | HP:0000131 | CHEBI:106982 |
| 1 | HP:0000131 | CHEBI:126532 |
| 1 | HP:0000131 | CHEBI:128499 |
| 1 | HP:0000131 | CHEBI:130316 |
| 1 | HP:0000131 | CHEBI:106958 |
| 1 | HP:0000131 | CHEBI:106975 |
| 1 | HP:0000131 | CHEBI:106978 |
| 1 | HP:0000131 | CHEBI:106969 |
| 1 | HP:0000131 | CHEBI:126492 |
| 1 | HP:0000131 | CHEBI:110537 |
| 1 | HP:0000131 | CHEBI:110581 |
| 1 | HP:0000131 | CHEBI:110595 |
| 1 | HP:0000131 | CHEBI:110562 |
| 1 | HP:0000131 | CHEBI:99223 |
| 1 | HP:0000131 | CHEBI:128560 |
| 1 | HP:0000131 | CHEBI:110834 |
| 1 | HP:0000131 | CHEBI:110908 |
| 1 | HP:0000131 | HP:0002640 |
| 1 | HP:0000131 | CHEBI:110968 |
| 1 | HP:0000131 | CHEBI:106787 |
| 1 | HP:0000131 | CHEBI:110970 |
| 1 | HP:0000131 | CHEBI:110979 |
| 1 | HP:0000131 | CHEBI:112519 |
| 1 | HP:0000131 | CHEBI:112511 |
| 1 | HP:0000131 | CHEBI:110901 |
| 1 | HP:0000131 | CHEBI:110848 |
| 1 | HP:0000131 | CHEBI:110713 |
| 1 | HP:0000131 | CHEBI:110844 |
| 1 | HP:0000131 | CHEBI:106815 |
| 1 | HP:0000131 | CHEBI:106809 |
| 1 | HP:0000131 | CHEBI:106803 |
| 1 | HP:0000131 | CHEBI:110876 |
| 1 | HP:0000131 | CHEBI:91381 |
| 1 | HP:0000131 | CHEBI:112590 |
| 1 | HP:0000131 | CHEBI:110814 |
| 1 | HP:0000131 | CHEBI:106791 |
| 1 | HP:0000131 | CHEBI:110995 |
| 1 | HP:0000131 | CHEBI:110984 |
| 1 | HP:0000131 | CHEBI:110935 |
| 1 | HP:0000131 | CHEBI:110707 |
| 1 | HP:0000131 | CHEBI:110703 |
| 1 | HP:0000131 | CHEBI:106756 |
| 1 | HP:0000131 | CHEBI:110775 |
| 1 | HP:0000131 | CHEBI:110948 |
| 1 | HP:0000131 | CHEBI:106778 |
| 1 | HP:0000131 | CHEBI:110958 |
| 1 | HP:0000131 | CHEBI:110769 |
| 1 | HP:0000131 | CHEBI:106719 |
| 1 | HP:0000131 | CHEBI:106710 |
| 1 | HP:0000131 | CHEBI:106715 |
| 1 | HP:0000131 | CHEBI:106701 |
| 1 | HP:0000131 | CHEBI:110924 |
| 1 | HP:0000131 | CHEBI:99117 |
| 1 | HP:0000131 | CHEBI:106732 |
| 1 | HP:0000131 | CHEBI:106735 |
| 1 | HP:0000131 | CHEBI:110799 |
| 1 | HP:0000131 | CHEBI:110974 |
| 1 | HP:0000131 | CHEBI:110965 |
| 1 | HP:0000131 | CHEBI:126805 |
| 1 | HP:0000131 | CHEBI:100294 |
| 1 | HP:0000131 | CHEBI:110823 |
| 1 | HP:0000131 | CHEBI:110826 |
| 1 | HP:0000131 | CHEBI:110831 |
| 1 | HP:0000131 | CHEBI:110801 |
| 1 | HP:0000131 | CHEBI:106988 |
| 1 | HP:0000131 | CHEBI:110809 |
| 1 | HP:0000131 | CHEBI:106984 |
| 1 | HP:0000131 | CHEBI:110811 |
| 1 | HP:0000131 | CHEBI:130112 |
| 1 | HP:0000131 | CHEBI:106939 |
| 1 | HP:0000131 | CHEBI:110843 |
| 1 | HP:0000131 | CHEBI:100010 |
| 1 | HP:0000131 | CHEBI:110850 |
| 1 | HP:0000131 | CHEBI:110859 |
| 1 | HP:0000131 | CHEBI:100030 |
| 1 | HP:0000131 | CHEBI:106924 |
| 1 | HP:0000131 | CHEBI:106920 |
| 1 | HP:0000131 | CHEBI:106938 |
| 1 | HP:0000131 | CHEBI:125296 |
| 1 | HP:0000131 | CHEBI:106950 |
| 1 | HP:0000131 | CHEBI:183858 |
| 1 | HP:0000131 | CHEBI:110893 |
| 1 | HP:0000131 | CHEBI:110860 |
| 1 | HP:0000131 | CHEBI:130107 |
| 1 | HP:0000131 | CHEBI:110810 |
| 1 | HP:0000131 | CHEBI:106637 |
| 1 | HP:0000131 | CHEBI:110838 |
| 1 | HP:0000131 | CHEBI:110836 |
| 1 | HP:0000131 | CHEBI:110829 |
| 1 | HP:0000131 | CHEBI:128993 |
| 1 | HP:0000131 | CHEBI:110898 |
| 1 | HP:0000131 | CHEBI:110892 |
| 1 | HP:0000131 | CHEBI:110890 |
| 1 | HP:0000131 | CHEBI:110886 |
| 1 | HP:0000131 | CHEBI:110881 |
| 1 | HP:0000131 | CHEBI:127751 |
| 1 | HP:0000131 | CHEBI:106977 |
| 1 | HP:0000131 | CHEBI:106974 |
| 1 | HP:0000131 | CHEBI:125302 |
| 1 | HP:0000131 | CHEBI:106931 |
| 1 | HP:0000131 | CHEBI:106906 |
| 1 | HP:0000131 | CHEBI:106655 |
| 1 | HP:0000131 | CHEBI:126898 |
| 1 | HP:0000131 | CHEBI:106893 |
| 1 | HP:0000131 | CHEBI:110721 |
| 1 | HP:0000131 | CHEBI:110727 |
| 1 | HP:0000131 | CHEBI:110732 |
| 1 | HP:0000131 | CHEBI:110736 |
| 1 | HP:0000131 | CHEBI:110739 |
| 1 | HP:0000131 | CHEBI:126852 |
| 1 | HP:0000131 | CHEBI:113322 |
| 1 | HP:0000131 | CHEBI:113316 |
| 1 | HP:0000131 | CHEBI:106918 |
| 1 | HP:0000131 | CHEBI:29584 |
| 1 | HP:0000131 | CHEBI:125828 |
| 1 | HP:0000131 | CHEBI:113312 |
| 1 | HP:0000131 | CHEBI:125800 |
| 1 | HP:0000131 | CHEBI:128998 |
| 1 | HP:0000131 | CHEBI:128999 |
| 1 | HP:0000131 | CHEBI:66256 |
| 1 | HP:0000131 | CHEBI:113342 |
| 1 | HP:0000131 | CHEBI:113267 |
| 1 | HP:0000131 | CHEBI:110745 |
| 1 | HP:0000131 | CHEBI:110741 |
| 1 | HP:0000131 | CHEBI:110740 |
| 1 | HP:0000131 | CHEBI:110786 |
| 1 | HP:0000131 | CHEBI:3654 |
| 1 | HP:0000131 | CHEBI:110794 |
| 1 | HP:0000131 | CHEBI:130121 |
| 1 | HP:0000131 | CHEBI:110797 |
| 1 | HP:0000131 | CHEBI:113286 |
| 1 | HP:0000131 | CHEBI:113274 |
| 1 | HP:0000131 | CHEBI:100307 |
| 1 | HP:0000131 | CHEBI:106840 |
| 1 | HP:0000131 | CHEBI:106856 |
| 1 | HP:0000131 | CHEBI:106853 |
| 1 | HP:0000131 | CHEBI:113295 |
| 1 | HP:0000131 | CHEBI:110705 |
| 1 | HP:0000131 | CHEBI:106839 |
| 1 | HP:0000131 | CHEBI:110710 |
| 1 | HP:0000131 | CHEBI:106889 |
| 1 | HP:0000131 | CHEBI:100372 |
| 1 | HP:0000131 | CHEBI:110868 |
| 1 | HP:0000131 | CHEBI:110871 |
| 1 | HP:0000131 | CHEBI:110963 |
| 1 | HP:0000131 | CHEBI:111037 |
| 1 | HP:0000131 | CHEBI:99651 |
| 1 | HP:0000131 | CHEBI:111094 |
| 1 | HP:0000131 | CHEBI:99677 |
| 1 | HP:0000131 | CHEBI:111098 |
| 1 | HP:0000131 | CHEBI:128882 |
| 1 | HP:0000131 | CHEBI:99693 |
| 1 | HP:0000131 | CHEBI:111004 |
| 1 | HP:0000131 | CHEBI:111009 |
| 1 | HP:0000131 | CHEBI:106817 |
| 1 | HP:0000131 | CHEBI:130217 |
| 1 | HP:0000131 | CHEBI:111047 |
| 1 | HP:0000131 | CHEBI:106819 |
| 1 | HP:0000131 | CHEBI:130259 |
| 1 | HP:0000131 | CHEBI:111011 |
| 1 | HP:0000131 | CHEBI:111017 |
| 1 | HP:0000131 | CHEBI:111021 |
| 1 | HP:0000131 | CHEBI:111027 |
| 1 | HP:0000131 | CHEBI:106799 |
| 1 | HP:0000131 | CHEBI:125921 |
| 1 | HP:0000131 | CHEBI:99668 |
| 1 | HP:0000131 | CHEBI:111057 |
| 1 | HP:0000131 | CHEBI:111090 |
| 1 | HP:0000131 | CHEBI:110990 |
| 1 | HP:0000131 | CHEBI:125709 |
| 1 | HP:0000131 | CHEBI:110980 |
| 1 | HP:0000131 | CHEBI:110938 |
| 1 | HP:0000131 | CHEBI:131280 |
| 1 | HP:0000131 | CHEBI:126059 |
| 1 | HP:0000131 | CHEBI:110950 |
| 1 | HP:0000131 | CHEBI:126080 |
| 1 | HP:0000131 | CHEBI:107010 |
| 1 | HP:0000131 | CHEBI:131269 |
| 1 | HP:0000131 | CHEBI:107007 |
| 1 | HP:0000131 | CHEBI:125976 |
| 1 | HP:0000131 | CHEBI:107027 |
| 1 | HP:0000131 | CHEBI:100004 |
| 1 | HP:0000131 | CHEBI:107014 |
| 1 | HP:0000131 | CHEBI:131217 |
| 1 | HP:0000131 | CHEBI:31526 |
| 1 | HP:0000131 | CHEBI:106795 |
| 1 | HP:0000131 | CHEBI:106727 |
| 1 | HP:0000131 | CHEBI:106642 |
| 1 | HP:0000131 | CHEBI:130089 |
| 1 | HP:0000131 | CHEBI:110776 |
| 1 | HP:0000131 | CHEBI:110773 |
| 1 | HP:0000131 | CHEBI:128912 |
| 1 | HP:0000131 | CHEBI:110796 |
| 1 | HP:0000131 | CHEBI:110792 |
| 1 | HP:0000131 | CHEBI:130000 |
| 1 | HP:0000131 | CHEBI:106685 |
| 1 | HP:0000131 | CHEBI:106666 |
| 1 | HP:0000131 | CHEBI:106605 |
| 1 | HP:0000131 | CHEBI:106720 |
| 1 | HP:0000131 | CHEBI:106602 |
| 1 | HP:0000131 | CHEBI:106616 |
| 1 | HP:0000131 | CHEBI:106612 |
| 1 | HP:0000131 | CHEBI:110856 |
| 1 | HP:0000131 | CHEBI:110853 |
| 1 | HP:0000131 | CHEBI:110849 |
| 1 | HP:0000131 | CHEBI:110845 |
| 1 | HP:0000131 | CHEBI:106649 |
| 1 | HP:0000131 | CHEBI:110779 |
| 1 | HP:0000131 | CHEBI:125847 |
| 1 | HP:0000131 | CHEBI:99538 |
| 1 | HP:0000131 | CHEBI:125816 |
| 1 | HP:0000131 | CHEBI:99599 |
| 1 | HP:0000131 | CHEBI:106734 |
| 1 | HP:0000131 | CHEBI:106708 |
| 1 | HP:0000131 | CHEBI:106767 |
| 1 | HP:0000131 | CHEBI:106762 |
| 1 | HP:0000131 | CHEBI:110731 |
| 1 | HP:0000131 | CHEBI:110720 |
| 1 | HP:0000131 | CHEBI:106775 |
| 1 | HP:0000131 | CHEBI:110758 |
| 1 | HP:0000131 | CHEBI:110751 |
| 1 | HP:0000131 | CHEBI:110748 |
| 1 | HP:0000131 | CHEBI:110744 |
| 1 | HP:0000131 | CHEBI:110717 |
| 1 | HP:0000131 | CHEBI:110714 |
| 1 | HP:0000131 | CHEBI:110708 |
| 1 | HP:0000131 | CHEBI:110704 |
| 1 | HP:0000131 | HP:0040160 |
| 1 | HP:0000131 | HP:0000875 |
| 1 | HP:0000131 | CHEBI:113394 |
| 1 | HP:0000131 | CHEBI:113425 |
| 1 | HP:0000131 | CHEBI:96911 |
| 1 | HP:0000131 | CHEBI:101905 |
| 1 | HP:0000131 | CHEBI:113364 |
| 1 | HP:0000131 | CHEBI:96342 |
| 1 | HP:0000131 | CHEBI:96347 |
| 1 | HP:0000131 | CHEBI:115154 |
| 1 | HP:0000131 | CHEBI:115158 |
| 1 | HP:0000131 | CHEBI:102590 |
| 1 | HP:0000131 | CHEBI:96975 |
| 1 | HP:0000131 | CHEBI:96013 |
| 1 | HP:0000131 | CHEBI:115162 |
| 1 | HP:0000131 | CHEBI:113357 |
| 1 | HP:0000131 | CHEBI:95847 |
| 1 | HP:0000131 | CHEBI:113350 |
| 1 | HP:0000131 | CHEBI:95634 |
| 1 | HP:0000131 | CHEBI:128003 |
| 1 | HP:0000131 | CHEBI:115166 |
| 1 | HP:0000131 | CHEBI:95840 |
| 1 | HP:0000131 | CHEBI:96945 |
| 1 | HP:0000131 | CHEBI:110528 |
| 1 | HP:0000131 | CHEBI:96984 |
| 1 | HP:0000131 | CHEBI:113385 |
| 1 | HP:0000131 | CHEBI:115972 |
| 1 | HP:0000131 | CHEBI:113383 |
| 1 | HP:0000131 | CHEBI:96995 |
| 1 | HP:0000131 | CHEBI:127357 |
| 1 | HP:0000131 | CHEBI:129326 |
| 1 | HP:0000131 | CHEBI:116003 |
| 1 | HP:0000131 | CHEBI:113392 |
| 1 | HP:0000131 | CHEBI:115998 |
| 1 | HP:0000131 | CHEBI:116038 |
| 1 | HP:0000131 | CHEBI:96396 |
| 1 | HP:0000131 | CHEBI:116034 |
| 1 | HP:0000131 | CHEBI:116020 |
| 1 | HP:0000131 | HP:0030035 |
| 1 | HP:0000131 | CHEBI:116058 |
| 1 | HP:0000131 | CHEBI:96906 |
| 1 | HP:0000131 | CHEBI:116052 |
| 1 | HP:0000131 | CHEBI:96301 |
| 1 | HP:0000131 | CHEBI:6375 |
| 1 | HP:0000131 | CHEBI:102571 |
| 1 | HP:0000131 | CHEBI:96356 |
| 1 | HP:0000131 | CHEBI:116019 |
| 1 | HP:0000131 | CHEBI:100346 |
| 1 | HP:0000131 | CHEBI:96990 |
| 1 | HP:0000131 | CHEBI:100967 |
| 1 | HP:0000131 | CHEBI:116016 |
| 1 | HP:0000131 | CHEBI:96320 |
| 1 | HP:0000131 | CHEBI:100992 |
| 1 | HP:0000131 | CHEBI:96962 |
| 1 | HP:0000131 | CHEBI:115317 |
| 1 | HP:0000131 | CHEBI:101924 |
| 1 | HP:0000131 | CHEBI:130525 |
| 1 | HP:0000131 | CHEBI:127882 |
| 1 | HP:0000131 | CHEBI:115979 |
| 1 | HP:0000131 | CHEBI:110592 |
| 1 | HP:0000131 | CHEBI:115775 |
| 1 | HP:0000131 | CHEBI:128357 |
| 1 | HP:0000131 | CHEBI:128359 |
| 1 | HP:0000131 | CHEBI:115781 |
| 1 | HP:0000131 | CHEBI:101132 |
| 1 | HP:0000131 | CHEBI:94856 |
| 1 | HP:0000131 | HP:0012625 |
| 1 | HP:0000131 | CHEBI:96927 |
| 1 | HP:0000131 | CHEBI:115779 |
| 1 | HP:0000131 | HP:0012623 |
| 1 | HP:0000131 | CHEBI:134743 |
| 1 | HP:0000131 | CHEBI:115895 |
| 1 | HP:0000131 | CHEBI:100240 |
| 1 | HP:0000131 | CHEBI:115784 |
| 1 | HP:0000131 | CHEBI:115899 |
| 1 | HP:0000131 | CHEBI:115817 |
| 1 | HP:0000131 | CHEBI:95802 |
| 1 | HP:0000131 | CHEBI:113440 |
| 1 | HP:0000131 | CHEBI:113438 |
| 1 | HP:0000131 | CHEBI:96937 |
| 1 | HP:0000131 | CHEBI:115365 |
| 1 | HP:0000131 | CHEBI:115199 |
| 1 | HP:0000131 | CHEBI:113466 |
| 1 | HP:0000131 | CHEBI:113460 |
| 1 | HP:0000131 | CHEBI:113406 |
| 1 | HP:0000131 | CHEBI:129670 |
| 1 | HP:0000131 | CHEBI:128334 |
| 1 | HP:0000131 | CHEBI:96963 |
| 1 | HP:0000131 | CHEBI:115909 |
| 1 | HP:0000131 | CHEBI:113374 |
| 1 | HP:0000131 | CHEBI:94876 |
| 1 | HP:0000131 | CHEBI:115986 |
| 1 | HP:0000131 | CHEBI:95867 |
| 1 | HP:0000131 | CHEBI:96976 |
| 1 | HP:0000131 | CHEBI:115134 |
| 1 | HP:0000131 | CHEBI:115913 |
| 1 | HP:0000131 | CHEBI:52900 |
| 1 | HP:0000131 | CHEBI:115923 |
| 1 | HP:0000131 | CHEBI:96956 |
| 1 | HP:0000131 | CHEBI:115901 |
| 1 | HP:0000131 | CHEBI:96924 |
| 1 | HP:0000131 | CHEBI:113281 |
| 1 | HP:0000131 | CHEBI:115356 |
| 1 | HP:0000131 | CHEBI:96930 |
| 1 | HP:0000131 | CHEBI:113297 |
| 1 | HP:0000131 | HP:0003574 |
| 1 | HP:0000131 | CHEBI:96955 |
| 1 | HP:0000131 | CHEBI:116072 |
| 1 | HP:0000131 | CHEBI:101149 |
| 1 | HP:0000131 | CHEBI:113277 |
| 1 | HP:0000131 | CHEBI:115141 |
| 1 | HP:0000131 | CHEBI:125305 |
| 1 | HP:0000131 | CHEBI:125300 |
| 1 | HP:0000131 | CHEBI:115902 |
| 1 | HP:0000131 | CHEBI:115924 |
| 1 | HP:0000131 | CHEBI:115918 |
| 1 | HP:0000131 | CHEBI:110617 |
| 1 | HP:0000131 | CHEBI:110624 |
| 1 | HP:0000131 | CHEBI:127964 |
| 1 | HP:0000131 | CHEBI:110638 |
| 1 | HP:0000131 | CHEBI:113344 |
| 1 | HP:0000131 | CHEBI:101593 |
| 1 | HP:0000131 | CHEBI:110602 |
| 1 | HP:0000131 | CHEBI:110606 |
| 1 | HP:0000131 | CHEBI:95972 |
| 1 | HP:0000131 | CHEBI:110612 |
| 1 | HP:0000131 | CHEBI:110616 |
| 1 | HP:0000131 | CHEBI:110613 |
| 1 | HP:0000131 | CHEBI:113314 |
| 1 | HP:0000131 | CHEBI:110627 |
| 1 | HP:0000131 | CHEBI:113315 |
| 1 | HP:0000131 | CHEBI:112648 |
| 1 | HP:0000131 | CHEBI:101300 |
| 1 | HP:0000131 | CHEBI:112644 |
| 1 | HP:0000131 | CHEBI:115391 |
| 1 | HP:0000131 | CHEBI:129425 |
| 1 | HP:0000131 | CHEBI:115993 |
| 1 | HP:0000131 | CHEBI:100913 |
| 1 | HP:0000131 | CHEBI:96825 |
| 1 | HP:0000131 | CHEBI:113325 |
| 1 | HP:0000131 | HP:0004724 |
| 1 | HP:0000131 | CHEBI:115941 |
| 1 | HP:0000131 | CHEBI:110623 |
| 1 | HP:0000131 | CHEBI:101569 |
| 1 | HP:0000131 | CHEBI:95950 |
| 1 | HP:0000131 | CHEBI:128525 |
| 1 | HP:0000131 | CHEBI:115762 |
| 1 | HP:0000131 | CHEBI:113302 |
| 1 | HP:0000131 | CHEBI:128162 |
| 1 | HP:0000131 | CHEBI:113307 |
| 1 | HP:0000131 | CHEBI:115768 |
| 1 | HP:0000131 | CHEBI:115964 |
| 1 | HP:0000131 | CHEBI:110681 |
| 1 | HP:0000131 | CHEBI:110688 |
| 1 | HP:0000131 | CHEBI:127949 |
| 1 | HP:0000131 | CHEBI:115839 |
| 1 | HP:0000131 | CHEBI:130287 |
| 1 | HP:0000131 | CHEBI:115935 |
| 1 | HP:0000131 | CHEBI:127202 |
| 1 | HP:0000131 | CHEBI:101612 |
| 1 | HP:0000131 | CHEBI:101626 |
| 1 | HP:0000131 | CHEBI:113333 |
| 1 | HP:0000131 | CHEBI:130647 |
| 1 | HP:0000131 | CHEBI:96854 |
| 1 | HP:0000131 | CHEBI:110698 |
| 1 | HP:0000131 | CHEBI:96856 |
| 1 | HP:0000131 | CHEBI:101271 |
| 1 | HP:0000131 | CHEBI:113337 |
| 1 | HP:0000131 | CHEBI:115932 |
| 1 | HP:0000131 | CHEBI:113503 |
| 1 | HP:0000131 | CHEBI:115752 |
| 1 | HP:0000131 | CHEBI:110549 |
| 1 | HP:0000131 | CHEBI:113510 |
| 1 | HP:0000131 | CHEBI:96987 |
| 1 | HP:0000131 | CHEBI:127147 |
| 1 | HP:0000131 | CHEBI:93755 |
| 1 | HP:0000131 | CHEBI:129574 |
| 1 | HP:0000131 | CHEBI:100512 |
| 1 | HP:0000131 | CHEBI:128096 |
| 1 | HP:0000131 | CHEBI:100506 |
| 1 | HP:0000131 | CHEBI:101338 |
| 1 | HP:0000131 | CHEBI:101307 |
| 1 | HP:0000131 | CHEBI:129570 |
| 1 | HP:0000131 | CHEBI:113522 |
| 1 | HP:0000131 | CHEBI:129537 |
| 1 | HP:0000131 | CHEBI:115310 |
| 1 | HP:0000131 | CHEBI:129547 |
| 1 | HP:0000131 | CHEBI:115323 |
| 1 | HP:0000131 | CHEBI:115949 |
| 1 | HP:0000131 | CHEBI:115982 |
| 1 | HP:0000131 | CHEBI:96998 |
| 1 | HP:0000131 | CHEBI:115990 |
| 1 | HP:0000131 | CHEBI:115991 |
| 1 | HP:0000131 | CHEBI:100399 |
| 1 | HP:0000131 | CHEBI:100337 |
| 1 | HP:0000131 | CHEBI:110544 |
| 1 | HP:0000131 | CHEBI:96367 |
| 1 | HP:0000131 | CHEBI:115318 |
| 1 | HP:0000131 | CHEBI:115977 |
| 1 | HP:0000131 | CHEBI:96090 |
| 1 | HP:0000131 | CHEBI:127427 |
| 1 | HP:0000131 | CHEBI:128055 |
| 1 | HP:0000131 | CHEBI:94902 |
| 1 | HP:0000131 | CHEBI:96800 |
| 1 | HP:0000131 | CHEBI:96807 |
| 1 | HP:0000131 | CHEBI:115758 |
| 1 | HP:0000131 | CHEBI:129392 |
| 1 | HP:0000131 | CHEBI:101554 |
| 1 | HP:0000131 | CHEBI:101363 |
| 1 | HP:0000131 | CHEBI:127387 |
| 1 | HP:0000131 | CHEBI:96410 |
| 1 | HP:0000131 | CHEBI:129512 |
| 1 | HP:0000131 | CHEBI:115847 |
| 1 | HP:0000131 | CHEBI:127443 |
| 1 | HP:0000131 | CHEBI:129528 |
| 1 | HP:0000131 | CHEBI:112617 |
| 1 | HP:0000131 | CHEBI:96909 |
| 1 | HP:0000131 | CHEBI:127122 |
| 1 | HP:0000131 | CHEBI:94950 |
| 1 | HP:0000131 | CHEBI:96450 |
| 1 | HP:0000131 | CHEBI:127127 |
| 1 | HP:0000131 | CHEBI:115951 |
| 1 | HP:0000131 | CHEBI:101311 |
| 1 | HP:0000131 | CHEBI:115963 |
| 1 | HP:0000131 | CHEBI:96815 |
| 1 | HP:0000131 | CHEBI:101250 |
| 1 | HP:0000131 | CHEBI:113424 |
| 1 | HP:0000131 | CHEBI:112627 |
| 1 | HP:0000131 | CHEBI:101258 |
| 1 | HP:0000131 | CHEBI:100946 |
| 1 | HP:0000131 | CHEBI:95729 |
| 1 | HP:0000131 | CHEBI:101245 |
| 1 | HP:0000131 | CHEBI:115879 |
| 1 | HP:0000131 | CHEBI:95712 |
| 1 | HP:0000131 | CHEBI:111139 |
| 1 | HP:0000131 | CHEBI:95745 |
| 1 | HP:0000131 | CHEBI:101325 |
| 1 | HP:0000131 | CHEBI:115797 |
| 1 | HP:0000131 | CHEBI:95730 |
| 1 | HP:0000131 | CHEBI:100918 |
| 1 | HP:0000131 | CHEBI:96794 |
| 1 | HP:0000131 | CHEBI:96479 |
| 1 | HP:0000131 | CHEBI:115857 |
| 1 | HP:0000131 | CHEBI:111149 |
| 1 | HP:0000131 | CHEBI:101041 |
| 1 | HP:0000131 | CHEBI:129446 |
| 1 | HP:0000131 | CHEBI:129416 |
| 1 | HP:0000131 | CHEBI:115965 |
| 1 | HP:0000131 | CHEBI:95752 |
| 1 | HP:0000131 | CHEBI:115953 |
| 1 | HP:0000131 | CHEBI:115886 |
| 1 | HP:0000131 | CHEBI:96897 |
| 1 | HP:0000131 | CHEBI:96086 |
| 1 | HP:0000131 | CHEBI:96775 |
| 1 | HP:0000131 | CHEBI:107016 |
| 1 | HP:0000131 | CHEBI:115787 |
| 1 | HP:0000131 | CHEBI:96783 |
| 1 | HP:0000131 | CHEBI:95426 |
| 1 | HP:0000131 | CHEBI:95581 |
| 1 | HP:0000131 | CHEBI:115815 |
| 1 | HP:0000131 | CHEBI:100042 |
| 1 | HP:0000131 | CHEBI:101138 |
| 1 | HP:0000131 | CHEBI:111101 |
| 1 | HP:0000131 | CHEBI:111109 |
| 1 | HP:0000131 | CHEBI:115411 |
| 1 | HP:0000131 | CHEBI:95575 |
| 1 | HP:0000131 | CHEBI:101157 |
| 1 | HP:0000131 | CHEBI:115778 |
| 1 | HP:0000131 | CHEBI:115416 |
| 1 | HP:0000131 | CHEBI:107026 |
| 1 | HP:0000131 | CHEBI:115930 |
| 1 | HP:0000131 | CHEBI:96117 |
| 1 | HP:0000131 | CHEBI:96888 |
| 1 | HP:0000131 | CHEBI:115421 |
| 1 | HP:0000131 | CHEBI:111151 |
| 1 | HP:0000131 | CHEBI:115429 |
| 1 | HP:0000131 | CHEBI:115887 |
| 1 | HP:0000131 | CHEBI:115770 |
| 1 | HP:0000131 | CHEBI:115823 |
| 1 | HP:0000131 | CHEBI:115773 |
| 1 | HP:0000131 | CHEBI:125787 |
| 1 | HP:0000131 | CHEBI:125771 |
| 1 | HP:0000131 | CHEBI:102064 |
| 1 | HP:0000131 | CHEBI:128242 |
| 1 | HP:0000131 | CHEBI:101219 |
| 1 | HP:0000131 | CHEBI:96817 |
| 1 | HP:0000131 | CHEBI:115802 |
| 1 | HP:0000131 | CHEBI:129642 |
| 1 | HP:0000131 | CHEBI:115367 |
| 1 | HP:0000131 | CHEBI:115382 |
| 1 | HP:0000131 | CHEBI:115389 |
| 1 | HP:0000131 | CHEBI:96820 |
| 1 | HP:0000131 | CHEBI:127385 |
| 1 | HP:0000131 | CHEBI:96828 |
| 1 | HP:0000131 | CHEBI:115355 |
| 1 | HP:0000131 | CHEBI:96837 |
| 1 | HP:0000131 | CHEBI:127093 |
| 1 | HP:0000131 | CHEBI:111072 |
| 1 | HP:0000131 | CHEBI:96848 |
| 1 | HP:0000131 | CHEBI:111052 |
| 1 | HP:0000131 | CHEBI:115307 |
| 1 | HP:0000131 | CHEBI:115303 |
| 1 | HP:0000131 | CHEBI:111050 |
| 1 | HP:0000131 | CHEBI:111085 |
| 1 | HP:0000131 | CHEBI:111082 |
| 1 | HP:0000131 | CHEBI:127078 |
| 1 | HP:0000131 | CHEBI:111074 |
| 1 | HP:0000131 | CHEBI:128220 |
| 1 | HP:0000131 | CHEBI:129940 |
| 1 | HP:0000131 | CHEBI:115348 |
| 1 | HP:0000131 | CHEBI:96887 |
| 1 | HP:0000131 | CHEBI:115373 |
| 1 | HP:0000131 | CHEBI:100990 |
| 1 | HP:0000131 | CHEBI:96037 |
| 1 | HP:0000131 | CHEBI:111023 |
| 1 | HP:0000131 | CHEBI:129559 |
| 1 | HP:0000131 | CHEBI:111012 |
| 1 | HP:0000131 | CHEBI:127057 |
| 1 | HP:0000131 | CHEBI:115854 |
| 1 | HP:0000131 | CHEBI:101186 |
| 1 | HP:0000131 | CHEBI:111042 |
| 1 | HP:0000131 | CHEBI:115804 |
| 1 | HP:0000131 | CHEBI:102317 |
| 1 | HP:0000131 | CHEBI:96866 |
| 1 | HP:0000131 | CHEBI:125720 |
| 1 | HP:0000131 | CHEBI:111038 |
| 1 | HP:0000131 | CHEBI:115869 |
| 1 | HP:0000131 | CHEBI:111032 |
| 1 | HP:0000131 | CHEBI:96047 |
| 1 | HP:0000131 | CHEBI:96040 |
| 1 | HP:0000131 | CHEBI:96892 |
| 1 | HP:0000131 | CHEBI:111007 |
| 1 | HP:0000131 | CHEBI:129993 |
| 1 | HP:0000131 | CHEBI:115809 |
| 1 | HP:0000131 | CHEBI:96862 |
| 1 | HP:0000131 | CHEBI:96896 |
| 1 | HP:0000131 | CHEBI:115863 |
| 1 | HP:0000131 | CHEBI:96393 |
| 1 | HP:0000131 | CHEBI:115793 |
| 1 | HP:0000131 | CHEBI:129400 |
| 1 | HP:0000131 | CHEBI:107009 |
| 1 | HP:0000131 | CHEBI:115830 |
| 1 | HP:0000131 | CHEBI:101849 |
| 1 | HP:0000131 | CHEBI:115825 |
| 1 | HP:0000131 | CHEBI:102930 |
| 1 | HP:0000131 | CHEBI:96699 |
| 1 | HP:0000131 | CHEBI:115989 |
| 1 | HP:0000131 | CHEBI:127022 |
| 1 | HP:0000131 | CHEBI:127412 |
| 1 | HP:0000131 | CHEBI:129712 |
| 1 | HP:0000131 | CHEBI:115984 |
| 1 | HP:0000131 | CHEBI:96668 |
| 1 | HP:0000131 | CHEBI:102262 |
| 1 | HP:0000131 | CHEBI:129747 |
| 1 | HP:0000131 | CHEBI:129722 |
| 1 | HP:0000131 | CHEBI:115842 |
| 1 | HP:0000131 | CHEBI:115849 |
| 1 | HP:0000131 | CHEBI:130049 |
| 1 | HP:0000131 | CHEBI:101206 |
| 1 | HP:0000131 | CHEBI:115127 |
| 1 | HP:0000131 | CHEBI:115983 |
| 1 | HP:0000131 | CHEBI:96239 |
| 1 | HP:0000131 | CHEBI:115816 |
| 1 | HP:0000131 | CHEBI:100447 |
| 1 | HP:0000131 | CHEBI:100478 |
| 1 | HP:0000131 | CHEBI:115828 |
| 1 | HP:0000131 | CHEBI:127073 |
| 1 | HP:0000131 | CHEBI:113493 |
| 1 | HP:0000131 | CHEBI:115871 |
| 1 | HP:0000131 | CHEBI:115925 |
| 1 | HP:0000131 | CHEBI:113419 |
| 1 | HP:0000131 | CHEBI:113411 |
| 1 | HP:0000131 | CHEBI:101028 |
| 1 | HP:0000131 | CHEBI:101286 |
| 1 | HP:0000131 | CHEBI:100266 |
| 1 | HP:0000131 | HP:0000976 |
| 1 | HP:0000131 | CHEBI:115891 |
| 1 | HP:0000131 | CHEBI:96017 |
| 1 | HP:0000131 | CHEBI:101268 |
| 1 | HP:0000131 | CHEBI:115903 |
| 1 | HP:0000131 | CHEBI:115896 |
| 1 | HP:0000131 | CHEBI:115898 |
| 1 | HP:0000131 | CHEBI:113499 |
| 1 | HP:0000131 | CHEBI:115181 |
| 1 | HP:0000131 | CHEBI:115187 |
| 1 | HP:0000131 | CHEBI:115915 |
| 1 | HP:0000131 | CHEBI:96297 |
| 1 | HP:0000131 | CHEBI:96009 |
| 1 | HP:0000131 | CHEBI:113489 |
| 1 | HP:0000131 | CHEBI:102621 |
| 1 | HP:0000131 | CHEBI:100226 |
| 1 | HP:0000131 | CHEBI:115820 |
| 1 | HP:0000131 | CHEBI:113475 |
| 1 | HP:0000131 | CHEBI:101294 |
| 1 | HP:0000131 | CHEBI:113501 |
| 1 | HP:0000131 | CHEBI:115877 |
| 1 | HP:0000131 | CHEBI:95411 |
| 1 | HP:0000131 | CHEBI:96738 |
| 1 | HP:0000131 | CHEBI:115803 |
| 1 | HP:0000131 | CHEBI:96749 |
| 1 | HP:0000131 | CHEBI:115837 |
| 1 | HP:0000131 | CHEBI:127453 |
| 1 | HP:0000131 | CHEBI:96754 |
| 1 | HP:0000131 | CHEBI:96078 |
| 1 | HP:0000131 | CHEBI:102038 |
| 1 | HP:0000131 | CHEBI:115840 |
| 1 | HP:0000131 | CHEBI:115845 |
| 1 | HP:0000131 | CHEBI:102222 |
| 1 | HP:0000131 | CHEBI:95396 |
| 1 | HP:0000131 | CHEBI:101353 |
| 1 | HP:0000131 | CHEBI:102978 |
| 1 | HP:0000131 | CHEBI:96193 |
| 1 | HP:0000131 | CHEBI:96070 |
| 1 | HP:0000131 | CHEBI:111118 |
| 1 | HP:0000131 | CHEBI:115994 |
| 1 | HP:0000131 | CHEBI:101195 |
| 1 | HP:0000131 | CHEBI:111121 |
| 1 | HP:0000131 | CHEBI:111127 |
| 1 | HP:0000131 | CHEBI:115432 |
| 1 | HP:0000131 | CHEBI:102958 |
| 1 | HP:0000131 | CHEBI:107001 |
| 1 | HP:0000131 | CHEBI:115436 |
| 1 | HP:0000131 | CHEBI:96120 |
| 1 | HP:0000131 | CHEBI:96134 |
| 1 | HP:0000131 | CHEBI:101496 |
| 1 | HP:0000131 | CHEBI:129302 |
| 1 | HP:0000131 | CHEBI:129371 |
| 1 | HP:0000131 | CHEBI:115878 |
| 1 | HP:0000131 | CHEBI:129761 |
| 1 | HP:0000131 | CHEBI:113520 |
| 1 | HP:0000131 | CHEBI:101174 |
| 1 | HP:0000131 | CHEBI:113516 |
| 1 | HP:0000131 | CHEBI:115976 |
| 1 | HP:0000131 | CHEBI:95681 |
| 1 | HP:0000131 | CHEBI:96657 |
| 1 | HP:0000131 | CHEBI:95327 |
| 1 | HP:0000131 | CHEBI:101504 |
| 1 | HP:0000131 | CHEBI:100088 |
| 1 | HP:0000131 | CHEBI:130004 |
| 1 | HP:0000131 | CHEBI:96280 |
| 1 | HP:0000131 | CHEBI:115851 |
| 1 | HP:0000131 | CHEBI:115852 |
| 1 | HP:0000131 | CHEBI:115794 |
| 1 | HP:0000131 | CHEBI:115858 |
| 1 | HP:0000131 | CHEBI:96289 |
| 1 | HP:0000131 | CHEBI:115861 |
| 1 | HP:0000131 | CHEBI:96261 |
| 1 | HP:0000131 | CHEBI:102446 |
| 1 | HP:0000131 | CHEBI:101519 |
| 1 | HP:0000131 | CHEBI:96275 |
| 1 | HP:0000131 | CHEBI:112629 |
| 1 | HP:0000131 | CHEBI:116009 |
| 1 | HP:0000131 | CHEBI:95697 |
| 1 | HP:0000131 | CHEBI:115299 |
| 1 | HP:0000131 | CHEBI:100891 |
| 1 | HP:0000131 | CHEBI:102714 |
| 1 | HP:0000131 | CHEBI:96860 |
| 1 | HP:0000131 | CHEBI:112542 |
| 1 | HP:0000131 | CHEBI:115208 |
| 1 | HP:0000131 | CHEBI:112549 |
| 1 | HP:0000131 | CHEBI:94851 |
| 1 | HP:0000131 | CHEBI:112552 |
| 1 | HP:0000131 | CHEBI:100839 |
| 1 | HP:0000131 | CHEBI:101283 |
| 1 | HP:0000131 | HP:0025081 |
| 1 | HP:0000131 | CHEBI:129458 |
| 1 | HP:0000131 | CHEBI:96191 |
| 1 | HP:0000131 | CHEBI:96868 |
| 1 | HP:0000131 | CHEBI:96863 |
| 1 | HP:0000131 | CHEBI:100813 |
| 1 | HP:0000131 | CHEBI:127611 |
| 1 | HP:0000131 | CHEBI:112584 |
| 1 | HP:0000131 | CHEBI:128643 |
| 1 | HP:0000131 | CHEBI:101023 |
| 1 | HP:0000131 | CHEBI:95691 |
| 1 | HP:0000131 | CHEBI:101749 |
| 1 | HP:0000131 | CHEBI:101734 |
| 1 | HP:0000131 | CHEBI:101014 |
| 1 | HP:0000131 | CHEBI:113447 |
| 1 | HP:0000131 | CHEBI:113441 |
| 1 | HP:0000131 | CHEBI:113477 |
| 1 | HP:0000131 | CHEBI:110640 |
| 1 | HP:0000131 | CHEBI:113483 |
| 1 | HP:0000131 | CHEBI:129437 |
| 1 | HP:0000131 | CHEBI:129862 |
| 1 | HP:0000131 | CHEBI:115831 |
| 1 | HP:0000131 | CHEBI:96872 |
| 1 | HP:0000131 | CHEBI:115236 |
| 1 | HP:0000131 | CHEBI:110656 |
| 1 | HP:0000131 | CHEBI:127251 |
| 1 | HP:0000131 | CHEBI:115230 |
| 1 | HP:0000131 | CHEBI:113410 |
| 1 | HP:0000131 | CHEBI:112563 |
| 1 | HP:0000131 | CHEBI:127272 |
| 1 | HP:0000131 | CHEBI:128178 |
| 1 | HP:0000131 | CHEBI:96149 |
| 1 | HP:0000131 | CHEBI:113427 |
| 1 | HP:0000131 | CHEBI:129466 |
| 1 | HP:0000131 | CHEBI:96577 |
| 1 | HP:0000131 | CHEBI:128171 |
| 1 | HP:0000131 | CHEBI:113400 |
| 1 | HP:0000131 | CHEBI:113452 |
| 1 | HP:0000131 | CHEBI:110662 |
| 1 | HP:0000131 | CHEBI:113468 |
| 1 | HP:0000131 | CHEBI:113432 |
| 1 | HP:0000131 | CHEBI:112577 |
| 1 | HP:0000131 | CHEBI:127695 |
| 1 | HP:0000131 | CHEBI:96492 |
| 1 | HP:0000131 | CHEBI:112593 |
| 1 | HP:0000131 | CHEBI:113360 |
| 1 | HP:0000131 | CHEBI:96974 |
| 1 | HP:0000131 | CHEBI:127644 |
| 1 | HP:0000131 | CHEBI:96969 |
| 1 | HP:0000131 | CHEBI:129898 |
| 1 | HP:0000131 | CHEBI:127996 |
| 1 | HP:0000131 | CHEBI:5778 |
| 1 | HP:0000131 | CHEBI:96982 |
| 1 | HP:0000131 | CHEBI:130172 |
| 1 | HP:0000131 | CHEBI:102812 |
| 1 | HP:0000131 | CHEBI:96915 |
| 1 | HP:0000131 | CHEBI:115287 |
| 1 | HP:0000131 | CHEBI:100710 |
| 1 | HP:0000131 | CHEBI:112597 |
| 1 | HP:0000131 | CHEBI:115275 |
| 1 | HP:0000131 | CHEBI:115280 |
| 1 | HP:0000131 | CHEBI:100641 |
| 1 | HP:0000131 | HP:0005681 |
| 1 | HP:0000131 | CHEBI:102375 |
| 1 | HP:0000131 | CHEBI:115766 |
| 1 | HP:0000131 | CHEBI:101389 |
| 1 | HP:0000131 | CHEBI:100736 |
| 1 | HP:0000131 | CHEBI:115755 |
| 1 | HP:0000131 | HP:0001413 |
| 1 | HP:0000131 | CHEBI:100780 |
| 1 | HP:0000131 | CHEBI:115291 |
| 1 | HP:0000131 | CHEBI:115253 |
| 1 | HP:0000131 | CHEBI:112518 |
| 1 | HP:0000131 | CHEBI:96940 |
| 1 | HP:0000131 | CHEBI:129075 |
| 1 | HP:0000131 | CHEBI:115409 |
| 1 | HP:0000131 | CHEBI:128683 |
| 1 | HP:0000131 | CHEBI:101635 |
| 1 | HP:0000131 | CHEBI:100637 |
| 1 | HP:0000131 | CHEBI:127301 |
| 1 | HP:0000131 | CHEBI:115404 |
| 1 | HP:0000131 | CHEBI:101705 |
| 1 | HP:0000131 | CHEBI:129021 |
| 1 | HP:0000131 | CHEBI:95306 |
| 1 | HP:0000131 | CHEBI:130137 |
| 1 | HP:0000131 | CHEBI:94887 |
| 1 | HP:0000131 | CHEBI:96935 |
| 1 | HP:0000131 | CHEBI:115257 |
| 1 | HP:0000131 | CHEBI:127655 |
| 1 | HP:0000131 | CHEBI:96922 |
| 1 | HP:0000131 | CHEBI:94895 |
| 1 | HP:0000131 | CHEBI:96954 |
| 1 | HP:0000131 | CHEBI:34669 |
| 1 | HP:0000131 | CHEBI:113376 |
| 1 | HP:0000131 | CHEBI:128714 |
| 1 | HP:0000131 | CHEBI:100667 |
| 1 | HP:0000131 | CHEBI:96949 |
| 1 | HP:0000131 | CHEBI:101935 |
| 1 | HP:0000131 | CHEBI:115311 |
| 1 | HP:0000131 | CHEBI:115248 |
| 1 | HP:0000131 | CHEBI:113471 |
| 1 | HP:0000131 | CHEBI:113495 |
| 1 | HP:0000131 | CHEBI:115956 |
| 1 | HP:0000131 | CHEBI:115757 |
| 1 | HP:0000131 | CHEBI:102467 |
| 1 | HP:0000131 | CHEBI:112639 |
| 1 | HP:0000131 | CHEBI:115761 |
| 1 | HP:0000131 | CHEBI:94756 |
| 1 | HP:0000131 | CHEBI:127498 |
| 1 | HP:0000131 | CHEBI:101110 |
| 1 | HP:0000131 | CHEBI:96845 |
